# Indigenous, integrative, and biomedical perimenopause and postmenopause healthcare for Indigenous midlife peoples residing in the United States and Canada: A scoping review

**DOI:** 10.1101/2025.07.10.25331221

**Authors:** Jacqueline Kent-Marvick, Lisa Taylor-Swanson, Lydia A. Howes, Po-Yu Hsu, Sharon Austin, Tumilara Aderibigbe, Leslie Crandall, Madison Elizabeth Maughan, Jessica Ann Ellis, Stephanie Quist, Sarah Rose Robertson, Lorna Khemchand, Mary M. McFarland, Sara E. Simonsen

## Abstract

**Background:** Little is known about the menopause-related healthcare of Indigenous people of the United States and Canada. This scoping review mapped the existing literature on Indigenous, integrative, and biomedical healthcare for U.S.- and Canada-based perimenopausal Indigenous populations.

**Methods:** We followed the JBI Manual for Evidence Synthesis. The protocol was registered on Open Science Framework. Eligibility criteria included records focused on Indigenous people of the U.S. and Canada perimenopause/postmenopause healthcare. Medline and eight other data bases were searched on June 1, 2023, and updated November 21, 2024. Covidence was used for selecting and extracting data.

**Findings:** After screening 6,684 records, 45 met inclusion criteria. These focused on Indigenous people’s experiences, care barriers, symptom reporting, and treatment preferences. Traditionality, community roles, and cultural beliefs influenced symptom experience and healthcare-seeking. Many participants preferred Indigenous or integrative interventions, sometimes in combination with biomedical care. Barriers included rushed encounters, language and cultural disconnects, and systemic inequities.

**Interpretation:** Findings highlight the need for culturally responsive, patient-centered care during the menopausal transition with an emphasis on evidence-based interventions including Indigenous and integrative health approaches. Additional research, stronger community engagement, and healthcare-provider training will help improve perimenopause health outcomes for Indigenous populations in the United States and Canada

**Funding:** The authors disclose receipt of the following financial support for the research, authorship, and/or publication of this article. The first author received support from the National Clinician Scholars Program (NCSP) as a postdoctoral fellow at the University of Pennsylvania. Additionally, she was supported by the National Institute of Nursing Research of the National Institutes of Health under Award Number F31NR020431. The content is solely the responsibility of the authors and does not necessarily represent the official views of the National Institutes of Health or the Indian Health Service. The authors were supported by a grant from the University of Utah office of the Vice President for Research.

**Panel: Research in Context:** *Evidence before this study:* Before undertaking this scoping review, the review team identified several sentinel articles. No prior evidence reviews nor protocols on our topic were identified in searches conducted on February 22, 2023 in PROSPERO, Open Science Framework, PubMed, Cochrane Library, Campbell Systematic Reviews, CINAHL, and Sociological Abstracts. We then conducted comprehensive searches across nine databases—Medline (Ovid) 1946-2024, Embase (Elsevier) 1974-2024, CINAHL Complete (EBSCOhost) 1937-2024, Ageline (EBSCOhost) 1966-2024, Alt HealthWatch (EBSCOhost) 1984-2024, APA PsycInfo (EBSCOhost) 1872-2024, Scopus (Elsevier) 1977-2024, Web of Science Core Collection (Clarivate) 1900-2024, and the Native Health Database (1900-2024). Searches were initially conducted on June 1, 2023, and repeated on November 21, 2024. We also reviewed reference lists of included studies. Search strategies combined keywords and subject headings related to menopause, healthcare, and Indigenous populations. Only English-language records were included due to funding constraints. We excluded books and cancer-related studies to focus on natural or surgical menopause not induced by chemotherapy. No quality assessment of included studies was conducted in accordance with scoping-review methodology.

*Added value of this study:* This is the first known scoping review to synthesize literature on perimenopause and postmenopause healthcare among Indigenous peoples in the United States and Canada. It maps Indigenous, integrative, and biomedical treatment practices; highlights structural and cultural barriers to care; and identifies the influence of traditional knowledge, community roles, and colonization on symptom interpretation and treatment-seeking. It also summarizes patient-centered recommendations for improving perimenopause healthcare delivery, and outlines key research gaps.

*Implications of all the available evidence:* Findings from this review, combined with existing literature, emphasize the urgent need for culturally responsive, community-informed healthcare for Indigenous midlife populations. Interventions should consider traditional knowledge systems, language and literacy needs, and preferences for Indigenous and integrative treatments. Healthcare providers must engage in shared decision-making and address systemic barriers to care. Future research should center Indigenous perspectives, investigate disparities in symptom experiences, and help to improve access to perimenopause healthcare for Indigenous peoples.

## INTRODUCTION

American Indian and Alaska Native (AI/AN) females in the United States (U.S.) have a lower life expectancy than any other racial or ethnic group of females, living an average of 75 years— six years fewer than their non-Hispanic White conterparts.^1^ Similarly, Indigenous females in Canada (i.e., First Nations, Métis, and Inuit) have shorter life expectancies than non-Indigenous females: 77·7 years for First Nations, 82·3 for Métis, and 76·1 for Inuit, compared to 87·3 years for non-Indigenous females.^2^

Indigenous people in both nations face disproportionately high rates of chronic disease and health-risk factors.^3,4^ For example, diabetes, heart disease, and cancer prevalence—and associated mortality—are higher in these populations, with AI/AN and Indigenous females experiencing the lowest cancer-survival rates of any racial or ethnic group in the U.S. and Canada.^5–9^ One contributing factor to these disparities is limited access to and use of preventive healthcare and screenings in midlife.^10–14^

Midlife Indigenous females (aged 40 to 55) are often regarded as the backbone of their families and communities. As part of the “sandwich generation,” they frequently care for children or grandchildren and aging parents while also working outside the home.^15^ Although traditionally revered as “givers of life,” their health is often deprioritized due to time constraints, resulting in missed annual checkups and screenings for conditions such as diabetes, heart disease, and cancer.^10–13^

During the menopausal transition (MT), the few studies that include AI/AN females indicate that this population is among the most likely to report vasomotor symptoms (VMS) compared to other racial and ethnic groups.^16^ Emerging evidence indicate that VMS may serve as a biomarker for chronic conditions,^17,18^ including cardiovascular disease. These findings underscore the urgency of improving access to midlife MT-related care and preventive care, as Indigenous females are less likely than non-Hispanic White females to receive regular screenings such as cervical cancer tests and mammograms,^11,13^ and face unique barriers to doing so.

To address gaps in knowledge about perimenopause and postmenopause care for AI/AN and Indigenous females (herein collectively referred to as Indigenous), we conducted a scoping review to examine the breadth and nature of existing literature on Indigenous, integrative, and biomedical healthcare in the U.S. and Canada. A scoping review was selected to map the range of available evidence, identify key themes and knowledge gaps, and clarify how this body of literature addressed the healthcare needs of Indigenous midlife populations.

### Objective

This scoping review aimed to map the current literature on Indigenous, integrative, and biomedical perimenopause and postmenopause healthcare among Indigenous peoples residing in the U.S. and Canada. This review addresses a critical gap in the literature and will inform recommendations for healthcare settings and future research.

Our guiding research question is: *What is the research or evidence about perimenopause and postmenopause Indigenous, integrative, and biomedical healthcare among Indigenous peoples of North America residing in the United States or Canada?*

## METHODS

The protocol for this scoping review was published on Open Science Framework (osf.io/hwna5). We conducted our scoping review with guidance from the JBI Manual for Evidence Synthesis.^19^ We followed the Arksey and O’Malley framework, which outlines five stages of scoping-review methodology: 1) identifying the research question, 2) identifying relevant studies, 3) study selection, 4) charting the data, and 5) collating, summarizing, and reporting the results.^20^ For transparency and reproducibility, we’ve adhered to the PRISMA-ScR reporting guidelines for scoping reviews and searches.^21,22^

We used Covidence (Veritas Health Innovation),^23^ an online systematic reviewing platform, to screen records, select records, and chart data. Covidence uses active learning to prioritize records most likely to be included, displaying these first during screening. Citation management and duplicate detection and removal were accomplished with EndNote 20 Version (Clarivate Analytics).^24^

### Search strategy and selection criteria

A University of Utah librarian (LAH) developed the search strategy for the primary database (Medline) using a combination of keywords and subject headings from sentinel studies,^14,16,25–27^ team feedback, and U.S. and Canadian government resources.^28,29^ Search terms included variations of menopause, vasomotor symptoms, Indigenous Americans, and First Nations. A library colleague (MMM) peer-reviewed the strategy according to PRESS guidelines to ensure an adequate evidence base.^30^ The primary strategy was then translated to use in other databases.^30^

The following databases were searched: Medline (Ovid) 1946–2024, Embase (Elsevier) 1974– 2024, CINAHL Complete (Ebscohost) 1937–2024, Ageline (Ebscohost) 1966–2024, Alt HealthWatch (Ebscohost) 1984–2024, APA PsycInfo (Ebscohost) 1872–2024, Scopus (Elsevier) 1970–2024, and Web of Science Core Collection (Clarivate) 1900–2024.

Searches were initially conducted between June 1 and 9, 2023 and then updated on November 21, 2024. No date limits were applied other than database inception or institutional license dates. In CINAHL, the publication types of “Book Review,” “Brief Item,” “Obituary,” or “Poetry” were excluded. Book reviews were excluded in Web of Science Core Collection. Otherwise, no search filters or limiters were applied to initial searches. Searches were updated on November 21, 2024, and limited to publication date or database entry date after the initial searches in June 2023. Databases on the Ebscohost platform were searched independently rather than simultaneously. The search strategies in Appendix A contain the full database searches.

Reference lists of included studies were also screened (JK-M, SA, P-YH, TA) to identify additional relevant studies not found with our database searches, and these were added to Covidence for screening and selection. In a deviation from the protocol, the team did not search for grey literature.

One reviewer (JK-M) independently screened all titles and abstracts. Each record was also independently reviewed by a second team member from a pool of ten (SES, SA, LT-S, SQ, SRR, LK, JE, TA, MEM, P-YH). This process was repeated for full-text review. Any discrepancies between reviewers were discussed during team meetings and resolved by consensus.

### Inclusion criteria

We used the Population, Concept, and Context (PCC) framework.^19^ The population included AI/AN and Canadian Indigenous individuals experiencing perimenopause or postmenopause. Please see the search strategies in Appendix A to view a list of all populations included under the broader categories of AI/AN and Canadian Indigenous.

The concept focused on both menopause and healthcare (including the provision of care and access to care). Articles were included if they addressed aspects of menopause—such as the MT or VMS—and any form of healthcare. This included Indigenous medicine (treatments traditionally used by Indigenous peoples), integrative approaches (e.g., herbal treatments, acupressure, acupuncture), and biomedical care (also referred to as allopathic medicine or conventional medicine). We use menopause-related definitions outlined by Harlow et al. (2012) in their updated Stages of Reproductive Aging Workshop +10 (STRAW+10) guidance.^31^ Perimenopause refers to the time surrounding menopause, and includes both the early and late stages of MT, as well as the first year following the final menstrual period.^31^ During the early MT, individuals experience hormonal fluctuations and changes in menstrual-cycle length— specifically, a variation of seven days or more between cycles within a span of 10 consecutive cycles. The late MT is characterized by more pronounced hormonal fluctuations, greater variability in cycle length, more frequent anovulation (i.e., absence of ovulation), and periods of amenorrhea lasting 60 days or longer.^31^ Postmenopause begins after 12 consecutive months of amenorrhea and continues for the remainder of the person’s life.^31^

The context was limited to U.S.- or Canada-based studies. We selected the U.S. and Canada as the context for this review because both are developed nations with comparable free-market economies, yet they differ significantly in the structure of their healthcare systems. Canada has a publicly funded, universal healthcare system, while the U.S. relies on a mixed model of public and private insurance. Examining studies from these two countries allows for an exploration of how structural differences in healthcare access and delivery may shape the perimenopause and postmenopause experiences of Indigenous peoples.

All types of sources were considered. Study types are listed in Table 1.

**Table 1:**
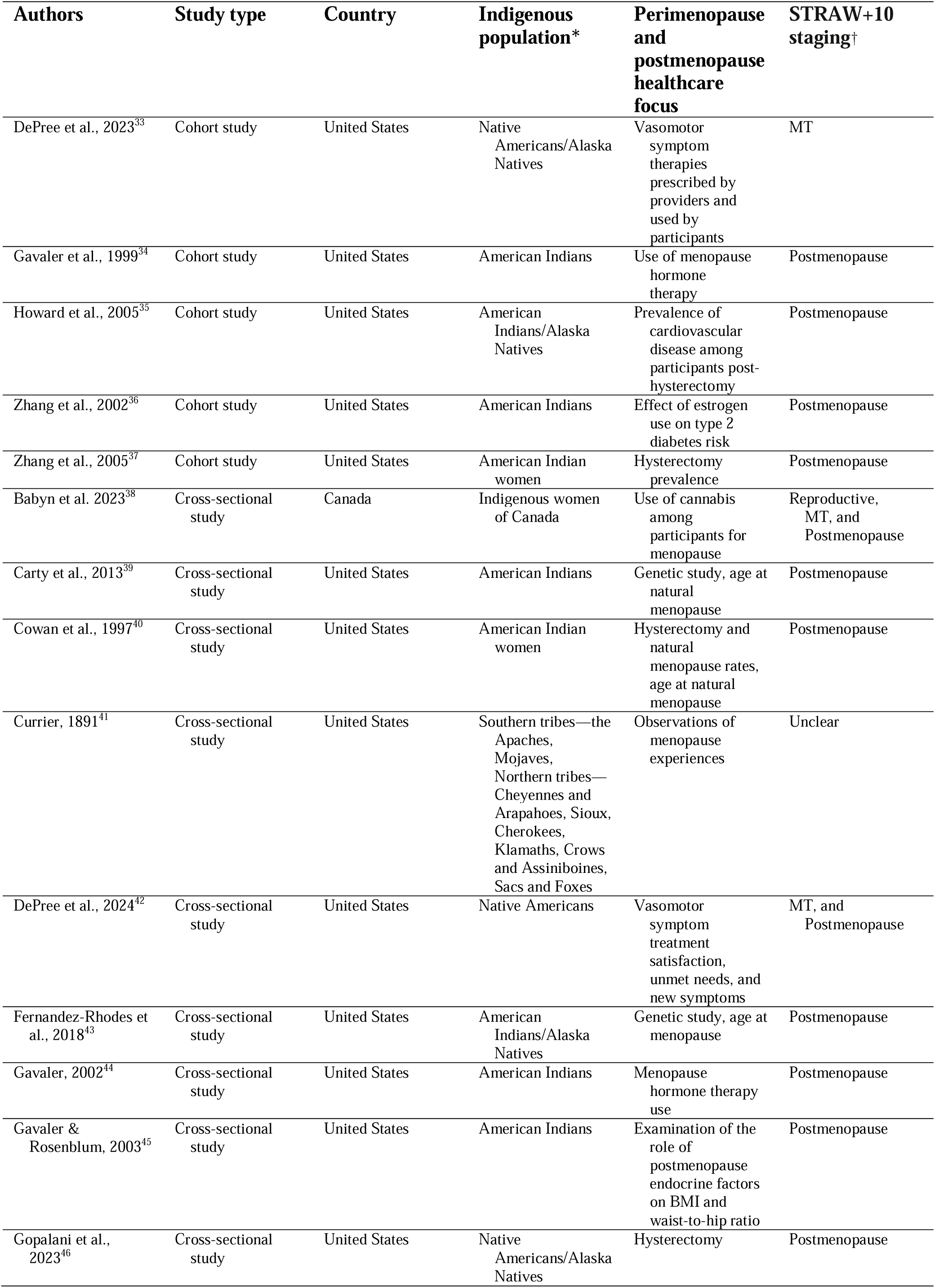
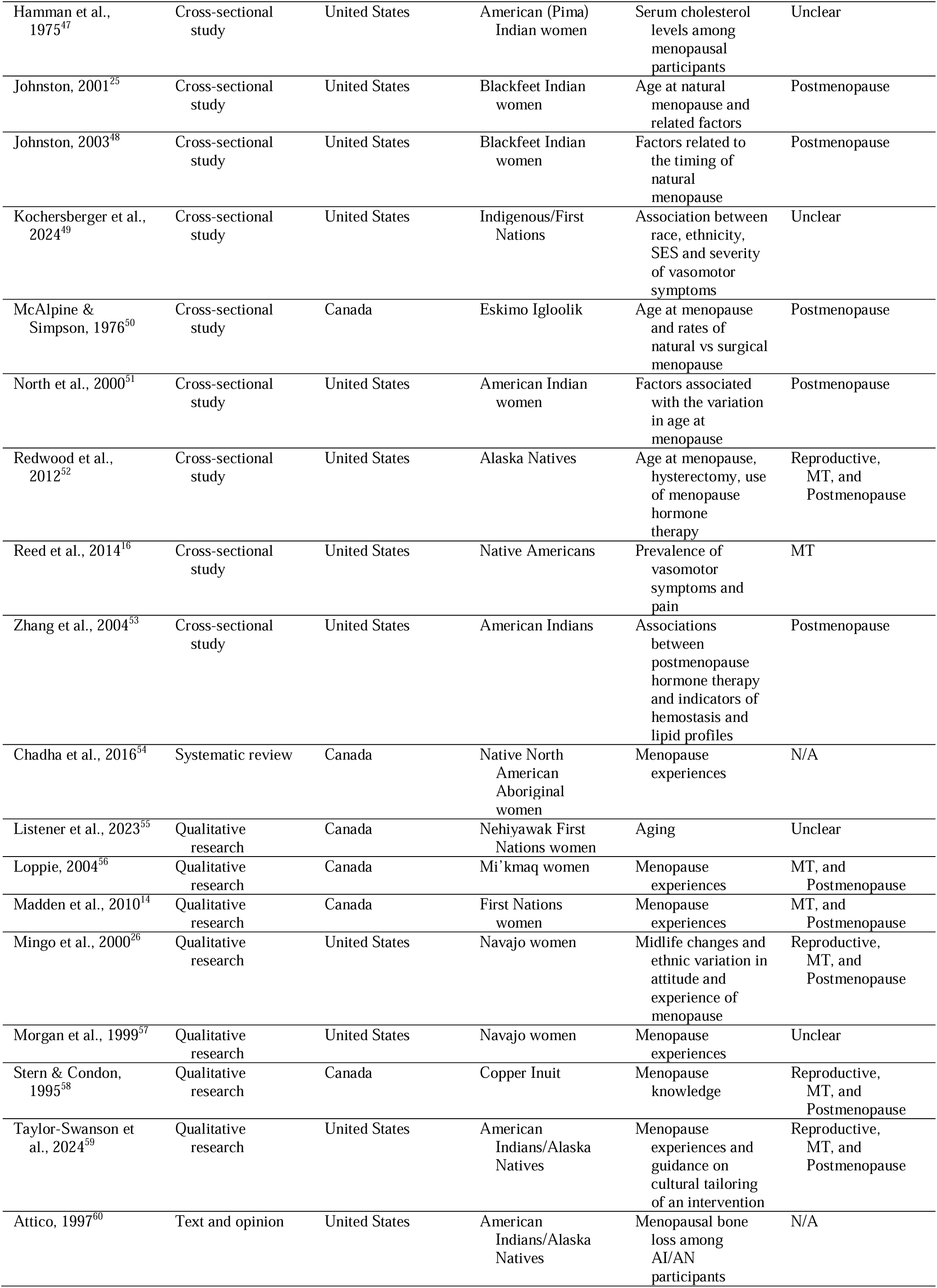
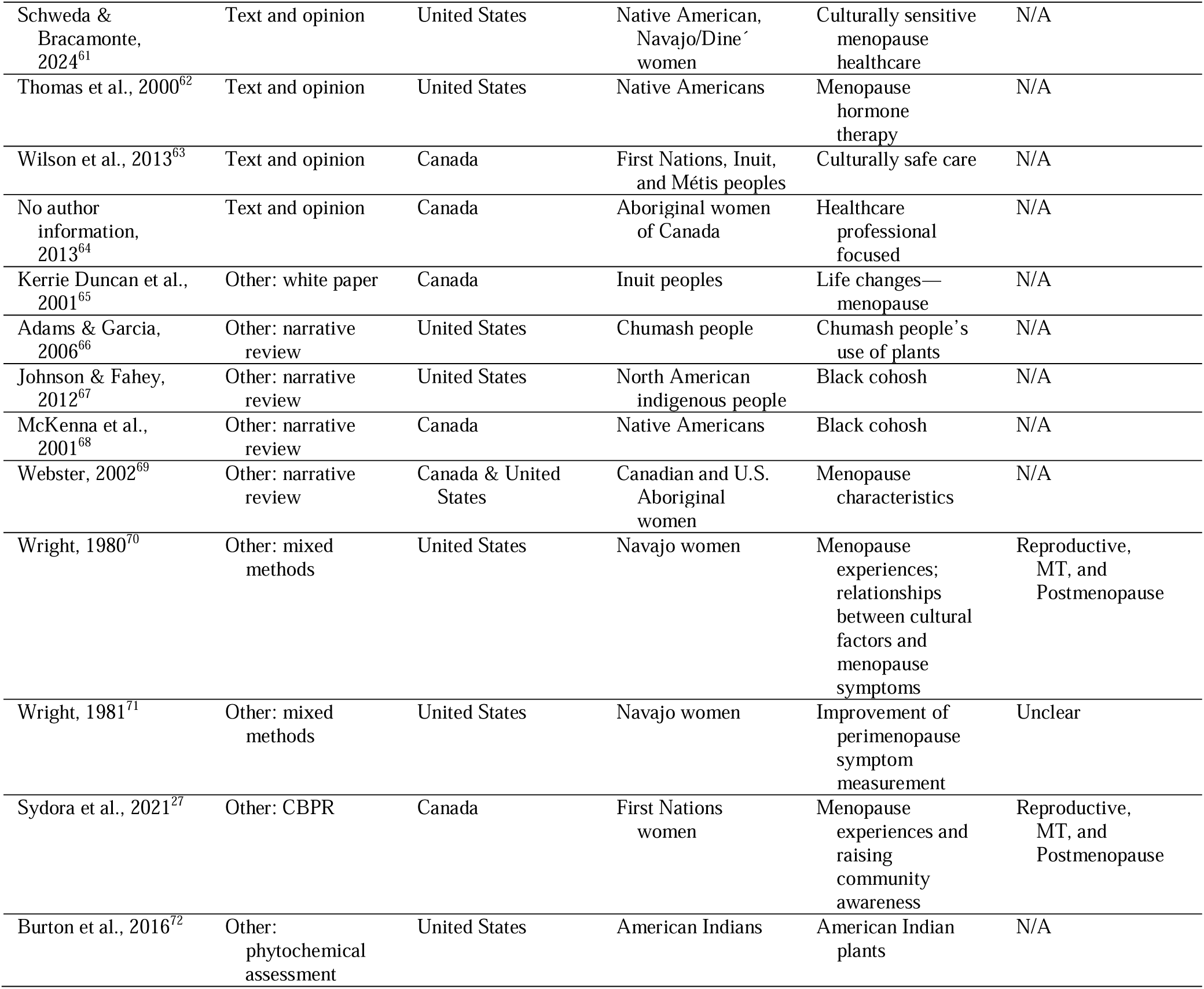
Characteristics of the sources of evidence. *The description of Indigenous populations reflects the terminology used in original records. †No records included in this scoping review formally applied STRAW+10 staging criteria. The authors of this scoping review applied the STRAW+10 criteria to sample descriptions retrospectively to estimate participants’ stage of reproductive aging.

### Exclusion criteria

We excluded studies related to cancer, as VMS in these contexts may be treatment-induced (e.g., from chemotherapy). Records not available in English were excluded at the full-text review stage. We listed “non-English record” as the reason for exclusion. The rationale for not including non-English records was that this project lacked funding for translation services. Books were excluded at full-text review for time feasibility and funding, a difference from the protocol.

### Quality assessment

Consistent with JBI guidance for scoping reviews—which aim to map the breadth of existing literature, we did not assess the methodological quality of included literature.^19^

### Data extraction

The lead author (JK-M) developed the extraction form, which was piloted by other team members (SES, SA, LT-S, JAE) using sentinel articles prior to protocol finalization. We customized Covidence’s data-charting Extraction 2 form to the PCC framework, capturing the following elements: title, authors, country, study aims, design, conflicts of interest, inclusion and exclusion criteria, population and sampling details, Indigenous sample description, total sample size (and number of Indigenous participants), findings relevant to the review aim, use of the STRAW+10 criteria,^31^ menopausal-transition staging using STRAW+10, author-reported barriers to and facilitators of treatment, and study limitations.

Two reviewers from a pool of five reviewers (JK-M, SA, P-YH, TA, MEM) independently extracted and charted data from the included literature (i.e., two reviewers independently extracted and charted data for each included record). Conflicts in data extraction were addressed and resolved at team meetings.

### Synthesis of results

In addition to narrative synthesis of results organized by PCC categories and thematic groupings, we used numerical counts and frequencies. Key findings are presented in tables and figures.

Author-reported barriers and facilitators to treatment, as well as rural versus urban differences are summarized in narrative form. Barriers and facilitators are also visually represented using an Indian Health Service-adapted version of the socioecological model.^31^ Based on statements from the included literature, we offer recommendations to improve healthcare settings and identify directions for future research in the Discussion section of this review. No funder had a role in the review’s design, data collection, data synthesis, data interpretation, or writing of the report.

Please note that terminology in this review reflects both the language used in the cited literature and a commitment to inclusivity. For the most part, “female” is used in referring to the sex-based findings reported in studies, while broader references use inclusive terms such as “perimenopausal people” to acknowledge gender diversity. When reporting the findings from specific records, we have applied the sample descriptors used by the authors of those records.

When discussing our review’s population more broadly, we use the term “Indigenous.”

Additionally, this academic team includes a member from an American Indian tribe from the U.S. southwest, as well as healthcare providers who have collaborated closely with American Indian and Alaska Native communities.

## RESULTS

A total of 6,684 records were identified through database and reference searches. After full-text screening, 45 records met the final inclusion criteria. The full selection process is detailed in the PRISMA flow diagram (Figure 1).

**Fig. 1.**
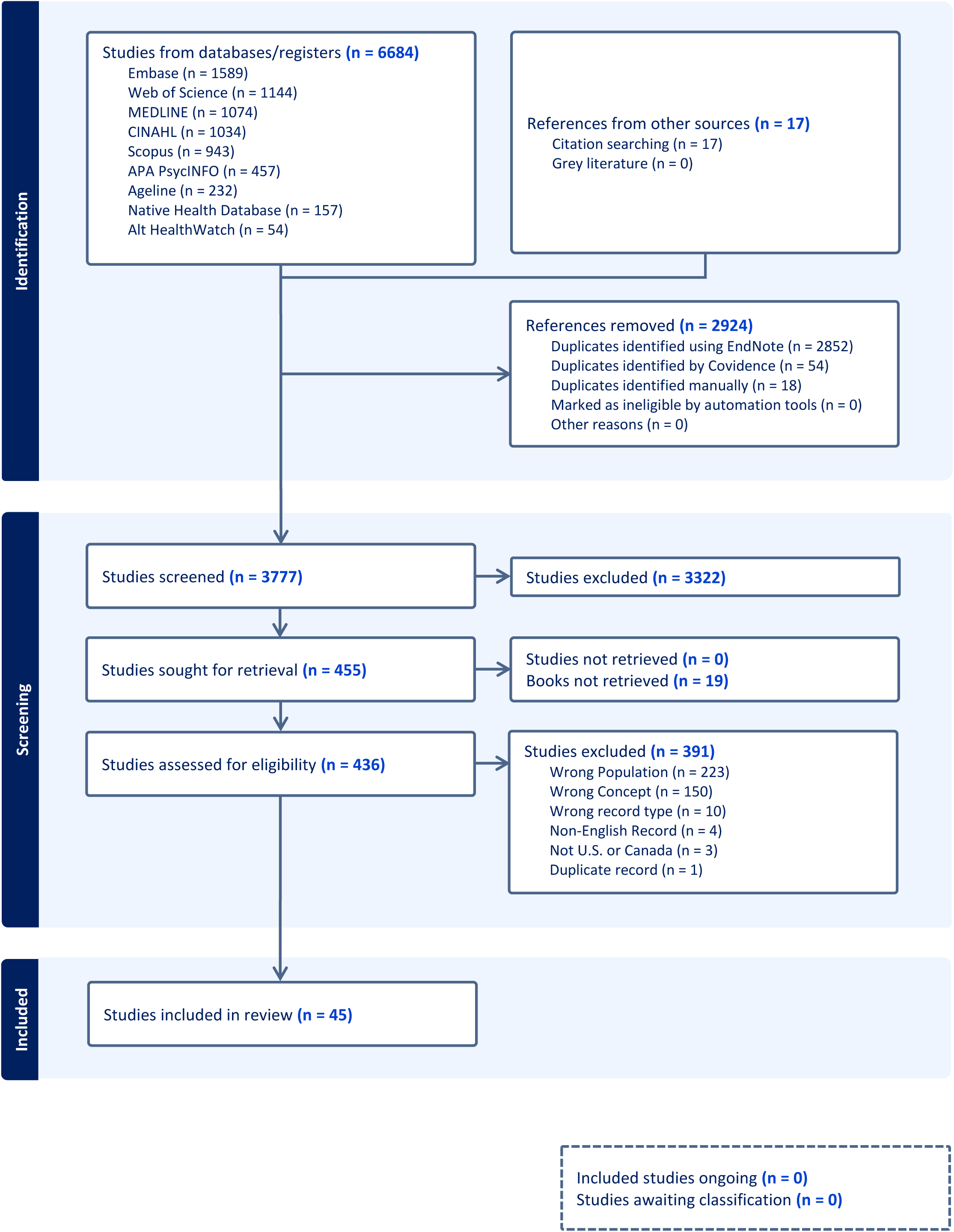
PRISMA Flow Diagram.

The included records encompassed a range of study designs and publication types: cohort or longitudinal studies (k=5), cross-sectional studies (k=18), a systematic review (k=1), qualitative research (k=7), text and opinion pieces (k=5), and other formats including a white paper (k=1), narrative reviews (k=4), mixed-methods studies (k=2), community-based participatory research (k=1), and a phytochemical assessment (k=1).

Table 1 summarizes the geographical context, Indigenous populations, and perimenopause and postmenopause healthcare focus of all included records. Table 1 is organized by type of research study. A complete record of extracted information is available in the Supplementary Materials of Appendix A.

### PCC summary: Population and context findings—Indigenous populations, age at hysterectomy and menopause, and geographic location

Of the 45 records included in this review, 14 (31·1%) examined Indigenous perimenopausal people as a subgroup within a broader population;^16,26,32–34,37,38,41–45,48,61^ 27 (60·0%) focused primarily on Indigenous perimenopausal populations;^14,25,27,35,36,39,40,46,47,49–60,62–64,68–70^ and four (8·8%) were ethnobotanical studies.^65–67,71^ Although these ethnobotanical records did not include participants, they contributed primary research on Indigenous plant use relevant to perimenopause and postmenopause care and were therefore included.

A U.S.-based study from 2018 found a median age of natural menopause of 50 among AI/AN participants.^42^ The range of mean age at menopause in the included literature was 47 to 51·2 years. A 2005 study reported a hysterectomy prevalence of 49·2%, and a 2023 study found a hysterectomy prevalence of 21·1%.^45^ The range of hysterectomy prevalence across the included literature was 22% to 49·2%. This highlights racial and ethnic disparities: while the literature revealed that hysterectomy rates declined across other groups, prevalence remained high among AI/AN participants. For records with a primary focus on Indigenous populations, Table 2 details the Indigenous groups represented, and reports mean or median age at natural or surgical menopause.

**Table 2:**
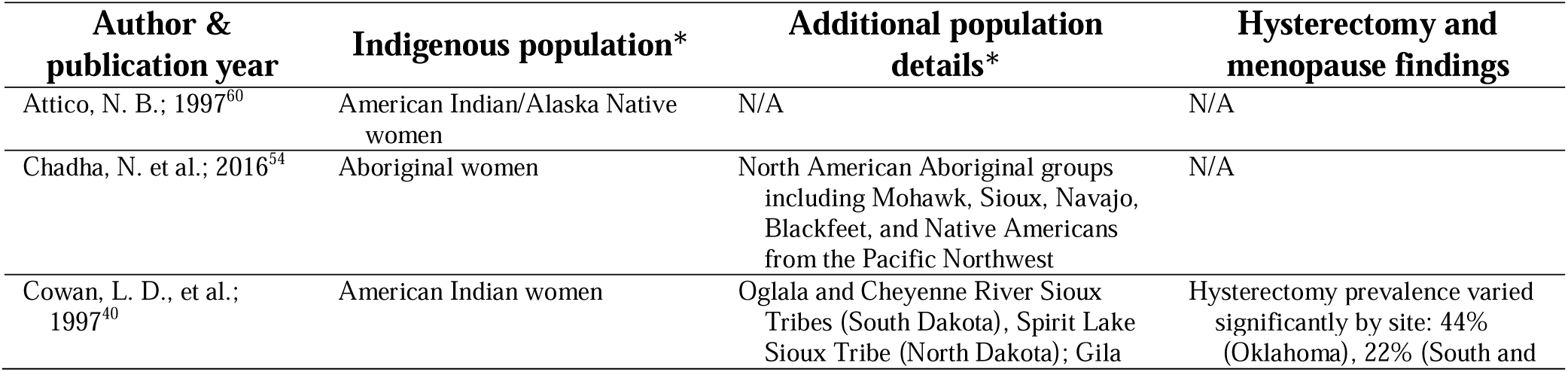
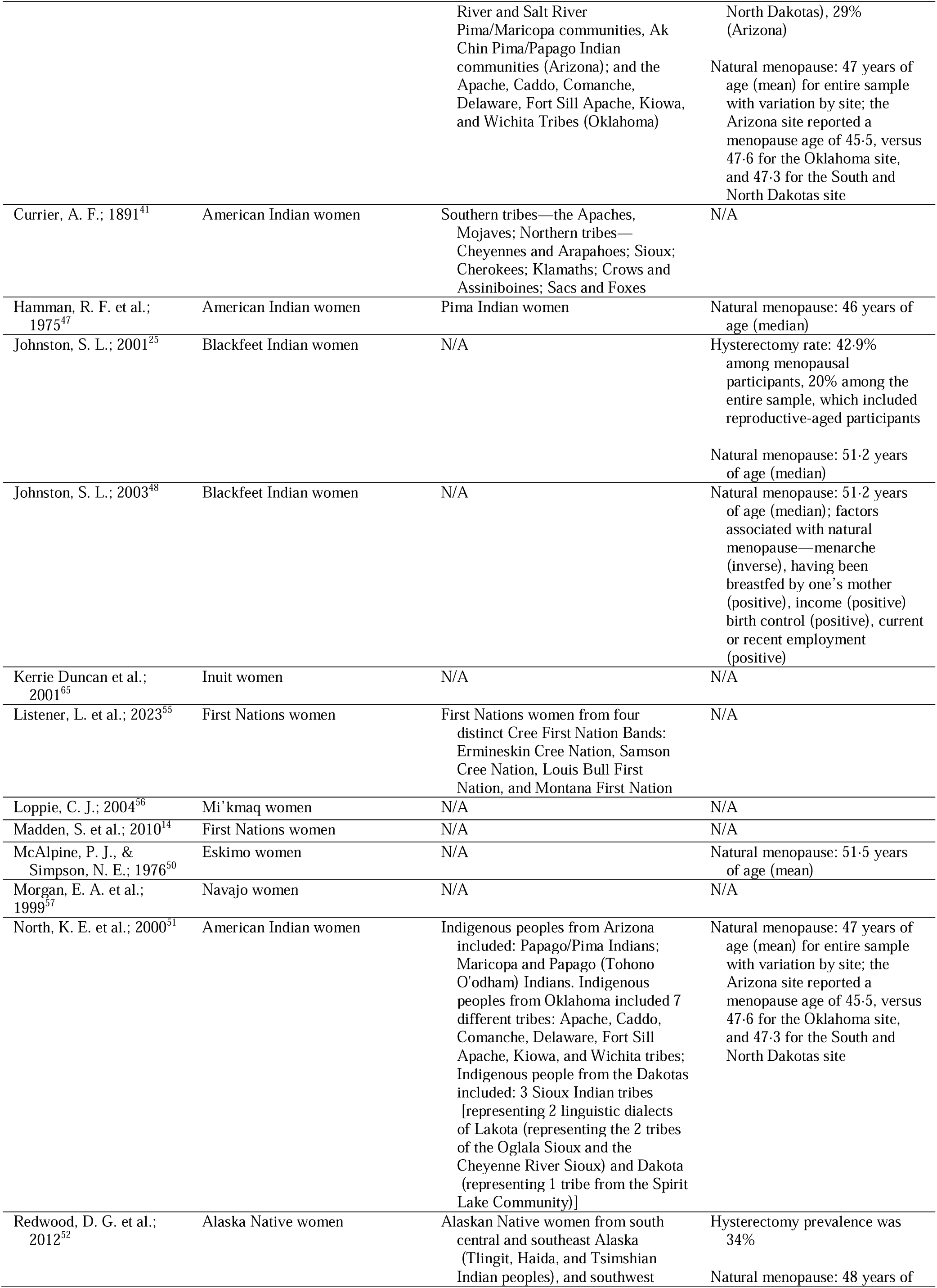
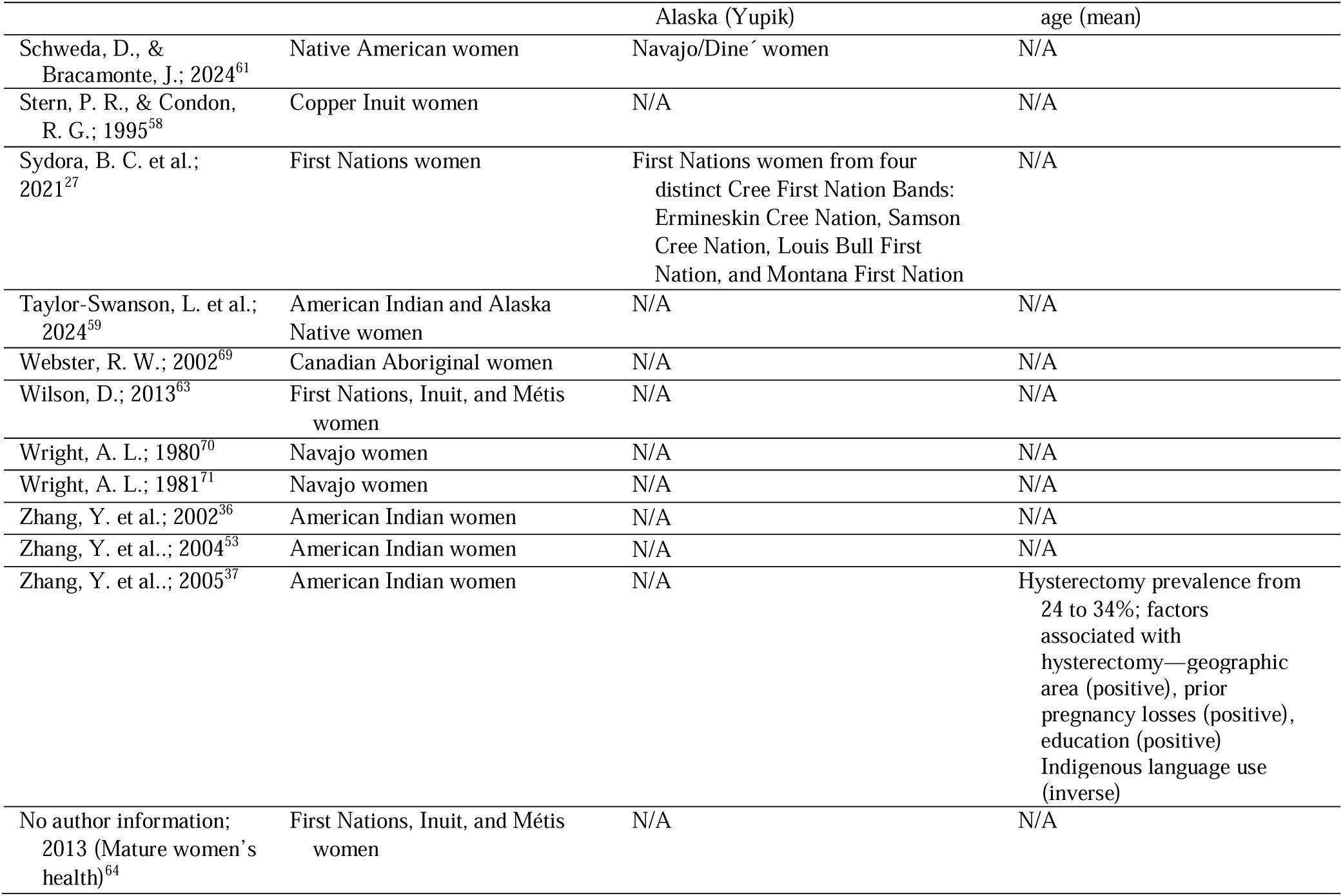
Indigenous populations who were included in reports with a primary focus on Indigenous health. *The description of Indigenous populations reflects the terminology used in original records.

Geographically, 32 records were conducted in the U.S.,12 in Canada, and one was based in the U.S., Canada, and Mexico. For this latter record, only results relevant to the U.S. and Canada were extracted. (See Table 1 for country context of all included records).

### PCC summary: Concept findings—perimenopause and postmenopause healthcare for Indigenous peoples in the United States and Canada

The literature included in this review underscores the importance of the cultural context in shaping the health experiences of midlife Indigenous individuals. One author described the MT as a “biocultural experience,” emphasizing that while physiological symptoms may occur across all cultures, the meaning, interpretation, and experience of this life stage are shaped by cultural context.^69^ For individuals not socialized within Western biomedical frameworks, the concept of “menopause” itself may be unfamiliar and may impact experiences of the midlife.^69^ Authors highlighted several contextual factors that healthcare providers and researchers should consider, including 1) the extent to which an individual’s beliefs and lifestyle align with their ancestors’ traditional beliefs and lifestyle, and 2) the role of community and family in shaping individual understandings of midlife.

#### Traditional Indigenous lifestyles

Several records in this review explored how traditional lifestyles shape midlife experiences among Indigenous peoples. In one study, females who lived a more traditional lifestyle—defined as rural residence, no formal education, income from welfare or sheep-herding, and Navajo-only language—conceptualized midlife differently from the Western biomedical framing of menopause.^69^ For example, Navajo participants described this life stage as marking the end of fertility and the beginning of grandparenthood.^69^ Similarly, a study of First Nations perimenopausal participants noted that in the Ojibway and Oji-Cree languages, there is no term for menopause; instead it is referred to simply as “the time when periods stop.”^14^ The absence of the Western biomedical framing may contribute to the underreporting of symptoms, particularly among more traditional participants.^69^ This was confirmed in a later qualitative study, which found that traditional Navajo participants were less likely to report symptoms to a provider. “Traditional lifestyle” was not defined in this record.^56^ In another mixed-methods study, most traditional Navajo participants sought Indigenous healers for the treatment of specific ailments— such as aching legs—rather than viewing menopause itself as a condition requiring treatment.^69^

Authors also suggest that life-course perspectives shape how the MT is experienced and interpreted. For example, in one study of First Nations females, participants frequently began their narratives about menopause by describing menarche (i.e., their first period).^14^ The authors posited that this life-stage continuity may reflect a holistic Indigenous view of womanhood and could inform culturally-aligned clinical communication.^14^ Similarly, among traditional Navajo participants, those who had participated in the Kinaaldá ceremony—a puberty rite celebrating entry into womanhood—were less likely to report menopause symptoms.^56^ Some participants believed this ceremony prepared them for life’s hardships and instilled resilience useful during the MT.^26^ This aligns with Indigenous knowledge systems that value the strength of females and emphasize the female role in caring for family and community.^55^ In one urban sample, AI/AN participants noted that traditional matriarchal teachings from grandmothers and mothers shaped their health behaviors; they were often seen as the backbone of the family, which led them to put their perimenopause and postmenopause struggles “on the back burner.”^58^ At the same time, this life stage was also associated with increased community respect and status,^14^ including the opportunity to participate in ceremonies^27^ or become medicine women.^55,69^

A mixed-methods study exploring cultural differences in perimenopause symptoms found that traditionality and acculturation influenced attitudes toward symptoms and treatment-seeking behaviors.^69^ In this study, Navajo participants were classified as either traditional (rural residents with no formal education, welfare or sheep-herding income, and Navajo-only language) or acculturated (small-town residents with at least a high-school education, wage-labor income, and both Navajo- and English-language use). Acculturated participants were more likely to discuss menopause, and, among those who sought care, to receive biomedical treatments. Additionally, acculturated participants with negative attitudes toward menopause reported more symptoms, while traditional participants who complained of poor health and poverty were more likely to experience greater symptom burden. These findings highlight the complex ways in which cultural orientation and social context shape the menopause experience. However, it should be noted that the bulk of this evidence was drawn from Navajo participants. As such, additional research is needed to explore how other traditional Indigenous lifestyles may uniquely impact responses to menopause.

#### Community and family

Records in this review emphasize the central role of Indigenous culture and kinship networks in shaping people’s understanding of the MT.^26,27,55^ However, many Indigenous participants reported not discussing menarche or menopause with their mothers, grandmothers, or elder females, complicating the issue.^14,26,27,54,55,58,60^ This silence may be self-perpetuating, as Indigenous people often refrain from sharing their discomfort during perimenopause and postmenopause, missing opportunities to share wisdom or normalize the experience.^55^

Colonialism has further exacerbated this issue. Participants linked cultural oppression and efforts to suppress Indigenous knowledge systems to reduced openness in discussing menopause outside their families or communities.^55^ The erosion of traditional remedies and teachings was also attributed to the legacy of residential schools and forced Westernization, which severed family and cultural connections.^27^

Despite these challenges, participants in several studies noted a generational shift, where more people are beginning to talk about menopause.^14^ They emphasized the importance of sharing information with younger generations,^55,58^ including daughters and sons, to foster supportive partnerships.^55^ Educating partners and family members was seen as a key strategy for promoting understanding and reducing stigma around menopause.^27^

Participants who did engage in conversations about menopause often found these discussions affirming. Observing symptoms in others and sharing personal experiences helped many better understand and accept their own symptoms.^27,58^ Common sources of information included mothers,^14,69^ other female relatives,^14,69^ and community members.^14,58^

### Perimenopause and postmenopause education

In addition to mothers, female relatives, and community members, some participants received perimenopause and postmenopause education from healthcare providers,^14,57^ and from books.^14^ However, many expressed limited knowledge of the MT and a desire for more comprehensive education.^54,58^ Participants called for more information on common symptoms,^14,54,58^ guidance on when to seek care,^54^ and available medications and treatment options.^14,58^ They emphasized that education should be delivered by female community members and should incorporate a well-rounded discussion of biomedical, integrative, and Indigenous treatments.^26,27,55,58,69^

A Navajo women’s healthcare provider noted that cultural perspectives influence a patient’s willingness to discuss menopause and openness to various treatment options.^60^ Some community-driven education efforts have already emerged. For example, a project with First Nations participants in Maskwacis, Alberta developed two publicly available pamphlets—one for significant others of those experiencing the MT and another for people going through the MT and their families.^27^ Similarly, a community-based initiative with urban AI/AN community-advisory board members tailored a series of menopause education modules in presentation format.^58^

### Perimenopause and postmenopause treatment

Several records examined patterns of treatment use for perimenopause and postmenopause symptoms among Indigenous participants. Two studies found that most participants did not use any treatment for symptoms,^27,69^ while others reported that participants relied either exclusively on culturally specific Indigenous forms of relief,^26,27,55,58,69^ or used these in combination with biomedical treatments.^26,27,32,55,58,69^ Although access to Indigenous treatments was sometimes limited, participants reported that Indigenous and integrative treatments were preferred as first-line interventions, followed by biomedical treatments.^27,58^

Reported Indigenous treatments included the use of traditional herbs,^26,27,55,58^ and spiritual practices such as faith,^55^ ceremonies, and other traditional practices.^26,27,58,69^ Integrative approaches included acupuncture,^58^ breathing and relaxation techniques,^27,58^ cannabis,^37^ chiropractic care,^58^ dietician support,^55,58^ healthy behaviors (e.g., diet and exercise),^26,27,32,58^ herbal supplements,^26,27,55,58^ massage,^58^ and meditation.^58^

Across records, participants frequently expressed dissatisfaction with biomedical care during perimenopause and postmenopause. Healthcare encounters were often described as rushed, impersonal, overly focused on medications, and lacking in educational support.^55,58^ Many participants were reluctant to use medications for menopause,^27,58^ instead preferring holistic and culturally aligned treatment options.^14,27,58^ Participants also emphasized the importance of provider awareness of language and cultural barriers.^14,58^ Expert consensus guidelines recommend that providers clarify Indigenous patients’ treatment preferences early in the decision-making process and offer transparent information on the benefits and risks of biomedical therapies.^60,61^

With respect to biomedical treatment use, studies found that Indigenous participants were less likely to have medications documented in their medical records compared to other racial and ethnic groups,^32^ and more likely to use over-the-counter or non-prescription therapies.^32,69^

Menopause hormone therapy (MHT) was a focus in several studies,^27,32,33,35,39,43,44,51,52,55,69^ which provided evidence of MHT use among Indigenous females. However, qualitative findings suggest that awareness and understanding of MHT may remain limited.^26^

### Perimenopause and postmenopause symptom experience

Four descriptive studies (three cross-sectional and one cohort) reported on perimenopause and postmenopause symptoms among Indigenous populations. Table 3 presents the proportions of the Indigenous subsamples reporting symptoms. While some studies identified statistically significant differences in symptom prevalence across racial and ethnic groups, several authors cautioned against interpreting these associational findings as biologically driven. For example, Kochersberger et al. (2024) noted that their Indigenous and First Nations participants were more likely to have low socioeconomic status.^48^ After adjusting for socioeconomic factors, the authors observed reductions in odds ratios, suggesting that social determinants play a key role in symptom prevalence.

**Table 3:**
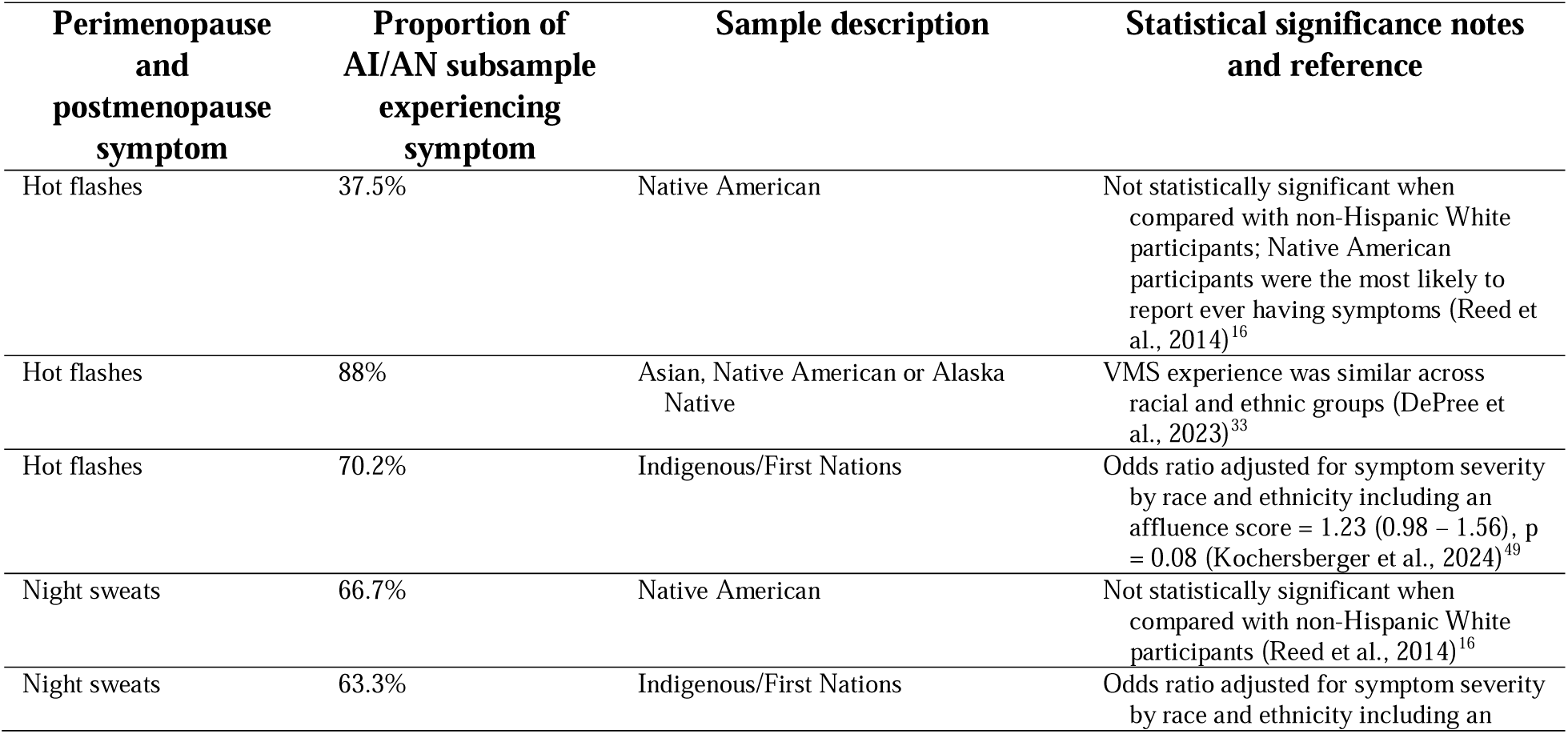
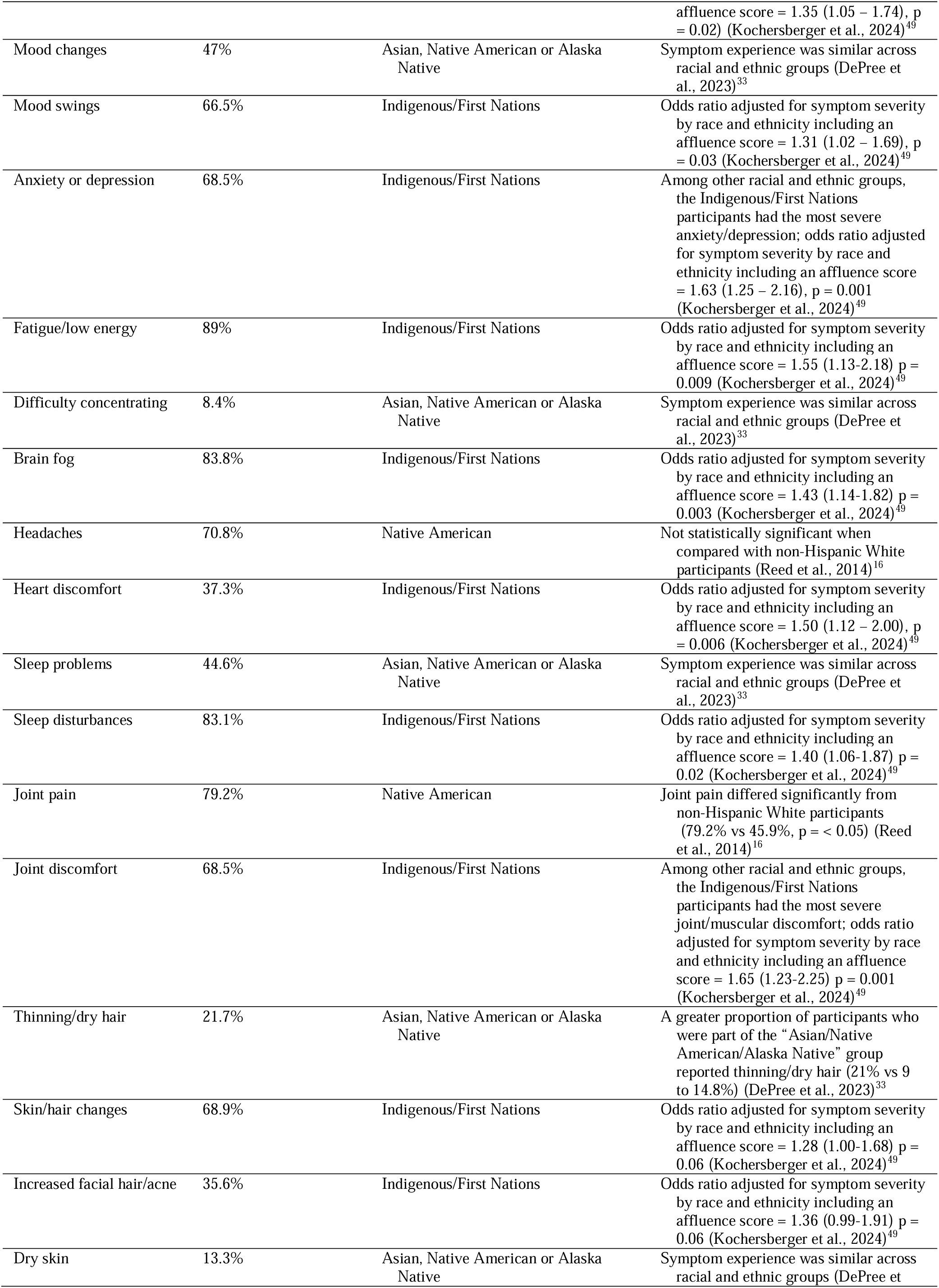
Proportion of Indigenous samples experiencing perimenopause and postmenopause symptoms.

Mixed-methods and qualitative studies included in this review offer important context for understanding how symptoms are experienced and reported. In one mixed-methods study, Navajo participants initially reported low levels of menopause-related changes. However, when asked about specific symptoms using a checklist, reporting increased.^70^ The author suggested that cultural expectations surrounding females’ midlife health may influence whether symptoms are disclosed. Similarly, a qualitative study of Navajo participants found variation in symptom reporting based on cultural and linguistic background. Among more traditional participants (those who spoke only Navajo and lived rurally), only one participant reported experiencing menopause symptoms, whereas symptoms were more commonly reported among less traditional participants (those who spoke English or were bilingual and lived in urban areas).^26^

### Rural versus urban differences in perimenopause and postmenopause healthcare

The differences in rural versus urban healthcare outcomes were reported in five studies.^26,39,45,50,58^ In addition to the rural and urban differences in symptom reporting outlined in the previous section (i.e., perimenopause and postmenopause experience), differences were found in patterns of hysterectomy prevalence. Surgical menopause was nearly twice as prevalent among urban participants in an Oklahoma center (44%) than among rural participants in Arizona (29%) and in the Dakotas (22%).^39^ Lastly, rural participants had access to Indigenous treatments on their reservation lands, which were difficult to access in urban/suburban areas.^58^

### Facilitators of and barriers to perimenopause and postmenopause healthcare among Indigenous populations

Authors of the records included in this scoping review identified facilitators of and barriers to perimenopause and postmenopause healthcare for Indigenous people. These facilitators and barriers are illustrated in Figure 2.

**Fig. 2.**
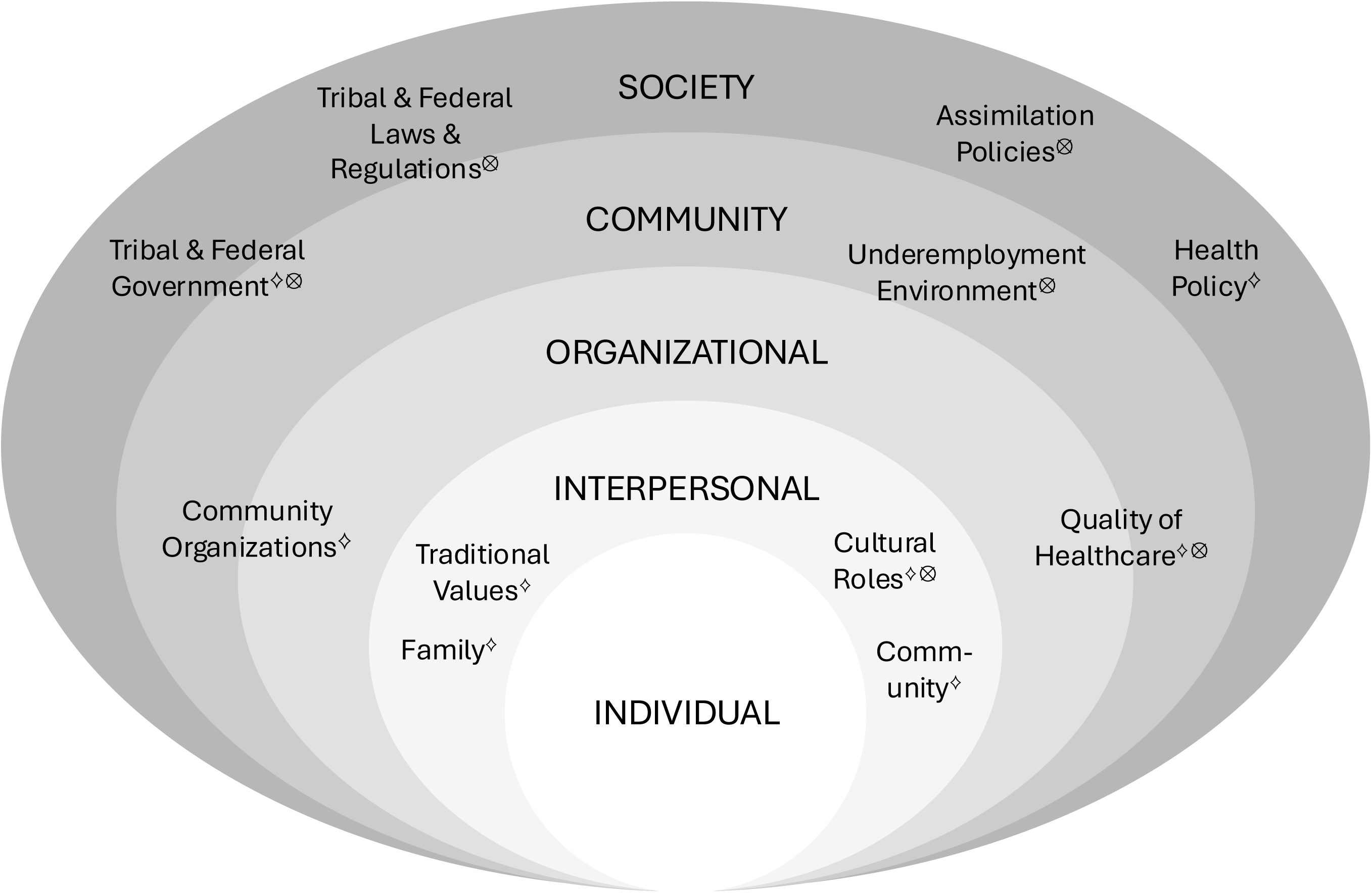
Barriers to and facilitators of Indigenous perimenopause and postmenopause health mapped to the socioecological model. ^⟡^The ^⟡^ symbol indicates a facilitator. ^11^The ^11^ symbol indicates a barrier.

Facilitators of healthcare access were identified at the societal, organizational, and interpersonal levels.^72^ At the societal level, two Canada-based studies highlighted policy and community collaboration as key facilitators of perimenopause healthcare for Indigenous populations. Wilson et al. (2013) emphasized the role of healthcare professionals in identifying the needs of First Nations and Inuit perimenopausal individuals and connecting them to the Non-Insured Health Benefits program.^62^ They also noted that Métis peoples are not eligible for this program, and stressed the importance of provider awareness concerning access challenges faced by this population.^62^ In a consensus report for the Inuit Tapiriit Kanatami Health Department, Kerrie Duncan (2001) recommended that Inuit peoples be directly involved in setting health priorities and determining the strategies used to address them.^64^

Organizational-level factors centered on features of the healthcare setting that can support healthcare uptake among Indigenous peoples. Providers can promote care by sharing information and discussing treatment options,^26,56,60^ incorporating traditional Indigenous treatments and practices,^55,56,58,64^ fostering environments of trust and cultural safety,^60,62^ and ensuring staff deliver culturally sensitive care.^62^ Providers should also recognize the diversity among Indigenous populations and prioritize building personal connections as a foundation for culturally centered care that fosters open communication and mutual understanding.^60^ Culturally responsive care includes slowing down and taking time to connect with patients.^26,58,60^ Tailored educational resources about perimenopause healthcare should be made available so individuals can revisit information and decide which treatment options are best for them.^26^ Similarly, new treatment instructions should be clearly explained and provided in take-home formats.^26^ At the interpersonal level, one study emphasized the importance of intergenerational capacity-building as a facilitator of healthcare uptake. Loppie (2004) recommended that both research and healthcare efforts support the transmission of Indigenous knowledge and practices related to midlife care and embed community values within healthcare delivery.^55^

Barriers to healthcare access were identified at the societal, community, organizational, and interpersonal levels. At the societal level, four Canada-based studies described structural conditions that hinder Indigenous perimenopause and postmenopause healthcare. Stern and Condon (1995) reported that federal policies aimed at forced assimilation disrupted the intergenerational transfer of health knowledge by weakening Indigenous language skills.^57^ Similarly, Loppie (2004) noted that the historical criminalization of traditional healing practices led to efforts within the Mi’kmaq community to safeguard Indigenous healing knowledge from appropriation by mainstream systems.^55^ Healthcare professionals working with First Nations communities also identified jurisdictional conflicts among federal, provincial, territorial, and band governments as barriers to delivering comprehensive care.^62^ Finally, limited access to government health reports—particularly those written in plain language and translated into Indigenous languages—was cited as an additional barrier to community-informed healthcare.^64^

At the community level, higher rates of unemployment among Indigenous people have contributed to socioeconomic disparities that limit access to healthcare.^62^ Several organizational-level barriers were also identified, particularly features of the healthcare system that hinder care uptake among Indigenous peoples. American Indian and Alaska Native participants reported that many providers lacked familiarity with Indigenous populations and their specific health needs. This included perceptions of cultural disconnect, and limited treatment options beyond biomedical care.^58^

Interpersonal-level barriers were often rooted in cultural roles and historical experiences. Among a sample of Inuit participants, self-preservation traits—such as willingness to adopt settler medicine and reluctance to engage in conflict—were described as adaptive strategies for survival that simultaneously limited continued engagement in traditional healing practices.^57^ Among Blackfeet participants, those with more traditional lifestyles were less likely to use the Indian Health Service.^25^ In an urban sample of AI/AN participants, participants described their matriarchal roles as a barrier to healthcare, explaining that family needs often took precedence over their own health.^58^

## DISCUSSION

This scoping review mapped evidence on perimenopause and postmenopause healthcare experiences, treatment preferences, and access among Indigenous peoples in the United States and Canada. The findings underscore the need for culturally grounded, patient-centered care and highlight important directions for future research and advocacy. In this discussion, we highlight recommendations from the reviewed literature regarding healthcare practice and research promotion to support Indigenous midlife health.

### Perimenopause and postmenopause healthcare advocacy for Indigenous populations

Several records advocated for comprehensive, culturally sensitive healthcare and called for additional research to support the health of Indigenous peoples of the United States and Canada. A key theme was importance of recognizing the heterogeneity of Indigenous peoples and cultures. Authors noted that factors such as acculturalization to Western lifestyles may shape individuals’ understanding of menopause, which is itself a Western medical concept.^60^ Cultural perspectives and treatment preferences must be central to patient-centered care for Indigenous midlife individuals.^60^ For instance, Navajo individuals have been reported to conceptualize the cessation of the menses in ways that differ greatly from Western framings of menopause.^26,69^ As a result, reporting of symptoms may be influenced by patients’ awareness of and beliefs about their connection to menopause.^70^

Healthcare providers should ensure patients are informed about common perimenopause and postmenopause changes, when to seek medical care, and the range of available treatment options.^54^ Providers should also consider individual beliefs, health-literacy levels, and potential misconceptions—particularly as they relate to MHT—and tailor education accordingly.^61^ Patient preference and readiness for treatment should be central to care-planning.^60^ This includes involving patients in shared decision-making, clearly communicating risks and benefits of treatment options, and providing follow-up care after treatment is initiated.^61^ Finally, providers should recognize the current lack of research focused on Indigenous midlife healthcare and advocate for more studies in this area.^62^

### Research promotion for Indigenous midlife health

The reviewed literature also highlights priorities for advancing research on Indigenous perimenopause and postmenopause health, which can contribute back to Indigenous peoples. Authors emphasized the importance of engaging with Indigenous people and organizations to collaboratively establish research priorities.^58,64^ Given that expectations and interpretations of midlife changes vary, researchers are encouraged to ask about VMS by name—such as hot flashes and night sweats—rather than relying on general questions about health changes during the MT.^70^

Authors called for additional research on Indigenous midlife health, disease prevention, and health promotion, including research investigating the links between estrogen and long-term outcomes such as cardiovascular and bone health.^53,54,59,62,64,68,70^ Studies are also needed to examine both the types of perimenopause healthcare Indigenous people receive and their lived experience of the MT, in order to better capture the diversity of needs and perspectives.^53,68^ In addition, researchers should investigate disparities in access to care, experiences of discrimination within the healthcare setting, and root causes of symptom severity during the MT.^48^ These lines of inquiry are crucial for understanding inequities in symptoms prevalence (including symptom clusters such as pain, sleep, mood, and cognition) and treatment engagement among Indigenous midlife populations.

Finally, associational findings should be interpreted with caution. While some studies report a higher prevalence of VMS among Indigenous subsamples, these findings are correlational and should not be interpreted as evidence of a causal link with race or ethnicity. Instead, further research is needed to explore the underlying social determinants of health that may contribute to these disparities. This scoping review has limitations. First, although we searched nine databases and references of included literature, unpublished studies and studies not indexed or cited with our selected sources will have been missed. Reporting bias was increased when we excluded books at full-text review for feasibility. Second, we limited inclusion to English-language records, as our team did not have the resources to review non-English materials. As a result, it is possible that some relevant records could have been missed. These limitations, combined with scarce data on Indigenous perimenopause and postmenopause health, limited the evidence for our study. Strengths of this review include its methodological rigor and focus on an underexplored topic. Indigenous midlife peoples and perimenopause health are underrepresented in the literature. To reduce this gap, we broadened our scope to include Indigenous, integrative, and biomedical healthcare, recognizing that many Indigenous people prefer traditional or holistic approaches to care. In addition, our review team took intentional steps to center Indigenous ways of knowing throughout the research process with the goal of developing recommendations for practice and research recommendations that support patient-centered midlife care for Indigenous peoples.

## CONCLUSION

This scoping review highlights the limited body of literature on perimenopause and postmenopause healthcare among Indigenous peoples in the United States and Canada. The findings underscore the need for culturally responsive, patient-centered care that acknowledges diverse perspectives on midlife health. Indigenous individuals face unique barriers to care and often prefer traditional and integrative treatments, yet biomedical systems remain underprepared to meet these needs. Addressing these gaps will require collaboration with Indigenous people and organizations, expanded research, and systemic changes to ensure equitable access to care during the menopausal transition and beyond.

## Contributors

JK-M—conception, analysis, data interpretation, writing (original draft), reviewing, editing.

LT-S—conception, analysis, data interpretation, writing (original draft), reviewing, editing.

P-YH—analysis, data interpretation, writing, reviewing, editing.

SES—conception, analysis, data interpretation, writing (original draft), reviewing, editing.

SA—conception, analysis, data interpretation, writing, reviewing, editing.

TA—conception, analysis, data interpretation, writing, reviewing, editing.

LC—data interpretation, writing, reviewing, editing.

MEM—analysis, data interpretation, writing, reviewing, editing.

JAE—conception, analysis, data interpretation, writing, reviewing, editing.

SQ—analysis, writing, reviewing, editing.

SRR—analysis, writing, reviewing, editing.

LK—analysis, writing, reviewing, editing.

MMM—conception, curation, writing, reviewing, editing.

LAH—conception, curation, investigation, writing (original draft), reviewing, editing. All authors had full access to all the data in the review and had final responsibility for the decision to submit for publication.

## Data Sharing

The protocol for this review can be found on Open Science Framework at: osf.io/hwna5. Please see the Search Strategies in the Supplementary Materials document for a complete search strategy and the data extracted from all records included in this review.

## Acknowledgments

JK-M was funded by the National Clinician Scholars Program (NCSP) as a postdoctoral fellow at the University of Pennsylvania. Additionally, she was supported by the National Institute of Nursing Research of the National Institutes of Health under Award Number F31NR020431. The content is solely the responsibility of the authors and does not necessarily represent the official views of the National Institutes of Health. The funders were not involved in the design and writing of this review. The authors were supported by a grant from the University of Utah Office of the Vice President for Research.

## Appendix A. Supplemental Material

Supplementary materials related to this review can be found at [enter doi].

During the preparation of this work the authors used Covidence software in order to filter the records most likely to be included. Although this feature of the Covidence software uses AI, all records were screened and given full consideration during the screening process. After using this tool, the authors reviewed and edited the content as needed and take full responsibility for the content of the publication.

## References

1. Arias E, Xu J, Curtin S, Bastian B, Tejada-Vera B. Mortality profile of the non-Hispanic American Indian or Alaska Native population, 2019. Natl Vital Stat Rep. 2021;70(12):1– 27.

2. Tjepkema M, Bushnik T, Bougie E. Life expectancy of First Nations, Métis and Inuit household populations in Canada. Health Rep. 2019;30(12):3–10.

3. Indian Health Service. Disparities. https://www.ihs.gov/newsroom/factsheets/disparities/. Published 2019. Accessed February 27, 2023.

4. King M. Chronic diseases and mortality in Canadian Aboriginal peoples: learning from the knowledge. Chronic Dis Can. 2010;31(1):2–3.

5. Espey DK, Jim MA, Cobb N, et al. Leading causes of death and all-cause mortality in American Indians and Alaska Natives. Am J Public Health. 2014;104 Suppl 3:S303–311.

6. Turin TC, Saad N, Jun M, et al. Lifetime risk of diabetes among First Nations and non-First Nations people. CMAJ. 2016;188(16):1147–1153.

7. Vervoort D, Kimmaliardjuk DM, Ross HJ, Fremes SE, Ouzounian M, Mashford-Pringle A. Access to cardiovascular care for indigenous peoples in Canada: a rapid review. CJC Open. 2022;4(9):782–791.

8. Mazereeuw MV, Withrow DR, Diane Nishri E, Tjepkema M, Marrett LD. Cancer incidence among First Nations adults in Canada: follow-up of the 1991 Census Mortality Cohort (1992-2009). Can J Public Health. 2018;109(5-6):700–709.

9. Nishri ED, Sheppard AJ, Withrow DR, Marrett LD. Cancer survival among First Nations people of Ontario, Canada (1968-2007). Int J Cancer. 2015;136(3):639–645.

10. Jerome-D’Emilia B, Gachupin FC, Suplee PD. A systematic review of barriers and facilitators to mammography in American Indian/Alaska Native women. J Transcult Nurs. 2019;30(2):173–186.

11. Risendal B, DeZapien J, Fowler B, Papenfuss M, Giuliano A. Pap smear screening among urban Southwestern American Indian women. Prev Med. 1999;29(6 Pt 1):510–518.

12. Mannix TR, Austin SD, Baayd JL, Simonsen SE. A community needs assessment of urban Utah American Indians and Alaska Natives. J Community Health. 2018;43(6):1217–1227.

13. Roubidoux MA, Richards B, Honey NE, Begay JA. Adherence to screening among American Indian women accessing a mobile mammography unit. Acad Radiol. 2021;28(7):944–949.

14. Madden S, St Pierre-Hansen N, Kelly L, Cromarty H, Linkewich B, Payne L. First Nations women’s knowledge of menopause: experiences and perspectives. Can Fam Physician. 2010;56(9):e331–337.

15. Lei L, Leggett AN, Maust DT. A national profile of sandwich generation caregivers providing care to both older adults and children. J Am Geriatr Soc. 2022;71(3):799–809.

16. Reed SD, Lampe JW, Qu C, et al. Premenopausal vasomotor symptoms in an ethnically diverse population. Menopause. 2014;21(2):153–158.

17. Biglia N, Cagnacci A, Gambacciani M, Lello S, Maffei S, Nappi RE. Vasomotor symptoms in menopause: a biomarker of cardiovascular disease risk and other chronic diseases? Climacteric. 2017;20(4):306–312.

18. Muka T, Oliver-Williams C, Colpani V, et al. Association of vasomotor and other menopausal symptoms with risk of cardiovascular disease: a systematic review and meta-analysis. PLoS One. 2016;11(6):e0157417.

19. Peters MDJ GC, McInerney P, Munn Z, Tricco AC, Khalil H. JBI Manual for Evidence Synthesis, JBI, 2020. https://synthesismanual.jbi.global. Published 2020. Accessed March, 21, 2023.

20. Arksey H, O’Malley L. Scoping studies: towards a methodological framework. Int J Soc Res Methodol. 2005;8(1):19–32.

21. Tricco AC, Lillie E, Zarin W, et al. PRISMA extension for scoping reviews (PRISMA-ScR): checklist and explanation. Ann Intern Med. 2018;169(7):467–473.

22. Rethlefsen ML, Kirtley S, Waffenschmidt S, et al. PRISMA-S: an extension to the PRISMA Statement for Reporting Literature Searches in Systematic Reviews. Syst Rev. 2021;10(1):39.

23. Veritas Health Innovation [computer program]. Covidence systematic review software. Melbourne, Australia. Available at www.covidence.org

24. EndNote [computer program]. Version EndNote X9. Philadelphia, PA: Clarivate; 2013.

25. Johnston SL. Associations with age at natural menopause in Blackfeet women. Am J Hum Biol. 2001;13(4):512–520.

26. Mingo C, Herman CJ, Jasperse M. Women’s stories: ethnic variations in women’s attitudes and experiences of menopause, hysterectomy, and hormone replacement therapy. J Womens Health Gend Based Med. 2000;9 Suppl 2:S27–38.

27. Sydora BC, Graham B, Oster RT, Ross S. Menopause experience in First Nations women and initiatives for menopause symptom awareness; a community-based participatory research approach. BMC Womens Health. 2021;21(1):179.

28. United States Bureau of Indian Affairs. Indian entities recognized by and eligible to receive services from the United States Bureau of Indian Affairs. Fed Regist. 2024;89:944–8.

29. Government of Canada. Crown-indigenous relations and northern affairs Canada. Search by First Nations [internet].

30. McGowan J, Sampson M, Salzwedel DM, Cogo E, Foerster V, Lefebvre C. PRESS Peer Review of Electronic Search Strategies: 2015 guideline statement. J Clin Epidemiol. 2016;75:40–46.

31. Harlow SD, Gass M, Hall JE, et al. Executive summary of the Stages of Reproductive Aging Workshop + 10: addressing the unfinished agenda of staging reproductive aging. J Clin Endocrinol Metab. 2012;97(4):1159–1168.

32. DePree B, Houghton K, Shiozawa A, et al. Treatment and resource utilization for menopausal symptoms in the United States: a retrospective review of real-world evidence from US electronic health records. Menopause. 2023;30(1):70–79.

33. Gavaler JS, Bonham-Leyba M, Castro CA, Harman SE. The Oklahoma Postmenopausal Women’s Health Study: recruitment and characteristics of American Indian, Asian, Black, Hispanic, and Caucasian women. Alcohol Clin Exp Res. 1999;23(2):220–223.

34. Howard BV, Kuller L, Langer R, et al. Risk of cardiovascular disease by hysterectomy status, with and without oophorectomy: the Women’s Health Initiative Observational Study. Circulation. 2005;111(12):1462–1470.

35. Zhang Y, Howard BV, Cowan LD, et al. The effect of estrogen use on levels of glucose and insulin and the risk of type 2 diabetes in American Indian postmenopausal women: the Strong Heart Study. Diabetes Care. 2002;25(3):500–504.

36. Zhang Y, Lee ET, Cowan LD, North KE, Wild RA, Howard BV. Hysterectomy prevalence and cardiovascular disease risk factors in American Indian women. Maturitas. 2005;52(3-4):328–336.

37. Babyn K, Ross S, Makowsky M, Kiang T, Yuksel N. Cannabis use for menopause in women aged 35 and over: a cross-sectional survey on usage patterns and perceptions in Alberta, Canada. BMJ Open. 2023;13(6):e069197.

38. Carty CL, Spencer KL, Setiawan VW, et al. Replication of genetic loci for ages at menarche and menopause in the multi-ethnic Population Architecture using Genomics and Epidemiology (PAGE) study. Hum Reprod. 2013;28(6):1695–1706.

39. Cowan LD, Go OT, Howard BV, et al. Parity, postmenopausal estrogen use, and cardiovascular disease risk factors in American Indian women: the Strong Heart Study. J Womens Health. 1997;6(4):441–449.

40. Currier AF. A study relative to the functions of the reproductive apparatus in American Indian women. Med News. 1891.

41. DePree BJ, Shiozawa A, Kim J, Wang Y, Yang H, Mancuso S. Treatment satisfaction, unmet needs, and new treatment expectations for vasomotor symptoms due to menopause: women’s and physicians’ opinions. Menopause. 2024;31(9):769–780.

42. Fernández-Rhodes L, Malinowski JR, Wang Y, et al. The genetic underpinnings of variation in ages at menarche and natural menopause among women from the multi-ethnic Population Architecture using Genomics and Epidemiology (PAGE) Study: a trans-ethnic meta-analysis. PLoS One. 2018;13(7):e0200486.

43. Gavaler JS. Oral hormone replacement therapy: factors that influence the estradiol concentrations achieved in a multiracial study population. J Clin Pharmacol. 2002;42(2):137–144.

44. Gavaler JS, Rosenblum E. Predictors of postmenopausal body mass index and waist hip ratio in the Oklahoma postmenopausal health disparities study. J Am Coll Nutr. 2003;22(4):269–276.

45. Gopalani SV, Dasari SR, Adam EE, Thompson TD, White MC, Saraiya M. Variation in hysterectomy prevalence and trends among U.S. States and territories-Behavioral Risk Factor Surveillance System, 2012-2020. Cancer Causes Control. 2023;34(10):829–835.

46. Hamman RF, Bennett PH, Miller M. The effect of menopause on serum cholesterol in American (Pima) Indian women. Am J Epidemiol. 1975;102(2):164–169.

47. Johnston SL. Menopause in Blackfeet women--a life span perspective. Coll Antropol. 2003;27(1):57–66.

48. Kochersberger A, Coakley A, Millheiser L, et al. The association of race, ethnicity, and socioeconomic status on the severity of menopause symptoms: a study of 68,864 women. Menopause. 2024;31(6):476–483.

49. McAlpine PJ, Simpson NE. Fertility and other demographic aspects of the Canadian Eskimo communities of Igloolik and Hall Beach. Hum Biol. 1976;48(1):114–138.

50. North KE, MacCluer JW, Cowan LD, Howard BV. Gravidity and parity in postmenopausal American Indian women: the Strong Heart Study. Hum Biol. 2000;72(3):397–414.

51. Redwood DG, Lanier AP, Johnston JM, Murphy N, Murtaugh MA. Reproductive cancer risk factors among Alaska Native women: the Alaska Education and Research Towards Health (EARTH) Study. Womens Health Issues. 2012;22(4):e387–393.

52. Zhang Y, Howard BV, Cowan LD, et al. Associations of postmenopausal hormone therapy with markers of hemostasis and inflammation and lipid profiles in diabetic and nondiabetic American Indian women: the Strong Heart Study. J Womens Health (Larchmt). 2004;13(2):155–163.

53. Chadha N, Chadha V, Ross S, Sydora BC. Experience of menopause in aboriginal women: a systematic review. Climacteric. 2016;19(1):17–26.

54. Listener L, Ross S, Oster R, Graham B, Heckman S, Voyageur C. Nehiyawak (Cree) women’s strategies for aging well: community-based participatory research in Maskwacîs, Alberta, Canada, by the Sohkitehew (Strong Heart) group. BMC Womens Health. 2023;23(1):341.

55. Loppie CJ. Grandmothers’ Voices: Mi’kmaq women and menopause [dissertation]. Halifax (NS): Dalhousie University; 2004. Available from: ProQuest Dissertations & Theses Global.

56. Morgan EA, Herman CJ, Coriz M, Whalawitsa M, Mingo C. The effect of traditionality on menopausal symptoms [abstract]. JIM. 1999;47(2):11.

57. Stern PR, Condon RG. Puberty, pregnancy, and menopause: lifecycle acculturation in a Copper Inuit community. Arctic Med Res. 1995;54(1):21–31.

58. Taylor-Swanson L, Kent-Marvick J, Austin SD, et al. Developing a menopausal transition health promotion intervention with indigenous, integrative, and biomedical health education: a community-based approach with urban American Indian/Alaska Native women. Glob Adv Integr Med Health. 2024;13:27536130241268232.

59. Attico N. The elder female: a preventative care plan and health watch. IHS Primary Care Provider. 1997;22(5):80–87.

60. Schweda D, Bracamonte J. Of a “certain age” - menopause in Native American women: a cultural perspective. Maturitas. 2024;190:107947.

61. Thomas MA, Kessel B, Miles T, et al. Strategies to encourage hormone replacement therapy use in minority or diverse populations: a consensus opinion. J Womens Health (Larchmt). 2000;1(1):46–52.

62. Wilson D, de la Ronde S, Brascoupé S, et al. Health professionals working with First Nations, Inuit, and Métis consensus guideline. J Obstet Gynaecol Can. 2013;35(6):550– 558.

63. RETIRED: Mature women’s health. JOGC. 2013;35(6):S37–S37. Retired.

64. Kerrie Duncan; Inuit Tappiriit Kanatami. National Inuit Health Information Conference: Inuit defined health information needs and directions; 2001 Jun; Inuvik, NT. Inuvik (NT): National Inuit Health Information Conference; 2001.

65. Adams JD, Jr., Garcia C. Women’s health among the Chumash. Evid Based Complement Alternat Med. 2006;3(1):125–131.

66. Johnson TL, Fahey JW. Black cohosh: coming full circle? J Ethnopharmacol. 2012;141(3):775–779.

67. McKenna DJ, Jones K, Humphrey S, Hughes K. Black cohosh: efficacy, safety, and use in clinical and preclinical applications. Altern Ther Health Med. 2001;7(3):93–100.

68. Webster RW. Aboriginal women and menopause. J Obstet Gynaecol Can. 2002;24(12):938–940.

69. Wright AL. *Cultural Variability in the experience of menopause: a comparison of Navajo and Western data* [dissertation]. Ann Arbor (MI): University of Michigan; 1980. Available from: ProQuest Dissertations & Theses.

70. Wright AL. On the calculation of climacteric symptoms. Maturitas. 1981;3(1):55–63.

71. Burton TD, T.; Dong, H.; Li, G.; Bolton, J.; Soejarto, D.; & Van Breemen, R. B. American Indian botanicals as possible alternatives to hormone therapy during menopause [abstract]. Planta Medica. 2016;82(5).

72. Bronfenbrenner U. Toward an experimental ecology of human development. Am Psychol. 1977;32(7):513–31.

