## Supplement 1 for "Indigenous, integrative, and biomedical perimenopause and postmenopause healthcare for Indigenous midlife peoples residing in the United States and Canada: A scoping review"

Appendix a: Search Strategies

### Strategy Development and Notes

Keywords and subject headings were harvested from sentinel studies^1,2,3,4,5^, database controlled vocabularies, team feedback, and U.S. and Canadian government resources^6,7^. Search terms included variations of menopause, vasomotor symptoms, Indigenous Americans, and First Nations.

The following databases were searched from inception or institutional license: Medline (Ovid) 1946-2024, Embase (Elsevier) 1974 – 2024, CINAHL Complete (Ebscohost) 1937-2024, Ageline (Ebscohost) 1966-2024, Alt HealthWatch (Ebscohost) 1984-2024, APA PsycInfo (Ebscohost) 1872-2024, Scopus (Elsevier) 1970-2024, Web of Science Core Collection (Clarivate) 1900-2024, and Native Health Database (1900-2024).

Searches were initially conducted between June 1st and 9th 2023 and then updated on November 21st, 2024. In CINAHL, the publication types of “Book Review,” “Brief Item,” “Obituary,” or “Poetry” were excluded. Book reviews were excluded in Web of Science Core Collection. Otherwise, no search filters or limiters were applied to initial searches. Searches were updated on November 21st, 2024, and limited to publication date (Ageline, Alt HealthWatch, Web of Science Core Collection) or database entry date (Medline, Embase, CINAHL, PsycInfo, Scopus) after the initial searches in June 2023. Databases on the Ebscohost platform were searched independently rather than simultaneously.

1. Johnston SL. Associations with age at natural menopause in Blackfeet women. Am J Hum Biol. 200;13(4):512-20. doi: 10.1002/ajhb.1083. PMID: 11400222.

2. Mingo C, Herman CJ, Jasperse M. Women's stories: ethnic variations in women's attitudes and experiences of menopause, hysterectomy, and hormone replacement therapy. J Womens Health Gend Based Med. 2000;9 Suppl 2:S27-38. doi: 10.1089/152460900318740. PMID: 10714743.

3. Reed SD, Lampe JW, Qu C, Copeland WK, Gundersen G, Fuller S, Newton KM. Premenopausal vasomotor symptoms in an ethnically diverse population. Menopause. 2014;21(2):153-8. doi: 10.1097/GME.0b013e3182952228. PMID: 23760434.

4. Madden S, St Pierre-Hansen N, Kelly L, Cromarty H, Linkewich B, Payne L. First Nations women's knowledge of menopause: experiences and perspectives. Can Fam Physician. 2010;56(9):e331-7. PMID: 20841572; PMCID: PMC2939134.

5. Sydora BC, Graham B, Oster RT, Ross S. Menopause experience in First Nations women and initiatives for menopause symptom awareness; a community-based participatory research approach. BMC Womens Health. 2021;21(1):179. doi: 10.1186/s12905-021-01303-7.

6. Search by First Nation. Crown-Indigenous Relations and Northern Affairs Canada. Accessed 1 April 2023, <https://fnp-ppn.aadnc-aandc.gc.ca/FNP/Main/Search/SearchFN.aspx?lang=eng>

7. United States Bureau of Indian Affairs. Indian Entities Recognized by and Eligible To Receive Services From the United States Bureau of Indian Affairs. *Fed Regist*. 89(5):944-948.

### Ovid Medline

**Field codes:** / = Medical Subject Heading (MeSH); ti = article title; ab = abstract; kf = keyword heading word; kw = keyword heading (author keywords); ui = unique identifier; ez = entrez date (date added to Medline).
**Proximity operator:** adj# 
**Truncation:**  *

Ovid MEDLINE(R) and Epub Ahead of Print, In-Process, In-Data-Review & Other Non-Indexed Citations, Daily and Versions <1946 to November 20, 2024>

1	"American Indian or Alaska Native"/ or Indians, North American/ or Alaska Natives/ or (((American* or Alaska*) adj2 (Native* or Indian or Indians or indigenous or aborigin*)) or Amerind*).ti,ab,kf,kw. [Native American / Alaska Natives, general]	29407

2	Indigenous Canadians/ or Inuit/ or (((Canada or Canadian*) adj2 (native* or indigenous or aborigin*)) or Metis or "Métis" or Michif* or Mitchif or Mechif or "Métif" or "Métchif" or Metif or Metchif or Inuit or "First Nations" or "First Nation").ti,ab,kf,kw.	8447

3	Navajo People/ or Pima People/ or Inuit/ [Tribe Index Terms]	4105

4	("Asa’carsarmiut" or "Dee-ni’" or "Mi’kmaq" or "S’Klallam" or "Shoonaq’" or "Sun’aq" or (("Ivanof Bay" or "Shoalwater Bay" or "Spirit Lake" or "Squaxin Island" or Dakota or Dakotah or Wichita or keechi or waco or Tawakonie or Cheyenne or Creek or Crow or Hoonah or Kasigluk or Maricopa or Miami or Omaha or Oneida or Ottawa or Peoria or Salamatof or Sitka or Snoqualmie or Spokane or Winnebago or Wyandotte) adj2 (Indian or Indians or Tribe or Tribes or native or natives or nation or people*)) or ((Native or indian or traditional) adj2 (village or council or community or association)) or Absentee-Shawnee or Achomawi or Agdaagux or "Agua Caliente" or Ajumawi or Ak-Chin or Akiachak or Akiak or Alabama-Coushatta or Alabama-Quassarte or Alatna or Algaaciq or Algin or Allakaket or "Alturas Indian Rancheria" or Alutiiq or Angoon or Anvik or Apache or Aporige or Aquinnah or arapaho or Aroostook or Assiniboine or Astarawi or Atsugewi or Atwamsini or Bdewakantunwan or Caddo or Cahto or Cahuilla or Catawba or Cayuga or Chalkyitsik or Cheesh-Na or Chehalis or Chemehuevi or Cher-Ae or Chevak or Cheyenne-Arapaho or Chickahominy or Chickaloon or Chickasaw or Chignik or Chilkat or Chilkoot or Chinik or Chinook or Chippewa* or Chitimacha or Choctaw or Chukchansi or Chuloonawick or Chumash or Cocopah or Colusa or Comanche or Coos or Cosmit or Coushatta or Cowlitz or "Cross Village" or Cupeno or Curyung or Diegueno or Dine' or Egegik or Eklutna or Elwha or "Emmonak Village" or "Evansville Village" or Ewiiaapaayp or "Galena Village" or Goshute or "Gulkana Village Council" or Haida or Hammawi or Havasupai or "Healy Lake Village" or Hewisedawi or Ho-Chunk or Hoh or Hokan or Holikachuk or "Holy Cross Tribe" or Hualapai or "Hughes Village" or "Huslia Village" or "Hydaburg Cooperative Association" or "Igiugig Village" or Iipay or Ilmawi or Inaja or Inupiat or Ione or Itsatawi or "Kaguyak Village" or Kaibab or "Kaktovik Village" or Kalispel or Kanosh or Karuk or Kashia or Kaw or Keetoowah or "Kenaitze Indian Tribe" or "Ketchikan Indian Community" or "Keweenaw Bay" or Kialegee or Kickapoo or "King Island Native Community" or "King Salmon Tribe" or Kiowa or Klamath or "Klawock Cooperative Association" or Kletsel or "Knik Tribe" or "Koi Nation" or "Kokhanok Village" or Koosharem or Kootenai or Kosalektawi or "Koyukuk Native Village" or Kumeyaay or Kwalhioqua or "Lesnoi Village" or "Levelock Village" or "Lime Village" or "Louden Village" or Luiseno or Lummi or Madesi or Maidu or Makah or Maliseet or "Manley Hot Springs Village" or "Manokotak Village" or Manzanita or Mashantucket or Match-e-be-nash-she-wish or Mattaponi or "McGrath Native Village" or Mdewakanton or Mechoopda or Menominee or Me-Wuk or Miccosukee or Micmacs or "Mission Indians" or Miwok or Miwuk or Modoc or Mohegan or Mojave or Monacan or "Mono Indians" or Muckleshoot or Muscogee or "Na Dené" or Nansemond or Narragansett or Navajo or "New Koliganek Village" or "New Stuyahok Village" or "Newhalen Village" or "Newtok Village" or "Nez Perce" or "Nikolai Village" or "Ninilchik Village" or Nisqually or Nomlaki or "Nondalton Village" or Nooksack or "Northway Village" or Nottawaseppi or "Nulato Village" or Nunakauyarmiut or Odawa or Ohgsenakale or "Ohkay Owingeh" or Onondaga or Oreilles or "Organized Village of Grayling" or "Organized Village of Kake" or "Organized Village of Kasaan" or "Organized Village of Kwethluk" or "Organized Village of Saxman" or Orutsararmiut or Osage or Otoe-Missouria or Paiute or Paiute-Shoshone or Pala or Pamunkey or Pascua or Paskenta or Passamaquoddy or "Pauloff Harbor Village" or Pauma or Pawnee or Pechanga or "Pechanga " or "Peck Indian Reservation" or "Pedro Bay Village" or Pee-Posh or Penobscot or Penutian or Perquot or "Pima People" or Pima-Maricopa or Poarch or Pokagon or Pomo or Ponca or "Portage Creek Village" or Potawatomi or Pottawatomi or Pueblo or Puyallup or "Qagan Tayagungin" or Qawalangin or Quapaw or Quechan or "Rampart Village" or Rappahannock or Resighini or "Sac & Fox" or "Sac and Fox" or "Saint George Island" or "Saint Paul Island" or Salish or Samish or Santee or Sauk-Suiattle or "Seldovia Village Tribe" or Seminole or Seneca or Seneca-Cayuga or Shinnecock or Shivwits or Shoshone or Shoshone-Bannock or Shoshone-Paiute or Siuslaw or Skagit or "Skagway Village" or Skokomish or "South Naknek Village" or "Stebbins Community Association" or Stillaguamish or Suquamish or Swinomish or "Takotna Village" or Tangirnaq or Tejon or "Telida Village" or Te-Moak or Thlopthlocco or "Tlingit & Haida" or "Tohono O’odham" or Tolowa or Tonawanda or Tonkawa or Tulalip or Tunica-Biloxi or Tuolumne or Tuscarora or "Twin Hills Village" or "Ugashik Village" or Umatilla or Umkumiut or Umpqua or Ute or "Utu Utu Gwaitu" or "Village of Alakanuk" or "Village of Anaktuvuk Pass" or "Village of Aniak" or "Village of Atmautluak" or "Village of Bill Moore’s Slough" or "Village of Chefornak" or "Village of Clarks Point" or "Village of Crooked Creek" or "Village of Dot Lake" or "Village of Iliamna" or "Village of Kalskag" or "Village of Kaltag" or "Village of Kotlik" or "Village of Lower Kalskag" or "Village of Ohogamiut" or "Village of Red Devil" or "Village of Sleetmute" or "Village of Solomon" or "Village of Stony River" or "Village of Venetie" or "Village of Wainwright" or Wailaki or Wampanoag or "Warm Springs Reservation" or Washoe or Willapa or Winnemucca or Wintun or Wiyot or "Wrangell Cooperative Association" or Yakama or Yakutat or Yaqui or Yavapai or Yokuts or Yuhaaviatam or Yuima or Yukian or Yukut or Yupiit or Zuni).ti,ab,kf,kw. [registered NA Tribes adapted from https://www.govinfo.gov/content/pkg/FR-2022-01-28/pdf/2022-01789.pdf]	24417

5	("Degexit'an" or "Yup'ik" or ("Bear River" adj2 (Indian or Indians or Tribe or Tribes or native or natives or nation or people*)) or Abenaki or Achumawi or Ahtna or "Akimel Oodham" or "Akimel O'odham" or "Akimel Ootham" or "Akimel O'otham" or Aleut* or Algonkian* or Alsea or Aluutiq or Apalachee or Arikara or Atakapa or Athabascan or Athabaskan or Athapascan or Athapaskan or Bannock or Bidai or Biloxi or Blackfeet or Blackfoot or Caddoan or Calice or Calusa or Carrizo or Cayuse or Chemakum or Cheraw or Cherokee or Chetco or Cheyenne or Chicora or Chilula or Chimariko or Clatskanie or Coahuiltecan or "Coeur d’Alene" or Costanoan or Cree or Croatan or Croatoan or Cusabo or Edisto or Eskaleut* or Eskimo or Eskimo-Aleut or Esselen or Eyak or "Fox and Sauk" or Gabrielino or "Gros Ventre" or Guale or Gwich'in or Halchidhoma or Hidatsa or Hitchiti or Hopi or Houma or Huichol* or Hupa or Ingalik or Inupiaq or Ioway or Iroquoi* or Jacome or Jano or Jeaga or Juaneno or Jumano or Kalapuya or Kansa or Karankawa or Karok or Kashaya or Kato or Kawaiisu or Kitanemuk or Kitsai or Klallam or Koasati or Kolchan or Konkow or Koyukon or Lakota* or Lassik or Laurentian or Lenape or "Lenni Lenape" or Mahican or Mandan or Maricopa or Massachusett or Mattole or Minisink or Missouria or Mohave or Mohawk or Mohican or Molalla or Multnomah or Munsee or Muscoge* or Naabeeho or Nakoda or Nanticoke or Natchez or Nauset or Navaho or Niantic or Nipmuc or Nisenan or Nongatl or Oconee or Ofo or Ohlone or Ojibw* or Okanagan or Oneida or Otoe or Palouse or Panamint or Papago or Patwin or "Pee Dee" or Pennacook or Pequot or Pima or Piro or Pocasset or Pocomtuc or Pokanoket or Poospatuck or Powhatan or Quechan or Quileute or Quinault or Quiripi or Salinan or Samich or Saponi or Serrano or Shasta or Shawnee or Siksikaitsitapi or Sinkyone or Siouan or Sioux or Snohomish or Snoqualmie or Squaxin or Suma or Susquehannock or Takelma or Tanacross or Tanana or Tataviam or Tawakoni or Tequesta or Tillamook or Timucua or Tlingit or "Tohono Oodham" or Tongva or Tsimshian or Tubatulabal or Tutelo or Tututni or Unalachtigo or Unami or Unangan or Unangas or Unkechaug or Waccamaw or Walapai or Wappo or Wasco or Whilkut or Wichita or Winnebago or Wintu or Wishram or Yahi or Yakima or Yamasee or Yana or Yaqu or Yuchi or Yuki or Yuma or Yupik or Yurok).ti,ab,kf,kw. [Additional NA Tribes and Groups]	13973

6	(Anishnawbek or "?Akisq'nuk" or "?aq'am" or "?Esdilagh" or "Abénakis" or "Akisq'nuk" or "aq'am" or "Che:k:tles7et'h'" or "Da'naxda'xw" or "Dzawada'enuxw" or "Esk'etemc" or "Gitga'at" or "Gitlaxt'aamiks" or "Gwa'Sala-Nakwaxda'xw" or "Gwich'in" or "Ka:'yu:'k't'h'" or "Kanyen'kehaka" or "Kapawe'no" or "K'atlodeeche" or "K'ómoks" or "Leq'á:mel" or "Lil'wat" or "Malécite" or "Mi'kmaq" or "Mowachaht/Muchalaht" or "Nisga'a" or "N'Quatqua" or "O'Chiese" or "Ononda'gega'" or "Onondowága'" or "Onyota'a:ka" or "Saik'uz" or "Sambaa K'e " or "Shxwhá:y" or "Shxw'ow'hamel" or "SIPEKNE'KATIK" or "Skaru:reh" or "Sq'éwlets" or "Stellat'en" or "Sts'ailes" or "Stz'uminus" or "T'it'q'et" or "Tla'amin" or "Tl'azt'en" or "Tl'etinqox" or "Tsal'alh" or "Tŝideldel" or "Ts'kw'aylaxw" or "T'Sou-ke" or "Ts'uubaa-asatx" or "Tsuut'ina" or "We'koqma'q" or "Wet'suwet'en" or "Xai'xais" or "Xaxli'p" or "Yunesit'in" or ((Acadia or Carmacks or Champagne or Creek or Delaware or Douglas or Montana or Similkameen) adj2 (Indian or Indians or Tribe or Tribes or native or natives or nation or people*)) or Aamjiwnaang or Abegweit or Abitibiwinni or Ahousaht or Ahtahkakoop or Aishihik or Aitchelitz or Aklavik or Algonquin* or Anicinape* or "Animakee Wa Zhing" or Anishina* or aqam or Atikamekw or Atjanasca* or Attawapiskat or Aundeck-Omni-Kaning or Batchewana or "Bearfoot Onondaga" or Beausoleil or "Behdzi Ahda*" or "Biigtigong Nishnaabeg" or "Biinjitiwaabik Zaaging Anishinaabek" or Bilijk or "Binche Whut'en" or "Bingwi Neyaashi Anishinaabek" or "Canupawakpa Dakota" or Cayuga or Chawathil or Cheam or "Cheslatta Carrier" or Chiniki or Chipew* or Cote or Couchiching or Cowessess or Cowichan or Cree or "Dakota Plains" or "Dakota Tipi" or "Dechi Laot'i" or Deline or Dena or Dene or Denesuline or "Deninu K'ue" or Ditidaht or "Dog Rib Rae" or Dokis or Eabametoong or Ehattesaht or Elsipogtog or Ermineskin or Esdilagh or Esgenoopetitj or Eskasoni or Esquimalt or Gameti or Gayogohono or Ginoogaming or Gitanmaax or Gitanyow or Gitsegukla or Gitwangak or Gitxaala or Glooscap or Gwawaenuk or "Gwichya Gwich'in" or Gwitchin or Hagwilget or Haisla or Halalt or Haudenosaunee or Heiltsuk or Hesquiaht or Hiawatha or Homalco or Hupacasath or Huu-ay-aht or Inninuwug or Innu or Innue or Innus or Inuvik or Iskatewizaagegan or Iskut or "Ka'a'gee Tu" or Kahkewistahaw or "Kasabonika Lake" or Kashechewan or Katzie or Kawacatoose or Kebaowek or Keeseekoose or Keeseekoowenin or Kee-Way-Win or "Kinistin Saulteaux" or Kinonjeoshtegon or Kispiox or Kitselas or Kitsumkalum or Kluane or Kutchin or Kwadacha or Kwakiutl or "Kwanlin Dun" or Kwaw-kwaw-Apilt or Kwiakah or "Kwikwasut'inuxw Haxwa'mis" or Kwikwetlem or "Lax Kw'alaams" or "Lheidli T'enneh" or "Liidlii Kue" or "Listuguj Mi'gmaq" or Lyackson or Magnetawan or Malahat or Maliseet or Mamalilikulla or Matachewan or Matsqui or Mattagami or M'Chigeeng or Membertou or Metepenagiag or Metlakatla or Miawpukek or Michipicoten or Micmacs or Millbrook or Mishkeegogamang or Mississauga* or Mitaanjigamiing or Mohawk* or Montagnais or Moosomin or Moravian or Mowachaht or Muchalaht or Munsee-Delaware or Muscowpetung or Muskoday or Muskowekwan or Musqueam or "Nacho Nyak Dun" or "Nadleh Whuten" or Naicatchewenin or "Nak'azdli Whut'en" or Nakota or Naotkamegwanning or Naskapi or Natoaganeg or Nee-Tahi-Buhn or Nehiyawak or Nekaneet or Neskantaga or Neskonlith or Nibinamik or Nigigoonsiminikaaning or Nipissing or Nishnaabeg or Nooaitch or Nuchatlaht or Nuxalk or Ochapowace or O-Chi-Chak-Ko-Sipi or Odanak or Ojibw* or Okanagan or Okanese or Okemasis or Oneida or Oromocto or Osoyoos or Pabineau or Pacheedaht or "Paqtnkek Mi'kmaw" or Pasqua or Pauingassi or Pauquachin or "Pays Plat" or Peguis or "Pehdzeh Ki" or Pekuakamiulnuatsh or Penelakut or Piapot or Piikani or Pikangikum or Pinaymootang or Potlotek or "Qalipu Mi'kmaq" or Qualicum or Quatsino or Sagamok or Sahgaiehcan or Samahquam or Saugeen or Saulteau or Saulteaux or Sechelt or Secwepemc* or Semiahmoo or Seneca or Shackan or Shamattawa or Shawanaga or Sheguiandah or Sheshegwaning or Shuswap or Siksika or Simpcw or Sioux or Siska or Skatin or Skawahlook or Skeetchestn or Skidegate or "Skin Tyee" or Skowkale or Skownan or Skuppah or Skwah or Snuneymuxw or Songhees or Soowahlie or Splatsin or Spuzzum or Squamish or Squiala or "Stswecem'c Xget'tem " or Sumas or "Ta'an Kwach'an" or Tagish or Tahltan or Takla or Tallcree or "Taykwa Tagamou" or "Teetl'it Gwich'in" or Temagami or Thessalon or Timiskaming or "Tk'emlúps te Secwépemc" or Tla-o-qui-aht or Tlatlasikwala or Tlingit or Tlowitsis or Tootinaowaziibeeng or Toquaht or "Tr'ondëk Hwëch'in" or Tsartlip or Tsawout or Tsawwassen or Tseshaht or Tseycum or "Ts'il Kaz Koh" or Tsleil-Waututh or Tuscarora or Tutchone or Tzeachten or Uchucklesaht or Ucluelet or Ulkatcho or Wabaseemoong or Wabauskang or Wagmatcook or Wahnapitae or "Wahpeton Dakota" or Wahsipekuk or Wapekeka or Wasagamack or Wasauksing or Waswanipi or Wawakapewin or Waywayseecappo or Webequie or Weenusk or Wemotaci or Wendat or "Wha Ti" or Whapmagoostui or "Whitecap Dakota" or Wikwemikong or Witset or Wolastoqiyik or Wuikinuxv or Wunnumin or "Wuskwi Sipihk" or Xatsull or "Xeni Gwet'in" or Yakweakwioose or Yekooche or "Yellow Quill" or Yellowknife or Yellowknives or Zhiibaahaasing).ti,ab,kf,kw. [Registered FNs adapted from https://fnp-ppn.aadnc-aandc.gc.ca/fnp/Main/Search/SearchFN.aspx?lang=eng]	8883

7	("Kwakwaka'wakw" or "Kwak'wala" or "L’uml’umuluts" or "Lheidli T'enneh " or "Nlaka'pamux" or "Nuxálk" or "Quw'utsun" or "Qwum’yiqun" or "S’amuna" or "Sm'algyax" or "St'at'imc" or "Stl'atl'imx" or "Sto:lo" or "Tl’ulpalus" or "Tsilhqot'in" or "T'Souke" or "Xa'xtsa" or (Labrador adj2 (Indian or Indians or Tribe or Tribes or native or natives or nation or people*)) or "Áa Tlein Kwáan" or Aivilingmiut or Algonkian or Algonquian or Algonquin or Anishinaabe or "Atlin people" or "Baffin Island" or "Bella Coola" or Burrard or Cheklesahht or Chilcotin or Chippew* or Clayoquot or Clemclemaluts or Comiaken or Comox or Coquitlam or Dakelh or Dane-zaa or "Deisleen Kwáan" or Dunne-za or Gitxsan or Gitxsanimaax or Guyohkohnyo or "Haad kil" or Halkomelem or Halqemeylem or Henaksiala or Hinupsum or Hunquminum or Hwlitsum or Hwulqwselu or Hydin or Iglulik or Iglulingmuit or Ininew or Inuinnait or Inuk or Inupiat or Inuvialuit or Iroquois or Kainai or Kaska or Kelsemeht or Khenipsen or Kilpahlas or Kimsquit or Kitamaat or Kitlinermiut or Kivallirmiut or Klahoose or Koksilah or Kootenay or Koskimo or Ktunaxa or Kwantlen or Kwatna or Kyuquot or Labradormiut or Laich-kwil-tach or Lamalch* or Lekwungen or Lillooet or Loucheaux or Lummi or Micmac or Mississaugas or Mohawk or Mowachaht-Muchalaht or Nadene or Na-Dene or Namgis or Nanaimo or Nanoose or Nequatque or Netsilik or Netsilingmiut or Niitsitapi or Nimpkish or Nootka or Nunavimiut or Nunavimmiut or Nuu-chah-nulth or Odawa or Ohiaht or Oji-cree or Onayotekaono or Onondaga or Onondowahgah or Onundagaono or Opetchesaht or Owekeeno or Passamaquoddy or Peigan or Petun or Plains-Cree or Popkum or Potawatomi or Quamichan or Sahtu or Salish or Sarcee or Scowlitz or "Seabird Island" or Secwepemc or Sekani or Semahquam or Shishalh or Sinixt or Ska-Ruh-Reh or Skway or Sliammon or Snaw-naw-as or Somena or Songish or Sooke or Stoney or Stuie or Tallheo or Tasttine or Teslin or Tlicho or Tsimshianic or Tununirmiut or Ungava or Wakashan or Weewaikai or Wewaykum or Wyandot or Yuculta).ti,ab,kf,kw. [Additional FN Tribes]	2157

8	or/1-2 [Indigenous North Americans, general]	35006

9	or/3-7 [Indigenous North Americans, specific]	47987

10	Climacteric/ or Menopause/ or Menopause, Premature/ or Perimenopause/ or Postmenopause/ or Premenopause/ or (Climacteric or Climacterium or Menopause or Menopaus* or "Menopause, Premature" or "climacterium praecox" or Perimenopause or Perimenopaus* or Postmenopause or Postmenopaus* or "Post menopaus*" or Post-menopaus* or Premenopause or Premenopause* or Pre-menopaus* or "Pre menopaus*" or Climacteric* or "Life Change*" or "Change of life").ti,ab,kf,kw. [Menopause]	132160

11	Hot Flashes/ or ("Hot Flashes" or "Hot Flash*" or "Hot Flush*" or "night sweat*" or "nocturnal hyperhidrosis" or "vasomotor symptom*" or VMS).ti,ab,kf,kw. [MP Symptoms]	12059

12	or/10-11 [Menopause set]	137965

13	(elder or elders).ti,ab,kf,kw. [elders]	20542

14	female/ or women/ or (female or woman or women).ti,ab,kf,kw. [women]	10412214

15	and/13-14 [women elders]	11384

16	Health Services, Indigenous/ or Traditional Medicine Practitioners/ or Shamanism/ or ("Health Services, Indigenous" or ((Indigenous or native or "first nation" or aborigin* or Indian) adj2 (health or healthcare or healing or medicine or practices)) or ethnomedicin* or "native healing" or Limpia or "indigenous healer" or medicineman or medicinemen or "medicine man" or "medicine men" or Curandero or "folk healer" or shaman* or shamen).ti,ab,kf,kw. [Indigenous medicine]	15458

17	Medicine, Traditional/ or Complementary Therapies/ or Herbal Medicine/ or (((Complementary or alternative or traditional) adj2 (medicine* or therap* or healing or treatment*)) or "Herbal Medicine" or herbalism or ((botanical or ethnobotanical or herb* or plant or plant-based) adj2 (medicine* or remedy or remedies or healing or therap* or treatment*)) or phyto-medicine or phytomedic* or "folk medicine" or "folk healer").ti,ab,kf,kw. [alternative medicine]	222230

18	North America/ or exp Canada/ or exp United States/ or ("North America" or Canada or Alberta or "British Columbia" or Manitoba or "New Brunswick" or "Newfoundland" or "Labrador" or "Northwest Territories" or "Nova Scotia" or Nunavut or Ontario or "Prince Edward Island" or Quebec or Saskatchewan or Yukon or "United States" or Appalachia* or "New England" or "Pacific States" or Alabama or Alaska or Arizona or Arkansas or California or Colorado or Connecticut or Delaware or Florida or Georgia or Hawaii or Idaho or Illinois or Indiana or Iowa or Kansas or Kentucky or Louisiana or Maine or Maryland or Massachusetts or Michigan or Minnesota or Mississippi or Missouri or Montana or Nebraska or Nevada or "New Hampshire" or "New Jersey" or "New Mexico" or "New York" or "North Carolina" or "North Dakota" or Ohio or Oklahoma or Oregon or Pennsylvania or "Rhode Island" or "South Carolina" or "South Dakota" or Tennessee or Texas or Utah or Vermont or Virginia or Washington or "West Virginia" or Wisconsin or Wyoming or "District of Columbia" or "Washington DC").ti,ab,kf,kw. [North America]	2180325

19	Women's Health Services/ or ((women* or woman* or female*) adj2 (health or healthcare or service* or clinic or clinics)).ti,ab,kf,kw. [Women's health services]	49567

20	("1989960" or "20841572" or "7253935" or "10714743" or "11400222" or "12974133" or "23760434" or "33902542").ui. [Exemplars]	8

21	and/8,12 [Indigenous North Americans, general + Menopause Set]	188

22	(and/9,12) not 21 [Indigenous North Americans, specific + Menopause Set]	57

23	(and/12,16) not (or/21-22) [Indigenous medicine + Menopause Set]	22

24	(15 and (or/8-9)) not (or/21-23) [women elders + Indigenous North Americans]	288

25	(19 and (or/8-9)) not (or/21-24) [Women's health services + indigenous North Americans]	341

26	(17 and (or/12,15) and 18) not (or/21-25) [alternative medicine + menopause/women elders + North America]	178

27	or/21-26 [FINAL SET]	1074

28	27 or 20 [final set finds exemplars]	1074

29	remove duplicates from 27	1074

30	limit 29 to ez=20230530-20241121	89

### Embase (Elsevier)

**Field codes:** ti = article title, ab = abstract, kw = keyword; /de = Index (descriptor) term ; /exp = explode index term; /sd = entry date.
**Proximity operator:** NEAR/# 
**Truncation:**  *

**Date**: 21 November 2024

| **No.** | **Query** | **Results** |
| --- | --- | --- |
| #27 | #26 AND [01-06-2023]/sd | 195 |
| #26 | #20 OR #21 OR #22 OR #23 OR #24 OR #25 | 1565 |
| #25 | #17 AND (#12 OR #15) AND #18 | 302 |
| #24 | #19 AND (#8 OR #9) | 424 |
| #23 | #15 AND (#8 OR #9) | 386 |
| #22 | #12 AND #16 | 66 |
| #21 | #9 AND #12 | 189 |
| #20 | #8 AND #12 | 319 |
| #19 | ((women* OR woman* OR female*) NEAR/2 (health OR healthcare OR service* OR clinic OR clinics)):ti,ab,kw | 55685 |
| #18 | 'north america'/de OR 'canada'/exp OR 'united states'/exp OR 'north america':ti,ab,kw OR canada:ti,ab,kw OR alberta:ti,ab,kw OR 'british columbia':ti,ab,kw OR manitoba:ti,ab,kw OR 'new brunswick':ti,ab,kw OR 'newfoundland':ti,ab,kw OR 'labrador':ti,ab,kw OR 'northwest territories':ti,ab,kw OR 'nova scotia':ti,ab,kw OR nunavut:ti,ab,kw OR ontario:ti,ab,kw OR 'prince edward island':ti,ab,kw OR quebec:ti,ab,kw OR saskatchewan:ti,ab,kw OR yukon:ti,ab,kw OR 'united states':ti,ab,kw OR appalachia*:ti,ab,kw OR 'new england':ti,ab,kw OR 'pacific states':ti,ab,kw OR alabama:ti,ab,kw OR alaska:ti,ab,kw OR arizona:ti,ab,kw OR arkansas:ti,ab,kw OR california:ti,ab,kw OR colorado:ti,ab,kw OR connecticut:ti,ab,kw OR delaware:ti,ab,kw OR florida:ti,ab,kw OR georgia:ti,ab,kw OR hawaii:ti,ab,kw OR idaho:ti,ab,kw OR illinois:ti,ab,kw OR indiana:ti,ab,kw OR iowa:ti,ab,kw OR kansas:ti,ab,kw OR kentucky:ti,ab,kw OR louisiana:ti,ab,kw OR maine:ti,ab,kw OR maryland:ti,ab,kw OR massachusetts:ti,ab,kw OR michigan:ti,ab,kw OR minnesota:ti,ab,kw OR mississippi:ti,ab,kw OR missouri:ti,ab,kw OR montana:ti,ab,kw OR nebraska:ti,ab,kw OR nevada:ti,ab,kw OR 'new hampshire':ti,ab,kw OR 'new jersey':ti,ab,kw OR 'new mexico':ti,ab,kw OR 'new york':ti,ab,kw OR 'north carolina':ti,ab,kw OR 'north dakota':ti,ab,kw OR ohio:ti,ab,kw OR oklahoma:ti,ab,kw OR oregon:ti,ab,kw OR pennsylvania:ti,ab,kw OR 'rhode island':ti,ab,kw OR 'south carolina':ti,ab,kw OR 'south dakota':ti,ab,kw OR tennessee:ti,ab,kw OR texas:ti,ab,kw OR utah:ti,ab,kw OR vermont:ti,ab,kw OR virginia:ti,ab,kw OR washington:ti,ab,kw OR 'west virginia':ti,ab,kw OR wisconsin:ti,ab,kw OR wyoming:ti,ab,kw OR 'district of columbia':ti,ab,kw OR 'washington dc':ti,ab,kw | 2305311 |
| #17 | 'traditional medicine'/de OR 'alternative medicine'/de OR 'herbal medicine'/de OR (((complementary OR alternative OR traditional) NEAR/2 (medicine* OR therap* OR healing OR treatment*)):ti,ab,kw) OR 'herbal medicine':ti,ab,kw OR herbalism:ti,ab,kw OR (((botanical OR ethnobotanical OR herb* OR plant OR 'plant based') NEAR/2 (medicine* OR remedy OR remedies OR healing OR therap* OR treatment*)):ti,ab,kw) OR 'phyto medicine':ti,ab,kw OR phytomedic*:ti,ab,kw OR 'folk medicine':ti,ab,kw OR 'folk healer':ti,ab,kw | 343292 |
| #16 | 'indigenous health care'/de OR 'native american medicine'/de OR 'traditional healer'/de OR 'health services, indigenous':ti,ab,kw OR (((indigenous OR native OR 'first nation' OR aborigin* OR indian) NEAR/2 (health OR healthcare OR healing OR medicine OR practices)):ti,ab,kw) OR ethnomedicin*:ti,ab,kw OR 'native healing':ti,ab,kw OR limpia:ti,ab,kw OR 'indigenous healer':ti,ab,kw OR medicineman:ti,ab,kw OR medicinemen:ti,ab,kw OR 'medicine man':ti,ab,kw OR 'medicine men':ti,ab,kw OR curandero:ti,ab,kw OR 'folk healer':ti,ab,kw OR shaman*:ti,ab,kw OR shamen:ti,ab,kw | 19902 |
| #15 | #13 AND #14 | 17143 |
| #14 | 'female'/de OR female:ti,ab,kw OR woman:ti,ab,kw OR women:ti,ab,kw | 13227490 |
| #13 | elder:ti,ab,kw OR elders:ti,ab,kw | 29442 |
| #12 | #10 OR #11 | 234794 |
| #11 | 'hot flush'/de OR 'hot flashes':ti,ab,kw OR 'hot flash*':ti,ab,kw OR 'hot flush*':ti,ab,kw OR 'night sweat*':ti,ab,kw OR 'nocturnal hyperhidrosis':ti,ab,kw OR 'vasomotor symptom*':ti,ab,kw OR vms:ti,ab,kw | 32729 |
| #10 | 'climacterium'/de OR 'menopause and climacterium'/de OR 'menopause'/de OR 'early menopause'/de OR 'postmenopause'/de OR 'premenopause'/de OR climacteric:ti,ab,kw OR climacterium:ti,ab,kw OR menopause:ti,ab,kw OR menopaus*:ti,ab,kw OR 'menopause, premature':ti,ab,kw OR 'climacterium praecox':ti,ab,kw OR perimenopause:ti,ab,kw OR perimenopaus*:ti,ab,kw OR postmenopause:ti,ab,kw OR postmenopaus*:ti,ab,kw OR 'post menopaus*':ti,ab,kw OR premenopause:ti,ab,kw OR premenopause*:ti,ab,kw OR 'pre menopaus*':ti,ab,kw OR climacteric*:ti,ab,kw OR 'life change*':ti,ab,kw OR 'change of life':ti,ab,kw | 213806 |
| #9 | #3 OR #4 OR #5 OR #6 OR #7 | 68819 |
| #8 | #1 OR #2 | 43965 |
| #7 | 'kwakwaka` wakw':ti,ab,kw OR 'kwak` wala':ti,ab,kw OR 'l` uml` umuluts':ti,ab,kw OR 'lheidli t` enneh':ti,ab,kw OR 'nlaka` pamux':ti,ab,kw OR 'nuxÃ¡lk':ti,ab,kw OR 'quw` utsun':ti,ab,kw OR 'qwum` yiqun':ti,ab,kw OR 's` amuna':ti,ab,kw OR 'sm` algyax':ti,ab,kw OR 'st` at` imc':ti,ab,kw OR 'stl` atl` imx':ti,ab,kw OR 'sto:lo':ti,ab,kw OR 'tl` ulpalus':ti,ab,kw OR 'tsilhqot` in':ti,ab,kw OR 't` souke':ti,ab,kw OR 'xa` xtsa':ti,ab,kw OR ((labrador NEAR/2 (indian OR indians OR tribe OR tribes OR native OR natives OR nation OR people*)):ti,ab,kw) OR 'Ã¡a tlein kwÃ¡an':ti,ab,kw OR aivilingmiut:ti,ab,kw OR algonkian:ti,ab,kw OR algonquian:ti,ab,kw OR algonquin:ti,ab,kw OR anishinaabe:ti,ab,kw OR 'atlin people':ti,ab,kw OR 'baffin island':ti,ab,kw OR 'bella coola':ti,ab,kw OR burrard:ti,ab,kw OR cheklesahht:ti,ab,kw OR chilcotin:ti,ab,kw OR chippew*:ti,ab,kw OR clayoquot:ti,ab,kw OR clemclemaluts:ti,ab,kw OR comiaken:ti,ab,kw OR comox:ti,ab,kw OR coquitlam:ti,ab,kw OR dakelh:ti,ab,kw OR 'dane zaa':ti,ab,kw OR 'deisleen kwÃ¡an':ti,ab,kw OR 'dunne za':ti,ab,kw OR gitxsan:ti,ab,kw OR gitxsanimaax:ti,ab,kw OR guyohkohnyo:ti,ab,kw OR 'haad kil':ti,ab,kw OR halkomelem:ti,ab,kw OR halqemeylem:ti,ab,kw OR henaksiala:ti,ab,kw OR hinupsum:ti,ab,kw OR hunquminum:ti,ab,kw OR hwlitsum:ti,ab,kw OR hwulqwselu:ti,ab,kw OR hydin:ti,ab,kw OR iglulik:ti,ab,kw OR iglulingmuit:ti,ab,kw OR ininew:ti,ab,kw OR inuinnait:ti,ab,kw OR inuk:ti,ab,kw OR inupiat:ti,ab,kw OR inuvialuit:ti,ab,kw OR iroquois:ti,ab,kw OR kainai:ti,ab,kw OR kaska:ti,ab,kw OR kelsemeht:ti,ab,kw OR khenipsen:ti,ab,kw OR kilpahlas:ti,ab,kw OR kimsquit:ti,ab,kw OR kitamaat:ti,ab,kw OR kitlinermiut:ti,ab,kw OR kivallirmiut:ti,ab,kw OR klahoose:ti,ab,kw OR koksilah:ti,ab,kw OR kootenay:ti,ab,kw OR koskimo:ti,ab,kw OR ktunaxa:ti,ab,kw OR kwantlen:ti,ab,kw OR kwatna:ti,ab,kw OR kyuquot:ti,ab,kw OR labradormiut:ti,ab,kw OR 'laich kwil tach':ti,ab,kw OR lamalch*:ti,ab,kw OR lekwungen:ti,ab,kw OR lillooet:ti,ab,kw OR loucheaux:ti,ab,kw OR lummi:ti,ab,kw OR micmac:ti,ab,kw OR mississaugas:ti,ab,kw OR mohawk:ti,ab,kw OR 'mowachaht muchalaht':ti,ab,kw OR nadene:ti,ab,kw OR 'na dene':ti,ab,kw OR namgis:ti,ab,kw OR nanaimo:ti,ab,kw OR nanoose:ti,ab,kw OR nequatque:ti,ab,kw OR netsilik:ti,ab,kw OR netsilingmiut:ti,ab,kw OR niitsitapi:ti,ab,kw OR nimpkish:ti,ab,kw OR nootka:ti,ab,kw OR nunavimiut:ti,ab,kw OR nunavimmiut:ti,ab,kw OR 'nuu chah nulth':ti,ab,kw OR odawa:ti,ab,kw OR ohiaht:ti,ab,kw OR 'oji cree':ti,ab,kw OR onayotekaono:ti,ab,kw OR onondaga:ti,ab,kw OR onondowahgah:ti,ab,kw OR onundagaono:ti,ab,kw OR opetchesaht:ti,ab,kw OR owekeeno:ti,ab,kw OR passamaquoddy:ti,ab,kw OR peigan:ti,ab,kw OR petun:ti,ab,kw OR 'plains cree':ti,ab,kw OR popkum:ti,ab,kw OR potawatomi:ti,ab,kw OR quamichan:ti,ab,kw OR sahtu:ti,ab,kw OR salish:ti,ab,kw OR sarcee:ti,ab,kw OR scowlitz:ti,ab,kw OR 'seabird island':ti,ab,kw OR secwepemc:ti,ab,kw OR sekani:ti,ab,kw OR semahquam:ti,ab,kw OR shishalh:ti,ab,kw OR sinixt:ti,ab,kw OR 'ska ruh reh':ti,ab,kw OR skway:ti,ab,kw OR sliammon:ti,ab,kw OR 'snaw naw as':ti,ab,kw OR somena:ti,ab,kw OR songish:ti,ab,kw OR sooke:ti,ab,kw OR stoney:ti,ab,kw OR stuie:ti,ab,kw OR tallheo:ti,ab,kw OR tasttine:ti,ab,kw OR teslin:ti,ab,kw OR tlicho:ti,ab,kw OR tsimshianic:ti,ab,kw OR tununirmiut:ti,ab,kw OR ungava:ti,ab,kw OR wakashan:ti,ab,kw OR weewaikai:ti,ab,kw OR wewaykum:ti,ab,kw OR wyandot:ti,ab,kw OR yuculta:ti,ab,kw | 2430 |
| #6 | anishnawbek:ti,ab,kw OR '?akisq` nuk':ti,ab,kw OR '?aq` am':ti,ab,kw OR '?esdilagh':ti,ab,kw OR 'abÃ©nakis':ti,ab,kw OR 'akisq` nuk':ti,ab,kw OR 'aq` am':ti,ab,kw OR 'che:k:tles7et` h`':ti,ab,kw OR 'da` naxda` xw':ti,ab,kw OR 'dzawada` enuxw':ti,ab,kw OR 'esk` etemc':ti,ab,kw OR 'gitga` at':ti,ab,kw OR 'gitlaxt` aamiks':ti,ab,kw OR 'gwa` sala-nakwaxda` xw':ti,ab,kw OR 'gwich` in':ti,ab,kw OR 'ka:` yu:` k` t` h`':ti,ab,kw OR 'kanyen` kehaka':ti,ab,kw OR 'kapawe` no':ti,ab,kw OR 'k` atlodeeche':ti,ab,kw OR 'k` Ã³moks':ti,ab,kw OR 'leq` Ã¡:mel':ti,ab,kw OR 'lil` wat':ti,ab,kw OR 'malÃ©cite':ti,ab,kw OR 'mi` kmaq':ti,ab,kw OR 'mowachaht/muchalaht':ti,ab,kw OR 'nisga` a':ti,ab,kw OR 'n` quatqua':ti,ab,kw OR 'o` chiese':ti,ab,kw OR 'ononda` gega`':ti,ab,kw OR 'onondowÃ¡ga`':ti,ab,kw OR 'onyota` a:ka':ti,ab,kw OR 'saik` uz':ti,ab,kw OR 'sambaa k` e':ti,ab,kw OR 'shxwhÃ¡:y':ti,ab,kw OR 'shxw` ow` hamel':ti,ab,kw OR 'sipekne` katik':ti,ab,kw OR 'skaru:reh':ti,ab,kw OR 'sq` Ã©wlets':ti,ab,kw OR 'stellat` en':ti,ab,kw OR 'sts` ailes':ti,ab,kw OR 'stz` uminus':ti,ab,kw OR 't` it` q` et':ti,ab,kw OR 'tla` amin':ti,ab,kw OR 'tl` azt` en':ti,ab,kw OR 'tl` etinqox':ti,ab,kw OR 'tsal` alh':ti,ab,kw OR 'tÅideldel':ti,ab,kw OR 'ts` kw` aylaxw':ti,ab,kw OR 't` sou-ke':ti,ab,kw OR 'ts` uubaa-asatx':ti,ab,kw OR 'tsuut` ina':ti,ab,kw OR 'we` koqma` q':ti,ab,kw OR 'wet` suwet` en':ti,ab,kw OR 'xai` xais':ti,ab,kw OR 'xaxli` p':ti,ab,kw OR 'yunesit` in':ti,ab,kw OR (((acadia OR carmacks OR champagne OR creek OR delaware OR douglas OR montana OR similkameen) NEAR/2 (indian OR indians OR tribe OR tribes OR native OR natives OR nation OR people*)):ti,ab,kw) OR aamjiwnaang:ti,ab,kw OR abegweit:ti,ab,kw OR abitibiwinni:ti,ab,kw OR ahousaht:ti,ab,kw OR ahtahkakoop:ti,ab,kw OR aishihik:ti,ab,kw OR aitchelitz:ti,ab,kw OR aklavik:ti,ab,kw OR algonquin*:ti,ab,kw OR anicinape*:ti,ab,kw OR 'animakee wa zhing':ti,ab,kw OR anishina*:ti,ab,kw OR aqam:ti,ab,kw OR atikamekw:ti,ab,kw OR atjanasca*:ti,ab,kw OR attawapiskat:ti,ab,kw OR 'aundeck omni kaning':ti,ab,kw OR batchewana:ti,ab,kw OR 'bearfoot onondaga':ti,ab,kw OR beausoleil:ti,ab,kw OR 'behdzi ahda*':ti,ab,kw OR 'biigtigong nishnaabeg':ti,ab,kw OR 'biinjitiwaabik zaaging anishinaabek':ti,ab,kw OR bilijk:ti,ab,kw OR 'binche whut` en':ti,ab,kw OR 'bingwi neyaashi anishinaabek':ti,ab,kw OR 'canupawakpa dakota':ti,ab,kw OR cayuga:ti,ab,kw OR chawathil:ti,ab,kw OR cheam:ti,ab,kw OR 'cheslatta carrier':ti,ab,kw OR chiniki:ti,ab,kw OR chipew*:ti,ab,kw OR cote:ti,ab,kw OR couchiching:ti,ab,kw OR cowessess:ti,ab,kw OR cowichan:ti,ab,kw OR cree:ti,ab,kw OR 'dakota plains':ti,ab,kw OR 'dakota tipi':ti,ab,kw OR 'dechi laot` i':ti,ab,kw OR deline:ti,ab,kw OR dena:ti,ab,kw OR dene:ti,ab,kw OR denesuline:ti,ab,kw OR 'deninu k` ue':ti,ab,kw OR ditidaht:ti,ab,kw OR 'dog rib rae':ti,ab,kw OR dokis:ti,ab,kw OR eabametoong:ti,ab,kw OR ehattesaht:ti,ab,kw OR elsipogtog:ti,ab,kw OR ermineskin:ti,ab,kw OR esdilagh:ti,ab,kw OR esgenoopetitj:ti,ab,kw OR eskasoni:ti,ab,kw OR esquimalt:ti,ab,kw OR gameti:ti,ab,kw OR gayogohono:ti,ab,kw OR ginoogaming:ti,ab,kw OR gitanmaax:ti,ab,kw OR gitanyow:ti,ab,kw OR gitsegukla:ti,ab,kw OR gitwangak:ti,ab,kw OR gitxaala:ti,ab,kw OR glooscap:ti,ab,kw OR gwawaenuk:ti,ab,kw OR 'gwichya gwich` in':ti,ab,kw OR gwitchin:ti,ab,kw OR hagwilget:ti,ab,kw OR haisla:ti,ab,kw OR halalt:ti,ab,kw OR haudenosaunee:ti,ab,kw OR heiltsuk:ti,ab,kw OR hesquiaht:ti,ab,kw OR hiawatha:ti,ab,kw OR homalco:ti,ab,kw OR hupacasath:ti,ab,kw OR 'huu ay aht':ti,ab,kw OR inninuwug:ti,ab,kw OR innu:ti,ab,kw OR innue:ti,ab,kw OR innus:ti,ab,kw OR inuvik:ti,ab,kw OR iskatewizaagegan:ti,ab,kw OR iskut:ti,ab,kw OR 'ka` a` gee tu':ti,ab,kw OR kahkewistahaw:ti,ab,kw OR 'kasabonika lake':ti,ab,kw OR kashechewan:ti,ab,kw OR katzie:ti,ab,kw OR kawacatoose:ti,ab,kw OR kebaowek:ti,ab,kw OR keeseekoose:ti,ab,kw OR keeseekoowenin:ti,ab,kw OR 'kee way win':ti,ab,kw OR 'kinistin saulteaux':ti,ab,kw OR kinonjeoshtegon:ti,ab,kw OR kispiox:ti,ab,kw OR kitselas:ti,ab,kw OR kitsumkalum:ti,ab,kw OR kluane:ti,ab,kw OR kutchin:ti,ab,kw OR kwadacha:ti,ab,kw OR kwakiutl:ti,ab,kw OR 'kwanlin dun':ti,ab,kw OR 'kwaw kwaw apilt':ti,ab,kw OR kwiakah:ti,ab,kw OR 'kwikwasut` inuxw haxwa` mis':ti,ab,kw OR kwikwetlem:ti,ab,kw OR 'lax kw` alaams':ti,ab,kw OR 'lheidli t` enneh':ti,ab,kw OR 'liidlii kue':ti,ab,kw OR 'listuguj mi` gmaq':ti,ab,kw OR lyackson:ti,ab,kw OR magnetawan:ti,ab,kw OR malahat:ti,ab,kw OR maliseet:ti,ab,kw OR mamalilikulla:ti,ab,kw OR matachewan:ti,ab,kw OR matsqui:ti,ab,kw OR mattagami:ti,ab,kw OR 'm` chigeeng':ti,ab,kw OR membertou:ti,ab,kw OR metepenagiag:ti,ab,kw OR metlakatla:ti,ab,kw OR miawpukek:ti,ab,kw OR michipicoten:ti,ab,kw OR micmacs:ti,ab,kw OR millbrook:ti,ab,kw OR mishkeegogamang:ti,ab,kw OR mississauga*:ti,ab,kw OR mitaanjigamiing:ti,ab,kw OR mohawk*:ti,ab,kw OR montagnais:ti,ab,kw OR moosomin:ti,ab,kw OR moravian:ti,ab,kw OR mowachaht:ti,ab,kw OR muchalaht:ti,ab,kw OR 'munsee delaware':ti,ab,kw OR muscowpetung:ti,ab,kw OR muskoday:ti,ab,kw OR muskowekwan:ti,ab,kw OR musqueam:ti,ab,kw OR 'nacho nyak dun':ti,ab,kw OR 'nadleh whuten':ti,ab,kw OR naicatchewenin:ti,ab,kw OR 'nak` azdli whut` en':ti,ab,kw OR nakota:ti,ab,kw OR naotkamegwanning:ti,ab,kw OR naskapi:ti,ab,kw OR natoaganeg:ti,ab,kw OR 'nee tahi buhn':ti,ab,kw OR nehiyawak:ti,ab,kw OR nekaneet:ti,ab,kw OR neskantaga:ti,ab,kw OR neskonlith:ti,ab,kw OR nibinamik:ti,ab,kw OR nigigoonsiminikaaning:ti,ab,kw OR nipissing:ti,ab,kw OR nishnaabeg:ti,ab,kw OR nooaitch:ti,ab,kw OR nuchatlaht:ti,ab,kw OR nuxalk:ti,ab,kw OR ochapowace:ti,ab,kw OR 'o chi chak ko sipi':ti,ab,kw OR odanak:ti,ab,kw OR ojibw*:ti,ab,kw OR okanagan:ti,ab,kw OR okanese:ti,ab,kw OR okemasis:ti,ab,kw OR oneida:ti,ab,kw OR oromocto:ti,ab,kw OR osoyoos:ti,ab,kw OR pabineau:ti,ab,kw OR pacheedaht:ti,ab,kw OR 'paqtnkek mi` kmaw':ti,ab,kw OR pasqua:ti,ab,kw OR pauingassi:ti,ab,kw OR pauquachin:ti,ab,kw OR 'pays plat':ti,ab,kw OR peguis:ti,ab,kw OR 'pehdzeh ki':ti,ab,kw OR pekuakamiulnuatsh:ti,ab,kw OR penelakut:ti,ab,kw OR piapot:ti,ab,kw OR piikani:ti,ab,kw OR pikangikum:ti,ab,kw OR pinaymootang:ti,ab,kw OR potlotek:ti,ab,kw OR 'qalipu mi` kmaq':ti,ab,kw OR qualicum:ti,ab,kw OR quatsino:ti,ab,kw OR sagamok:ti,ab,kw OR sahgaiehcan:ti,ab,kw OR samahquam:ti,ab,kw OR saugeen:ti,ab,kw OR saulteau:ti,ab,kw OR saulteaux:ti,ab,kw OR sechelt:ti,ab,kw OR secwepemc*:ti,ab,kw OR semiahmoo:ti,ab,kw OR seneca:ti,ab,kw OR shackan:ti,ab,kw OR shamattawa:ti,ab,kw OR shawanaga:ti,ab,kw OR sheguiandah:ti,ab,kw OR sheshegwaning:ti,ab,kw OR shuswap:ti,ab,kw OR siksika:ti,ab,kw OR simpcw:ti,ab,kw OR sioux:ti,ab,kw OR siska:ti,ab,kw OR skatin:ti,ab,kw OR skawahlook:ti,ab,kw OR skeetchestn:ti,ab,kw OR skidegate:ti,ab,kw OR 'skin tyee':ti,ab,kw OR skowkale:ti,ab,kw OR skownan:ti,ab,kw OR skuppah:ti,ab,kw OR skwah:ti,ab,kw OR snuneymuxw:ti,ab,kw OR songhees:ti,ab,kw OR soowahlie:ti,ab,kw OR splatsin:ti,ab,kw OR spuzzum:ti,ab,kw OR squamish:ti,ab,kw OR squiala:ti,ab,kw OR 'stswecem` c xget` tem':ti,ab,kw OR sumas:ti,ab,kw OR 'ta` an kwach` an':ti,ab,kw OR tagish:ti,ab,kw OR tahltan:ti,ab,kw OR takla:ti,ab,kw OR tallcree:ti,ab,kw OR 'taykwa tagamou':ti,ab,kw OR 'teetl` it gwich` in':ti,ab,kw OR temagami:ti,ab,kw OR thessalon:ti,ab,kw OR timiskaming:ti,ab,kw OR 'tk` emlÃºps te secwÃ©pemc':ti,ab,kw OR 'tla o qui aht':ti,ab,kw OR tlatlasikwala:ti,ab,kw OR tlingit:ti,ab,kw OR tlowitsis:ti,ab,kw OR tootinaowaziibeeng:ti,ab,kw OR toquaht:ti,ab,kw OR 'tr` ondÃ«k hwÃ«ch` in':ti,ab,kw OR tsartlip:ti,ab,kw OR tsawout:ti,ab,kw OR tsawwassen:ti,ab,kw OR tseshaht:ti,ab,kw OR tseycum:ti,ab,kw OR 'ts` il kaz koh':ti,ab,kw OR 'tsleil waututh':ti,ab,kw OR tuscarora:ti,ab,kw OR tutchone:ti,ab,kw OR tzeachten:ti,ab,kw OR uchucklesaht:ti,ab,kw OR ucluelet:ti,ab,kw OR ulkatcho:ti,ab,kw OR wabaseemoong:ti,ab,kw OR wabauskang:ti,ab,kw OR wagmatcook:ti,ab,kw OR wahnapitae:ti,ab,kw OR 'wahpeton dakota':ti,ab,kw OR wahsipekuk:ti,ab,kw OR wapekeka:ti,ab,kw OR wasagamack:ti,ab,kw OR wasauksing:ti,ab,kw OR waswanipi:ti,ab,kw OR wawakapewin:ti,ab,kw OR waywayseecappo:ti,ab,kw OR webequie:ti,ab,kw OR weenusk:ti,ab,kw OR wemotaci:ti,ab,kw OR wendat:ti,ab,kw OR 'wha ti':ti,ab,kw OR whapmagoostui:ti,ab,kw OR 'whitecap dakota':ti,ab,kw OR wikwemikong:ti,ab,kw OR witset:ti,ab,kw OR wolastoqiyik:ti,ab,kw OR wuikinuxv:ti,ab,kw OR wunnumin:ti,ab,kw OR 'wuskwi sipihk':ti,ab,kw OR xatsull:ti,ab,kw OR 'xeni gwet` in':ti,ab,kw OR yakweakwioose:ti,ab,kw OR yekooche:ti,ab,kw OR 'yellow quill':ti,ab,kw OR yellowknife:ti,ab,kw OR yellowknives:ti,ab,kw OR zhiibaahaasing:ti,ab,kw | 10107 |
| #5 | 'degexit` an':ti,ab,kw OR 'yup` ik':ti,ab,kw OR (('bear river' NEAR/2 (indian OR indians OR tribe OR tribes OR native OR natives OR nation OR people*)):ti,ab,kw) OR abenaki:ti,ab,kw OR achumawi:ti,ab,kw OR ahtna:ti,ab,kw OR 'akimel oodham':ti,ab,kw OR 'akimel o` odham':ti,ab,kw OR 'akimel ootham':ti,ab,kw OR 'akimel o` otham':ti,ab,kw OR aleut*:ti,ab,kw OR algonkian*:ti,ab,kw OR alsea:ti,ab,kw OR aluutiq:ti,ab,kw OR apalachee:ti,ab,kw OR arikara:ti,ab,kw OR atakapa:ti,ab,kw OR athabascan:ti,ab,kw OR athabaskan:ti,ab,kw OR athapascan:ti,ab,kw OR athapaskan:ti,ab,kw OR bannock:ti,ab,kw OR bidai:ti,ab,kw OR biloxi:ti,ab,kw OR blackfeet:ti,ab,kw OR blackfoot:ti,ab,kw OR caddoan:ti,ab,kw OR calice:ti,ab,kw OR calusa:ti,ab,kw OR carrizo:ti,ab,kw OR cayuse:ti,ab,kw OR chemakum:ti,ab,kw OR cheraw:ti,ab,kw OR cherokee:ti,ab,kw OR chetco:ti,ab,kw OR cheyenne:ti,ab,kw OR chicora:ti,ab,kw OR chilula:ti,ab,kw OR chimariko:ti,ab,kw OR clatskanie:ti,ab,kw OR coahuiltecan:ti,ab,kw OR 'coeur d` alene':ti,ab,kw OR costanoan:ti,ab,kw OR cree:ti,ab,kw OR croatan:ti,ab,kw OR croatoan:ti,ab,kw OR cusabo:ti,ab,kw OR edisto:ti,ab,kw OR eskaleut*:ti,ab,kw OR eskimo:ti,ab,kw OR 'eskimo aleut':ti,ab,kw OR esselen:ti,ab,kw OR eyak:ti,ab,kw OR 'fox and sauk':ti,ab,kw OR gabrielino:ti,ab,kw OR 'gros ventre':ti,ab,kw OR guale:ti,ab,kw OR 'gwich` in':ti,ab,kw OR halchidhoma:ti,ab,kw OR hidatsa:ti,ab,kw OR hitchiti:ti,ab,kw OR hopi:ti,ab,kw OR houma:ti,ab,kw OR huichol*:ti,ab,kw OR hupa:ti,ab,kw OR ingalik:ti,ab,kw OR inupiaq:ti,ab,kw OR ioway:ti,ab,kw OR iroquoi*:ti,ab,kw OR jacome:ti,ab,kw OR jano:ti,ab,kw OR jeaga:ti,ab,kw OR juaneno:ti,ab,kw OR jumano:ti,ab,kw OR kalapuya:ti,ab,kw OR kansa:ti,ab,kw OR karankawa:ti,ab,kw OR karok:ti,ab,kw OR kashaya:ti,ab,kw OR kato:ti,ab,kw OR kawaiisu:ti,ab,kw OR kitanemuk:ti,ab,kw OR kitsai:ti,ab,kw OR klallam:ti,ab,kw OR koasati:ti,ab,kw OR kolchan:ti,ab,kw OR konkow:ti,ab,kw OR koyukon:ti,ab,kw OR lakota*:ti,ab,kw OR lassik:ti,ab,kw OR laurentian:ti,ab,kw OR lenape:ti,ab,kw OR 'lenni lenape':ti,ab,kw OR mahican:ti,ab,kw OR mandan:ti,ab,kw OR maricopa:ti,ab,kw OR massachusett:ti,ab,kw OR mattole:ti,ab,kw OR minisink:ti,ab,kw OR missouria:ti,ab,kw OR mohave:ti,ab,kw OR mohawk:ti,ab,kw OR mohican:ti,ab,kw OR molalla:ti,ab,kw OR multnomah:ti,ab,kw OR munsee:ti,ab,kw OR muscoge*:ti,ab,kw OR naabeeho:ti,ab,kw OR nakoda:ti,ab,kw OR nanticoke:ti,ab,kw OR natchez:ti,ab,kw OR nauset:ti,ab,kw OR navaho:ti,ab,kw OR niantic:ti,ab,kw OR nipmuc:ti,ab,kw OR nisenan:ti,ab,kw OR nongatl:ti,ab,kw OR oconee:ti,ab,kw OR ofo:ti,ab,kw OR ohlone:ti,ab,kw OR ojibw*:ti,ab,kw OR okanagan:ti,ab,kw OR oneida:ti,ab,kw OR otoe:ti,ab,kw OR palouse:ti,ab,kw OR panamint:ti,ab,kw OR papago:ti,ab,kw OR patwin:ti,ab,kw OR 'pee dee':ti,ab,kw OR pennacook:ti,ab,kw OR pequot:ti,ab,kw OR pima:ti,ab,kw OR piro:ti,ab,kw OR pocasset:ti,ab,kw OR pocomtuc:ti,ab,kw OR pokanoket:ti,ab,kw OR poospatuck:ti,ab,kw OR powhatan:ti,ab,kw OR quechan:ti,ab,kw OR quileute:ti,ab,kw OR quinault:ti,ab,kw OR quiripi:ti,ab,kw OR salinan:ti,ab,kw OR samich:ti,ab,kw OR saponi:ti,ab,kw OR serrano:ti,ab,kw OR shasta:ti,ab,kw OR shawnee:ti,ab,kw OR siksikaitsitapi:ti,ab,kw OR sinkyone:ti,ab,kw OR siouan:ti,ab,kw OR sioux:ti,ab,kw OR snohomish:ti,ab,kw OR snoqualmie:ti,ab,kw OR squaxin:ti,ab,kw OR suma:ti,ab,kw OR susquehannock:ti,ab,kw OR takelma:ti,ab,kw OR tanacross:ti,ab,kw OR tanana:ti,ab,kw OR tataviam:ti,ab,kw OR tawakoni:ti,ab,kw OR tequesta:ti,ab,kw OR tillamook:ti,ab,kw OR timucua:ti,ab,kw OR tlingit:ti,ab,kw OR 'tohono oodham':ti,ab,kw OR tongva:ti,ab,kw OR tsimshian:ti,ab,kw OR tubatulabal:ti,ab,kw OR tutelo:ti,ab,kw OR tututni:ti,ab,kw OR unalachtigo:ti,ab,kw OR unami:ti,ab,kw OR unangan:ti,ab,kw OR unangas:ti,ab,kw OR unkechaug:ti,ab,kw OR waccamaw:ti,ab,kw OR walapai:ti,ab,kw OR wappo:ti,ab,kw OR wasco:ti,ab,kw OR whilkut:ti,ab,kw OR wichita:ti,ab,kw OR winnebago:ti,ab,kw OR wintu:ti,ab,kw OR wishram:ti,ab,kw OR yahi:ti,ab,kw OR yakima:ti,ab,kw OR yamasee:ti,ab,kw OR yana:ti,ab,kw OR yaqu:ti,ab,kw OR yuchi:ti,ab,kw OR yuki:ti,ab,kw OR yuma:ti,ab,kw OR yupik:ti,ab,kw OR yurok:ti,ab,kw | 16860 |
| #4 | 'asa` carsarmiut':ti,ab,kw OR 'dee-ni`':ti,ab,kw OR 'mi` kmaq':ti,ab,kw OR 's` klallam':ti,ab,kw OR 'shoonaq`':ti,ab,kw OR 'sun` aq':ti,ab,kw OR ((('ivanof bay' OR 'shoalwater bay' OR 'spirit lake' OR 'squaxin island' OR dakota OR dakotah OR wichita OR keechi OR waco OR tawakonie OR cheyenne OR creek OR crow OR hoonah OR kasigluk OR maricopa OR miami OR omaha OR oneida OR ottawa OR peoria OR salamatof OR sitka OR snoqualmie OR spokane OR winnebago OR wyandotte) NEAR/2 (indian OR indians OR tribe OR tribes OR native OR natives OR nation OR people*)):ti,ab,kw) OR (((native OR indian OR traditional) NEAR/2 (village OR council OR community OR association)):ti,ab,kw) OR 'absentee shawnee':ti,ab,kw OR achomawi:ti,ab,kw OR agdaagux:ti,ab,kw OR 'agua caliente':ti,ab,kw OR ajumawi:ti,ab,kw OR 'ak chin':ti,ab,kw OR akiachak:ti,ab,kw OR akiak:ti,ab,kw OR 'alabama coushatta':ti,ab,kw OR 'alabama quassarte':ti,ab,kw OR alatna:ti,ab,kw OR algaaciq:ti,ab,kw OR algin:ti,ab,kw OR allakaket:ti,ab,kw OR 'alturas indian rancheria':ti,ab,kw OR alutiiq:ti,ab,kw OR angoon:ti,ab,kw OR anvik:ti,ab,kw OR apache:ti,ab,kw OR aporige:ti,ab,kw OR aquinnah:ti,ab,kw OR arapaho:ti,ab,kw OR aroostook:ti,ab,kw OR assiniboine:ti,ab,kw OR astarawi:ti,ab,kw OR atsugewi:ti,ab,kw OR atwamsini:ti,ab,kw OR bdewakantunwan:ti,ab,kw OR caddo:ti,ab,kw OR cahto:ti,ab,kw OR cahuilla:ti,ab,kw OR catawba:ti,ab,kw OR cayuga:ti,ab,kw OR chalkyitsik:ti,ab,kw OR 'cheesh na':ti,ab,kw OR chehalis:ti,ab,kw OR chemehuevi:ti,ab,kw OR 'cher ae':ti,ab,kw OR chevak:ti,ab,kw OR 'cheyenne arapaho':ti,ab,kw OR chickahominy:ti,ab,kw OR chickaloon:ti,ab,kw OR chickasaw:ti,ab,kw OR chignik:ti,ab,kw OR chilkat:ti,ab,kw OR chilkoot:ti,ab,kw OR chinik:ti,ab,kw OR chinook:ti,ab,kw OR chippewa*:ti,ab,kw OR chitimacha:ti,ab,kw OR choctaw:ti,ab,kw OR chukchansi:ti,ab,kw OR chuloonawick:ti,ab,kw OR chumash:ti,ab,kw OR cocopah:ti,ab,kw OR colusa:ti,ab,kw OR comanche:ti,ab,kw OR coos:ti,ab,kw OR cosmit:ti,ab,kw OR coushatta:ti,ab,kw OR cowlitz:ti,ab,kw OR 'cross village':ti,ab,kw OR cupeno:ti,ab,kw OR curyung:ti,ab,kw OR diegueno:ti,ab,kw OR dine`:ti,ab,kw OR egegik:ti,ab,kw OR eklutna:ti,ab,kw OR elwha:ti,ab,kw OR 'emmonak village':ti,ab,kw OR 'evansville village':ti,ab,kw OR ewiiaapaayp:ti,ab,kw OR 'galena village':ti,ab,kw OR goshute:ti,ab,kw OR 'gulkana village council':ti,ab,kw OR haida:ti,ab,kw OR hammawi:ti,ab,kw OR havasupai:ti,ab,kw OR 'healy lake village':ti,ab,kw OR hewisedawi:ti,ab,kw OR 'ho chunk':ti,ab,kw OR hoh:ti,ab,kw OR hokan:ti,ab,kw OR holikachuk:ti,ab,kw OR 'holy cross tribe':ti,ab,kw OR hualapai:ti,ab,kw OR 'hughes village':ti,ab,kw OR 'huslia village':ti,ab,kw OR 'hydaburg cooperative association':ti,ab,kw OR 'igiugig village':ti,ab,kw OR iipay:ti,ab,kw OR ilmawi:ti,ab,kw OR inaja:ti,ab,kw OR inupiat:ti,ab,kw OR ione:ti,ab,kw OR itsatawi:ti,ab,kw OR 'kaguyak village':ti,ab,kw OR kaibab:ti,ab,kw OR 'kaktovik village':ti,ab,kw OR kalispel:ti,ab,kw OR kanosh:ti,ab,kw OR karuk:ti,ab,kw OR kashia:ti,ab,kw OR kaw:ti,ab,kw OR keetoowah:ti,ab,kw OR 'kenaitze indian tribe':ti,ab,kw OR 'ketchikan indian community':ti,ab,kw OR 'keweenaw bay':ti,ab,kw OR kialegee:ti,ab,kw OR kickapoo:ti,ab,kw OR 'king island native community':ti,ab,kw OR 'king salmon tribe':ti,ab,kw OR kiowa:ti,ab,kw OR klamath:ti,ab,kw OR 'klawock cooperative association':ti,ab,kw OR kletsel:ti,ab,kw OR 'knik tribe':ti,ab,kw OR 'koi nation':ti,ab,kw OR 'kokhanok village':ti,ab,kw OR koosharem:ti,ab,kw OR kootenai:ti,ab,kw OR kosalektawi:ti,ab,kw OR 'koyukuk native village':ti,ab,kw OR kumeyaay:ti,ab,kw OR kwalhioqua:ti,ab,kw OR 'lesnoi village':ti,ab,kw OR 'levelock village':ti,ab,kw OR 'lime village':ti,ab,kw OR 'louden village':ti,ab,kw OR luiseno:ti,ab,kw OR lummi:ti,ab,kw OR madesi:ti,ab,kw OR maidu:ti,ab,kw OR makah:ti,ab,kw OR maliseet:ti,ab,kw OR 'manley hot springs village':ti,ab,kw OR 'manokotak village':ti,ab,kw OR manzanita:ti,ab,kw OR mashantucket:ti,ab,kw OR 'match e be nash she wish':ti,ab,kw OR mattaponi:ti,ab,kw OR 'mcgrath native village':ti,ab,kw OR mdewakanton:ti,ab,kw OR mechoopda:ti,ab,kw OR menominee:ti,ab,kw OR 'me wuk':ti,ab,kw OR miccosukee:ti,ab,kw OR micmacs:ti,ab,kw OR 'mission indians':ti,ab,kw OR miwok:ti,ab,kw OR miwuk:ti,ab,kw OR modoc:ti,ab,kw OR mohegan:ti,ab,kw OR mojave:ti,ab,kw OR monacan:ti,ab,kw OR 'mono indians':ti,ab,kw OR muckleshoot:ti,ab,kw OR muscogee:ti,ab,kw OR 'na denÃ©':ti,ab,kw OR nansemond:ti,ab,kw OR narragansett:ti,ab,kw OR navajo:ti,ab,kw OR 'new koliganek village':ti,ab,kw OR 'new stuyahok village':ti,ab,kw OR 'newhalen village':ti,ab,kw OR 'newtok village':ti,ab,kw OR 'nez perce':ti,ab,kw OR 'nikolai village':ti,ab,kw OR 'ninilchik village':ti,ab,kw OR nisqually:ti,ab,kw OR nomlaki:ti,ab,kw OR 'nondalton village':ti,ab,kw OR nooksack:ti,ab,kw OR 'northway village':ti,ab,kw OR nottawaseppi:ti,ab,kw OR 'nulato village':ti,ab,kw OR nunakauyarmiut:ti,ab,kw OR odawa:ti,ab,kw OR ohgsenakale:ti,ab,kw OR 'ohkay owingeh':ti,ab,kw OR onondaga:ti,ab,kw OR oreilles:ti,ab,kw OR 'organized village of grayling':ti,ab,kw OR 'organized village of kake':ti,ab,kw OR 'organized village of kasaan':ti,ab,kw OR 'organized village of kwethluk':ti,ab,kw OR 'organized village of saxman':ti,ab,kw OR orutsararmiut:ti,ab,kw OR osage:ti,ab,kw OR 'otoe missouria':ti,ab,kw OR paiute:ti,ab,kw OR 'paiute shoshone':ti,ab,kw OR pala:ti,ab,kw OR pamunkey:ti,ab,kw OR pascua:ti,ab,kw OR paskenta:ti,ab,kw OR passamaquoddy:ti,ab,kw OR 'pauloff harbor village':ti,ab,kw OR pauma:ti,ab,kw OR pawnee:ti,ab,kw OR pechanga:ti,ab,kw OR 'pechanga':ti,ab,kw OR 'peck indian reservation':ti,ab,kw OR 'pedro bay village':ti,ab,kw OR 'pee posh':ti,ab,kw OR penobscot:ti,ab,kw OR penutian:ti,ab,kw OR perquot:ti,ab,kw OR 'pima people':ti,ab,kw OR 'pima maricopa':ti,ab,kw OR poarch:ti,ab,kw OR pokagon:ti,ab,kw OR pomo:ti,ab,kw OR ponca:ti,ab,kw OR 'portage creek village':ti,ab,kw OR potawatomi:ti,ab,kw OR pottawatomi:ti,ab,kw OR pueblo:ti,ab,kw OR puyallup:ti,ab,kw OR 'qagan tayagungin':ti,ab,kw OR qawalangin:ti,ab,kw OR quapaw:ti,ab,kw OR quechan:ti,ab,kw OR 'rampart village':ti,ab,kw OR rappahannock:ti,ab,kw OR resighini:ti,ab,kw OR 'sac & fox':ti,ab,kw OR 'sac and fox':ti,ab,kw OR 'saint george island':ti,ab,kw OR 'saint paul island':ti,ab,kw OR salish:ti,ab,kw OR samish:ti,ab,kw OR santee:ti,ab,kw OR 'sauk suiattle':ti,ab,kw OR 'seldovia village tribe':ti,ab,kw OR seminole:ti,ab,kw OR seneca:ti,ab,kw OR 'seneca cayuga':ti,ab,kw OR shinnecock:ti,ab,kw OR shivwits:ti,ab,kw OR shoshone:ti,ab,kw OR 'shoshone bannock':ti,ab,kw OR 'shoshone paiute':ti,ab,kw OR siuslaw:ti,ab,kw OR skagit:ti,ab,kw OR 'skagway village':ti,ab,kw OR skokomish:ti,ab,kw OR 'south naknek village':ti,ab,kw OR 'stebbins community association':ti,ab,kw OR stillaguamish:ti,ab,kw OR suquamish:ti,ab,kw OR swinomish:ti,ab,kw OR 'takotna village':ti,ab,kw OR tangirnaq:ti,ab,kw OR tejon:ti,ab,kw OR 'telida village':ti,ab,kw OR 'te moak':ti,ab,kw OR thlopthlocco:ti,ab,kw OR 'tlingit & haida':ti,ab,kw OR 'tohono o` odham':ti,ab,kw OR tolowa:ti,ab,kw OR tonawanda:ti,ab,kw OR tonkawa:ti,ab,kw OR tulalip:ti,ab,kw OR 'tunica biloxi':ti,ab,kw OR tuolumne:ti,ab,kw OR tuscarora:ti,ab,kw OR 'twin hills village':ti,ab,kw OR 'ugashik village':ti,ab,kw OR umatilla:ti,ab,kw OR umkumiut:ti,ab,kw OR umpqua:ti,ab,kw OR ute:ti,ab,kw OR 'utu utu gwaitu':ti,ab,kw OR 'village of alakanuk':ti,ab,kw OR 'village of anaktuvuk pass':ti,ab,kw OR 'village of aniak':ti,ab,kw OR 'village of atmautluak':ti,ab,kw OR 'village of bill moore` s slough':ti,ab,kw OR 'village of chefornak':ti,ab,kw OR 'village of clarks point':ti,ab,kw OR 'village of crooked creek':ti,ab,kw OR 'village of dot lake':ti,ab,kw OR 'village of iliamna':ti,ab,kw OR 'village of kalskag':ti,ab,kw OR 'village of kaltag':ti,ab,kw OR 'village of kotlik':ti,ab,kw OR 'village of lower kalskag':ti,ab,kw OR 'village of ohogamiut':ti,ab,kw OR 'village of red devil':ti,ab,kw OR 'village of sleetmute':ti,ab,kw OR 'village of solomon':ti,ab,kw OR 'village of stony river':ti,ab,kw OR 'village of venetie':ti,ab,kw OR 'village of wainwright':ti,ab,kw OR wailaki:ti,ab,kw OR wampanoag:ti,ab,kw OR 'warm springs reservation':ti,ab,kw OR washoe:ti,ab,kw OR willapa:ti,ab,kw OR winnemucca:ti,ab,kw OR wintun:ti,ab,kw OR wiyot:ti,ab,kw OR 'wrangell cooperative association':ti,ab,kw OR yakama:ti,ab,kw OR yakutat:ti,ab,kw OR yaqui:ti,ab,kw OR yavapai:ti,ab,kw OR yokuts:ti,ab,kw OR yuhaaviatam:ti,ab,kw OR yuima:ti,ab,kw OR yukian:ti,ab,kw OR yukut:ti,ab,kw OR yupiit:ti,ab,kw OR zuni:ti,ab,kw | 39424 |
| #3 | 'aleut (people)'/de OR 'algonkian people'/de OR 'algonquin (people)'/de OR 'apache (people)'/de OR 'arikara (people)'/de OR 'athabaskan (people)'/de OR 'caddoan people'/de OR 'cherokee (people)'/de OR 'chippewa (people)'/de OR 'choctaw (people)'/de OR 'cree (people)'/de OR 'eskimo'/de OR 'eskimo-aleut people'/de OR 'haida (people)'/de OR 'hopi (people)'/de OR 'inuit'/de OR 'inupiat (people)'/de OR 'iroquois (people)'/de OR 'lakota (people)'/de OR 'maya (people)'/de OR 'mayo (people)'/de OR 'mazahua (people)'/de OR 'mohawk (people)'/de OR 'muscogee (people)'/de OR 'na-dene people'/de OR 'navajo (people)'/de OR 'ojibway (people)'/de OR 'oji-cree (people)'/de OR 'penutian people'/de OR 'pima (people)'/de OR 'seminole (people)'/de OR 'sioux (people)'/de OR 'tlingit (people)'/de OR 'tohono o`odham'/de OR 'uto-aztecan people'/de OR 'white mountain apache'/de OR 'yaqui (people)'/de OR 'yupik (people)'/de OR 'zuni (people)'/de | 6279 |
| #2 | 'first nations'/de OR 'canadian aboriginal'/de OR 'metis'/de OR (((canada OR canadian*) NEAR/2 (native* OR indigenous OR aborigin*)):ti,ab,kw) OR metis:ti,ab,kw OR 'mÃ©tis':ti,ab,kw OR michif*:ti,ab,kw OR mitchif:ti,ab,kw OR mechif:ti,ab,kw OR 'mÃ©tif':ti,ab,kw OR 'mÃ©tchif':ti,ab,kw OR metif:ti,ab,kw OR metchif:ti,ab,kw OR inuit:ti,ab,kw OR 'first nations':ti,ab,kw OR 'first nation':ti,ab,kw | 6771 |
| #1 | 'american indian'/de OR 'alaska native'/de OR 'amerind people'/de OR 'north-central amerind people'/de OR (((american* OR alaska*) NEAR/2 (native* OR indian OR indians OR indigenous OR aborigin*)):ti,ab,kw) OR amerind*:ti,ab,kw | 38261 |

### CINAHL Complete (EBSCOhost)

**Field codes:** TI = title, AB= abstract, MH = CINAHL Exact Subject Headings, EM = Entry Date
**Proximity operator:** N# 
**Truncation:**  *

**Database** - CINAHL Complete

**Interface** - EBSCOhost Research Databases

**Search Screen** - Advanced Search

**Date** - Thu, November 21, 2024 2:27:00 PM

| **#** | **Query** | **Limiters/Expanders** | **Results** |
| --- | --- | --- | --- |
| S27 | S26 AND EM 20230509- |  | 66 |
| S26 | S24 NOT S25 |  | 992 |
| S25 | S19 OR S20 OR S21 OR S22 OR S23 | Limiters - Publication Type: Book Review, Brief Item, Obituary, Poetry | 7 |
| S24 | S19 OR S20 OR S21 OR S22 OR S23 |  | 999 |
| S23 | S16 AND (S11 OR S14) AND S17 |  | 155 |
| S22 | S18 AND S8 |  | 521 |
| S21 | S14 AND S8 |  | 206 |
| S20 | S15 AND S11 |  | 24 |
| S19 | S8 AND S11 |  | 160 |
| S18 | MH("Women's Health Services") OR TI((women* OR woman* OR female*) N2 (health OR healthcare OR service* OR clinic OR clinics)) OR AB((women* OR woman* OR female*) N2 (health OR healthcare OR service* OR clinic OR clinics)) |  | 40328 |
| S17 | MH("North America" OR "Canada+" OR "United States+") OR TI("North America" OR Canada OR Alberta OR "British Columbia" OR Manitoba OR "New Brunswick" OR "Newfoundland" OR "Labrador" OR "Northwest Territories" OR "Nova Scotia" OR Nunavut OR Ontario OR "Prince Edward Island" OR Quebec OR Saskatchewan OR Yukon OR "United States" OR Appalachia* OR "New England" OR "Pacific States" OR Alabama OR Alaska OR Arizona OR Arkansas OR California OR Colorado OR Connecticut OR Delaware OR Florida OR Georgia OR Hawaii OR Idaho OR Illinois OR Indiana OR Iowa OR Kansas OR Kentucky OR Louisiana OR Maine OR Maryland OR Massachusetts OR Michigan OR Minnesota OR Mississippi OR Missouri OR Montana OR Nebraska OR Nevada OR "New Hampshire" OR "New Jersey" OR "New Mexico" OR "New York" OR "North Carolina" OR "North Dakota" OR Ohio OR Oklahoma OR Oregon OR Pennsylvania OR "Rhode Island" OR "South Carolina" OR "South Dakota" OR Tennessee OR Texas OR Utah OR Vermont OR Virginia OR Washington OR "West Virginia" OR Wisconsin OR Wyoming OR "District of Columbia" OR "Washington DC") OR AB("North America" OR Canada OR Alberta OR "British Columbia" OR Manitoba OR "New Brunswick" OR "Newfoundland" OR "Labrador" OR "Northwest Territories" OR "Nova Scotia" OR Nunavut OR Ontario OR "Prince Edward Island" OR Quebec OR Saskatchewan OR Yukon OR "United States" OR Appalachia* OR "New England" OR "Pacific States" OR Alabama OR Alaska OR Arizona OR Arkansas OR California OR Colorado OR Connecticut OR Delaware OR Florida OR Georgia OR Hawaii OR Idaho OR Illinois OR Indiana OR Iowa OR Kansas OR Kentucky OR Louisiana OR Maine OR Maryland OR Massachusetts OR Michigan OR Minnesota OR Mississippi OR Missouri OR Montana OR Nebraska OR Nevada OR "New Hampshire" OR "New Jersey" OR "New Mexico" OR "New York" OR "North Carolina" OR "North Dakota" OR Ohio OR Oklahoma OR Oregon OR Pennsylvania OR "Rhode Island" OR "South Carolina" OR "South Dakota" OR Tennessee OR Texas OR Utah OR Vermont OR Virginia OR Washington OR "West Virginia" OR Wisconsin OR Wyoming OR "District of Columbia" OR "Washington DC") |  | 1041824 |
| S16 | MH("Medicine, Traditional" OR "Herbal Medicine" OR "Alternative Therapies") OR TI(((Complementary OR alternative OR traditional) N2 (medicine* OR therap* OR healing OR treatment*)) OR "Herbal Medicine" OR herbalism OR ((botanical OR ethnobotanical OR herb* OR plant OR plant-based) N2 (medicine* OR remedy OR remedies OR healing OR therap* OR treatment*)) OR phyto-medicine OR phytomedic* OR "folk medicine" OR "folk healer") OR AB(((Complementary OR alternative OR traditional) N2 (medicine* OR therap* OR healing OR treatment*)) OR "Herbal Medicine" OR herbalism OR ((botanical OR ethnobotanical OR herb* OR plant OR plant-based) N2 (medicine* OR remedy OR remedies OR healing OR therap* OR treatment*)) OR phyto-medicine OR phytomedic* OR "folk medicine" OR "folk healer") |  | 94100 |
| S15 | MH("Medicine, Native American Traditional" OR "Health Services, Indigenous" OR "Traditional Healers" OR Shamanism) OR TI(((Indigenous OR native OR "first nation" OR aborigin* OR Indian) N2 (health OR healthcare OR healing OR medicine OR practices)) OR ethnomedicin* OR "native healing" OR Limpia OR "indigenous healer" OR medicineman OR medicinemen OR "medicine man" OR "medicine men" OR Curandero OR "folk healer" OR shaman* OR shamen) OR AB(((Indigenous OR native OR "first nation" OR aborigin* OR Indian) N2 (health OR healthcare OR healing OR medicine OR practices)) OR ethnomedicin* OR "native healing" OR Limpia OR "indigenous healer" OR medicineman OR medicinemen OR "medicine man" OR "medicine men" OR Curandero OR "folk healer" OR shaman* OR shamen) |  | 10394 |
| S14 | S12 AND S13 |  | 5521 |
| S13 | MH(female or "women+") OR TI(female OR woman OR women) OR AB(female OR woman OR women) |  | 2455028 |
| S12 | TI(elder OR elders) OR AB(elder OR elders) |  | 12544 |
| S11 | S9 OR S10 |  | 45756 |
| S10 | MH("Hot Flashes") OR TI("Hot Flashes" OR "Hot Flash*" OR "Hot Flush*" OR "night sweat*" OR "nocturnal hyperhidrosis" OR "vasomotor symptom*" OR VMS) OR AB("Hot Flashes" OR "Hot Flash*" OR "Hot Flush*" OR "night sweat*" OR "nocturnal hyperhidrosis" OR "vasomotor symptom*" OR VMS) |  | 5291 |
| S9 | MH(Climacteric OR Menopause OR "Menopause, Premature" OR Perimenopause OR Postmenopause OR Premenopause) OR TI(Climacteric OR Climacterium OR Menopause OR Menopaus* OR "Menopause, Premature" OR "climacterium praecox" OR Perimenopause OR Perimenopaus* OR Postmenopause OR Postmenopaus* OR "Post menopaus*" OR Post-menopaus* OR Premenopause OR Premenopause* OR Pre-menopaus* OR "Pre menopaus*" OR Climacteric* OR "Life Change*" OR "Change of life") OR AB(Climacteric OR Climacterium OR Menopause OR Menopaus* OR "Menopause, Premature" OR "climacterium praecox" OR Perimenopause OR Perimenopaus* OR Postmenopause OR Postmenopaus* OR "Post menopaus*" OR Post-menopaus* OR Premenopause OR Premenopause* OR Pre-menopaus* OR "Pre menopaus*" OR Climacteric* OR "Life Change*" OR "Change of life") |  | 43637 |
| S8 | S1 OR S2 OR S3 OR S4 OR S5 OR S6 OR S7 |  | 36813 |
| S7 | TI("Kwakwaka'wakw" OR "Kwak'wala" OR "L'uml'umuluts" OR "Lheidli T'enneh " OR "Nlaka'pamux" OR "Nuxálk" OR "Quw'utsun" OR "Qwum'yiqun" OR "S'amuna" OR "Sm'algyax" OR "St'at'imc" OR "Stl'atl'imx" OR "Sto:lo" OR "Tl'ulpalus" OR "Tsilhqot'in" OR "T'Souke" OR "Xa'xtsa" OR (Labrador N2 (Indian OR Indians OR Tribe OR Tribes OR native OR natives OR nation OR people*)) OR "Áa Tlein Kwáan" OR Aivilingmiut OR Algonkian OR Algonquian OR Algonquin OR Anishinaabe OR "Atlin people" OR "Baffin Island" OR "Bella Coola" OR Burrard OR Cheklesahht OR Chilcotin OR Chippew* OR Clayoquot OR Clemclemaluts OR Comiaken OR Comox OR Coquitlam OR Dakelh OR Dane-zaa OR "Deisleen Kwáan" OR Dunne-za OR Gitxsan OR Gitxsanimaax OR Guyohkohnyo OR "Haad kil" OR Halkomelem OR Halqemeylem OR Henaksiala OR Hinupsum OR Hunquminum OR Hwlitsum OR Hwulqwselu OR Hydin OR Iglulik OR Iglulingmuit OR Ininew OR Inuinnait OR Inuk OR Inupiat OR Inuvialuit OR Iroquois OR Kainai OR Kaska OR Kelsemeht OR Khenipsen OR Kilpahlas OR Kimsquit OR Kitamaat OR Kitlinermiut OR Kivallirmiut OR Klahoose OR Koksilah OR Kootenay OR Koskimo OR Ktunaxa OR Kwantlen OR Kwatna OR Kyuquot OR Labradormiut OR Laich-kwil-tach OR Lamalch* OR Lekwungen OR Lillooet OR Loucheaux OR Lummi OR Micmac OR Mississaugas OR Mohawk OR Mowachaht-Muchalaht OR Nadene OR Na-Dene OR Namgis OR Nanaimo OR Nanoose OR Nequatque OR Netsilik OR Netsilingmiut OR Niitsitapi OR Nimpkish OR Nootka OR Nunavimiut OR Nunavimmiut OR Nuu-chah-nulth OR Odawa OR Ohiaht OR Oji-cree OR Onayotekaono OR Onondaga OR Onondowahgah OR Onundagaono OR Opetchesaht OR Owekeeno OR Passamaquoddy OR Peigan OR Petun OR Plains-Cree OR Popkum OR Potawatomi OR Quamichan OR Sahtu OR Salish OR Sarcee OR Scowlitz OR "Seabird Island" OR Secwepemc OR Sekani OR Semahquam OR Shishalh OR Sinixt OR Ska-Ruh-Reh OR Skway OR Sliammon OR Snaw-naw-as OR Somena OR Songish OR Sooke OR Stoney OR Stuie OR Tallheo OR Tasttine OR Teslin OR Tlicho OR Tsimshianic OR Tununirmiut OR Ungava OR Wakashan OR Weewaikai OR Wewaykum OR Wyandot OR Yuculta) OR AB("Kwakwaka'wakw" OR "Kwak'wala" OR "L'uml'umuluts" OR "Lheidli T'enneh " OR "Nlaka'pamux" OR "Nuxálk" OR "Quw'utsun" OR "Qwum'yiqun" OR "S'amuna" OR "Sm'algyax" OR "St'at'imc" OR "Stl'atl'imx" OR "Sto:lo" OR "Tl'ulpalus" OR "Tsilhqot'in" OR "T'Souke" OR "Xa'xtsa" OR (Labrador N2 (Indian OR Indians OR Tribe OR Tribes OR native OR natives OR nation OR people*)) OR "Áa Tlein Kwáan" OR Aivilingmiut OR Algonkian OR Algonquian OR Algonquin OR Anishinaabe OR "Atlin people" OR "Baffin Island" OR "Bella Coola" OR Burrard OR Cheklesahht OR Chilcotin OR Chippew* OR Clayoquot OR Clemclemaluts OR Comiaken OR Comox OR Coquitlam OR Dakelh OR Dane-zaa OR "Deisleen Kwáan" OR Dunne-za OR Gitxsan OR Gitxsanimaax OR Guyohkohnyo OR "Haad kil" OR Halkomelem OR Halqemeylem OR Henaksiala OR Hinupsum OR Hunquminum OR Hwlitsum OR Hwulqwselu OR Hydin OR Iglulik OR Iglulingmuit OR Ininew OR Inuinnait OR Inuk OR Inupiat OR Inuvialuit OR Iroquois OR Kainai OR Kaska OR Kelsemeht OR Khenipsen OR Kilpahlas OR Kimsquit OR Kitamaat OR Kitlinermiut OR Kivallirmiut OR Klahoose OR Koksilah OR Kootenay OR Koskimo OR Ktunaxa OR Kwantlen OR Kwatna OR Kyuquot OR Labradormiut OR Laich-kwil-tach OR Lamalch* OR Lekwungen OR Lillooet OR Loucheaux OR Lummi OR Micmac OR Mississaugas OR Mohawk OR Mowachaht-Muchalaht OR Nadene OR Na-Dene OR Namgis OR Nanaimo OR Nanoose OR Nequatque OR Netsilik OR Netsilingmiut OR Niitsitapi OR Nimpkish OR Nootka OR Nunavimiut OR Nunavimmiut OR Nuu-chah-nulth OR Odawa OR Ohiaht OR Oji-cree OR Onayotekaono OR Onondaga OR Onondowahgah OR Onundagaono OR Opetchesaht OR Owekeeno OR Passamaquoddy OR Peigan OR Petun OR Plains-Cree OR Popkum OR Potawatomi OR Quamichan OR Sahtu OR Salish OR Sarcee OR Scowlitz OR "Seabird Island" OR Secwepemc OR Sekani OR Semahquam OR Shishalh OR Sinixt OR Ska-Ruh-Reh OR Skway OR Sliammon OR Snaw-naw-as OR Somena OR Songish OR Sooke OR Stoney OR Stuie OR Tallheo OR Tasttine OR Teslin OR Tlicho OR Tsimshianic OR Tununirmiut OR Ungava OR Wakashan OR Weewaikai OR Wewaykum OR Wyandot OR Yuculta) |  | 388 |
| S6 | TI(Anishnawbek OR "?Akisq'nuk" OR "?aq'am" OR "?Esdilagh" OR "Abénakis" OR "Akisq'nuk" OR "aq'am" OR "Che:k:tles7et'h'" OR "Da'naxda'xw" OR "Dzawada'enuxw" OR "Esk'etemc" OR "Gitga'at" OR "Gitlaxt'aamiks" OR "Gwa'Sala-Nakwaxda'xw" OR "Gwich'in" OR "Ka:'yu:'k't'h'" OR "Kanyen'kehaka" OR "Kapawe'no" OR "K'atlodeeche" OR "K'ómoks" OR "Leq'á:mel" OR "Lil'wat" OR "Malécite" OR "Mi'kmaq" OR "Mowachaht/Muchalaht" OR "Nisga'a" OR "N'Quatqua" OR "O'Chiese" OR "Ononda'gega'" OR "Onondowága'" OR "Onyota'a:ka" OR "Saik'uz" OR "Sambaa K'e " OR "Shxwhá:y" OR "Shxw'ow'hamel" OR "SIPEKNE'KATIK" OR "Skaru:reh" OR "Sq'éwlets" OR "Stellat'en" OR "Sts'ailes" OR "Stz'uminus" OR "T'it'q'et" OR "Tla'amin" OR "Tl'azt'en" OR "Tl'etinqox" OR "Tsal'alh" OR "Tsideldel" OR "Ts'kw'aylaxw" OR "T'Sou-ke" OR "Ts'uubaa-asatx" OR "Tsuut'ina" OR "We'koqma'q" OR "Wet'suwet'en" OR "Xai'xais" OR "Xaxli'p" OR "Yunesit'in" OR ((Acadia OR Carmacks OR Champagne OR Creek OR Delaware OR Douglas OR Montana OR Similkameen) N2 (Indian OR Indians OR Tribe OR Tribes OR native OR natives OR nation OR people*)) OR Aamjiwnaang OR Abegweit OR Abitibiwinni OR Ahousaht OR Ahtahkakoop OR Aishihik OR Aitchelitz OR Aklavik OR Algonquin* OR Anicinape* OR "Animakee Wa Zhing" OR Anishina* OR aqam OR Atikamekw OR Atjanasca* OR Attawapiskat OR Aundeck-Omni-Kaning OR Batchewana OR "Bearfoot Onondaga" OR Beausoleil OR "Behdzi Ahda*" OR "Biigtigong Nishnaabeg" OR "Biinjitiwaabik Zaaging Anishinaabek" OR Bilijk OR "Binche Whut'en" OR "Bingwi Neyaashi Anishinaabek" OR "Canupawakpa Dakota" OR Cayuga OR Chawathil OR Cheam OR "Cheslatta Carrier" OR Chiniki OR Chipew* OR Cote OR Couchiching OR Cowessess OR Cowichan OR Cree OR "Dakota Plains" OR "Dakota Tipi" OR "Dechi Laot'i" OR Deline OR Dena OR Dene OR Denesuline OR "Deninu K'ue" OR Ditidaht OR "Dog Rib Rae" OR Dokis OR Eabametoong OR Ehattesaht OR Elsipogtog OR Ermineskin OR Esdilagh OR Esgenoopetitj OR Eskasoni OR Esquimalt OR Gameti OR Gayogohono OR Ginoogaming OR Gitanmaax OR Gitanyow OR Gitsegukla OR Gitwangak OR Gitxaala OR Glooscap OR Gwawaenuk OR "Gwichya Gwich'in" OR Gwitchin OR Hagwilget OR Haisla OR Halalt OR Haudenosaunee OR Heiltsuk OR Hesquiaht OR Hiawatha OR Homalco OR Hupacasath OR Huu-ay-aht OR Inninuwug OR Innu OR Innue OR Innus OR Inuvik OR Iskatewizaagegan OR Iskut OR "Ka'a'gee Tu" OR Kahkewistahaw OR "Kasabonika Lake" OR Kashechewan OR Katzie OR Kawacatoose OR Kebaowek OR Keeseekoose OR Keeseekoowenin OR Kee-Way-Win OR "Kinistin Saulteaux" OR Kinonjeoshtegon OR Kispiox OR Kitselas OR Kitsumkalum OR Kluane OR Kutchin OR Kwadacha OR Kwakiutl OR "Kwanlin Dun" OR Kwaw-kwaw-Apilt OR Kwiakah OR "Kwikwasut'inuxw Haxwa'mis" OR Kwikwetlem OR "Lax Kw'alaams" OR "Lheidli T'enneh" OR "Liidlii Kue" OR "Listuguj Mi'gmaq" OR Lyackson OR Magnetawan OR Malahat OR Maliseet OR Mamalilikulla OR Matachewan OR Matsqui OR Mattagami OR "M'Chigeeng" OR Membertou OR Metepenagiag OR Metlakatla OR Miawpukek OR Michipicoten OR Micmacs OR Millbrook OR Mishkeegogamang OR Mississauga* OR Mitaanjigamiing OR Mohawk* OR Montagnais OR Moosomin OR Moravian OR Mowachaht OR Muchalaht OR Munsee-Delaware OR Muscowpetung OR Muskoday OR Muskowekwan OR Musqueam OR "Nacho Nyak Dun" OR "Nadleh Whuten" OR Naicatchewenin OR "Nak'azdli Whut'en" OR Nakota OR Naotkamegwanning OR Naskapi OR Natoaganeg OR Nee-Tahi-Buhn OR Nehiyawak OR Nekaneet OR Neskantaga OR Neskonlith OR Nibinamik OR Nigigoonsiminikaaning OR Nipissing OR Nishnaabeg OR Nooaitch OR Nuchatlaht OR Nuxalk OR Ochapowace OR O-Chi-Chak-Ko-Sipi OR Odanak OR Ojibw* OR Okanagan OR Okanese OR Okemasis OR Oneida OR Oromocto OR Osoyoos OR Pabineau OR Pacheedaht OR "Paqtnkek Mi'kmaw" OR Pasqua OR Pauingassi OR Pauquachin OR "Pays Plat" OR Peguis OR "Pehdzeh Ki" OR Pekuakamiulnuatsh OR Penelakut OR Piapot OR Piikani OR Pikangikum OR Pinaymootang OR Potlotek OR "Qalipu Mi'kmaq" OR Qualicum OR Quatsino OR Sagamok OR Sahgaiehcan OR Samahquam OR Saugeen OR Saulteau OR Saulteaux OR Sechelt OR Secwepemc* OR Semiahmoo OR Seneca OR Shackan OR Shamattawa OR Shawanaga OR Sheguiandah OR Sheshegwaning OR Shuswap OR Siksika OR Simpcw OR Sioux OR Siska OR Skatin OR Skawahlook OR Skeetchestn OR Skidegate OR "Skin Tyee" OR Skowkale OR Skownan OR Skuppah OR Skwah OR Snuneymuxw OR Songhees OR Soowahlie OR Splatsin OR Spuzzum OR Squamish OR Squiala OR "Stswecem'c Xget'tem" OR Sumas OR "Ta'an Kwach'an" OR Tagish OR Tahltan OR Takla OR Tallcree OR "Taykwa Tagamou" OR "Teetl'it Gwich'in" OR Temagami OR Thessalon OR Timiskaming OR "Tk'emlúps te Secwépemc" OR Tla-o-qui-aht OR Tlatlasikwala OR Tlingit OR Tlowitsis OR Tootinaowaziibeeng OR Toquaht OR "Tr'ondëk Hwëch'in" OR Tsartlip OR Tsawout OR Tsawwassen OR Tseshaht OR Tseycum OR "Ts'il Kaz Koh" OR Tsleil-Waututh OR Tuscarora OR Tutchone OR Tzeachten OR Uchucklesaht OR Ucluelet OR Ulkatcho OR Wabaseemoong OR Wabauskang OR Wagmatcook OR Wahnapitae OR "Wahpeton Dakota" OR Wahsipekuk OR Wapekeka OR Wasagamack OR Wasauksing OR Waswanipi OR Wawakapewin OR Waywayseecappo OR Webequie OR Weenusk OR Wemotaci OR Wendat OR "Wha Ti" OR Whapmagoostui OR "Whitecap Dakota" OR Wikwemikong OR Witset OR Wolastoqiyik OR Wuikinuxv OR Wunnumin OR "Wuskwi Sipihk" OR Xatsull OR "Xeni Gwet'in" OR Yakweakwioose OR Yekooche OR "Yellow Quill" OR Yellowknife OR Yellowknives OR Zhiibaahaasing) OR AB(Anishnawbek OR "?Akisq'nuk" OR "?aq'am" OR "?Esdilagh" OR "Abénakis" OR "Akisq'nuk" OR "aq'am" OR "Che:k:tles7et'h'" OR "Da'naxda'xw" OR "Dzawada'enuxw" OR "Esk'etemc" OR "Gitga'at" OR "Gitlaxt'aamiks" OR "Gwa'Sala-Nakwaxda'xw" OR "Gwich'in" OR "Ka:'yu:'k't'h'" OR "Kanyen'kehaka" OR "Kapawe'no" OR "K'atlodeeche" OR "K'ómoks" OR "Leq'á:mel" OR "Lil'wat" OR "Malécite" OR "Mi'kmaq" OR "Mowachaht/Muchalaht" OR "Nisga'a" OR "N'Quatqua" OR "O'Chiese" OR "Ononda'gega'" OR "Onondowága'" OR "Onyota'a:ka" OR "Saik'uz" OR "Sambaa K'e " OR "Shxwhá:y" OR "Shxw'ow'hamel" OR "SIPEKNE'KATIK" OR "Skaru:reh" OR "Sq'éwlets" OR "Stellat'en" OR "Sts'ailes" OR "Stz'uminus" OR "T'it'q'et" OR "Tla'amin" OR "Tl'azt'en" OR "Tl'etinqox" OR "Tsal'alh" OR "Tsideldel" OR "Ts'kw'aylaxw" OR "T'Sou-ke" OR "Ts'uubaa-asatx" OR "Tsuut'ina" OR "We'koqma'q" OR "Wet'suwet'en" OR "Xai'xais" OR "Xaxli'p" OR "Yunesit'in" OR ((Acadia OR Carmacks OR Champagne OR Creek OR Delaware OR Douglas OR Montana OR Similkameen) N2 (Indian OR Indians OR Tribe OR Tribes OR native OR natives OR nation OR people*)) OR Aamjiwnaang OR Abegweit OR Abitibiwinni OR Ahousaht OR Ahtahkakoop OR Aishihik OR Aitchelitz OR Aklavik OR Algonquin* OR Anicinape* OR "Animakee Wa Zhing" OR Anishina* OR aqam OR Atikamekw OR Atjanasca* OR Attawapiskat OR Aundeck-Omni-Kaning OR Batchewana OR "Bearfoot Onondaga" OR Beausoleil OR "Behdzi Ahda*" OR "Biigtigong Nishnaabeg" OR "Biinjitiwaabik Zaaging Anishinaabek" OR Bilijk OR "Binche Whut'en" OR "Bingwi Neyaashi Anishinaabek" OR "Canupawakpa Dakota" OR Cayuga OR Chawathil OR Cheam OR "Cheslatta Carrier" OR Chiniki OR Chipew* OR Cote OR Couchiching OR Cowessess OR Cowichan OR Cree OR "Dakota Plains" OR "Dakota Tipi" OR "Dechi Laot'i" OR Deline OR Dena OR Dene OR Denesuline OR "Deninu K'ue" OR Ditidaht OR "Dog Rib Rae" OR Dokis OR Eabametoong OR Ehattesaht OR Elsipogtog OR Ermineskin OR Esdilagh OR Esgenoopetitj OR Eskasoni OR Esquimalt OR Gameti OR Gayogohono OR Ginoogaming OR Gitanmaax OR Gitanyow OR Gitsegukla OR Gitwangak OR Gitxaala OR Glooscap OR Gwawaenuk OR "Gwichya Gwich'in" OR Gwitchin OR Hagwilget OR Haisla OR Halalt OR Haudenosaunee OR Heiltsuk OR Hesquiaht OR Hiawatha OR Homalco OR Hupacasath OR Huu-ay-aht OR Inninuwug OR Innu OR Innue OR Innus OR Inuvik OR Iskatewizaagegan OR Iskut OR "Ka'a'gee Tu" OR Kahkewistahaw OR "Kasabonika Lake" OR Kashechewan OR Katzie OR Kawacatoose OR Kebaowek OR Keeseekoose OR Keeseekoowenin OR Kee-Way-Win OR "Kinistin Saulteaux" OR Kinonjeoshtegon OR Kispiox OR Kitselas OR Kitsumkalum OR Kluane OR Kutchin OR Kwadacha OR Kwakiutl OR "Kwanlin Dun" OR Kwaw-kwaw-Apilt OR Kwiakah OR "Kwikwasut'inuxw Haxwa'mis" OR Kwikwetlem OR "Lax Kw'alaams" OR "Lheidli T'enneh" OR "Liidlii Kue" OR "Listuguj Mi'gmaq" OR Lyackson OR Magnetawan OR Malahat OR Maliseet OR Mamalilikulla OR Matachewan OR Matsqui OR Mattagami OR "M'Chigeeng" OR Membertou OR Metepenagiag OR Metlakatla OR Miawpukek OR Michipicoten OR Micmacs OR Millbrook OR Mishkeegogamang OR Mississauga* OR Mitaanjigamiing OR Mohawk* OR Montagnais OR Moosomin OR Moravian OR Mowachaht OR Muchalaht OR Munsee-Delaware OR Muscowpetung OR Muskoday OR Muskowekwan OR Musqueam OR "Nacho Nyak Dun" OR "Nadleh Whuten" OR Naicatchewenin OR "Nak'azdli Whut'en" OR Nakota OR Naotkamegwanning OR Naskapi OR Natoaganeg OR Nee-Tahi-Buhn OR Nehiyawak OR Nekaneet OR Neskantaga OR Neskonlith OR Nibinamik OR Nigigoonsiminikaaning OR Nipissing OR Nishnaabeg OR Nooaitch OR Nuchatlaht OR Nuxalk OR Ochapowace OR O-Chi-Chak-Ko-Sipi OR Odanak OR Ojibw* OR Okanagan OR Okanese OR Okemasis OR Oneida OR Oromocto OR Osoyoos OR Pabineau OR Pacheedaht OR "Paqtnkek Mi'kmaw" OR Pasqua OR Pauingassi OR Pauquachin OR "Pays Plat" OR Peguis OR "Pehdzeh Ki" OR Pekuakamiulnuatsh OR Penelakut OR Piapot OR Piikani OR Pikangikum OR Pinaymootang OR Potlotek OR "Qalipu Mi'kmaq" OR Qualicum OR Quatsino OR Sagamok OR Sahgaiehcan OR Samahquam OR Saugeen OR Saulteau OR Saulteaux OR Sechelt OR Secwepemc* OR Semiahmoo OR Seneca OR Shackan OR Shamattawa OR Shawanaga OR Sheguiandah OR Sheshegwaning OR Shuswap OR Siksika OR Simpcw OR Sioux OR Siska OR Skatin OR Skawahlook OR Skeetchestn OR Skidegate OR "Skin Tyee" OR Skowkale OR Skownan OR Skuppah OR Skwah OR Snuneymuxw OR Songhees OR Soowahlie OR Splatsin OR Spuzzum OR Squamish OR Squiala OR "Stswecem'c Xget'tem" OR Sumas OR "Ta'an Kwach'an" OR Tagish OR Tahltan OR Takla OR Tallcree OR "Taykwa Tagamou" OR "Teetl'it Gwich'in" OR Temagami OR Thessalon OR Timiskaming OR "Tk'emlúps te Secwépemc" OR Tla-o-qui-aht OR Tlatlasikwala OR Tlingit OR Tlowitsis OR Tootinaowaziibeeng OR Toquaht OR "Tr'ondëk Hwëch'in" OR Tsartlip OR Tsawout OR Tsawwassen OR Tseshaht OR Tseycum OR "Ts'il Kaz Koh" OR Tsleil-Waututh OR Tuscarora OR Tutchone OR Tzeachten OR Uchucklesaht OR Ucluelet OR Ulkatcho OR Wabaseemoong OR Wabauskang OR Wagmatcook OR Wahnapitae OR "Wahpeton Dakota" OR Wahsipekuk OR Wapekeka OR Wasagamack OR Wasauksing OR Waswanipi OR Wawakapewin OR Waywayseecappo OR Webequie OR Weenusk OR Wemotaci OR Wendat OR "Wha Ti" OR Whapmagoostui OR "Whitecap Dakota" OR Wikwemikong OR Witset OR Wolastoqiyik OR Wuikinuxv OR Wunnumin OR "Wuskwi Sipihk" OR Xatsull OR "Xeni Gwet'in" OR Yakweakwioose OR Yekooche OR "Yellow Quill" OR Yellowknife OR Yellowknives OR Zhiibaahaasing) |  | 3086 |
| S5 | TI("Degexit'an" OR "Yup'ik" OR ("Bear River" N2 (Indian OR Indians OR Tribe OR Tribes OR native OR natives OR nation OR people*)) OR Abenaki OR Achumawi OR Ahtna OR "Akimel Oodham" OR "Akimel O'odham" OR "Akimel Ootham" OR "Akimel O'otham" OR Aleut* OR Algonkian* OR Alsea OR Aluutiq OR Apalachee OR Arikara OR Atakapa OR Athabascan OR Athabaskan OR Athapascan OR Athapaskan OR Bannock OR Bidai OR Biloxi OR Blackfeet OR Blackfoot OR Caddoan OR Calice OR Calusa OR Carrizo OR Cayuse OR Chemakum OR Cheraw OR Cherokee OR Chetco OR Cheyenne OR Chicora OR Chilula OR Chimariko OR Clatskanie OR Coahuiltecan OR "Coeur d'Alene" OR Costanoan OR Cree OR Croatan OR Croatoan OR Cusabo OR Edisto OR Eskaleut* OR Eskimo OR Eskimo-Aleut OR Esselen OR Eyak OR "Fox and Sauk" OR Gabrielino OR "Gros Ventre" OR Guale OR "Gwich'in" OR Halchidhoma OR Hidatsa OR Hitchiti OR Hopi OR Houma OR Huichol* OR Hupa OR Ingalik OR Inupiaq OR Ioway OR Iroquoi* OR Jacome OR Jano OR Jeaga OR Juaneno OR Jumano OR Kalapuya OR Kansa OR Karankawa OR Karok OR Kashaya OR Kato OR Kawaiisu OR Kitanemuk OR Kitsai OR Klallam OR Koasati OR Kolchan OR Konkow OR Koyukon OR Lakota* OR Lassik OR Laurentian OR Lenape OR "Lenni Lenape" OR Mahican OR Mandan OR Maricopa OR Massachusett OR Mattole OR Minisink OR Missouria OR Mohave OR Mohawk OR Mohican OR Molalla OR Multnomah OR Munsee OR Muscoge* OR Naabeeho OR Nakoda OR Nanticoke OR Natchez OR Nauset OR Navaho OR Niantic OR Nipmuc OR Nisenan OR Nongatl OR Oconee OR Ofo OR Ohlone OR Ojibw* OR Okanagan OR Oneida OR Otoe OR Palouse OR Panamint OR Papago OR Patwin OR "Pee Dee" OR Pennacook OR Pequot OR Pima OR Piro OR Pocasset OR Pocomtuc OR Pokanoket OR Poospatuck OR Powhatan OR Quechan OR Quileute OR Quinault OR Quiripi OR Salinan OR Samich OR Saponi OR Serrano OR Shasta OR Shawnee OR Siksikaitsitapi OR Sinkyone OR Siouan OR Sioux OR Snohomish OR Snoqualmie OR Squaxin OR Suma OR Susquehannock OR Takelma OR Tanacross OR Tanana OR Tataviam OR Tawakoni OR Tequesta OR Tillamook OR Timucua OR Tlingit OR "Tohono Oodham" OR Tongva OR Tsimshian OR Tubatulabal OR Tutelo OR Tututni OR Unalachtigo OR Unami OR Unangan OR Unangas OR Unkechaug OR Waccamaw OR Walapai OR Wappo OR Wasco OR Whilkut OR Wichita OR Winnebago OR Wintu OR Wishram OR Yahi OR Yakima OR Yamasee OR Yana OR Yaqu OR Yuchi OR Yuki OR Yuma OR Yupik OR Yurok) OR AB("Degexit'an" OR "Yup'ik" OR ("Bear River" N2 (Indian OR Indians OR Tribe OR Tribes OR native OR natives OR nation OR people*)) OR Abenaki OR Achumawi OR Ahtna OR "Akimel Oodham" OR "Akimel O'odham" OR "Akimel Ootham" OR "Akimel O'otham" OR Aleut* OR Algonkian* OR Alsea OR Aluutiq OR Apalachee OR Arikara OR Atakapa OR Athabascan OR Athabaskan OR Athapascan OR Athapaskan OR Bannock OR Bidai OR Biloxi OR Blackfeet OR Blackfoot OR Caddoan OR Calice OR Calusa OR Carrizo OR Cayuse OR Chemakum OR Cheraw OR Cherokee OR Chetco OR Cheyenne OR Chicora OR Chilula OR Chimariko OR Clatskanie OR Coahuiltecan OR "Coeur d'Alene" OR Costanoan OR Cree OR Croatan OR Croatoan OR Cusabo OR Edisto OR Eskaleut* OR Eskimo OR Eskimo-Aleut OR Esselen OR Eyak OR "Fox and Sauk" OR Gabrielino OR "Gros Ventre" OR Guale OR "Gwich'in" OR Halchidhoma OR Hidatsa OR Hitchiti OR Hopi OR Houma OR Huichol* OR Hupa OR Ingalik OR Inupiaq OR Ioway OR Iroquoi* OR Jacome OR Jano OR Jeaga OR Juaneno OR Jumano OR Kalapuya OR Kansa OR Karankawa OR Karok OR Kashaya OR Kato OR Kawaiisu OR Kitanemuk OR Kitsai OR Klallam OR Koasati OR Kolchan OR Konkow OR Koyukon OR Lakota* OR Lassik OR Laurentian OR Lenape OR "Lenni Lenape" OR Mahican OR Mandan OR Maricopa OR Massachusett OR Mattole OR Minisink OR Missouria OR Mohave OR Mohawk OR Mohican OR Molalla OR Multnomah OR Munsee OR Muscoge* OR Naabeeho OR Nakoda OR Nanticoke OR Natchez OR Nauset OR Navaho OR Niantic OR Nipmuc OR Nisenan OR Nongatl OR Oconee OR Ofo OR Ohlone OR Ojibw* OR Okanagan OR Oneida OR Otoe OR Palouse OR Panamint OR Papago OR Patwin OR "Pee Dee" OR Pennacook OR Pequot OR Pima OR Piro OR Pocasset OR Pocomtuc OR Pokanoket OR Poospatuck OR Powhatan OR Quechan OR Quileute OR Quinault OR Quiripi OR Salinan OR Samich OR Saponi OR Serrano OR Shasta OR Shawnee OR Siksikaitsitapi OR Sinkyone OR Siouan OR Sioux OR Snohomish OR Snoqualmie OR Squaxin OR Suma OR Susquehannock OR Takelma OR Tanacross OR Tanana OR Tataviam OR Tawakoni OR Tequesta OR Tillamook OR Timucua OR Tlingit OR "Tohono Oodham" OR Tongva OR Tsimshian OR Tubatulabal OR Tutelo OR Tututni OR Unalachtigo OR Unami OR Unangan OR Unangas OR Unkechaug OR Waccamaw OR Walapai OR Wappo OR Wasco OR Whilkut OR Wichita OR Winnebago OR Wintu OR Wishram OR Yahi OR Yakima OR Yamasee OR Yana OR Yaqu OR Yuchi OR Yuki OR Yuma OR Yupik OR Yurok) |  | 14082 |
| S4 | TI("Asa'carsarmiut" OR "Dee-ni'" OR "Mi'kmaq" OR "S'Klallam" OR "Shoonaq'" OR "Sun'aq" OR (("Ivanof Bay" OR "Shoalwater Bay" OR "Spirit Lake" OR "Squaxin Island" OR Dakota OR Dakotah OR Wichita OR keechi OR waco OR Tawakonie OR Cheyenne OR Creek OR Crow OR Hoonah OR Kasigluk OR Maricopa OR Miami OR Omaha OR Oneida OR Ottawa OR Peoria OR Salamatof OR Sitka OR Snoqualmie OR Spokane OR Winnebago OR Wyandotte) N2 (Indian OR Indians OR Tribe OR Tribes OR native OR natives OR nation OR people*)) OR ((Native OR indian OR traditional) N2 (village OR council OR community OR association)) OR Absentee-Shawnee OR Achomawi OR Agdaagux OR "Agua Caliente" OR Ajumawi OR Ak-Chin OR Akiachak OR Akiak OR Alabama-Coushatta OR Alabama-Quassarte OR Alatna OR Algaaciq OR Algin OR Allakaket OR "Alturas Indian Rancheria" OR Alutiiq OR Angoon OR Anvik OR Apache OR Aporige OR Aquinnah OR arapaho OR Aroostook OR Assiniboine OR Astarawi OR Atsugewi OR Atwamsini OR Bdewakantunwan OR Caddo OR Cahto OR Cahuilla OR Catawba OR Cayuga OR Chalkyitsik OR Cheesh-Na OR Chehalis OR Chemehuevi OR Cher-Ae OR Chevak OR Cheyenne-Arapaho OR Chickahominy OR Chickaloon OR Chickasaw OR Chignik OR Chilkat OR Chilkoot OR Chinik OR Chinook OR Chippewa* OR Chitimacha OR Choctaw OR Chukchansi OR Chuloonawick OR Chumash OR Cocopah OR Colusa OR Comanche OR Coos OR Cosmit OR Coushatta OR Cowlitz OR "Cross Village" OR Cupeno OR Curyung OR Diegueno OR Dine' OR Egegik OR Eklutna OR Elwha OR "Emmonak Village" OR "Evansville Village" OR Ewiiaapaayp OR "Galena Village" OR Goshute OR "Gulkana Village Council" OR Haida OR Hammawi OR Havasupai OR "Healy Lake Village" OR Hewisedawi OR Ho-Chunk OR Hoh OR Hokan OR Holikachuk OR "Holy Cross Tribe" OR Hualapai OR "Hughes Village" OR "Huslia Village" OR "Hydaburg Cooperative Association" OR "Igiugig Village" OR Iipay OR Ilmawi OR Inaja OR Inupiat OR Ione OR Itsatawi OR "Kaguyak Village" OR Kaibab OR "Kaktovik Village" OR Kalispel OR Kanosh OR Karuk OR Kashia OR Kaw OR Keetoowah OR "Kenaitze Indian Tribe" OR "Ketchikan Indian Community" OR "Keweenaw Bay" OR Kialegee OR Kickapoo OR "King Island Native Community" OR "King Salmon Tribe" OR Kiowa OR Klamath OR "Klawock Cooperative Association" OR Kletsel OR "Knik Tribe" OR "Koi Nation" OR "Kokhanok Village" OR Koosharem OR Kootenai OR Kosalektawi OR "Koyukuk Native Village" OR Kumeyaay OR Kwalhioqua OR "Lesnoi Village" OR "Levelock Village" OR "Lime Village" OR "Louden Village" OR Luiseno OR Lummi OR Madesi OR Maidu OR Makah OR Maliseet OR "Manley Hot Springs Village" OR "Manokotak Village" OR Manzanita OR Mashantucket OR Match-e-be-nash-she-wish OR Mattaponi OR "McGrath Native Village" OR Mdewakanton OR Mechoopda OR Menominee OR Me-Wuk OR Miccosukee OR Micmacs OR "Mission Indians" OR Miwok OR Miwuk OR Modoc OR Mohegan OR Mojave OR Monacan OR "Mono Indians" OR Muckleshoot OR Muscogee OR "Na Dené" OR Nansemond OR Narragansett OR Navajo OR "New Koliganek Village" OR "New Stuyahok Village" OR "Newhalen Village" OR "Newtok Village" OR "Nez Perce" OR "Nikolai Village" OR "Ninilchik Village" OR Nisqually OR Nomlaki OR "Nondalton Village" OR Nooksack OR "Northway Village" OR Nottawaseppi OR "Nulato Village" OR Nunakauyarmiut OR Odawa OR Ohgsenakale OR "Ohkay Owingeh" OR Onondaga OR Oreilles OR "Organized Village of Grayling" OR "Organized Village of Kake" OR "Organized Village of Kasaan" OR "Organized Village of Kwethluk" OR "Organized Village of Saxman" OR Orutsararmiut OR Osage OR Otoe-Missouria OR Paiute OR Paiute-Shoshone OR Pala OR Pamunkey OR Pascua OR Paskenta OR Passamaquoddy OR "Pauloff Harbor Village" OR Pauma OR Pawnee OR Pechanga OR "Pechanga " OR "Peck Indian Reservation" OR "Pedro Bay Village" OR Pee-Posh OR Penobscot OR Penutian OR Perquot OR "Pima People" OR Pima-Maricopa OR Poarch OR Pokagon OR Pomo OR Ponca OR "Portage Creek Village" OR Potawatomi OR Pottawatomi OR Pueblo OR Puyallup OR "Qagan Tayagungin" OR Qawalangin OR Quapaw OR Quechan OR "Rampart Village" OR Rappahannock OR Resighini OR "Sac & Fox" OR "Sac and Fox" OR "Saint George Island" OR "Saint Paul Island" OR Salish OR Samish OR Santee OR Sauk-Suiattle OR "Seldovia Village Tribe" OR Seminole OR Seneca OR Seneca-Cayuga OR Shinnecock OR Shivwits OR Shoshone OR Shoshone-Bannock OR Shoshone-Paiute OR Siuslaw OR Skagit OR "Skagway Village" OR Skokomish OR "South Naknek Village" OR "Stebbins Community Association" OR Stillaguamish OR Suquamish OR Swinomish OR "Takotna Village" OR Tangirnaq OR Tejon OR "Telida Village" OR Te-Moak OR Thlopthlocco OR "Tlingit & Haida" OR "Tohono O'odham" OR Tolowa OR Tonawanda OR Tonkawa OR Tulalip OR Tunica-Biloxi OR Tuolumne OR Tuscarora OR "Twin Hills Village" OR "Ugashik Village" OR Umatilla OR Umkumiut OR Umpqua OR Ute OR "Utu Utu Gwaitu" OR "Village of Alakanuk" OR "Village of Anaktuvuk Pass" OR "Village of Aniak" OR "Village of Atmautluak" OR "Village of Bill Moore's Slough" OR "Village of Chefornak" OR "Village of Clarks Point" OR "Village of Crooked Creek" OR "Village of Dot Lake" OR "Village of Iliamna" OR "Village of Kalskag" OR "Village of Kaltag" OR "Village of Kotlik" OR "Village of Lower Kalskag" OR "Village of Ohogamiut" OR "Village of Red Devil" OR "Village of Sleetmute" OR "Village of Solomon" OR "Village of Stony River" OR "Village of Venetie" OR "Village of Wainwright" OR Wailaki OR Wampanoag OR "Warm Springs Reservation" OR Washoe OR Willapa OR Winnemucca OR Wintun OR Wiyot OR "Wrangell Cooperative Association" OR Yakama OR Yakutat OR Yaqui OR Yavapai OR Yokuts OR Yuhaaviatam OR Yuima OR Yukian OR Yukut OR Yupiit OR Zuni) OR AB("Asa'carsarmiut" OR "Dee-ni'" OR "Mi'kmaq" OR "S'Klallam" OR "Shoonaq'" OR "Sun'aq" OR (("Ivanof Bay" OR "Shoalwater Bay" OR "Spirit Lake" OR "Squaxin Island" OR Dakota OR Dakotah OR Wichita OR keechi OR waco OR Tawakonie OR Cheyenne OR Creek OR Crow OR Hoonah OR Kasigluk OR Maricopa OR Miami OR Omaha OR Oneida OR Ottawa OR Peoria OR Salamatof OR Sitka OR Snoqualmie OR Spokane OR Winnebago OR Wyandotte) N2 (Indian OR Indians OR Tribe OR Tribes OR native OR natives OR nation OR people*)) OR ((Native OR indian OR traditional) N2 (village OR council OR community OR association)) OR Absentee-Shawnee OR Achomawi OR Agdaagux OR "Agua Caliente" OR Ajumawi OR Ak-Chin OR Akiachak OR Akiak OR Alabama-Coushatta OR Alabama-Quassarte OR Alatna OR Algaaciq OR Algin OR Allakaket OR "Alturas Indian Rancheria" OR Alutiiq OR Angoon OR Anvik OR Apache OR Aporige OR Aquinnah OR arapaho OR Aroostook OR Assiniboine OR Astarawi OR Atsugewi OR Atwamsini OR Bdewakantunwan OR Caddo OR Cahto OR Cahuilla OR Catawba OR Cayuga OR Chalkyitsik OR Cheesh-Na OR Chehalis OR Chemehuevi OR Cher-Ae OR Chevak OR Cheyenne-Arapaho OR Chickahominy OR Chickaloon OR Chickasaw OR Chignik OR Chilkat OR Chilkoot OR Chinik OR Chinook OR Chippewa* OR Chitimacha OR Choctaw OR Chukchansi OR Chuloonawick OR Chumash OR Cocopah OR Colusa OR Comanche OR Coos OR Cosmit OR Coushatta OR Cowlitz OR "Cross Village" OR Cupeno OR Curyung OR Diegueno OR Dine' OR Egegik OR Eklutna OR Elwha OR "Emmonak Village" OR "Evansville Village" OR Ewiiaapaayp OR "Galena Village" OR Goshute OR "Gulkana Village Council" OR Haida OR Hammawi OR Havasupai OR "Healy Lake Village" OR Hewisedawi OR Ho-Chunk OR Hoh OR Hokan OR Holikachuk OR "Holy Cross Tribe" OR Hualapai OR "Hughes Village" OR "Huslia Village" OR "Hydaburg Cooperative Association" OR "Igiugig Village" OR Iipay OR Ilmawi OR Inaja OR Inupiat OR Ione OR Itsatawi OR "Kaguyak Village" OR Kaibab OR "Kaktovik Village" OR Kalispel OR Kanosh OR Karuk OR Kashia OR Kaw OR Keetoowah OR "Kenaitze Indian Tribe" OR "Ketchikan Indian Community" OR "Keweenaw Bay" OR Kialegee OR Kickapoo OR "King Island Native Community" OR "King Salmon Tribe" OR Kiowa OR Klamath OR "Klawock Cooperative Association" OR Kletsel OR "Knik Tribe" OR "Koi Nation" OR "Kokhanok Village" OR Koosharem OR Kootenai OR Kosalektawi OR "Koyukuk Native Village" OR Kumeyaay OR Kwalhioqua OR "Lesnoi Village" OR "Levelock Village" OR "Lime Village" OR "Louden Village" OR Luiseno OR Lummi OR Madesi OR Maidu OR Makah OR Maliseet OR "Manley Hot Springs Village" OR "Manokotak Village" OR Manzanita OR Mashantucket OR Match-e-be-nash-she-wish OR Mattaponi OR "McGrath Native Village" OR Mdewakanton OR Mechoopda OR Menominee OR Me-Wuk OR Miccosukee OR Micmacs OR "Mission Indians" OR Miwok OR Miwuk OR Modoc OR Mohegan OR Mojave OR Monacan OR "Mono Indians" OR Muckleshoot OR Muscogee OR "Na Dené" OR Nansemond OR Narragansett OR Navajo OR "New Koliganek Village" OR "New Stuyahok Village" OR "Newhalen Village" OR "Newtok Village" OR "Nez Perce" OR "Nikolai Village" OR "Ninilchik Village" OR Nisqually OR Nomlaki OR "Nondalton Village" OR Nooksack OR "Northway Village" OR Nottawaseppi OR "Nulato Village" OR Nunakauyarmiut OR Odawa OR Ohgsenakale OR "Ohkay Owingeh" OR Onondaga OR Oreilles OR "Organized Village of Grayling" OR "Organized Village of Kake" OR "Organized Village of Kasaan" OR "Organized Village of Kwethluk" OR "Organized Village of Saxman" OR Orutsararmiut OR Osage OR Otoe-Missouria OR Paiute OR Paiute-Shoshone OR Pala OR Pamunkey OR Pascua OR Paskenta OR Passamaquoddy OR "Pauloff Harbor Village" OR Pauma OR Pawnee OR Pechanga OR "Pechanga " OR "Peck Indian Reservation" OR "Pedro Bay Village" OR Pee-Posh OR Penobscot OR Penutian OR Perquot OR "Pima People" OR Pima-Maricopa OR Poarch OR Pokagon OR Pomo OR Ponca OR "Portage Creek Village" OR Potawatomi OR Pottawatomi OR Pueblo OR Puyallup OR "Qagan Tayagungin" OR Qawalangin OR Quapaw OR Quechan OR "Rampart Village" OR Rappahannock OR Resighini OR "Sac & Fox" OR "Sac and Fox" OR "Saint George Island" OR "Saint Paul Island" OR Salish OR Samish OR Santee OR Sauk-Suiattle OR "Seldovia Village Tribe" OR Seminole OR Seneca OR Seneca-Cayuga OR Shinnecock OR Shivwits OR Shoshone OR Shoshone-Bannock OR Shoshone-Paiute OR Siuslaw OR Skagit OR "Skagway Village" OR Skokomish OR "South Naknek Village" OR "Stebbins Community Association" OR Stillaguamish OR Suquamish OR Swinomish OR "Takotna Village" OR Tangirnaq OR Tejon OR "Telida Village" OR Te-Moak OR Thlopthlocco OR "Tlingit & Haida" OR "Tohono O'odham" OR Tolowa OR Tonawanda OR Tonkawa OR Tulalip OR Tunica-Biloxi OR Tuolumne OR Tuscarora OR "Twin Hills Village" OR "Ugashik Village" OR Umatilla OR Umkumiut OR Umpqua OR Ute OR "Utu Utu Gwaitu" OR "Village of Alakanuk" OR "Village of Anaktuvuk Pass" OR "Village of Aniak" OR "Village of Atmautluak" OR "Village of Bill Moore's Slough" OR "Village of Chefornak" OR "Village of Clarks Point" OR "Village of Crooked Creek" OR "Village of Dot Lake" OR "Village of Iliamna" OR "Village of Kalskag" OR "Village of Kaltag" OR "Village of Kotlik" OR "Village of Lower Kalskag" OR "Village of Ohogamiut" OR "Village of Red Devil" OR "Village of Sleetmute" OR "Village of Solomon" OR "Village of Stony River" OR "Village of Venetie" OR "Village of Wainwright" OR Wailaki OR Wampanoag OR "Warm Springs Reservation" OR Washoe OR Willapa OR Winnemucca OR Wintun OR Wiyot OR "Wrangell Cooperative Association" OR Yakama OR Yakutat OR Yaqui OR Yavapai OR Yokuts OR Yuhaaviatam OR Yuima OR Yukian OR Yukut OR Yupiit OR Zuni) |  | 7268 |
| S3 | MH("Arctic Peoples" OR "Inuit") or TI(inuit or (arctic N2 people*)) OR AB(inuit or (arctic N2 people*)) |  | 1283 |
| S2 | MH("Aboriginal Canadians" OR "First Nations of Canada") OR TI(((Canada OR Canadian*) N2 (native* OR indigenous OR aborigin*)) OR Metis OR "Métis" OR Michif* OR Mitchif OR Mechif OR "Métif" OR "Métchif" OR Metif OR Metchif OR Inuit OR "First Nations" OR "First Nation") OR AB(((Canada OR Canadian*) N2 (native* OR indigenous OR aborigin*)) OR Metis OR "Métis" OR Michif* OR Mitchif OR Mechif OR "Métif" OR "Métchif" OR Metif OR Metchif OR Inuit OR "First Nations" OR "First Nation") |  | 2937 |
| S1 | MH("Native Americans" OR "Alaska Natives") OR TI(((American* OR Alaska*) N2 (Native* OR Indian OR Indians OR indigenous OR aborigin*)) OR Amerind*) OR AB(((American* OR Alaska*) N2 (Native* OR Indian OR Indians OR indigenous OR aborigin*)) OR Amerind*) |  | 13305 |

### APA PsycInfo (EBSCOhost)

**Field Codes:** TI = title, AB= abstract, DE = subjects [exact], RD = release date (date added).

**Proximity operator:** N# 
**Truncation:**  *

**Database** - APA PsycInfo

**Interface** - EBSCOhost Research Databases

**Search Screen** - Advanced Search

**Search modes** – Proximity

**Date** - Thu, November 21, 2024 2:43:05 PM

| **#** | **Query** | **Results** |
| --- | --- | --- |
| S27 | S26 AND RD 20230506- | 32 |
| S26 | S20 OR S21 OR S22 OR S23 OR S24 OR S25 | 457 |
| S25 | S17 AND (S12 OR S15) AND S18 | 26 |
| S24 | S19 AND (S8 OR S9) | 291 |
| S23 | S15 AND (S8 OR S9) | 96 |
| S22 | S12 AND S16 | 7 |
| S21 | S9 and S12 | 31 |
| S20 | S8 and S12 | 29 |
| S19 | TI((women* OR woman* OR female*) N2 (health OR healthcare OR service* OR clinic OR clinics)) OR AB((women* OR woman* OR female*) N2 (health OR healthcare OR service* OR clinic OR clinics)) | 21627 |
| S18 | TI("North America" OR Canada OR Alberta OR "British Columbia" OR Manitoba OR "New Brunswick" OR "Newfoundland" OR "Labrador" OR "Northwest Territories" OR "Nova Scotia" OR Nunavut OR Ontario OR "Prince Edward Island" OR Quebec OR Saskatchewan OR Yukon OR "United States" OR Appalachia* OR "New England" OR "Pacific States" OR Alabama OR Alaska OR Arizona OR Arkansas OR California OR Colorado OR Connecticut OR Delaware OR Florida OR Georgia OR Hawaii OR Idaho OR Illinois OR Indiana OR Iowa OR Kansas OR Kentucky OR Louisiana OR Maine OR Maryland OR Massachusetts OR Michigan OR Minnesota OR Mississippi OR Missouri OR Montana OR Nebraska OR Nevada OR "New Hampshire" OR "New Jersey" OR "New Mexico" OR "New York" OR "North Carolina" OR "North Dakota" OR Ohio OR Oklahoma OR Oregon OR Pennsylvania OR "Rhode Island" OR "South Carolina" OR "South Dakota" OR Tennessee OR Texas OR Utah OR Vermont OR Virginia OR Washington OR "West Virginia" OR Wisconsin OR Wyoming OR "District of Columbia" OR "Washington DC") OR AB("North America" OR Canada OR Alberta OR "British Columbia" OR Manitoba OR "New Brunswick" OR "Newfoundland" OR "Labrador" OR "Northwest Territories" OR "Nova Scotia" OR Nunavut OR Ontario OR "Prince Edward Island" OR Quebec OR Saskatchewan OR Yukon OR "United States" OR Appalachia* OR "New England" OR "Pacific States" OR Alabama OR Alaska OR Arizona OR Arkansas OR California OR Colorado OR Connecticut OR Delaware OR Florida OR Georgia OR Hawaii OR Idaho OR Illinois OR Indiana OR Iowa OR Kansas OR Kentucky OR Louisiana OR Maine OR Maryland OR Massachusetts OR Michigan OR Minnesota OR Mississippi OR Missouri OR Montana OR Nebraska OR Nevada OR "New Hampshire" OR "New Jersey" OR "New Mexico" OR "New York" OR "North Carolina" OR "North Dakota" OR Ohio OR Oklahoma OR Oregon OR Pennsylvania OR "Rhode Island" OR "South Carolina" OR "South Dakota" OR Tennessee OR Texas OR Utah OR Vermont OR Virginia OR Washington OR "West Virginia" OR Wisconsin OR Wyoming OR "District of Columbia" OR "Washington DC") | 376587 |
| S17 | DE("Alternative Medicine" OR "Medicinal Herbs and Plants") OR TI(((Complementary OR alternative OR traditional) N2 (medicine* OR therap* OR healing OR treatment*)) OR "Herbal Medicine" OR herbalism OR ((botanical OR ethnobotanical OR herb* OR plant OR plant-based) N2 (medicine* OR remedy OR remedies OR healing OR therap* OR treatment*)) OR phyto-medicine OR phytomedic* OR "folk medicine" OR "folk healer") OR AB(((Complementary OR alternative OR traditional) N2 (medicine* OR therap* OR healing OR treatment*)) OR "Herbal Medicine" OR herbalism OR ((botanical OR ethnobotanical OR herb* OR plant OR plant-based) N2 (medicine* OR remedy OR remedies OR healing OR therap* OR treatment*)) OR phyto-medicine OR phytomedic* OR "folk medicine" OR "folk healer") | 24377 |
| S16 | DE(Shamanism) OR TI(((Indigenous OR native OR "first nation" OR aborigin* OR Indian) N2 (health OR healthcare OR healing OR medicine OR practices)) OR ethnomedicin* OR "native healing" OR Limpia OR "indigenous healer" OR medicineman OR medicinemen OR "medicine man" OR "medicine men" OR Curandero OR "folk healer" OR shaman* OR shamen) OR AB(((Indigenous OR native OR "first nation" OR aborigin* OR Indian) N2 (health OR healthcare OR healing OR medicine OR practices)) OR ethnomedicin* OR "native healing" OR Limpia OR "indigenous healer" OR medicineman OR medicinemen OR "medicine man" OR "medicine men" OR Curandero OR "folk healer" OR shaman* OR shamen) | 5739 |
| S15 | S13 AND S14 | 2054 |
| S14 | DE("Human Females") OR TI(female OR woman OR women) OR AB(female OR woman OR women) | 689380 |
| S13 | TI(elder OR elders) OR AB(elder OR elders) | 12148 |
| S12 | S10 OR S11 | 11415 |
| S11 | TI("Hot Flashes" OR "Hot Flash*" OR "Hot Flush*" OR "night sweat*" OR "nocturnal hyperhidrosis" OR "vasomotor symptom*" OR VMS) OR AB("Hot Flashes" OR "Hot Flash*" OR "Hot Flush*" OR "night sweat*" OR "nocturnal hyperhidrosis" OR "vasomotor symptom*" OR VMS) | 1135 |
| S10 | DE(Menopause) OR TI(Climacteric OR Climacterium OR Menopause OR Menopaus* OR "Menopause, Premature" OR "climacterium praecox" OR Perimenopause OR Perimenopaus* OR Postmenopause OR Postmenopaus* OR "Post menopaus*" OR Post-menopaus* OR Premenopause OR Premenopause* OR Pre-menopaus* OR "Pre menopaus*" OR Climacteric* OR "Life Change*" OR "Change of life") OR AB(Climacteric OR Climacterium OR Menopause OR Menopaus* OR "Menopause, Premature" OR "climacterium praecox" OR Perimenopause OR Perimenopaus* OR Postmenopause OR Postmenopaus* OR "Post menopaus*" OR Post-menopaus* OR Premenopause OR Premenopause* OR Pre-menopaus* OR "Pre menopaus*" OR Climacteric* OR "Life Change*" OR "Change of life") | 10978 |
| S9 | S3 OR S4 OR S5 OR S6 OR S7 | 20125 |
| S8 | S1 OR S2 | 16534 |
| S7 | TI("Kwakwaka'wakw" OR "Kwak'wala" OR "L'uml'umuluts" OR "Lheidli T'enneh " OR "Nlaka'pamux" OR "Nuxálk" OR "Quw'utsun" OR "Qwum'yiqun" OR "S'amuna" OR "Sm'algyax" OR "St'at'imc" OR "Stl'atl'imx" OR "Sto:lo" OR "Tl'ulpalus" OR "Tsilhqot'in" OR "T'Souke" OR "Xa'xtsa" OR (Labrador N2 (Indian OR Indians OR Tribe OR Tribes OR native OR natives OR nation OR people*)) OR "Áa Tlein Kwáan" OR Aivilingmiut OR Algonkian OR Algonquian OR Algonquin OR Anishinaabe OR "Atlin people" OR "Baffin Island" OR "Bella Coola" OR Burrard OR Cheklesahht OR Chilcotin OR Chippew* OR Clayoquot OR Clemclemaluts OR Comiaken OR Comox OR Coquitlam OR Dakelh OR Dane-zaa OR "Deisleen Kwáan" OR Dunne-za OR Gitxsan OR Gitxsanimaax OR Guyohkohnyo OR "Haad kil" OR Halkomelem OR Halqemeylem OR Henaksiala OR Hinupsum OR Hunquminum OR Hwlitsum OR Hwulqwselu OR Hydin OR Iglulik OR Iglulingmuit OR Ininew OR Inuinnait OR Inuk OR Inupiat OR Inuvialuit OR Iroquois OR Kainai OR Kaska OR Kelsemeht OR Khenipsen OR Kilpahlas OR Kimsquit OR Kitamaat OR Kitlinermiut OR Kivallirmiut OR Klahoose OR Koksilah OR Kootenay OR Koskimo OR Ktunaxa OR Kwantlen OR Kwatna OR Kyuquot OR Labradormiut OR Laich-kwil-tach OR Lamalch* OR Lekwungen OR Lillooet OR Loucheaux OR Lummi OR Micmac OR Mississaugas OR Mohawk OR Mowachaht-Muchalaht OR Nadene OR Na-Dene OR Namgis OR Nanaimo OR Nanoose OR Nequatque OR Netsilik OR Netsilingmiut OR Niitsitapi OR Nimpkish OR Nootka OR Nunavimiut OR Nunavimmiut OR Nuu-chah-nulth OR Odawa OR Ohiaht OR Oji-cree OR Onayotekaono OR Onondaga OR Onondowahgah OR Onundagaono OR Opetchesaht OR Owekeeno OR Passamaquoddy OR Peigan OR Petun OR Plains-Cree OR Popkum OR Potawatomi OR Quamichan OR Sahtu OR Salish OR Sarcee OR Scowlitz OR "Seabird Island" OR Secwepemc OR Sekani OR Semahquam OR Shishalh OR Sinixt OR Ska-Ruh-Reh OR Skway OR Sliammon OR Snaw-naw-as OR Somena OR Songish OR Sooke OR Stoney OR Stuie OR Tallheo OR Tasttine OR Teslin OR Tlicho OR Tsimshianic OR Tununirmiut OR Ungava OR Wakashan OR Weewaikai OR Wewaykum OR Wyandot OR Yuculta) OR AB("Kwakwaka'wakw" OR "Kwak'wala" OR "L'uml'umuluts" OR "Lheidli T'enneh " OR "Nlaka'pamux" OR "Nuxálk" OR "Quw'utsun" OR "Qwum'yiqun" OR "S'amuna" OR "Sm'algyax" OR "St'at'imc" OR "Stl'atl'imx" OR "Sto:lo" OR "Tl'ulpalus" OR "Tsilhqot'in" OR "T'Souke" OR "Xa'xtsa" OR (Labrador N2 (Indian OR Indians OR Tribe OR Tribes OR native OR natives OR nation OR people*)) OR "Áa Tlein Kwáan" OR Aivilingmiut OR Algonkian OR Algonquian OR Algonquin OR Anishinaabe OR "Atlin people" OR "Baffin Island" OR "Bella Coola" OR Burrard OR Cheklesahht OR Chilcotin OR Chippew* OR Clayoquot OR Clemclemaluts OR Comiaken OR Comox OR Coquitlam OR Dakelh OR Dane-zaa OR "Deisleen Kwáan" OR Dunne-za OR Gitxsan OR Gitxsanimaax OR Guyohkohnyo OR "Haad kil" OR Halkomelem OR Halqemeylem OR Henaksiala OR Hinupsum OR Hunquminum OR Hwlitsum OR Hwulqwselu OR Hydin OR Iglulik OR Iglulingmuit OR Ininew OR Inuinnait OR Inuk OR Inupiat OR Inuvialuit OR Iroquois OR Kainai OR Kaska OR Kelsemeht OR Khenipsen OR Kilpahlas OR Kimsquit OR Kitamaat OR Kitlinermiut OR Kivallirmiut OR Klahoose OR Koksilah OR Kootenay OR Koskimo OR Ktunaxa OR Kwantlen OR Kwatna OR Kyuquot OR Labradormiut OR Laich-kwil-tach OR Lamalch* OR Lekwungen OR Lillooet OR Loucheaux OR Lummi OR Micmac OR Mississaugas OR Mohawk OR Mowachaht-Muchalaht OR Nadene OR Na-Dene OR Namgis OR Nanaimo OR Nanoose OR Nequatque OR Netsilik OR Netsilingmiut OR Niitsitapi OR Nimpkish OR Nootka OR Nunavimiut OR Nunavimmiut OR Nuu-chah-nulth OR Odawa OR Ohiaht OR Oji-cree OR Onayotekaono OR Onondaga OR Onondowahgah OR Onundagaono OR Opetchesaht OR Owekeeno OR Passamaquoddy OR Peigan OR Petun OR Plains-Cree OR Popkum OR Potawatomi OR Quamichan OR Sahtu OR Salish OR Sarcee OR Scowlitz OR "Seabird Island" OR Secwepemc OR Sekani OR Semahquam OR Shishalh OR Sinixt OR Ska-Ruh-Reh OR Skway OR Sliammon OR Snaw-naw-as OR Somena OR Songish OR Sooke OR Stoney OR Stuie OR Tallheo OR Tasttine OR Teslin OR Tlicho OR Tsimshianic OR Tununirmiut OR Ungava OR Wakashan OR Weewaikai OR Wewaykum OR Wyandot OR Yuculta) | 658 |
| S6 | TI(Anishnawbek OR "?Akisq'nuk" OR "?aq'am" OR "?Esdilagh" OR "Abénakis" OR "Akisq'nuk" OR "aq'am" OR "Che:k:tles7et'h'" OR "Da'naxda'xw" OR "Dzawada'enuxw" OR "Esk'etemc" OR "Gitga'at" OR "Gitlaxt'aamiks" OR "Gwa'Sala-Nakwaxda'xw" OR "Gwich'in" OR "Ka:'yu:'k't'h'" OR "Kanyen'kehaka" OR "Kapawe'no" OR "K'atlodeeche" OR "K'ómoks" OR "Leq'á:mel" OR "Lil'wat" OR "Malécite" OR "Mi'kmaq" OR "Mowachaht/Muchalaht" OR "Nisga'a" OR "N'Quatqua" OR "O'Chiese" OR "Ononda'gega'" OR "Onondowága'" OR "Onyota'a:ka" OR "Saik'uz" OR "Sambaa K'e " OR "Shxwhá:y" OR "Shxw'ow'hamel" OR "SIPEKNE'KATIK" OR "Skaru:reh" OR "Sq'éwlets" OR "Stellat'en" OR "Sts'ailes" OR "Stz'uminus" OR "T'it'q'et" OR "Tla'amin" OR "Tl'azt'en" OR "Tl'etinqox" OR "Tsal'alh" OR "Tsideldel" OR "Ts'kw'aylaxw" OR "T'Sou-ke" OR "Ts'uubaa-asatx" OR "Tsuut'ina" OR "We'koqma'q" OR "Wet'suwet'en" OR "Xai'xais" OR "Xaxli'p" OR "Yunesit'in" OR ((Acadia OR Carmacks OR Champagne OR Creek OR Delaware OR Douglas OR Montana OR Similkameen) N2 (Indian OR Indians OR Tribe OR Tribes OR native OR natives OR nation OR people*)) OR Aamjiwnaang OR Abegweit OR Abitibiwinni OR Ahousaht OR Ahtahkakoop OR Aishihik OR Aitchelitz OR Aklavik OR Algonquin* OR Anicinape* OR "Animakee Wa Zhing" OR Anishina* OR aqam OR Atikamekw OR Atjanasca* OR Attawapiskat OR Aundeck-Omni-Kaning OR Batchewana OR "Bearfoot Onondaga" OR Beausoleil OR "Behdzi Ahda*" OR "Biigtigong Nishnaabeg" OR "Biinjitiwaabik Zaaging Anishinaabek" OR Bilijk OR "Binche Whut'en" OR "Bingwi Neyaashi Anishinaabek" OR "Canupawakpa Dakota" OR Cayuga OR Chawathil OR Cheam OR "Cheslatta Carrier" OR Chiniki OR Chipew* OR Cote OR Couchiching OR Cowessess OR Cowichan OR Cree OR "Dakota Plains" OR "Dakota Tipi" OR "Dechi Laot'i" OR Deline OR Dena OR Dene OR Denesuline OR "Deninu K'ue" OR Ditidaht OR "Dog Rib Rae" OR Dokis OR Eabametoong OR Ehattesaht OR Elsipogtog OR Ermineskin OR Esdilagh OR Esgenoopetitj OR Eskasoni OR Esquimalt OR Gameti OR Gayogohono OR Ginoogaming OR Gitanmaax OR Gitanyow OR Gitsegukla OR Gitwangak OR Gitxaala OR Glooscap OR Gwawaenuk OR "Gwichya Gwich'in" OR Gwitchin OR Hagwilget OR Haisla OR Halalt OR Haudenosaunee OR Heiltsuk OR Hesquiaht OR Hiawatha OR Homalco OR Hupacasath OR Huu-ay-aht OR Inninuwug OR Innu OR Innue OR Innus OR Inuvik OR Iskatewizaagegan OR Iskut OR "Ka'a'gee Tu" OR Kahkewistahaw OR "Kasabonika Lake" OR Kashechewan OR Katzie OR Kawacatoose OR Kebaowek OR Keeseekoose OR Keeseekoowenin OR Kee-Way-Win OR "Kinistin Saulteaux" OR Kinonjeoshtegon OR Kispiox OR Kitselas OR Kitsumkalum OR Kluane OR Kutchin OR Kwadacha OR Kwakiutl OR "Kwanlin Dun" OR Kwaw-kwaw-Apilt OR Kwiakah OR "Kwikwasut'inuxw Haxwa'mis" OR Kwikwetlem OR "Lax Kw'alaams" OR "Lheidli T'enneh" OR "Liidlii Kue" OR "Listuguj Mi'gmaq" OR Lyackson OR Magnetawan OR Malahat OR Maliseet OR Mamalilikulla OR Matachewan OR Matsqui OR Mattagami OR "M'Chigeeng" OR Membertou OR Metepenagiag OR Metlakatla OR Miawpukek OR Michipicoten OR Micmacs OR Millbrook OR Mishkeegogamang OR Mississauga* OR Mitaanjigamiing OR Mohawk* OR Montagnais OR Moosomin OR Moravian OR Mowachaht OR Muchalaht OR Munsee-Delaware OR Muscowpetung OR Muskoday OR Muskowekwan OR Musqueam OR "Nacho Nyak Dun" OR "Nadleh Whuten" OR Naicatchewenin OR "Nak'azdli Whut'en" OR Nakota OR Naotkamegwanning OR Naskapi OR Natoaganeg OR Nee-Tahi-Buhn OR Nehiyawak OR Nekaneet OR Neskantaga OR Neskonlith OR Nibinamik OR Nigigoonsiminikaaning OR Nipissing OR Nishnaabeg OR Nooaitch OR Nuchatlaht OR Nuxalk OR Ochapowace OR O-Chi-Chak-Ko-Sipi OR Odanak OR Ojibw* OR Okanagan OR Okanese OR Okemasis OR Oneida OR Oromocto OR Osoyoos OR Pabineau OR Pacheedaht OR "Paqtnkek Mi'kmaw" OR Pasqua OR Pauingassi OR Pauquachin OR "Pays Plat" OR Peguis OR "Pehdzeh Ki" OR Pekuakamiulnuatsh OR Penelakut OR Piapot OR Piikani OR Pikangikum OR Pinaymootang OR Potlotek OR "Qalipu Mi'kmaq" OR Qualicum OR Quatsino OR Sagamok OR Sahgaiehcan OR Samahquam OR Saugeen OR Saulteau OR Saulteaux OR Sechelt OR Secwepemc* OR Semiahmoo OR Seneca OR Shackan OR Shamattawa OR Shawanaga OR Sheguiandah OR Sheshegwaning OR Shuswap OR Siksika OR Simpcw OR Sioux OR Siska OR Skatin OR Skawahlook OR Skeetchestn OR Skidegate OR "Skin Tyee" OR Skowkale OR Skownan OR Skuppah OR Skwah OR Snuneymuxw OR Songhees OR Soowahlie OR Splatsin OR Spuzzum OR Squamish OR Squiala OR "Stswecem'c Xget'tem" OR Sumas OR "Ta'an Kwach'an" OR Tagish OR Tahltan OR Takla OR Tallcree OR "Taykwa Tagamou" OR "Teetl'it Gwich'in" OR Temagami OR Thessalon OR Timiskaming OR "Tk'emlúps te Secwépemc" OR Tla-o-qui-aht OR Tlatlasikwala OR Tlingit OR Tlowitsis OR Tootinaowaziibeeng OR Toquaht OR "Tr'ondëk Hwëch'in" OR Tsartlip OR Tsawout OR Tsawwassen OR Tseshaht OR Tseycum OR "Ts'il Kaz Koh" OR Tsleil-Waututh OR Tuscarora OR Tutchone OR Tzeachten OR Uchucklesaht OR Ucluelet OR Ulkatcho OR Wabaseemoong OR Wabauskang OR Wagmatcook OR Wahnapitae OR "Wahpeton Dakota" OR Wahsipekuk OR Wapekeka OR Wasagamack OR Wasauksing OR Waswanipi OR Wawakapewin OR Waywayseecappo OR Webequie OR Weenusk OR Wemotaci OR Wendat OR "Wha Ti" OR Whapmagoostui OR "Whitecap Dakota" OR Wikwemikong OR Witset OR Wolastoqiyik OR Wuikinuxv OR Wunnumin OR "Wuskwi Sipihk" OR Xatsull OR "Xeni Gwet'in" OR Yakweakwioose OR Yekooche OR "Yellow Quill" OR Yellowknife OR Yellowknives OR Zhiibaahaasing) OR AB(Anishnawbek OR "?Akisq'nuk" OR "?aq'am" OR "?Esdilagh" OR "Abénakis" OR "Akisq'nuk" OR "aq'am" OR "Che:k:tles7et'h'" OR "Da'naxda'xw" OR "Dzawada'enuxw" OR "Esk'etemc" OR "Gitga'at" OR "Gitlaxt'aamiks" OR "Gwa'Sala-Nakwaxda'xw" OR "Gwich'in" OR "Ka:'yu:'k't'h'" OR "Kanyen'kehaka" OR "Kapawe'no" OR "K'atlodeeche" OR "K'ómoks" OR "Leq'á:mel" OR "Lil'wat" OR "Malécite" OR "Mi'kmaq" OR "Mowachaht/Muchalaht" OR "Nisga'a" OR "N'Quatqua" OR "O'Chiese" OR "Ononda'gega'" OR "Onondowága'" OR "Onyota'a:ka" OR "Saik'uz" OR "Sambaa K'e " OR "Shxwhá:y" OR "Shxw'ow'hamel" OR "SIPEKNE'KATIK" OR "Skaru:reh" OR "Sq'éwlets" OR "Stellat'en" OR "Sts'ailes" OR "Stz'uminus" OR "T'it'q'et" OR "Tla'amin" OR "Tl'azt'en" OR "Tl'etinqox" OR "Tsal'alh" OR "Tsideldel" OR "Ts'kw'aylaxw" OR "T'Sou-ke" OR "Ts'uubaa-asatx" OR "Tsuut'ina" OR "We'koqma'q" OR "Wet'suwet'en" OR "Xai'xais" OR "Xaxli'p" OR "Yunesit'in" OR ((Acadia OR Carmacks OR Champagne OR Creek OR Delaware OR Douglas OR Montana OR Similkameen) N2 (Indian OR Indians OR Tribe OR Tribes OR native OR natives OR nation OR people*)) OR Aamjiwnaang OR Abegweit OR Abitibiwinni OR Ahousaht OR Ahtahkakoop OR Aishihik OR Aitchelitz OR Aklavik OR Algonquin* OR Anicinape* OR "Animakee Wa Zhing" OR Anishina* OR aqam OR Atikamekw OR Atjanasca* OR Attawapiskat OR Aundeck-Omni-Kaning OR Batchewana OR "Bearfoot Onondaga" OR Beausoleil OR "Behdzi Ahda*" OR "Biigtigong Nishnaabeg" OR "Biinjitiwaabik Zaaging Anishinaabek" OR Bilijk OR "Binche Whut'en" OR "Bingwi Neyaashi Anishinaabek" OR "Canupawakpa Dakota" OR Cayuga OR Chawathil OR Cheam OR "Cheslatta Carrier" OR Chiniki OR Chipew* OR Cote OR Couchiching OR Cowessess OR Cowichan OR Cree OR "Dakota Plains" OR "Dakota Tipi" OR "Dechi Laot'i" OR Deline OR Dena OR Dene OR Denesuline OR "Deninu K'ue" OR Ditidaht OR "Dog Rib Rae" OR Dokis OR Eabametoong OR Ehattesaht OR Elsipogtog OR Ermineskin OR Esdilagh OR Esgenoopetitj OR Eskasoni OR Esquimalt OR Gameti OR Gayogohono OR Ginoogaming OR Gitanmaax OR Gitanyow OR Gitsegukla OR Gitwangak OR Gitxaala OR Glooscap OR Gwawaenuk OR "Gwichya Gwich'in" OR Gwitchin OR Hagwilget OR Haisla OR Halalt OR Haudenosaunee OR Heiltsuk OR Hesquiaht OR Hiawatha OR Homalco OR Hupacasath OR Huu-ay-aht OR Inninuwug OR Innu OR Innue OR Innus OR Inuvik OR Iskatewizaagegan OR Iskut OR "Ka'a'gee Tu" OR Kahkewistahaw OR "Kasabonika Lake" OR Kashechewan OR Katzie OR Kawacatoose OR Kebaowek OR Keeseekoose OR Keeseekoowenin OR Kee-Way-Win OR "Kinistin Saulteaux" OR Kinonjeoshtegon OR Kispiox OR Kitselas OR Kitsumkalum OR Kluane OR Kutchin OR Kwadacha OR Kwakiutl OR "Kwanlin Dun" OR Kwaw-kwaw-Apilt OR Kwiakah OR "Kwikwasut'inuxw Haxwa'mis" OR Kwikwetlem OR "Lax Kw'alaams" OR "Lheidli T'enneh" OR "Liidlii Kue" OR "Listuguj Mi'gmaq" OR Lyackson OR Magnetawan OR Malahat OR Maliseet OR Mamalilikulla OR Matachewan OR Matsqui OR Mattagami OR "M'Chigeeng" OR Membertou OR Metepenagiag OR Metlakatla OR Miawpukek OR Michipicoten OR Micmacs OR Millbrook OR Mishkeegogamang OR Mississauga* OR Mitaanjigamiing OR Mohawk* OR Montagnais OR Moosomin OR Moravian OR Mowachaht OR Muchalaht OR Munsee-Delaware OR Muscowpetung OR Muskoday OR Muskowekwan OR Musqueam OR "Nacho Nyak Dun" OR "Nadleh Whuten" OR Naicatchewenin OR "Nak'azdli Whut'en" OR Nakota OR Naotkamegwanning OR Naskapi OR Natoaganeg OR Nee-Tahi-Buhn OR Nehiyawak OR Nekaneet OR Neskantaga OR Neskonlith OR Nibinamik OR Nigigoonsiminikaaning OR Nipissing OR Nishnaabeg OR Nooaitch OR Nuchatlaht OR Nuxalk OR Ochapowace OR O-Chi-Chak-Ko-Sipi OR Odanak OR Ojibw* OR Okanagan OR Okanese OR Okemasis OR Oneida OR Oromocto OR Osoyoos OR Pabineau OR Pacheedaht OR "Paqtnkek Mi'kmaw" OR Pasqua OR Pauingassi OR Pauquachin OR "Pays Plat" OR Peguis OR "Pehdzeh Ki" OR Pekuakamiulnuatsh OR Penelakut OR Piapot OR Piikani OR Pikangikum OR Pinaymootang OR Potlotek OR "Qalipu Mi'kmaq" OR Qualicum OR Quatsino OR Sagamok OR Sahgaiehcan OR Samahquam OR Saugeen OR Saulteau OR Saulteaux OR Sechelt OR Secwepemc* OR Semiahmoo OR Seneca OR Shackan OR Shamattawa OR Shawanaga OR Sheguiandah OR Sheshegwaning OR Shuswap OR Siksika OR Simpcw OR Sioux OR Siska OR Skatin OR Skawahlook OR Skeetchestn OR Skidegate OR "Skin Tyee" OR Skowkale OR Skownan OR Skuppah OR Skwah OR Snuneymuxw OR Songhees OR Soowahlie OR Splatsin OR Spuzzum OR Squamish OR Squiala OR "Stswecem'c Xget'tem" OR Sumas OR "Ta'an Kwach'an" OR Tagish OR Tahltan OR Takla OR Tallcree OR "Taykwa Tagamou" OR "Teetl'it Gwich'in" OR Temagami OR Thessalon OR Timiskaming OR "Tk'emlúps te Secwépemc" OR Tla-o-qui-aht OR Tlatlasikwala OR Tlingit OR Tlowitsis OR Tootinaowaziibeeng OR Toquaht OR "Tr'ondëk Hwëch'in" OR Tsartlip OR Tsawout OR Tsawwassen OR Tseshaht OR Tseycum OR "Ts'il Kaz Koh" OR Tsleil-Waututh OR Tuscarora OR Tutchone OR Tzeachten OR Uchucklesaht OR Ucluelet OR Ulkatcho OR Wabaseemoong OR Wabauskang OR Wagmatcook OR Wahnapitae OR "Wahpeton Dakota" OR Wahsipekuk OR Wapekeka OR Wasagamack OR Wasauksing OR Waswanipi OR Wawakapewin OR Waywayseecappo OR Webequie OR Weenusk OR Wemotaci OR Wendat OR "Wha Ti" OR Whapmagoostui OR "Whitecap Dakota" OR Wikwemikong OR Witset OR Wolastoqiyik OR Wuikinuxv OR Wunnumin OR "Wuskwi Sipihk" OR Xatsull OR "Xeni Gwet'in" OR Yakweakwioose OR Yekooche OR "Yellow Quill" OR Yellowknife OR Yellowknives OR Zhiibaahaasing) | 3001 |
| S5 | TI("Degexit'an" OR "Yup'ik" OR ("Bear River" N2 (Indian OR Indians OR Tribe OR Tribes OR native OR natives OR nation OR people*)) OR Abenaki OR Achumawi OR Ahtna OR "Akimel Oodham" OR "Akimel O'odham" OR "Akimel Ootham" OR "Akimel O'otham" OR Aleut* OR Algonkian* OR Alsea OR Aluutiq OR Apalachee OR Arikara OR Atakapa OR Athabascan OR Athabaskan OR Athapascan OR Athapaskan OR Bannock OR Bidai OR Biloxi OR Blackfeet OR Blackfoot OR Caddoan OR Calice OR Calusa OR Carrizo OR Cayuse OR Chemakum OR Cheraw OR Cherokee OR Chetco OR Cheyenne OR Chicora OR Chilula OR Chimariko OR Clatskanie OR Coahuiltecan OR "Coeur d'Alene" OR Costanoan OR Cree OR Croatan OR Croatoan OR Cusabo OR Edisto OR Eskaleut* OR Eskimo OR Eskimo-Aleut OR Esselen OR Eyak OR "Fox and Sauk" OR Gabrielino OR "Gros Ventre" OR Guale OR "Gwich'in" OR Halchidhoma OR Hidatsa OR Hitchiti OR Hopi OR Houma OR Huichol* OR Hupa OR Ingalik OR Inupiaq OR Ioway OR Iroquoi* OR Jacome OR Jano OR Jeaga OR Juaneno OR Jumano OR Kalapuya OR Kansa OR Karankawa OR Karok OR Kashaya OR Kato OR Kawaiisu OR Kitanemuk OR Kitsai OR Klallam OR Koasati OR Kolchan OR Konkow OR Koyukon OR Lakota* OR Lassik OR Laurentian OR Lenape OR "Lenni Lenape" OR Mahican OR Mandan OR Maricopa OR Massachusett OR Mattole OR Minisink OR Missouria OR Mohave OR Mohawk OR Mohican OR Molalla OR Multnomah OR Munsee OR Muscoge* OR Naabeeho OR Nakoda OR Nanticoke OR Natchez OR Nauset OR Navaho OR Niantic OR Nipmuc OR Nisenan OR Nongatl OR Oconee OR Ofo OR Ohlone OR Ojibw* OR Okanagan OR Oneida OR Otoe OR Palouse OR Panamint OR Papago OR Patwin OR "Pee Dee" OR Pennacook OR Pequot OR Pima OR Piro OR Pocasset OR Pocomtuc OR Pokanoket OR Poospatuck OR Powhatan OR Quechan OR Quileute OR Quinault OR Quiripi OR Salinan OR Samich OR Saponi OR Serrano OR Shasta OR Shawnee OR Siksikaitsitapi OR Sinkyone OR Siouan OR Sioux OR Snohomish OR Snoqualmie OR Squaxin OR Suma OR Susquehannock OR Takelma OR Tanacross OR Tanana OR Tataviam OR Tawakoni OR Tequesta OR Tillamook OR Timucua OR Tlingit OR "Tohono Oodham" OR Tongva OR Tsimshian OR Tubatulabal OR Tutelo OR Tututni OR Unalachtigo OR Unami OR Unangan OR Unangas OR Unkechaug OR Waccamaw OR Walapai OR Wappo OR Wasco OR Whilkut OR Wichita OR Winnebago OR Wintu OR Wishram OR Yahi OR Yakima OR Yamasee OR Yana OR Yaqu OR Yuchi OR Yuki OR Yuma OR Yupik OR Yurok) OR AB("Degexit'an" OR "Yup'ik" OR ("Bear River" N2 (Indian OR Indians OR Tribe OR Tribes OR native OR natives OR nation OR people*)) OR Abenaki OR Achumawi OR Ahtna OR "Akimel Oodham" OR "Akimel O'odham" OR "Akimel Ootham" OR "Akimel O'otham" OR Aleut* OR Algonkian* OR Alsea OR Aluutiq OR Apalachee OR Arikara OR Atakapa OR Athabascan OR Athabaskan OR Athapascan OR Athapaskan OR Bannock OR Bidai OR Biloxi OR Blackfeet OR Blackfoot OR Caddoan OR Calice OR Calusa OR Carrizo OR Cayuse OR Chemakum OR Cheraw OR Cherokee OR Chetco OR Cheyenne OR Chicora OR Chilula OR Chimariko OR Clatskanie OR Coahuiltecan OR "Coeur d'Alene" OR Costanoan OR Cree OR Croatan OR Croatoan OR Cusabo OR Edisto OR Eskaleut* OR Eskimo OR Eskimo-Aleut OR Esselen OR Eyak OR "Fox and Sauk" OR Gabrielino OR "Gros Ventre" OR Guale OR "Gwich'in" OR Halchidhoma OR Hidatsa OR Hitchiti OR Hopi OR Houma OR Huichol* OR Hupa OR Ingalik OR Inupiaq OR Ioway OR Iroquoi* OR Jacome OR Jano OR Jeaga OR Juaneno OR Jumano OR Kalapuya OR Kansa OR Karankawa OR Karok OR Kashaya OR Kato OR Kawaiisu OR Kitanemuk OR Kitsai OR Klallam OR Koasati OR Kolchan OR Konkow OR Koyukon OR Lakota* OR Lassik OR Laurentian OR Lenape OR "Lenni Lenape" OR Mahican OR Mandan OR Maricopa OR Massachusett OR Mattole OR Minisink OR Missouria OR Mohave OR Mohawk OR Mohican OR Molalla OR Multnomah OR Munsee OR Muscoge* OR Naabeeho OR Nakoda OR Nanticoke OR Natchez OR Nauset OR Navaho OR Niantic OR Nipmuc OR Nisenan OR Nongatl OR Oconee OR Ofo OR Ohlone OR Ojibw* OR Okanagan OR Oneida OR Otoe OR Palouse OR Panamint OR Papago OR Patwin OR "Pee Dee" OR Pennacook OR Pequot OR Pima OR Piro OR Pocasset OR Pocomtuc OR Pokanoket OR Poospatuck OR Powhatan OR Quechan OR Quileute OR Quinault OR Quiripi OR Salinan OR Samich OR Saponi OR Serrano OR Shasta OR Shawnee OR Siksikaitsitapi OR Sinkyone OR Siouan OR Sioux OR Snohomish OR Snoqualmie OR Squaxin OR Suma OR Susquehannock OR Takelma OR Tanacross OR Tanana OR Tataviam OR Tawakoni OR Tequesta OR Tillamook OR Timucua OR Tlingit OR "Tohono Oodham" OR Tongva OR Tsimshian OR Tubatulabal OR Tutelo OR Tututni OR Unalachtigo OR Unami OR Unangan OR Unangas OR Unkechaug OR Waccamaw OR Walapai OR Wappo OR Wasco OR Whilkut OR Wichita OR Winnebago OR Wintu OR Wishram OR Yahi OR Yakima OR Yamasee OR Yana OR Yaqu OR Yuchi OR Yuki OR Yuma OR Yupik OR Yurok) | 11916 |
| S4 | TI("Asa'carsarmiut" OR "Dee-ni'" OR "Mi'kmaq" OR "S'Klallam" OR "Shoonaq'" OR "Sun'aq" OR (("Ivanof Bay" OR "Shoalwater Bay" OR "Spirit Lake" OR "Squaxin Island" OR Dakota OR Dakotah OR Wichita OR keechi OR waco OR Tawakonie OR Cheyenne OR Creek OR Crow OR Hoonah OR Kasigluk OR Maricopa OR Miami OR Omaha OR Oneida OR Ottawa OR Peoria OR Salamatof OR Sitka OR Snoqualmie OR Spokane OR Winnebago OR Wyandotte) N2 (Indian OR Indians OR Tribe OR Tribes OR native OR natives OR nation OR people*)) OR ((Native OR indian OR traditional) N2 (village OR council OR community OR association)) OR Absentee-Shawnee OR Achomawi OR Agdaagux OR "Agua Caliente" OR Ajumawi OR Ak-Chin OR Akiachak OR Akiak OR Alabama-Coushatta OR Alabama-Quassarte OR Alatna OR Algaaciq OR Algin OR Allakaket OR "Alturas Indian Rancheria" OR Alutiiq OR Angoon OR Anvik OR Apache OR Aporige OR Aquinnah OR arapaho OR Aroostook OR Assiniboine OR Astarawi OR Atsugewi OR Atwamsini OR Bdewakantunwan OR Caddo OR Cahto OR Cahuilla OR Catawba OR Cayuga OR Chalkyitsik OR Cheesh-Na OR Chehalis OR Chemehuevi OR Cher-Ae OR Chevak OR Cheyenne-Arapaho OR Chickahominy OR Chickaloon OR Chickasaw OR Chignik OR Chilkat OR Chilkoot OR Chinik OR Chinook OR Chippewa* OR Chitimacha OR Choctaw OR Chukchansi OR Chuloonawick OR Chumash OR Cocopah OR Colusa OR Comanche OR Coos OR Cosmit OR Coushatta OR Cowlitz OR "Cross Village" OR Cupeno OR Curyung OR Diegueno OR Dine' OR Egegik OR Eklutna OR Elwha OR "Emmonak Village" OR "Evansville Village" OR Ewiiaapaayp OR "Galena Village" OR Goshute OR "Gulkana Village Council" OR Haida OR Hammawi OR Havasupai OR "Healy Lake Village" OR Hewisedawi OR Ho-Chunk OR Hoh OR Hokan OR Holikachuk OR "Holy Cross Tribe" OR Hualapai OR "Hughes Village" OR "Huslia Village" OR "Hydaburg Cooperative Association" OR "Igiugig Village" OR Iipay OR Ilmawi OR Inaja OR Inupiat OR Ione OR Itsatawi OR "Kaguyak Village" OR Kaibab OR "Kaktovik Village" OR Kalispel OR Kanosh OR Karuk OR Kashia OR Kaw OR Keetoowah OR "Kenaitze Indian Tribe" OR "Ketchikan Indian Community" OR "Keweenaw Bay" OR Kialegee OR Kickapoo OR "King Island Native Community" OR "King Salmon Tribe" OR Kiowa OR Klamath OR "Klawock Cooperative Association" OR Kletsel OR "Knik Tribe" OR "Koi Nation" OR "Kokhanok Village" OR Koosharem OR Kootenai OR Kosalektawi OR "Koyukuk Native Village" OR Kumeyaay OR Kwalhioqua OR "Lesnoi Village" OR "Levelock Village" OR "Lime Village" OR "Louden Village" OR Luiseno OR Lummi OR Madesi OR Maidu OR Makah OR Maliseet OR "Manley Hot Springs Village" OR "Manokotak Village" OR Manzanita OR Mashantucket OR Match-e-be-nash-she-wish OR Mattaponi OR "McGrath Native Village" OR Mdewakanton OR Mechoopda OR Menominee OR Me-Wuk OR Miccosukee OR Micmacs OR "Mission Indians" OR Miwok OR Miwuk OR Modoc OR Mohegan OR Mojave OR Monacan OR "Mono Indians" OR Muckleshoot OR Muscogee OR "Na Dené" OR Nansemond OR Narragansett OR Navajo OR "New Koliganek Village" OR "New Stuyahok Village" OR "Newhalen Village" OR "Newtok Village" OR "Nez Perce" OR "Nikolai Village" OR "Ninilchik Village" OR Nisqually OR Nomlaki OR "Nondalton Village" OR Nooksack OR "Northway Village" OR Nottawaseppi OR "Nulato Village" OR Nunakauyarmiut OR Odawa OR Ohgsenakale OR "Ohkay Owingeh" OR Onondaga OR Oreilles OR "Organized Village of Grayling" OR "Organized Village of Kake" OR "Organized Village of Kasaan" OR "Organized Village of Kwethluk" OR "Organized Village of Saxman" OR Orutsararmiut OR Osage OR Otoe-Missouria OR Paiute OR Paiute-Shoshone OR Pala OR Pamunkey OR Pascua OR Paskenta OR Passamaquoddy OR "Pauloff Harbor Village" OR Pauma OR Pawnee OR Pechanga OR "Pechanga " OR "Peck Indian Reservation" OR "Pedro Bay Village" OR Pee-Posh OR Penobscot OR Penutian OR Perquot OR "Pima People" OR Pima-Maricopa OR Poarch OR Pokagon OR Pomo OR Ponca OR "Portage Creek Village" OR Potawatomi OR Pottawatomi OR Pueblo OR Puyallup OR "Qagan Tayagungin" OR Qawalangin OR Quapaw OR Quechan OR "Rampart Village" OR Rappahannock OR Resighini OR "Sac & Fox" OR "Sac and Fox" OR "Saint George Island" OR "Saint Paul Island" OR Salish OR Samish OR Santee OR Sauk-Suiattle OR "Seldovia Village Tribe" OR Seminole OR Seneca OR Seneca-Cayuga OR Shinnecock OR Shivwits OR Shoshone OR Shoshone-Bannock OR Shoshone-Paiute OR Siuslaw OR Skagit OR "Skagway Village" OR Skokomish OR "South Naknek Village" OR "Stebbins Community Association" OR Stillaguamish OR Suquamish OR Swinomish OR "Takotna Village" OR Tangirnaq OR Tejon OR "Telida Village" OR Te-Moak OR Thlopthlocco OR "Tlingit & Haida" OR "Tohono O'odham" OR Tolowa OR Tonawanda OR Tonkawa OR Tulalip OR Tunica-Biloxi OR Tuolumne OR Tuscarora OR "Twin Hills Village" OR "Ugashik Village" OR Umatilla OR Umkumiut OR Umpqua OR Ute OR "Utu Utu Gwaitu" OR "Village of Alakanuk" OR "Village of Anaktuvuk Pass" OR "Village of Aniak" OR "Village of Atmautluak" OR "Village of Bill Moore's Slough" OR "Village of Chefornak" OR "Village of Clarks Point" OR "Village of Crooked Creek" OR "Village of Dot Lake" OR "Village of Iliamna" OR "Village of Kalskag" OR "Village of Kaltag" OR "Village of Kotlik" OR "Village of Lower Kalskag" OR "Village of Ohogamiut" OR "Village of Red Devil" OR "Village of Sleetmute" OR "Village of Solomon" OR "Village of Stony River" OR "Village of Venetie" OR "Village of Wainwright" OR Wailaki OR Wampanoag OR "Warm Springs Reservation" OR Washoe OR Willapa OR Winnemucca OR Wintun OR Wiyot OR "Wrangell Cooperative Association" OR Yakama OR Yakutat OR Yaqui OR Yavapai OR Yokuts OR Yuhaaviatam OR Yuima OR Yukian OR Yukut OR Yupiit OR Zuni) OR AB("Asa'carsarmiut" OR "Dee-ni'" OR "Mi'kmaq" OR "S'Klallam" OR "Shoonaq'" OR "Sun'aq" OR (("Ivanof Bay" OR "Shoalwater Bay" OR "Spirit Lake" OR "Squaxin Island" OR Dakota OR Dakotah OR Wichita OR keechi OR waco OR Tawakonie OR Cheyenne OR Creek OR Crow OR Hoonah OR Kasigluk OR Maricopa OR Miami OR Omaha OR Oneida OR Ottawa OR Peoria OR Salamatof OR Sitka OR Snoqualmie OR Spokane OR Winnebago OR Wyandotte) N2 (Indian OR Indians OR Tribe OR Tribes OR native OR natives OR nation OR people*)) OR ((Native OR indian OR traditional) N2 (village OR council OR community OR association)) OR Absentee-Shawnee OR Achomawi OR Agdaagux OR "Agua Caliente" OR Ajumawi OR Ak-Chin OR Akiachak OR Akiak OR Alabama-Coushatta OR Alabama-Quassarte OR Alatna OR Algaaciq OR Algin OR Allakaket OR "Alturas Indian Rancheria" OR Alutiiq OR Angoon OR Anvik OR Apache OR Aporige OR Aquinnah OR arapaho OR Aroostook OR Assiniboine OR Astarawi OR Atsugewi OR Atwamsini OR Bdewakantunwan OR Caddo OR Cahto OR Cahuilla OR Catawba OR Cayuga OR Chalkyitsik OR Cheesh-Na OR Chehalis OR Chemehuevi OR Cher-Ae OR Chevak OR Cheyenne-Arapaho OR Chickahominy OR Chickaloon OR Chickasaw OR Chignik OR Chilkat OR Chilkoot OR Chinik OR Chinook OR Chippewa* OR Chitimacha OR Choctaw OR Chukchansi OR Chuloonawick OR Chumash OR Cocopah OR Colusa OR Comanche OR Coos OR Cosmit OR Coushatta OR Cowlitz OR "Cross Village" OR Cupeno OR Curyung OR Diegueno OR Dine' OR Egegik OR Eklutna OR Elwha OR "Emmonak Village" OR "Evansville Village" OR Ewiiaapaayp OR "Galena Village" OR Goshute OR "Gulkana Village Council" OR Haida OR Hammawi OR Havasupai OR "Healy Lake Village" OR Hewisedawi OR Ho-Chunk OR Hoh OR Hokan OR Holikachuk OR "Holy Cross Tribe" OR Hualapai OR "Hughes Village" OR "Huslia Village" OR "Hydaburg Cooperative Association" OR "Igiugig Village" OR Iipay OR Ilmawi OR Inaja OR Inupiat OR Ione OR Itsatawi OR "Kaguyak Village" OR Kaibab OR "Kaktovik Village" OR Kalispel OR Kanosh OR Karuk OR Kashia OR Kaw OR Keetoowah OR "Kenaitze Indian Tribe" OR "Ketchikan Indian Community" OR "Keweenaw Bay" OR Kialegee OR Kickapoo OR "King Island Native Community" OR "King Salmon Tribe" OR Kiowa OR Klamath OR "Klawock Cooperative Association" OR Kletsel OR "Knik Tribe" OR "Koi Nation" OR "Kokhanok Village" OR Koosharem OR Kootenai OR Kosalektawi OR "Koyukuk Native Village" OR Kumeyaay OR Kwalhioqua OR "Lesnoi Village" OR "Levelock Village" OR "Lime Village" OR "Louden Village" OR Luiseno OR Lummi OR Madesi OR Maidu OR Makah OR Maliseet OR "Manley Hot Springs Village" OR "Manokotak Village" OR Manzanita OR Mashantucket OR Match-e-be-nash-she-wish OR Mattaponi OR "McGrath Native Village" OR Mdewakanton OR Mechoopda OR Menominee OR Me-Wuk OR Miccosukee OR Micmacs OR "Mission Indians" OR Miwok OR Miwuk OR Modoc OR Mohegan OR Mojave OR Monacan OR "Mono Indians" OR Muckleshoot OR Muscogee OR "Na Dené" OR Nansemond OR Narragansett OR Navajo OR "New Koliganek Village" OR "New Stuyahok Village" OR "Newhalen Village" OR "Newtok Village" OR "Nez Perce" OR "Nikolai Village" OR "Ninilchik Village" OR Nisqually OR Nomlaki OR "Nondalton Village" OR Nooksack OR "Northway Village" OR Nottawaseppi OR "Nulato Village" OR Nunakauyarmiut OR Odawa OR Ohgsenakale OR "Ohkay Owingeh" OR Onondaga OR Oreilles OR "Organized Village of Grayling" OR "Organized Village of Kake" OR "Organized Village of Kasaan" OR "Organized Village of Kwethluk" OR "Organized Village of Saxman" OR Orutsararmiut OR Osage OR Otoe-Missouria OR Paiute OR Paiute-Shoshone OR Pala OR Pamunkey OR Pascua OR Paskenta OR Passamaquoddy OR "Pauloff Harbor Village" OR Pauma OR Pawnee OR Pechanga OR "Pechanga " OR "Peck Indian Reservation" OR "Pedro Bay Village" OR Pee-Posh OR Penobscot OR Penutian OR Perquot OR "Pima People" OR Pima-Maricopa OR Poarch OR Pokagon OR Pomo OR Ponca OR "Portage Creek Village" OR Potawatomi OR Pottawatomi OR Pueblo OR Puyallup OR "Qagan Tayagungin" OR Qawalangin OR Quapaw OR Quechan OR "Rampart Village" OR Rappahannock OR Resighini OR "Sac & Fox" OR "Sac and Fox" OR "Saint George Island" OR "Saint Paul Island" OR Salish OR Samish OR Santee OR Sauk-Suiattle OR "Seldovia Village Tribe" OR Seminole OR Seneca OR Seneca-Cayuga OR Shinnecock OR Shivwits OR Shoshone OR Shoshone-Bannock OR Shoshone-Paiute OR Siuslaw OR Skagit OR "Skagway Village" OR Skokomish OR "South Naknek Village" OR "Stebbins Community Association" OR Stillaguamish OR Suquamish OR Swinomish OR "Takotna Village" OR Tangirnaq OR Tejon OR "Telida Village" OR Te-Moak OR Thlopthlocco OR "Tlingit & Haida" OR "Tohono O'odham" OR Tolowa OR Tonawanda OR Tonkawa OR Tulalip OR Tunica-Biloxi OR Tuolumne OR Tuscarora OR "Twin Hills Village" OR "Ugashik Village" OR Umatilla OR Umkumiut OR Umpqua OR Ute OR "Utu Utu Gwaitu" OR "Village of Alakanuk" OR "Village of Anaktuvuk Pass" OR "Village of Aniak" OR "Village of Atmautluak" OR "Village of Bill Moore's Slough" OR "Village of Chefornak" OR "Village of Clarks Point" OR "Village of Crooked Creek" OR "Village of Dot Lake" OR "Village of Iliamna" OR "Village of Kalskag" OR "Village of Kaltag" OR "Village of Kotlik" OR "Village of Lower Kalskag" OR "Village of Ohogamiut" OR "Village of Red Devil" OR "Village of Sleetmute" OR "Village of Solomon" OR "Village of Stony River" OR "Village of Venetie" OR "Village of Wainwright" OR Wailaki OR Wampanoag OR "Warm Springs Reservation" OR Washoe OR Willapa OR Winnemucca OR Wintun OR Wiyot OR "Wrangell Cooperative Association" OR Yakama OR Yakutat OR Yaqui OR Yavapai OR Yokuts OR Yuhaaviatam OR Yuima OR Yukian OR Yukut OR Yupiit OR Zuni) | 6723 |
| S3 | DE("Inuit") or TI(inuit or (arctic N2 people*)) | 613 |
| S2 | TI(((Canada OR Canadian*) N2 (native* OR indigenous OR aborigin*)) OR Metis OR "Métis" OR Michif* OR Mitchif OR Mechif OR "Métif" OR "Métchif" OR Metif OR Metchif OR Inuit OR "First Nations" OR "First Nation") OR AB(((Canada OR Canadian*) N2 (native* OR indigenous OR aborigin*)) OR Metis OR "Métis" OR Michif* OR Mitchif OR Mechif OR "Métif" OR "Métchif" OR Metif OR Metchif OR Inuit OR "First Nations" OR "First Nation") | 2323 |
| S1 | DE("American Indians" OR "Alaska Natives") OR TI(((American* OR Alaska*) N2 (Native* OR Indian OR Indians OR indigenous OR aborigin*)) OR Amerind*) OR AB(((American* OR Alaska*) N2 (Native* OR Indian OR Indians OR indigenous OR aborigin*)) OR Amerind*) | 14557 |

### Alt Health Watch (EBSCOhost)

**Field Codes:** TI = title, AB= abstract, DE = subjects [exact].

**Proximity operator:** N# 
**Truncation:**  *

**Database** - Alt HealthWatch

**Interface** - EBSCOhost Research Databases

**Search Screen** - Basic Search

**Search modes** – Proximity

**Date**: Thursday, June 08, 2023 12:37:13 AM

| **#** | **Query** | **Limiters/Expanders** | **Results** |
| --- | --- | --- | --- |
| S29 | S28 | Publication Date: 20230101-20241231 | 0 |
| S28 | S22 OR S23 OR S24 OR S25 OR S26 OR S27 |  | 54 |
| S27 | ((S16 AND S13) OR ((S13 AND S15) AND (S8 OR S9)) |  | 2 |
| S26 | S19 AND (S12 OR S17) AND S20 |  | 28 |
| S25 | (S8 OR S9) AND S21 |  | 7 |
| S24 | S18 AND S12 |  | 0 |
| S23 | S9 AND S12 |  | 12 |
| S22 | S8 AND S12 |  | 8 |
| S21 | TI((women* OR woman* OR female*) N2 (health OR healthcare OR service* OR clinic OR clinics)) OR AB((women* OR woman* OR female*) N2 (health OR healthcare OR service* OR clinic OR clinics)) |  | 1709 |
| S20 | DE("NORTH Americans" OR "AMERICANS" OR "CANADIANS" OR "CANADIAN provinces" OR "U.S. states") OR TI("North America" OR Canada OR Alberta OR "British Columbia" OR Manitoba OR "New Brunswick" OR "Newfoundland" OR "Labrador" OR "Northwest Territories" OR "Nova Scotia" OR Nunavut OR Ontario OR "Prince Edward Island" OR Quebec OR Saskatchewan OR Yukon OR "United States" OR Appalachia* OR "New England" OR "Pacific States" OR Alabama OR Alaska OR Arizona OR Arkansas OR California OR Colorado OR Connecticut OR Delaware OR Florida OR Georgia OR Hawaii OR Idaho OR Illinois OR Indiana OR Iowa OR Kansas OR Kentucky OR Louisiana OR Maine OR Maryland OR Massachusetts OR Michigan OR Minnesota OR Mississippi OR Missouri OR Montana OR Nebraska OR Nevada OR "New Hampshire" OR "New Jersey" OR "New Mexico" OR "New York" OR "North Carolina" OR "North Dakota" OR Ohio OR Oklahoma OR Oregon OR Pennsylvania OR "Rhode Island" OR "South Carolina" OR "South Dakota" OR Tennessee OR Texas OR Utah OR Vermont OR Virginia OR Washington OR "West Virginia" OR Wisconsin OR Wyoming OR "District of Columbia" OR "Washington DC") OR AB("North America" OR Canada OR Alberta OR "British Columbia" OR Manitoba OR "New Brunswick" OR "Newfoundland" OR "Labrador" OR "Northwest Territories" OR "Nova Scotia" OR Nunavut OR Ontario OR "Prince Edward Island" OR Quebec OR Saskatchewan OR Yukon OR "United States" OR Appalachia* OR "New England" OR "Pacific States" OR Alabama OR Alaska OR Arizona OR Arkansas OR California OR Colorado OR Connecticut OR Delaware OR Florida OR Georgia OR Hawaii OR Idaho OR Illinois OR Indiana OR Iowa OR Kansas OR Kentucky OR Louisiana OR Maine OR Maryland OR Massachusetts OR Michigan OR Minnesota OR Mississippi OR Missouri OR Montana OR Nebraska OR Nevada OR "New Hampshire" OR "New Jersey" OR "New Mexico" OR "New York" OR "North Carolina" OR "North Dakota" OR Ohio OR Oklahoma OR Oregon OR Pennsylvania OR "Rhode Island" OR "South Carolina" OR "South Dakota" OR Tennessee OR Texas OR Utah OR Vermont OR Virginia OR Washington OR "West Virginia" OR Wisconsin OR Wyoming OR "District of Columbia" OR "Washington DC") |  | 25949 |
| S19 | DE("ALTERNATIVE medicine" OR "TRADITIONAL medicine" OR "HEALING gods" OR "HERBAL medicine") OR TI(((Complementary OR alternative OR traditional) N2 (medicine* OR therap* OR healing OR treatment*)) OR "Herbal Medicine" OR herbalism OR ((botanical OR ethnobotanical OR herb* OR plant OR plant-based) N2 (medicine* OR remedy OR remedies OR healing OR therap* OR treatment*)) OR phyto-medicine OR phytomedic* OR "folk medicine" OR "folk healer") OR AB(((Complementary OR alternative OR traditional) N2 (medicine* OR therap* OR healing OR treatment*)) OR "Herbal Medicine" OR herbalism OR ((botanical OR ethnobotanical OR herb* OR plant OR plant-based) N2 (medicine* OR remedy OR remedies OR healing OR therap* OR treatment*)) OR phyto-medicine OR phytomedic* OR "folk medicine" OR "folk healer") |  | 26968 |
| S18 | DE("NATIVE American medicine" OR "NATIVE American traditional medicine" OR "NATIVE American healers" OR "SHAMANS" OR "MALE shamans" OR "WOMEN shamans") OR TI(((Indigenous OR native OR "first nation" OR aborigin* OR Indian) N2 (health OR healthcare OR healing OR medicine OR practices)) OR ethnomedicin* OR "native healing" OR Limpia OR "indigenous healer" OR medicineman OR medicinemen OR "medicine man" OR "medicine men" OR Curandero OR "folk healer" OR shaman* OR shamen) OR AB(((Indigenous OR native OR "first nation" OR aborigin* OR Indian) N2 (health OR healthcare OR healing OR medicine OR practices)) OR ethnomedicin* OR "native healing" OR Limpia OR "indigenous healer" OR medicineman OR medicinemen OR "medicine man" OR "medicine men" OR Curandero OR "folk healer" OR shaman* OR shamen) |  | 828 |
| S17 | (S15 OR S16) AND S13 |  | 56 |
| S16 | DE("NATIVE American women" OR "ALASKA Native women" OR "ATHAPASCAN women" OR "CHINOOK women" OR "CHOCTAW women" OR "CHUMASH women" OR "CREEK women" OR OR "HIDATSA women" OR "IROQUOIAN women" OR "LAGUNA women" OR "MANDAN women" OR "MARICOPA women" OR "METIS women" OR "NATIVE American mothers" OR "NORTHERN Paiute women" OR "OGLALA women" OR "OJIBWA women" OR "OMAHA women" OR "PAIUTE women" OR "PENOBSCOT women" OR "PIMA women" OR "POMO women" OR "POWHATAN women" OR "PUEBLO women" OR "SEMINOLE women" OR "SENECA women" OR "SHOSHONI women" OR "TEWA women" OR "TLAKLUIT women" OR "TOHONO O'odham women" OR "UTE women" OR "WASHOE women" OR "WINNEBAGO women" OR "YAKAMA women" OR "YANKTON women" OR "YUROK women") |  | 2 |
| S15 | DE(WOMEN) OR TI(female OR woman OR women) OR AB(female OR woman OR women) |  | 20117 |
| S14 | DE("OLDER Alaska Natives" OR "OLDER Native Americans") |  | 250 |
| S13 | DE("OLDER people") OR TI(elder OR elders) OR AB(elder OR elders) |  | 589 |
| S12 | S10 OR S11 |  | 3060 |
| S11 | DE("HOT flashes" OR "NIGHT sweats") OR TI("Hot Flashes" OR "Hot Flash*" OR "Hot Flush*" OR "night sweat*" OR "nocturnal hyperhidrosis" OR "vasomotor symptom*" OR VMS) OR AB("Hot Flashes" OR "Hot Flash*" OR "Hot Flush*" OR "night sweat*" OR "nocturnal hyperhidrosis" OR "vasomotor symptom*" OR VMS) |  | 524 |
| S10 | DE("CLIMACTERIC" OR "MENOPAUSE" OR "PERIMENOPAUSE" OR "POSTMENOPAUSE" OR "PREMATURE menopause") OR TI(Climacteric OR Climacterium OR Menopause OR Menopaus* OR "Menopause, Premature" OR "climacterium praecox" OR Perimenopause OR Perimenopaus* OR Postmenopause OR Postmenopaus* OR "Post menopaus*" OR Post-menopaus* OR Premenopause OR Premenopause* OR Pre-menopaus* OR "Pre menopaus*" OR Climacteric* OR "Life Change*" OR "Change of life") OR AB(Climacteric OR Climacterium OR Menopause OR Menopaus* OR "Menopause, Premature" OR "climacterium praecox" OR Perimenopause OR Perimenopaus* OR Postmenopause OR Postmenopaus* OR "Post menopaus*" OR Post-menopaus* OR Premenopause OR Premenopause* OR Pre-menopaus* OR "Pre menopaus*" OR Climacteric* OR "Life Change*" OR "Change of life") |  | 2851 |
| S9 | S3 OR S4 OR S5 OR S6 OR S7 |  | 1564 |
| S8 | S1 OR S2 |  | 360 |
| S7 | TI("Kwakwaka'wakw" OR "Kwak'wala" OR "L'uml'umuluts" OR "Lheidli T'enneh " OR "Nlaka'pamux" OR "Nuxálk" OR "Quw'utsun" OR "Qwum'yiqun" OR "S'amuna" OR "Sm'algyax" OR "St'at'imc" OR "Stl'atl'imx" OR "Sto:lo" OR "Tl'ulpalus" OR "Tsilhqot'in" OR "T'Souke" OR "Xa'xtsa" OR (Labrador N2 (Indian OR Indians OR Tribe OR Tribes OR native OR natives OR nation OR people*)) OR "Áa Tlein Kwáan" OR Aivilingmiut OR Algonkian OR Algonquian OR Algonquin OR Anishinaabe OR "Atlin people" OR "Baffin Island" OR "Bella Coola" OR Burrard OR Cheklesahht OR Chilcotin OR Chippew* OR Clayoquot OR Clemclemaluts OR Comiaken OR Comox OR Coquitlam OR Dakelh OR Dane-zaa OR "Deisleen Kwáan" OR Dunne-za OR Gitxsan OR Gitxsanimaax OR Guyohkohnyo OR "Haad kil" OR Halkomelem OR Halqemeylem OR Henaksiala OR Hinupsum OR Hunquminum OR Hwlitsum OR Hwulqwselu OR Hydin OR Iglulik OR Iglulingmuit OR Ininew OR Inuinnait OR Inuk OR Inupiat OR Inuvialuit OR Iroquois OR Kainai OR Kaska OR Kelsemeht OR Khenipsen OR Kilpahlas OR Kimsquit OR Kitamaat OR Kitlinermiut OR Kivallirmiut OR Klahoose OR Koksilah OR Kootenay OR Koskimo OR Ktunaxa OR Kwantlen OR Kwatna OR Kyuquot OR Labradormiut OR Laich-kwil-tach OR Lamalch* OR Lekwungen OR Lillooet OR Loucheaux OR Lummi OR Micmac OR Mississaugas OR Mohawk OR Mowachaht-Muchalaht OR Nadene OR Na-Dene OR Namgis OR Nanaimo OR Nanoose OR Nequatque OR Netsilik OR Netsilingmiut OR Niitsitapi OR Nimpkish OR Nootka OR Nunavimiut OR Nunavimmiut OR Nuu-chah-nulth OR Odawa OR Ohiaht OR Oji-cree OR Onayotekaono OR Onondaga OR Onondowahgah OR Onundagaono OR Opetchesaht OR Owekeeno OR Passamaquoddy OR Peigan OR Petun OR Plains-Cree OR Popkum OR Potawatomi OR Quamichan OR Sahtu OR Salish OR Sarcee OR Scowlitz OR "Seabird Island" OR Secwepemc OR Sekani OR Semahquam OR Shishalh OR Sinixt OR Ska-Ruh-Reh OR Skway OR Sliammon OR Snaw-naw-as OR Somena OR Songish OR Sooke OR Stoney OR Stuie OR Tallheo OR Tasttine OR Teslin OR Tlicho OR Tsimshianic OR Tununirmiut OR Ungava OR Wakashan OR Weewaikai OR Wewaykum OR Wyandot OR Yuculta) OR AB("Kwakwaka'wakw" OR "Kwak'wala" OR "L'uml'umuluts" OR "Lheidli T'enneh " OR "Nlaka'pamux" OR "Nuxálk" OR "Quw'utsun" OR "Qwum'yiqun" OR "S'amuna" OR "Sm'algyax" OR "St'at'imc" OR "Stl'atl'imx" OR "Sto:lo" OR "Tl'ulpalus" OR "Tsilhqot'in" OR "T'Souke" OR "Xa'xtsa" OR (Labrador N2 (Indian OR Indians OR Tribe OR Tribes OR native OR natives OR nation OR people*)) OR "Áa Tlein Kwáan" OR Aivilingmiut OR Algonkian OR Algonquian OR Algonquin OR Anishinaabe OR "Atlin people" OR "Baffin Island" OR "Bella Coola" OR Burrard OR Cheklesahht OR Chilcotin OR Chippew* OR Clayoquot OR Clemclemaluts OR Comiaken OR Comox OR Coquitlam OR Dakelh OR Dane-zaa OR "Deisleen Kwáan" OR Dunne-za OR Gitxsan OR Gitxsanimaax OR Guyohkohnyo OR "Haad kil" OR Halkomelem OR Halqemeylem OR Henaksiala OR Hinupsum OR Hunquminum OR Hwlitsum OR Hwulqwselu OR Hydin OR Iglulik OR Iglulingmuit OR Ininew OR Inuinnait OR Inuk OR Inupiat OR Inuvialuit OR Iroquois OR Kainai OR Kaska OR Kelsemeht OR Khenipsen OR Kilpahlas OR Kimsquit OR Kitamaat OR Kitlinermiut OR Kivallirmiut OR Klahoose OR Koksilah OR Kootenay OR Koskimo OR Ktunaxa OR Kwantlen OR Kwatna OR Kyuquot OR Labradormiut OR Laich-kwil-tach OR Lamalch* OR Lekwungen OR Lillooet OR Loucheaux OR Lummi OR Micmac OR Mississaugas OR Mohawk OR Mowachaht-Muchalaht OR Nadene OR Na-Dene OR Namgis OR Nanaimo OR Nanoose OR Nequatque OR Netsilik OR Netsilingmiut OR Niitsitapi OR Nimpkish OR Nootka OR Nunavimiut OR Nunavimmiut OR Nuu-chah-nulth OR Odawa OR Ohiaht OR Oji-cree OR Onayotekaono OR Onondaga OR Onondowahgah OR Onundagaono OR Opetchesaht OR Owekeeno OR Passamaquoddy OR Peigan OR Petun OR Plains-Cree OR Popkum OR Potawatomi OR Quamichan OR Sahtu OR Salish OR Sarcee OR Scowlitz OR "Seabird Island" OR Secwepemc OR Sekani OR Semahquam OR Shishalh OR Sinixt OR Ska-Ruh-Reh OR Skway OR Sliammon OR Snaw-naw-as OR Somena OR Songish OR Sooke OR Stoney OR Stuie OR Tallheo OR Tasttine OR Teslin OR Tlicho OR Tsimshianic OR Tununirmiut OR Ungava OR Wakashan OR Weewaikai OR Wewaykum OR Wyandot OR Yuculta) |  | 42 |
| S6 | TI(Anishnawbek OR "?Akisq'nuk" OR "?aq'am" OR "?Esdilagh" OR "Abénakis" OR "Akisq'nuk" OR "aq'am" OR "Che:k:tles7et'h'" OR "Da'naxda'xw" OR "Dzawada'enuxw" OR "Esk'etemc" OR "Gitga'at" OR "Gitlaxt'aamiks" OR "Gwa'Sala-Nakwaxda'xw" OR "Gwich'in" OR "Ka:'yu:'k't'h'" OR "Kanyen'kehaka" OR "Kapawe'no" OR "K'atlodeeche" OR "K'ómoks" OR "Leq'á:mel" OR "Lil'wat" OR "Malécite" OR "Mi'kmaq" OR "Mowachaht/Muchalaht" OR "Nisga'a" OR "N'Quatqua" OR "O'Chiese" OR "Ononda'gega'" OR "Onondowága'" OR "Onyota'a:ka" OR "Saik'uz" OR "Sambaa K'e " OR "Shxwhá:y" OR "Shxw'ow'hamel" OR "SIPEKNE'KATIK" OR "Skaru:reh" OR "Sq'éwlets" OR "Stellat'en" OR "Sts'ailes" OR "Stz'uminus" OR "T'it'q'et" OR "Tla'amin" OR "Tl'azt'en" OR "Tl'etinqox" OR "Tsal'alh" OR "Tsideldel" OR "Ts'kw'aylaxw" OR "T'Sou-ke" OR "Ts'uubaa-asatx" OR "Tsuut'ina" OR "We'koqma'q" OR "Wet'suwet'en" OR "Xai'xais" OR "Xaxli'p" OR "Yunesit'in" OR ((Acadia OR Carmacks OR Champagne OR Creek OR Delaware OR Douglas OR Montana OR Similkameen) N2 (Indian OR Indians OR Tribe OR Tribes OR native OR natives OR nation OR people*)) OR Aamjiwnaang OR Abegweit OR Abitibiwinni OR Ahousaht OR Ahtahkakoop OR Aishihik OR Aitchelitz OR Aklavik OR Algonquin* OR Anicinape* OR "Animakee Wa Zhing" OR Anishina* OR aqam OR Atikamekw OR Atjanasca* OR Attawapiskat OR Aundeck-Omni-Kaning OR Batchewana OR "Bearfoot Onondaga" OR Beausoleil OR "Behdzi Ahda*" OR "Biigtigong Nishnaabeg" OR "Biinjitiwaabik Zaaging Anishinaabek" OR Bilijk OR "Binche Whut'en" OR "Bingwi Neyaashi Anishinaabek" OR "Canupawakpa Dakota" OR Cayuga OR Chawathil OR Cheam OR "Cheslatta Carrier" OR Chiniki OR Chipew* OR Cote OR Couchiching OR Cowessess OR Cowichan OR Cree OR "Dakota Plains" OR "Dakota Tipi" OR "Dechi Laot'i" OR Deline OR Dena OR Dene OR Denesuline OR "Deninu K'ue" OR Ditidaht OR "Dog Rib Rae" OR Dokis OR Eabametoong OR Ehattesaht OR Elsipogtog OR Ermineskin OR Esdilagh OR Esgenoopetitj OR Eskasoni OR Esquimalt OR Gameti OR Gayogohono OR Ginoogaming OR Gitanmaax OR Gitanyow OR Gitsegukla OR Gitwangak OR Gitxaala OR Glooscap OR Gwawaenuk OR "Gwichya Gwich'in" OR Gwitchin OR Hagwilget OR Haisla OR Halalt OR Haudenosaunee OR Heiltsuk OR Hesquiaht OR Hiawatha OR Homalco OR Hupacasath OR Huu-ay-aht OR Inninuwug OR Innu OR Innue OR Innus OR Inuvik OR Iskatewizaagegan OR Iskut OR "Ka'a'gee Tu" OR Kahkewistahaw OR "Kasabonika Lake" OR Kashechewan OR Katzie OR Kawacatoose OR Kebaowek OR Keeseekoose OR Keeseekoowenin OR Kee-Way-Win OR "Kinistin Saulteaux" OR Kinonjeoshtegon OR Kispiox OR Kitselas OR Kitsumkalum OR Kluane OR Kutchin OR Kwadacha OR Kwakiutl OR "Kwanlin Dun" OR Kwaw-kwaw-Apilt OR Kwiakah OR "Kwikwasut'inuxw Haxwa'mis" OR Kwikwetlem OR "Lax Kw'alaams" OR "Lheidli T'enneh" OR "Liidlii Kue" OR "Listuguj Mi'gmaq" OR Lyackson OR Magnetawan OR Malahat OR Maliseet OR Mamalilikulla OR Matachewan OR Matsqui OR Mattagami OR "M'Chigeeng" OR Membertou OR Metepenagiag OR Metlakatla OR Miawpukek OR Michipicoten OR Micmacs OR Millbrook OR Mishkeegogamang OR Mississauga* OR Mitaanjigamiing OR Mohawk* OR Montagnais OR Moosomin OR Moravian OR Mowachaht OR Muchalaht OR Munsee-Delaware OR Muscowpetung OR Muskoday OR Muskowekwan OR Musqueam OR "Nacho Nyak Dun" OR "Nadleh Whuten" OR Naicatchewenin OR "Nak'azdli Whut'en" OR Nakota OR Naotkamegwanning OR Naskapi OR Natoaganeg OR Nee-Tahi-Buhn OR Nehiyawak OR Nekaneet OR Neskantaga OR Neskonlith OR Nibinamik OR Nigigoonsiminikaaning OR Nipissing OR Nishnaabeg OR Nooaitch OR Nuchatlaht OR Nuxalk OR Ochapowace OR O-Chi-Chak-Ko-Sipi OR Odanak OR Ojibw* OR Okanagan OR Okanese OR Okemasis OR Oneida OR Oromocto OR Osoyoos OR Pabineau OR Pacheedaht OR "Paqtnkek Mi'kmaw" OR Pasqua OR Pauingassi OR Pauquachin OR "Pays Plat" OR Peguis OR "Pehdzeh Ki" OR Pekuakamiulnuatsh OR Penelakut OR Piapot OR Piikani OR Pikangikum OR Pinaymootang OR Potlotek OR "Qalipu Mi'kmaq" OR Qualicum OR Quatsino OR Sagamok OR Sahgaiehcan OR Samahquam OR Saugeen OR Saulteau OR Saulteaux OR Sechelt OR Secwepemc* OR Semiahmoo OR Seneca OR Shackan OR Shamattawa OR Shawanaga OR Sheguiandah OR Sheshegwaning OR Shuswap OR Siksika OR Simpcw OR Sioux OR Siska OR Skatin OR Skawahlook OR Skeetchestn OR Skidegate OR "Skin Tyee" OR Skowkale OR Skownan OR Skuppah OR Skwah OR Snuneymuxw OR Songhees OR Soowahlie OR Splatsin OR Spuzzum OR Squamish OR Squiala OR "Stswecem'c Xget'tem" OR Sumas OR "Ta'an Kwach'an" OR Tagish OR Tahltan OR Takla OR Tallcree OR "Taykwa Tagamou" OR "Teetl'it Gwich'in" OR Temagami OR Thessalon OR Timiskaming OR "Tk'emlúps te Secwépemc" OR Tla-o-qui-aht OR Tlatlasikwala OR Tlingit OR Tlowitsis OR Tootinaowaziibeeng OR Toquaht OR "Tr'ondëk Hwëch'in" OR Tsartlip OR Tsawout OR Tsawwassen OR Tseshaht OR Tseycum OR "Ts'il Kaz Koh" OR Tsleil-Waututh OR Tuscarora OR Tutchone OR Tzeachten OR Uchucklesaht OR Ucluelet OR Ulkatcho OR Wabaseemoong OR Wabauskang OR Wagmatcook OR Wahnapitae OR "Wahpeton Dakota" OR Wahsipekuk OR Wapekeka OR Wasagamack OR Wasauksing OR Waswanipi OR Wawakapewin OR Waywayseecappo OR Webequie OR Weenusk OR Wemotaci OR Wendat OR "Wha Ti" OR Whapmagoostui OR "Whitecap Dakota" OR Wikwemikong OR Witset OR Wolastoqiyik OR Wuikinuxv OR Wunnumin OR "Wuskwi Sipihk" OR Xatsull OR "Xeni Gwet'in" OR Yakweakwioose OR Yekooche OR "Yellow Quill" OR Yellowknife OR Yellowknives OR Zhiibaahaasing) OR AB(Anishnawbek OR "?Akisq'nuk" OR "?aq'am" OR "?Esdilagh" OR "Abénakis" OR "Akisq'nuk" OR "aq'am" OR "Che:k:tles7et'h'" OR "Da'naxda'xw" OR "Dzawada'enuxw" OR "Esk'etemc" OR "Gitga'at" OR "Gitlaxt'aamiks" OR "Gwa'Sala-Nakwaxda'xw" OR "Gwich'in" OR "Ka:'yu:'k't'h'" OR "Kanyen'kehaka" OR "Kapawe'no" OR "K'atlodeeche" OR "K'ómoks" OR "Leq'á:mel" OR "Lil'wat" OR "Malécite" OR "Mi'kmaq" OR "Mowachaht/Muchalaht" OR "Nisga'a" OR "N'Quatqua" OR "O'Chiese" OR "Ononda'gega'" OR "Onondowága'" OR "Onyota'a:ka" OR "Saik'uz" OR "Sambaa K'e " OR "Shxwhá:y" OR "Shxw'ow'hamel" OR "SIPEKNE'KATIK" OR "Skaru:reh" OR "Sq'éwlets" OR "Stellat'en" OR "Sts'ailes" OR "Stz'uminus" OR "T'it'q'et" OR "Tla'amin" OR "Tl'azt'en" OR "Tl'etinqox" OR "Tsal'alh" OR "Tsideldel" OR "Ts'kw'aylaxw" OR "T'Sou-ke" OR "Ts'uubaa-asatx" OR "Tsuut'ina" OR "We'koqma'q" OR "Wet'suwet'en" OR "Xai'xais" OR "Xaxli'p" OR "Yunesit'in" OR ((Acadia OR Carmacks OR Champagne OR Creek OR Delaware OR Douglas OR Montana OR Similkameen) N2 (Indian OR Indians OR Tribe OR Tribes OR native OR natives OR nation OR people*)) OR Aamjiwnaang OR Abegweit OR Abitibiwinni OR Ahousaht OR Ahtahkakoop OR Aishihik OR Aitchelitz OR Aklavik OR Algonquin* OR Anicinape* OR "Animakee Wa Zhing" OR Anishina* OR aqam OR Atikamekw OR Atjanasca* OR Attawapiskat OR Aundeck-Omni-Kaning OR Batchewana OR "Bearfoot Onondaga" OR Beausoleil OR "Behdzi Ahda*" OR "Biigtigong Nishnaabeg" OR "Biinjitiwaabik Zaaging Anishinaabek" OR Bilijk OR "Binche Whut'en" OR "Bingwi Neyaashi Anishinaabek" OR "Canupawakpa Dakota" OR Cayuga OR Chawathil OR Cheam OR "Cheslatta Carrier" OR Chiniki OR Chipew* OR Cote OR Couchiching OR Cowessess OR Cowichan OR Cree OR "Dakota Plains" OR "Dakota Tipi" OR "Dechi Laot'i" OR Deline OR Dena OR Dene OR Denesuline OR "Deninu K'ue" OR Ditidaht OR "Dog Rib Rae" OR Dokis OR Eabametoong OR Ehattesaht OR Elsipogtog OR Ermineskin OR Esdilagh OR Esgenoopetitj OR Eskasoni OR Esquimalt OR Gameti OR Gayogohono OR Ginoogaming OR Gitanmaax OR Gitanyow OR Gitsegukla OR Gitwangak OR Gitxaala OR Glooscap OR Gwawaenuk OR "Gwichya Gwich'in" OR Gwitchin OR Hagwilget OR Haisla OR Halalt OR Haudenosaunee OR Heiltsuk OR Hesquiaht OR Hiawatha OR Homalco OR Hupacasath OR Huu-ay-aht OR Inninuwug OR Innu OR Innue OR Innus OR Inuvik OR Iskatewizaagegan OR Iskut OR "Ka'a'gee Tu" OR Kahkewistahaw OR "Kasabonika Lake" OR Kashechewan OR Katzie OR Kawacatoose OR Kebaowek OR Keeseekoose OR Keeseekoowenin OR Kee-Way-Win OR "Kinistin Saulteaux" OR Kinonjeoshtegon OR Kispiox OR Kitselas OR Kitsumkalum OR Kluane OR Kutchin OR Kwadacha OR Kwakiutl OR "Kwanlin Dun" OR Kwaw-kwaw-Apilt OR Kwiakah OR "Kwikwasut'inuxw Haxwa'mis" OR Kwikwetlem OR "Lax Kw'alaams" OR "Lheidli T'enneh" OR "Liidlii Kue" OR "Listuguj Mi'gmaq" OR Lyackson OR Magnetawan OR Malahat OR Maliseet OR Mamalilikulla OR Matachewan OR Matsqui OR Mattagami OR "M'Chigeeng" OR Membertou OR Metepenagiag OR Metlakatla OR Miawpukek OR Michipicoten OR Micmacs OR Millbrook OR Mishkeegogamang OR Mississauga* OR Mitaanjigamiing OR Mohawk* OR Montagnais OR Moosomin OR Moravian OR Mowachaht OR Muchalaht OR Munsee-Delaware OR Muscowpetung OR Muskoday OR Muskowekwan OR Musqueam OR "Nacho Nyak Dun" OR "Nadleh Whuten" OR Naicatchewenin OR "Nak'azdli Whut'en" OR Nakota OR Naotkamegwanning OR Naskapi OR Natoaganeg OR Nee-Tahi-Buhn OR Nehiyawak OR Nekaneet OR Neskantaga OR Neskonlith OR Nibinamik OR Nigigoonsiminikaaning OR Nipissing OR Nishnaabeg OR Nooaitch OR Nuchatlaht OR Nuxalk OR Ochapowace OR O-Chi-Chak-Ko-Sipi OR Odanak OR Ojibw* OR Okanagan OR Okanese OR Okemasis OR Oneida OR Oromocto OR Osoyoos OR Pabineau OR Pacheedaht OR "Paqtnkek Mi'kmaw" OR Pasqua OR Pauingassi OR Pauquachin OR "Pays Plat" OR Peguis OR "Pehdzeh Ki" OR Pekuakamiulnuatsh OR Penelakut OR Piapot OR Piikani OR Pikangikum OR Pinaymootang OR Potlotek OR "Qalipu Mi'kmaq" OR Qualicum OR Quatsino OR Sagamok OR Sahgaiehcan OR Samahquam OR Saugeen OR Saulteau OR Saulteaux OR Sechelt OR Secwepemc* OR Semiahmoo OR Seneca OR Shackan OR Shamattawa OR Shawanaga OR Sheguiandah OR Sheshegwaning OR Shuswap OR Siksika OR Simpcw OR Sioux OR Siska OR Skatin OR Skawahlook OR Skeetchestn OR Skidegate OR "Skin Tyee" OR Skowkale OR Skownan OR Skuppah OR Skwah OR Snuneymuxw OR Songhees OR Soowahlie OR Splatsin OR Spuzzum OR Squamish OR Squiala OR "Stswecem'c Xget'tem" OR Sumas OR "Ta'an Kwach'an" OR Tagish OR Tahltan OR Takla OR Tallcree OR "Taykwa Tagamou" OR "Teetl'it Gwich'in" OR Temagami OR Thessalon OR Timiskaming OR "Tk'emlúps te Secwépemc" OR Tla-o-qui-aht OR Tlatlasikwala OR Tlingit OR Tlowitsis OR Tootinaowaziibeeng OR Toquaht OR "Tr'ondëk Hwëch'in" OR Tsartlip OR Tsawout OR Tsawwassen OR Tseshaht OR Tseycum OR "Ts'il Kaz Koh" OR Tsleil-Waututh OR Tuscarora OR Tutchone OR Tzeachten OR Uchucklesaht OR Ucluelet OR Ulkatcho OR Wabaseemoong OR Wabauskang OR Wagmatcook OR Wahnapitae OR "Wahpeton Dakota" OR Wahsipekuk OR Wapekeka OR Wasagamack OR Wasauksing OR Waswanipi OR Wawakapewin OR Waywayseecappo OR Webequie OR Weenusk OR Wemotaci OR Wendat OR "Wha Ti" OR Whapmagoostui OR "Whitecap Dakota" OR Wikwemikong OR Witset OR Wolastoqiyik OR Wuikinuxv OR Wunnumin OR "Wuskwi Sipihk" OR Xatsull OR "Xeni Gwet'in" OR Yakweakwioose OR Yekooche OR "Yellow Quill" OR Yellowknife OR Yellowknives OR Zhiibaahaasing) |  | 155 |
| S5 | TI("Degexit'an" OR "Yup'ik" OR ("Bear River" N2 (Indian OR Indians OR Tribe OR Tribes OR native OR natives OR nation OR people*)) OR Abenaki OR Achumawi OR Ahtna OR "Akimel Oodham" OR "Akimel O'odham" OR "Akimel Ootham" OR "Akimel O'otham" OR Aleut* OR Algonkian* OR Alsea OR Aluutiq OR Apalachee OR Arikara OR Atakapa OR Athabascan OR Athabaskan OR Athapascan OR Athapaskan OR Bannock OR Bidai OR Biloxi OR Blackfeet OR Blackfoot OR Caddoan OR Calice OR Calusa OR Carrizo OR Cayuse OR Chemakum OR Cheraw OR Cherokee OR Chetco OR Cheyenne OR Chicora OR Chilula OR Chimariko OR Clatskanie OR Coahuiltecan OR "Coeur d'Alene" OR Costanoan OR Cree OR Croatan OR Croatoan OR Cusabo OR Edisto OR Eskaleut* OR Eskimo OR Eskimo-Aleut OR Esselen OR Eyak OR "Fox and Sauk" OR Gabrielino OR "Gros Ventre" OR Guale OR "Gwich'in" OR Halchidhoma OR Hidatsa OR Hitchiti OR Hopi OR Houma OR Huichol* OR Hupa OR Ingalik OR Inupiaq OR Ioway OR Iroquoi* OR Jacome OR Jano OR Jeaga OR Juaneno OR Jumano OR Kalapuya OR Kansa OR Karankawa OR Karok OR Kashaya OR Kato OR Kawaiisu OR Kitanemuk OR Kitsai OR Klallam OR Koasati OR Kolchan OR Konkow OR Koyukon OR Lakota* OR Lassik OR Laurentian OR Lenape OR "Lenni Lenape" OR Mahican OR Mandan OR Maricopa OR Massachusett OR Mattole OR Minisink OR Missouria OR Mohave OR Mohawk OR Mohican OR Molalla OR Multnomah OR Munsee OR Muscoge* OR Naabeeho OR Nakoda OR Nanticoke OR Natchez OR Nauset OR Navaho OR Niantic OR Nipmuc OR Nisenan OR Nongatl OR Oconee OR Ofo OR Ohlone OR Ojibw* OR Okanagan OR Oneida OR Otoe OR Palouse OR Panamint OR Papago OR Patwin OR "Pee Dee" OR Pennacook OR Pequot OR Pima OR Piro OR Pocasset OR Pocomtuc OR Pokanoket OR Poospatuck OR Powhatan OR Quechan OR Quileute OR Quinault OR Quiripi OR Salinan OR Samich OR Saponi OR Serrano OR Shasta OR Shawnee OR Siksikaitsitapi OR Sinkyone OR Siouan OR Sioux OR Snohomish OR Snoqualmie OR Squaxin OR Suma OR Susquehannock OR Takelma OR Tanacross OR Tanana OR Tataviam OR Tawakoni OR Tequesta OR Tillamook OR Timucua OR Tlingit OR "Tohono Oodham" OR Tongva OR Tsimshian OR Tubatulabal OR Tutelo OR Tututni OR Unalachtigo OR Unami OR Unangan OR Unangas OR Unkechaug OR Waccamaw OR Walapai OR Wappo OR Wasco OR Whilkut OR Wichita OR Winnebago OR Wintu OR Wishram OR Yahi OR Yakima OR Yamasee OR Yana OR Yaqu OR Yuchi OR Yuki OR Yuma OR Yupik OR Yurok) OR AB("Degexit'an" OR "Yup'ik" OR ("Bear River" N2 (Indian OR Indians OR Tribe OR Tribes OR native OR natives OR nation OR people*)) OR Abenaki OR Achumawi OR Ahtna OR "Akimel Oodham" OR "Akimel O'odham" OR "Akimel Ootham" OR "Akimel O'otham" OR Aleut* OR Algonkian* OR Alsea OR Aluutiq OR Apalachee OR Arikara OR Atakapa OR Athabascan OR Athabaskan OR Athapascan OR Athapaskan OR Bannock OR Bidai OR Biloxi OR Blackfeet OR Blackfoot OR Caddoan OR Calice OR Calusa OR Carrizo OR Cayuse OR Chemakum OR Cheraw OR Cherokee OR Chetco OR Cheyenne OR Chicora OR Chilula OR Chimariko OR Clatskanie OR Coahuiltecan OR "Coeur d'Alene" OR Costanoan OR Cree OR Croatan OR Croatoan OR Cusabo OR Edisto OR Eskaleut* OR Eskimo OR Eskimo-Aleut OR Esselen OR Eyak OR "Fox and Sauk" OR Gabrielino OR "Gros Ventre" OR Guale OR "Gwich'in" OR Halchidhoma OR Hidatsa OR Hitchiti OR Hopi OR Houma OR Huichol* OR Hupa OR Ingalik OR Inupiaq OR Ioway OR Iroquoi* OR Jacome OR Jano OR Jeaga OR Juaneno OR Jumano OR Kalapuya OR Kansa OR Karankawa OR Karok OR Kashaya OR Kato OR Kawaiisu OR Kitanemuk OR Kitsai OR Klallam OR Koasati OR Kolchan OR Konkow OR Koyukon OR Lakota* OR Lassik OR Laurentian OR Lenape OR "Lenni Lenape" OR Mahican OR Mandan OR Maricopa OR Massachusett OR Mattole OR Minisink OR Missouria OR Mohave OR Mohawk OR Mohican OR Molalla OR Multnomah OR Munsee OR Muscoge* OR Naabeeho OR Nakoda OR Nanticoke OR Natchez OR Nauset OR Navaho OR Niantic OR Nipmuc OR Nisenan OR Nongatl OR Oconee OR Ofo OR Ohlone OR Ojibw* OR Okanagan OR Oneida OR Otoe OR Palouse OR Panamint OR Papago OR Patwin OR "Pee Dee" OR Pennacook OR Pequot OR Pima OR Piro OR Pocasset OR Pocomtuc OR Pokanoket OR Poospatuck OR Powhatan OR Quechan OR Quileute OR Quinault OR Quiripi OR Salinan OR Samich OR Saponi OR Serrano OR Shasta OR Shawnee OR Siksikaitsitapi OR Sinkyone OR Siouan OR Sioux OR Snohomish OR Snoqualmie OR Squaxin OR Suma OR Susquehannock OR Takelma OR Tanacross OR Tanana OR Tataviam OR Tawakoni OR Tequesta OR Tillamook OR Timucua OR Tlingit OR "Tohono Oodham" OR Tongva OR Tsimshian OR Tubatulabal OR Tutelo OR Tututni OR Unalachtigo OR Unami OR Unangan OR Unangas OR Unkechaug OR Waccamaw OR Walapai OR Wappo OR Wasco OR Whilkut OR Wichita OR Winnebago OR Wintu OR Wishram OR Yahi OR Yakima OR Yamasee OR Yana OR Yaqu OR Yuchi OR Yuki OR Yuma OR Yupik OR Yurok) |  | 1175 |
| S4 | TI("Asa'carsarmiut" OR "Dee-ni'" OR "Mi'kmaq" OR "S'Klallam" OR "Shoonaq'" OR "Sun'aq" OR (("Ivanof Bay" OR "Shoalwater Bay" OR "Spirit Lake" OR "Squaxin Island" OR Dakota OR Dakotah OR Wichita OR keechi OR waco OR Tawakonie OR Cheyenne OR Creek OR Crow OR Hoonah OR Kasigluk OR Maricopa OR Miami OR Omaha OR Oneida OR Ottawa OR Peoria OR Salamatof OR Sitka OR Snoqualmie OR Spokane OR Winnebago OR Wyandotte) N2 (Indian OR Indians OR Tribe OR Tribes OR native OR natives OR nation OR people*)) OR ((Native OR indian OR traditional) N2 (village OR council OR community OR association)) OR Absentee-Shawnee OR Achomawi OR Agdaagux OR "Agua Caliente" OR Ajumawi OR Ak-Chin OR Akiachak OR Akiak OR Alabama-Coushatta OR Alabama-Quassarte OR Alatna OR Algaaciq OR Algin OR Allakaket OR "Alturas Indian Rancheria" OR Alutiiq OR Angoon OR Anvik OR Apache OR Aporige OR Aquinnah OR arapaho OR Aroostook OR Assiniboine OR Astarawi OR Atsugewi OR Atwamsini OR Bdewakantunwan OR Caddo OR Cahto OR Cahuilla OR Catawba OR Cayuga OR Chalkyitsik OR Cheesh-Na OR Chehalis OR Chemehuevi OR Cher-Ae OR Chevak OR Cheyenne-Arapaho OR Chickahominy OR Chickaloon OR Chickasaw OR Chignik OR Chilkat OR Chilkoot OR Chinik OR Chinook OR Chippewa* OR Chitimacha OR Choctaw OR Chukchansi OR Chuloonawick OR Chumash OR Cocopah OR Colusa OR Comanche OR Coos OR Cosmit OR Coushatta OR Cowlitz OR "Cross Village" OR Cupeno OR Curyung OR Diegueno OR Dine' OR Egegik OR Eklutna OR Elwha OR "Emmonak Village" OR "Evansville Village" OR Ewiiaapaayp OR "Galena Village" OR Goshute OR "Gulkana Village Council" OR Haida OR Hammawi OR Havasupai OR "Healy Lake Village" OR Hewisedawi OR Ho-Chunk OR Hoh OR Hokan OR Holikachuk OR "Holy Cross Tribe" OR Hualapai OR "Hughes Village" OR "Huslia Village" OR "Hydaburg Cooperative Association" OR "Igiugig Village" OR Iipay OR Ilmawi OR Inaja OR Inupiat OR Ione OR Itsatawi OR "Kaguyak Village" OR Kaibab OR "Kaktovik Village" OR Kalispel OR Kanosh OR Karuk OR Kashia OR Kaw OR Keetoowah OR "Kenaitze Indian Tribe" OR "Ketchikan Indian Community" OR "Keweenaw Bay" OR Kialegee OR Kickapoo OR "King Island Native Community" OR "King Salmon Tribe" OR Kiowa OR Klamath OR "Klawock Cooperative Association" OR Kletsel OR "Knik Tribe" OR "Koi Nation" OR "Kokhanok Village" OR Koosharem OR Kootenai OR Kosalektawi OR "Koyukuk Native Village" OR Kumeyaay OR Kwalhioqua OR "Lesnoi Village" OR "Levelock Village" OR "Lime Village" OR "Louden Village" OR Luiseno OR Lummi OR Madesi OR Maidu OR Makah OR Maliseet OR "Manley Hot Springs Village" OR "Manokotak Village" OR Manzanita OR Mashantucket OR Match-e-be-nash-she-wish OR Mattaponi OR "McGrath Native Village" OR Mdewakanton OR Mechoopda OR Menominee OR Me-Wuk OR Miccosukee OR Micmacs OR "Mission Indians" OR Miwok OR Miwuk OR Modoc OR Mohegan OR Mojave OR Monacan OR "Mono Indians" OR Muckleshoot OR Muscogee OR "Na Dené" OR Nansemond OR Narragansett OR Navajo OR "New Koliganek Village" OR "New Stuyahok Village" OR "Newhalen Village" OR "Newtok Village" OR "Nez Perce" OR "Nikolai Village" OR "Ninilchik Village" OR Nisqually OR Nomlaki OR "Nondalton Village" OR Nooksack OR "Northway Village" OR Nottawaseppi OR "Nulato Village" OR Nunakauyarmiut OR Odawa OR Ohgsenakale OR "Ohkay Owingeh" OR Onondaga OR Oreilles OR "Organized Village of Grayling" OR "Organized Village of Kake" OR "Organized Village of Kasaan" OR "Organized Village of Kwethluk" OR "Organized Village of Saxman" OR Orutsararmiut OR Osage OR Otoe-Missouria OR Paiute OR Paiute-Shoshone OR Pala OR Pamunkey OR Pascua OR Paskenta OR Passamaquoddy OR "Pauloff Harbor Village" OR Pauma OR Pawnee OR Pechanga OR "Pechanga " OR "Peck Indian Reservation" OR "Pedro Bay Village" OR Pee-Posh OR Penobscot OR Penutian OR Perquot OR "Pima People" OR Pima-Maricopa OR Poarch OR Pokagon OR Pomo OR Ponca OR "Portage Creek Village" OR Potawatomi OR Pottawatomi OR Pueblo OR Puyallup OR "Qagan Tayagungin" OR Qawalangin OR Quapaw OR Quechan OR "Rampart Village" OR Rappahannock OR Resighini OR "Sac & Fox" OR "Sac and Fox" OR "Saint George Island" OR "Saint Paul Island" OR Salish OR Samish OR Santee OR Sauk-Suiattle OR "Seldovia Village Tribe" OR Seminole OR Seneca OR Seneca-Cayuga OR Shinnecock OR Shivwits OR Shoshone OR Shoshone-Bannock OR Shoshone-Paiute OR Siuslaw OR Skagit OR "Skagway Village" OR Skokomish OR "South Naknek Village" OR "Stebbins Community Association" OR Stillaguamish OR Suquamish OR Swinomish OR "Takotna Village" OR Tangirnaq OR Tejon OR "Telida Village" OR Te-Moak OR Thlopthlocco OR "Tlingit & Haida" OR "Tohono O'odham" OR Tolowa OR Tonawanda OR Tonkawa OR Tulalip OR Tunica-Biloxi OR Tuolumne OR Tuscarora OR "Twin Hills Village" OR "Ugashik Village" OR Umatilla OR Umkumiut OR Umpqua OR Ute OR "Utu Utu Gwaitu" OR "Village of Alakanuk" OR "Village of Anaktuvuk Pass" OR "Village of Aniak" OR "Village of Atmautluak" OR "Village of Bill Moore's Slough" OR "Village of Chefornak" OR "Village of Clarks Point" OR "Village of Crooked Creek" OR "Village of Dot Lake" OR "Village of Iliamna" OR "Village of Kalskag" OR "Village of Kaltag" OR "Village of Kotlik" OR "Village of Lower Kalskag" OR "Village of Ohogamiut" OR "Village of Red Devil" OR "Village of Sleetmute" OR "Village of Solomon" OR "Village of Stony River" OR "Village of Venetie" OR "Village of Wainwright" OR Wailaki OR Wampanoag OR "Warm Springs Reservation" OR Washoe OR Willapa OR Winnemucca OR Wintun OR Wiyot OR "Wrangell Cooperative Association" OR Yakama OR Yakutat OR Yaqui OR Yavapai OR Yokuts OR Yuhaaviatam OR Yuima OR Yukian OR Yukut OR Yupiit OR Zuni) OR AB("Asa'carsarmiut" OR "Dee-ni'" OR "Mi'kmaq" OR "S'Klallam" OR "Shoonaq'" OR "Sun'aq" OR (("Ivanof Bay" OR "Shoalwater Bay" OR "Spirit Lake" OR "Squaxin Island" OR Dakota OR Dakotah OR Wichita OR keechi OR waco OR Tawakonie OR Cheyenne OR Creek OR Crow OR Hoonah OR Kasigluk OR Maricopa OR Miami OR Omaha OR Oneida OR Ottawa OR Peoria OR Salamatof OR Sitka OR Snoqualmie OR Spokane OR Winnebago OR Wyandotte) N2 (Indian OR Indians OR Tribe OR Tribes OR native OR natives OR nation OR people*)) OR ((Native OR indian OR traditional) N2 (village OR council OR community OR association)) OR Absentee-Shawnee OR Achomawi OR Agdaagux OR "Agua Caliente" OR Ajumawi OR Ak-Chin OR Akiachak OR Akiak OR Alabama-Coushatta OR Alabama-Quassarte OR Alatna OR Algaaciq OR Algin OR Allakaket OR "Alturas Indian Rancheria" OR Alutiiq OR Angoon OR Anvik OR Apache OR Aporige OR Aquinnah OR arapaho OR Aroostook OR Assiniboine OR Astarawi OR Atsugewi OR Atwamsini OR Bdewakantunwan OR Caddo OR Cahto OR Cahuilla OR Catawba OR Cayuga OR Chalkyitsik OR Cheesh-Na OR Chehalis OR Chemehuevi OR Cher-Ae OR Chevak OR Cheyenne-Arapaho OR Chickahominy OR Chickaloon OR Chickasaw OR Chignik OR Chilkat OR Chilkoot OR Chinik OR Chinook OR Chippewa* OR Chitimacha OR Choctaw OR Chukchansi OR Chuloonawick OR Chumash OR Cocopah OR Colusa OR Comanche OR Coos OR Cosmit OR Coushatta OR Cowlitz OR "Cross Village" OR Cupeno OR Curyung OR Diegueno OR Dine' OR Egegik OR Eklutna OR Elwha OR "Emmonak Village" OR "Evansville Village" OR Ewiiaapaayp OR "Galena Village" OR Goshute OR "Gulkana Village Council" OR Haida OR Hammawi OR Havasupai OR "Healy Lake Village" OR Hewisedawi OR Ho-Chunk OR Hoh OR Hokan OR Holikachuk OR "Holy Cross Tribe" OR Hualapai OR "Hughes Village" OR "Huslia Village" OR "Hydaburg Cooperative Association" OR "Igiugig Village" OR Iipay OR Ilmawi OR Inaja OR Inupiat OR Ione OR Itsatawi OR "Kaguyak Village" OR Kaibab OR "Kaktovik Village" OR Kalispel OR Kanosh OR Karuk OR Kashia OR Kaw OR Keetoowah OR "Kenaitze Indian Tribe" OR "Ketchikan Indian Community" OR "Keweenaw Bay" OR Kialegee OR Kickapoo OR "King Island Native Community" OR "King Salmon Tribe" OR Kiowa OR Klamath OR "Klawock Cooperative Association" OR Kletsel OR "Knik Tribe" OR "Koi Nation" OR "Kokhanok Village" OR Koosharem OR Kootenai OR Kosalektawi OR "Koyukuk Native Village" OR Kumeyaay OR Kwalhioqua OR "Lesnoi Village" OR "Levelock Village" OR "Lime Village" OR "Louden Village" OR Luiseno OR Lummi OR Madesi OR Maidu OR Makah OR Maliseet OR "Manley Hot Springs Village" OR "Manokotak Village" OR Manzanita OR Mashantucket OR Match-e-be-nash-she-wish OR Mattaponi OR "McGrath Native Village" OR Mdewakanton OR Mechoopda OR Menominee OR Me-Wuk OR Miccosukee OR Micmacs OR "Mission Indians" OR Miwok OR Miwuk OR Modoc OR Mohegan OR Mojave OR Monacan OR "Mono Indians" OR Muckleshoot OR Muscogee OR "Na Dené" OR Nansemond OR Narragansett OR Navajo OR "New Koliganek Village" OR "New Stuyahok Village" OR "Newhalen Village" OR "Newtok Village" OR "Nez Perce" OR "Nikolai Village" OR "Ninilchik Village" OR Nisqually OR Nomlaki OR "Nondalton Village" OR Nooksack OR "Northway Village" OR Nottawaseppi OR "Nulato Village" OR Nunakauyarmiut OR Odawa OR Ohgsenakale OR "Ohkay Owingeh" OR Onondaga OR Oreilles OR "Organized Village of Grayling" OR "Organized Village of Kake" OR "Organized Village of Kasaan" OR "Organized Village of Kwethluk" OR "Organized Village of Saxman" OR Orutsararmiut OR Osage OR Otoe-Missouria OR Paiute OR Paiute-Shoshone OR Pala OR Pamunkey OR Pascua OR Paskenta OR Passamaquoddy OR "Pauloff Harbor Village" OR Pauma OR Pawnee OR Pechanga OR "Pechanga " OR "Peck Indian Reservation" OR "Pedro Bay Village" OR Pee-Posh OR Penobscot OR Penutian OR Perquot OR "Pima People" OR Pima-Maricopa OR Poarch OR Pokagon OR Pomo OR Ponca OR "Portage Creek Village" OR Potawatomi OR Pottawatomi OR Pueblo OR Puyallup OR "Qagan Tayagungin" OR Qawalangin OR Quapaw OR Quechan OR "Rampart Village" OR Rappahannock OR Resighini OR "Sac & Fox" OR "Sac and Fox" OR "Saint George Island" OR "Saint Paul Island" OR Salish OR Samish OR Santee OR Sauk-Suiattle OR "Seldovia Village Tribe" OR Seminole OR Seneca OR Seneca-Cayuga OR Shinnecock OR Shivwits OR Shoshone OR Shoshone-Bannock OR Shoshone-Paiute OR Siuslaw OR Skagit OR "Skagway Village" OR Skokomish OR "South Naknek Village" OR "Stebbins Community Association" OR Stillaguamish OR Suquamish OR Swinomish OR "Takotna Village" OR Tangirnaq OR Tejon OR "Telida Village" OR Te-Moak OR Thlopthlocco OR "Tlingit & Haida" OR "Tohono O'odham" OR Tolowa OR Tonawanda OR Tonkawa OR Tulalip OR Tunica-Biloxi OR Tuolumne OR Tuscarora OR "Twin Hills Village" OR "Ugashik Village" OR Umatilla OR Umkumiut OR Umpqua OR Ute OR "Utu Utu Gwaitu" OR "Village of Alakanuk" OR "Village of Anaktuvuk Pass" OR "Village of Aniak" OR "Village of Atmautluak" OR "Village of Bill Moore's Slough" OR "Village of Chefornak" OR "Village of Clarks Point" OR "Village of Crooked Creek" OR "Village of Dot Lake" OR "Village of Iliamna" OR "Village of Kalskag" OR "Village of Kaltag" OR "Village of Kotlik" OR "Village of Lower Kalskag" OR "Village of Ohogamiut" OR "Village of Red Devil" OR "Village of Sleetmute" OR "Village of Solomon" OR "Village of Stony River" OR "Village of Venetie" OR "Village of Wainwright" OR Wailaki OR Wampanoag OR "Warm Springs Reservation" OR Washoe OR Willapa OR Winnemucca OR Wintun OR Wiyot OR "Wrangell Cooperative Association" OR Yakama OR Yakutat OR Yaqui OR Yavapai OR Yokuts OR Yuhaaviatam OR Yuima OR Yukian OR Yukut OR Yupiit OR Zuni) |  | 288 |
| S3 | DE("PIMA (North American people)" OR "PLAINS peoples (North American peoples)" OR "APACHE (North American people)" OR "ARIKARA (North American people)" OR "COMANCHE (North American people)" OR "CROW (North American people)" OR "DAKOTA (North American people)" OR "DHEGIHA (North American people)" OR "GROS Ventre (Montanan people)" OR "HIDATSA (North American people)" OR "HUNKPAPA (North American people)" OR "IOWA (North American people)" OR "KEECHE (North American people)" OR "KIOWA (North American people)" OR "KIOWA Apache (North American people)" OR "LAKOTA (North American people)" OR "MANDAN (North American people)" OR "MILL Creek Indians (Plains people)" OR "OGLALA Lakota (North American people)" OR "OMAHA (North American people)" OR "ONEONTA (Plains people)" OR "OTO (North American people)" OR "PONCA (North American people)" OR "SANTEE (North American people)" OR "SARSI (North American people)" OR "SIOUANS (North American peoples)" OR "SISSETON Indians (North American people)" OR "TONKAWA (North American people)" OR "YANKTON Indians (North American people)" OR "ANISHINAABE (North American people)" OR "ATHAPASCANS (North American people)" OR "CADDOANS (North American peoples)" OR "CALIFORNIA Indians (North American people)" OR "CIBECUE Apache (North American people)" OR "GREAT Basin peoples (North American peoples)" OR "HALCHIDHOMA (North American people)" OR "HAVASUPAI (North American people)" OR "HIA-Ced O'odham (North American people)" OR "HUALAPAI (North American people)" OR "IROQUOIANS (North American peoples)" OR "MARICOPA (North American people)" OR "MASSAWOMECK (North American people)" OR "MIMBRENO (North American people)" OR "MONONGAHELA (North American people)" OR "MOUND-builders" OR "OFF-reservation Indians (Native Americans)" OR "PACIFIC Northwest peoples (North American peoples)" OR "PIMAN peoples (North American peoples)" OR "RESERVATION Indians (Native Americans)" OR "SOBAIPURI (North American people)" OR "SOUTHERN Paiute (North American people)" OR "SOUTHWEST Indians (North American peoples)" OR "SUBARCTIC peoples (North Americans)" OR "TEWA (North American people)" OR "TOHONO O'odham (North American people)" OR "UTO-Aztecan peoples (North American peoples)" OR "WESTERN Apache (North American people)" OR "WHITE Mountain Apache (North American people)" OR "WOODLAND peoples (North American peoples)" OR "YAQUI (North American people)" OR "YAVAPAI (North American people)" OR "YUMA (North American people)" OR "CREE (North American people)" OR ESKIMOS OR "AHTENA (North American people)" OR "ALASKAN Eskimos" OR "ALEUTS" OR "CHILKAT (North American people)" OR "CHIMMESYAN (North American people)" OR "DEG Hit'an (North American people)" OR "DENA'INA (North American people)" OR "EYAK (North American people)" OR "GWICH'IN (North American people)" OR "HAIDA (North American people)" OR "HAN (North American people)" OR "KOYUKON (North American people)" OR "NEETS'AII Gwich'in (North American people)" OR "OLDER Alaska Natives" OR "TAHLTAN (North American people)" OR "TANANA (North American people)" OR "TLINGIT (North American people)" OR "TSETSAUT (North American people)" OR "TSIMSHIAN (North American people)" OR "VUNTUT Gwich'in (North American people)") |  | 16 |
| S2 | DE("FIRST Nations of Canada" OR "ABORIGINAL Canadians" OR "CANADIAN Inuit" OR "METIS") OR TI(((Canada OR Canadian*) N2 (native* OR indigenous OR aborigin*)) OR Metis OR "Métis" OR Michif* OR Mitchif OR Mechif OR "Métif" OR "Métchif" OR Metif OR Metchif OR Inuit OR "First Nations" OR "First Nation") OR AB(((Canada OR Canadian*) N2 (native* OR indigenous OR aborigin*)) OR Metis OR "Métis" OR Michif* OR Mitchif OR Mechif OR "Métif" OR "Métchif" OR Metif OR Metchif OR Inuit OR "First Nations" OR "First Nation") |  | 61 |
| S1 | DE("NATIVE Americans" OR "ALASKA Natives") OR TI(((American* OR Alaska*) N2 (Native* OR Indian OR Indians OR indigenous OR aborigin*)) OR Amerind*) OR AB(((American* OR Alaska*) N2 (Native* OR Indian OR Indians OR indigenous OR aborigin*)) OR Amerind*) |  | 303 |

### AgeLine (EBSCOHost)

Results for both the initial search and search update are provided as the database coverage changed significantly between the two. In 2023 Ageline (EBSCO) indexed 207 journals^1^, but indexed only 88 in 2024^2^.

1.	Abstracting & Indexing Database: Ageline. EBSCO. 21 March 2023. <https://www.ebsco.com/products/research-databases/ageline>. Accessed 16 April 2025 via the Internet Archive: <https://web.archive.org/web/20230321125059/https://www.ebsco.com/products/research-databases/ageline>

2.	Abstracting & Indexing Database: Ageline. EBSCO. 12 December 2024. <https://www.ebsco.com/products/research-databases/ageline>. Accessed 16 April 2025 via the Internet Archive: <https://web.archive.org/web/20241212070814/https://www.ebsco.com/products/research-databases/ageline>

**Field Codes:** TI = title, AB= abstract, DE = subjects [exact].

**Proximity operator:** N# 
**Truncation:**  *

**Database** - AgeLine

**Interface** - EBSCOhost Research Databases

**Search Screen** - Advanced Search

**Search modes** - Proximity

**Initial Search Date** - Wednesday, June 07, 2023 11:41:47 PM

**Updated**: Thu, November 21, 2024 4:08:46 PM

| **#** | **Query** | **Limiters** | **6/7/23 Results** | **11/21/24 Results** |
| --- | --- | --- | --- | --- |
| S24 | S17 OR S28 OR S19 OR S20 OR S21 OR S22 | Publication Date: 20230501-20241231 | N/A | 0 |
| S23 | S17 OR S28 OR S19 OR S20 OR S21 OR S22 |  | 232 | 85 |
| S22 | S14 AND (S9 OR S12) AND S15 |  | 90 | 21 |
| S21 | S16 AND (S7 OR S8) |  | 37 | 26 |
| S20 | S12 AND (S7 OR S8) |  | 94 | 28 |
| S19 | S9 AND S13 |  | 2 | 3 |
| S18 | S8 AND S9 |  | 13 | 8 |
| S17 | S7 AND S9 |  | 10 | 7 |
| S16 | TI((women* OR woman* OR female*) N2 (health OR healthcare OR service* OR clinic OR clinics)) OR AB((women* OR woman* OR female*) N2 (health OR healthcare OR service* OR clinic OR clinics)) |  | 1613 | 2101 |
| S15 | DE("North America" OR Canada OR Canadians OR Alberta OR "British Columbia" OR Manitoba OR "New Brunswick" OR "Newfoundland" OR "Labrador" OR "Northwest Territories" OR "Nova Scotia" OR Nunavut OR Ontario OR "Prince Edward Island" OR Quebec OR Saskatchewan OR Yukon OR "United States" OR Appalachia* OR "New England" OR "Pacific States" OR Alabama OR Alaska OR Arizona OR Arkansas OR California OR Colorado OR Connecticut OR Delaware OR Florida OR Georgia OR Hawaii OR Idaho OR Illinois OR Indiana OR Iowa OR Kansas OR Kentucky OR Louisiana OR Maine OR Maryland OR Massachusetts OR Michigan OR Minnesota OR Mississippi OR Missouri OR Montana OR Nebraska OR Nevada OR "New Hampshire" OR "New Jersey" OR "New Mexico" OR "New York" OR "North Carolina" OR "North Dakota" OR Ohio OR Oklahoma OR Oregon OR Pennsylvania OR "Rhode Island" OR "South Carolina" OR "South Dakota" OR Tennessee OR Texas OR Utah OR Vermont OR Virginia OR Washington OR "West Virginia" OR Wisconsin OR Wyoming OR "District of Columbia" OR "Washington DC") OR TI("North America" OR Canada OR Alberta OR "British Columbia" OR Manitoba OR "New Brunswick" OR "Newfoundland" OR "Labrador" OR "Northwest Territories" OR "Nova Scotia" OR Nunavut OR Ontario OR "Prince Edward Island" OR Quebec OR Saskatchewan OR Yukon OR "United States" OR Appalachia* OR "New England" OR "Pacific States" OR Alabama OR Alaska OR Arizona OR Arkansas OR California OR Colorado OR Connecticut OR Delaware OR Florida OR Georgia OR Hawaii OR Idaho OR Illinois OR Indiana OR Iowa OR Kansas OR Kentucky OR Louisiana OR Maine OR Maryland OR Massachusetts OR Michigan OR Minnesota OR Mississippi OR Missouri OR Montana OR Nebraska OR Nevada OR "New Hampshire" OR "New Jersey" OR "New Mexico" OR "New York" OR "North Carolina" OR "North Dakota" OR Ohio OR Oklahoma OR Oregon OR Pennsylvania OR "Rhode Island" OR "South Carolina" OR "South Dakota" OR Tennessee OR Texas OR Utah OR Vermont OR Virginia OR Washington OR "West Virginia" OR Wisconsin OR Wyoming OR "District of Columbia" OR "Washington DC") OR AB("North America" OR Canada OR Alberta OR "British Columbia" OR Manitoba OR "New Brunswick" OR "Newfoundland" OR "Labrador" OR "Northwest Territories" OR "Nova Scotia" OR Nunavut OR Ontario OR "Prince Edward Island" OR Quebec OR Saskatchewan OR Yukon OR "United States" OR Appalachia* OR "New England" OR "Pacific States" OR Alabama OR Alaska OR Arizona OR Arkansas OR California OR Colorado OR Connecticut OR Delaware OR Florida OR Georgia OR Hawaii OR Idaho OR Illinois OR Indiana OR Iowa OR Kansas OR Kentucky OR Louisiana OR Maine OR Maryland OR Massachusetts OR Michigan OR Minnesota OR Mississippi OR Missouri OR Montana OR Nebraska OR Nevada OR "New Hampshire" OR "New Jersey" OR "New Mexico" OR "New York" OR "North Carolina" OR "North Dakota" OR Ohio OR Oklahoma OR Oregon OR Pennsylvania OR "Rhode Island" OR "South Carolina" OR "South Dakota" OR Tennessee OR Texas OR Utah OR Vermont OR Virginia OR Washington OR "West Virginia" OR Wisconsin OR Wyoming OR "District of Columbia" OR "Washington DC") |  | 107783 | 77653 |
| S14 | DE("Alternative Medicine" OR "Dietary Supplements") OR TI(((Complementary OR alternative OR traditional) N2 (medicine* OR therap* OR healing OR treatment*)) OR "Herbal Medicine" OR herbalism OR ((botanical OR ethnobotanical OR herb* OR plant OR plant-based) N2 (medicine* OR remedy OR remedies OR healing OR therap* OR treatment*)) OR phyto-medicine OR phytomedic* OR "folk medicine" OR "folk healer") OR AB(((Complementary OR alternative OR traditional) N2 (medicine* OR therap* OR healing OR treatment*)) OR "Herbal Medicine" OR herbalism OR ((botanical OR ethnobotanical OR herb* OR plant OR plant-based) N2 (medicine* OR remedy OR remedies OR healing OR therap* OR treatment*)) OR phyto-medicine OR phytomedic* OR "folk medicine" OR "folk healer") |  | 1608 | 1652 |
| S13 | DE("National Indian Council on Aging" OR "Indian Health Service") OR TI(((Indigenous OR native OR "first nation" OR aborigin* OR Indian) N2 (health OR healthcare OR healing OR medicine OR practices)) OR ethnomedicin* OR "native healing" OR Limpia OR "indigenous healer" OR medicineman OR medicinemen OR "medicine man" OR "medicine men" OR Curandero OR "folk healer" OR shaman* OR shamen) OR AB(((Indigenous OR native OR "first nation" OR aborigin* OR Indian) N2 (health OR healthcare OR healing OR medicine OR practices)) OR ethnomedicin* OR "native healing" OR Limpia OR "indigenous healer" OR medicineman OR medicinemen OR "medicine man" OR "medicine men" OR Curandero OR "folk healer" OR shaman* OR shamen) |  | 170 | 225 |
| S12 | S10 AND S11 |  | 1482 | 1067 |
| S11 | DE(Females) OR TI(female OR woman OR women) OR AB(female OR woman OR women) |  | 34025 | 37011 |
| S10 | TI(elder OR elders) OR AB(elder OR elders) |  | 8722 | 8524 |
| S9 | DE(Menopause OR Perimenopause) OR TI(Climacteric OR Climacterium OR Menopause OR Menopaus* OR "Menopause, Premature" OR "climacterium praecox" OR Perimenopause OR Perimenopaus* OR Postmenopause OR Postmenopaus* OR "Post menopaus*" OR Post-menopaus* OR Premenopause OR Premenopause* OR Pre-menopaus* OR "Pre menopaus*" OR Climacteric* OR "Life Change*" OR "Change of life") OR AB(Climacteric OR Climacterium OR Menopause OR Menopaus* OR "Menopause, Premature" OR "climacterium praecox" OR Perimenopause OR Perimenopaus* OR Postmenopause OR Postmenopaus* OR "Post menopaus*" OR Post-menopaus* OR Premenopause OR Premenopause* OR Pre-menopaus* OR "Pre menopaus*" OR Climacteric* OR "Life Change*" OR "Change of life") |  | 1753 | 1839 |
| S8 | S3 OR S4 OR S5 OR S6 |  | 2704 | 2334 |
| S7 | S1 OR S2 |  | 973 | 800 |
| S6 | TI("Kwakwaka'wakw" OR "Kwak'wala" OR "L'uml'umuluts" OR "Lheidli T'enneh " OR "Nlaka'pamux" OR "Nuxálk" OR "Quw'utsun" OR "Qwum'yiqun" OR "S'amuna" OR "Sm'algyax" OR "St'at'imc" OR "Stl'atl'imx" OR "Sto:lo" OR "Tl'ulpalus" OR "Tsilhqot'in" OR "T'Souke" OR "Xa'xtsa" OR (Labrador N2 (Indian OR Indians OR Tribe OR Tribes OR native OR natives OR nation OR people*)) OR "Áa Tlein Kwáan" OR Aivilingmiut OR Algonkian OR Algonquian OR Algonquin OR Anishinaabe OR "Atlin people" OR "Baffin Island" OR "Bella Coola" OR Burrard OR Cheklesahht OR Chilcotin OR Chippew* OR Clayoquot OR Clemclemaluts OR Comiaken OR Comox OR Coquitlam OR Dakelh OR Dane-zaa OR "Deisleen Kwáan" OR Dunne-za OR Gitxsan OR Gitxsanimaax OR Guyohkohnyo OR "Haad kil" OR Halkomelem OR Halqemeylem OR Henaksiala OR Hinupsum OR Hunquminum OR Hwlitsum OR Hwulqwselu OR Hydin OR Iglulik OR Iglulingmuit OR Ininew OR Inuinnait OR Inuk OR Inupiat OR Inuvialuit OR Iroquois OR Kainai OR Kaska OR Kelsemeht OR Khenipsen OR Kilpahlas OR Kimsquit OR Kitamaat OR Kitlinermiut OR Kivallirmiut OR Klahoose OR Koksilah OR Kootenay OR Koskimo OR Ktunaxa OR Kwantlen OR Kwatna OR Kyuquot OR Labradormiut OR Laich-kwil-tach OR Lamalch* OR Lekwungen OR Lillooet OR Loucheaux OR Lummi OR Micmac OR Mississaugas OR Mohawk OR Mowachaht-Muchalaht OR Nadene OR Na-Dene OR Namgis OR Nanaimo OR Nanoose OR Nequatque OR Netsilik OR Netsilingmiut OR Niitsitapi OR Nimpkish OR Nootka OR Nunavimiut OR Nunavimmiut OR Nuu-chah-nulth OR Odawa OR Ohiaht OR Oji-cree OR Onayotekaono OR Onondaga OR Onondowahgah OR Onundagaono OR Opetchesaht OR Owekeeno OR Passamaquoddy OR Peigan OR Petun OR Plains-Cree OR Popkum OR Potawatomi OR Quamichan OR Sahtu OR Salish OR Sarcee OR Scowlitz OR "Seabird Island" OR Secwepemc OR Sekani OR Semahquam OR Shishalh OR Sinixt OR Ska-Ruh-Reh OR Skway OR Sliammon OR Snaw-naw-as OR Somena OR Songish OR Sooke OR Stoney OR Stuie OR Tallheo OR Tasttine OR Teslin OR Tlicho OR Tsimshianic OR Tununirmiut OR Ungava OR Wakashan OR Weewaikai OR Wewaykum OR Wyandot OR Yuculta) OR AB("Kwakwaka'wakw" OR "Kwak'wala" OR "L'uml'umuluts" OR "Lheidli T'enneh " OR "Nlaka'pamux" OR "Nuxálk" OR "Quw'utsun" OR "Qwum'yiqun" OR "S'amuna" OR "Sm'algyax" OR "St'at'imc" OR "Stl'atl'imx" OR "Sto:lo" OR "Tl'ulpalus" OR "Tsilhqot'in" OR "T'Souke" OR "Xa'xtsa" OR (Labrador N2 (Indian OR Indians OR Tribe OR Tribes OR native OR natives OR nation OR people*)) OR "Áa Tlein Kwáan" OR Aivilingmiut OR Algonkian OR Algonquian OR Algonquin OR Anishinaabe OR "Atlin people" OR "Baffin Island" OR "Bella Coola" OR Burrard OR Cheklesahht OR Chilcotin OR Chippew* OR Clayoquot OR Clemclemaluts OR Comiaken OR Comox OR Coquitlam OR Dakelh OR Dane-zaa OR "Deisleen Kwáan" OR Dunne-za OR Gitxsan OR Gitxsanimaax OR Guyohkohnyo OR "Haad kil" OR Halkomelem OR Halqemeylem OR Henaksiala OR Hinupsum OR Hunquminum OR Hwlitsum OR Hwulqwselu OR Hydin OR Iglulik OR Iglulingmuit OR Ininew OR Inuinnait OR Inuk OR Inupiat OR Inuvialuit OR Iroquois OR Kainai OR Kaska OR Kelsemeht OR Khenipsen OR Kilpahlas OR Kimsquit OR Kitamaat OR Kitlinermiut OR Kivallirmiut OR Klahoose OR Koksilah OR Kootenay OR Koskimo OR Ktunaxa OR Kwantlen OR Kwatna OR Kyuquot OR Labradormiut OR Laich-kwil-tach OR Lamalch* OR Lekwungen OR Lillooet OR Loucheaux OR Lummi OR Micmac OR Mississaugas OR Mohawk OR Mowachaht-Muchalaht OR Nadene OR Na-Dene OR Namgis OR Nanaimo OR Nanoose OR Nequatque OR Netsilik OR Netsilingmiut OR Niitsitapi OR Nimpkish OR Nootka OR Nunavimiut OR Nunavimmiut OR Nuu-chah-nulth OR Odawa OR Ohiaht OR Oji-cree OR Onayotekaono OR Onondaga OR Onondowahgah OR Onundagaono OR Opetchesaht OR Owekeeno OR Passamaquoddy OR Peigan OR Petun OR Plains-Cree OR Popkum OR Potawatomi OR Quamichan OR Sahtu OR Salish OR Sarcee OR Scowlitz OR "Seabird Island" OR Secwepemc OR Sekani OR Semahquam OR Shishalh OR Sinixt OR Ska-Ruh-Reh OR Skway OR Sliammon OR Snaw-naw-as OR Somena OR Songish OR Sooke OR Stoney OR Stuie OR Tallheo OR Tasttine OR Teslin OR Tlicho OR Tsimshianic OR Tununirmiut OR Ungava OR Wakashan OR Weewaikai OR Wewaykum OR Wyandot OR Yuculta) |  | 41 | 29 |
| S5 | TI(Anishnawbek OR "?Akisq'nuk" OR "?aq'am" OR "?Esdilagh" OR "Abénakis" OR "Akisq'nuk" OR "aq'am" OR "Che:k:tles7et'h'" OR "Da'naxda'xw" OR "Dzawada'enuxw" OR "Esk'etemc" OR "Gitga'at" OR "Gitlaxt'aamiks" OR "Gwa'Sala-Nakwaxda'xw" OR "Gwich'in" OR "Ka:'yu:'k't'h'" OR "Kanyen'kehaka" OR "Kapawe'no" OR "K'atlodeeche" OR "K'ómoks" OR "Leq'á:mel" OR "Lil'wat" OR "Malécite" OR "Mi'kmaq" OR "Mowachaht/Muchalaht" OR "Nisga'a" OR "N'Quatqua" OR "O'Chiese" OR "Ononda'gega'" OR "Onondowága'" OR "Onyota'a:ka" OR "Saik'uz" OR "Sambaa K'e " OR "Shxwhá:y" OR "Shxw'ow'hamel" OR "SIPEKNE'KATIK" OR "Skaru:reh" OR "Sq'éwlets" OR "Stellat'en" OR "Sts'ailes" OR "Stz'uminus" OR "T'it'q'et" OR "Tla'amin" OR "Tl'azt'en" OR "Tl'etinqox" OR "Tsal'alh" OR "Tsideldel" OR "Ts'kw'aylaxw" OR "T'Sou-ke" OR "Ts'uubaa-asatx" OR "Tsuut'ina" OR "We'koqma'q" OR "Wet'suwet'en" OR "Xai'xais" OR "Xaxli'p" OR "Yunesit'in" OR ((Acadia OR Carmacks OR Champagne OR Creek OR Delaware OR Douglas OR Montana OR Similkameen) N2 (Indian OR Indians OR Tribe OR Tribes OR native OR natives OR nation OR people*)) OR Aamjiwnaang OR Abegweit OR Abitibiwinni OR Ahousaht OR Ahtahkakoop OR Aishihik OR Aitchelitz OR Aklavik OR Algonquin* OR Anicinape* OR "Animakee Wa Zhing" OR Anishina* OR aqam OR Atikamekw OR Atjanasca* OR Attawapiskat OR Aundeck-Omni-Kaning OR Batchewana OR "Bearfoot Onondaga" OR Beausoleil OR "Behdzi Ahda*" OR "Biigtigong Nishnaabeg" OR "Biinjitiwaabik Zaaging Anishinaabek" OR Bilijk OR "Binche Whut'en" OR "Bingwi Neyaashi Anishinaabek" OR "Canupawakpa Dakota" OR Cayuga OR Chawathil OR Cheam OR "Cheslatta Carrier" OR Chiniki OR Chipew* OR Cote OR Couchiching OR Cowessess OR Cowichan OR Cree OR "Dakota Plains" OR "Dakota Tipi" OR "Dechi Laot'i" OR Deline OR Dena OR Dene OR Denesuline OR "Deninu K'ue" OR Ditidaht OR "Dog Rib Rae" OR Dokis OR Eabametoong OR Ehattesaht OR Elsipogtog OR Ermineskin OR Esdilagh OR Esgenoopetitj OR Eskasoni OR Esquimalt OR Gameti OR Gayogohono OR Ginoogaming OR Gitanmaax OR Gitanyow OR Gitsegukla OR Gitwangak OR Gitxaala OR Glooscap OR Gwawaenuk OR "Gwichya Gwich'in" OR Gwitchin OR Hagwilget OR Haisla OR Halalt OR Haudenosaunee OR Heiltsuk OR Hesquiaht OR Hiawatha OR Homalco OR Hupacasath OR Huu-ay-aht OR Inninuwug OR Innu OR Innue OR Innus OR Inuvik OR Iskatewizaagegan OR Iskut OR "Ka'a'gee Tu" OR Kahkewistahaw OR "Kasabonika Lake" OR Kashechewan OR Katzie OR Kawacatoose OR Kebaowek OR Keeseekoose OR Keeseekoowenin OR Kee-Way-Win OR "Kinistin Saulteaux" OR Kinonjeoshtegon OR Kispiox OR Kitselas OR Kitsumkalum OR Kluane OR Kutchin OR Kwadacha OR Kwakiutl OR "Kwanlin Dun" OR Kwaw-kwaw-Apilt OR Kwiakah OR "Kwikwasut'inuxw Haxwa'mis" OR Kwikwetlem OR "Lax Kw'alaams" OR "Lheidli T'enneh" OR "Liidlii Kue" OR "Listuguj Mi'gmaq" OR Lyackson OR Magnetawan OR Malahat OR Maliseet OR Mamalilikulla OR Matachewan OR Matsqui OR Mattagami OR "M'Chigeeng" OR Membertou OR Metepenagiag OR Metlakatla OR Miawpukek OR Michipicoten OR Micmacs OR Millbrook OR Mishkeegogamang OR Mississauga* OR Mitaanjigamiing OR Mohawk* OR Montagnais OR Moosomin OR Moravian OR Mowachaht OR Muchalaht OR Munsee-Delaware OR Muscowpetung OR Muskoday OR Muskowekwan OR Musqueam OR "Nacho Nyak Dun" OR "Nadleh Whuten" OR Naicatchewenin OR "Nak'azdli Whut'en" OR Nakota OR Naotkamegwanning OR Naskapi OR Natoaganeg OR Nee-Tahi-Buhn OR Nehiyawak OR Nekaneet OR Neskantaga OR Neskonlith OR Nibinamik OR Nigigoonsiminikaaning OR Nipissing OR Nishnaabeg OR Nooaitch OR Nuchatlaht OR Nuxalk OR Ochapowace OR O-Chi-Chak-Ko-Sipi OR Odanak OR Ojibw* OR Okanagan OR Okanese OR Okemasis OR Oneida OR Oromocto OR Osoyoos OR Pabineau OR Pacheedaht OR "Paqtnkek Mi'kmaw" OR Pasqua OR Pauingassi OR Pauquachin OR "Pays Plat" OR Peguis OR "Pehdzeh Ki" OR Pekuakamiulnuatsh OR Penelakut OR Piapot OR Piikani OR Pikangikum OR Pinaymootang OR Potlotek OR "Qalipu Mi'kmaq" OR Qualicum OR Quatsino OR Sagamok OR Sahgaiehcan OR Samahquam OR Saugeen OR Saulteau OR Saulteaux OR Sechelt OR Secwepemc* OR Semiahmoo OR Seneca OR Shackan OR Shamattawa OR Shawanaga OR Sheguiandah OR Sheshegwaning OR Shuswap OR Siksika OR Simpcw OR Sioux OR Siska OR Skatin OR Skawahlook OR Skeetchestn OR Skidegate OR "Skin Tyee" OR Skowkale OR Skownan OR Skuppah OR Skwah OR Snuneymuxw OR Songhees OR Soowahlie OR Splatsin OR Spuzzum OR Squamish OR Squiala OR "Stswecem'c Xget'tem" OR Sumas OR "Ta'an Kwach'an" OR Tagish OR Tahltan OR Takla OR Tallcree OR "Taykwa Tagamou" OR "Teetl'it Gwich'in" OR Temagami OR Thessalon OR Timiskaming OR "Tk'emlúps te Secwépemc" OR Tla-o-qui-aht OR Tlatlasikwala OR Tlingit OR Tlowitsis OR Tootinaowaziibeeng OR Toquaht OR "Tr'ondëk Hwëch'in" OR Tsartlip OR Tsawout OR Tsawwassen OR Tseshaht OR Tseycum OR "Ts'il Kaz Koh" OR Tsleil-Waututh OR Tuscarora OR Tutchone OR Tzeachten OR Uchucklesaht OR Ucluelet OR Ulkatcho OR Wabaseemoong OR Wabauskang OR Wagmatcook OR Wahnapitae OR "Wahpeton Dakota" OR Wahsipekuk OR Wapekeka OR Wasagamack OR Wasauksing OR Waswanipi OR Wawakapewin OR Waywayseecappo OR Webequie OR Weenusk OR Wemotaci OR Wendat OR "Wha Ti" OR Whapmagoostui OR "Whitecap Dakota" OR Wikwemikong OR Witset OR Wolastoqiyik OR Wuikinuxv OR Wunnumin OR "Wuskwi Sipihk" OR Xatsull OR "Xeni Gwet'in" OR Yakweakwioose OR Yekooche OR "Yellow Quill" OR Yellowknife OR Yellowknives OR Zhiibaahaasing) OR AB(Anishnawbek OR "?Akisq'nuk" OR "?aq'am" OR "?Esdilagh" OR "Abénakis" OR "Akisq'nuk" OR "aq'am" OR "Che:k:tles7et'h'" OR "Da'naxda'xw" OR "Dzawada'enuxw" OR "Esk'etemc" OR "Gitga'at" OR "Gitlaxt'aamiks" OR "Gwa'Sala-Nakwaxda'xw" OR "Gwich'in" OR "Ka:'yu:'k't'h'" OR "Kanyen'kehaka" OR "Kapawe'no" OR "K'atlodeeche" OR "K'ómoks" OR "Leq'á:mel" OR "Lil'wat" OR "Malécite" OR "Mi'kmaq" OR "Mowachaht/Muchalaht" OR "Nisga'a" OR "N'Quatqua" OR "O'Chiese" OR "Ononda'gega'" OR "Onondowága'" OR "Onyota'a:ka" OR "Saik'uz" OR "Sambaa K'e " OR "Shxwhá:y" OR "Shxw'ow'hamel" OR "SIPEKNE'KATIK" OR "Skaru:reh" OR "Sq'éwlets" OR "Stellat'en" OR "Sts'ailes" OR "Stz'uminus" OR "T'it'q'et" OR "Tla'amin" OR "Tl'azt'en" OR "Tl'etinqox" OR "Tsal'alh" OR "Tsideldel" OR "Ts'kw'aylaxw" OR "T'Sou-ke" OR "Ts'uubaa-asatx" OR "Tsuut'ina" OR "We'koqma'q" OR "Wet'suwet'en" OR "Xai'xais" OR "Xaxli'p" OR "Yunesit'in" OR ((Acadia OR Carmacks OR Champagne OR Creek OR Delaware OR Douglas OR Montana OR Similkameen) N2 (Indian OR Indians OR Tribe OR Tribes OR native OR natives OR nation OR people*)) OR Aamjiwnaang OR Abegweit OR Abitibiwinni OR Ahousaht OR Ahtahkakoop OR Aishihik OR Aitchelitz OR Aklavik OR Algonquin* OR Anicinape* OR "Animakee Wa Zhing" OR Anishina* OR aqam OR Atikamekw OR Atjanasca* OR Attawapiskat OR Aundeck-Omni-Kaning OR Batchewana OR "Bearfoot Onondaga" OR Beausoleil OR "Behdzi Ahda*" OR "Biigtigong Nishnaabeg" OR "Biinjitiwaabik Zaaging Anishinaabek" OR Bilijk OR "Binche Whut'en" OR "Bingwi Neyaashi Anishinaabek" OR "Canupawakpa Dakota" OR Cayuga OR Chawathil OR Cheam OR "Cheslatta Carrier" OR Chiniki OR Chipew* OR Cote OR Couchiching OR Cowessess OR Cowichan OR Cree OR "Dakota Plains" OR "Dakota Tipi" OR "Dechi Laot'i" OR Deline OR Dena OR Dene OR Denesuline OR "Deninu K'ue" OR Ditidaht OR "Dog Rib Rae" OR Dokis OR Eabametoong OR Ehattesaht OR Elsipogtog OR Ermineskin OR Esdilagh OR Esgenoopetitj OR Eskasoni OR Esquimalt OR Gameti OR Gayogohono OR Ginoogaming OR Gitanmaax OR Gitanyow OR Gitsegukla OR Gitwangak OR Gitxaala OR Glooscap OR Gwawaenuk OR "Gwichya Gwich'in" OR Gwitchin OR Hagwilget OR Haisla OR Halalt OR Haudenosaunee OR Heiltsuk OR Hesquiaht OR Hiawatha OR Homalco OR Hupacasath OR Huu-ay-aht OR Inninuwug OR Innu OR Innue OR Innus OR Inuvik OR Iskatewizaagegan OR Iskut OR "Ka'a'gee Tu" OR Kahkewistahaw OR "Kasabonika Lake" OR Kashechewan OR Katzie OR Kawacatoose OR Kebaowek OR Keeseekoose OR Keeseekoowenin OR Kee-Way-Win OR "Kinistin Saulteaux" OR Kinonjeoshtegon OR Kispiox OR Kitselas OR Kitsumkalum OR Kluane OR Kutchin OR Kwadacha OR Kwakiutl OR "Kwanlin Dun" OR Kwaw-kwaw-Apilt OR Kwiakah OR "Kwikwasut'inuxw Haxwa'mis" OR Kwikwetlem OR "Lax Kw'alaams" OR "Lheidli T'enneh" OR "Liidlii Kue" OR "Listuguj Mi'gmaq" OR Lyackson OR Magnetawan OR Malahat OR Maliseet OR Mamalilikulla OR Matachewan OR Matsqui OR Mattagami OR "M'Chigeeng" OR Membertou OR Metepenagiag OR Metlakatla OR Miawpukek OR Michipicoten OR Micmacs OR Millbrook OR Mishkeegogamang OR Mississauga* OR Mitaanjigamiing OR Mohawk* OR Montagnais OR Moosomin OR Moravian OR Mowachaht OR Muchalaht OR Munsee-Delaware OR Muscowpetung OR Muskoday OR Muskowekwan OR Musqueam OR "Nacho Nyak Dun" OR "Nadleh Whuten" OR Naicatchewenin OR "Nak'azdli Whut'en" OR Nakota OR Naotkamegwanning OR Naskapi OR Natoaganeg OR Nee-Tahi-Buhn OR Nehiyawak OR Nekaneet OR Neskantaga OR Neskonlith OR Nibinamik OR Nigigoonsiminikaaning OR Nipissing OR Nishnaabeg OR Nooaitch OR Nuchatlaht OR Nuxalk OR Ochapowace OR O-Chi-Chak-Ko-Sipi OR Odanak OR Ojibw* OR Okanagan OR Okanese OR Okemasis OR Oneida OR Oromocto OR Osoyoos OR Pabineau OR Pacheedaht OR "Paqtnkek Mi'kmaw" OR Pasqua OR Pauingassi OR Pauquachin OR "Pays Plat" OR Peguis OR "Pehdzeh Ki" OR Pekuakamiulnuatsh OR Penelakut OR Piapot OR Piikani OR Pikangikum OR Pinaymootang OR Potlotek OR "Qalipu Mi'kmaq" OR Qualicum OR Quatsino OR Sagamok OR Sahgaiehcan OR Samahquam OR Saugeen OR Saulteau OR Saulteaux OR Sechelt OR Secwepemc* OR Semiahmoo OR Seneca OR Shackan OR Shamattawa OR Shawanaga OR Sheguiandah OR Sheshegwaning OR Shuswap OR Siksika OR Simpcw OR Sioux OR Siska OR Skatin OR Skawahlook OR Skeetchestn OR Skidegate OR "Skin Tyee" OR Skowkale OR Skownan OR Skuppah OR Skwah OR Snuneymuxw OR Songhees OR Soowahlie OR Splatsin OR Spuzzum OR Squamish OR Squiala OR "Stswecem'c Xget'tem" OR Sumas OR "Ta'an Kwach'an" OR Tagish OR Tahltan OR Takla OR Tallcree OR "Taykwa Tagamou" OR "Teetl'it Gwich'in" OR Temagami OR Thessalon OR Timiskaming OR "Tk'emlúps te Secwépemc" OR Tla-o-qui-aht OR Tlatlasikwala OR Tlingit OR Tlowitsis OR Tootinaowaziibeeng OR Toquaht OR "Tr'ondëk Hwëch'in" OR Tsartlip OR Tsawout OR Tsawwassen OR Tseshaht OR Tseycum OR "Ts'il Kaz Koh" OR Tsleil-Waututh OR Tuscarora OR Tutchone OR Tzeachten OR Uchucklesaht OR Ucluelet OR Ulkatcho OR Wabaseemoong OR Wabauskang OR Wagmatcook OR Wahnapitae OR "Wahpeton Dakota" OR Wahsipekuk OR Wapekeka OR Wasagamack OR Wasauksing OR Waswanipi OR Wawakapewin OR Waywayseecappo OR Webequie OR Weenusk OR Wemotaci OR Wendat OR "Wha Ti" OR Whapmagoostui OR "Whitecap Dakota" OR Wikwemikong OR Witset OR Wolastoqiyik OR Wuikinuxv OR Wunnumin OR "Wuskwi Sipihk" OR Xatsull OR "Xeni Gwet'in" OR Yakweakwioose OR Yekooche OR "Yellow Quill" OR Yellowknife OR Yellowknives OR Zhiibaahaasing) |  | 126 | 136 |
| S4 | TI("Degexit'an" OR "Yup'ik" OR ("Bear River" N2 (Indian OR Indians OR Tribe OR Tribes OR native OR natives OR nation OR people*)) OR Abenaki OR Achumawi OR Ahtna OR "Akimel Oodham" OR "Akimel O'odham" OR "Akimel Ootham" OR "Akimel O'otham" OR Aleut* OR Algonkian* OR Alsea OR Aluutiq OR Apalachee OR Arikara OR Atakapa OR Athabascan OR Athabaskan OR Athapascan OR Athapaskan OR Bannock OR Bidai OR Biloxi OR Blackfeet OR Blackfoot OR Caddoan OR Calice OR Calusa OR Carrizo OR Cayuse OR Chemakum OR Cheraw OR Cherokee OR Chetco OR Cheyenne OR Chicora OR Chilula OR Chimariko OR Clatskanie OR Coahuiltecan OR "Coeur d'Alene" OR Costanoan OR Cree OR Croatan OR Croatoan OR Cusabo OR Edisto OR Eskaleut* OR Eskimo OR Eskimo-Aleut OR Esselen OR Eyak OR "Fox and Sauk" OR Gabrielino OR "Gros Ventre" OR Guale OR "Gwich'in" OR Halchidhoma OR Hidatsa OR Hitchiti OR Hopi OR Houma OR Huichol* OR Hupa OR Ingalik OR Inupiaq OR Ioway OR Iroquoi* OR Jacome OR Jano OR Jeaga OR Juaneno OR Jumano OR Kalapuya OR Kansa OR Karankawa OR Karok OR Kashaya OR Kato OR Kawaiisu OR Kitanemuk OR Kitsai OR Klallam OR Koasati OR Kolchan OR Konkow OR Koyukon OR Lakota* OR Lassik OR Laurentian OR Lenape OR "Lenni Lenape" OR Mahican OR Mandan OR Maricopa OR Massachusett OR Mattole OR Minisink OR Missouria OR Mohave OR Mohawk OR Mohican OR Molalla OR Multnomah OR Munsee OR Muscoge* OR Naabeeho OR Nakoda OR Nanticoke OR Natchez OR Nauset OR Navaho OR Niantic OR Nipmuc OR Nisenan OR Nongatl OR Oconee OR Ofo OR Ohlone OR Ojibw* OR Okanagan OR Oneida OR Otoe OR Palouse OR Panamint OR Papago OR Patwin OR "Pee Dee" OR Pennacook OR Pequot OR Pima OR Piro OR Pocasset OR Pocomtuc OR Pokanoket OR Poospatuck OR Powhatan OR Quechan OR Quileute OR Quinault OR Quiripi OR Salinan OR Samich OR Saponi OR Serrano OR Shasta OR Shawnee OR Siksikaitsitapi OR Sinkyone OR Siouan OR Sioux OR Snohomish OR Snoqualmie OR Squaxin OR Suma OR Susquehannock OR Takelma OR Tanacross OR Tanana OR Tataviam OR Tawakoni OR Tequesta OR Tillamook OR Timucua OR Tlingit OR "Tohono Oodham" OR Tongva OR Tsimshian OR Tubatulabal OR Tutelo OR Tututni OR Unalachtigo OR Unami OR Unangan OR Unangas OR Unkechaug OR Waccamaw OR Walapai OR Wappo OR Wasco OR Whilkut OR Wichita OR Winnebago OR Wintu OR Wishram OR Yahi OR Yakima OR Yamasee OR Yana OR Yaqu OR Yuchi OR Yuki OR Yuma OR Yupik OR Yurok) OR AB("Degexit'an" OR "Yup'ik" OR ("Bear River" N2 (Indian OR Indians OR Tribe OR Tribes OR native OR natives OR nation OR people*)) OR Abenaki OR Achumawi OR Ahtna OR "Akimel Oodham" OR "Akimel O'odham" OR "Akimel Ootham" OR "Akimel O'otham" OR Aleut* OR Algonkian* OR Alsea OR Aluutiq OR Apalachee OR Arikara OR Atakapa OR Athabascan OR Athabaskan OR Athapascan OR Athapaskan OR Bannock OR Bidai OR Biloxi OR Blackfeet OR Blackfoot OR Caddoan OR Calice OR Calusa OR Carrizo OR Cayuse OR Chemakum OR Cheraw OR Cherokee OR Chetco OR Cheyenne OR Chicora OR Chilula OR Chimariko OR Clatskanie OR Coahuiltecan OR "Coeur d'Alene" OR Costanoan OR Cree OR Croatan OR Croatoan OR Cusabo OR Edisto OR Eskaleut* OR Eskimo OR Eskimo-Aleut OR Esselen OR Eyak OR "Fox and Sauk" OR Gabrielino OR "Gros Ventre" OR Guale OR "Gwich'in" OR Halchidhoma OR Hidatsa OR Hitchiti OR Hopi OR Houma OR Huichol* OR Hupa OR Ingalik OR Inupiaq OR Ioway OR Iroquoi* OR Jacome OR Jano OR Jeaga OR Juaneno OR Jumano OR Kalapuya OR Kansa OR Karankawa OR Karok OR Kashaya OR Kato OR Kawaiisu OR Kitanemuk OR Kitsai OR Klallam OR Koasati OR Kolchan OR Konkow OR Koyukon OR Lakota* OR Lassik OR Laurentian OR Lenape OR "Lenni Lenape" OR Mahican OR Mandan OR Maricopa OR Massachusett OR Mattole OR Minisink OR Missouria OR Mohave OR Mohawk OR Mohican OR Molalla OR Multnomah OR Munsee OR Muscoge* OR Naabeeho OR Nakoda OR Nanticoke OR Natchez OR Nauset OR Navaho OR Niantic OR Nipmuc OR Nisenan OR Nongatl OR Oconee OR Ofo OR Ohlone OR Ojibw* OR Okanagan OR Oneida OR Otoe OR Palouse OR Panamint OR Papago OR Patwin OR "Pee Dee" OR Pennacook OR Pequot OR Pima OR Piro OR Pocasset OR Pocomtuc OR Pokanoket OR Poospatuck OR Powhatan OR Quechan OR Quileute OR Quinault OR Quiripi OR Salinan OR Samich OR Saponi OR Serrano OR Shasta OR Shawnee OR Siksikaitsitapi OR Sinkyone OR Siouan OR Sioux OR Snohomish OR Snoqualmie OR Squaxin OR Suma OR Susquehannock OR Takelma OR Tanacross OR Tanana OR Tataviam OR Tawakoni OR Tequesta OR Tillamook OR Timucua OR Tlingit OR "Tohono Oodham" OR Tongva OR Tsimshian OR Tubatulabal OR Tutelo OR Tututni OR Unalachtigo OR Unami OR Unangan OR Unangas OR Unkechaug OR Waccamaw OR Walapai OR Wappo OR Wasco OR Whilkut OR Wichita OR Winnebago OR Wintu OR Wishram OR Yahi OR Yakima OR Yamasee OR Yana OR Yaqu OR Yuchi OR Yuki OR Yuma OR Yupik OR Yurok) |  | 1892 | 1836 |
| S3 | TI("Asa'carsarmiut" OR "Dee-ni'" OR "Mi'kmaq" OR "S'Klallam" OR "Shoonaq'" OR "Sun'aq" OR (("Ivanof Bay" OR "Shoalwater Bay" OR "Spirit Lake" OR "Squaxin Island" OR Dakota OR Dakotah OR Wichita OR keechi OR waco OR Tawakonie OR Cheyenne OR Creek OR Crow OR Hoonah OR Kasigluk OR Maricopa OR Miami OR Omaha OR Oneida OR Ottawa OR Peoria OR Salamatof OR Sitka OR Snoqualmie OR Spokane OR Winnebago OR Wyandotte) N2 (Indian OR Indians OR Tribe OR Tribes OR native OR natives OR nation OR people*)) OR ((Native OR indian OR traditional) N2 (village OR council OR community OR association)) OR Absentee-Shawnee OR Achomawi OR Agdaagux OR "Agua Caliente" OR Ajumawi OR Ak-Chin OR Akiachak OR Akiak OR Alabama-Coushatta OR Alabama-Quassarte OR Alatna OR Algaaciq OR Algin OR Allakaket OR "Alturas Indian Rancheria" OR Alutiiq OR Angoon OR Anvik OR Apache OR Aporige OR Aquinnah OR arapaho OR Aroostook OR Assiniboine OR Astarawi OR Atsugewi OR Atwamsini OR Bdewakantunwan OR Caddo OR Cahto OR Cahuilla OR Catawba OR Cayuga OR Chalkyitsik OR Cheesh-Na OR Chehalis OR Chemehuevi OR Cher-Ae OR Chevak OR Cheyenne-Arapaho OR Chickahominy OR Chickaloon OR Chickasaw OR Chignik OR Chilkat OR Chilkoot OR Chinik OR Chinook OR Chippewa* OR Chitimacha OR Choctaw OR Chukchansi OR Chuloonawick OR Chumash OR Cocopah OR Colusa OR Comanche OR Coos OR Cosmit OR Coushatta OR Cowlitz OR "Cross Village" OR Cupeno OR Curyung OR Diegueno OR Dine' OR Egegik OR Eklutna OR Elwha OR "Emmonak Village" OR "Evansville Village" OR Ewiiaapaayp OR "Galena Village" OR Goshute OR "Gulkana Village Council" OR Haida OR Hammawi OR Havasupai OR "Healy Lake Village" OR Hewisedawi OR Ho-Chunk OR Hoh OR Hokan OR Holikachuk OR "Holy Cross Tribe" OR Hualapai OR "Hughes Village" OR "Huslia Village" OR "Hydaburg Cooperative Association" OR "Igiugig Village" OR Iipay OR Ilmawi OR Inaja OR Inupiat OR Ione OR Itsatawi OR "Kaguyak Village" OR Kaibab OR "Kaktovik Village" OR Kalispel OR Kanosh OR Karuk OR Kashia OR Kaw OR Keetoowah OR "Kenaitze Indian Tribe" OR "Ketchikan Indian Community" OR "Keweenaw Bay" OR Kialegee OR Kickapoo OR "King Island Native Community" OR "King Salmon Tribe" OR Kiowa OR Klamath OR "Klawock Cooperative Association" OR Kletsel OR "Knik Tribe" OR "Koi Nation" OR "Kokhanok Village" OR Koosharem OR Kootenai OR Kosalektawi OR "Koyukuk Native Village" OR Kumeyaay OR Kwalhioqua OR "Lesnoi Village" OR "Levelock Village" OR "Lime Village" OR "Louden Village" OR Luiseno OR Lummi OR Madesi OR Maidu OR Makah OR Maliseet OR "Manley Hot Springs Village" OR "Manokotak Village" OR Manzanita OR Mashantucket OR Match-e-be-nash-she-wish OR Mattaponi OR "McGrath Native Village" OR Mdewakanton OR Mechoopda OR Menominee OR Me-Wuk OR Miccosukee OR Micmacs OR "Mission Indians" OR Miwok OR Miwuk OR Modoc OR Mohegan OR Mojave OR Monacan OR "Mono Indians" OR Muckleshoot OR Muscogee OR "Na Dené" OR Nansemond OR Narragansett OR Navajo OR "New Koliganek Village" OR "New Stuyahok Village" OR "Newhalen Village" OR "Newtok Village" OR "Nez Perce" OR "Nikolai Village" OR "Ninilchik Village" OR Nisqually OR Nomlaki OR "Nondalton Village" OR Nooksack OR "Northway Village" OR Nottawaseppi OR "Nulato Village" OR Nunakauyarmiut OR Odawa OR Ohgsenakale OR "Ohkay Owingeh" OR Onondaga OR Oreilles OR "Organized Village of Grayling" OR "Organized Village of Kake" OR "Organized Village of Kasaan" OR "Organized Village of Kwethluk" OR "Organized Village of Saxman" OR Orutsararmiut OR Osage OR Otoe-Missouria OR Paiute OR Paiute-Shoshone OR Pala OR Pamunkey OR Pascua OR Paskenta OR Passamaquoddy OR "Pauloff Harbor Village" OR Pauma OR Pawnee OR Pechanga OR "Pechanga " OR "Peck Indian Reservation" OR "Pedro Bay Village" OR Pee-Posh OR Penobscot OR Penutian OR Perquot OR "Pima People" OR Pima-Maricopa OR Poarch OR Pokagon OR Pomo OR Ponca OR "Portage Creek Village" OR Potawatomi OR Pottawatomi OR Pueblo OR Puyallup OR "Qagan Tayagungin" OR Qawalangin OR Quapaw OR Quechan OR "Rampart Village" OR Rappahannock OR Resighini OR "Sac & Fox" OR "Sac and Fox" OR "Saint George Island" OR "Saint Paul Island" OR Salish OR Samish OR Santee OR Sauk-Suiattle OR "Seldovia Village Tribe" OR Seminole OR Seneca OR Seneca-Cayuga OR Shinnecock OR Shivwits OR Shoshone OR Shoshone-Bannock OR Shoshone-Paiute OR Siuslaw OR Skagit OR "Skagway Village" OR Skokomish OR "South Naknek Village" OR "Stebbins Community Association" OR Stillaguamish OR Suquamish OR Swinomish OR "Takotna Village" OR Tangirnaq OR Tejon OR "Telida Village" OR Te-Moak OR Thlopthlocco OR "Tlingit & Haida" OR "Tohono O'odham" OR Tolowa OR Tonawanda OR Tonkawa OR Tulalip OR Tunica-Biloxi OR Tuolumne OR Tuscarora OR "Twin Hills Village" OR "Ugashik Village" OR Umatilla OR Umkumiut OR Umpqua OR Ute OR "Utu Utu Gwaitu" OR "Village of Alakanuk" OR "Village of Anaktuvuk Pass" OR "Village of Aniak" OR "Village of Atmautluak" OR "Village of Bill Moore's Slough" OR "Village of Chefornak" OR "Village of Clarks Point" OR "Village of Crooked Creek" OR "Village of Dot Lake" OR "Village of Iliamna" OR "Village of Kalskag" OR "Village of Kaltag" OR "Village of Kotlik" OR "Village of Lower Kalskag" OR "Village of Ohogamiut" OR "Village of Red Devil" OR "Village of Sleetmute" OR "Village of Solomon" OR "Village of Stony River" OR "Village of Venetie" OR "Village of Wainwright" OR Wailaki OR Wampanoag OR "Warm Springs Reservation" OR Washoe OR Willapa OR Winnemucca OR Wintun OR Wiyot OR "Wrangell Cooperative Association" OR Yakama OR Yakutat OR Yaqui OR Yavapai OR Yokuts OR Yuhaaviatam OR Yuima OR Yukian OR Yukut OR Yupiit OR Zuni) OR AB("Asa'carsarmiut" OR "Dee-ni'" OR "Mi'kmaq" OR "S'Klallam" OR "Shoonaq'" OR "Sun'aq" OR (("Ivanof Bay" OR "Shoalwater Bay" OR "Spirit Lake" OR "Squaxin Island" OR Dakota OR Dakotah OR Wichita OR keechi OR waco OR Tawakonie OR Cheyenne OR Creek OR Crow OR Hoonah OR Kasigluk OR Maricopa OR Miami OR Omaha OR Oneida OR Ottawa OR Peoria OR Salamatof OR Sitka OR Snoqualmie OR Spokane OR Winnebago OR Wyandotte) N2 (Indian OR Indians OR Tribe OR Tribes OR native OR natives OR nation OR people*)) OR ((Native OR indian OR traditional) N2 (village OR council OR community OR association)) OR Absentee-Shawnee OR Achomawi OR Agdaagux OR "Agua Caliente" OR Ajumawi OR Ak-Chin OR Akiachak OR Akiak OR Alabama-Coushatta OR Alabama-Quassarte OR Alatna OR Algaaciq OR Algin OR Allakaket OR "Alturas Indian Rancheria" OR Alutiiq OR Angoon OR Anvik OR Apache OR Aporige OR Aquinnah OR arapaho OR Aroostook OR Assiniboine OR Astarawi OR Atsugewi OR Atwamsini OR Bdewakantunwan OR Caddo OR Cahto OR Cahuilla OR Catawba OR Cayuga OR Chalkyitsik OR Cheesh-Na OR Chehalis OR Chemehuevi OR Cher-Ae OR Chevak OR Cheyenne-Arapaho OR Chickahominy OR Chickaloon OR Chickasaw OR Chignik OR Chilkat OR Chilkoot OR Chinik OR Chinook OR Chippewa* OR Chitimacha OR Choctaw OR Chukchansi OR Chuloonawick OR Chumash OR Cocopah OR Colusa OR Comanche OR Coos OR Cosmit OR Coushatta OR Cowlitz OR "Cross Village" OR Cupeno OR Curyung OR Diegueno OR Dine' OR Egegik OR Eklutna OR Elwha OR "Emmonak Village" OR "Evansville Village" OR Ewiiaapaayp OR "Galena Village" OR Goshute OR "Gulkana Village Council" OR Haida OR Hammawi OR Havasupai OR "Healy Lake Village" OR Hewisedawi OR Ho-Chunk OR Hoh OR Hokan OR Holikachuk OR "Holy Cross Tribe" OR Hualapai OR "Hughes Village" OR "Huslia Village" OR "Hydaburg Cooperative Association" OR "Igiugig Village" OR Iipay OR Ilmawi OR Inaja OR Inupiat OR Ione OR Itsatawi OR "Kaguyak Village" OR Kaibab OR "Kaktovik Village" OR Kalispel OR Kanosh OR Karuk OR Kashia OR Kaw OR Keetoowah OR "Kenaitze Indian Tribe" OR "Ketchikan Indian Community" OR "Keweenaw Bay" OR Kialegee OR Kickapoo OR "King Island Native Community" OR "King Salmon Tribe" OR Kiowa OR Klamath OR "Klawock Cooperative Association" OR Kletsel OR "Knik Tribe" OR "Koi Nation" OR "Kokhanok Village" OR Koosharem OR Kootenai OR Kosalektawi OR "Koyukuk Native Village" OR Kumeyaay OR Kwalhioqua OR "Lesnoi Village" OR "Levelock Village" OR "Lime Village" OR "Louden Village" OR Luiseno OR Lummi OR Madesi OR Maidu OR Makah OR Maliseet OR "Manley Hot Springs Village" OR "Manokotak Village" OR Manzanita OR Mashantucket OR Match-e-be-nash-she-wish OR Mattaponi OR "McGrath Native Village" OR Mdewakanton OR Mechoopda OR Menominee OR Me-Wuk OR Miccosukee OR Micmacs OR "Mission Indians" OR Miwok OR Miwuk OR Modoc OR Mohegan OR Mojave OR Monacan OR "Mono Indians" OR Muckleshoot OR Muscogee OR "Na Dené" OR Nansemond OR Narragansett OR Navajo OR "New Koliganek Village" OR "New Stuyahok Village" OR "Newhalen Village" OR "Newtok Village" OR "Nez Perce" OR "Nikolai Village" OR "Ninilchik Village" OR Nisqually OR Nomlaki OR "Nondalton Village" OR Nooksack OR "Northway Village" OR Nottawaseppi OR "Nulato Village" OR Nunakauyarmiut OR Odawa OR Ohgsenakale OR "Ohkay Owingeh" OR Onondaga OR Oreilles OR "Organized Village of Grayling" OR "Organized Village of Kake" OR "Organized Village of Kasaan" OR "Organized Village of Kwethluk" OR "Organized Village of Saxman" OR Orutsararmiut OR Osage OR Otoe-Missouria OR Paiute OR Paiute-Shoshone OR Pala OR Pamunkey OR Pascua OR Paskenta OR Passamaquoddy OR "Pauloff Harbor Village" OR Pauma OR Pawnee OR Pechanga OR "Pechanga " OR "Peck Indian Reservation" OR "Pedro Bay Village" OR Pee-Posh OR Penobscot OR Penutian OR Perquot OR "Pima People" OR Pima-Maricopa OR Poarch OR Pokagon OR Pomo OR Ponca OR "Portage Creek Village" OR Potawatomi OR Pottawatomi OR Pueblo OR Puyallup OR "Qagan Tayagungin" OR Qawalangin OR Quapaw OR Quechan OR "Rampart Village" OR Rappahannock OR Resighini OR "Sac & Fox" OR "Sac and Fox" OR "Saint George Island" OR "Saint Paul Island" OR Salish OR Samish OR Santee OR Sauk-Suiattle OR "Seldovia Village Tribe" OR Seminole OR Seneca OR Seneca-Cayuga OR Shinnecock OR Shivwits OR Shoshone OR Shoshone-Bannock OR Shoshone-Paiute OR Siuslaw OR Skagit OR "Skagway Village" OR Skokomish OR "South Naknek Village" OR "Stebbins Community Association" OR Stillaguamish OR Suquamish OR Swinomish OR "Takotna Village" OR Tangirnaq OR Tejon OR "Telida Village" OR Te-Moak OR Thlopthlocco OR "Tlingit & Haida" OR "Tohono O'odham" OR Tolowa OR Tonawanda OR Tonkawa OR Tulalip OR Tunica-Biloxi OR Tuolumne OR Tuscarora OR "Twin Hills Village" OR "Ugashik Village" OR Umatilla OR Umkumiut OR Umpqua OR Ute OR "Utu Utu Gwaitu" OR "Village of Alakanuk" OR "Village of Anaktuvuk Pass" OR "Village of Aniak" OR "Village of Atmautluak" OR "Village of Bill Moore's Slough" OR "Village of Chefornak" OR "Village of Clarks Point" OR "Village of Crooked Creek" OR "Village of Dot Lake" OR "Village of Iliamna" OR "Village of Kalskag" OR "Village of Kaltag" OR "Village of Kotlik" OR "Village of Lower Kalskag" OR "Village of Ohogamiut" OR "Village of Red Devil" OR "Village of Sleetmute" OR "Village of Solomon" OR "Village of Stony River" OR "Village of Venetie" OR "Village of Wainwright" OR Wailaki OR Wampanoag OR "Warm Springs Reservation" OR Washoe OR Willapa OR Winnemucca OR Wintun OR Wiyot OR "Wrangell Cooperative Association" OR Yakama OR Yakutat OR Yaqui OR Yavapai OR Yokuts OR Yuhaaviatam OR Yuima OR Yukian OR Yukut OR Yupiit OR Zuni) |  | 824 | 463 |
| S2 | TI(((Canada OR Canadian*) N2 (native* OR indigenous OR aborigin*)) OR Metis OR "Métis" OR Michif* OR Mitchif OR Mechif OR "Métif" OR "Métchif" OR Metif OR Metchif OR Inuit OR "First Nations" OR "First Nation") OR AB(((Canada OR Canadian*) N2 (native* OR indigenous OR aborigin*)) OR Metis OR "Métis" OR Michif* OR Mitchif OR Mechif OR "Métif" OR "Métchif" OR Metif OR Metchif OR Inuit OR "First Nations" OR "First Nation") |  | 83 | 94 |
| S1 | DE("American Indians") OR TI(((American* OR Alaska*) N2 (Native* OR Indian OR Indians OR indigenous OR aborigin*)) OR Amerind*) OR AB(((American* OR Alaska*) N2 (Native* OR Indian OR Indians OR indigenous OR aborigin*)) OR Amerind*) |  | 913 | 711 |

### Web of Science (Clarivate)

**Field codes:** TI = article title; AB = abstract; kf = keyword heading word; AK = author supplied keywords; KP = Keywords Plus
**Proximity operator:** NEAR/# 
**Truncation:**  *

**Entitlements:**
- WOS.SCI: 1900 to 2024
- WOS.AHCI: 1975 to 2024
- WOS.ESCI: 2018 to 2024
- WOS.SSCI: 1900 to 2024

**Initial Search Date**: Thu Jun 08 2023 10:19:43 GMT-0600 (Mountain Daylight Time)
**Update Date:** Thu Nov 21 2024 15:12:51 GMT-0700 (Mountain Standard Time)

| **#** | **Search Query** | **6/08/23**  **Results** | **11/21/24**  **Results** |
| --- | --- | --- | --- |
| 1 | TI=(((American* OR Alaska*) NEAR/2 (Native* OR Indian OR Indians OR indigenous OR aborigin*)) OR Amerind*) OR AB=(((American* OR Alaska*) NEAR/2 (Native* OR Indian OR Indians OR indigenous OR aborigin*)) OR Amerind*) OR AK=(((American* OR Alaska*) NEAR/2 (Native* OR Indian OR Indians OR indigenous OR aborigin*)) OR Amerind*) OR KP=(((American* OR Alaska*) NEAR/2 (Native* OR Indian OR Indians OR indigenous OR aborigin*)) OR Amerind*) | 37434 | 41214 |
| 2 | TI=(((Canada OR Canadian*) NEAR/2 (native* OR indigenous OR aborigin*)) OR Metis OR "Métis" OR Michif* OR Mitchif OR Mechif OR "Métif" OR "Métchif" OR Metif OR Metchif OR Inuit OR "First Nations" OR "First Nation") OR AB=(((Canada OR Canadian*) NEAR/2 (native* OR indigenous OR aborigin*)) OR Metis OR "Métis" OR Michif* OR Mitchif OR Mechif OR "Métif" OR "Métchif" OR Metif OR Metchif OR Inuit OR "First Nations" OR "First Nation") OR AK=(((Canada OR Canadian*) NEAR/2 (native* OR indigenous OR aborigin*)) OR Metis OR "Métis" OR Michif* OR Mitchif OR Mechif OR "Métif" OR "Métchif" OR Metif OR Metchif OR Inuit OR "First Nations" OR "First Nation") OR KP=(((Canada OR Canadian*) NEAR/2 (native* OR indigenous OR aborigin*)) OR Metis OR "Métis" OR Michif* OR Mitchif OR Mechif OR "Métif" OR "Métchif" OR Metif OR Metchif OR Inuit OR "First Nations" OR "First Nation") | 11411 | 12982 |
| 3 | TI=(inuit or (arctic NEAR/2 people*)) OR AB=(inuit or (arctic NEAR/2 people*)) OR AK=(inuit or (arctic NEAR/2 people*)) OR KP=(inuit or (arctic NEAR/2 people*)) | 4348 | 4800 |
| 4 | TI=("Asa'carsarmiut" OR "Dee-ni'" OR "Mi'kmaq" OR "S'Klallam" OR "Shoonaq'" OR "Sun'aq" OR (("Ivanof Bay" OR "Shoalwater Bay" OR "Spirit Lake" OR "Squaxin Island" OR Dakota OR Dakotah OR Wichita OR keechi OR waco OR Tawakonie OR Cheyenne OR Creek OR Crow OR Hoonah OR Kasigluk OR Maricopa OR Miami OR Omaha OR Oneida OR Ottawa OR Peoria OR Salamatof OR Sitka OR Snoqualmie OR Spokane OR Winnebago OR Wyandotte) NEAR/2 (Indian OR Indians OR Tribe OR Tribes OR native OR natives OR nation OR people*)) OR ((Native OR indian OR traditional) NEAR/2 (village OR council OR community OR association)) OR Absentee-Shawnee OR Achomawi OR Agdaagux OR "Agua Caliente" OR Ajumawi OR Ak-Chin OR Akiachak OR Akiak OR Alabama-Coushatta OR Alabama-Quassarte OR Alatna OR Algaaciq OR Algin OR Allakaket OR "Alturas Indian Rancheria" OR Alutiiq OR Angoon OR Anvik OR Apache OR Aporige OR Aquinnah OR arapaho OR Aroostook OR Assiniboine OR Astarawi OR Atsugewi OR Atwamsini OR Bdewakantunwan OR Caddo OR Cahto OR Cahuilla OR Catawba OR Cayuga OR Chalkyitsik OR Cheesh-Na OR Chehalis OR Chemehuevi OR Cher-Ae OR Chevak OR Cheyenne-Arapaho OR Chickahominy OR Chickaloon OR Chickasaw OR Chignik OR Chilkat OR Chilkoot OR Chinik OR Chinook OR Chippewa* OR Chitimacha OR Choctaw OR Chukchansi OR Chuloonawick OR Chumash OR Cocopah OR Colusa OR Comanche OR Coos OR Cosmit OR Coushatta OR Cowlitz OR "Cross Village" OR Cupeno OR Curyung OR Diegueno OR Dine' OR Egegik OR Eklutna OR Elwha OR "Emmonak Village" OR "Evansville Village" OR Ewiiaapaayp OR "Galena Village" OR Goshute OR "Gulkana Village Council" OR Haida OR Hammawi OR Havasupai OR "Healy Lake Village" OR Hewisedawi OR Ho-Chunk OR Hoh OR Hokan OR Holikachuk OR "Holy Cross Tribe" OR Hualapai OR "Hughes Village" OR "Huslia Village" OR "Hydaburg Cooperative Association" OR "Igiugig Village" OR Iipay OR Ilmawi OR Inaja OR Inupiat OR Ione OR Itsatawi OR "Kaguyak Village" OR Kaibab OR "Kaktovik Village" OR Kalispel OR Kanosh OR Karuk OR Kashia OR Kaw OR Keetoowah OR "Kenaitze Indian Tribe" OR "Ketchikan Indian Community" OR "Keweenaw Bay" OR Kialegee OR Kickapoo OR "King Island Native Community" OR "King Salmon Tribe" OR Kiowa OR Klamath OR "Klawock Cooperative Association" OR Kletsel OR "Knik Tribe" OR "Koi Nation" OR "Kokhanok Village" OR Koosharem OR Kootenai OR Kosalektawi OR "Koyukuk Native Village" OR Kumeyaay OR Kwalhioqua OR "Lesnoi Village" OR "Levelock Village" OR "Lime Village" OR "Louden Village" OR Luiseno OR Lummi OR Madesi OR Maidu OR Makah OR Maliseet OR "Manley Hot Springs Village" OR "Manokotak Village" OR Manzanita OR Mashantucket OR Match-e-be-nash-she-wish OR Mattaponi OR "McGrath Native Village" OR Mdewakanton OR Mechoopda OR Menominee OR Me-Wuk OR Miccosukee OR Micmacs OR "Mission Indians" OR Miwok OR Miwuk OR Modoc OR Mohegan OR Mojave OR Monacan OR "Mono Indians" OR Muckleshoot OR Muscogee OR "Na Dené" OR Nansemond OR Narragansett OR Navajo OR "New Koliganek Village" OR "New Stuyahok Village" OR "Newhalen Village" OR "Newtok Village" OR "Nez Perce" OR "Nikolai Village" OR "Ninilchik Village" OR Nisqually OR Nomlaki OR "Nondalton Village" OR Nooksack OR "Northway Village" OR Nottawaseppi OR "Nulato Village" OR Nunakauyarmiut OR Odawa OR Ohgsenakale OR "Ohkay Owingeh" OR Onondaga OR Oreilles OR "Organized Village of Grayling" OR "Organized Village of Kake" OR "Organized Village of Kasaan" OR "Organized Village of Kwethluk" OR "Organized Village of Saxman" OR Orutsararmiut OR Osage OR Otoe-Missouria OR Paiute OR Paiute-Shoshone OR Pala OR Pamunkey OR Pascua OR Paskenta OR Passamaquoddy OR "Pauloff Harbor Village" OR Pauma OR Pawnee OR Pechanga OR "Pechanga " OR "Peck Indian Reservation" OR "Pedro Bay Village" OR Pee-Posh OR Penobscot OR Penutian OR Perquot OR "Pima People" OR Pima-Maricopa OR Poarch OR Pokagon OR Pomo OR Ponca OR "Portage Creek Village" OR Potawatomi OR Pottawatomi OR Pueblo OR Puyallup OR "Qagan Tayagungin" OR Qawalangin OR Quapaw OR Quechan OR "Rampart Village" OR Rappahannock OR Resighini OR "Sac & Fox" OR "Sac and Fox" OR "Saint George Island" OR "Saint Paul Island" OR Salish OR Samish OR Santee OR Sauk-Suiattle OR "Seldovia Village Tribe" OR Seminole OR Seneca OR Seneca-Cayuga OR Shinnecock OR Shivwits OR Shoshone OR Shoshone-Bannock OR Shoshone-Paiute OR Siuslaw OR Skagit OR "Skagway Village" OR Skokomish OR "South Naknek Village" OR "Stebbins Community Association" OR Stillaguamish OR Suquamish OR Swinomish OR "Takotna Village" OR Tangirnaq OR Tejon OR "Telida Village" OR Te-Moak OR Thlopthlocco OR "Tlingit & Haida" OR "Tohono O'odham" OR Tolowa OR Tonawanda OR Tonkawa OR Tulalip OR Tunica-Biloxi OR Tuolumne OR Tuscarora OR "Twin Hills Village" OR "Ugashik Village" OR Umatilla OR Umkumiut OR Umpqua OR Ute OR "Utu Utu Gwaitu" OR "Village of Alakanuk" OR "Village of Anaktuvuk Pass" OR "Village of Aniak" OR "Village of Atmautluak" OR "Village of Bill Moore's Slough" OR "Village of Chefornak" OR "Village of Clarks Point" OR "Village of Crooked Creek" OR "Village of Dot Lake" OR "Village of Iliamna" OR "Village of Kalskag" OR "Village of Kaltag" OR "Village of Kotlik" OR "Village of Lower Kalskag" OR "Village of Ohogamiut" OR "Village of Red Devil" OR "Village of Sleetmute" OR "Village of Solomon" OR "Village of Stony River" OR "Village of Venetie" OR "Village of Wainwright" OR Wailaki OR Wampanoag OR "Warm Springs Reservation" OR Washoe OR Willapa OR Winnemucca OR Wintun OR Wiyot OR "Wrangell Cooperative Association" OR Yakama OR Yakutat OR Yaqui OR Yavapai OR Yokuts OR Yuhaaviatam OR Yuima OR Yukian OR Yukut OR Yupiit OR Zuni) OR AB=("Asa'carsarmiut" OR "Dee-ni'" OR "Mi'kmaq" OR "S'Klallam" OR "Shoonaq'" OR "Sun'aq" OR (("Ivanof Bay" OR "Shoalwater Bay" OR "Spirit Lake" OR "Squaxin Island" OR Dakota OR Dakotah OR Wichita OR keechi OR waco OR Tawakonie OR Cheyenne OR Creek OR Crow OR Hoonah OR Kasigluk OR Maricopa OR Miami OR Omaha OR Oneida OR Ottawa OR Peoria OR Salamatof OR Sitka OR Snoqualmie OR Spokane OR Winnebago OR Wyandotte) NEAR/2 (Indian OR Indians OR Tribe OR Tribes OR native OR natives OR nation OR people*)) OR ((Native OR indian OR traditional) NEAR/2 (village OR council OR community OR association)) OR Absentee-Shawnee OR Achomawi OR Agdaagux OR "Agua Caliente" OR Ajumawi OR Ak-Chin OR Akiachak OR Akiak OR Alabama-Coushatta OR Alabama-Quassarte OR Alatna OR Algaaciq OR Algin OR Allakaket OR "Alturas Indian Rancheria" OR Alutiiq OR Angoon OR Anvik OR Apache OR Aporige OR Aquinnah OR arapaho OR Aroostook OR Assiniboine OR Astarawi OR Atsugewi OR Atwamsini OR Bdewakantunwan OR Caddo OR Cahto OR Cahuilla OR Catawba OR Cayuga OR Chalkyitsik OR Cheesh-Na OR Chehalis OR Chemehuevi OR Cher-Ae OR Chevak OR Cheyenne-Arapaho OR Chickahominy OR Chickaloon OR Chickasaw OR Chignik OR Chilkat OR Chilkoot OR Chinik OR Chinook OR Chippewa* OR Chitimacha OR Choctaw OR Chukchansi OR Chuloonawick OR Chumash OR Cocopah OR Colusa OR Comanche OR Coos OR Cosmit OR Coushatta OR Cowlitz OR "Cross Village" OR Cupeno OR Curyung OR Diegueno OR Dine' OR Egegik OR Eklutna OR Elwha OR "Emmonak Village" OR "Evansville Village" OR Ewiiaapaayp OR "Galena Village" OR Goshute OR "Gulkana Village Council" OR Haida OR Hammawi OR Havasupai OR "Healy Lake Village" OR Hewisedawi OR Ho-Chunk OR Hoh OR Hokan OR Holikachuk OR "Holy Cross Tribe" OR Hualapai OR "Hughes Village" OR "Huslia Village" OR "Hydaburg Cooperative Association" OR "Igiugig Village" OR Iipay OR Ilmawi OR Inaja OR Inupiat OR Ione OR Itsatawi OR "Kaguyak Village" OR Kaibab OR "Kaktovik Village" OR Kalispel OR Kanosh OR Karuk OR Kashia OR Kaw OR Keetoowah OR "Kenaitze Indian Tribe" OR "Ketchikan Indian Community" OR "Keweenaw Bay" OR Kialegee OR Kickapoo OR "King Island Native Community" OR "King Salmon Tribe" OR Kiowa OR Klamath OR "Klawock Cooperative Association" OR Kletsel OR "Knik Tribe" OR "Koi Nation" OR "Kokhanok Village" OR Koosharem OR Kootenai OR Kosalektawi OR "Koyukuk Native Village" OR Kumeyaay OR Kwalhioqua OR "Lesnoi Village" OR "Levelock Village" OR "Lime Village" OR "Louden Village" OR Luiseno OR Lummi OR Madesi OR Maidu OR Makah OR Maliseet OR "Manley Hot Springs Village" OR "Manokotak Village" OR Manzanita OR Mashantucket OR Match-e-be-nash-she-wish OR Mattaponi OR "McGrath Native Village" OR Mdewakanton OR Mechoopda OR Menominee OR Me-Wuk OR Miccosukee OR Micmacs OR "Mission Indians" OR Miwok OR Miwuk OR Modoc OR Mohegan OR Mojave OR Monacan OR "Mono Indians" OR Muckleshoot OR Muscogee OR "Na Dené" OR Nansemond OR Narragansett OR Navajo OR "New Koliganek Village" OR "New Stuyahok Village" OR "Newhalen Village" OR "Newtok Village" OR "Nez Perce" OR "Nikolai Village" OR "Ninilchik Village" OR Nisqually OR Nomlaki OR "Nondalton Village" OR Nooksack OR "Northway Village" OR Nottawaseppi OR "Nulato Village" OR Nunakauyarmiut OR Odawa OR Ohgsenakale OR "Ohkay Owingeh" OR Onondaga OR Oreilles OR "Organized Village of Grayling" OR "Organized Village of Kake" OR "Organized Village of Kasaan" OR "Organized Village of Kwethluk" OR "Organized Village of Saxman" OR Orutsararmiut OR Osage OR Otoe-Missouria OR Paiute OR Paiute-Shoshone OR Pala OR Pamunkey OR Pascua OR Paskenta OR Passamaquoddy OR "Pauloff Harbor Village" OR Pauma OR Pawnee OR Pechanga OR "Pechanga " OR "Peck Indian Reservation" OR "Pedro Bay Village" OR Pee-Posh OR Penobscot OR Penutian OR Perquot OR "Pima People" OR Pima-Maricopa OR Poarch OR Pokagon OR Pomo OR Ponca OR "Portage Creek Village" OR Potawatomi OR Pottawatomi OR Pueblo OR Puyallup OR "Qagan Tayagungin" OR Qawalangin OR Quapaw OR Quechan OR "Rampart Village" OR Rappahannock OR Resighini OR "Sac & Fox" OR "Sac and Fox" OR "Saint George Island" OR "Saint Paul Island" OR Salish OR Samish OR Santee OR Sauk-Suiattle OR "Seldovia Village Tribe" OR Seminole OR Seneca OR Seneca-Cayuga OR Shinnecock OR Shivwits OR Shoshone OR Shoshone-Bannock OR Shoshone-Paiute OR Siuslaw OR Skagit OR "Skagway Village" OR Skokomish OR "South Naknek Village" OR "Stebbins Community Association" OR Stillaguamish OR Suquamish OR Swinomish OR "Takotna Village" OR Tangirnaq OR Tejon OR "Telida Village" OR Te-Moak OR Thlopthlocco OR "Tlingit & Haida" OR "Tohono O'odham" OR Tolowa OR Tonawanda OR Tonkawa OR Tulalip OR Tunica-Biloxi OR Tuolumne OR Tuscarora OR "Twin Hills Village" OR "Ugashik Village" OR Umatilla OR Umkumiut OR Umpqua OR Ute OR "Utu Utu Gwaitu" OR "Village of Alakanuk" OR "Village of Anaktuvuk Pass" OR "Village of Aniak" OR "Village of Atmautluak" OR "Village of Bill Moore's Slough" OR "Village of Chefornak" OR "Village of Clarks Point" OR "Village of Crooked Creek" OR "Village of Dot Lake" OR "Village of Iliamna" OR "Village of Kalskag" OR "Village of Kaltag" OR "Village of Kotlik" OR "Village of Lower Kalskag" OR "Village of Ohogamiut" OR "Village of Red Devil" OR "Village of Sleetmute" OR "Village of Solomon" OR "Village of Stony River" OR "Village of Venetie" OR "Village of Wainwright" OR Wailaki OR Wampanoag OR "Warm Springs Reservation" OR Washoe OR Willapa OR Winnemucca OR Wintun OR Wiyot OR "Wrangell Cooperative Association" OR Yakama OR Yakutat OR Yaqui OR Yavapai OR Yokuts OR Yuhaaviatam OR Yuima OR Yukian OR Yukut OR Yupiit OR Zuni) OR AK=("Asa'carsarmiut" OR "Dee-ni'" OR "Mi'kmaq" OR "S'Klallam" OR "Shoonaq'" OR "Sun'aq" OR (("Ivanof Bay" OR "Shoalwater Bay" OR "Spirit Lake" OR "Squaxin Island" OR Dakota OR Dakotah OR Wichita OR keechi OR waco OR Tawakonie OR Cheyenne OR Creek OR Crow OR Hoonah OR Kasigluk OR Maricopa OR Miami OR Omaha OR Oneida OR Ottawa OR Peoria OR Salamatof OR Sitka OR Snoqualmie OR Spokane OR Winnebago OR Wyandotte) NEAR/2 (Indian OR Indians OR Tribe OR Tribes OR native OR natives OR nation OR people*)) OR ((Native OR indian OR traditional) NEAR/2 (village OR council OR community OR association)) OR Absentee-Shawnee OR Achomawi OR Agdaagux OR "Agua Caliente" OR Ajumawi OR Ak-Chin OR Akiachak OR Akiak OR Alabama-Coushatta OR Alabama-Quassarte OR Alatna OR Algaaciq OR Algin OR Allakaket OR "Alturas Indian Rancheria" OR Alutiiq OR Angoon OR Anvik OR Apache OR Aporige OR Aquinnah OR arapaho OR Aroostook OR Assiniboine OR Astarawi OR Atsugewi OR Atwamsini OR Bdewakantunwan OR Caddo OR Cahto OR Cahuilla OR Catawba OR Cayuga OR Chalkyitsik OR Cheesh-Na OR Chehalis OR Chemehuevi OR Cher-Ae OR Chevak OR Cheyenne-Arapaho OR Chickahominy OR Chickaloon OR Chickasaw OR Chignik OR Chilkat OR Chilkoot OR Chinik OR Chinook OR Chippewa* OR Chitimacha OR Choctaw OR Chukchansi OR Chuloonawick OR Chumash OR Cocopah OR Colusa OR Comanche OR Coos OR Cosmit OR Coushatta OR Cowlitz OR "Cross Village" OR Cupeno OR Curyung OR Diegueno OR Dine' OR Egegik OR Eklutna OR Elwha OR "Emmonak Village" OR "Evansville Village" OR Ewiiaapaayp OR "Galena Village" OR Goshute OR "Gulkana Village Council" OR Haida OR Hammawi OR Havasupai OR "Healy Lake Village" OR Hewisedawi OR Ho-Chunk OR Hoh OR Hokan OR Holikachuk OR "Holy Cross Tribe" OR Hualapai OR "Hughes Village" OR "Huslia Village" OR "Hydaburg Cooperative Association" OR "Igiugig Village" OR Iipay OR Ilmawi OR Inaja OR Inupiat OR Ione OR Itsatawi OR "Kaguyak Village" OR Kaibab OR "Kaktovik Village" OR Kalispel OR Kanosh OR Karuk OR Kashia OR Kaw OR Keetoowah OR "Kenaitze Indian Tribe" OR "Ketchikan Indian Community" OR "Keweenaw Bay" OR Kialegee OR Kickapoo OR "King Island Native Community" OR "King Salmon Tribe" OR Kiowa OR Klamath OR "Klawock Cooperative Association" OR Kletsel OR "Knik Tribe" OR "Koi Nation" OR "Kokhanok Village" OR Koosharem OR Kootenai OR Kosalektawi OR "Koyukuk Native Village" OR Kumeyaay OR Kwalhioqua OR "Lesnoi Village" OR "Levelock Village" OR "Lime Village" OR "Louden Village" OR Luiseno OR Lummi OR Madesi OR Maidu OR Makah OR Maliseet OR "Manley Hot Springs Village" OR "Manokotak Village" OR Manzanita OR Mashantucket OR Match-e-be-nash-she-wish OR Mattaponi OR "McGrath Native Village" OR Mdewakanton OR Mechoopda OR Menominee OR Me-Wuk OR Miccosukee OR Micmacs OR "Mission Indians" OR Miwok OR Miwuk OR Modoc OR Mohegan OR Mojave OR Monacan OR "Mono Indians" OR Muckleshoot OR Muscogee OR "Na Dené" OR Nansemond OR Narragansett OR Navajo OR "New Koliganek Village" OR "New Stuyahok Village" OR "Newhalen Village" OR "Newtok Village" OR "Nez Perce" OR "Nikolai Village" OR "Ninilchik Village" OR Nisqually OR Nomlaki OR "Nondalton Village" OR Nooksack OR "Northway Village" OR Nottawaseppi OR "Nulato Village" OR Nunakauyarmiut OR Odawa OR Ohgsenakale OR "Ohkay Owingeh" OR Onondaga OR Oreilles OR "Organized Village of Grayling" OR "Organized Village of Kake" OR "Organized Village of Kasaan" OR "Organized Village of Kwethluk" OR "Organized Village of Saxman" OR Orutsararmiut OR Osage OR Otoe-Missouria OR Paiute OR Paiute-Shoshone OR Pala OR Pamunkey OR Pascua OR Paskenta OR Passamaquoddy OR "Pauloff Harbor Village" OR Pauma OR Pawnee OR Pechanga OR "Pechanga " OR "Peck Indian Reservation" OR "Pedro Bay Village" OR Pee-Posh OR Penobscot OR Penutian OR Perquot OR "Pima People" OR Pima-Maricopa OR Poarch OR Pokagon OR Pomo OR Ponca OR "Portage Creek Village" OR Potawatomi OR Pottawatomi OR Pueblo OR Puyallup OR "Qagan Tayagungin" OR Qawalangin OR Quapaw OR Quechan OR "Rampart Village" OR Rappahannock OR Resighini OR "Sac & Fox" OR "Sac and Fox" OR "Saint George Island" OR "Saint Paul Island" OR Salish OR Samish OR Santee OR Sauk-Suiattle OR "Seldovia Village Tribe" OR Seminole OR Seneca OR Seneca-Cayuga OR Shinnecock OR Shivwits OR Shoshone OR Shoshone-Bannock OR Shoshone-Paiute OR Siuslaw OR Skagit OR "Skagway Village" OR Skokomish OR "South Naknek Village" OR "Stebbins Community Association" OR Stillaguamish OR Suquamish OR Swinomish OR "Takotna Village" OR Tangirnaq OR Tejon OR "Telida Village" OR Te-Moak OR Thlopthlocco OR "Tlingit & Haida" OR "Tohono O'odham" OR Tolowa OR Tonawanda OR Tonkawa OR Tulalip OR Tunica-Biloxi OR Tuolumne OR Tuscarora OR "Twin Hills Village" OR "Ugashik Village" OR Umatilla OR Umkumiut OR Umpqua OR Ute OR "Utu Utu Gwaitu" OR "Village of Alakanuk" OR "Village of Anaktuvuk Pass" OR "Village of Aniak" OR "Village of Atmautluak" OR "Village of Bill Moore's Slough" OR "Village of Chefornak" OR "Village of Clarks Point" OR "Village of Crooked Creek" OR "Village of Dot Lake" OR "Village of Iliamna" OR "Village of Kalskag" OR "Village of Kaltag" OR "Village of Kotlik" OR "Village of Lower Kalskag" OR "Village of Ohogamiut" OR "Village of Red Devil" OR "Village of Sleetmute" OR "Village of Solomon" OR "Village of Stony River" OR "Village of Venetie" OR "Village of Wainwright" OR Wailaki OR Wampanoag OR "Warm Springs Reservation" OR Washoe OR Willapa OR Winnemucca OR Wintun OR Wiyot OR "Wrangell Cooperative Association" OR Yakama OR Yakutat OR Yaqui OR Yavapai OR Yokuts OR Yuhaaviatam OR Yuima OR Yukian OR Yukut OR Yupiit OR Zuni) OR KP=("Asa'carsarmiut" OR "Dee-ni'" OR "Mi'kmaq" OR "S'Klallam" OR "Shoonaq'" OR "Sun'aq" OR (("Ivanof Bay" OR "Shoalwater Bay" OR "Spirit Lake" OR "Squaxin Island" OR Dakota OR Dakotah OR Wichita OR keechi OR waco OR Tawakonie OR Cheyenne OR Creek OR Crow OR Hoonah OR Kasigluk OR Maricopa OR Miami OR Omaha OR Oneida OR Ottawa OR Peoria OR Salamatof OR Sitka OR Snoqualmie OR Spokane OR Winnebago OR Wyandotte) NEAR/2 (Indian OR Indians OR Tribe OR Tribes OR native OR natives OR nation OR people*)) OR ((Native OR indian OR traditional) NEAR/2 (village OR council OR community OR association)) OR Absentee-Shawnee OR Achomawi OR Agdaagux OR "Agua Caliente" OR Ajumawi OR Ak-Chin OR Akiachak OR Akiak OR Alabama-Coushatta OR Alabama-Quassarte OR Alatna OR Algaaciq OR Algin OR Allakaket OR "Alturas Indian Rancheria" OR Alutiiq OR Angoon OR Anvik OR Apache OR Aporige OR Aquinnah OR arapaho OR Aroostook OR Assiniboine OR Astarawi OR Atsugewi OR Atwamsini OR Bdewakantunwan OR Caddo OR Cahto OR Cahuilla OR Catawba OR Cayuga OR Chalkyitsik OR Cheesh-Na OR Chehalis OR Chemehuevi OR Cher-Ae OR Chevak OR Cheyenne-Arapaho OR Chickahominy OR Chickaloon OR Chickasaw OR Chignik OR Chilkat OR Chilkoot OR Chinik OR Chinook OR Chippewa* OR Chitimacha OR Choctaw OR Chukchansi OR Chuloonawick OR Chumash OR Cocopah OR Colusa OR Comanche OR Coos OR Cosmit OR Coushatta OR Cowlitz OR "Cross Village" OR Cupeno OR Curyung OR Diegueno OR Dine' OR Egegik OR Eklutna OR Elwha OR "Emmonak Village" OR "Evansville Village" OR Ewiiaapaayp OR "Galena Village" OR Goshute OR "Gulkana Village Council" OR Haida OR Hammawi OR Havasupai OR "Healy Lake Village" OR Hewisedawi OR Ho-Chunk OR Hoh OR Hokan OR Holikachuk OR "Holy Cross Tribe" OR Hualapai OR "Hughes Village" OR "Huslia Village" OR "Hydaburg Cooperative Association" OR "Igiugig Village" OR Iipay OR Ilmawi OR Inaja OR Inupiat OR Ione OR Itsatawi OR "Kaguyak Village" OR Kaibab OR "Kaktovik Village" OR Kalispel OR Kanosh OR Karuk OR Kashia OR Kaw OR Keetoowah OR "Kenaitze Indian Tribe" OR "Ketchikan Indian Community" OR "Keweenaw Bay" OR Kialegee OR Kickapoo OR "King Island Native Community" OR "King Salmon Tribe" OR Kiowa OR Klamath OR "Klawock Cooperative Association" OR Kletsel OR "Knik Tribe" OR "Koi Nation" OR "Kokhanok Village" OR Koosharem OR Kootenai OR Kosalektawi OR "Koyukuk Native Village" OR Kumeyaay OR Kwalhioqua OR "Lesnoi Village" OR "Levelock Village" OR "Lime Village" OR "Louden Village" OR Luiseno OR Lummi OR Madesi OR Maidu OR Makah OR Maliseet OR "Manley Hot Springs Village" OR "Manokotak Village" OR Manzanita OR Mashantucket OR Match-e-be-nash-she-wish OR Mattaponi OR "McGrath Native Village" OR Mdewakanton OR Mechoopda OR Menominee OR Me-Wuk OR Miccosukee OR Micmacs OR "Mission Indians" OR Miwok OR Miwuk OR Modoc OR Mohegan OR Mojave OR Monacan OR "Mono Indians" OR Muckleshoot OR Muscogee OR "Na Dené" OR Nansemond OR Narragansett OR Navajo OR "New Koliganek Village" OR "New Stuyahok Village" OR "Newhalen Village" OR "Newtok Village" OR "Nez Perce" OR "Nikolai Village" OR "Ninilchik Village" OR Nisqually OR Nomlaki OR "Nondalton Village" OR Nooksack OR "Northway Village" OR Nottawaseppi OR "Nulato Village" OR Nunakauyarmiut OR Odawa OR Ohgsenakale OR "Ohkay Owingeh" OR Onondaga OR Oreilles OR "Organized Village of Grayling" OR "Organized Village of Kake" OR "Organized Village of Kasaan" OR "Organized Village of Kwethluk" OR "Organized Village of Saxman" OR Orutsararmiut OR Osage OR Otoe-Missouria OR Paiute OR Paiute-Shoshone OR Pala OR Pamunkey OR Pascua OR Paskenta OR Passamaquoddy OR "Pauloff Harbor Village" OR Pauma OR Pawnee OR Pechanga OR "Pechanga " OR "Peck Indian Reservation" OR "Pedro Bay Village" OR Pee-Posh OR Penobscot OR Penutian OR Perquot OR "Pima People" OR Pima-Maricopa OR Poarch OR Pokagon OR Pomo OR Ponca OR "Portage Creek Village" OR Potawatomi OR Pottawatomi OR Pueblo OR Puyallup OR "Qagan Tayagungin" OR Qawalangin OR Quapaw OR Quechan OR "Rampart Village" OR Rappahannock OR Resighini OR "Sac & Fox" OR "Sac and Fox" OR "Saint George Island" OR "Saint Paul Island" OR Salish OR Samish OR Santee OR Sauk-Suiattle OR "Seldovia Village Tribe" OR Seminole OR Seneca OR Seneca-Cayuga OR Shinnecock OR Shivwits OR Shoshone OR Shoshone-Bannock OR Shoshone-Paiute OR Siuslaw OR Skagit OR "Skagway Village" OR Skokomish OR "South Naknek Village" OR "Stebbins Community Association" OR Stillaguamish OR Suquamish OR Swinomish OR "Takotna Village" OR Tangirnaq OR Tejon OR "Telida Village" OR Te-Moak OR Thlopthlocco OR "Tlingit & Haida" OR "Tohono O'odham" OR Tolowa OR Tonawanda OR Tonkawa OR Tulalip OR Tunica-Biloxi OR Tuolumne OR Tuscarora OR "Twin Hills Village" OR "Ugashik Village" OR Umatilla OR Umkumiut OR Umpqua OR Ute OR "Utu Utu Gwaitu" OR "Village of Alakanuk" OR "Village of Anaktuvuk Pass" OR "Village of Aniak" OR "Village of Atmautluak" OR "Village of Bill Moore's Slough" OR "Village of Chefornak" OR "Village of Clarks Point" OR "Village of Crooked Creek" OR "Village of Dot Lake" OR "Village of Iliamna" OR "Village of Kalskag" OR "Village of Kaltag" OR "Village of Kotlik" OR "Village of Lower Kalskag" OR "Village of Ohogamiut" OR "Village of Red Devil" OR "Village of Sleetmute" OR "Village of Solomon" OR "Village of Stony River" OR "Village of Venetie" OR "Village of Wainwright" OR Wailaki OR Wampanoag OR "Warm Springs Reservation" OR Washoe OR Willapa OR Winnemucca OR Wintun OR Wiyot OR "Wrangell Cooperative Association" OR Yakama OR Yakutat OR Yaqui OR Yavapai OR Yokuts OR Yuhaaviatam OR Yuima OR Yukian OR Yukut OR Yupiit OR Zuni) | 77643 | 86842 |
| 5 | TI=("Degexit'an" OR "Yup'ik" OR ("Bear River" NEAR/2 (Indian OR Indians OR Tribe OR Tribes OR native OR natives OR nation OR people*)) OR Abenaki OR Achumawi OR Ahtna OR "Akimel Oodham" OR "Akimel O'odham" OR "Akimel Ootham" OR "Akimel O'otham" OR Aleut* OR Algonkian* OR Alsea OR Aluutiq OR Apalachee OR Arikara OR Atakapa OR Athabascan OR Athabaskan OR Athapascan OR Athapaskan OR Bannock OR Bidai OR Biloxi OR Blackfeet OR Blackfoot OR Caddoan OR Calice OR Calusa OR Carrizo OR Cayuse OR Chemakum OR Cheraw OR Cherokee OR Chetco OR Cheyenne OR Chicora OR Chilula OR Chimariko OR Clatskanie OR Coahuiltecan OR "Coeur d'Alene" OR Costanoan OR Cree OR Croatan OR Croatoan OR Cusabo OR Edisto OR Eskaleut* OR Eskimo OR Eskimo-Aleut OR Esselen OR Eyak OR "Fox and Sauk" OR Gabrielino OR "Gros Ventre" OR Guale OR "Gwich'in" OR Halchidhoma OR Hidatsa OR Hitchiti OR Hopi OR Houma OR Huichol* OR Hupa OR Ingalik OR Inupiaq OR Ioway OR Iroquoi* OR Jacome OR Jano OR Jeaga OR Juaneno OR Jumano OR Kalapuya OR Kansa OR Karankawa OR Karok OR Kashaya OR Kato OR Kawaiisu OR Kitanemuk OR Kitsai OR Klallam OR Koasati OR Kolchan OR Konkow OR Koyukon OR Lakota* OR Lassik OR Laurentian OR Lenape OR "Lenni Lenape" OR Mahican OR Mandan OR Maricopa OR Massachusett OR Mattole OR Minisink OR Missouria OR Mohave OR Mohawk OR Mohican OR Molalla OR Multnomah OR Munsee OR Muscoge* OR Naabeeho OR Nakoda OR Nanticoke OR Natchez OR Nauset OR Navaho OR Niantic OR Nipmuc OR Nisenan OR Nongatl OR Oconee OR Ofo OR Ohlone OR Ojibw* OR Okanagan OR Oneida OR Otoe OR Palouse OR Panamint OR Papago OR Patwin OR "Pee Dee" OR Pennacook OR Pequot OR Pima OR Piro OR Pocasset OR Pocomtuc OR Pokanoket OR Poospatuck OR Powhatan OR Quechan OR Quileute OR Quinault OR Quiripi OR Salinan OR Samich OR Saponi OR Serrano OR Shasta OR Shawnee OR Siksikaitsitapi OR Sinkyone OR Siouan OR Sioux OR Snohomish OR Snoqualmie OR Squaxin OR Suma OR Susquehannock OR Takelma OR Tanacross OR Tanana OR Tataviam OR Tawakoni OR Tequesta OR Tillamook OR Timucua OR Tlingit OR "Tohono Oodham" OR Tongva OR Tsimshian OR Tubatulabal OR Tutelo OR Tututni OR Unalachtigo OR Unami OR Unangan OR Unangas OR Unkechaug OR Waccamaw OR Walapai OR Wappo OR Wasco OR Whilkut OR Wichita OR Winnebago OR Wintu OR Wishram OR Yahi OR Yakima OR Yamasee OR Yana OR Yaqu OR Yuchi OR Yuki OR Yuma OR Yupik OR Yurok) OR AB=("Degexit'an" OR "Yup'ik" OR ("Bear River" NEAR/2 (Indian OR Indians OR Tribe OR Tribes OR native OR natives OR nation OR people*)) OR Abenaki OR Achumawi OR Ahtna OR "Akimel Oodham" OR "Akimel O'odham" OR "Akimel Ootham" OR "Akimel O'otham" OR Aleut* OR Algonkian* OR Alsea OR Aluutiq OR Apalachee OR Arikara OR Atakapa OR Athabascan OR Athabaskan OR Athapascan OR Athapaskan OR Bannock OR Bidai OR Biloxi OR Blackfeet OR Blackfoot OR Caddoan OR Calice OR Calusa OR Carrizo OR Cayuse OR Chemakum OR Cheraw OR Cherokee OR Chetco OR Cheyenne OR Chicora OR Chilula OR Chimariko OR Clatskanie OR Coahuiltecan OR "Coeur d'Alene" OR Costanoan OR Cree OR Croatan OR Croatoan OR Cusabo OR Edisto OR Eskaleut* OR Eskimo OR Eskimo-Aleut OR Esselen OR Eyak OR "Fox and Sauk" OR Gabrielino OR "Gros Ventre" OR Guale OR "Gwich'in" OR Halchidhoma OR Hidatsa OR Hitchiti OR Hopi OR Houma OR Huichol* OR Hupa OR Ingalik OR Inupiaq OR Ioway OR Iroquoi* OR Jacome OR Jano OR Jeaga OR Juaneno OR Jumano OR Kalapuya OR Kansa OR Karankawa OR Karok OR Kashaya OR Kato OR Kawaiisu OR Kitanemuk OR Kitsai OR Klallam OR Koasati OR Kolchan OR Konkow OR Koyukon OR Lakota* OR Lassik OR Laurentian OR Lenape OR "Lenni Lenape" OR Mahican OR Mandan OR Maricopa OR Massachusett OR Mattole OR Minisink OR Missouria OR Mohave OR Mohawk OR Mohican OR Molalla OR Multnomah OR Munsee OR Muscoge* OR Naabeeho OR Nakoda OR Nanticoke OR Natchez OR Nauset OR Navaho OR Niantic OR Nipmuc OR Nisenan OR Nongatl OR Oconee OR Ofo OR Ohlone OR Ojibw* OR Okanagan OR Oneida OR Otoe OR Palouse OR Panamint OR Papago OR Patwin OR "Pee Dee" OR Pennacook OR Pequot OR Pima OR Piro OR Pocasset OR Pocomtuc OR Pokanoket OR Poospatuck OR Powhatan OR Quechan OR Quileute OR Quinault OR Quiripi OR Salinan OR Samich OR Saponi OR Serrano OR Shasta OR Shawnee OR Siksikaitsitapi OR Sinkyone OR Siouan OR Sioux OR Snohomish OR Snoqualmie OR Squaxin OR Suma OR Susquehannock OR Takelma OR Tanacross OR Tanana OR Tataviam OR Tawakoni OR Tequesta OR Tillamook OR Timucua OR Tlingit OR "Tohono Oodham" OR Tongva OR Tsimshian OR Tubatulabal OR Tutelo OR Tututni OR Unalachtigo OR Unami OR Unangan OR Unangas OR Unkechaug OR Waccamaw OR Walapai OR Wappo OR Wasco OR Whilkut OR Wichita OR Winnebago OR Wintu OR Wishram OR Yahi OR Yakima OR Yamasee OR Yana OR Yaqu OR Yuchi OR Yuki OR Yuma OR Yupik OR Yurok) OR AK=("Degexit'an" OR "Yup'ik" OR ("Bear River" NEAR/2 (Indian OR Indians OR Tribe OR Tribes OR native OR natives OR nation OR people*)) OR Abenaki OR Achumawi OR Ahtna OR "Akimel Oodham" OR "Akimel O'odham" OR "Akimel Ootham" OR "Akimel O'otham" OR Aleut* OR Algonkian* OR Alsea OR Aluutiq OR Apalachee OR Arikara OR Atakapa OR Athabascan OR Athabaskan OR Athapascan OR Athapaskan OR Bannock OR Bidai OR Biloxi OR Blackfeet OR Blackfoot OR Caddoan OR Calice OR Calusa OR Carrizo OR Cayuse OR Chemakum OR Cheraw OR Cherokee OR Chetco OR Cheyenne OR Chicora OR Chilula OR Chimariko OR Clatskanie OR Coahuiltecan OR "Coeur d'Alene" OR Costanoan OR Cree OR Croatan OR Croatoan OR Cusabo OR Edisto OR Eskaleut* OR Eskimo OR Eskimo-Aleut OR Esselen OR Eyak OR "Fox and Sauk" OR Gabrielino OR "Gros Ventre" OR Guale OR "Gwich'in" OR Halchidhoma OR Hidatsa OR Hitchiti OR Hopi OR Houma OR Huichol* OR Hupa OR Ingalik OR Inupiaq OR Ioway OR Iroquoi* OR Jacome OR Jano OR Jeaga OR Juaneno OR Jumano OR Kalapuya OR Kansa OR Karankawa OR Karok OR Kashaya OR Kato OR Kawaiisu OR Kitanemuk OR Kitsai OR Klallam OR Koasati OR Kolchan OR Konkow OR Koyukon OR Lakota* OR Lassik OR Laurentian OR Lenape OR "Lenni Lenape" OR Mahican OR Mandan OR Maricopa OR Massachusett OR Mattole OR Minisink OR Missouria OR Mohave OR Mohawk OR Mohican OR Molalla OR Multnomah OR Munsee OR Muscoge* OR Naabeeho OR Nakoda OR Nanticoke OR Natchez OR Nauset OR Navaho OR Niantic OR Nipmuc OR Nisenan OR Nongatl OR Oconee OR Ofo OR Ohlone OR Ojibw* OR Okanagan OR Oneida OR Otoe OR Palouse OR Panamint OR Papago OR Patwin OR "Pee Dee" OR Pennacook OR Pequot OR Pima OR Piro OR Pocasset OR Pocomtuc OR Pokanoket OR Poospatuck OR Powhatan OR Quechan OR Quileute OR Quinault OR Quiripi OR Salinan OR Samich OR Saponi OR Serrano OR Shasta OR Shawnee OR Siksikaitsitapi OR Sinkyone OR Siouan OR Sioux OR Snohomish OR Snoqualmie OR Squaxin OR Suma OR Susquehannock OR Takelma OR Tanacross OR Tanana OR Tataviam OR Tawakoni OR Tequesta OR Tillamook OR Timucua OR Tlingit OR "Tohono Oodham" OR Tongva OR Tsimshian OR Tubatulabal OR Tutelo OR Tututni OR Unalachtigo OR Unami OR Unangan OR Unangas OR Unkechaug OR Waccamaw OR Walapai OR Wappo OR Wasco OR Whilkut OR Wichita OR Winnebago OR Wintu OR Wishram OR Yahi OR Yakima OR Yamasee OR Yana OR Yaqu OR Yuchi OR Yuki OR Yuma OR Yupik OR Yurok) OR KP=("Degexit'an" OR "Yup'ik" OR ("Bear River" NEAR/2 (Indian OR Indians OR Tribe OR Tribes OR native OR natives OR nation OR people*)) OR Abenaki OR Achumawi OR Ahtna OR "Akimel Oodham" OR "Akimel O'odham" OR "Akimel Ootham" OR "Akimel O'otham" OR Aleut* OR Algonkian* OR Alsea OR Aluutiq OR Apalachee OR Arikara OR Atakapa OR Athabascan OR Athabaskan OR Athapascan OR Athapaskan OR Bannock OR Bidai OR Biloxi OR Blackfeet OR Blackfoot OR Caddoan OR Calice OR Calusa OR Carrizo OR Cayuse OR Chemakum OR Cheraw OR Cherokee OR Chetco OR Cheyenne OR Chicora OR Chilula OR Chimariko OR Clatskanie OR Coahuiltecan OR "Coeur d'Alene" OR Costanoan OR Cree OR Croatan OR Croatoan OR Cusabo OR Edisto OR Eskaleut* OR Eskimo OR Eskimo-Aleut OR Esselen OR Eyak OR "Fox and Sauk" OR Gabrielino OR "Gros Ventre" OR Guale OR "Gwich'in" OR Halchidhoma OR Hidatsa OR Hitchiti OR Hopi OR Houma OR Huichol* OR Hupa OR Ingalik OR Inupiaq OR Ioway OR Iroquoi* OR Jacome OR Jano OR Jeaga OR Juaneno OR Jumano OR Kalapuya OR Kansa OR Karankawa OR Karok OR Kashaya OR Kato OR Kawaiisu OR Kitanemuk OR Kitsai OR Klallam OR Koasati OR Kolchan OR Konkow OR Koyukon OR Lakota* OR Lassik OR Laurentian OR Lenape OR "Lenni Lenape" OR Mahican OR Mandan OR Maricopa OR Massachusett OR Mattole OR Minisink OR Missouria OR Mohave OR Mohawk OR Mohican OR Molalla OR Multnomah OR Munsee OR Muscoge* OR Naabeeho OR Nakoda OR Nanticoke OR Natchez OR Nauset OR Navaho OR Niantic OR Nipmuc OR Nisenan OR Nongatl OR Oconee OR Ofo OR Ohlone OR Ojibw* OR Okanagan OR Oneida OR Otoe OR Palouse OR Panamint OR Papago OR Patwin OR "Pee Dee" OR Pennacook OR Pequot OR Pima OR Piro OR Pocasset OR Pocomtuc OR Pokanoket OR Poospatuck OR Powhatan OR Quechan OR Quileute OR Quinault OR Quiripi OR Salinan OR Samich OR Saponi OR Serrano OR Shasta OR Shawnee OR Siksikaitsitapi OR Sinkyone OR Siouan OR Sioux OR Snohomish OR Snoqualmie OR Squaxin OR Suma OR Susquehannock OR Takelma OR Tanacross OR Tanana OR Tataviam OR Tawakoni OR Tequesta OR Tillamook OR Timucua OR Tlingit OR "Tohono Oodham" OR Tongva OR Tsimshian OR Tubatulabal OR Tutelo OR Tututni OR Unalachtigo OR Unami OR Unangan OR Unangas OR Unkechaug OR Waccamaw OR Walapai OR Wappo OR Wasco OR Whilkut OR Wichita OR Winnebago OR Wintu OR Wishram OR Yahi OR Yakima OR Yamasee OR Yana OR Yaqu OR Yuchi OR Yuki OR Yuma OR Yupik OR Yurok) | 48030 | 52749 |
| 6 | TI=(Anishnawbek OR "?Akisq'nuk" OR "?aq'am" OR "?Esdilagh" OR "Abénakis" OR "Akisq'nuk" OR "aq'am" OR "Che:k:tles7et'h'" OR "Da'naxda'xw" OR "Dzawada'enuxw" OR "Esk'etemc" OR "Gitga'at" OR "Gitlaxt'aamiks" OR "Gwa'Sala-Nakwaxda'xw" OR "Gwich'in" OR "Ka:'yu:'k't'h'" OR "Kanyen'kehaka" OR "Kapawe'no" OR "K'atlodeeche" OR "K'ómoks" OR "Leq'á:mel" OR "Lil'wat" OR "Malécite" OR "Mi'kmaq" OR "Mowachaht/Muchalaht" OR "Nisga'a" OR "N'Quatqua" OR "O'Chiese" OR "Ononda'gega'" OR "Onondowága'" OR "Onyota'a:ka" OR "Saik'uz" OR "Sambaa K'e " OR "Shxwhá:y" OR "Shxw'ow'hamel" OR "SIPEKNE'KATIK" OR "Skaru:reh" OR "Sq'éwlets" OR "Stellat'en" OR "Sts'ailes" OR "Stz'uminus" OR "T'it'q'et" OR "Tla'amin" OR "Tl'azt'en" OR "Tl'etinqox" OR "Tsal'alh" OR "Tsideldel" OR "Ts'kw'aylaxw" OR "T'Sou-ke" OR "Ts'uubaa-asatx" OR "Tsuut'ina" OR "We'koqma'q" OR "Wet'suwet'en" OR "Xai'xais" OR "Xaxli'p" OR "Yunesit'in" OR ((Acadia OR Carmacks OR Champagne OR Creek OR Delaware OR Douglas OR Montana OR Similkameen) NEAR/2 (Indian OR Indians OR Tribe OR Tribes OR native OR natives OR nation OR people*)) OR Aamjiwnaang OR Abegweit OR Abitibiwinni OR Ahousaht OR Ahtahkakoop OR Aishihik OR Aitchelitz OR Aklavik OR Algonquin* OR Anicinape* OR "Animakee Wa Zhing" OR Anishina* OR aqam OR Atikamekw OR Atjanasca* OR Attawapiskat OR Aundeck-Omni-Kaning OR Batchewana OR "Bearfoot Onondaga" OR Beausoleil OR "Behdzi Ahda*" OR "Biigtigong Nishnaabeg" OR "Biinjitiwaabik Zaaging Anishinaabek" OR Bilijk OR "Binche Whut'en" OR "Bingwi Neyaashi Anishinaabek" OR "Canupawakpa Dakota" OR Cayuga OR Chawathil OR Cheam OR "Cheslatta Carrier" OR Chiniki OR Chipew* OR Cote OR Couchiching OR Cowessess OR Cowichan OR Cree OR "Dakota Plains" OR "Dakota Tipi" OR "Dechi Laot'i" OR Deline OR Dena OR Dene OR Denesuline OR "Deninu K'ue" OR Ditidaht OR "Dog Rib Rae" OR Dokis OR Eabametoong OR Ehattesaht OR Elsipogtog OR Ermineskin OR Esdilagh OR Esgenoopetitj OR Eskasoni OR Esquimalt OR Gameti OR Gayogohono OR Ginoogaming OR Gitanmaax OR Gitanyow OR Gitsegukla OR Gitwangak OR Gitxaala OR Glooscap OR Gwawaenuk OR "Gwichya Gwich'in" OR Gwitchin OR Hagwilget OR Haisla OR Halalt OR Haudenosaunee OR Heiltsuk OR Hesquiaht OR Hiawatha OR Homalco OR Hupacasath OR Huu-ay-aht OR Inninuwug OR Innu OR Innue OR Innus OR Inuvik OR Iskatewizaagegan OR Iskut OR "Ka'a'gee Tu" OR Kahkewistahaw OR "Kasabonika Lake" OR Kashechewan OR Katzie OR Kawacatoose OR Kebaowek OR Keeseekoose OR Keeseekoowenin OR Kee-Way-Win OR "Kinistin Saulteaux" OR Kinonjeoshtegon OR Kispiox OR Kitselas OR Kitsumkalum OR Kluane OR Kutchin OR Kwadacha OR Kwakiutl OR "Kwanlin Dun" OR Kwaw-kwaw-Apilt OR Kwiakah OR "Kwikwasut'inuxw Haxwa'mis" OR Kwikwetlem OR "Lax Kw'alaams" OR "Lheidli T'enneh" OR "Liidlii Kue" OR "Listuguj Mi'gmaq" OR Lyackson OR Magnetawan OR Malahat OR Maliseet OR Mamalilikulla OR Matachewan OR Matsqui OR Mattagami OR "M'Chigeeng" OR Membertou OR Metepenagiag OR Metlakatla OR Miawpukek OR Michipicoten OR Micmacs OR Millbrook OR Mishkeegogamang OR Mississauga* OR Mitaanjigamiing OR Mohawk* OR Montagnais OR Moosomin OR Moravian OR Mowachaht OR Muchalaht OR Munsee-Delaware OR Muscowpetung OR Muskoday OR Muskowekwan OR Musqueam OR "Nacho Nyak Dun" OR "Nadleh Whuten" OR Naicatchewenin OR "Nak'azdli Whut'en" OR Nakota OR Naotkamegwanning OR Naskapi OR Natoaganeg OR Nee-Tahi-Buhn OR Nehiyawak OR Nekaneet OR Neskantaga OR Neskonlith OR Nibinamik OR Nigigoonsiminikaaning OR Nipissing OR Nishnaabeg OR Nooaitch OR Nuchatlaht OR Nuxalk OR Ochapowace OR O-Chi-Chak-Ko-Sipi OR Odanak OR Ojibw* OR Okanagan OR Okanese OR Okemasis OR Oneida OR Oromocto OR Osoyoos OR Pabineau OR Pacheedaht OR "Paqtnkek Mi'kmaw" OR Pasqua OR Pauingassi OR Pauquachin OR "Pays Plat" OR Peguis OR "Pehdzeh Ki" OR Pekuakamiulnuatsh OR Penelakut OR Piapot OR Piikani OR Pikangikum OR Pinaymootang OR Potlotek OR "Qalipu Mi'kmaq" OR Qualicum OR Quatsino OR Sagamok OR Sahgaiehcan OR Samahquam OR Saugeen OR Saulteau OR Saulteaux OR Sechelt OR Secwepemc* OR Semiahmoo OR Seneca OR Shackan OR Shamattawa OR Shawanaga OR Sheguiandah OR Sheshegwaning OR Shuswap OR Siksika OR Simpcw OR Sioux OR Siska OR Skatin OR Skawahlook OR Skeetchestn OR Skidegate OR "Skin Tyee" OR Skowkale OR Skownan OR Skuppah OR Skwah OR Snuneymuxw OR Songhees OR Soowahlie OR Splatsin OR Spuzzum OR Squamish OR Squiala OR "Stswecem'c Xget'tem" OR Sumas OR "Ta'an Kwach'an" OR Tagish OR Tahltan OR Takla OR Tallcree OR "Taykwa Tagamou" OR "Teetl'it Gwich'in" OR Temagami OR Thessalon OR Timiskaming OR "Tk'emlúps te Secwépemc" OR Tla-o-qui-aht OR Tlatlasikwala OR Tlingit OR Tlowitsis OR Tootinaowaziibeeng OR Toquaht OR "Tr'ondëk Hwëch'in" OR Tsartlip OR Tsawout OR Tsawwassen OR Tseshaht OR Tseycum OR "Ts'il Kaz Koh" OR Tsleil-Waututh OR Tuscarora OR Tutchone OR Tzeachten OR Uchucklesaht OR Ucluelet OR Ulkatcho OR Wabaseemoong OR Wabauskang OR Wagmatcook OR Wahnapitae OR "Wahpeton Dakota" OR Wahsipekuk OR Wapekeka OR Wasagamack OR Wasauksing OR Waswanipi OR Wawakapewin OR Waywayseecappo OR Webequie OR Weenusk OR Wemotaci OR Wendat OR "Wha Ti" OR Whapmagoostui OR "Whitecap Dakota" OR Wikwemikong OR Witset OR Wolastoqiyik OR Wuikinuxv OR Wunnumin OR "Wuskwi Sipihk" OR Xatsull OR "Xeni Gwet'in" OR Yakweakwioose OR Yekooche OR "Yellow Quill" OR Yellowknife OR Yellowknives OR Zhiibaahaasing) OR AB=(Anishnawbek OR "Akisq'nuk" OR "aq'am" OR "Esdilagh" OR "Abénakis" OR "Akisq'nuk" OR "aq'am" OR "Che:k:tles7et'h'" OR "Da'naxda'xw" OR "Dzawada'enuxw" OR "Esk'etemc" OR "Gitga'at" OR "Gitlaxt'aamiks" OR "Gwa'Sala-Nakwaxda'xw" OR "Gwich'in" OR "Ka:'yu:'k't'h'" OR "Kanyen'kehaka" OR "Kapawe'no" OR "K'atlodeeche" OR "K'ómoks" OR "Leq'á:mel" OR "Lil'wat" OR "Malécite" OR "Mi'kmaq" OR "Mowachaht/Muchalaht" OR "Nisga'a" OR "N'Quatqua" OR "O'Chiese" OR "Ononda'gega'" OR "Onondowága'" OR "Onyota'a:ka" OR "Saik'uz" OR "Sambaa K'e " OR "Shxwhá:y" OR "Shxw'ow'hamel" OR "SIPEKNE'KATIK" OR "Skaru:reh" OR "Sq'éwlets" OR "Stellat'en" OR "Sts'ailes" OR "Stz'uminus" OR "T'it'q'et" OR "Tla'amin" OR "Tl'azt'en" OR "Tl'etinqox" OR "Tsal'alh" OR "Tsideldel" OR "Ts'kw'aylaxw" OR "T'Sou-ke" OR "Ts'uubaa-asatx" OR "Tsuut'ina" OR "We'koqma'q" OR "Wet'suwet'en" OR "Xai'xais" OR "Xaxli'p" OR "Yunesit'in" OR ((Acadia OR Carmacks OR Champagne OR Creek OR Delaware OR Douglas OR Montana OR Similkameen) NEAR/2 (Indian OR Indians OR Tribe OR Tribes OR native OR natives OR nation OR people*)) OR Aamjiwnaang OR Abegweit OR Abitibiwinni OR Ahousaht OR Ahtahkakoop OR Aishihik OR Aitchelitz OR Aklavik OR Algonquin* OR Anicinape* OR "Animakee Wa Zhing" OR Anishina* OR aqam OR Atikamekw OR Atjanasca* OR Attawapiskat OR Aundeck-Omni-Kaning OR Batchewana OR "Bearfoot Onondaga" OR Beausoleil OR "Behdzi Ahda*" OR "Biigtigong Nishnaabeg" OR "Biinjitiwaabik Zaaging Anishinaabek" OR Bilijk OR "Binche Whut'en" OR "Bingwi Neyaashi Anishinaabek" OR "Canupawakpa Dakota" OR Cayuga OR Chawathil OR Cheam OR "Cheslatta Carrier" OR Chiniki OR Chipew* OR Cote OR Couchiching OR Cowessess OR Cowichan OR Cree OR "Dakota Plains" OR "Dakota Tipi" OR "Dechi Laot'i" OR Deline OR Dena OR Dene OR Denesuline OR "Deninu K'ue" OR Ditidaht OR "Dog Rib Rae" OR Dokis OR Eabametoong OR Ehattesaht OR Elsipogtog OR Ermineskin OR Esdilagh OR Esgenoopetitj OR Eskasoni OR Esquimalt OR Gameti OR Gayogohono OR Ginoogaming OR Gitanmaax OR Gitanyow OR Gitsegukla OR Gitwangak OR Gitxaala OR Glooscap OR Gwawaenuk OR "Gwichya Gwich'in" OR Gwitchin OR Hagwilget OR Haisla OR Halalt OR Haudenosaunee OR Heiltsuk OR Hesquiaht OR Hiawatha OR Homalco OR Hupacasath OR Huu-ay-aht OR Inninuwug OR Innu OR Innue OR Innus OR Inuvik OR Iskatewizaagegan OR Iskut OR "Ka'a'gee Tu" OR Kahkewistahaw OR "Kasabonika Lake" OR Kashechewan OR Katzie OR Kawacatoose OR Kebaowek OR Keeseekoose OR Keeseekoowenin OR Kee-Way-Win OR "Kinistin Saulteaux" OR Kinonjeoshtegon OR Kispiox OR Kitselas OR Kitsumkalum OR Kluane OR Kutchin OR Kwadacha OR Kwakiutl OR "Kwanlin Dun" OR Kwaw-kwaw-Apilt OR Kwiakah OR "Kwikwasut'inuxw Haxwa'mis" OR Kwikwetlem OR "Lax Kw'alaams" OR "Lheidli T'enneh" OR "Liidlii Kue" OR "Listuguj Mi'gmaq" OR Lyackson OR Magnetawan OR Malahat OR Maliseet OR Mamalilikulla OR Matachewan OR Matsqui OR Mattagami OR "M'Chigeeng" OR Membertou OR Metepenagiag OR Metlakatla OR Miawpukek OR Michipicoten OR Micmacs OR Millbrook OR Mishkeegogamang OR Mississauga* OR Mitaanjigamiing OR Mohawk* OR Montagnais OR Moosomin OR Moravian OR Mowachaht OR Muchalaht OR Munsee-Delaware OR Muscowpetung OR Muskoday OR Muskowekwan OR Musqueam OR "Nacho Nyak Dun" OR "Nadleh Whuten" OR Naicatchewenin OR "Nak'azdli Whut'en" OR Nakota OR Naotkamegwanning OR Naskapi OR Natoaganeg OR Nee-Tahi-Buhn OR Nehiyawak OR Nekaneet OR Neskantaga OR Neskonlith OR Nibinamik OR Nigigoonsiminikaaning OR Nipissing OR Nishnaabeg OR Nooaitch OR Nuchatlaht OR Nuxalk OR Ochapowace OR O-Chi-Chak-Ko-Sipi OR Odanak OR Ojibw* OR Okanagan OR Okanese OR Okemasis OR Oneida OR Oromocto OR Osoyoos OR Pabineau OR Pacheedaht OR "Paqtnkek Mi'kmaw" OR Pasqua OR Pauingassi OR Pauquachin OR "Pays Plat" OR Peguis OR "Pehdzeh Ki" OR Pekuakamiulnuatsh OR Penelakut OR Piapot OR Piikani OR Pikangikum OR Pinaymootang OR Potlotek OR "Qalipu Mi'kmaq" OR Qualicum OR Quatsino OR Sagamok OR Sahgaiehcan OR Samahquam OR Saugeen OR Saulteau OR Saulteaux OR Sechelt OR Secwepemc* OR Semiahmoo OR Seneca OR Shackan OR Shamattawa OR Shawanaga OR Sheguiandah OR Sheshegwaning OR Shuswap OR Siksika OR Simpcw OR Sioux OR Siska OR Skatin OR Skawahlook OR Skeetchestn OR Skidegate OR "Skin Tyee" OR Skowkale OR Skownan OR Skuppah OR Skwah OR Snuneymuxw OR Songhees OR Soowahlie OR Splatsin OR Spuzzum OR Squamish OR Squiala OR "Stswecem'c Xget'tem" OR Sumas OR "Ta'an Kwach'an" OR Tagish OR Tahltan OR Takla OR Tallcree OR "Taykwa Tagamou" OR "Teetl'it Gwich'in" OR Temagami OR Thessalon OR Timiskaming OR "Tk'emlúps te Secwépemc" OR Tla-o-qui-aht OR Tlatlasikwala OR Tlingit OR Tlowitsis OR Tootinaowaziibeeng OR Toquaht OR "Tr'ondëk Hwëch'in" OR Tsartlip OR Tsawout OR Tsawwassen OR Tseshaht OR Tseycum OR "Ts'il Kaz Koh" OR Tsleil-Waututh OR Tuscarora OR Tutchone OR Tzeachten OR Uchucklesaht OR Ucluelet OR Ulkatcho OR Wabaseemoong OR Wabauskang OR Wagmatcook OR Wahnapitae OR "Wahpeton Dakota" OR Wahsipekuk OR Wapekeka OR Wasagamack OR Wasauksing OR Waswanipi OR Wawakapewin OR Waywayseecappo OR Webequie OR Weenusk OR Wemotaci OR Wendat OR "Wha Ti" OR Whapmagoostui OR "Whitecap Dakota" OR Wikwemikong OR Witset OR Wolastoqiyik OR Wuikinuxv OR Wunnumin OR "Wuskwi Sipihk" OR Xatsull OR "Xeni Gwet'in" OR Yakweakwioose OR Yekooche OR "Yellow Quill" OR Yellowknife OR Yellowknives OR Zhiibaahaasing) OR AK=(Anishnawbek OR "Akisq'nuk" OR "aq'am" OR "Esdilagh" OR "Abénakis" OR "Akisq'nuk" OR "aq'am" OR "Che:k:tles7et'h'" OR "Da'naxda'xw" OR "Dzawada'enuxw" OR "Esk'etemc" OR "Gitga'at" OR "Gitlaxt'aamiks" OR "Gwa'Sala-Nakwaxda'xw" OR "Gwich'in" OR "Ka:'yu:'k't'h'" OR "Kanyen'kehaka" OR "Kapawe'no" OR "K'atlodeeche" OR "K'ómoks" OR "Leq'á:mel" OR "Lil'wat" OR "Malécite" OR "Mi'kmaq" OR "Mowachaht/Muchalaht" OR "Nisga'a" OR "N'Quatqua" OR "O'Chiese" OR "Ononda'gega'" OR "Onondowága'" OR "Onyota'a:ka" OR "Saik'uz" OR "Sambaa K'e " OR "Shxwhá:y" OR "Shxw'ow'hamel" OR "SIPEKNE'KATIK" OR "Skaru:reh" OR "Sq'éwlets" OR "Stellat'en" OR "Sts'ailes" OR "Stz'uminus" OR "T'it'q'et" OR "Tla'amin" OR "Tl'azt'en" OR "Tl'etinqox" OR "Tsal'alh" OR "Tsideldel" OR "Ts'kw'aylaxw" OR "T'Sou-ke" OR "Ts'uubaa-asatx" OR "Tsuut'ina" OR "We'koqma'q" OR "Wet'suwet'en" OR "Xai'xais" OR "Xaxli'p" OR "Yunesit'in" OR ((Acadia OR Carmacks OR Champagne OR Creek OR Delaware OR Douglas OR Montana OR Similkameen) NEAR/2 (Indian OR Indians OR Tribe OR Tribes OR native OR natives OR nation OR people*)) OR Aamjiwnaang OR Abegweit OR Abitibiwinni OR Ahousaht OR Ahtahkakoop OR Aishihik OR Aitchelitz OR Aklavik OR Algonquin* OR Anicinape* OR "Animakee Wa Zhing" OR Anishina* OR aqam OR Atikamekw OR Atjanasca* OR Attawapiskat OR Aundeck-Omni-Kaning OR Batchewana OR "Bearfoot Onondaga" OR Beausoleil OR "Behdzi Ahda*" OR "Biigtigong Nishnaabeg" OR "Biinjitiwaabik Zaaging Anishinaabek" OR Bilijk OR "Binche Whut'en" OR "Bingwi Neyaashi Anishinaabek" OR "Canupawakpa Dakota" OR Cayuga OR Chawathil OR Cheam OR "Cheslatta Carrier" OR Chiniki OR Chipew* OR Cote OR Couchiching OR Cowessess OR Cowichan OR Cree OR "Dakota Plains" OR "Dakota Tipi" OR "Dechi Laot'i" OR Deline OR Dena OR Dene OR Denesuline OR "Deninu K'ue" OR Ditidaht OR "Dog Rib Rae" OR Dokis OR Eabametoong OR Ehattesaht OR Elsipogtog OR Ermineskin OR Esdilagh OR Esgenoopetitj OR Eskasoni OR Esquimalt OR Gameti OR Gayogohono OR Ginoogaming OR Gitanmaax OR Gitanyow OR Gitsegukla OR Gitwangak OR Gitxaala OR Glooscap OR Gwawaenuk OR "Gwichya Gwich'in" OR Gwitchin OR Hagwilget OR Haisla OR Halalt OR Haudenosaunee OR Heiltsuk OR Hesquiaht OR Hiawatha OR Homalco OR Hupacasath OR Huu-ay-aht OR Inninuwug OR Innu OR Innue OR Innus OR Inuvik OR Iskatewizaagegan OR Iskut OR "Ka'a'gee Tu" OR Kahkewistahaw OR "Kasabonika Lake" OR Kashechewan OR Katzie OR Kawacatoose OR Kebaowek OR Keeseekoose OR Keeseekoowenin OR Kee-Way-Win OR "Kinistin Saulteaux" OR Kinonjeoshtegon OR Kispiox OR Kitselas OR Kitsumkalum OR Kluane OR Kutchin OR Kwadacha OR Kwakiutl OR "Kwanlin Dun" OR Kwaw-kwaw-Apilt OR Kwiakah OR "Kwikwasut'inuxw Haxwa'mis" OR Kwikwetlem OR "Lax Kw'alaams" OR "Lheidli T'enneh" OR "Liidlii Kue" OR "Listuguj Mi'gmaq" OR Lyackson OR Magnetawan OR Malahat OR Maliseet OR Mamalilikulla OR Matachewan OR Matsqui OR Mattagami OR "M'Chigeeng" OR Membertou OR Metepenagiag OR Metlakatla OR Miawpukek OR Michipicoten OR Micmacs OR Millbrook OR Mishkeegogamang OR Mississauga* OR Mitaanjigamiing OR Mohawk* OR Montagnais OR Moosomin OR Moravian OR Mowachaht OR Muchalaht OR Munsee-Delaware OR Muscowpetung OR Muskoday OR Muskowekwan OR Musqueam OR "Nacho Nyak Dun" OR "Nadleh Whuten" OR Naicatchewenin OR "Nak'azdli Whut'en" OR Nakota OR Naotkamegwanning OR Naskapi OR Natoaganeg OR Nee-Tahi-Buhn OR Nehiyawak OR Nekaneet OR Neskantaga OR Neskonlith OR Nibinamik OR Nigigoonsiminikaaning OR Nipissing OR Nishnaabeg OR Nooaitch OR Nuchatlaht OR Nuxalk OR Ochapowace OR O-Chi-Chak-Ko-Sipi OR Odanak OR Ojibw* OR Okanagan OR Okanese OR Okemasis OR Oneida OR Oromocto OR Osoyoos OR Pabineau OR Pacheedaht OR "Paqtnkek Mi'kmaw" OR Pasqua OR Pauingassi OR Pauquachin OR "Pays Plat" OR Peguis OR "Pehdzeh Ki" OR Pekuakamiulnuatsh OR Penelakut OR Piapot OR Piikani OR Pikangikum OR Pinaymootang OR Potlotek OR "Qalipu Mi'kmaq" OR Qualicum OR Quatsino OR Sagamok OR Sahgaiehcan OR Samahquam OR Saugeen OR Saulteau OR Saulteaux OR Sechelt OR Secwepemc* OR Semiahmoo OR Seneca OR Shackan OR Shamattawa OR Shawanaga OR Sheguiandah OR Sheshegwaning OR Shuswap OR Siksika OR Simpcw OR Sioux OR Siska OR Skatin OR Skawahlook OR Skeetchestn OR Skidegate OR "Skin Tyee" OR Skowkale OR Skownan OR Skuppah OR Skwah OR Snuneymuxw OR Songhees OR Soowahlie OR Splatsin OR Spuzzum OR Squamish OR Squiala OR "Stswecem'c Xget'tem" OR Sumas OR "Ta'an Kwach'an" OR Tagish OR Tahltan OR Takla OR Tallcree OR "Taykwa Tagamou" OR "Teetl'it Gwich'in" OR Temagami OR Thessalon OR Timiskaming OR "Tk'emlúps te Secwépemc" OR Tla-o-qui-aht OR Tlatlasikwala OR Tlingit OR Tlowitsis OR Tootinaowaziibeeng OR Toquaht OR "Tr'ondëk Hwëch'in" OR Tsartlip OR Tsawout OR Tsawwassen OR Tseshaht OR Tseycum OR "Ts'il Kaz Koh" OR Tsleil-Waututh OR Tuscarora OR Tutchone OR Tzeachten OR Uchucklesaht OR Ucluelet OR Ulkatcho OR Wabaseemoong OR Wabauskang OR Wagmatcook OR Wahnapitae OR "Wahpeton Dakota" OR Wahsipekuk OR Wapekeka OR Wasagamack OR Wasauksing OR Waswanipi OR Wawakapewin OR Waywayseecappo OR Webequie OR Weenusk OR Wemotaci OR Wendat OR "Wha Ti" OR Whapmagoostui OR "Whitecap Dakota" OR Wikwemikong OR Witset OR Wolastoqiyik OR Wuikinuxv OR Wunnumin OR "Wuskwi Sipihk" OR Xatsull OR "Xeni Gwet'in" OR Yakweakwioose OR Yekooche OR "Yellow Quill" OR Yellowknife OR Yellowknives OR Zhiibaahaasing) OR KP=(Anishnawbek OR "Akisq'nuk" OR "aq'am" OR "Esdilagh" OR "Abénakis" OR "Akisq'nuk" OR "aq'am" OR "Che:k:tles7et'h'" OR "Da'naxda'xw" OR "Dzawada'enuxw" OR "Esk'etemc" OR "Gitga'at" OR "Gitlaxt'aamiks" OR "Gwa'Sala-Nakwaxda'xw" OR "Gwich'in" OR "Ka:'yu:'k't'h'" OR "Kanyen'kehaka" OR "Kapawe'no" OR "K'atlodeeche" OR "K'ómoks" OR "Leq'á:mel" OR "Lil'wat" OR "Malécite" OR "Mi'kmaq" OR "Mowachaht/Muchalaht" OR "Nisga'a" OR "N'Quatqua" OR "O'Chiese" OR "Ononda'gega'" OR "Onondowága'" OR "Onyota'a:ka" OR "Saik'uz" OR "Sambaa K'e " OR "Shxwhá:y" OR "Shxw'ow'hamel" OR "SIPEKNE'KATIK" OR "Skaru:reh" OR "Sq'éwlets" OR "Stellat'en" OR "Sts'ailes" OR "Stz'uminus" OR "T'it'q'et" OR "Tla'amin" OR "Tl'azt'en" OR "Tl'etinqox" OR "Tsal'alh" OR "Tsideldel" OR "Ts'kw'aylaxw" OR "T'Sou-ke" OR "Ts'uubaa-asatx" OR "Tsuut'ina" OR "We'koqma'q" OR "Wet'suwet'en" OR "Xai'xais" OR "Xaxli'p" OR "Yunesit'in" OR ((Acadia OR Carmacks OR Champagne OR Creek OR Delaware OR Douglas OR Montana OR Similkameen) NEAR/2 (Indian OR Indians OR Tribe OR Tribes OR native OR natives OR nation OR people*)) OR Aamjiwnaang OR Abegweit OR Abitibiwinni OR Ahousaht OR Ahtahkakoop OR Aishihik OR Aitchelitz OR Aklavik OR Algonquin* OR Anicinape* OR "Animakee Wa Zhing" OR Anishina* OR aqam OR Atikamekw OR Atjanasca* OR Attawapiskat OR Aundeck-Omni-Kaning OR Batchewana OR "Bearfoot Onondaga" OR Beausoleil OR "Behdzi Ahda*" OR "Biigtigong Nishnaabeg" OR "Biinjitiwaabik Zaaging Anishinaabek" OR Bilijk OR "Binche Whut'en" OR "Bingwi Neyaashi Anishinaabek" OR "Canupawakpa Dakota" OR Cayuga OR Chawathil OR Cheam OR "Cheslatta Carrier" OR Chiniki OR Chipew* OR Cote OR Couchiching OR Cowessess OR Cowichan OR Cree OR "Dakota Plains" OR "Dakota Tipi" OR "Dechi Laot'i" OR Deline OR Dena OR Dene OR Denesuline OR "Deninu K'ue" OR Ditidaht OR "Dog Rib Rae" OR Dokis OR Eabametoong OR Ehattesaht OR Elsipogtog OR Ermineskin OR Esdilagh OR Esgenoopetitj OR Eskasoni OR Esquimalt OR Gameti OR Gayogohono OR Ginoogaming OR Gitanmaax OR Gitanyow OR Gitsegukla OR Gitwangak OR Gitxaala OR Glooscap OR Gwawaenuk OR "Gwichya Gwich'in" OR Gwitchin OR Hagwilget OR Haisla OR Halalt OR Haudenosaunee OR Heiltsuk OR Hesquiaht OR Hiawatha OR Homalco OR Hupacasath OR Huu-ay-aht OR Inninuwug OR Innu OR Innue OR Innus OR Inuvik OR Iskatewizaagegan OR Iskut OR "Ka'a'gee Tu" OR Kahkewistahaw OR "Kasabonika Lake" OR Kashechewan OR Katzie OR Kawacatoose OR Kebaowek OR Keeseekoose OR Keeseekoowenin OR Kee-Way-Win OR "Kinistin Saulteaux" OR Kinonjeoshtegon OR Kispiox OR Kitselas OR Kitsumkalum OR Kluane OR Kutchin OR Kwadacha OR Kwakiutl OR "Kwanlin Dun" OR Kwaw-kwaw-Apilt OR Kwiakah OR "Kwikwasut'inuxw Haxwa'mis" OR Kwikwetlem OR "Lax Kw'alaams" OR "Lheidli T'enneh" OR "Liidlii Kue" OR "Listuguj Mi'gmaq" OR Lyackson OR Magnetawan OR Malahat OR Maliseet OR Mamalilikulla OR Matachewan OR Matsqui OR Mattagami OR "M'Chigeeng" OR Membertou OR Metepenagiag OR Metlakatla OR Miawpukek OR Michipicoten OR Micmacs OR Millbrook OR Mishkeegogamang OR Mississauga* OR Mitaanjigamiing OR Mohawk* OR Montagnais OR Moosomin OR Moravian OR Mowachaht OR Muchalaht OR Munsee-Delaware OR Muscowpetung OR Muskoday OR Muskowekwan OR Musqueam OR "Nacho Nyak Dun" OR "Nadleh Whuten" OR Naicatchewenin OR "Nak'azdli Whut'en" OR Nakota OR Naotkamegwanning OR Naskapi OR Natoaganeg OR Nee-Tahi-Buhn OR Nehiyawak OR Nekaneet OR Neskantaga OR Neskonlith OR Nibinamik OR Nigigoonsiminikaaning OR Nipissing OR Nishnaabeg OR Nooaitch OR Nuchatlaht OR Nuxalk OR Ochapowace OR O-Chi-Chak-Ko-Sipi OR Odanak OR Ojibw* OR Okanagan OR Okanese OR Okemasis OR Oneida OR Oromocto OR Osoyoos OR Pabineau OR Pacheedaht OR "Paqtnkek Mi'kmaw" OR Pasqua OR Pauingassi OR Pauquachin OR "Pays Plat" OR Peguis OR "Pehdzeh Ki" OR Pekuakamiulnuatsh OR Penelakut OR Piapot OR Piikani OR Pikangikum OR Pinaymootang OR Potlotek OR "Qalipu Mi'kmaq" OR Qualicum OR Quatsino OR Sagamok OR Sahgaiehcan OR Samahquam OR Saugeen OR Saulteau OR Saulteaux OR Sechelt OR Secwepemc* OR Semiahmoo OR Seneca OR Shackan OR Shamattawa OR Shawanaga OR Sheguiandah OR Sheshegwaning OR Shuswap OR Siksika OR Simpcw OR Sioux OR Siska OR Skatin OR Skawahlook OR Skeetchestn OR Skidegate OR "Skin Tyee" OR Skowkale OR Skownan OR Skuppah OR Skwah OR Snuneymuxw OR Songhees OR Soowahlie OR Splatsin OR Spuzzum OR Squamish OR Squiala OR "Stswecem'c Xget'tem" OR Sumas OR "Ta'an Kwach'an" OR Tagish OR Tahltan OR Takla OR Tallcree OR "Taykwa Tagamou" OR "Teetl'it Gwich'in" OR Temagami OR Thessalon OR Timiskaming OR "Tk'emlúps te Secwépemc" OR Tla-o-qui-aht OR Tlatlasikwala OR Tlingit OR Tlowitsis OR Tootinaowaziibeeng OR Toquaht OR "Tr'ondëk Hwëch'in" OR Tsartlip OR Tsawout OR Tsawwassen OR Tseshaht OR Tseycum OR "Ts'il Kaz Koh" OR Tsleil-Waututh OR Tuscarora OR Tutchone OR Tzeachten OR Uchucklesaht OR Ucluelet OR Ulkatcho OR Wabaseemoong OR Wabauskang OR Wagmatcook OR Wahnapitae OR "Wahpeton Dakota" OR Wahsipekuk OR Wapekeka OR Wasagamack OR Wasauksing OR Waswanipi OR Wawakapewin OR Waywayseecappo OR Webequie OR Weenusk OR Wemotaci OR Wendat OR "Wha Ti" OR Whapmagoostui OR "Whitecap Dakota" OR Wikwemikong OR Witset OR Wolastoqiyik OR Wuikinuxv OR Wunnumin OR "Wuskwi Sipihk" OR Xatsull OR "Xeni Gwet'in" OR Yakweakwioose OR Yekooche OR "Yellow Quill" OR Yellowknife OR Yellowknives OR Zhiibaahaasing) | 27523 | 30527 |
| 7 | TI=("Kwakwaka'wakw" OR "Kwak'wala" OR "L'uml'umuluts" OR "Lheidli T'enneh " OR "Nlaka'pamux" OR "Nuxálk" OR "Quw'utsun" OR "Qwum'yiqun" OR "S'amuna" OR "Sm'algyax" OR "St'at'imc" OR "Stl'atl'imx" OR "Sto:lo" OR "Tl'ulpalus" OR "Tsilhqot'in" OR "T'Souke" OR "Xa'xtsa" OR (Labrador NEAR/2 (Indian OR Indians OR Tribe OR Tribes OR native OR natives OR nation OR people*)) OR "Áa Tlein Kwáan" OR Aivilingmiut OR Algonkian OR Algonquian OR Algonquin OR Anishinaabe OR "Atlin people" OR "Baffin Island" OR "Bella Coola" OR Burrard OR Cheklesahht OR Chilcotin OR Chippew* OR Clayoquot OR Clemclemaluts OR Comiaken OR Comox OR Coquitlam OR Dakelh OR Dane-zaa OR "Deisleen Kwáan" OR Dunne-za OR Gitxsan OR Gitxsanimaax OR Guyohkohnyo OR "Haad kil" OR Halkomelem OR Halqemeylem OR Henaksiala OR Hinupsum OR Hunquminum OR Hwlitsum OR Hwulqwselu OR Hydin OR Iglulik OR Iglulingmuit OR Ininew OR Inuinnait OR Inuk OR Inupiat OR Inuvialuit OR Iroquois OR Kainai OR Kaska OR Kelsemeht OR Khenipsen OR Kilpahlas OR Kimsquit OR Kitamaat OR Kitlinermiut OR Kivallirmiut OR Klahoose OR Koksilah OR Kootenay OR Koskimo OR Ktunaxa OR Kwantlen OR Kwatna OR Kyuquot OR Labradormiut OR Laich-kwil-tach OR Lamalch* OR Lekwungen OR Lillooet OR Loucheaux OR Lummi OR Micmac OR Mississaugas OR Mohawk OR Mowachaht-Muchalaht OR Nadene OR Na-Dene OR Namgis OR Nanaimo OR Nanoose OR Nequatque OR Netsilik OR Netsilingmiut OR Niitsitapi OR Nimpkish OR Nootka OR Nunavimiut OR Nunavimmiut OR Nuu-chah-nulth OR Odawa OR Ohiaht OR Oji-cree OR Onayotekaono OR Onondaga OR Onondowahgah OR Onundagaono OR Opetchesaht OR Owekeeno OR Passamaquoddy OR Peigan OR Petun OR Plains-Cree OR Popkum OR Potawatomi OR Quamichan OR Sahtu OR Salish OR Sarcee OR Scowlitz OR "Seabird Island" OR Secwepemc OR Sekani OR Semahquam OR Shishalh OR Sinixt OR Ska-Ruh-Reh OR Skway OR Sliammon OR Snaw-naw-as OR Somena OR Songish OR Sooke OR Stoney OR Stuie OR Tallheo OR Tasttine OR Teslin OR Tlicho OR Tsimshianic OR Tununirmiut OR Ungava OR Wakashan OR Weewaikai OR Wewaykum OR Wyandot OR Yuculta) OR AB=("Kwakwaka'wakw" OR "Kwak'wala" OR "L'uml'umuluts" OR "Lheidli T'enneh " OR "Nlaka'pamux" OR "Nuxálk" OR "Quw'utsun" OR "Qwum'yiqun" OR "S'amuna" OR "Sm'algyax" OR "St'at'imc" OR "Stl'atl'imx" OR "Sto:lo" OR "Tl'ulpalus" OR "Tsilhqot'in" OR "T'Souke" OR "Xa'xtsa" OR (Labrador NEAR/2 (Indian OR Indians OR Tribe OR Tribes OR native OR natives OR nation OR people*)) OR "Áa Tlein Kwáan" OR Aivilingmiut OR Algonkian OR Algonquian OR Algonquin OR Anishinaabe OR "Atlin people" OR "Baffin Island" OR "Bella Coola" OR Burrard OR Cheklesahht OR Chilcotin OR Chippew* OR Clayoquot OR Clemclemaluts OR Comiaken OR Comox OR Coquitlam OR Dakelh OR Dane-zaa OR "Deisleen Kwáan" OR Dunne-za OR Gitxsan OR Gitxsanimaax OR Guyohkohnyo OR "Haad kil" OR Halkomelem OR Halqemeylem OR Henaksiala OR Hinupsum OR Hunquminum OR Hwlitsum OR Hwulqwselu OR Hydin OR Iglulik OR Iglulingmuit OR Ininew OR Inuinnait OR Inuk OR Inupiat OR Inuvialuit OR Iroquois OR Kainai OR Kaska OR Kelsemeht OR Khenipsen OR Kilpahlas OR Kimsquit OR Kitamaat OR Kitlinermiut OR Kivallirmiut OR Klahoose OR Koksilah OR Kootenay OR Koskimo OR Ktunaxa OR Kwantlen OR Kwatna OR Kyuquot OR Labradormiut OR Laich-kwil-tach OR Lamalch* OR Lekwungen OR Lillooet OR Loucheaux OR Lummi OR Micmac OR Mississaugas OR Mohawk OR Mowachaht-Muchalaht OR Nadene OR Na-Dene OR Namgis OR Nanaimo OR Nanoose OR Nequatque OR Netsilik OR Netsilingmiut OR Niitsitapi OR Nimpkish OR Nootka OR Nunavimiut OR Nunavimmiut OR Nuu-chah-nulth OR Odawa OR Ohiaht OR Oji-cree OR Onayotekaono OR Onondaga OR Onondowahgah OR Onundagaono OR Opetchesaht OR Owekeeno OR Passamaquoddy OR Peigan OR Petun OR Plains-Cree OR Popkum OR Potawatomi OR Quamichan OR Sahtu OR Salish OR Sarcee OR Scowlitz OR "Seabird Island" OR Secwepemc OR Sekani OR Semahquam OR Shishalh OR Sinixt OR Ska-Ruh-Reh OR Skway OR Sliammon OR Snaw-naw-as OR Somena OR Songish OR Sooke OR Stoney OR Stuie OR Tallheo OR Tasttine OR Teslin OR Tlicho OR Tsimshianic OR Tununirmiut OR Ungava OR Wakashan OR Weewaikai OR Wewaykum OR Wyandot OR Yuculta) OR AK=("Kwakwaka'wakw" OR "Kwak'wala" OR "L'uml'umuluts" OR "Lheidli T'enneh " OR "Nlaka'pamux" OR "Nuxálk" OR "Quw'utsun" OR "Qwum'yiqun" OR "S'amuna" OR "Sm'algyax" OR "St'at'imc" OR "Stl'atl'imx" OR "Sto:lo" OR "Tl'ulpalus" OR "Tsilhqot'in" OR "T'Souke" OR "Xa'xtsa" OR (Labrador NEAR/2 (Indian OR Indians OR Tribe OR Tribes OR native OR natives OR nation OR people*)) OR "Áa Tlein Kwáan" OR Aivilingmiut OR Algonkian OR Algonquian OR Algonquin OR Anishinaabe OR "Atlin people" OR "Baffin Island" OR "Bella Coola" OR Burrard OR Cheklesahht OR Chilcotin OR Chippew* OR Clayoquot OR Clemclemaluts OR Comiaken OR Comox OR Coquitlam OR Dakelh OR Dane-zaa OR "Deisleen Kwáan" OR Dunne-za OR Gitxsan OR Gitxsanimaax OR Guyohkohnyo OR "Haad kil" OR Halkomelem OR Halqemeylem OR Henaksiala OR Hinupsum OR Hunquminum OR Hwlitsum OR Hwulqwselu OR Hydin OR Iglulik OR Iglulingmuit OR Ininew OR Inuinnait OR Inuk OR Inupiat OR Inuvialuit OR Iroquois OR Kainai OR Kaska OR Kelsemeht OR Khenipsen OR Kilpahlas OR Kimsquit OR Kitamaat OR Kitlinermiut OR Kivallirmiut OR Klahoose OR Koksilah OR Kootenay OR Koskimo OR Ktunaxa OR Kwantlen OR Kwatna OR Kyuquot OR Labradormiut OR Laich-kwil-tach OR Lamalch* OR Lekwungen OR Lillooet OR Loucheaux OR Lummi OR Micmac OR Mississaugas OR Mohawk OR Mowachaht-Muchalaht OR Nadene OR Na-Dene OR Namgis OR Nanaimo OR Nanoose OR Nequatque OR Netsilik OR Netsilingmiut OR Niitsitapi OR Nimpkish OR Nootka OR Nunavimiut OR Nunavimmiut OR Nuu-chah-nulth OR Odawa OR Ohiaht OR Oji-cree OR Onayotekaono OR Onondaga OR Onondowahgah OR Onundagaono OR Opetchesaht OR Owekeeno OR Passamaquoddy OR Peigan OR Petun OR Plains-Cree OR Popkum OR Potawatomi OR Quamichan OR Sahtu OR Salish OR Sarcee OR Scowlitz OR "Seabird Island" OR Secwepemc OR Sekani OR Semahquam OR Shishalh OR Sinixt OR Ska-Ruh-Reh OR Skway OR Sliammon OR Snaw-naw-as OR Somena OR Songish OR Sooke OR Stoney OR Stuie OR Tallheo OR Tasttine OR Teslin OR Tlicho OR Tsimshianic OR Tununirmiut OR Ungava OR Wakashan OR Weewaikai OR Wewaykum OR Wyandot OR Yuculta) OR KP=("Kwakwaka'wakw" OR "Kwak'wala" OR "L'uml'umuluts" OR "Lheidli T'enneh " OR "Nlaka'pamux" OR "Nuxálk" OR "Quw'utsun" OR "Qwum'yiqun" OR "S'amuna" OR "Sm'algyax" OR "St'at'imc" OR "Stl'atl'imx" OR "Sto:lo" OR "Tl'ulpalus" OR "Tsilhqot'in" OR "T'Souke" OR "Xa'xtsa" OR (Labrador NEAR/2 (Indian OR Indians OR Tribe OR Tribes OR native OR natives OR nation OR people*)) OR "Áa Tlein Kwáan" OR Aivilingmiut OR Algonkian OR Algonquian OR Algonquin OR Anishinaabe OR "Atlin people" OR "Baffin Island" OR "Bella Coola" OR Burrard OR Cheklesahht OR Chilcotin OR Chippew* OR Clayoquot OR Clemclemaluts OR Comiaken OR Comox OR Coquitlam OR Dakelh OR Dane-zaa OR "Deisleen Kwáan" OR Dunne-za OR Gitxsan OR Gitxsanimaax OR Guyohkohnyo OR "Haad kil" OR Halkomelem OR Halqemeylem OR Henaksiala OR Hinupsum OR Hunquminum OR Hwlitsum OR Hwulqwselu OR Hydin OR Iglulik OR Iglulingmuit OR Ininew OR Inuinnait OR Inuk OR Inupiat OR Inuvialuit OR Iroquois OR Kainai OR Kaska OR Kelsemeht OR Khenipsen OR Kilpahlas OR Kimsquit OR Kitamaat OR Kitlinermiut OR Kivallirmiut OR Klahoose OR Koksilah OR Kootenay OR Koskimo OR Ktunaxa OR Kwantlen OR Kwatna OR Kyuquot OR Labradormiut OR Laich-kwil-tach OR Lamalch* OR Lekwungen OR Lillooet OR Loucheaux OR Lummi OR Micmac OR Mississaugas OR Mohawk OR Mowachaht-Muchalaht OR Nadene OR Na-Dene OR Namgis OR Nanaimo OR Nanoose OR Nequatque OR Netsilik OR Netsilingmiut OR Niitsitapi OR Nimpkish OR Nootka OR Nunavimiut OR Nunavimmiut OR Nuu-chah-nulth OR Odawa OR Ohiaht OR Oji-cree OR Onayotekaono OR Onondaga OR Onondowahgah OR Onundagaono OR Opetchesaht OR Owekeeno OR Passamaquoddy OR Peigan OR Petun OR Plains-Cree OR Popkum OR Potawatomi OR Quamichan OR Sahtu OR Salish OR Sarcee OR Scowlitz OR "Seabird Island" OR Secwepemc OR Sekani OR Semahquam OR Shishalh OR Sinixt OR Ska-Ruh-Reh OR Skway OR Sliammon OR Snaw-naw-as OR Somena OR Songish OR Sooke OR Stoney OR Stuie OR Tallheo OR Tasttine OR Teslin OR Tlicho OR Tsimshianic OR Tununirmiut OR Ungava OR Wakashan OR Weewaikai OR Wewaykum OR Wyandot OR Yuculta) | 10800 | 12204 |
| 8 | #1 OR #2 | 48171 | 53472 |
| 9 | #3 OR #4 OR #5 OR #6 OR #7 | 147009 | 163006 |
| 10 | TI=(Climacteric OR Climacterium OR Menopause OR Menopaus* OR "Menopause, Premature" OR "climacterium praecox" OR Perimenopause OR Perimenopaus* OR Postmenopause OR Postmenopaus* OR "Post menopaus*" OR Post-menopaus* OR Premenopause OR Premenopause* OR Pre-menopaus* OR "Pre menopaus*" OR Climacteric* OR "Life Change*" OR "Change of life") OR AB=(Climacteric OR Climacterium OR Menopause OR Menopaus* OR "Menopause, Premature" OR "climacterium praecox" OR Perimenopause OR Perimenopaus* OR Postmenopause OR Postmenopaus* OR "Post menopaus*" OR Post-menopaus* OR Premenopause OR Premenopause* OR Pre-menopaus* OR "Pre menopaus*" OR Climacteric* OR "Life Change*" OR "Change of life") OR AK=(Climacteric OR Climacterium OR Menopause OR Menopaus* OR "Menopause, Premature" OR "climacterium praecox" OR Perimenopause OR Perimenopaus* OR Postmenopause OR Postmenopaus* OR "Post menopaus*" OR Post-menopaus* OR Premenopause OR Premenopause* OR Pre-menopaus* OR "Pre menopaus*" OR Climacteric* OR "Life Change*" OR "Change of life") OR KP=(Climacteric OR Climacterium OR Menopause OR Menopaus* OR "Menopause, Premature" OR "climacterium praecox" OR Perimenopause OR Perimenopaus* OR Postmenopause OR Postmenopaus* OR "Post menopaus*" OR Post-menopaus* OR Premenopause OR Premenopause* OR Pre-menopaus* OR "Pre menopaus*" OR Climacteric* OR "Life Change*" OR "Change of life") | 145108 | 158968 |
| 11 | TI=("Hot Flashes" OR "Hot Flash*" OR "Hot Flush*" OR "night sweat*" OR "nocturnal hyperhidrosis" OR "vasomotor symptom*" OR VMS) OR AB=("Hot Flashes" OR "Hot Flash*" OR "Hot Flush*" OR "night sweat*" OR "nocturnal hyperhidrosis" OR "vasomotor symptom*" OR VMS) OR AK=("Hot Flashes" OR "Hot Flash*" OR "Hot Flush*" OR "night sweat*" OR "nocturnal hyperhidrosis" OR "vasomotor symptom*" OR VMS) OR KP=("Hot Flashes" OR "Hot Flash*" OR "Hot Flush*" OR "night sweat*" OR "nocturnal hyperhidrosis" OR "vasomotor symptom*" OR VMS) | 14447 | 16312 |
| 12 | #10 OR #11 | 153798 | 168923 |
| 13 | TI=(female OR woman OR women) OR AB=(female OR woman OR women) OR AK=(female OR woman OR women) OR KP=(female OR woman OR women) | 2741321 | 3163786 |
| 14 | TI=("elder" OR "elders") OR AB=("elder" OR "elders") OR AK=("elder" OR "elders") OR KP=("elder" OR "elders") | 28672 | 31088 |
| 15 | #13 AND #14 | 4848 | 5302 |
| 16 | TI=(((Indigenous OR native OR "first nation" OR aborigin* OR Indian) NEAR/2 (health OR healthcare OR healing OR medicine OR practices)) OR ethnomedicin* OR "native healing" OR Limpia OR "indigenous healer" OR medicineman OR medicinemen OR "medicine man" OR "medicine men" OR Curandero OR "folk healer" OR shaman* OR shamen) OR AB=(((Indigenous OR native OR "first nation" OR aborigin* OR Indian) NEAR/2 (health OR healthcare OR healing OR medicine OR practices)) OR ethnomedicin* OR "native healing" OR Limpia OR "indigenous healer" OR medicineman OR medicinemen OR "medicine man" OR "medicine men" OR Curandero OR "folk healer" OR shaman* OR shamen) OR AK=(((Indigenous OR native OR "first nation" OR aborigin* OR Indian) NEAR/2 (health OR healthcare OR healing OR medicine OR practices)) OR ethnomedicin* OR "native healing" OR Limpia OR "indigenous healer" OR medicineman OR medicinemen OR "medicine man" OR "medicine men" OR Curandero OR "folk healer" OR shaman* OR shamen) OR KP=(((Indigenous OR native OR "first nation" OR aborigin* OR Indian) NEAR/2 (health OR healthcare OR healing OR medicine OR practices)) OR ethnomedicin* OR "native healing" OR Limpia OR "indigenous healer" OR medicineman OR medicinemen OR "medicine man" OR "medicine men" OR Curandero OR "folk healer" OR shaman* OR shamen) | 22639 | 25767 |
| 17 | TI=((traditional NEAR/2 (medicine* OR therap* OR healing OR treatment*)) OR "folk medicine" OR "folk healer") OR AB=((traditional NEAR/2 (medicine* OR therap* OR healing OR treatment*)) OR "folk medicine" OR "folk healer") OR AK=((traditional NEAR/2 (medicine* OR therap* OR healing OR treatment*)) OR "folk medicine" OR "folk healer") OR KP=((traditional NEAR/2 (medicine* OR therap* OR healing OR treatment*)) OR "folk medicine" OR "folk healer") | 90349 | 107747 |
| 18 | TI=("North America" OR Canada OR Alberta OR "British Columbia" OR Manitoba OR "New Brunswick" OR "Newfoundland" OR "Labrador" OR "Northwest Territories" OR "Nova Scotia" OR Nunavut OR Ontario OR "Prince Edward Island" OR Quebec OR Saskatchewan OR Yukon OR "United States" OR Appalachia* OR "New England" OR "Pacific States" OR Alabama OR Alaska OR Arizona OR Arkansas OR California OR Colorado OR Connecticut OR Delaware OR Florida OR Georgia OR Hawaii OR Idaho OR Illinois OR Indiana OR Iowa OR Kansas OR Kentucky OR Louisiana OR Maine OR Maryland OR Massachusetts OR Michigan OR Minnesota OR Mississippi OR Missouri OR Montana OR Nebraska OR Nevada OR "New Hampshire" OR "New Jersey" OR "New Mexico" OR "New York" OR "North Carolina" OR "North Dakota" OR Ohio OR Oklahoma OR Oregon OR Pennsylvania OR "Rhode Island" OR "South Carolina" OR "South Dakota" OR Tennessee OR Texas OR Utah OR Vermont OR Virginia OR Washington OR "West Virginia" OR Wisconsin OR Wyoming OR "District of Columbia" OR "Washington DC") OR AB=("North America" OR Canada OR Alberta OR "British Columbia" OR Manitoba OR "New Brunswick" OR "Newfoundland" OR "Labrador" OR "Northwest Territories" OR "Nova Scotia" OR Nunavut OR Ontario OR "Prince Edward Island" OR Quebec OR Saskatchewan OR Yukon OR "United States" OR Appalachia* OR "New England" OR "Pacific States" OR Alabama OR Alaska OR Arizona OR Arkansas OR California OR Colorado OR Connecticut OR Delaware OR Florida OR Georgia OR Hawaii OR Idaho OR Illinois OR Indiana OR Iowa OR Kansas OR Kentucky OR Louisiana OR Maine OR Maryland OR Massachusetts OR Michigan OR Minnesota OR Mississippi OR Missouri OR Montana OR Nebraska OR Nevada OR "New Hampshire" OR "New Jersey" OR "New Mexico" OR "New York" OR "North Carolina" OR "North Dakota" OR Ohio OR Oklahoma OR Oregon OR Pennsylvania OR "Rhode Island" OR "South Carolina" OR "South Dakota" OR Tennessee OR Texas OR Utah OR Vermont OR Virginia OR Washington OR "West Virginia" OR Wisconsin OR Wyoming OR "District of Columbia" OR "Washington DC") OR AK=("North America" OR Canada OR Alberta OR "British Columbia" OR Manitoba OR "New Brunswick" OR "Newfoundland" OR "Labrador" OR "Northwest Territories" OR "Nova Scotia" OR Nunavut OR Ontario OR "Prince Edward Island" OR Quebec OR Saskatchewan OR Yukon OR "United States" OR Appalachia* OR "New England" OR "Pacific States" OR Alabama OR Alaska OR Arizona OR Arkansas OR California OR Colorado OR Connecticut OR Delaware OR Florida OR Georgia OR Hawaii OR Idaho OR Illinois OR Indiana OR Iowa OR Kansas OR Kentucky OR Louisiana OR Maine OR Maryland OR Massachusetts OR Michigan OR Minnesota OR Mississippi OR Missouri OR Montana OR Nebraska OR Nevada OR "New Hampshire" OR "New Jersey" OR "New Mexico" OR "New York" OR "North Carolina" OR "North Dakota" OR Ohio OR Oklahoma OR Oregon OR Pennsylvania OR "Rhode Island" OR "South Carolina" OR "South Dakota" OR Tennessee OR Texas OR Utah OR Vermont OR Virginia OR Washington OR "West Virginia" OR Wisconsin OR Wyoming OR "District of Columbia" OR "Washington DC") OR KP=("North America" OR Canada OR Alberta OR "British Columbia" OR Manitoba OR "New Brunswick" OR "Newfoundland" OR "Labrador" OR "Northwest Territories" OR "Nova Scotia" OR Nunavut OR Ontario OR "Prince Edward Island" OR Quebec OR Saskatchewan OR Yukon OR "United States" OR Appalachia* OR "New England" OR "Pacific States" OR Alabama OR Alaska OR Arizona OR Arkansas OR California OR Colorado OR Connecticut OR Delaware OR Florida OR Georgia OR Hawaii OR Idaho OR Illinois OR Indiana OR Iowa OR Kansas OR Kentucky OR Louisiana OR Maine OR Maryland OR Massachusetts OR Michigan OR Minnesota OR Mississippi OR Missouri OR Montana OR Nebraska OR Nevada OR "New Hampshire" OR "New Jersey" OR "New Mexico" OR "New York" OR "North Carolina" OR "North Dakota" OR Ohio OR Oklahoma OR Oregon OR Pennsylvania OR "Rhode Island" OR "South Carolina" OR "South Dakota" OR Tennessee OR Texas OR Utah OR Vermont OR Virginia OR Washington OR "West Virginia" OR Wisconsin OR Wyoming OR "District of Columbia" OR "Washington DC") | 2135864 | 2349942 |
| 19 | TI=((women* OR woman* OR female*) NEAR/2 (health OR healthcare OR service* OR clinic OR clinics)) OR AB=((women* OR woman* OR female*) NEAR/2 (health OR healthcare OR service* OR clinic OR clinics)) OR AK=((women* OR woman* OR female*) NEAR/2 (health OR healthcare OR service* OR clinic OR clinics)) OR KP=((women* OR woman* OR female*) NEAR/2 (health OR healthcare OR service* OR clinic OR clinics)) | 62386 | 71677 |
| 20 | #8 AND #12 | 177 | 189 |
| 21 | #9 AND #12 | 196 | 222 |
| 22 | #16 AND #12 | 48 | 51 |
| 23 | #15 AND (#8 OR #9) | 165 | 186 |
| 24 | #19 AND (#8 OR #9) | 490 | 567 |
| 25 | #17 AND (#12 OR #15) AND #18 | 30 | 33 |
| 26 | #20 OR #21 OR #22 OR #23 OR #24 OR #25 and Book Review (Exclude – Document Types) Timespan: 2023-06-08 to 2024-11-21 | N/A | 133 |
| 26 | #20 OR #21 OR #22 OR #23 OR #24 OR #25 | 1014 | N/A |
| 27 | #20 OR #21 OR #22 OR #23 OR #24 OR #25 and Book Review (Exclude – Document Types) | 1011 | N/A |

### Scopus (Elsevier)

**Field codes:** TITLE-ABS-KEY = Document Title, Abstract, Keywords; ORIG-LOAD-DATE = the day the record was added to Scopus
**Proximity operator:** W/#
**Truncation:** *

**Date of Initial Search:** 8 June 2023
**Results:** 867
**Updated (Shown):** 21 November 2023
**Results of Update:** 76

( ( ( ( TITLE-ABS-KEY ( ( ( american* OR alaska* ) W/2 ( native* OR indian OR indians OR indigenous OR aborigin* ) ) OR amerind* ) ) OR ( TITLE-ABS-KEY ( ( ( canada OR canadian* ) W/2 ( native* OR indigenous OR aborigin* ) ) OR metis OR "M&#233;tis" OR michif* OR mitchif OR mechif OR "M&#233;tif" OR "M&#233;tchif" OR metif OR metchif OR inuit OR "First Nations" OR "First Nation" ) ) OR ( TITLE-ABS-KEY ( inuit OR ( arctic W/2 people* ) ) ) OR ( TITLE-ABS-KEY ( "Asa&apos;carsarmiut" OR "Dee-ni&apos;" OR "Mi&apos;kmaq" OR "S&apos;Klallam" OR "Shoonaq&apos;" OR "Sun&apos;aq" OR ( ( "Ivanof Bay" OR "Shoalwater Bay" OR "Spirit Lake" OR "Squaxin Island" OR dakota OR dakotah OR wichita OR keechi OR waco OR tawakonie OR cheyenne OR creek OR crow OR hoonah OR kasigluk OR maricopa OR miami OR omaha OR oneida OR ottawa OR peoria OR salamatof OR sitka OR snoqualmie OR spokane OR winnebago OR wyandotte ) W/2 ( indian OR indians OR tribe OR tribes OR native OR natives OR nation OR people* ) ) OR ( ( native OR indian OR traditional ) W/2 ( village OR council OR community OR association ) ) OR absentee-shawnee OR achomawi OR agdaagux OR "Agua Caliente" OR ajumawi OR ak-chin OR akiachak OR akiak OR alabama-coushatta OR alabama-quassarte OR alatna OR algaaciq OR algin OR allakaket OR "Alturas Indian Rancheria" OR alutiiq OR angoon OR anvik OR apache OR aporige OR aquinnah OR arapaho OR aroostook OR assiniboine OR astarawi OR atsugewi OR atwamsini OR bdewakantunwan OR caddo OR cahto OR cahuilla OR catawba OR cayuga OR chalkyitsik OR cheesh-na OR chehalis OR chemehuevi OR cher-ae OR chevak OR cheyenne-arapaho OR chickahominy OR chickaloon OR chickasaw OR chignik OR chilkat OR chilkoot OR chinik OR chinook OR chippewa* OR chitimacha OR choctaw OR chukchansi OR chuloonawick OR chumash OR cocopah OR colusa OR comanche OR coos OR cosmit OR coushatta OR cowlitz OR "Cross Village" OR cupeno OR curyung OR diegueno OR dine&apos; OR egegik OR eklutna OR elwha OR "Emmonak Village" OR "Evansville Village" OR ewiiaapaayp OR "Galena Village" OR goshute OR "Gulkana Village Council" OR haida OR hammawi OR havasupai OR "Healy Lake Village" OR hewisedawi OR ho-chunk OR hoh OR hokan OR holikachuk OR "Holy Cross Tribe" OR hualapai OR "Hughes Village" OR "Huslia Village" OR "Hydaburg Cooperative Association" OR "Igiugig Village" OR iipay OR ilmawi OR inaja OR inupiat OR ione OR itsatawi OR "Kaguyak Village" OR kaibab OR "Kaktovik Village" OR kalispel OR kanosh OR karuk OR kashia OR kaw OR keetoowah OR "Kenaitze Indian Tribe" OR "Ketchikan Indian Community" OR "Keweenaw Bay" OR kialegee OR kickapoo OR "King Island Native Community" OR "King Salmon Tribe" OR kiowa OR klamath OR "Klawock Cooperative Association" OR kletsel OR "Knik Tribe" OR "Koi Nation" OR "Kokhanok Village" OR koosharem OR kootenai OR kosalektawi OR "Koyukuk Native Village" OR kumeyaay OR kwalhioqua OR "Lesnoi Village" OR "Levelock Village" OR "Lime Village" OR "Louden Village" OR luiseno OR lummi OR madesi OR maidu OR makah OR maliseet OR "Manley Hot Springs Village" OR "Manokotak Village" OR manzanita OR mashantucket OR match-e-be-nash-she-wish OR mattaponi OR "McGrath Native Village" OR mdewakanton OR mechoopda OR menominee OR me-wuk OR miccosukee OR micmacs OR "Mission Indians" OR miwok OR miwuk OR modoc OR mohegan OR mojave OR monacan OR "Mono Indians" OR muckleshoot OR muscogee OR "Na Den&#233;" OR nansemond OR narragansett OR navajo OR "New Koliganek Village" OR "New Stuyahok Village" OR "Newhalen Village" OR "Newtok Village" OR "Nez Perce" OR "Nikolai Village" OR "Ninilchik Village" OR nisqually OR nomlaki OR "Nondalton Village" OR nooksack OR "Northway Village" OR nottawaseppi OR "Nulato Village" OR nunakauyarmiut OR odawa OR ohgsenakale OR "Ohkay Owingeh" OR onondaga OR oreilles OR "Organized Village of Grayling" OR "Organized Village of Kake" OR "Organized Village of Kasaan" OR "Organized Village of Kwethluk" OR "Organized Village of Saxman" OR orutsararmiut OR osage OR otoe-missouria OR paiute OR paiute-shoshone OR pala OR pamunkey OR pascua OR paskenta OR passamaquoddy OR "Pauloff Harbor Village" OR pauma OR pawnee OR pechanga OR "Pechanga " OR "Peck Indian Reservation" OR "Pedro Bay Village" OR pee-posh OR penobscot OR penutian OR perquot OR "Pima People" OR pima-maricopa OR poarch OR pokagon OR pomo OR ponca OR "Portage Creek Village" OR potawatomi OR pottawatomi OR pueblo OR puyallup OR "Qagan Tayagungin" OR qawalangin OR quapaw OR quechan OR "Rampart Village" OR rappahannock OR resighini OR "Sac &amp; Fox" OR "Sac and Fox" OR "Saint George Island" OR "Saint Paul Island" OR salish OR samish OR santee OR sauk-suiattle OR "Seldovia Village Tribe" OR seminole OR seneca OR seneca-cayuga OR shinnecock OR shivwits OR shoshone OR shoshone-bannock OR shoshone-paiute OR siuslaw OR skagit OR "Skagway Village" OR skokomish OR "South Naknek Village" OR "Stebbins Community Association" OR stillaguamish OR suquamish OR swinomish OR "Takotna Village" OR tangirnaq OR tejon OR "Telida Village" OR te-moak OR thlopthlocco OR "Tlingit &amp; Haida" OR "Tohono O&apos;odham" OR tolowa OR tonawanda OR tonkawa OR tulalip OR tunica-biloxi OR tuolumne OR tuscarora OR "Twin Hills Village" OR "Ugashik Village" OR umatilla OR umkumiut OR umpqua OR ute OR "Utu Utu Gwaitu" OR "Village of Alakanuk" OR "Village of Anaktuvuk Pass" OR "Village of Aniak" OR "Village of Atmautluak" OR "Village of Bill Moore&apos;s Slough" OR "Village of Chefornak" OR "Village of Clarks Point" OR "Village of Crooked Creek" OR "Village of Dot Lake" OR "Village of Iliamna" OR "Village of Kalskag" OR "Village of Kaltag" OR "Village of Kotlik" OR "Village of Lower Kalskag" OR "Village of Ohogamiut" OR "Village of Red Devil" OR "Village of Sleetmute" OR "Village of Solomon" OR "Village of Stony River" OR "Village of Venetie" OR "Village of Wainwright" OR wailaki OR wampanoag OR "Warm Springs Reservation" OR washoe OR willapa OR winnemucca OR wintun OR wiyot OR "Wrangell Cooperative Association" OR yakama OR yakutat OR yaqui OR yavapai OR yokuts OR yuhaaviatam OR yuima OR yukian OR yukut OR yupiit OR zuni ) ) OR ( TITLE-ABS-KEY ( "Degexit&apos;an" OR "Yup&apos;ik" OR ( "Bear River" W/2 ( indian OR indians OR tribe OR tribes OR native OR natives OR nation OR people* ) ) OR abenaki OR achumawi OR ahtna OR "Akimel Oodham" OR "Akimel O&apos;odham" OR "Akimel Ootham" OR "Akimel O&apos;otham" OR aleut* OR algonkian* OR alsea OR aluutiq OR apalachee OR arikara OR atakapa OR athabascan OR athabaskan OR athapascan OR athapaskan OR bannock OR bidai OR biloxi OR blackfeet OR blackfoot OR caddoan OR calice OR calusa OR carrizo OR cayuse OR chemakum OR cheraw OR cherokee OR chetco OR cheyenne OR chicora OR chilula OR chimariko OR clatskanie OR coahuiltecan OR "Coeur d&apos;Alene" OR costanoan OR cree OR croatan OR croatoan OR cusabo OR edisto OR eskaleut* OR eskimo OR eskimo-aleut OR esselen OR eyak OR "Fox and Sauk" OR gabrielino OR "Gros Ventre" OR guale OR "Gwich&apos;in" OR halchidhoma OR hidatsa OR hitchiti OR hopi OR houma OR huichol* OR hupa OR ingalik OR inupiaq OR ioway OR iroquoi* OR jacome OR jano OR jeaga OR juaneno OR jumano OR kalapuya OR kansa OR karankawa OR karok OR kashaya OR kato OR kawaiisu OR kitanemuk OR kitsai OR klallam OR koasati OR kolchan OR konkow OR koyukon OR lakota* OR lassik OR laurentian OR lenape OR "Lenni Lenape" OR mahican OR mandan OR maricopa OR massachusett OR mattole OR minisink OR missouria OR mohave OR mohawk OR mohican OR molalla OR multnomah OR munsee OR muscoge* OR naabeeho OR nakoda OR nanticoke OR natchez OR nauset OR navaho OR niantic OR nipmuc OR nisenan OR nongatl OR oconee OR ofo OR ohlone OR ojibw* OR okanagan OR oneida OR otoe OR palouse OR panamint OR papago OR patwin OR "Pee Dee" OR pennacook OR pequot OR pima OR piro OR pocasset OR pocomtuc OR pokanoket OR poospatuck OR powhatan OR quechan OR quileute OR quinault OR quiripi OR salinan OR samich OR saponi OR serrano OR shasta OR shawnee OR siksikaitsitapi OR sinkyone OR siouan OR sioux OR snohomish OR snoqualmie OR squaxin OR suma OR susquehannock OR takelma OR tanacross OR tanana OR tataviam OR tawakoni OR tequesta OR tillamook OR timucua OR tlingit OR "Tohono Oodham" OR tongva OR tsimshian OR tubatulabal OR tutelo OR tututni OR unalachtigo OR unami OR unangan OR unangas OR unkechaug OR waccamaw OR walapai OR wappo OR wasco OR whilkut OR wichita OR winnebago OR wintu OR wishram OR yahi OR yakima OR yamasee OR yana OR yaqu OR yuchi OR yuki OR yuma OR yupik OR yurok ) ) OR ( TITLE-ABS-KEY ( anishnawbek OR "?Akisq&apos;nuk" OR "?aq&apos;am" OR "?Esdilagh" OR "Ab&#233;nakis" OR "Akisq&apos;nuk" OR "aq&apos;am" OR "Che:k:tles7et&apos;h&apos;" OR "Da&apos;naxda&apos;xw" OR "Dzawada&apos;enuxw" OR "Esk&apos;etemc" OR "Gitga&apos;at" OR "Gitlaxt&apos;aamiks" OR "Gwa&apos;Sala-Nakwaxda&apos;xw" OR "Gwich&apos;in" OR "Ka:&apos;yu:&apos;k&apos;t&apos;h&apos;" OR "Kanyen&apos;kehaka" OR "Kapawe&apos;no" OR "K&apos;atlodeeche" OR "K&apos;&#243;moks" OR "Leq&apos;&#225;:mel" OR "Lil&apos;wat" OR "Mal&#233;cite" OR "Mi&apos;kmaq" OR "Mowachaht/Muchalaht" OR "Nisga&apos;a" OR "N&apos;Quatqua" OR "O&apos;Chiese" OR "Ononda&apos;gega&apos;" OR "Onondow&#225;ga&apos;" OR "Onyota&apos;a:ka" OR "Saik&apos;uz" OR "Sambaa K&apos;e " OR "Shxwh&#225;:y" OR "Shxw&apos;ow&apos;hamel" OR "SIPEKNE&apos;KATIK" OR "Skaru:reh" OR "Sq&apos;&#233;wlets" OR "Stellat&apos;en" OR "Sts&apos;ailes" OR "Stz&apos;uminus" OR "T&apos;it&apos;q&apos;et" OR "Tla&apos;amin" OR "Tl&apos;azt&apos;en" OR "Tl&apos;etinqox" OR "Tsal&apos;alh" OR "Tsideldel" OR "Ts&apos;kw&apos;aylaxw" OR "T&apos;Sou-ke" OR "Ts&apos;uubaa-asatx" OR "Tsuut&apos;ina" OR "We&apos;koqma&apos;q" OR "Wet&apos;suwet&apos;en" OR "Xai&apos;xais" OR "Xaxli&apos;p" OR "Yunesit&apos;in" OR ( ( acadia OR carmacks OR champagne OR creek OR delaware OR douglas OR montana OR similkameen ) W/2 ( indian OR indians OR tribe OR tribes OR native OR natives OR nation OR people* ) ) OR aamjiwnaang OR abegweit OR abitibiwinni OR ahousaht OR ahtahkakoop OR aishihik OR aitchelitz OR aklavik OR algonquin* OR anicinape* OR "Animakee Wa Zhing" OR anishina* OR aqam OR atikamekw OR atjanasca* OR attawapiskat OR aundeck-omni-kaning OR batchewana OR "Bearfoot Onondaga" OR beausoleil OR "Behdzi Ahda*" OR "Biigtigong Nishnaabeg" OR "Biinjitiwaabik Zaaging Anishinaabek" OR bilijk OR "Binche Whut&apos;en" OR "Bingwi Neyaashi Anishinaabek" OR "Canupawakpa Dakota" OR cayuga OR chawathil OR cheam OR "Cheslatta Carrier" OR chiniki OR chipew* OR cote OR couchiching OR cowessess OR cowichan OR cree OR "Dakota Plains" OR "Dakota Tipi" OR "Dechi Laot&apos;i" OR deline OR dena OR dene OR denesuline OR "Deninu K&apos;ue" OR ditidaht OR "Dog Rib Rae" OR dokis OR eabametoong OR ehattesaht OR elsipogtog OR ermineskin OR esdilagh OR esgenoopetitj OR eskasoni OR esquimalt OR gameti OR gayogohono OR ginoogaming OR gitanmaax OR gitanyow OR gitsegukla OR gitwangak OR gitxaala OR glooscap OR gwawaenuk OR "Gwichya Gwich&apos;in" OR gwitchin OR hagwilget OR haisla OR halalt OR haudenosaunee OR heiltsuk OR hesquiaht OR hiawatha OR homalco OR hupacasath OR huu-ay-aht OR inninuwug OR innu OR innue OR innus OR inuvik OR iskatewizaagegan OR iskut OR "Ka&apos;a&apos;gee Tu" OR kahkewistahaw OR "Kasabonika Lake" OR kashechewan OR katzie OR kawacatoose OR kebaowek OR keeseekoose OR keeseekoowenin OR kee-way-win OR "Kinistin Saulteaux" OR kinonjeoshtegon OR kispiox OR kitselas OR kitsumkalum OR kluane OR kutchin OR kwadacha OR kwakiutl OR "Kwanlin Dun" OR kwaw-kwaw-apilt OR kwiakah OR "Kwikwasut&apos;inuxw Haxwa&apos;mis" OR kwikwetlem OR "Lax Kw&apos;alaams" OR "Lheidli T&apos;enneh" OR "Liidlii Kue" OR "Listuguj Mi&apos;gmaq" OR lyackson OR magnetawan OR malahat OR maliseet OR mamalilikulla OR matachewan OR matsqui OR mattagami OR "M&apos;Chigeeng" OR membertou OR metepenagiag OR metlakatla OR miawpukek OR michipicoten OR micmacs OR millbrook OR mishkeegogamang OR mississauga* OR mitaanjigamiing OR mohawk* OR montagnais OR moosomin OR moravian OR mowachaht OR muchalaht OR munsee-delaware OR muscowpetung OR muskoday OR muskowekwan OR musqueam OR "Nacho Nyak Dun" OR "Nadleh Whuten" OR naicatchewenin OR "Nak&apos;azdli Whut&apos;en" OR nakota OR naotkamegwanning OR naskapi OR natoaganeg OR nee-tahi-buhn OR nehiyawak OR nekaneet OR neskantaga OR neskonlith OR nibinamik OR nigigoonsiminikaaning OR nipissing OR nishnaabeg OR nooaitch OR nuchatlaht OR nuxalk OR ochapowace OR o-chi-chak-ko-sipi OR odanak OR ojibw* OR okanagan OR okanese OR okemasis OR oneida OR oromocto OR osoyoos OR pabineau OR pacheedaht OR "Paqtnkek Mi&apos;kmaw" OR pasqua OR pauingassi OR pauquachin OR "Pays Plat" OR peguis OR "Pehdzeh Ki" OR pekuakamiulnuatsh OR penelakut OR piapot OR piikani OR pikangikum OR pinaymootang OR potlotek OR "Qalipu Mi&apos;kmaq" OR qualicum OR quatsino OR sagamok OR sahgaiehcan OR samahquam OR saugeen OR saulteau OR saulteaux OR sechelt OR secwepemc* OR semiahmoo OR seneca OR shackan OR shamattawa OR shawanaga OR sheguiandah OR sheshegwaning OR shuswap OR siksika OR simpcw OR sioux OR siska OR skatin OR skawahlook OR skeetchestn OR skidegate OR "Skin Tyee" OR skowkale OR skownan OR skuppah OR skwah OR snuneymuxw OR songhees OR soowahlie OR splatsin OR spuzzum OR squamish OR squiala OR "Stswecem&apos;c Xget&apos;tem" OR sumas OR "Ta&apos;an Kwach&apos;an" OR tagish OR tahltan OR takla OR tallcree OR "Taykwa Tagamou" OR "Teetl&apos;it Gwich&apos;in" OR temagami OR thessalon OR timiskaming OR "Tk&apos;eml&#250;ps te Secw&#233;pemc" OR tla-o-qui-aht OR tlatlasikwala OR tlingit OR tlowitsis OR tootinaowaziibeeng OR toquaht OR "Tr&apos;ond&#235;k Hw&#235;ch&apos;in" OR tsartlip OR tsawout OR tsawwassen OR tseshaht OR tseycum OR "Ts&apos;il Kaz Koh" OR tsleil-waututh OR tuscarora OR tutchone OR tzeachten OR uchucklesaht OR ucluelet OR ulkatcho OR wabaseemoong OR wabauskang OR wagmatcook OR wahnapitae OR "Wahpeton Dakota" OR wahsipekuk OR wapekeka OR wasagamack OR wasauksing OR waswanipi OR wawakapewin OR waywayseecappo OR webequie OR weenusk OR wemotaci OR wendat OR "Wha Ti" OR whapmagoostui OR "Whitecap Dakota" OR wikwemikong OR witset OR wolastoqiyik OR wuikinuxv OR wunnumin OR "Wuskwi Sipihk" OR xatsull OR "Xeni Gwet&apos;in" OR yakweakwioose OR yekooche OR "Yellow Quill" OR yellowknife OR yellowknives OR zhiibaahaasing ) ) OR ( TITLE-ABS-KEY ( "Kwakwaka&apos;wakw" OR "Kwak&apos;wala" OR "L&apos;uml&apos;umuluts" OR "Lheidli T&apos;enneh " OR "Nlaka&apos;pamux" OR "Nux&#225;lk" OR "Quw&apos;utsun" OR "Qwum&apos;yiqun" OR "S&apos;amuna" OR "Sm&apos;algyax" OR "St&apos;at&apos;imc" OR "Stl&apos;atl&apos;imx" OR "Sto:lo" OR "Tl&apos;ulpalus" OR "Tsilhqot&apos;in" OR "T&apos;Souke" OR "Xa&apos;xtsa" OR ( labrador W/2 ( indian OR indians OR tribe OR tribes OR native OR natives OR nation OR people* ) ) OR "&#193;a Tlein Kw&#225;an" OR aivilingmiut OR algonkian OR algonquian OR algonquin OR anishinaabe OR "Atlin people" OR "Baffin Island" OR "Bella Coola" OR burrard OR cheklesahht OR chilcotin OR chippew* OR clayoquot OR clemclemaluts OR comiaken OR comox OR coquitlam OR dakelh OR dane-zaa OR "Deisleen Kw&#225;an" OR dunne-za OR gitxsan OR gitxsanimaax OR guyohkohnyo OR "Haad kil" OR halkomelem OR halqemeylem OR henaksiala OR hinupsum OR hunquminum OR hwlitsum OR hwulqwselu OR hydin OR iglulik OR iglulingmuit OR ininew OR inuinnait OR inuk OR inupiat OR inuvialuit OR iroquois OR kainai OR kaska OR kelsemeht OR khenipsen OR kilpahlas OR kimsquit OR kitamaat OR kitlinermiut OR kivallirmiut OR klahoose OR koksilah OR kootenay OR koskimo OR ktunaxa OR kwantlen OR kwatna OR kyuquot OR labradormiut OR laich-kwil-tach OR lamalch* OR lekwungen OR lillooet OR loucheaux OR lummi OR micmac OR mississaugas OR mohawk OR mowachaht-muchalaht OR nadene OR na-dene OR namgis OR nanaimo OR nanoose OR nequatque OR netsilik OR netsilingmiut OR niitsitapi OR nimpkish OR nootka OR nunavimiut OR nunavimmiut OR nuu-chah-nulth OR odawa OR ohiaht OR oji-cree OR onayotekaono OR onondaga OR onondowahgah OR onundagaono OR opetchesaht OR owekeeno OR passamaquoddy OR peigan OR petun OR plains-cree OR popkum OR potawatomi OR quamichan OR sahtu OR salish OR sarcee OR scowlitz OR "Seabird Island" OR secwepemc OR sekani OR semahquam OR shishalh OR sinixt OR ska-ruh-reh OR skway OR sliammon OR snaw-naw-as OR somena OR songish OR sooke OR stoney OR stuie OR tallheo OR tasttine OR teslin OR tlicho OR tsimshianic OR tununirmiut OR ungava OR wakashan OR weewaikai OR wewaykum OR wyandot OR yuculta ) ) OR ( TITLE-ABS-KEY ( ( ( indigenous OR native OR "first nation" OR aborigin* OR indian ) W/2 ( health OR healthcare OR healing OR medicine OR practices ) ) OR ethnomedicin* OR "native healing" OR limpia OR "indigenous healer" OR medicineman OR medicinemen OR "medicine man" OR "medicine men" OR curandero OR "folk healer" OR shaman* OR shamen ) ) ) AND ( TITLE-ABS-KEY ( climacteric OR climacterium OR menopause OR menopaus* OR "Menopause, Premature" OR "climacterium praecox" OR perimenopause OR perimenopaus* OR postmenopause OR postmenopaus* OR "Post menopaus*" OR post-menopaus* OR premenopause OR premenopause* OR pre-menopaus* OR "Pre menopaus*" OR climacteric* OR "Life Change*" OR "Change of life" OR "Hot Flashes" OR "Hot Flash*" OR "Hot Flush*" OR "night sweat*" OR "nocturnal hyperhidrosis" OR "vasomotor symptom*" OR vms ) ) ) OR ( ( ( TITLE-ABS-KEY ( ( ( american* OR alaska* ) W/2 ( native* OR indian OR indians OR indigenous OR aborigin* ) ) OR amerind* ) ) OR ( TITLE-ABS-KEY ( ( ( canada OR canadian* ) W/2 ( native* OR indigenous OR aborigin* ) ) OR metis OR "M&#233;tis" OR michif* OR mitchif OR mechif OR "M&#233;tif" OR "M&#233;tchif" OR metif OR metchif OR inuit OR "First Nations" OR "First Nation" ) ) OR ( TITLE-ABS-KEY ( inuit OR ( arctic W/2 people* ) ) ) OR ( TITLE-ABS-KEY ( "Asa&apos;carsarmiut" OR "Dee-ni&apos;" OR "Mi&apos;kmaq" OR "S&apos;Klallam" OR "Shoonaq&apos;" OR "Sun&apos;aq" OR ( ( "Ivanof Bay" OR "Shoalwater Bay" OR "Spirit Lake" OR "Squaxin Island" OR dakota OR dakotah OR wichita OR keechi OR waco OR tawakonie OR cheyenne OR creek OR crow OR hoonah OR kasigluk OR maricopa OR miami OR omaha OR oneida OR ottawa OR peoria OR salamatof OR sitka OR snoqualmie OR spokane OR winnebago OR wyandotte ) W/2 ( indian OR indians OR tribe OR tribes OR native OR natives OR nation OR people* ) ) OR ( ( native OR indian OR traditional ) W/2 ( village OR council OR community OR association ) ) OR absentee-shawnee OR achomawi OR agdaagux OR "Agua Caliente" OR ajumawi OR ak-chin OR akiachak OR akiak OR alabama-coushatta OR alabama-quassarte OR alatna OR algaaciq OR algin OR allakaket OR "Alturas Indian Rancheria" OR alutiiq OR angoon OR anvik OR apache OR aporige OR aquinnah OR arapaho OR aroostook OR assiniboine OR astarawi OR atsugewi OR atwamsini OR bdewakantunwan OR caddo OR cahto OR cahuilla OR catawba OR cayuga OR chalkyitsik OR cheesh-na OR chehalis OR chemehuevi OR cher-ae OR chevak OR cheyenne-arapaho OR chickahominy OR chickaloon OR chickasaw OR chignik OR chilkat OR chilkoot OR chinik OR chinook OR chippewa* OR chitimacha OR choctaw OR chukchansi OR chuloonawick OR chumash OR cocopah OR colusa OR comanche OR coos OR cosmit OR coushatta OR cowlitz OR "Cross Village" OR cupeno OR curyung OR diegueno OR dine&apos; OR egegik OR eklutna OR elwha OR "Emmonak Village" OR "Evansville Village" OR ewiiaapaayp OR "Galena Village" OR goshute OR "Gulkana Village Council" OR haida OR hammawi OR havasupai OR "Healy Lake Village" OR hewisedawi OR ho-chunk OR hoh OR hokan OR holikachuk OR "Holy Cross Tribe" OR hualapai OR "Hughes Village" OR "Huslia Village" OR "Hydaburg Cooperative Association" OR "Igiugig Village" OR iipay OR ilmawi OR inaja OR inupiat OR ione OR itsatawi OR "Kaguyak Village" OR kaibab OR "Kaktovik Village" OR kalispel OR kanosh OR karuk OR kashia OR kaw OR keetoowah OR "Kenaitze Indian Tribe" OR "Ketchikan Indian Community" OR "Keweenaw Bay" OR kialegee OR kickapoo OR "King Island Native Community" OR "King Salmon Tribe" OR kiowa OR klamath OR "Klawock Cooperative Association" OR kletsel OR "Knik Tribe" OR "Koi Nation" OR "Kokhanok Village" OR koosharem OR kootenai OR kosalektawi OR "Koyukuk Native Village" OR kumeyaay OR kwalhioqua OR "Lesnoi Village" OR "Levelock Village" OR "Lime Village" OR "Louden Village" OR luiseno OR lummi OR madesi OR maidu OR makah OR maliseet OR "Manley Hot Springs Village" OR "Manokotak Village" OR manzanita OR mashantucket OR match-e-be-nash-she-wish OR mattaponi OR "McGrath Native Village" OR mdewakanton OR mechoopda OR menominee OR me-wuk OR miccosukee OR micmacs OR "Mission Indians" OR miwok OR miwuk OR modoc OR mohegan OR mojave OR monacan OR "Mono Indians" OR muckleshoot OR muscogee OR "Na Den&#233;" OR nansemond OR narragansett OR navajo OR "New Koliganek Village" OR "New Stuyahok Village" OR "Newhalen Village" OR "Newtok Village" OR "Nez Perce" OR "Nikolai Village" OR "Ninilchik Village" OR nisqually OR nomlaki OR "Nondalton Village" OR nooksack OR "Northway Village" OR nottawaseppi OR "Nulato Village" OR nunakauyarmiut OR odawa OR ohgsenakale OR "Ohkay Owingeh" OR onondaga OR oreilles OR "Organized Village of Grayling" OR "Organized Village of Kake" OR "Organized Village of Kasaan" OR "Organized Village of Kwethluk" OR "Organized Village of Saxman" OR orutsararmiut OR osage OR otoe-missouria OR paiute OR paiute-shoshone OR pala OR pamunkey OR pascua OR paskenta OR passamaquoddy OR "Pauloff Harbor Village" OR pauma OR pawnee OR pechanga OR "Pechanga " OR "Peck Indian Reservation" OR "Pedro Bay Village" OR pee-posh OR penobscot OR penutian OR perquot OR "Pima People" OR pima-maricopa OR poarch OR pokagon OR pomo OR ponca OR "Portage Creek Village" OR potawatomi OR pottawatomi OR pueblo OR puyallup OR "Qagan Tayagungin" OR qawalangin OR quapaw OR quechan OR "Rampart Village" OR rappahannock OR resighini OR "Sac &amp; Fox" OR "Sac and Fox" OR "Saint George Island" OR "Saint Paul Island" OR salish OR samish OR santee OR sauk-suiattle OR "Seldovia Village Tribe" OR seminole OR seneca OR seneca-cayuga OR shinnecock OR shivwits OR shoshone OR shoshone-bannock OR shoshone-paiute OR siuslaw OR skagit OR "Skagway Village" OR skokomish OR "South Naknek Village" OR "Stebbins Community Association" OR stillaguamish OR suquamish OR swinomish OR "Takotna Village" OR tangirnaq OR tejon OR "Telida Village" OR te-moak OR thlopthlocco OR "Tlingit &amp; Haida" OR "Tohono O&apos;odham" OR tolowa OR tonawanda OR tonkawa OR tulalip OR tunica-biloxi OR tuolumne OR tuscarora OR "Twin Hills Village" OR "Ugashik Village" OR umatilla OR umkumiut OR umpqua OR ute OR "Utu Utu Gwaitu" OR "Village of Alakanuk" OR "Village of Anaktuvuk Pass" OR "Village of Aniak" OR "Village of Atmautluak" OR "Village of Bill Moore&apos;s Slough" OR "Village of Chefornak" OR "Village of Clarks Point" OR "Village of Crooked Creek" OR "Village of Dot Lake" OR "Village of Iliamna" OR "Village of Kalskag" OR "Village of Kaltag" OR "Village of Kotlik" OR "Village of Lower Kalskag" OR "Village of Ohogamiut" OR "Village of Red Devil" OR "Village of Sleetmute" OR "Village of Solomon" OR "Village of Stony River" OR "Village of Venetie" OR "Village of Wainwright" OR wailaki OR wampanoag OR "Warm Springs Reservation" OR washoe OR willapa OR winnemucca OR wintun OR wiyot OR "Wrangell Cooperative Association" OR yakama OR yakutat OR yaqui OR yavapai OR yokuts OR yuhaaviatam OR yuima OR yukian OR yukut OR yupiit OR zuni ) ) OR ( TITLE-ABS-KEY ( "Degexit&apos;an" OR "Yup&apos;ik" OR ( "Bear River" W/2 ( indian OR indians OR tribe OR tribes OR native OR natives OR nation OR people* ) ) OR abenaki OR achumawi OR ahtna OR "Akimel Oodham" OR "Akimel O&apos;odham" OR "Akimel Ootham" OR "Akimel O&apos;otham" OR aleut* OR algonkian* OR alsea OR aluutiq OR apalachee OR arikara OR atakapa OR athabascan OR athabaskan OR athapascan OR athapaskan OR bannock OR bidai OR biloxi OR blackfeet OR blackfoot OR caddoan OR calice OR calusa OR carrizo OR cayuse OR chemakum OR cheraw OR cherokee OR chetco OR cheyenne OR chicora OR chilula OR chimariko OR clatskanie OR coahuiltecan OR "Coeur d&apos;Alene" OR costanoan OR cree OR croatan OR croatoan OR cusabo OR edisto OR eskaleut* OR eskimo OR eskimo-aleut OR esselen OR eyak OR "Fox and Sauk" OR gabrielino OR "Gros Ventre" OR guale OR "Gwich&apos;in" OR halchidhoma OR hidatsa OR hitchiti OR hopi OR houma OR huichol* OR hupa OR ingalik OR inupiaq OR ioway OR iroquoi* OR jacome OR jano OR jeaga OR juaneno OR jumano OR kalapuya OR kansa OR karankawa OR karok OR kashaya OR kato OR kawaiisu OR kitanemuk OR kitsai OR klallam OR koasati OR kolchan OR konkow OR koyukon OR lakota* OR lassik OR laurentian OR lenape OR "Lenni Lenape" OR mahican OR mandan OR maricopa OR massachusett OR mattole OR minisink OR missouria OR mohave OR mohawk OR mohican OR molalla OR multnomah OR munsee OR muscoge* OR naabeeho OR nakoda OR nanticoke OR natchez OR nauset OR navaho OR niantic OR nipmuc OR nisenan OR nongatl OR oconee OR ofo OR ohlone OR ojibw* OR okanagan OR oneida OR otoe OR palouse OR panamint OR papago OR patwin OR "Pee Dee" OR pennacook OR pequot OR pima OR piro OR pocasset OR pocomtuc OR pokanoket OR poospatuck OR powhatan OR quechan OR quileute OR quinault OR quiripi OR salinan OR samich OR saponi OR serrano OR shasta OR shawnee OR siksikaitsitapi OR sinkyone OR siouan OR sioux OR snohomish OR snoqualmie OR squaxin OR suma OR susquehannock OR takelma OR tanacross OR tanana OR tataviam OR tawakoni OR tequesta OR tillamook OR timucua OR tlingit OR "Tohono Oodham" OR tongva OR tsimshian OR tubatulabal OR tutelo OR tututni OR unalachtigo OR unami OR unangan OR unangas OR unkechaug OR waccamaw OR walapai OR wappo OR wasco OR whilkut OR wichita OR winnebago OR wintu OR wishram OR yahi OR yakima OR yamasee OR yana OR yaqu OR yuchi OR yuki OR yuma OR yupik OR yurok ) ) OR ( TITLE-ABS-KEY ( anishnawbek OR "?Akisq&apos;nuk" OR "?aq&apos;am" OR "?Esdilagh" OR "Ab&#233;nakis" OR "Akisq&apos;nuk" OR "aq&apos;am" OR "Che:k:tles7et&apos;h&apos;" OR "Da&apos;naxda&apos;xw" OR "Dzawada&apos;enuxw" OR "Esk&apos;etemc" OR "Gitga&apos;at" OR "Gitlaxt&apos;aamiks" OR "Gwa&apos;Sala-Nakwaxda&apos;xw" OR "Gwich&apos;in" OR "Ka:&apos;yu:&apos;k&apos;t&apos;h&apos;" OR "Kanyen&apos;kehaka" OR "Kapawe&apos;no" OR "K&apos;atlodeeche" OR "K&apos;&#243;moks" OR "Leq&apos;&#225;:mel" OR "Lil&apos;wat" OR "Mal&#233;cite" OR "Mi&apos;kmaq" OR "Mowachaht/Muchalaht" OR "Nisga&apos;a" OR "N&apos;Quatqua" OR "O&apos;Chiese" OR "Ononda&apos;gega&apos;" OR "Onondow&#225;ga&apos;" OR "Onyota&apos;a:ka" OR "Saik&apos;uz" OR "Sambaa K&apos;e " OR "Shxwh&#225;:y" OR "Shxw&apos;ow&apos;hamel" OR "SIPEKNE&apos;KATIK" OR "Skaru:reh" OR "Sq&apos;&#233;wlets" OR "Stellat&apos;en" OR "Sts&apos;ailes" OR "Stz&apos;uminus" OR "T&apos;it&apos;q&apos;et" OR "Tla&apos;amin" OR "Tl&apos;azt&apos;en" OR "Tl&apos;etinqox" OR "Tsal&apos;alh" OR "Tsideldel" OR "Ts&apos;kw&apos;aylaxw" OR "T&apos;Sou-ke" OR "Ts&apos;uubaa-asatx" OR "Tsuut&apos;ina" OR "We&apos;koqma&apos;q" OR "Wet&apos;suwet&apos;en" OR "Xai&apos;xais" OR "Xaxli&apos;p" OR "Yunesit&apos;in" OR ( ( acadia OR carmacks OR champagne OR creek OR delaware OR douglas OR montana OR similkameen ) W/2 ( indian OR indians OR tribe OR tribes OR native OR natives OR nation OR people* ) ) OR aamjiwnaang OR abegweit OR abitibiwinni OR ahousaht OR ahtahkakoop OR aishihik OR aitchelitz OR aklavik OR algonquin* OR anicinape* OR "Animakee Wa Zhing" OR anishina* OR aqam OR atikamekw OR atjanasca* OR attawapiskat OR aundeck-omni-kaning OR batchewana OR "Bearfoot Onondaga" OR beausoleil OR "Behdzi Ahda*" OR "Biigtigong Nishnaabeg" OR "Biinjitiwaabik Zaaging Anishinaabek" OR bilijk OR "Binche Whut&apos;en" OR "Bingwi Neyaashi Anishinaabek" OR "Canupawakpa Dakota" OR cayuga OR chawathil OR cheam OR "Cheslatta Carrier" OR chiniki OR chipew* OR cote OR couchiching OR cowessess OR cowichan OR cree OR "Dakota Plains" OR "Dakota Tipi" OR "Dechi Laot&apos;i" OR deline OR dena OR dene OR denesuline OR "Deninu K&apos;ue" OR ditidaht OR "Dog Rib Rae" OR dokis OR eabametoong OR ehattesaht OR elsipogtog OR ermineskin OR esdilagh OR esgenoopetitj OR eskasoni OR esquimalt OR gameti OR gayogohono OR ginoogaming OR gitanmaax OR gitanyow OR gitsegukla OR gitwangak OR gitxaala OR glooscap OR gwawaenuk OR "Gwichya Gwich&apos;in" OR gwitchin OR hagwilget OR haisla OR halalt OR haudenosaunee OR heiltsuk OR hesquiaht OR hiawatha OR homalco OR hupacasath OR huu-ay-aht OR inninuwug OR innu OR innue OR innus OR inuvik OR iskatewizaagegan OR iskut OR "Ka&apos;a&apos;gee Tu" OR kahkewistahaw OR "Kasabonika Lake" OR kashechewan OR katzie OR kawacatoose OR kebaowek OR keeseekoose OR keeseekoowenin OR kee-way-win OR "Kinistin Saulteaux" OR kinonjeoshtegon OR kispiox OR kitselas OR kitsumkalum OR kluane OR kutchin OR kwadacha OR kwakiutl OR "Kwanlin Dun" OR kwaw-kwaw-apilt OR kwiakah OR "Kwikwasut&apos;inuxw Haxwa&apos;mis" OR kwikwetlem OR "Lax Kw&apos;alaams" OR "Lheidli T&apos;enneh" OR "Liidlii Kue" OR "Listuguj Mi&apos;gmaq" OR lyackson OR magnetawan OR malahat OR maliseet OR mamalilikulla OR matachewan OR matsqui OR mattagami OR "M&apos;Chigeeng" OR membertou OR metepenagiag OR metlakatla OR miawpukek OR michipicoten OR micmacs OR millbrook OR mishkeegogamang OR mississauga* OR mitaanjigamiing OR mohawk* OR montagnais OR moosomin OR moravian OR mowachaht OR muchalaht OR munsee-delaware OR muscowpetung OR muskoday OR muskowekwan OR musqueam OR "Nacho Nyak Dun" OR "Nadleh Whuten" OR naicatchewenin OR "Nak&apos;azdli Whut&apos;en" OR nakota OR naotkamegwanning OR naskapi OR natoaganeg OR nee-tahi-buhn OR nehiyawak OR nekaneet OR neskantaga OR neskonlith OR nibinamik OR nigigoonsiminikaaning OR nipissing OR nishnaabeg OR nooaitch OR nuchatlaht OR nuxalk OR ochapowace OR o-chi-chak-ko-sipi OR odanak OR ojibw* OR okanagan OR okanese OR okemasis OR oneida OR oromocto OR osoyoos OR pabineau OR pacheedaht OR "Paqtnkek Mi&apos;kmaw" OR pasqua OR pauingassi OR pauquachin OR "Pays Plat" OR peguis OR "Pehdzeh Ki" OR pekuakamiulnuatsh OR penelakut OR piapot OR piikani OR pikangikum OR pinaymootang OR potlotek OR "Qalipu Mi&apos;kmaq" OR qualicum OR quatsino OR sagamok OR sahgaiehcan OR samahquam OR saugeen OR saulteau OR saulteaux OR sechelt OR secwepemc* OR semiahmoo OR seneca OR shackan OR shamattawa OR shawanaga OR sheguiandah OR sheshegwaning OR shuswap OR siksika OR simpcw OR sioux OR siska OR skatin OR skawahlook OR skeetchestn OR skidegate OR "Skin Tyee" OR skowkale OR skownan OR skuppah OR skwah OR snuneymuxw OR songhees OR soowahlie OR splatsin OR spuzzum OR squamish OR squiala OR "Stswecem&apos;c Xget&apos;tem" OR sumas OR "Ta&apos;an Kwach&apos;an" OR tagish OR tahltan OR takla OR tallcree OR "Taykwa Tagamou" OR "Teetl&apos;it Gwich&apos;in" OR temagami OR thessalon OR timiskaming OR "Tk&apos;eml&#250;ps te Secw&#233;pemc" OR tla-o-qui-aht OR tlatlasikwala OR tlingit OR tlowitsis OR tootinaowaziibeeng OR toquaht OR "Tr&apos;ond&#235;k Hw&#235;ch&apos;in" OR tsartlip OR tsawout OR tsawwassen OR tseshaht OR tseycum OR "Ts&apos;il Kaz Koh" OR tsleil-waututh OR tuscarora OR tutchone OR tzeachten OR uchucklesaht OR ucluelet OR ulkatcho OR wabaseemoong OR wabauskang OR wagmatcook OR wahnapitae OR "Wahpeton Dakota" OR wahsipekuk OR wapekeka OR wasagamack OR wasauksing OR waswanipi OR wawakapewin OR waywayseecappo OR webequie OR weenusk OR wemotaci OR wendat OR "Wha Ti" OR whapmagoostui OR "Whitecap Dakota" OR wikwemikong OR witset OR wolastoqiyik OR wuikinuxv OR wunnumin OR "Wuskwi Sipihk" OR xatsull OR "Xeni Gwet&apos;in" OR yakweakwioose OR yekooche OR "Yellow Quill" OR yellowknife OR yellowknives OR zhiibaahaasing ) ) OR ( TITLE-ABS-KEY ( "Kwakwaka&apos;wakw" OR "Kwak&apos;wala" OR "L&apos;uml&apos;umuluts" OR "Lheidli T&apos;enneh " OR "Nlaka&apos;pamux" OR "Nux&#225;lk" OR "Quw&apos;utsun" OR "Qwum&apos;yiqun" OR "S&apos;amuna" OR "Sm&apos;algyax" OR "St&apos;at&apos;imc" OR "Stl&apos;atl&apos;imx" OR "Sto:lo" OR "Tl&apos;ulpalus" OR "Tsilhqot&apos;in" OR "T&apos;Souke" OR "Xa&apos;xtsa" OR ( labrador W/2 ( indian OR indians OR tribe OR tribes OR native OR natives OR nation OR people* ) ) OR "&#193;a Tlein Kw&#225;an" OR aivilingmiut OR algonkian OR algonquian OR algonquin OR anishinaabe OR "Atlin people" OR "Baffin Island" OR "Bella Coola" OR burrard OR cheklesahht OR chilcotin OR chippew* OR clayoquot OR clemclemaluts OR comiaken OR comox OR coquitlam OR dakelh OR dane-zaa OR "Deisleen Kw&#225;an" OR dunne-za OR gitxsan OR gitxsanimaax OR guyohkohnyo OR "Haad kil" OR halkomelem OR halqemeylem OR henaksiala OR hinupsum OR hunquminum OR hwlitsum OR hwulqwselu OR hydin OR iglulik OR iglulingmuit OR ininew OR inuinnait OR inuk OR inupiat OR inuvialuit OR iroquois OR kainai OR kaska OR kelsemeht OR khenipsen OR kilpahlas OR kimsquit OR kitamaat OR kitlinermiut OR kivallirmiut OR klahoose OR koksilah OR kootenay OR koskimo OR ktunaxa OR kwantlen OR kwatna OR kyuquot OR labradormiut OR laich-kwil-tach OR lamalch* OR lekwungen OR lillooet OR loucheaux OR lummi OR micmac OR mississaugas OR mohawk OR mowachaht-muchalaht OR nadene OR na-dene OR namgis OR nanaimo OR nanoose OR nequatque OR netsilik OR netsilingmiut OR niitsitapi OR nimpkish OR nootka OR nunavimiut OR nunavimmiut OR nuu-chah-nulth OR odawa OR ohiaht OR oji-cree OR onayotekaono OR onondaga OR onondowahgah OR onundagaono OR opetchesaht OR owekeeno OR passamaquoddy OR peigan OR petun OR plains-cree OR popkum OR potawatomi OR quamichan OR sahtu OR salish OR sarcee OR scowlitz OR "Seabird Island" OR secwepemc OR sekani OR semahquam OR shishalh OR sinixt OR ska-ruh-reh OR skway OR sliammon OR snaw-naw-as OR somena OR songish OR sooke OR stoney OR stuie OR tallheo OR tasttine OR teslin OR tlicho OR tsimshianic OR tununirmiut OR ungava OR wakashan OR weewaikai OR wewaykum OR wyandot OR yuculta ) ) ) AND ( ( TITLE ( "elder" OR "elders" ) OR KEY ( "elder" OR "elders" ) ) AND TITLE-ABS-KEY ( female OR woman OR women ) ) ) OR ( ( TITLE-ABS-KEY ( ( traditional W/2 ( medicine* OR therap* OR healing OR treatment* ) ) OR "folk medicine" OR "folk healer" ) ) AND ( ( TITLE-ABS-KEY ( climacteric OR climacterium OR menopause OR menopaus* OR "Menopause, Premature" OR "climacterium praecox" OR perimenopause OR perimenopaus* OR postmenopause OR postmenopaus* OR "Post menopaus*" OR post-menopaus* OR premenopause OR premenopause* OR pre-menopaus* OR "Pre menopaus*" OR climacteric* OR "Life Change*" OR "Change of life" OR "Hot Flashes" OR "Hot Flash*" OR "Hot Flush*" OR "night sweat*" OR "nocturnal hyperhidrosis" OR "vasomotor symptom*" OR vms ) ) OR ( ( TITLE ( "elder" OR "elders" ) OR KEY ( "elder" OR "elders" ) ) AND TITLE-ABS-KEY ( female OR woman OR women ) ) ) AND ( TITLE-ABS-KEY ( "North America" OR canada OR alberta OR "British Columbia" OR manitoba OR "New Brunswick" OR "Newfoundland" OR "Labrador" OR "Northwest Territories" OR "Nova Scotia" OR nunavut OR ontario OR "Prince Edward Island" OR quebec OR saskatchewan OR yukon OR "United States" OR appalachia* OR "New England" OR "Pacific States" OR alabama OR alaska OR arizona OR arkansas OR california OR colorado OR connecticut OR delaware OR florida OR georgia OR hawaii OR idaho OR illinois OR indiana OR iowa OR kansas OR kentucky OR louisiana OR maine OR maryland OR massachusetts OR michigan OR minnesota OR mississippi OR missouri OR montana OR nebraska OR nevada OR "New Hampshire" OR "New Jersey" OR "New Mexico" OR "New York" OR "North Carolina" OR "North Dakota" OR ohio OR oklahoma OR oregon OR pennsylvania OR "Rhode Island" OR "South Carolina" OR "South Dakota" OR tennessee OR texas OR utah OR vermont OR virginia OR washington OR "West Virginia" OR wisconsin OR wyoming OR "District of Columbia" OR "Washington DC" ) ) ) ) AND ( ORIG-LOAD-DATE > 20230607 )

### Native Health Database (nativehealthdatabase.net/)

**Date:** 21 November 2024

Each line represents a separate search. There were a total of 157 cumulative records found across all searches and reviewed by the team.

| **Search** | **Results 11/21/24** |
| --- | --- |
| Menopause | 11 |
| Menopausal | 4 |
| peri-menopause | 1 |
| perimenopause | 1 |
| pre-menopause | 0 |
| premenopause | 0 |
| post-menopause | 0 |
| postmenopause | 0 |
| post-menopausal | 12 |
| perimneopausal | 1 |
| premneopausal | 3 |
| Life Change | 26 |
| Life Changes | 31 |
| Change of Life | 31 |
| Life transition | 1 |
| female elder | 3 |
| women elder | 7 |
| midlife | 8 |
| Elder | 83 |

| **Record ID** | **Title** | | | | |
| --- | --- | --- | --- | --- | --- |
| #3513 – 2001 | National Inuit Health Information Conference: Inuit defined health information needs and directions | | | | |
| **Author information** | | | | | **Country** |
| Kerrie Duncan; & Inuit Tapiriit Kanatami | | | | | Canada |
| **Study aims** | | | | | |
| “In June 2001, over 40 delegates from across Canada gathered in Inuvik, NWT to learn, discuss, review and advise the Inuit Tapiriit Kanatami (ITK) Health Department on directions required with regard to Health Information and Health Information Initiatives for Inuit.” Page 3.  “The objectives of the conference were: • To address the relative absence of Health Information about Inuit. • To identify key issues for Inuit on the challenges involved in addressing the relative absence of Inuit health information. • To outline resources needed to take action on issues identified during the conference. • To make recommendations on directions, activities, resources and key issues related to Health Information. • To make recommendations on Inuit participation in ongoing activities.” Page 3. | | | | | |
| **Study design** | **Conflicts of interest** | | **Inclusion criteria** | | **Exclusion criteria** |
| Other: White paper. | N/A; none listed. | | N/A; this was not a study. | | N/A. |
| **Population and sampling description** | | **Indigenous description** | | **Total number of participants** | |
| Inuit peoples were the focus of this white paper. | | Inuit peoples. | | N/A. | |
| **Study outcomes and important results** | | | | | |
| Under the “Women’s perspective” section of the paper, there is a table called “Health Issues for Women” that included the bullet “Life Changes — Menopause.” These Health Issues for Women were outlined by Donna Kisoun and Reepa Evic-Carleton, who were representatives of Pauktuutit, Inuit Women’s Organization. They began their presentation by stating it is important to consult with all sectors of society when identifying priorities and setting health research priorities—including Inuit women. Page 9. | | | | | |
| **Use of STRAW+10** | **Menopausal transition staging of sample using STRAW+10** | | | | |
| The STRAW+10 not used. | N/A. | | | | |
| **Author reported facilitators to treatment** | | | **Author reported barriers to treatment** | | |
| Collecting data about culture and community to tailor healthcare and public-health planning: “What questions are most important to ask in an Inuit context? How can the health planning process be more responsive to Inuit health priorities? There are unique situations and needs in the North that require Inuit participation to ensure the information collected is true and meaningful. Until now, the elements of Inuit culture that Inuit use to determine health have not been considered. Inuit have the right to access health services equal to other Canadians. This includes the right to define and develop methods that respond to their cultural and community needs, as well as the right to holistic treatment and preventative services. Information collection and management can help to facilitate responses to ensure these rights are met.” Pages 6 and 7.  “Data is meaningful when it makes sense to the people who provided it. When the data makes sense to them, we know it is true, because it matches their reality. Data would have more  meaning if Inuit designed, collected and interpreted their own research. Information is only valuable when we know about it. An ongoing commitment to inform communities about current knowledge must be part of a complete approach.” Page 7.  “Treatment centers must better complement Inuit concepts of healthcare.” Page 13.  Incorporating Indigenous knowledge into healthcare: “Incorporating Inuit traditional knowledge into health treatment and healing programs is both economic, effective and accessible. There are Elders in every community who could teach us how Inuit healed themselves before the Southern physicians came with their Aspirins and bandages. —Working Group Discussion” Page 16. | | | Language as a barrier to healthcare: “Language is always a major barrier to effective healthcare and data maintenance. Diversity of Inuktitut dialects makes communication difficult within each region and community. Information and reports from the governments should be translated, not only Inuktitut syllabics, but all Inuktitut dialects, including Inuvialuktun, Inuinnaqtun and Labradorian. Information and government reports should also be written in plain, easily understood language. Regular training should be made available to all Inuit interpreter/translators in the field to update and standardize clinical, medical and technical terminology.” Page 13.  Care that isn’t patient centered and culturally grounded: “Inuit need the self-confidence to seek treatment and make themselves clearly understood. They often assume their doctor’s diagnosis and prescribed treatment is correct even if they know they haven’t communicated their symptoms properly. A lot of the time, especially the Elders will just accept the treatment without arguing, even though they know it might not be the right one. —Working Group Discussion. Cultural awareness workshops should be mandatory orientation for healthcare workers providing service for Inuit both north and south. Ensure Inuit are informed of their individual rights within the healthcare system. Ensure ongoing training for existing personnel in the communities regarding medical procedures and treatments. Inuit patients should be provided with better explanations of prescribed treatments and procedures. Encourage new healthcare professionals to recognize participation in community life is part of a positive and holistic approach to healthcare. Describe and develop a holistic approach to healthcare that reflects Inuit Traditional Knowledge.” Page 15. | | |
| **Study limitations** | | | | | |
| N/A; none described. | | | | | |

| **Record ID** | **Title** | | | | |
| --- | --- | --- | --- | --- | --- |
| #3341 – 2000 | Strategies to encourage hormone replacement therapy use in minority or diverse populations: A consensus opinion | | | | |
| **Author information** | | | | | **Country** |
| Thomas, M. A.; Kessel, B.; Miles, T.; Owens, O. M.; Rice, V. M.; Romaguera, J.; Shulman, L.; Thornton, K. L.; Maclin, V.; Walton, E. W.; & Rebar, R. W. | | | | | United States |
| **Study aims** | | | | | |
| “Objective. Hormone replacement therapy (HRT) has the potential to provide benefits for minority women (African American, Asian American, Hispanic American, and Native American) as it does for Caucasian women as they approach menopause and beyond. When HRT is indicated, women in minority groups should be encouraged to start to become educated in strategies to continue this medication for all of the important preventive medical benefits (prevention of osteoporosis, cardiovascular events, and cognitive dysfunction). Design. A group of experts convened to write a consensus opinion on how to achieve a long-term compliance of HRT use during menopause in minority and diverse populations. The topics discussed included whether minority women are not prescribed HRT as often as white women, and why minorities who are prescribed HRT discontinue this medication. This panel also made recommendations to help minority women achieve long-term continuation as well as how to increase participation in clinical trials and improve education for these women and the physicians who care for them.” Abstract. | | | | | |
| **Study design** | **Conflicts of interest** | | **Inclusion criteria** | | **Exclusion criteria** |
| Text and opinion. | N/A; none listed. | | N/A; this was not a study. | | N/A. |
| **Population and sampling description** | | **Indigenous description** | | **Total number of participants** | |
| The populations of interest for this consensus opinion paper were: African American, Asian American, Hispanic American, and Native American.  No participants were sampled because this is not primary research. | | Native American.  The authors do not discuss how they would define the population of people who have a Native American racial/ethnic background. Because this is a consensus opinion paper and not primary research, this record does not include a sample of Native American participants. | | N/A. | |
| **Study outcomes and important results** | | | | | |
| “Results. On the basis of current knowledge, the experts established the following recommendations for helping minority women achieve long-term continuation of HRT: (1) include minority women in clinical trials of HRT, (2) educate physicians and other health care providers on the myths and misconceptions about HRT use by minorities, (3) involve the woman in the decision-making process, (4) explain benefits and risks with clarity and personalize them, (5) clarify the woman’s beliefs and preferences early in decision making, (6) provide educational information that the woman can understand, and (7) follow up with the woman after HRT has been prescribed. Although these recommendations are similar to those for all women after menopause, the implementation of these recommendations needs to be culturally specific.” Abstract. | | | | | |
| **Use of STRAW+10** | **Menopausal transition staging of sample using STRAW+10** | | | | |
| The STRAW+10 not used. | N/A. There is no study sample, as this is a consensus opinion paper. However, the authors do state they are focused on HRT use “during menopause.” | | | | |
| **Author reported facilitators to treatment** | | | **Author reported barriers to treatment** | | |
| N/A. | | | N/A. | | |
| **Study limitations** | | | | | |
| N/A; none described. | | | | | |

| **Record ID** | **Title** | | | | |
| --- | --- | --- | --- | --- | --- |
| #2621 – 1999 | The effect of traditionality on menopausal symptoms | | | | |
| **Author information** | | | | | **Country** |
| Morgan, E. A.; Herman, C. J.; Coriz, M.; Whalawitsa, M.; & Mingo, C. | | | | | United States |
| **Extraction note** | | | | | |
| This is a conference abstract published in the Journal of Investigative Medicine for the Western Student Medical Research Forum, Oral Session III on Thursday, January 28th, 1999. | | | | | |
| **Study aims** | | | | | |
| “Purpose: To better understand Navajo women’s experience of and attitudes toward menopause in order to develop education materials for women and their health care providers.” | | | | | |
| **Study design** | **Conflicts of interest** | | **Inclusion criteria** | | **Exclusion criteria** |
| Qualitative research. | N/A; none listed. | | N/A; the conference abstract does not list inclusion or exclusion criteria. | | N/A. |
| **Population and sampling description** | | **Indigenous description** | | **Total number of participants** | |
| The population of interest was Navajo women.  The conference abstract lists no sampling details. | | The participants were Navajo women; the focus groups “were conducted throughout the Eastern Navajo Nation”.  The conference abstract does not describe how the racial/ethnic background of Navajo was defined.  The percentage of the sample who were Indigenous was 100%. | | The total number of participants is 57 and the total number of Indigenous participants is 57. | |
| **Study outcomes and important results** | | | | | |
| “Overall 57 women participated in the focus groups. Data were analyzed using QRS Nud*ist software from the 26 women who were near the age of menopause or older. The mean age of the participants was 61 years. 49.1% of the women were married. Those women who lived a more traditional lifestyle (n=15) were less likely to report menopausal symptoms. In addition, those women who had kinaalda did not report symptoms of menopause as they grew older.” | | | | | |
| **Use of STRAW+10** | **Menopausal transition staging of sample using STRAW+10** | | | | |
| The STRAW+10 not used. | N/A; this cannot be determined as no details about menopause status are listed in the conference abstract. | | | | |
| **Author reported facilitators to treatment** | | | **Author reported barriers to treatment** | | |
| “Sharing information and supporting traditional ways may assist women in their transition through menopause.” | | | N/A. | | |
| **Study limitations** | | | | | |
| N/A; none described in the conference abstract. | | | | | |

| **Record ID** | **Title** | | | | |
| --- | --- | --- | --- | --- | --- |
| #2393 – 2012 | Black cohosh: Coming full circle? | | | | |
| **Author information** | | | | | **Country** |
| Johnson, T. L.; & Fahey, J. W. | | | | | United States |
| **Study aims** | | | | | |
| “To highlight black cohosh as an example in which disregard for the ethnobotanical and ethnopharmacologic usages of a plant has perhaps hindered modern scientific attempts to understand the mechanism of action of its bioactive phytochemical, and ascribe cause to effect.” Abstract. | | | | | |
| **Study design** | **Conflicts of interest** | | **Inclusion criteria** | | **Exclusion criteria** |
| Other: narrative review. | N/A; none listed. | | N/A; this is not a study. | | N/A. |
| **Population and sampling description** | | **Indigenous description** | | **Total number of participants** | |
| N/A; this is a narrative review of the plant black cohosh. | | North American indigenous people. | | N/A. | |
| **Study outcomes and important results** | | | | | |
| “Anecdotally, cohosh is believed to be an Algonquian word for pregnancy (Gladstar and Hirsch, 2000).” Page 776.  “North American indigenous people’s treatment of the herb was somewhat more pragmatic, however, referring to it simply as “squaw root,” a descriptor more in keeping with its preferential application as a treatment for gynecological conditions such as pain associated with premenstrual syndrome (PMS), pain during childbirth, and menopausal complaints, including migraine (Lighthall, 1883).” Page 776. Reference: Lighthall, J. I., 1883. The Indian household medical guide. Kissinger, Whitefish. | | | | | |
| **Use of STRAW+10** | **Menopausal transition staging of sample using STRAW+10** | | | | |
| The STRAW+10 not used. | N/A; narrative review. | | | | |
| **Author reported facilitators to treatment** | | | **Author reported barriers to treatment** | | |
| N/A. | | | N/A. | | |
| **Study limitations** | | | | | |
| N/A; none described. | | | | | |

| **Record ID** | **Title** | | | | |
| --- | --- | --- | --- | --- | --- |
| #2155 – 1980 | Cultural variability in the experience of menopause: A comparison of Navajo and Western data | | | | |
| **Author information** | | | | | **Country** |
| Wright, A. L. | | | | | United States |
| **Study aims** | | | | | |
| “Despite its universality, it has been neglected by social scientists as a topic of study. This dissertation aims to partially fill this data gap in two ways. First, it contributes to our cross-cultural knowledge of this phenomenon by describing the menopausal experiences of Navajo women. Its second goal is to test hypotheses regarding the relationship between certain cultural factors and individual and cultural variability in the experience of menopausal symptoms.” Abstract.  “This dissertation, then, attempts to consider all aspects of the experience of menopause, from symptomatology to its overall signifi­cance in a particular culture, that of the Navajo. ... A second section then broadens the focus, by comparing these data with those reported for other cultures.” Page 5.  “Given the paucity of ethnographic data concerning menopause, the present project was designed with two aims. First, data were collected to provide a thorough description of the experience of menopause among the Navajo, the first such description for any non-European group. The second goal was to test and evaluate hypotheses which purport to account for variability in the experience of menopausal symptoms in an attempt to explain the observed variability. Hypotheses regarding variability were to be tested at two levels, the individual and the group.” Pages 23 and 24.  “In response, the present project was designed to: (1) describe the experience of menopause for a non-Western society; and (2) test hypotheses regarding individual and cultural variability in the incidence of symptoms in an attempt to isolate contributing factors. Descriptive interests included all aspects of the menopausal experience: the culturally-specific conception of menopause, symptom experience, individual attitudes toward menopause and life changes. For the second goal, factors which had previously been suggested as influencing the experience of symptoms were formulated as hypotheses and tested for relationship with symptom experience for Navajo women.” Page 25. | | | | | |
| **Study design** | **Conflicts of interest** | | **Inclusion criteria** | | **Exclusion criteria** |
| Other: Mixed methods dissertation; exploratory sequential (participant interviews preceded the development of a questionnaire). | N/A; none listed. | | To categorize participants into either the “traditional” or “acculturated” groups, “Residence”, “Education”, “Income”, and “Language” were considered. Inclusion criteria for the “traditional” group included rural residence, no education, income from welfare or sheep herding, and Navajo only language. Inclusion criteria for the “acculturated” group included residence in a small town, education at the level of high school or more, income from wage-labor employment, and Navajo and English language. | | N/A; none listed. |
| **Population and sampling description** | | **Indigenous description** | | **Total number of participants** | |
| The population of interest was middle-aged Navajo women.  “An attempt was made to interview all Kit Silee women known to be between the ages of 40-60. Only those who refused cooperation or who were repeatedly unavailable were not interviewed.” Page 31.  “[‘Acculturated’] women were recruited by asking acquaintances in Chinle and respondents for the names of working women between the ages of 40-60. The ‘acculturated’ sample is not, therefore, random with respect to friendship networks, a factor unlikely to affect the results in any way.” Page 32. | | Middle-aged Navajo women.  The author does not describe how “Navajo” was defined. The author recruited from Navajo reservation areas. Additionally, the author states: “Seventeen clan affiliations were reported by the 80 women. Some clans were represented only among the acculturated women, including Ta'neeszahnii, Tótsohnii, Kiyaa'áanii and Tábaaha. Others, such as Honágháahnii and Dibé łízhiní, were found only among the traditional women. More than half (24) of the traditional women were members of three clans: Tó'ahani (14), Áshįįhí (5) and Honágháahnii (5). In contrast, the acculturated women were spread out over the spectrum of clans, with the largest number (7 each) aligned with Tó'dích'íín'ii or Tó'aheedlíinii clans.” Page 59.  The percentage of the sample who were Indigenous is 100%.  “The women were of two types: “traditional” and “acculturated.” The former were uneducated, spoke no English, lived in a rural area and were not employed in the wage economy. “Acculturated” women, on the other hand, held wage paying jobs, spoke English, had completed high school and lived in towns on the reservation.” From the abstract  Interviews among “traditional participants” were conducted in Navajo with the aid of a very capable female interpreter, whereas “acculturated” respondents were interviewed in English.” Page 31.  “The ‘traditional’ group consisted of 40 women from the Kit Silee area of Black Mesa (Kit Silee, Tachee, Rough Rock and Pinon).” Page 31.  “This first sample was matched with a second, or ‘acculturated’ group of 40 respondents. These women lived in nearby wage settlements (Chinle, Many Farms, and Pinon), areas selected because funds for transportation were limited. Two criteria, education and employment, were used to isolate the ‘acculturated’ sample, factors which assured that the two groups differed substantially in many respects." Page 31.  “Two inferences should not be made regarding the two samples. First, the ‘traditional’ sample is not simply a throwback to a previous time. Rather, it represents a modern adaptation to the influence (among other things) of Anglo ideas which increasingly filter out to the more isolated areas of the reservation. Likewise, the ‘acculturated’ group represents not an intermediate stage blending ‘traditional’ Navajo and Anglo customs (which are themselves tremendously varied), but a unique adaptation founded on a specific historical process. Secondly, there is no modal ‘traditional’ or ‘acculturated’ community. The reservation with its varied lifestyles contains many such ‘traditional’ and ‘acculturated’ areas, each of which differs in some respects from those chosen here. Thus, these glosses gain significance only after the differences and similarities between the samples are carefully considered” Page 34. | | The original “field sample” size was 80. However, symptoms were only analyzed for women who had not undergone surgical menopause. Therefore, the total sample size was 66 and the total number of Indigenous participants was 66 (38 “traditional” and 28 “acculturated”). Page 37. | |
| **Study outcomes and important results** | | | | | |
| “It became apparent that ‘traditional’ Navajo respondents viewed this event differently than do Anglos, as a consequence of lack of communication and the few role changes which take place at this time. They tend to emphasize the end of fertility rather than the cessation of menstruation, and associate ‘the change’ with achievement of grandparenthood. It was also demonstrated that Navajo women suffer from the same symptoms in roughly the same proportion as Anglo women. As to the second goal, it was predicted that ‘traditional’ Navajo women would experience fewer symptoms as a consequence of: (1) retention of a childrearing role; (2) greater integration into kin networks and community; (3) more positive attitudes toward menopause; and (4) access to high status roles at menopause. This prediction was not fulfilled. Subsequently, each of these factors was tested for relationship to individual variability in symptom experience. Different patterns emerged for the two groups. ‘Traditional’ women who complained of poor health and poverty were more likely to experience more symptoms, whereas ‘acculturated’ women reporting negative attitudes suffered the most menopausal symptoms. While these results are somewhat ambiguous, they suggest that the experience of menopause may differ cross-culturally and that social factors influence the experience of menopausal symptoms.” Abstract  “Choice of the treatment sought for menopause related symptoms also differed substantially for the samples. Most (9) of the acculturated women who sought medical help for their menopausal complaints went to doctors or nurses at the clinic, and received advice, aspirin, tranquilizers and estrogens for their pains (Table 18). One of these women (A13) subsequently went to a hataałii when her complaints were unresponsive to Anglo medicines. This woman, who suffered extensively from menopausal symptoms, had one five day (Lightning) ceremony, an unspecified two day ceremony, a Blessingway and several peyote meetings, in addition to regular injections of estrogens.” Page 62 (Note for the meaning of the word hataałii: “a medicine woman” from page 99).  Table 18. “Type of treatment sought for menopausal symptoms, by sample” on page 63 categorizes women by acculturated and traditional and then by their choice of “Medical System.” For the participants categorized as acculturated, 19 sought no treatment, 8 sought “Anglo Only” treatment, 0 sought “Navajo Only” treatment, and 1 sought “Both” (n=28). For the participants categorized as traditional, 22 sought no treatment, 8 sought “Anglo Only” treatment, 4 sought “Navajo Only” treatment, and 4 sought “Both” (n=38).  “Traditional women were more inclined to seek the help of native healers. Eight women went to the clinic only, four to traditional Navajo healers only and in four cases, traditional women sought help from both medical systems. These women reported receiving pain pills, aspirin and other unidentified pills from the clinic. Treatment by traditional healers were much more varied and elaborate. Five women attended peyote meetings for their symptoms. In one case (T19) a Hopi stargazer sucked pebbles from a foot which were causing leg aches. A Blessingway ceremony was prescribed for one woman’s (T33) symptoms, attributed to violation of a taboo. All but one woman (T29), who had sat for four days through a Red Antway ceremony for her leg aches, felt that the ceremonies provided relief from their symptoms. This lack of a specific ceremony for menopause in the traditional Navajo healing system reflects the accurate perception that “menopausal” symptoms are not unique to that state. Therefore, depending on the symptoms a woman suffers from and discussions of the illness (Morgan 1977), the cause will be diagnosed and the appropriate ceremony will be performed.” Page 64.  “Menopause. Data from several sources leave the impression that menopause is not an elaborated concept for traditional Navajo. While most of these women know that menopause occurs, the only major change associated with this event is the end of fertility. The more restricted traditional concept appears to be a consequence of two factors, patterns of communication and types of role changes, each of which is discussed in greater detail below. The first factor is that communication about menopause among traditional respondents is rare. While communication about sexual matters is generally restricted, the lack of a specific word for ‘menopause’ in Navajo (my interpreter asked her questions in terms of ‘the end of the thing the ladies get,’ or the end of menstrual periods) makes reference to this event somewhat difficult, and tends to inhibit communication. ... Limited communication results in each woman knowing only her own reactions, both physical and attitudinal, to menopause, thus impeding the creation and sharing of a unified cultural concept of menopause.” Pages 94 and 96.  “Almost half (18) of the traditional women had never discussed this topic with anyone. (This discussion excludes communication with husbands. All ten traditional women who were married and post-menopausal claimed their husbands ‘knew’ they had stopped menstruating, while 8 of 12 comparable acculturated women had told their husbands.) As one such respondent (T22) noted: ‘We’re bashful about such things.’ Virtually all of those who discussed menopause did so with female relatives, particularly mothers (10) and sisters (6). Age at which menopause occurs was the most common topic discussed (7). ... ‘They say that with the end of the periods, you’re through with having children’ (T37). Five traditional respondents also mentioned that menopause is the beginning of health problems. Commonly the problems were unspecified, however, one woman (T27) referred to aches and pains in the joints, while another (T35) mentioned heavy menstrual flow. When specifically asked if they had ever heard of women experiencing problems with menopause, only five answered in the affirmative. These women had heard of ‘fever,’ aches and pains, and blood storing up in the body.” Pages 96 and 97.  “Acculturated women were much more likely to have talked with someone about menopause — only eight had not. They also exhibited a different pattern concerning with whom and about what they talked. Mothers and friends were equally likely to be sources of information (15 responses each), while nine acculturated women mentioned discussing aspects of menopause with sisters. Doctors, nurses, patients or health workers were consulted in seven cases, as compared with only twice among traditional women.” Page 97.  “Topics were likewise different. Discussion of symptoms was common (16 cases). Psychological symptoms, including moodiness, depression and ‘craziness,’ were mentioned by 7 respondents, while only 4 mentioned hot flashes. Other symptoms mentioned included period problems (2) and physical symptoms (2), especially aches and pains. Age was also a popular topic, however, only rarely was menopause associated with old age. Only 11 acculturated women had never heard of women experiencing problems with menopause. Nine had heard of flushing, 5 of psychological problems and another 5 specified painful and frequent periods.” Pages 97 and 98.  “A second factor contributes to the lack of an elaborated concept of menopause among traditional women: this event signifies relatively few changes, only two of any magnitude, for traditional Navajo women. Obviously, it does mark the end of the ability to conceive. Aside from the end of this reproductive role, which is generally viewed simply as a stage in the life cycle, a traditional woman’s major roles remain the same after menopause. The childbearing function, particularly, is unaltered. Because most traditional women produce children until late in their reproductive lives, there are still young children around at this time who need attention. Further, residence in multi-family camps where siblings’ children and, more importantly, a woman’s own grandchildren abound, promotes retention of childrearing functions until late in life. Thus, traditional women are unlikely to face the ‘empty nest,’ which is characteristic of some industrial societies (Spence and Lonner 1971) and often coincides with menopause.” Page 98.  “Secondly, a woman’s normal productive activities, such as sheepherding and weaving, are unaffected by menopause. They, like the domestic activities of cooking and cleaning, continue beyond menopause until the woman is physically unable to pursue them. Thus, an older Navajo woman continues to contribute both food and labor to her family. Finally, experience is highly valued among traditional Navajos. Consequently, both women and men gain power as they age, and the woman’s role in decision making may grow as her experience is increasingly respected.” Pages 98 and 99.  “The second change menopause renders is the end of menstruation and consequently menstrual taboos. For Navajo women who observe the taboos, this commonly represents the end of a great burden. However, for a few women this change is especially important because is presents the opportunity to become a medicine woman, or hataałii. With the end of menstruation, a woman is freed from the ceremonial taboo which would normally interrupt her pursuit of this elaborate ritual-medical knowledge.” Page 99.  “In sum, for traditional Navajo women excepting those who become hataałii, menopause signifies few role changes. Acculturated women, on the other hand, face a greater change. They tend to have fewer children and to complete their families earlier (Kunitz 1976). Thus, like Anglo women, whose developmental cycle is similar, they are faced with children growing up and leaving home at roughly the time they reach menopause. Residence in single family dwellings increases their isolation from other people’s children. Thus, the loss of the childbearing function becomes even more obvious because the child-rearing role is simultaneously terminated.” Page 100.  “What, then, is the traditional Navajo interpretation of menopause, given patterns of communication and role changes which differ from those reported for acculturated women? As implied above, many traditional respondents associated the bearing of children with menstruation, with pregnancy involving the transformation of menstrual blood into a baby. The relevance of this point here is that menopause is defined among the Navajo, not with reference to menstruation as it appears to be in our society, but rather in terms of the inability to bear more children. ‘It just happens that we were made like that. You have children, then you stop. This is what the old folks say: that you get periods to have babies and there are no more periods after you stop having babies’ (T32).” Page 101.  “The cultural explanation of the relationship between menopause and the end of fertility was obtained from a female hataałii. Women are created with the ability to bear children by means of impregnated menstruation. By the time they become 45-50, a woman has borne numerous children, and her daughter is already having her own babies. The woman no longer needs to produce children: the childbearing role has been passed on to her daughter. Menstruation, which is no longer necessary, is also terminated. Thus, menopause, as the end of the fertile years, is associated with becoming a grandmother. While she recognizes that some women continue bearing children after becoming grandmothers, this is considered an exception which is due to unusual strength and good health.” Page 101.  “How then does this different emphasis, relating menopause to the achievement of grandparenthood and the end of fertility, affect attitudes toward menopause? Of obvious importance here are attitudes toward old age. Traditional reactions toward this stage in the life cycle were mixed. Of the 25 women who expressed an opinion on the topic, half expressed positive sentiments: Lots of times we pray to live until we are old, so it is good to reach that age, not to die before your time (T37). One respondent (T29) stated that ‘life begins when you get older.’ On the other hand, nine women voiced fears of being disabled, with particular concern for how they would manage and who would care for them: I wonder about it /old age/ alot. Like who’s going to care for me? Age is as bad as being disabled, like being sick all the time (T15).” Page 102.  From Table 33. “Expectation of changes at menopause, by sample.” on page 104, the author categorizes responses from the traditional and acculturated participant groups into “None” and “Some.” And, among those who expected changes, the author categorizes the “Symptoms mentioned” by participant group. Twenty-one participants of the traditional group reported “None” for expectations of change at menopause, while 12 of the acculturated group reported “None.” Six participants of the traditional group reported “Some” for expectations of change at menopause, while 16 of the acculturated group reported “Some.” Of the participants who reported expecting some changes at menopause, the symptoms mentioned by the traditional group included: “Worry, aches and pains, sickness.” The symptoms mentioned by the acculturated group who expected some changes at menopause included: “Flushes or sweats, fatigue, aches and pains, nervousness, depression, sickness, weight gain, dizziness.”  “One doctor noted that he had seen approximately five patients, out of 500-1000 women aged 40-60 that year, who reported what appeared to be menopausal complaints. Another Navajo nurse noted that ‘you never see the symptoms as a whole package.’ Rather, women complain of one or two symptoms which they themselves do not recognize are related to menopause and thus may go unreported. A second Navajo nurse, presently menopausal herself, estimated that only 20% of 40-60 year old Navajo women experience the symptoms, with hot flushes, depression and lethargy being the most common complaints. She believes that the dearth of menopausal complaints among traditional women resulted from their greater tolerance of symptoms. It is more likely that the restricted concept, including the lack of association between symptoms and menopause, influences the likelihood of reporting symptoms to health professionals.” Page 105.  “Three major points have emerged which may be relevant to the experience of menopause and its symptoms as discussed in the next chapter. First, the heralding of menarche in the Kinaalda clearly proclaims that the attainment of womanhood is a highly valued achievement. High fertility has been, until recently, adaptive and motherhood remains a major symbol of life and relations between kinspeople. Secondly, menstrual blood itself, while symbolizing fertility, is also considered dangerous. The simultaneous existence of these two beliefs might contribute to conflicting attitudes toward menopause. Finally, menopause itself is approached with less trepidation than it is in our society. Instead, it is viewed as simply the next stage in the reproductive cycle, one which may be accompanied by positive status changes: the termination of menstrual taboos, the achievement of old age, and the possible access to the role of hataałii.” Page 108.  Table 35. “Median age at menopause, by sample.” on page 114 categorizes the acculturated and traditional participant groups into “Currently Menopausal” and “At Menopause (Post-menopausal)”. For the acculturated group, the median age for the “Currently Menopausal” group was 49 (n=5), and the median age “At Menopause” was 50 (n=8). For the traditional group, the median age for the “Currently Menopausal” group was 48 (n=7), and the median age “At Menopause” was 49 (n=16).  “One of the major findings is that the issues related to experience of menopausal symptoms are much more complex than initial work suggests. Now, it appears unlikely that one culture will experience significantly more or less symptoms on the basis of gross social differences. Statements regarding these social differences are clearly simplistic, and leave many potentially relevant variables beyond the control of the researcher. For example, in the present case, health emerged as an important variable in predicting symptom experience although it was not one of the original hypotheses.” Page 184.  “In sum, the basic assumption under which this study was conducted, namely that menopause, like other human events, is a bio-cultural experience, has been supported. It was found that menopausal symptoms of the type reported for European women do exist in other cultures, supporting a physiological relationship with hormone production. However, cultural factors apparently influence this experience, as may genetic and other environmental factors.” Page 186. | | | | | |
| **Use of STRAW+10** | **Menopausal transition staging of sample using STRAW+10** | | | | |
| The STRAW+10 not used. | Reproductive, Menopausal Transition, and Postmenopause.  “An attempt was made to achieve a similar breakdown according to menopausal status (pre-, currently or post-menopausal)” by “traditional” or “acculturated.” Page 32.  Table 34. “Menopausal status, by sample.” on page 112 sorts acculturated and traditional groups into the menopausal statuses of “Pre-”, “Currently”, “Post-1-5 yrs.”, and, “Post-5+ yrs.”. For the acculturated group, 15 reported they were “Pre-”, 5 were “Currently”, 4 were “Post-1-5 yrs.”, and 4 were “Post-5+ yrs.” (n=28). For the traditional group, 15 reported they were “Pre-”, 7 were “Currently”, 9 were “Post-1-5 yrs.”, and 7 were “Post-5+ yrs.” (n=38). | | | | |
| **Author reported facilitators to treatment** | | | **Author reported barriers to treatment** | | |
| N/A. | | | N/A. | | |
| **Study limitations** | | | | | |
| N/A; none described. | | | | | |

| **Record ID** | **Title** | | | | |
| --- | --- | --- | --- | --- | --- |
| #2101 – 1997 | The elder female: A preventative care plan and health watch | | | | |
| **Author information** | | | | | **Country** |
| Attico, N. B. | | | | | United States |
| **Study aims** | | | | | |
| “In this article, comments will be specifically directed towards the female elder, looking at diseases or conditions that have a predilection to attack the female at different rates than the male, or those that are predetermined by female anatomy.” Page 80. | | | | | |
| **Study design** | **Conflicts of interest** | | **Inclusion criteria** | | **Exclusion criteria** |
| Text and opinion. | N/A; none listed. | | N/A; this is not a study. | | N/A. |
| **Population and sampling description** | | **Indigenous description** | | **Total number of participants** | |
| The population of interest was elder American Indian/Alaska Native females.  The author defines the population of interest in Table 1. Population data, elder age groups, American Indians/Alaska Natives. “This table was derived from IHS service population data, based upon the 1994 revision of the 1990 census data. Multiple population age values were calculated because of lack of a standard definition for ‘elder,’ varying from age 50 years (American Association of Retired Persons) through age 65 years (Social Security Administration). There are grants that tribes may become eligible for based upon ages 60 or 65, and some studies may collect data based upon ages 55 and above. Most of the health recommendations in this paper are based upon age 60 years. Regardless of the age criterion used, females exceed males in absolute numbers by 25-30%.” Page 80. | | American Indians/Alaska Natives. | | N/A. | |
| **Study outcomes and important results** | | | | | |
| “As the Indian Health Service presently lacks bone density measurement equipment, there are no IHS data or studies on post­menopausal bone loss in Indian women, but specific data may be forthcoming from the Women’s Health Initiative (WHI), a 12-year, multi-institutional, National Institutes of Health (NIH) study presently underway nationwide in postmenopausal women. At least two of the research institutions in the WHI are actively recruiting Indian women for participation in the study. It is known that fat tissue metabolizes estrogen differently than lean tissue, mostly via an estrone (E1) rather than estradiol (E2) pathway, with estrone being a more potent estrogen in many of its features. There is a clinical impression that Indian women do not develop osteoporosis; Is this true? And, if so, is the high incidence of obesity in Indian women a reason? Do Indian women have higher or more effective levels of postmenopausal estrogen and/or androgen? Do the generally lower rates of smoking in this population have any influence? Answers to these questions, and others, are expected from the WHI. In any event, until more is known, most clinicians today advise the use of estrogen in all postmenopausal women, including Indian women, for the known cardioprotective and osteoprotective features.” Page 83. | | | | | |
| **Use of STRAW+10** | **Menopausal transition staging of sample using STRAW+10** | | | | |
| The STRAW+10 not used. | N/A. | | | | |
| **Author reported facilitators to treatment** | | | **Author reported barriers to treatment** | | |
| N/A. | | | N/A. | | |
| **Study limitations** | | | | | |
| N/A; none described. | | | | | |

| **Record ID** | **Title** | | | | |
| --- | --- | --- | --- | --- | --- |
| #2051 – 2004 | Grandmothers’ Voices: Mi'kmaq Women and Menopause | | | | |
| **Author information** | | | | | **Country** |
| Loppie, C. J. | | | | | Canada |
| **Study aims** | | | | | |
| “The purpose of this naturalistic inquiry was to explore the mid-life experiences of Mi'kmaq women.” Abstract  “The purpose of this study is to explore Mi'kmaq women’s perceptions of mid-life health, with particular emphasis on menopause. Specifically, through this study, I have attempted to answer the following questions: 1. What issues become most important to Mi'kmaq women during midlife? 2. What are Mi'kmaq women’s beliefs and attitudes about menopause? 3. What contributes to Mi'kmaq women’s understanding of mid-life health and menopause? 4. What socio-cultural determinants influence Mi'kmaq women’s experiences of mid-life health and menopause?” Page 5.  “The purpose of this research is to explore the perceptions (physical, psychological, emotional, and spiritual) of mid-life health, with particular emphasis on the menopausal transition among mid-life Mi'kmaq women. In particular, my objective was to gain an in-depth, holistic description of the mid-life transition from a First Nations’ perspective. The ultimate goal of this study is to provide an account that evokes ‘verisimilitude’ (being there) in readers with respect to the experiences of research participants (Creswell, 1998). This position is based on the assumption that ‘if we can appreciate each other’s views, we can see the whole picture more clearly. To heal ourselves or to help heal others, we need to reconnect magic and science...’ (Hammerschleg, 1988, pg14).” Page 50. | | | | | |
| **Study design** | **Conflicts of interest** | | **Inclusion criteria** | | **Exclusion criteria** |
| Qualitative research (dissertation). | N/A; none listed. | | Aged 38 to 83; experiencing or had experienced menopause. Page 60. | | N/A; none listed. |
| **Population and sampling description** | | **Indigenous description** | | **Total number of participants** | |
| The population of interest was “Mi’kmaq women from five First Nations in Nova Scotia”.  “This study included six groups, totaling 42 Mi’kmaq women, ranging from 38 to 83 years of age, from five First Nation communities in Nova Scotia. A number of sampling techniques, including purposive, snowball and opportunistic sampling were employed. Facilitators from each participating community were asked to contact five or six women and invite them to participate in a group discussion about mid-life health and menopause. Each of the facilitators purposefully recruited women who were within the specified age group and who were experiencing or who had experienced menopause. They also recruited these women using snowball sampling (one person suggested another who might be interested in participating) or opportunistic sampling (whoever happened to be available and wanted to participate), or both” Page 60. | | Mi’kmaq women.  The author does not describe how Mi’kmaq racial/ethnic background was defined.  The sample was 100% Indigenous. | | The total number of participants was 42 and 42 were Indigenous. | |
| **Study outcomes and important results** | | | | | |
| “Findings revealed themes of vision, balance, and relationships. Women’s mid-life vision encompasses an evolving perspective of life, self, and the peri-menopausal transition. This vision is informed by past generations, by women’s life experiences, and by the limited resources available to First Nations women. Balance emerged as a central determinant of women’s perspective, experience, and response to mid-life change. The holism with which First Nations women view health was evident in their construction of experiences within physical, emotional, social, and spiritual domains. The context and substance of women’s relationships also plays an important role in shaping their mid-life experiences. Community and kinship networks represent the foundation upon which women understand and experience all aspects of health, including those related to peri-menopausal change.” Abstract  “Women’s life vision is rooted in past values that become increasingly meaningful as they experience the change of life. Maturity and wisdom correspond with a more positive perspective and an acceptance of change in many forms. This enhanced vision not only brings women closer to themselves but also to those individuals who have traveled with them. During this life transition, the values learned in childhood become not only a source of strength but also a source of reflection.” Page 86.  “During mid-life, these women begin to seek knowledge with a new respect for their own capacity to learn and in their ability to guide their own learning. With humility that only comes with true maturity, they seek out new ways of knowing, some rooted in the tradition of their ancestors, and some based on the science of modern medicine.” Page 87.  “Women’s vision of menopause is shaped by myriad influences; by the silence surrounding the experiences of their mother and grandmother, by their expectations of what changes they will experience, the freedom that peri-menopause represents, as well as their own understanding of the physical, emotional and social determinants of menopausal change.” Page 88.  “The women in this study describe a collective silence about women’s bodies in general and menopause in particular. Among these groups of women, almost no one’s mother ever spoke about menopause, nor did their grandmother or most older women they know. Many of the women told stories of a somewhat traumatic menarche in which no one told them what was happening to their young bodies. The silence of women's menarche seems to have sent a subtle but powerful message more clearly than loud words, that women’s reproduction is not a topic of conversation.” Page 89.  “The edict of silence might be perpetuated by women’s beliefs in the power of mothers as role models for behavior. Women view their mothers as strong and resourceful and in attempting to emulate them, remain silent. In this way, silence is passed from one generation to another, from mothers to daughters. Silence, however, can also perpetuate ignorance - women may become prey to misinformation, fear and uncertainty about their ability to cope with change. Misunderstandings and fear of menarche may be carried through a lifetime and menopause can become the corresponding bookend of that experience.” Page 89.  “Women seemed to recognize that the energy and focus required to resist cultural oppression and retrain traditional ways of knowing might have kept past generations of women from pursing information outside their family or community. They acknowledge that silence of menopause comes from a different place and time, and that the tide is turning with a new generation of women who know what questions to ask and where to find answers.” Page 90.  “Many women recognize that their expectations of mid-life change are rooted in the knowledge and experience of others. For some, their expectations are based on stories, which are not altogether positive. In fact, it seems as though silence is broken only with stories of suffering. It is not surprising therefore, to discover that women’s expectations often include negative elements of change. Lack of knowledge about menopause might introduce uncertainty into women’s vision. Some women may even become fearful of change when the information they receive is misunderstood or misrepresented. This cycle of misinformation and misattribution becomes a self-fulfilling prophecy. Fear itself can create negative experience - fear can affect what women do to care for themselves and their families and how they view themselves in terms of changing perspectives and behaviors.” Pages 90 and 91.  “Observing the often-silent experiences of their mothers forms the foundation of many women’s expectations about their own peri-menopausal experience. These expectations however, are not limited to the physical and emotional changes they might experience. In addition to representing the foundation of women’s beliefs about what to expect of the experience, mothers and grandmothers inform women’s expectations about what to expect of themselves, about their ability to cope with change in a way that fulfils [sic] others’ expectations of them.” Page 91.  “Women’s vision also includes the concept of freedom; specifically, freedom from the constraints of childbearing and child rearing as well as repeated cycles of menstrual cramping, bloating and bleeding. Many women emphasized the relative freedom contemporary women have around controlling childbirth. In the past, most women had children well into their forties, so that menopause occurred at a time when they were still caring for small children and many times, large extended families.” Pages 91 and 92.  “Many of these women look forward to a time of rest, with less pressure and responsibilities, and with more leisure time during their old age to pursue activities they did not have time for during the busy years of work and/or caring for young children. They also look forward to a time when they can focus more on themselves, of free time to discover what else they can achieve outside the role of ‘mother’. They embrace this new-found freedom to choose what avenues they wish to pursue - what pleasures they wish to indulge.” Page 92.  “Women seem to have mixed feelings about the issue of mid-life sexual freedom. Although most women appreciate liberation from contraceptives and unplanned pregnancies, they also disclose that their view of sexuality has been altered by misinformation about sexual function during later years. Consequently, some are not able to take full advantage of the relative sexual freedom this time of life offers them. According to this group of women, lack of a sexual partner also impacts women’s ability to pursue a satisfying sex life during mid and later life.” Page 93.  “As discussed earlier, whether through heredity or conditioning, many women believe that their mothers’ experience influences their own. Heredity is also tied to understandings of the past and of ancestors as having an influence on current experiences. However, according to some women, the number and timing of children may also affect a woman’s reproductive system in such a way that menopausal changes will differ, even among women in the same family.” Pages 94 and 95.  “Despite their acceptance of the genetic, biological and physiological components of peri-menopausal change, women emphasize contextual elements, which play a meaningful role in this experience. For instance, large families and a busy lifestyle, filled with hard work and many responsibilities, may distract women from changes occurring in their bodies. Even if these changes are perceived, they may not become prioritized in such a way that they demand little of women's attention and therefore, are relegated to the category of ‘non-event’.” Page 95.  “Family relationships are revealed as having a significant impact on women’s vision of peri-menopausal change. Women express particular concern about how this change might influence their relationships and their care of others. Specifically, women propose that the elements of tension versus support, stress versus relaxation, and self-care versus other care in their social relationships, play a significant role in their perception of peri-menopausal change.” Page 96.  “Lived experience is rooted in relationships; whether women traverse the change alone or with the support of others, connection and caring are brought more closely into the focus of their vision. As is the tradition, vision includes everyone — the entire community and family - the past, present and future. During peri-menopausal change, these women report experiencing a greater connection to others; to those closest to them, but also to those who have gone before them.” Page 99.  “Women’s vision also involves an element of reciprocity - of sharing their thoughts and experiences, their fears and their challenges. This vision embraces those younger women who will follow them through this stage of life. Women maintain that with increased knowledge, comes increased responsibility. They view their own experience as a critical element in the learning of younger women. ... As elder women, it is their responsibility to pass along for the wisdom of their years - so that they might create a more enriched experience for their daughters, their granddaughters, their nieces and other young women in their family and community.” Page 99.  “Women’s vision of themselves is sometimes formed as a result of comparing their experiences with those of other women and determining the degree to which they ‘fit the mould.’ Comparing their physical experience, particularly hot flashes, with those of other women represents an attempt to determine a ‘normal’ menopausal experience. They appear to gain some comfort in knowing that their experience is not out of the ordinary - one that is not so unique as to warrant much of anyone’s attention. This issue was more often than not referred to as balance, in the sense that normal refers to equilibrium - of a process that occurs as it was designed. In order to situate their own experience, some women attempt to gauge it upon what they see and hear from sisters, aunts, cousins and friends. It is interesting to note that they rarely depend on written material to inform their perspective but instead, rely on the lived experience of other women, who are like them and sometimes, of women who they might perceive as very different.” Page 100.  “Euro-Canadian Women. Some women viewed the peri-menopausal experiences of First Nations and Euro-Canadian women as somewhat similar. However, many women believed that their experiences differed from Euro-Canadian women in relation to social and educational issues, social norms, relationships, and belief systems.” Page 101.  “Access to information is viewed as a major difference between First Nations and Euro-Canadian women, with most Euro-Canadian women having more access to information about their bodies in general as well as specific material related to changes that occur during mid-life change. Moreover, interns of general health, Euro-Canadian women are perceived to benefit from a health system, which is set up to serve their needs and not necessarily those of First Nations women. Chronic and degenerative diseases, many of which become more problematic as women age, also become part of their experience of mid-life health and menopause. More and higher quality resources in Euro-Canadian communities are perceived as advantaging many Euro-Canadian women with respect to their physical health. Socio-economic status in general is also seen as a resource for better health, in which First Nations women are disadvantaged.” Pages 101 and 102.  “Close relationships with grandmothers as well as veneration and respect, which represent an essential element of that relationship, are believed to explain First Nations women’s acceptance of aging as well as their connection with and embrace of the grandmother role. According to these women, close connection to one’s grandmothers creates a foundation of understanding for the perspective of elders; learning and guidance are part of the wisdom, which is translated into women's experience. Some women perceive Euro-Canadian culture as lacking in respect and care for elders, while First Nations people view elders as an important resource. This cultural difference provides the basis of women’s belief about why aging may not be as ‘traumatizing’ for First Nations women, as it may be among many Euro-Canadian women” Page 103.  “Women’s vision of life, of change and of themselves may be understood in relation to its source and its substance. Women revealed the source of their vision as indirect as well as overt information they receive from both informal and formal sources. Women cited elders as an especially important source of wisdom in terms of influencing their perspective and contributing to their knowledge. Women also acknowledge the past, both their own and past traditions that have been passed on to them, as an important source of their vision. The substance of women’s vision concerns the specific messages they reveal as having influenced their vision of life and self and menopause.” Page 104.  “Education. Vision is best informed through multiple learning opportunities that relate to all the senses and encompass the holistic nature of this experience. The pluralism of women’s vision honors the gifts of science and medicine as well as the tradition of their ancestors. They inform their vision through investigation on their own, and by talking to friends and family members. They search for information that is balanced in terms of medicine and tradition; they also search for knowledge that accommodates their enhanced capacity for understanding.” Page 105.  “As a source of information, which might enhance women’s vision, doctors tend to fall short of the mark. Consequently, women often search for information through media such as the internet because they perceive that doctors know little about menopause, beyond symptoms and details about HRT. Moreover, women are often made to feel as though their views are not respected and that this experience does not totally belong to them. These women insist that they need an opportunity to learn as much as they can, so they will feel completely informed. Yet, doctors seldom encourage women to pursue alternative forms of healing, nor do they often take time to listen or demonstrate respect for women’s capacity to inform themselves. Thus, women often feel discouraged from attempting to gain greater insight and may forgo further discussion with their doctor.” Page 105 and 106.  “Lessons learned from elders are an important source of women's menopausal vision. In part, grandmothers shape a vision of life that settles more deeply into women's spirit as they journey toward the stage in their own lives. Beyond the practical lessons related to life, elders instill values, which become the fabric of community, family and individual life.” Page 106.  “Among many First Nations people, lessons are taught through stories, into which metaphors for life are intricately woven. One such story, shared by an elder woman in this study, involved a basket maker, performing a task that combines skill, patience and time. Circular and interwoven, the basket takes shape through a combination of the physical elements of nature, gently coaxed into something more beautiful, which seems to take on a life of its own. Sometimes the weaver plans the design and remains true to the form of that arrangement. At other times, the weaver achieves such harmony with the basket that its shape emerges in concert with the interplay of the elements, of each and spirit and caring. The process requires the weaver’s undivided attention and the outcome is often ‘fine art.’ This story provides a metaphor for women’s change and the message is as beautifully and carefully woven as the basket. The journey begins with elements of the earth, of human creation and female fertility, both gifts of nature. The journey takes shape over time, involving creativity and patience. The design may be planned or it may emerge through a relaxed interplay, which combines the plasticity of women’s physical experience with emotional and spiritual elements of change.” Page 108.  “Negative talk. Women ‘hear people talk’ - not so much about menopause in general, but about physical and emotional responses to change. Women hear about hot flashes, weight gain, mental health problems, lack of sex drive, vaginal dryness, painful intercourse, accelerated aging, infidelity (on the part of women as well as their partners), sadness, loss of emotional control, loss of memory, and sleep problems - the list is endless.” Page 110.  “The women who partnered in this study conceptualize health as involving physical, emotional, social and spiritual constituents. Women recognize that good health can only be achieved when all of these elements are balanced. Through this conceptualization, they reveal an appreciation for the myriad dimensions of human existence, which engage in bi-directional influence of women’s mid-life change. The findings also reveal women’s understanding - that describing the event of menopause is not sufficient to explain mid-life health.” Page 112.  “Traditional ceremonies are premised on the notion of balancing physical, mental and spiritual elements of human experience. For those women who practice traditional healing, these ceremonies may become particularly meaningful at mid-life. Most recognize that medical intervention alone will not fully address every health concern. Even those who don’t practice traditional healing appreciate the holistic and interrelated concept of health. In fact, all of the women appreciate and report that optimal emotional health is unlikely without good physical health and vice versa, for both depend on health behaviours that respect the body, mind and spirit.” Page 112 and 113.  “Menopause is seen by some women as a catalyst for change - a renewal of spirit - of a zest for life - a life more centered on them - as a woman and as a human being. Many find themselves with renewed enthusiasm for activities they have not pursued since their youth. Women discover that they still have an interest in activities they once enjoyed, before the responsibilities of a home, and work and family captured most of their time and attention. Many women see it as a time to relax, when worries over work and children diminish; a time when those for whom they have cared in the past will care for them. Women are generally more relaxed about the future than they were in their youth - they live each day to the fullest because they have learned the wisdom of taking life slowly - of savoring small delights and not waiting for future pleasures that may not materialize. Most reveal this as a time when they are better able to let go of worry and become more open to new beginnings.” Page 115.  “Many women viewed freedom from monthly bleeding as one of the physical benefits of menopause. Those who wish to, may now participate in ceremonies, from which they were once restricted during their menstrual cycles. Women's enhanced comfort with themselves also represents a gift of menopause, which is viewed as a milestone toward increased power and wisdom as elders. Finally, mid-life change is viewed as a gift of time - time to slow down - time to think about one's own needs, one's own health and happiness. Time becomes women's ally, rather than something to feel pressures by or battle against, as they might have done in their youth.” Page 116.  “Health. The women who partnered in this research claim that health is one of the most important considerations during mid-life. It seems as though peri-menopausal changes prompt women to think about their physical health more often than they might have when their bodies were younger and stronger. As they become increasingly concerned with their health, women begin to search for information that will help them prevent disease and maintain health.” Page 117.  “Health conditions occurring at mid-life may coincide with the experience of menopause. It is not surprising therefore, to discover that women may associate menopause with illness and declining health. This perspective is supported by medical information, which emphasizes the impact of hormonal change on women’s physical and emotional health.” Page 117.  “Disease. Cultural nuances notwithstanding, menopause is viewed as a sign of old age. Like the experience of menopause, female aging may be perceived and experienced on a number of levels and women report experiences ‘getting old’ within physical, psychological, spiritual and social dimensions. Yet, physical aging, and its corresponding reduction in overall health is likely of more concern for First Nations women, than middle-class, Euro-Canadian women. Marginalized health systems and disadvantaged socioeconomic conditions contribute to the relatively poor health of First Nations women, so that old age typically brings significant risk of chronic degenerative disease, experienced within a health care system this inadequately funded and delivered.” Page 118.  “Finally, the high rates of hysterectomy among these First Nations women (approximately two-thirds of the sample) create concerns about estrogen causing cancers.” Page 119.  “Some women report no discernible changes in their menstrual cycle until it abruptly stops. Likewise, some report no mood swings, no hot flashes, and no memory disturbances. The experiences reported by other women include changes in personality, eating habits, interpersonal tolerance and health; hot flashes, weight gain, mood swings, heart palpitations, hot or cold sweats, changes in libido, vaginal dryness and/or painful intercourse, sleep disturbances, irregular periods, dizziness, depression, fatigue, memory loss, incontinence, and reduced energy. The gradation and variation in reported experiences is best viewed on many continua, related to the frequency, duration and intensity of these experiences.” Page 121.  “While perhaps not as much a focus of attention for First Nations women, body image did emerge as one of the factors which influence women’s vision of themselves during mid-life. In particular, weight control seems to become more important to some women after natural menopause or hysterectomy. Although they observed that First Nations women may not be as self conscious about the ‘signs’ of aging, they did acknowledge that weight gain can create feelings that range from discomfort to depression. In general, women recognize the importance of feeling good about the way they look and of pursing sensible activities (exercise, proper diet, hair and skin care) that enhance self-esteem and well being. It is interesting to note however, that body image also emerged as an issue of physical fragility and declining health, which was associated with women's ability to fulfil [sic] work responsibilities.” Pages 122 and 123.  “Hot Flashes. The diversity with which women experience hot flashes provides the most dramatic example of the plasticity of this peri-menopausal phenomenon. In general, women report experiences that range from no hot flashes at all to those, which are very mild to quite extreme.” Page 123.  “Hysterectomy. A surprising number of women in this study reported having undergone hysterectomy. Whether they represent increased use of this procedure among First Nations women or whether this large number is an artifact of the recruitment process, only future research will discern. These women do however, provide valuable insight into the experience of reproductive and hormonal change. The two most common experiences of these women include sudden hot flashes and weight gain. However, it is difficult to know whether weight gain is an outcome of hysterectomy or a product of dietary changes that might be related to coping with the diagnosis of uterine disease and subsequent surgical intervention. These women did report that they were told prior to their surgery that they would gain weight - and so, the power of suggestion might also serve to explain their weight gain. Women reported drastic reduction in physical activity after hysterectomy, providing yet another potential explanation. They were also told that they would lose their sex drive after surgery and, low and behold - some did! This presumption of diminished sex drives seems to be pervasive.” Page 125.  “One can only speculate about whether women’s emotions are a product of sudden hormonal change or whether they were rooted in the changes wrought by this experience to women’s self concept - to their sexuality, femininity, fertility, or wholeness as a person. They report receiving little or no information and that what little they did receive, was written and/or presented in such a way as to alienate or confuse them. Most of the women conceptualize their experience as ‘no normal’ and felt unsure about where they fit in discussions of menopause. It seems that hysterecomized [sic] women tend to be excluded or to be separated from ‘normal’ menopausal women - thus further fragmenting the experiences of this group.” Page 126.  “It is important to note that the post-menopausal women in these groups claim that the emotional liability of peri-menopausal change is transient and often followed by a well-earned sense of calm.” Page 128.  “A few of the women in this study revealed feelings of loss during peri-menopausal change. These women often exist in a bi-cultural state that is infused with western values of youth and beauty, so they may perceive a sense of loss during mid-life. They are often not prepared for the physical changes brought on by age.” Page 130.  “Female strength is valued in most Aboriginal cultures. For women who practice the traditional spirituality of the Mi'kmaq people, menopause is viewed as a sign of strength, unlike the expectations for many Euro-Canadian women, who have traditionally been seen as the weaker sex (even though this label is fallacious in the extreme). Before the intrusion of European culture, First Nations women worked side-by-side with their male partners during times when egalitarianism was the norm. To be a woman is to be strong. This mantle of strength is born with pride and with some measure of ambivalence, for it is accompanied by inequitable responsibilities, for self, family and community. The expectation is often that exceptional responsibilities will be shouldered without comment or complaint, and without thanks or appreciation to the benefit of others, despite the detriment to women.” Page 133.  “Several theories emerge to explain this sustained edict of female strength. The first relates to cultural values of stoicism. The next relates to cultural norms around complaining or drawing attention to oneself. The third relates to weakness - historically, the challenges faced by Aboriginal peoples left little room for error. Everyone, women included, had to contribute to their family community. Complaints did not contribute and were thus discouraged. Acceptance of what must be done in order to adapt to changes and attend to the daily rigors of life was expected. The forth relates to the third but deals specifically with the harsh realities of colonialism and the ensuing encroachment on Aboriginal lands and intrusion into Aboriginal lives, which created difficult realities that could not - until recently, be changed - so, had to be borne. ‘That which does not kill us makes us stronger’ – ‘that which cannot be changed, must be accepted’ - both may be aptly applied to the cultural imperatives of the Mi'kmaq people, upon which women address the change of life. Strength, which is always present, must be harnessed during times of challenge. Taught the harsh reality of an Aboriginal existence early in life, mothers and grandmothers attempted to instill the same determination and tenacity in their children, so that they might draw on that strength during the hard reality of residential school, the hardships brought on by a colonial government and the blood born, historic trauma of attempted ethnocide.” Page 134.  “Women who are raising sons while going through the change sometimes take the opportunity to teach these young men about the reproductive changes women experience and encourage them to be gentle and compassionate with women. This connection and teaching, particularly between mothers and sons is another Aboriginal practice that stands as an example for western culture. Strong family connections encourage empathy in boys and a greater likelihood that they will grow into men who are equally empathetic and supportive with their life partner.” Page 144.  “Spirituality emerged as an important determinant of women's overall health, which becomes increasingly emphasized during the peri-menopausal transition. Faith, in whatever form it takes, seems to sustain women through this sometimes-difficult transition. For many women, spirituality has always been a foundational component of life and provides the value system upon which they conduct themselves and raise their children.” Page 146.  “The enlightenment and clarity, which emerges during mid-life, prompts many women to re-discover their spirituality. Some women practice Euro-Canadian religious traditions, while others practice traditional ceremonies such as the sweat lodge, pow wow and traditional prayer, all of which contribute to their spiritual strength and help them connect more deeply with themselves and others.” Page 147.  “The re-emergence of Aboriginal spirituality was reflected in the perspectives and practices of several women. Beyond their involvement in traditional ceremonies, they also reveal traditional metaphysical beliefs about a connection between the natural world, human existence and multiple domains of spirituality. Women express particular pride in the spirituality of First Nations people and the way it benefits them in general as well as in specific ways relative to the peri-menopausal transition.” Page 147.  “For those who practice Aboriginal spirituality, the emphasis seems to be on connecting with emotional or spiritual stress and releasing it through the body. This ideology works especially well during the peri-menopausal transition, when emotional and physical experiences respond well to traditional healing. For instance, heat and sweating, often pathologized in Euro-Canadian culture, are emphasized as an avenue of health and spiritual growth.” Pages 147 and 148.  “Women seem to practice stoicism not only to cope with their own discomfort but also to protect their families from the negative ‘trickle’ effects of their experience. First Nations women, like most women, are often the center of family life - this fact is widely acknowledged. Despite inequalities in social, political and economic power, women’s role and responsibility in stabilizing families is rarely questioned. Women recognize that this time of change may be especially difficult because of the pivotal role they play in families and their position as emotional compass for family life. This responsibility however, creates tremendous pressure on women to ‘hold it together’, not only for their own sake, but for the sake of their family and through extended kinship networks, their community.” Page 153.  “Women recognize that the social conditions and subsequent health problems experienced by First Nations people, impact women’s experience of mid-life health and that healing must also address those domains in order to be effective. To a lesser extent, women talked about changing their diets, particularly adding food and vitamins that are known to benefit health and/or have estrogenic properties (e.g. soy products). Very few women took medication other than HRT to maintain balance and only one woman revealed that she occasionally smoked to relieve the stress brought on by hormonal change.” Page 156.  “Traditional healing is available, and in varying degrees practiced, by several of the women. The extent to which they pursue traditional healing was often a function of the degree of cultural accommodation within their community, but often depended on individual women’s own distinct cultural identity. Those who do not ascribe to traditional healing practices, however, sometimes pursue it when western medicine fails to provide the healing they seek.” Page 157.  “Herbal Medicine. A combination of adherence to traditional healing practices as well as recent evidence about the deleterious effects of HRT, has led many First Nations women to seek out Aboriginal and Euro-Canadian herbal preparations to address peri-menopausal change and to improve overall health. Herbal remedies typically work with, rather than against, the body and are more bioavailable and bioacceptable. Herbal medicine is sometimes used to relieve anxiety and stress - making it an ideal choice for peri-menopausal women. Herbal teas were especially popular as a sleep aid or for calming jittery nerves. Some women described herbal medicine that will improve your sex life - comments such as these were met with much friendly teasing and laughter.” Page 158.  “Many women see the use of Aboriginal medicine, provided by the earth, and untouched by pharmaceutical entrepreneurs, as a means of restoring balance in a natural way. The oldest woman (83), who was very knowledgeable about traditional healing, used teas made from rose hips or punch berries extensively. She advised that punch berries, brewed into a light tea, help to thin the blood. This medicine is very old and was used extensively by the Mi'kmaq people in the not-so-distant past.” Pages 158 and 159.  “According to one woman, there are ‘Indian medicines’ that help women go through menopausal change. However, it is almost impossible to find out what these medicines are because most healers will not share their secrets. This is due, in part, to years when Aboriginal medicine and other healing practices were outlawed in North America. Healers had to practice ‘underground’, away from the prying eyes of Indian agents and reserve doctors. This is also a way to protect Aboriginal medicine from Euro-Canadians who might try to exploit this knowledge, stripping it of its spiritual context and extracting the biopharmaceutical elements.” Page 160.  “Most doctors do not offer alternatives to HRT and many downplay the risks. Consequently, women do not feel as though they receive much help from their doctor in the form of information or support around alternatives to HRT. Women recognize the disparity of information about the health of men and women. This is particularly true for First Nations people, who are typically aggregated into gender neutral groups for the purpose of examining health issues. According to these women, the aging of men is more accurately understood and, thus women do not always value male doctors as a credible source of information about menopause.” Pages 163 and 164.  “Women talked about the important changes, which have occurred over their generation - a time when new ways of thinking were not always encouraged. During those tumultuous times, First Nations traditions were in danger of annihilation and elders struggled to sustain Mi'kmaq language and culture. Yet, socio-economic pressure has forced many First Nations to adopt a more bi-cultural approach. It is into this environment, the next generation of First Nations women will traverse peri-menopausal change.” Page 169.  “Some women expressed concerns about the overzealous prescribing practices of physicians. Rather than working in collaboration with other health professionals to care for the whole person, women describe an approach to health care that more closely resembles a grocery store check out. Women receive a polite hello, a quick scan, and a concluding assessment - handled, bagged and sent on their way - slightly beleaguered but relieved to have it over with.” Page 176.  “Many of the women who have undergone hysterectomy seems especially disillusioned by their interaction with health care professionals. According to some of these women, they received relatively little information about their particular problem or the procedure required to improve their health. They also report a lack of information about the potential consequences and risks of the procedure or the hormone regimen that was to become a permanent fixture in their lives. Some women perceive this as an assumption on the part of health professionals that First Nations women do not wish to receive or cannot understand information about this aspect of their health.” Page 177.  “Women reveal the value of autonomy in their acknowledgement of the right of every woman to subjectively perceive the benefits and drawbacks of menopausal change. The notion of staying true to what feels right for the individual also emerged as a component as women’s autonomy. Everyone’s experience was accepted and validated as a meaningful expression of her individuality and no one's healing practice was received with disapproval or scorn. According to these women, to claim that menopause is definitively this or that, may demonstrate disrespect toward a woman who it not experiencing the same benefit or challenge. This belief provides some clues about why advice giving around menopause is so uncommon among First Nations women - as this type of opinion sharing or advice may be considered inappropriate and/or disrespectful.” Pages 187 and 188.  “Joking and teasing are an integral part of communication in many First Nations cultures, particularly around sensitive issues. Many of the women in this study used jokes as a vehicle for sharing information about menopause in a more palatable and appropriate way. The value of this communication strategy lies in its capacity to allow women to express their feelings and to highlight this issue in a context where it might otherwise be unfavorably received. In fact, any women report that jokes about menopause are the only context in which they have ever heard it mentioned.” Page 194. | | | | | |
| **Use of STRAW+10** | **Menopausal transition staging of sample using STRAW+10** | | | | |
| The STRAW+10 not used. | Menopausal Transition and Postmenopause. | | | | |
| **Author reported facilitators to treatment** | | | **Author reported barriers to treatment** | | |
| “The inclusion of a First Nations perspective contributes substantially to a multi-dimensional, inter-relational model of mid-life change, which can inform research, policy and practice. Further inquiry into the additional obstacles faced by Aboriginal women is essential and a more balances and interdisciplinary research agenda is required. Programs and services must be based on indigenous values and program inertia must be in the direction of intergenerational capacity building, rather than perpetuation of medical dependence. Finally, programs must incorporate opportunities for women to engage in diverse strategies toward health and healing.” Abstract | | | “Medical Contexts. The historical significance of relationships between First Nations people and the federal health care system cannot be underestimated when attempting to understand the influence of social contexts on women’s mid-life experience. According to the findings of this study, women who pursue traditional healing and/or herbal medicine seem to feel more empowered about this life experience. However, jurisdictional issues related to accessing health services outside those covered by the federal government, present barriers to First Nations women who wish to pursue alternatives to HRT and ‘medical treatment’ during this natural process of change. Historic relationships, in this case between First Nations women and physicians, differ substantially from those of most Euro-Canadian women, the cultural group to which most physicians belong. Interactions between physicians and First Nations women are premised on inter-cultural and inter-class inequities in social power, which may disadvantage women with respect to decisions about their mid-life health (Adams, 1995). Physicians may not be particularly sensitive to women’s past life experiences, particularly those, which might predispose First Nations women to difficulties during peri-menopause.” Pages 244 and 245. | | |
| **Study limitations** | | | | | |
| “Sampling. Research is full of limitations, not the least of which is represented by the undirected comments of a select group of mid-life First Nations women, who were recruited and decided to participate in a study, which asks them to talk about their subjective experience of peri-menopausal change. However, without this information, we cannot begin to appreciate the salient issues for these women during their middle years. In particular, the cultural lens used to interpret this information reveals some significant differences in the way First Nations women subjectively understand and experience this process, which we now understand, is in no way universally experienced. The findings of this study cannot necessarily be generalized to all First Nations women or even to Mi'kmaq women who did not participate in the research. Like most, these findings are based on a relatively small sample, from selected communities, of women who, for one reason or another, heard about and chose to participate in, this study.” Page 82.  “Use of a purposive sampling strategy restricts the generalization of the findings to this population of First Nations women. In fact, I must acknowledge that this phenomenon is too variable and context-bound to permit broad generalization of the findings. At best, I offer extrapolations of ‘modest speculations on the likely applicability of findings to other situations under similar, but not identical, conditions’” Page 82.  “Group Methods. Although the use of group discussion permitted me to increase the sample size over individual interviews, it did not confer generalizability of the research findings. ... One of the most important limitations of group discussion relates to the issue of confidentiality. While facilitators and I assured the confidentiality of participants' responses, we cannot guarantee that all group members will respect the privacy of others (Patton).” Page 83.  “Community/Personal Descriptors. Given the relatively small number of First Nations in Nova Scotia as well as the relatively few mid-life, First Nations women, I have not included specific descriptors related to participants or their home communities. Although community and/or personal level descriptors may not identify participants or communities to most Euro-Canadian readers, the likelihood of identification increases substantially among other Mi'kmaq people from Nova Scotia, who might easily recognize specific descriptors such as participants’ occupation or a community's main industry. While some may consider the absence of these descriptors a limitation of this research, I believe that emphasizing this type of description follows a Eurocentric model of identity that might actually detract from the essence of women’s lived experience. The rationale for this decision is based on my understanding of indigenous identity, which is not defined by one’s economic activity but rather, by one’s relationship to others and by the experiences that shape individual and collective life.” Page 84. | | | | | |

| **Record ID** | **Title** | | | | |
| --- | --- | --- | --- | --- | --- |
| #1717 – 1891 | A study relative to the functions of the reproductive apparatus in American Indian women | | | | |
| **Author information** | | | | | **Country** |
| Currier, A. F. | | | | | United States |
| **Study aims** | | | | | |
| The objective was to examine the functions of the reproductive apparatus in American Indian women. | | | | | |
| **Study design** | **Conflicts of interest** | | **Inclusion criteria** | | **Exclusion criteria** |
| Cross-sectional study. | N/A; none listed. | | N/A; none listed. | | N/A. |
| **Population and sampling description** | | **Indigenous description** | | **Total number of participants** | |
| The population of interest was American Indian women.  This record offers no details about sampling. The author does state “interesting facts were gathered” at “twenty-eight Indian agencies and army posts.” Additionally, “the report of the Indian Commissioner for 1890 was also freely consulted, and was the source of much valuable information.” Page 2. | | “Southern tribes—the Apaches, Mojaves”. Pages 2 and 8.  “Northern tribes ... Cheyennes and Arapahoes”. Pages 2 and 8.  “Sioux”. Pages 2 and 8.  “Quabaw Agency (Ind. Ter.)”. Page 3.  “Round Valley Agency (Cal.)”. Page 3.  “Neah Bay Agency(Wash.)”. Page 3.  “Cherokees”. Page 4.  “Klamaths (Oregon)”. Page 4.  “Crows and Assiniboines”. Pages 4, 5, and 8.  “Sacs and Foxes”. Page 7.  The author does not describe how racial/ethnic background was defined.  The percentage of the sample who were Indigenous is 100%. | | N/A. | |
| **Study outcomes and important results** | | | | | |
| “The menopause seldom causes any trouble. The age at which it comes shows as wide a diversity as it does among civilized women, and there was also the same variety as to duration. It may come abruptly, or it may require several years.” Page 4.  “The Menopause.—Indian women are exceptionally free from the nervous and vascular disturbances that so commonly accompany the menopause in civilized women. The duration of the menopause varies as greatly as it does in civilized life. It usually comes between the fortieth and fiftieth years, but not infrequently is delayed beyond the fiftieth year. Many gestations occurring in rapid succession, continuous hard work, and the exposure and physical suffering incidental to a savage life do not tend to shorten the menstrual and childbearing periods.” Pages 8 and 9.  “Finally, Indian women in the savage state undergo less physical suffering in connection with the reproductive apparatus than do civilized women. As a rule they menstruate, bear children, and pass the menopause with the minimum of discomfort. This is due to three causes: (1) Natural or racial insensitiveness compared with the far more sensitive Caucasian; (2) abundance of exercise; (3) life in the open air. With its complex conditions, civilized life will always present obstacles to the performance of the functions peculiar to women with the same ease with which they are experienced by savages, and when Indian women exchange the savage for the civilized state they must necessarily adopt also some of the ills that are inseparable from the latter.” Page 11.  Note: No specific tribes are referenced when presenting observations about menopause. Rather, the author makes a distinction between “savages” and the “civilized.” It should be noted that the author compared experiences between the groups of “savages” and “civilized.” The author does not find rural vs. urban differences in outcomes. However, the author state: “The menopause seldom causes any trouble. The age at which it comes shows as wide a diversity as it does among civilized women, and there was also the same variety as to duration. It may come abruptly, or it may require several years.” Pages 3 and 4 “Finally, Indian women in the savage state undergo less physical suffering in connection with the reproductive apparatus than do civilized women. As a rule they menstruate, bear children, and pass the menopause with the minimum of discomfort. This is due to three causes: (i) Natural or racial insensitiveness compared with the far more sensitive Caucasian; (2) abundance of exercise ; (3) life in the open air.” Page 11. | | | | | |
| **Use of STRAW+10** | **Menopausal transition staging of sample using STRAW+10** | | | | |
| The STRAW+10 not used. | Unclear. | | | | |
| **Author reported facilitators to treatment** | | | **Author reported barriers to treatment** | | |
| N/A. | | | N/A. | | |
| **Study limitations** | | | | | |
| N/A; none described. Author bias from the colonizer’s perspective is evident. | | | | | |

| **Record ID** | **Title** | | | | |
| --- | --- | --- | --- | --- | --- |
| #1617 – 2005 | Hysterectomy prevalence and cardiovascular disease risk factors in American Indian women | | | | |
| **Author information** | | | | | **Country** |
| Zhang, Y.; Lee, E. T.; Cowan, L. D.; North, K. E.; Wild, R. A.; & Howard, B. V. | | | | | United States |
| **Study aims** | | | | | |
| “Objectives: We investigated hysterectomy prevalence and associated demographic and reproductive factors among American Indian women. The association between hysterectomy and cardiovascular disease (CVD) risk factors was also examined.” Abstract  “The aims of this study were to: (1) examine the prevalence of hysterectomy prior to natural menopause and its associated demographic and reproductive factors in a large cohort of American Indian women, and (2) determine if premenopausal hysterectomy with ovaries conserved is associated with CVD risk factors (including lifestyle factors and blood analytes) among women who have reached the expected age of menopause.” Page 329. | | | | | |
| **Study design** | **Conflicts of interest** | | **Inclusion criteria** | | **Exclusion criteria** |
| Cohort study. | N/A; none listed. | | “Women who were postmenopausal, were not using estrogen, knew if their ovaries had been removed, and had no history of CVD at the baseline exam (n = 1726) were included in analyses of the associations between hysterectomy and CVD risk factors.” Page 330. | | Participants with missing data were excluded. Women using estrogen were excluded. Pages 329 and 330. |
| **Population and sampling description** | | **Indigenous description** | | **Total number of participants** | |
| The population of interest was American Indian women from “13 American Indian tribes/communities conducted in three study centers: southwestern Oklahoma, central Arizona, and North and South (N/S) Dakota.” Page 329.  Sampling details are not provided: “The study design, examination methods, and laboratory techniques were described previously [22–24].” Page 329. References:  22. Lee ET, Welty TK, Fabsitz R, et al. The Strong Heart Study. A study of cardiovascular disease in American Indians: design and methods. Am J Epidemiol 1990;132:1141–55.;  23. Howard BV, Welty TK, Fabsitz RR, et al. Risk factors for coronary heart disease in diabetic and nondiabetic native Americans. The Strong Heart Study. Diabetes 1992;41(Suppl 2):4–11.;  24. Cowan LD, Go OT, Howard BV, et al. Parity, postmenopausal estrogen use, and cardiovascular disease risk factors in American Indian women: the Strong Heart Study. J Womens Health 1997;6:441–9. | | American Indian women from “13 American Indian tribes/communities conducted in three study centers: southwestern Oklahoma, central Arizona, and North and South (N/S) Dakota.” Page 329.  The authors do not describe how American Indian race/ethnicity was defined.  The percentage of the sample who were Indigenous was 100%. | | Total sample size: “The final sample size for the hysterectomy prevalence analysis was 2689.” Page 329.  Total sample size for the analysis of premenopausal hysterectomy and CVD risk factors: n=1726. Page 330.  All were Indigenous. | |
| **Study outcomes and important results** | | | | | |
| “Results: Hysterectomy prevalence ranged from 24% to 34% across differing age groups and the percent with oophorectomy among those with a hysterectomy ranged from 43% to 63%. Geographic area, more prior pregnancy losses, more education, and less speaking of the native language were associated with increased hysterectomy prevalence. After adjustment for age, the women who had a hysterectomy with intact ovaries had higher total cholesterol and low-density lipoprotein cholesterol levels than those with a natural menopause.” Abstract  “Hysterectomy prevalence analysis. The overall prevalence of pre-menopausal hysterectomy (with or without oophorectomy) in this population was 30%, and age-specific prevalence proportions ranged from 24% to 34% (Fig. 1). ... The women who had hysterectomies tended to have more years of education, more prior fetal losses, spoke their native language less frequently, and were more likely to live in Oklahoma or Arizona (Table 1). The multivariate analyses shows that more education (odds ratio: 1.07 (per year increase of education), 95% confidence interval (CI): 1.04, 1.11), and a greater number of lost pregnancies (≥3 lost pregnancies versus <3, odds ratio: 1.57, 95% CI: 1.05, 2.36) were associated with higher hysterectomy prevalence (Table 2). More speaking of the native language (odds ratio: 0.90 (per level increase of native language speaking frequency), 95% CI: 0.84, 0.97) was associated with lower prevalence of hysterectomy (Table 2)." Page 331 "Overall, the women who had hysterectomy with preservation of ovaries had higher total cholesterol and LDL-C levels than women who had a natural menopause, while women who had hysterectomy plus oophorectomy had the lowest level of HDL-C among the three groups, although they did not differ significantly from women with natural menopause.” Page 333.  From the Discussion section: “Compared to previous studies conducted in different populations, American Indian women have either a similar or a lower hysterectomy proportion than non-Hispanic whites, a higher hysterectomy prevalence than Hispanics, and a lower hysterectomy proportion than African American women in certain age groups. The hysterectomy prevalence for 50–54-year-old SHS women is similar (p = 0.3) to the prevalence in non-Hispanic whites (31%) who joined the Wisconsin Longitudinal Study [28]. The SHS had lower prevalence rate in each corresponding age group (p < 0.0001) as compared to women who joined another study conducted in 16 states, including Oklahoma (majority non-Hispanic whites) [29]. As compared to National Health Interview Survey data (1998, 1999) [30], in both 45–64-years old and in those 65 years and older, American Indian women have a higher hysterectomy proportion than Hispanic women (27.8% for 45–64 years and 12.4% for 65 years and above) and a similar prevalence in comparison to non-Hispanic whites (33.7% and 22.8%, respectively).” Page 333. | | | | | |
| **Use of STRAW+10** | **Menopausal transition staging of sample using STRAW+10** | | | | |
| The STRAW+10 not used. | Postmenopause.  “To assess menopausal status, participants were asked if their menstrual cycles had stopped permanently, and if so, whether their menopause was natural or surgical. If the answer was ‘surgical’, the participant was classified as having had a premenopausal hysterectomy and she was questioned further as to whether or not her ovaries were also removed. If she indicated ‘yes’, she was classified as having a history of oophorectomy. Women were considered to be post-menopausal if they had a natural menopause or had a hysterectomy with concurrent oophorectomy. Women who reported hysterectomy without oophorectomy or who did not know whether their ovaries had been removed were classified as postmenopausal if their age was ≥53 years, the age at which 90% of women in this cohort had a natural menopause. If the answers to the menopause questions were missing and the woman’s age was ≥55 years at the time of exam, then they were classified as postmenopausal, since 95% of the women in this cohort had a natural menopause by this age.” Pages 329 and 330. | | | | |
| **Author reported facilitators to treatment** | | | **Author reported barriers to treatment** | | |
| N/A. | | | N/A. | | |
| **Study limitations** | | | | | |
| N/A; none described. | | | | | |

| **Record ID** | **Title** | | | | |
| --- | --- | --- | --- | --- | --- |
| #1616 – 2002 | The effect of estrogen use on levels of glucose and insulin and the risk of type 2 diabetes in American Indian postmenopausal women: The Strong Heart Study | | | | |
| **Author information** | | | | | **Country** |
| Zhang, Y.; Howard, B. V.; Cowan, L. D.; Yeh, J.; Schaefer, C. F.; Wild, R. A.; Wang, W.; & Lee, E. T. | | | | | United States |
| **Study aims** | | | | | |
| “Objective—To examine the associations between estrogen use and levels of insulin and glucose as well as the effect of estrogen use on the risk of type 2 diabetes.” Abstract.  “In the Strong Heart Study (SHS) population, which represents 13 tribes in three geographic areas, the prevalence rates of type 2 diabetes range from 40 to 70% in women aged 45-74 years (14). This high prevalence of diabetes provides a unique opportunity to help clarify the role of estrogen use.” Page 500. | | | | | |
| **Study design** | **Conflicts of interest** | | **Inclusion criteria** | | **Exclusion criteria** |
| Cohort study. | N/A; none listed. | | “Postmenopausal women who did not have a history of diabetes, did not take diabetic medication, and had a fasting plasma glucose level <7.0 mmol/l (126 mg/dl) and a 2-h post-challenge glucose level <11.1 mmol/l (200 mg/dl) at the baseline examination were eligible for the present analysis” Page 501. | | Incomplete data and “Inconsistent information on estrogen use at the baseline and second examination.” Page 501. |
| **Population and sampling description** | | **Indigenous description** | | **Total number of participants** | |
| The population of interest was American Indians from 13 tribes/communities living in Arizona, North and South Dakota, and Oklahoma. Page 501.  Sampling strategies are not described in this report. The author states: “The design, survey methods, and laboratory techniques of this study have been presented in previous reports (15-17).” Page 501.  References:  15. Lee ET, Welty TK, Fabsitz R, Cowan LD, Le NA, Oopik AJ, Cucchiara AJ, Savage PJ, Howard BV: The Strong Heart Study: a study of cardiovascular disease in American Indians: design and methods. AM J Epidemiol 132:1141-1155, 1990.;  16. Howard BV, Welty TK, Fabsitz RR, Cowan LD, Oopik AJ, Le NA, Yeh J, Savage PJ, Lee ET: Risk factors for coronary heart disease in diabetic and nondiabetic Native Americans: the Strong Heart Study. Diabetes 41:4-11, 1992.;  17. Lee ET, Howard BV, Savage PJ, Cowan LD, Fabsitz RR, Oopik AJ, Yeh J, Go O, Robbins DC, Welty TK: Diabetes and impaired glucose tolerance in three American Indian populations aged 45-74 years: the Strong Heart Study Diabetes Care 18:599-610, 1995. | | American Indians from 13 tribes/communities living in Arizona, North and South Dakota, and Oklahoma. Page 501.  The authors do not describe how they defined American Indian racial/ethnic background.  The percentage of the sample who were Indigenous was 100%. | | A total of 857 participants were eligible and 857 were Indigenous. Page 501. | |
| **Study outcomes and important results** | | | | | |
| “Results—Postmenopausal estrogen use was associated with lower fasting glucose (0.2 mmol/lower) but higher 2-h glucose levels (0.4 mmol/l higher) compared with never users. It was significantly associated with the risk of type 2 diabetes compared with past and never users, based on American Diabetes Association or World Health Organization definitions of diabetes or on only a 2-h glucose level ≥11.1 mmol/l. However, the risk of type 2 diabetes increased with increasing duration of estrogen use among current users, with an odds ratio of 1.10 per year of use (95% CI: 1.01-1.19). Conclusions—The data suggest that estrogen use in American Indian postmenopausal women may relate to deterioration of glucose tolerance. Longer duration of estrogen use among current users may relate to an increased risk of type 2 diabetes.” Abstract  “Cross-sectional associations between estrogen use and baseline levels of glucose and insulin. Current estrogen users had lower fasting insulin and fasting glucose levels than past users and never users, when other risk factors (age, obesity, etc.) were not considered (Table 1). After adjustment for obesity variables, sociodemographic variables, lifestyle factors, family history of diabetes, and reproductive variables, current users had lower fasting glucose than never users (0.2 mmol/l lowers) but higher 2-h glucose levels than never users (0.4 mmol/l higher). There was no difference in fasting insulin levels between current users and never users after adjustment for covariates. Insulin and glucose levels in past users were not different from never users. Results did not change when we conducted the same analyses by strata of BMI, family history, or baseline age (data not shown).” Page 501.  “Longitudinal analyses of the association between estrogen use and the risk of type 2 diabetes. Because past users and never users had similar insulin and glucose levels at the baseline survey, they were combined for analyses of the risk of type 2 diabetes. After adjustment for other risk factors, estrogen was not associated with increased risk of type 2 diabetes, using any of the three criteria for diabetes (Table 2). However, there was an increasing trend of ORs based on the inclusion of 2-h glucose. Before adjustment, the ORs for diabetes risk by criteria of only fasting glucose, fasting or 2-h glucose were 0.33, 066, and 0.82, respectively. After adjustment, the ORs for diabetes risk were 0.48, 1.11, and 1.58, respectively. Pages 501 and 502 Conclusions—In the cross-sectional analyses of baseline data, compared with never users, current users of estrogen had lower fasting glucose (0.2 mmol/l lower) but higher 2-h glucose levels (0.4 mmol/l higher). During a 4-year follow-up period, postmenopausal estrogen use was not significantly associated with increased incidence of type 2 diabetes, but ORs increased as 2-h glucose was added to the definitions of diabetes. With increasing duration of postmenopausal estrogen use, the risk of type 2 diabetes showed a significant increase among current users.” Pages 502 and 503. | | | | | |
| **Use of STRAW+10** | **Menopausal transition staging of sample using STRAW+10** | | | | |
| The STRAW+10 not used. | Postmenopause.  “Postmenopausal women included those who had undergone either natural or surgical menopause. Natural menopause was defined as at least 12 months since the last menstrual period. If menopause was surgical, the participants were asked if only their uterus had been removed. Women who reported hysterectomy but no oophorectomy or who did not know whether their ovaries had been removed were included if they were ≥ 53 years old.” Page 501. | | | | |
| **Author reported facilitators to treatment** | | | **Author reported barriers to treatment** | | |
| N/A. | | | N/A. | | |
| **Study limitations** | | | | | |
| N/A; none described. | | | | | |

| **Record ID** | **Title** | | | | |
| --- | --- | --- | --- | --- | --- |
| #1615 – 2004 | Associations of postmenopausal hormone therapy with markers of hemostasis and inflammation and lipid profiles in diabetic and nondiabetic American Indian women: The Strong Heart Study | | | | |
| **Author information** | | | | | **Country** |
| Zhang, Y.; Howard, B. V.; Cowan, L. D.; Welty, T. K.; Schaefer, C. F.; Wild, R. A.; Yeh, J.; & Lee, E. T. | | | | | United States |
| **Study aims** | | | | | |
| “Objectives: To examine the associations of postmenopausal hormone therapy (PHT) with indicators of hemostasis and inflammation and with lipid profiles in American Indian women and to determine if diabetes modifies these associations.” From the abstract.  “The present study examined the associations of PHT and hemostatic factors, an inflammatory marker, and lipid profiles in a large population of American Indian women with a high prevalence of both diabetes and CVD. ... PHT use is well known to bring about decreased levels of low-density lipoprotein cholesterol (LDL-C) and increased levels of HDL-C and tryglyceride. [16] Information on PHT and lipid profiles in diabetic and nondiabetic women is limited, however, especially in American Indian women. We investigated this issue in the present study.” Page 156. | | | | | |
| **Study design** | **Conflicts of interest** | | **Inclusion criteria** | | **Exclusion criteria** |
| Cross-sectional study. | N/A; none listed. | | American Indian female aged 45–74 years; free of CVD Abbreviation: CVD—cardiovascular disease. | | “Those who were using lipid-lowering drugs or who had missing lipid/lipoprotein values at the second examination were excluded from the analyses of lipid profiles.” Page 157. |
| **Population and sampling description** | | **Indigenous description** | | **Total number of participants** | |
| The population of interest was American Indian women.  This record represents a cross-sectional analysis from a larger longitudinal study. Details about sampling were not included in this report. | | “American Indians from 13 tribes/communities located in southwestern Oklahoma, central Arizona, and North and South Dakota.” Page 156.  The authors do not describe how they defined American Indian racial/ethnic background beyond stating that they sampled participants located in Oklahoma, Arizona, and the Dakotas.  The percentage of the sample who were Indigenous is 100%.  Note: About one third of the postmenopausal women reported hysterectomy.” Page 157. | | The total sample size was 1446 and the total number of Indigenous participants was 1446. Page 156. | |
| **Study outcomes and important results** | | | | | |
| “Results: Among nondiabetic women, current PHT users had lower mean fibrinogen, PAI-1, and low-density lipoprotein cholesterol (LDLC) levels than those in never users (38.4 mg/dl, 8.68 ng/ml, and 14.16 mg/dl lower, respectively) but higher CRP and triglyceride levels (1.53 mg/l and 31.43 mg/dl higher, respectively). Multivariate adjustment did not alter any of these associations. In diabetic women, current PHT use was associated only with lower PAI-1 (5.48 ng/ml lower) and higher high-density lipoprotein cholesterol (HDLC) levels (3.33 mg/dl higher) compared with never users. Conclusions: In American Indian women without diabetes, PHT was associated with lower levels of hemostatic markers but higher levels of an inflammatory marker. Associations were less marked in women with diabetes. The relation of PHT with lipid profiles also differed in nondiabetic and diabetic women. These data provide an additional rationale for considering diabetes status when deciding whether or not to use PHT.” From the abstract. Abbreviations: PHT—postmenopausal hormone therapy; PAI-1—plasminogen activator inhibitor type 1; CRP—C-reactive protein.  “Demographic characteristics of the PHT groups are shown in Table 1. Compared with never users, current users and past users were significantly younger and better educated, with a lesser degree of American Indian heritage, lower diabetes prevalence, higher proportion of surgical menopause, lower parity, and lower waist/hip ratio. Compared with never users, current users had a significantly higher proportion of alcohol use and a lower mean BMI. The distribution of never users, past users, and current users differed significantly among the three geographic areas. Oklahoma had the highest proportion of current users.” Page 158.  “After adjustment for all covariates, the average fibrinogen for current hormone users was 38.4 mg/dl lower than that of never users, and the average PAI-1 level for current users was 8.68 ng/ml lower. However, the average CRP level of current users was 1.53 mg/L higher than that of never users.” Page 158.  “In diabetic participants, current hormone use was also related to lower PAI-1 levels (5.48 ng/dl less than never users) (Table 2), and there was no significant interaction between PHT and diabetes in the association with PAI-1. The mean CRP level in current users was higher than that in never users in both univariate and adjusted models, but the differences were not statistically significant (p=0.19 and p=0.11, respectively). There was no significant association between hormone use and fibrinogen levels in diabetic postmenopausal women after multivariate adjustment. No relation between duration of hormone use and fibrinogen, CRP, and PAI-1 concentration was seen (data not shown).” Pages 158 and 159.  “Table 3 shows that in nondiabetic women, current PHT use is related to lower LDLC (14.16 mg/dl lower) and higher triglyceride (31.43 mg/dl higher) levels compared with never users. These associations were not changed after adjustment for other risk factors. After adjustment for other risk factors, however, PHT was not significantly related to HDLC. The results for HDLC were reversed in diabetic participants. Current PHT use was only related to increased HDLC (3.33 mg/dl higher than never users) in diabetic participants regardless of multivariate adjustment. The associations of PHT and LDLC and triglyceride were not statistically significant in diabetic participants.” Page 159.  “In nondiabetic women, we found that current PHT was associated with lower fibrinogen, PAI-1, and LDLC levels but higher CRP and triglyceride levels compared with never users, after adjustment for covariates. The mechanism of those changes needs to be studied. Some changes may be related to endothelial function. [5,27]” Page 159.  “In diabetic postmenopausal women, the relation between hormone use and hemostatic and inflammatory markers appeared to be attenuated. PAI-1 was significantly lower in current users than in never users, but there were no significant differences between current users and never users in mean fibrinogen or CRP levels. Also, the association of PHT and lipid profiles was different in diabetic women than in nondiabetic women. PHT was related to higher HDLC in diabetic women. LDLC and triglyceride levels were not associated with PHT in diabetic women.” Page 159. | | | | | |
| **Use of STRAW+10** | **Menopausal transition staging of sample using STRAW+10** | | | | |
| The STRAW+10 not used. | Postmenopause.  “Postmenopausal women included both those who had a natural menopause and those who had a surgical menopause. Natural menopause was defined as being at least 12 months beyond the last menstrual period. If a participant underwent a hysterectomy, she was asked if only her uterus had been removed. Women who reported hysterectomy without oophorectomy or who were not sure if their ovaries had been removed were included as postmenopausal if they were 53 years old, as 90% of the women in this cohort reached menopause by this age.” Page 157. | | | | |
| **Author reported facilitators to treatment** | | | **Author reported barriers to treatment** | | |
| N/A. | | | N/A. | | |
| **Study limitations** | | | | | |
| N/A; none described. | | | | | |

| **Record ID** | **Title** | | | | |
| --- | --- | --- | --- | --- | --- |
| #1583 – 1981 | On the calculation of climacteric symptoms | | | | |
| **Author information** | | | | | **Country** |
| Wright, A. L. | | | | | United States |
| **Extraction note** | | | | | |
| This is a methods paper to test a hypothesis. The author justifies their selection of the Navajo population. Although this paper did have a 100% Indigenous sample, this record addresses the author’s hypothesis. The author argues that the present approach, which uses qualitative data, is more accurate than using the survey approach used at the time of this publication: “data collection techniques used in the present study permit the required evaluation by (a) asking respondents about potential non-physiological causes of symptoms, (b) knowledge of their life situation, and (c) review of medical records, as well as controlling for the effect of expectation in symptom reporting. In addition, an indicator was developed for use in the present study which counted only climacteric-related symptoms, eliminating those complaints which stem from other non-physiological factors.” Page 6. | | | | | |
| **Study aims** | | | | | |
| “The question, therefore, arises as to how to distinguish symptoms attributable to the physiological changes of the climacteric from those which may stem from the process of ageing or other exogenous factors. ... The goal here is to suggest possible improvements in survey techniques which could permit such an evaluation. This paper opens with a consideration of some common methods of arriving at estimates of symptom experience and their disadvantages. It then discusses both data collection techniques and a means of incorporating the resultant information in measures of symptom prevalence, which were developed during a study of the climacteric experience among a group of American Indians. Symptom prevalence as estimated by the different counts is subsequently reported, and interpretations of these results are provided.” Pages 55 and 56. | | | | | |
| **Study design** | **Conflicts of interest** | | **Inclusion criteria** | | **Exclusion criteria** |
| Other: mixed methods—use of qualitative data to distinguish symptoms attributable to the physiological changes of the climacteric from those which may stem from the process of ageing or other exogenous factors. | N/A; none listed. | | N/A; none listed. | | N/A. |
| **Population and sampling description** | | **Indigenous description** | | **Total number of participants** | |
| The population of interest was “American Indians;” specifically “middle-aged Navajo women.” Page 57.  No details are described for the sampling procedures. A previous publication is cited: “Anthropological field-work was conducted on their reservation in northeastern Arizona on two occasions, in 1976 and 1978, with the latter visit focusing on the experience of menopause [l0].  Reference:  Wright, A.L. (1980) Cultural variability in the experience of menopause: a comparison of Navajo and western cases. University Microfilms International, Michigan. | | Navajo middle-aged women. Page 57.  Besides stating that participants were sampled on a “reservation in northeastern Arizona,” the author does not detail how Navajo was defined.  The percentage of the sample who were Indigenous was 100%.  “The women were of two types: ‘traditional’ and ‘acculturated’. The former were uneducated, spoke no English, lived in a rural area and were not employed in the wage economy. ‘Acculturated’ women, on the other hand, held wage-paying jobs, spoke English, had completed high school and lived in towns on the reservation. Given potentially different experiences for woman who experienced surgical menopause, the field sample of 80 women was reduced to 38 ‘traditional’ and 28 ‘acculturated’ women who achieved menopause naturally.” Page 57. | | The total number of participants was 66, of which 38 were “traditional” and 28 were “acculturated.” Page 57. | |
| **Study outcomes and important results** | | | | | |
| “Comparisons of these measures of symptom experience with each other led to the following results. First, virtually all respondents reported that there were no ‘bodily changes’ since menopause in response to the open-ended question. However, when they were subsequently asked if they experienced the symptoms in the checklist, most responded affirmatively.” Page 60.  “The results discussed above support several of the contentions made previously, as well as providing information concerning the reliability of these two types of symptom counts. The first result, that individual women reported symptoms in response to the checklist which were not elicited by an open-ended question, supports the notion that cultural expectations may affect the reporting of symptoms. As discussed elsewhere [ll], the Navajo have an unelaborated concept of the menopause, not associating any symptoms with the cessation of menstruation. The differing response of these women to the two questions may be attributed to this belief. This interpretation has implications for both research and health care. It suggests that researchers in all cultures should ask explicitly if a respondent has experienced specific symptoms, rather than asking generally if there have been any health problems or bodily changes since the ending of the periods. This finding also suggests that the reporting of climacteric symptoms to physicians is affected by culturally specific expectations and beliefs about the relationship of these complaints to the menopause. On the other hand, the decrease in symptom prevalence as estimated by the climacteric related counts suggests that gross symptom counts among these women were elevated by inclusion of complaints which were not physiologically related to the climacteric. This result may be applicable to other studies as well: symptom prevalence among the Navajo is very similar to that reported for western women only when gross counts are compared [12]. If symptoms attributable to non-physiological causes were eliminated in these other studies as well, our estimates of symptom prevalence in the climacteric might decrease substantially.” Page 61. | | | | | |
| **Use of STRAW+10** | **Menopausal transition staging of sample using STRAW+10** | | | | |
| The STRAW+10 not used. | Unclear.  The sample included middle-aged women. | | | | |
| **Author reported facilitators to treatment** | | | **Author reported barriers to treatment** | | |
| N/A. | | | N/A. | | |
| **Study limitations** | | | | | |
| “These techniques do not, of course, resolve the problem of over-estimation of symptoms by virtue of suggestion.” Page 62. | | | | | |

| **Record ID** | **Title** | | | | |
| --- | --- | --- | --- | --- | --- |
| #1567 – 2013 | Health professionals working with First Nations, Inuit, and Métis consensus guideline | | | | |
| **Author information** | | | | | **Country** |
| Wilson, D.; de la Ronde, S.; Brascoupe, S.; Apale, A. N.; Barney, L.; Guthrie, B.; Harrold, E.; Horn, O.; Johnson, R.; Rattray, D.; Robinson, N.; Alainga-Kango, N.; Becker, G.; Senikas, V.; Aningmiuq, A.; Bailey, G.; Birch, D.; Cook, K.; Danforth, J.; Daoust, M.; Kitty, D.; Koebel, J.; Kornelsen, J.; Tsatsa Kotwas, N.; Lawrence, A.; Mudry, A.; Senikas, V.; Turner, G. T.; Van Wagner, V.; Vides, E.; Wasekeesikaw, F. H.; Wolfe, S.; Aboriginal Health Initiative Committee, Special Contributors Endorsing Organizations Aboriginal Nurses Association of Canada Canadian Association of Midwives Canadian Association of Perinatal; Women's Health Nurses, Indigenous Physicians Association of Canada Inuit Tapiriit Kanatami Metis National Council Minwaashin Lodge National Aboriginal Council of Midwives National Aboriginal Health Organization Native Women’s Association of Canada Native Youth Sexual Health Network Pauktuutit Inuit Women of Canada Royal College of Physicians; & Surgeons of Canada, Society of Rural Physicians of Canada | | | | | Canada |
| **Study aims** | | | | | |
| “Objective: Our aim is to provide health care professionals in Canada with the knowledge and tools to provide culturally safe care to First Nations, Inuit, and Métis women and through them, to their families, in order to improve the health of First Nations, Inuit, and Métis.” Abstract. | | | | | |
| **Study design** | **Conflicts of interest** | | **Inclusion criteria** | | **Exclusion criteria** |
| Text and opinion. | “Disclosure statements have been received from all contributors.” Page 551. | | Inclusion criteria for the literature review: “Published literature was retrieved through searches of PubMed, CINAHL, Sociological Abstracts, and The Cochrane Library in 2011 using appropriate controlled vocabulary (e.g., cultural competency, health services, indigenous, transcultural nursing) and key words (e.g., indigenous health services, transcultural health care, cultural safety). Targeted searches on subtopics (e.g., ceremonial rites and sexual coming of age) were also performed. ... Searches were updated on a regular basis and incorporated in the guideline to May 2012. Grey (unpublished) literature was identified through searching the websites of selected related agencies (e.g., Campbell Collaboration, Social Care Online, Institute for Healthcare Improvement).” Abstract. | | “The PubMed search was restricted to the years 2005 and later because of the large number of records retrieved on this topic.” Abstract. |
| **Population and sampling description** | | **Indigenous description** | | **Total number of participants** | |
| The population of interest was First Nations, Inuit, and Métis women.  Sampling details are not applicable as this is a consensus guideline paper. | | First Nations, Inuit, and Métis peoples were the focus of this consensus guideline paper. | | N/A. | |
| **Study outcomes and important results** | | | | | |
| “Health professionals should be aware that there is a great lack of research, resources, and programming about mature women’s health issues, including menopause, that is specific to First Nations, Inuit, and Métis. Health professionals should advocate for further research in this area.” Page 553. | | | | | |
| **Use of STRAW+10** | **Menopausal transition staging of sample using STRAW+10** | | | | |
| The STRAW+10 not used. | N/A; menopause was not a primary focus of this consensus guideline paper. | | | | |
| **Author reported facilitators to treatment** | | | **Author reported barriers to treatment** | | |
| Establishing an environment of trust and cultural safety: “Demographically, First Nations, Inuit, and Métis peoples are younger and more mobile than non-Aboriginal people. This requires extra effort on the part of health care professionals to establish an environment of trust and cultural safety in their workplaces as the opportunity to provide care may be brief.” Page 551.  “Research has shown that where cultural competency strategies have been implemented, health outcomes and patient satisfaction have improved.” Page 552.  “Health professionals who provide care to First Nations and Inuit should be aware of the Non-Insured Health Benefits program, its eligibility and coverage requirements, and the exceptions and special permissions needed in some cases. Health professionals should recognize that they have a vital role in advocating for their First Nations and Inuit patients and assisting with obtaining these benefits. Health professionals should be aware that Métis do not have access to the Non-Insured Health Benefits and may face unique challenges accessing health care.” Page 552. | | | “The unemployment rate is much higher in Aboriginal communities than in those of non-Aboriginal Canadians. This is a major contributor to the gaps in socioeconomic status and access to equitable and quality health care.” Page 551.  “Jurisdictional conflicts between federal, provincial, territorial, and band governments make it difficult to provide comprehensive public health and health services to First Nations.” Page 551. | | |
| **Study limitations** | | | | | |
| “These techniques do not, of course, resolve the problem of over-estimation of symptoms by virtue of suggestion.” Page 62. | | | | | |

| **Record ID** | **Title** | | | | |
| --- | --- | --- | --- | --- | --- |
| #1542 – 2002 | Aboriginal women and menopause | | | | |
| **Author information** | | | | | **Country** |
| Webster, R. W. | | | | | Other: Results include Indigenous women living in Canada and the United States (and Mexico). |
| **Extraction note** | | | | | |
| A limitation of our study is that we did not include books in our results, such as those included in this review. | | | | | |
| **Study aims** | | | | | |
| “Objective: To determine the characteristics of menopause in Aboriginal women, in particular Canadian Aboriginal women.” Abstract.  “Little is known about menopause in Aboriginal women. The purpose of this review is to determine the characteristics of menopause in Aboriginal women, in particular Canadian Aboriginal women.” Page 938. | | | | | |
| **Study design** | **Conflicts of interest** | | **Inclusion criteria** | | **Exclusion criteria** |
| Other: narrative review. | “Competing interests: None declared.” Page 938. | | Inclusion criteria for the literature review: “An extensive review of medical and non-medical databases was undertaken. For each database, the search strategy combined the key word ‘menopause’ with at least one of the following terms: Aboriginals, Native Americans, Natives, Indians, Metis, Inuit, Eskimo, and Indigenous people. Eleven databases were searched: CINAHL, eHRAF Collection of Ethnography, First Nations Periodical Index, MEDLINE, Native Health History Database, Native Health Research Database, AgeLine, POPLINE, Periodical Contents Index (PCI), Social Science Abstracts Full Text, and the Cochrane Library. All articles were reviewed, and those that provided information with respect to the study objective were summarized and the key points recorded.” Page 939. | | N/A; none listed. |
| **Population and sampling description** | | **Indigenous description** | | **Total number of participants** | |
| The objective states Canadian Aboriginal women were the focus of this narrative review; however, the results include participants who lived in Canada, the U.S., and Mexico. As this is a narrative review and not primary research, this record does not include any primary participants of research. | | Although this narrative review aimed to focus on Canadian Aboriginal women, the review includes results for participants living in Canada, the U.S., and Mexico. The search strategy included the terms: “aboriginals, Native Americans, Natives, Indians, Metis, Inuit, Eskimo, and Indigenous people.” Page 938.  The author does not discuss details about racial/ethnic definitions.  As this is a narrative review and not primary research, there are no details to report about the percentage of the sample who were Indigenous. | | N/A; this is not primary research. However, the total number of records identified by the author’s search was 13. | |
| **Study outcomes and important results** | | | | | |
| “This review shows the limitations in our current knowledge about menopause in Aboriginal women and indicates the need for further study of Canadian Aboriginal women with respect to symptoms of menopause and the health consequences seen postmenopausally.” Page 940.  “The studies reviewed suggest the transition through menopause seems to have little effect on Aboriginal women’s lives, [4-10,13,16] and that if there is an effect, it seems to be a positive one in that it increases freedom to do activities the women deem important. In addition, while it appears that Aboriginal women experience fewer vasomotor symptoms than their Caucasian counterparts (level II-2 in rural New Mexico[I6]), it is not known if they experience other symptoms of menopause, such as those related to the urogenital tract or psychological issues. The fact that the difference between Aboriginal and non-Aboriginal women in symptoms such as hot flushes does not appear to be biologically caused suggests culture, tradition, and ethnicity play a role in the reporting of vasomotor symptoms. With respect to the age of menopause in Aboriginal women, there is little evidence to suggest a difference from non-Aboriginal women.” Page 940. | | | | | |
| **Use of STRAW+10** | **Menopausal transition staging of sample using STRAW+10** | | | | |
| The STRAW+10 not used. | Menopausal Transition and Postmenopause. | | | | |
| **Author reported facilitators to treatment** | | | **Author reported barriers to treatment** | | |
| N/A. | | | N/A. | | |
| **Study limitations** | | | | | |
| N/A; none listed in the context of this record. The author describes the limitations of the literature reviewed. | | | | | |

| **Record ID** | **Title** | | | | |
| --- | --- | --- | --- | --- | --- |
| #1440 – 2021 | Menopause experience in First Nations women and initiatives for menopause symptom awareness; A community-based participatory research approach | | | | |
| **Author information** | | | | | **Country** |
| Sydora, B. C.; Graham, B.; Oster, R. T.; & Ross, S. | | | | | Canada |
| **Study aims** | | | | | |
| “In response to the wishes of Cree women living in Maskwacis, Alberta, to start a dialogue on menopause, we undertook community-based participatory research (CBPR) to explore menopause experience and raise awareness of menopause symptoms in the community.” Abstract.  “Our preliminary community-based participatory research (CBPR) goals were to engage with women community members to explore in a series of workshops their perceptions and experiences of menopause and the impact of menopause on their QOL. The subsequent knowledge translation efforts responded to the findings of the initial research with the goal of benefiting community members by raising awareness about menopause symptoms and experience.” Page 2. | | | | | |
| **Study design** | **Conflicts of interest** | | **Inclusion criteria** | | **Exclusion criteria** |
| Other: CBPR multiple methods (focus groups and questionnaire). | “Competing interests. The authors declare that they have no competing interests.” Page 10. | | “Our research aimed to involve women in menopause transition age (40–65) from any of the four Nations who were interested in sharing their experiences of menopause with other women.” Page 3. | | N/A; none listed. |
| **Population and sampling description** | | **Indigenous description** | | **Total number of participants** | |
| The population of interest was “First Nations women living in the community of Maskwacis, Alberta.” Page 2.  “Recruitment posters were displayed in health centers in Maskwacis and Pigeon Lake. Women were also recruited by a local hired research assistant for the first two workshops; subsequent recruitment occurred through word of mouth.” Page 3. | | “First Nations women living in the community of Maskwacis, Alberta.” Page 2.  “The community of Maskwacis, which includes the hamlet of Pigeon Lake, is situated 90 km south of Edmonton and includes four distinct Cree First Nation Bands: Ermineskin Cree Nation, Samson Cree Nation, Louis Bull First Nation, and Montana First Nation.” Page 2.  The authors do not describe how they defined participants’ racial/ethnic background of Cree First Nations. They do state that, “All women lived on reserve.” Page 4 And, on page 5, Table 1 “Characteristics of participants from initial 5 menopause workshop,” “Native heritage” is described for the participants: 19 (55.9%) participants reported “Both parents Cree”; 13 (38.2%) participants reported “One parent Cree”; 1 (2.9%) participant reported “Both parents other native”; and, last, 1 (2.9%) participant did not report their Native heritage. Page 5 The percentage of the sample who were Indigenous was 100%. | | The total number of participants who participated in the menopause workshops was 37 and the total number of Indigenous participants was 37.  “Questionnaire data were available from 34 participants.” Page 4. | |
| **Study outcomes and important results** | | | | | |
| Workshop discussions “1. Menopause symptom experience. Participants talked very openly about their own experience of menopause: their experiences of menopause onset and symptoms varied widely. The majority of women talked about experiencing menopause symptoms: most could relate to hot flashes and night sweats. Hot flashes were sometimes extreme; one woman described her feeling as ‘burning inside and out’, while another woman described it as getting hot from the inside ‘like firecracker’. Sometimes the main symptom was their face flushing very red. Night sweats could also be extreme, causing women to be drenched in sweat. One woman described an occasion when a new nightdress leaked dye into clean sheets—truly ‘leaving your mark’. Many women complained about hot feet that required cooling off. One said that she could ‘start a fire’ with her feet when walking. Others complained they feel the cold much more now compared to when they were younger indicating the limited window of comfortable temperature commonly experienced during menopause. Participants described unpredictability in menstrual blood flow which caused embarrassment and anxiety. Blood flow changes could include both decreases, as well as significant increases in flow. In extreme cases, blood could flow ‘like a waterfall... need to change [menstrual] pads every 30 min’. For some women the variation in flow caused them to worry that it may indicate a more serious cause (e.g. cancer). Heavy bleeding was sometimes experienced in connection with severe pain that required medical interventions including hysterectomy. The impact of hysterectomy (with or without ovaries being removed) and ‘tied tubes’ was of concern; women discussed how these procedures affected menopause symptoms and how they may influence their future health. Vaginal dryness was experienced by a number of women and could affect their sex drive. Lack of desire for sex was worrying, particularly for women who had previously enjoyed a healthy and fulfilling sex life. They wondered whether and when their previous desires would return, and were concerned because their partner and relationship suffered. Many participants experienced physical symptoms related to weakness, stiff and achy joints, and sleep deprivation. Insomnia was often associated with night sweats leading to sleep disruptions. Uncontrollable weight gain was also experienced by some women much to their dismay. Other symptoms included changes to skin and hair texture and hair growth where it was not seen before, such as on face, chin or lengthening of eyebrows. These symptoms were not always recognized as being associated with menopause. Psychological symptoms such as mood swings, anger, depression, and anxiety were also common, unpredictable and worrisome. ‘Sometimes you cry all the time or get mad about trivial things’. These psychological symptoms and mood swings were a concern, specifically because of their impact on family life and relationships, and were often connected to other life events such as children leaving home and the start of a new phase in family life. One woman described her newly experienced contentment with being alone; another woman mentioned her withdrawal from gathering with people and socializing; one woman mentioned occasional suicidal thoughts. Some women complained about memory loss and forgetfulness. This prompted a discussion about menopause-specific versus age-related symptoms. During one workshop, benefits of menopause were discussed. One woman described how activities were less restricted post-menopause. A major constraint before menopause was not being able to take part in ceremonies (sweats, dancing, etc.) while having one’s moon time (menstruation) and for four days afterwards” Page 4.  “2. Menopause knowledge prior to own experience. Most participants felt they had not acquired sufficient information about menopause and symptoms before their own experience, because menopause was ‘not talked about’ by their mothers, aunts, grandmothers or by their doctors. Several women reported that their mothers, when asked directly about menopause symptoms, declared that menopause was not a problem, ‘it was really nothing’, or endured their symptoms stoically. The women discussed whether their mothers were in denial of menopause symptoms, viewed them as the consequences of a natural, invariable stage of their life, or whether they really had forgotten about their symptoms. One woman suggested that in the past, menopause symptoms may not have been as obvious because people were too busy to deal with symptoms, and sweating was usual when women were occupied doing physical work. Observing other women experiencing menopause has helped some women prepare for their own menopause. Participants remembered occasions when as young girls they were seeing their mothers, a class teacher, an older female friend or relative experiencing mood swings or hot flashes and being drenched in sweat, ‘like someone turning on a tap’, without knowing what was happening at the time. One woman said she felt grateful to have observed others experience these symptoms. A few women mentioned their older sister or a friend as the source of first menopause knowledge. In one workshop women discussed whether menopause disregard prevents worry about symptoms and symptom awareness. While this hypothesis was considered by some, the majority agreed with another woman’s statement: ‘it is the other way around: if we know what to expect, then we would worry less’.” Pages 6 and 7.  “3. Menopause symptom management. While going through their own menopause transition, some participants were prepared to deal with symptoms. One woman found that exercise (specifically walking) was very helpful to control mood swings and calm her thoughts during menopause. Strategies for hot flashes included going outside in winter, leaning against cold concrete walls, or walking on cold surfaces to cool feet. Some women stated that they take changes of clothes with them in case they sweat profusely; others were prepared with extra towels for night sweats. An older woman advised younger women: ‘Don’t think about it, and it won’t bother you. You can cope well... Open your windows.’ For unpredictable menstruation some peri-menopausal women carried extra menstrual products. Several women knew of others who used traditional herbs and other natural remedies for treatment of menopause symptoms. There were discussions about herbal and root teas used by Elders in the past for a variety of conditions. One woman said that her mother is a healer who knows about treatments for many illnesses. Another woman regretted not learning from her, now deceased, mother-in-law, who was very knowledgeable about plants and traditional Indigenous medicines. Other traditional practices to reduce the impact of menopause were taking good care of feet and not going out with wet hair during menopause. Some women had seen others eating dirt, ash or earth during menopause, perhaps in response to a deficiency that the body was trying to correct.” Page 7.  “4. Impact of menopause on family members. A major concern for workshop participants was the effect of severe symptoms on their family relationships, particularly their partner-relationships. It was acknowledged by all that menopause can be a strain on the relationship with a partner. The husband/partner may not understand symptoms such as significant mood swings and decreased libido, leading to matrimonial disharmony. One woman jokingly referenced the impact on male partners: ‘that’s why it’s called MENopause!’ Decline in sex drive was of significant concern for women—they felt this was of major importance to their partners, but could not force themselves to be interested in sex, despite knowing this could adversely impact their relationships and potentially cause partners to think their wives no longer care for them or might be unfaithful. One woman complained that she was sick of always apologizing to her husband for her mood swings. Participants noted that other family members could also be affected by menopause symptoms. Several women felt they were not understood by their family members, especially because of their unpredictable mood swings, depression and anger. These symptoms could cause strife within the family, particularly if the family members did not understand the cause, or if they were undergoing their own hormonal changes (e.g. puberty). Participants felt that they can only now relate to earlier uncomfortable observations of mother’s, aunt’s or grandmother’s menopausal symptoms. One woman said she has told her children, ‘Don’t get scared of me, I get scared of myself!’ Participants felt that, if male partners and family members learned about the effects of menopause, they would be more understanding about changes to women’s behavior as they go through the transition. They believed there was a need to educate spouses, children, and family members about menopause symptoms. ‘This is important. I want my family, my daughters and granddaughters to know what [physical changes are] coming’ to prepare family members to deal with apparently inexplicable changes that can occur.” Page 7.  From Table 1 “Characteristics of participants from initial 5 menopause workshop,” page 6, “Menopause therapies and symptom management” for the sample is described: 12 participants (35.2%) reported “Do not currently use anything in particular to manage symptoms,” 6 participants (17.6%) reported “Lifestyle activities for menopause therapy,” 4 participants (11.8%) reported “Cultural therapies (Sweats and Native Medicine),” 4 participants (11.8%) reported “Herbs or vitamin supplements,” 4 participants (11.8%) reported “Seek advice from friends or relatives,” 3 participants (8.8%) reported “Anti-depressants,” 1 participant (2.9%) reported “Seek advice from health care workers,” 1 participant (2.9%) reported “Menopausal Hormone Therapy (MHT; vaginal),” 1 participant (2.9%) reported “Over-the-counter remedies,” 1 participant (2.9%) reported “Relaxation therapy.” Multiple responses were allowed.  From Table 1 “Characteristics of participants from initial 5 menopause workshop,” page 6, “Considered management if symptoms become severe and bothersome” for the sample is described: 7 participants (20.6%) reported “Seek advice from health care workers,” 6 participants (17.6%) reported “Seek advice from friends or relatives,” 2 participants (5.9%) reported “Intend to practice cultural forms of relief,” 1 participant (2.4%) reported “Consider hormonal therapy,” 1 participant (2.4%) reported “Would consider over-the-counter,” 9 participants (26.5%) reported “Not sure yet,” and, last, 4 participants (14.7%) reported “Nothing in particular.” Multiple responses were allowed.  Questionnaire responses “1. Menopause treatment and symptom management. Questionnaire data revealed that the majority of women were reluctant to use drugs such as hormone therapy or over-the-counter remedies, rather choosing to make lifestyle changes, use cultural therapies or herb and vitamin supplements, or use no specific management option (Table 1). The questionnaire results corroborated the qualitative findings from the workshops that women preferred modest, non-medical strategies. Considering future managements, after participating in the workshops, women were more ready to discuss menopause symptoms with friends or relatives or seek advice from health care professionals (Table 1).” Page 7.  “2. Impact of menopause symptoms on QOL. The mean average MENQOL scores ranged from 3.3 (vasomotor domain) to 4.1 (sexual domain) indicating that quality of life on average was negatively impacted during the menopause transition period for workshop participants (Table 2). The MENQOL results also validate the experiences women discussed in the workshops, describing mainly the uncomfortable aspects of menopause.” Pages 7 and 8.  Knowledge translation “1. Formation of Sohki Tehew group and implementation of menopause awareness strategies. A key qualitative finding was a need for increased knowledge about menopause within the community.” Page 8.  “2. Development of two pamphlets. Two culturally-sensitive pamphlets were developed by the Sohki Tehew Group for use within the community, based on the findings of the qualitative study. The wording and layout for both pamphlets are very similar, but targeted at different audiences: one for husband/partner, the other for women and other family members (Additional files 2, 3: Appendix 1 - Menopause information for men and Appendix 2 - Menopause information for women and families). Several sections are designed to help understand about the "Change of Life" i.e. menopause” Page 8.  “3. Implementation. Following the development of the pamphlets, hundreds of copies have been distributed through Maskwacis Health Services and in the Health Clinics. In addition, the Sohki Tehew Group has taken pamphlets to many community events in the four Nations and Pigeon Lake, including Sohki Tehew Workshops, Powwows, Health Fairs, Indigenous Addictions Week, and Maskwacis Community College and school library events.” Page 8.  From the Discussion section: “Few used cultural or other medication to address those symptoms, preferring to use modest physical methods such as cooling strategies to reduce the effect of hot flashes. These choices may be partly due to the loss of a dialogue between generations about female topics as a result of residential schools and westernization that have deprived women of their status [29].” Page 9. | | | | | |
| **Use of STRAW+10** | **Menopausal transition staging of sample using STRAW+10** | | | | |
| The STRAW+10 not used. | Reproductive, Menopausal Transition, Postmenopause.  While the inclusion criteria were “women in menopause transition age (40–65)” (Page 3), the results section reveals the sample included women from a range of reproductive stages: “Among women who completed the questionnaire the majority of women were post-menopausal (absence of menses for>1 year) (n=23), six had irregular menses (peri-menopause) and five still experienced regular menses (pre-menopause).” Page 4 | | | | |
| **Author reported facilitators to treatment** | | | **Author reported barriers to treatment** | | |
| N/A. | | | N/A. | | |
| **Study limitations** | | | | | |
| “Our study has several limitations and strengths. The research was conducted in one Indigenous community, and involved only 37 women in the research workshops. The participating women may not be representative of the population of Maskwacis women in menopause transition stages; the workshops may have mostly attracted women with moderate to severe symptoms and menopause-related health issues. Although the MENQOL questionnaire result correlates with the perceived menopause symptom-related reduced QOL described by the women, this association should be viewed with caution as the MENQOL questionnaire to date has not been specifically validated for use in Indigenous groups. The small workshop size and community-specific location could be considered a limitation; however, it may also be a strength because the women were very open in their discussions, providing rich information about their experiences, perhaps because they were comfortable with each other.” Page 9. | | | | | |

| **Record ID** | **Title** | | | | |
| --- | --- | --- | --- | --- | --- |
| #1415 – 1995 | Puberty, pregnancy, and menopause: Lifecycle acculturation in a Copper Inuit community | | | | |
| **Author information** | | | | | **Country** |
| Stern, P. R.; & Condon, R. G. | | | | | Canada |
| **Study aims** | | | | | |
| “Adult women living today in the Inuit community of Holman, N.W.T. fall into three distinct age cohorts-- elders (women in their 50s, 60s and 70s), middle-aged women (in their mid-30s and 40s), and young adults (under 35). These three groups of women are defined not only by age, but, more importantly, by the social and physical context of their life histories. ... The three groups of women gained their knowledge about reproductive health under distinct social conditions and from very different sources. Consequently, their understandings and attitudes regarding menstruation, pregnancy, infant care and their own aging vary considerably. This paper will consider each group in terms of its knowledge and opinions regarding women’s health concerns. We will attempt to explain how both Inuit culture and specific historical factors have combined to create these disparities and consider the likely consequences for current and future generations of Inuit women.” Pages 21 and 22. | | | | | |
| **Study design** | **Conflicts of interest** | | **Inclusion criteria** | | **Exclusion criteria** |
| Qualitative research. | N/A; none listed. | | N/A; none listed. | | N/A. |
| **Population and sampling description** | | **Indigenous description** | | **Total number of participants** | |
| The population of interest was Copper Inuit women living in the Central Canadian Arctic village of Holman. | | Three generations for Copper Inuit women from the Canadian Arctic village of Holman: elders aged 50 and older, middle-aged women (aged 35 to 49), and young adults under the age of 35.  The authors do not describe how “Copper Inuit” was defined except that they were living in the village of Holman. “The community of Holman is located on the western coast of Victoria Island in the Central Canadian Arctic. Its approximately 400 residents are primarily descendants of the two northernmost groups of Copper Inuit who were among the last groups in the North American Arctic to be contacted by Eurocanadians. The community also includes a number of Inuit families from the Western Arctic (North Alaska and the Mackenzie Delta region) who arrived in the 1930s and 1940s.” Page 22.  The percentage of the sample who are Indigenous is 100%  Additional details about the sample: “The oldest group of women grew up in isolated hunting and fishing camps. The camps were seasonal and nomadic ... Most received no schooling, and though most understand spoken English, few of these women are comfortable conversing in anything other than Inuinaqtun. Almost without exception, these women are the primary generators of cash for their households, using their traditional skills to produce items for sale to consumers of native art” Page 24.  Middle-aged women: “Most of the women in this cohort are bilingual in English and Inuinaqtun. While they have managed to main­tain many traditional skills, for the most part the focus of their daily activities is within the settlement rather than on the land. Many hold non-traditional jobs that bridge the gap between Inuit and Western culture such as social workers, interpreters, and managers. While the youngest members of this group attended school (at least irregularly) in town, the oldest members spent late childhood and early adolescence in boarding schools in Aklavik, Inuvik, and Yellowknife. Some spent as little as two years away from their families, while others were gone up to six years, returning only for summer vacations. When they returned home to stay, many found that their families had given up camp life in favor of the security of the settlement.” Page 26.  Young adults under the age of 35: They were “raised entirely within the context of the settlement. ... They vary considerably in the extent of their mastery of traditional skills. ... Most speak little or no Inuinaqtun and less than half a dozen consider themselves fluent in the language spoken by their parents and grandparents. They have more formal education than any other group (and many have attended high school in Yellowknife), but a chronic shortage of wage labor jobs in Holman and their own lack of traditional skills severely limits their employment opportunities.” Page 27. | | The total sample size is 106 and the total number of Indigenous participants is 106.  The elders group included 17 participants. The group of middle-aged women, aged 35 to 49, included 31 participants. The group of young adults under the age of 35 included 58 participants. | |
| **Study outcomes and important results** | | | | | |
| For elders aged 50 and older: “Menopause, for all of this cohort, occurred well, after they were integrated into the western medical system. Some knew to expect the physical and emotional changes associated with menopause, but many were taken completely by surprise (Audrey Woodget 1993, personal communication). One woman reported that, after she went to the nurse to discuss the heaviness she felt in her limbs, she remembered being told long before that one day her menses would stop. Presumably, had camp life continued, these women would have received training for menopause in much the same way that they learned of menstruation and pregnancy.” Pages 25 and 26.  Middle-aged women: “The oldest members of this group are just now approaching menopause. Unfortunately, we have no data regarding their knowledge of or attitudes towards this lifestage.” Page 27.  Young adults under the age of 35: “Almost without exception, these women have received their knowledge of reproductive health from Eurocanadian sources, primarily nurses and teachers, but also magazines and television. Consequently, like the elderly women, their understanding appears to be far more thorough than that of the middle-aged women. The information, however, has been filtered through a Eurocanadian cultural template rather than an Inuit one. Most know nothing of the pregnancy and childbirth experiences of their mothers and grandmothers. Nor do they have any understanding of the way their ancestors utilized and manipulated the natural environment for medical purposes.” Page 27. | | | | | |
| **Use of STRAW+10** | **Menopausal transition staging of sample using STRAW+10** | | | | |
| The STRAW+10 not used. | Reproductive, Menopausal Transition, and Postmenopause. | | | | |
| **Author reported facilitators to treatment** | | | **Author reported barriers to treatment** | | |
| “Nearly all scholars of lnuit culture have remarked upon the great flexibility inherent in the society (13,14). This flexibility allowed Inuit culture to survive for centuries in one of the harshest, most unproductive environments on earth. It also permitted them to adapt their behavior and social relations to the demands of powerful outsiders. ... As new forms of treatment and medical technologies have become available, people have readily accepted them. ... In general, Holman residents express satisfaction with the quality of the current health care system. No doubt this is due, in large part, to the accessible and friendly environment established by Holman's nursing staff. ... Holman is quite unusual among communities in the Kitikmeot Region of the NWT. There has been extremely low turnover in nursing staff compared to the other communities in the region, and the current Nurse-in-Charge, Audrey Woodget, has served in the community for nearly a decade.” Page 29. | | | Barriers related to the transmission of Indigenous healthcare practices: “It is our belief that two particular characteristics of Inuit culture have led to the near total abandonment of traditional medical practices. These two factors are: 1) an openness to adopt novel technologies, and 2) an unwillingness to confront others or cause conflict. Nearly all scholars of lnuit culture have remarked upon the great flexibility inherent in the society (13,14). This flexibility allowed Inuit culture to survive for centuries in one of the harshest, most unproductive environ­ments on earth. It also permitted them to adapt their behavior and social relations to the demands of powerful outsiders.” Page 29.  “One additional factor must be weighed in any consideration of the generational transmission of health information, specifically the degree to which the various generations are willing and able to communicate with one another. In the past, according to one elder (Stem 1988, fieldnotes), ‘children would follow the way that they were taught by their parents and would carry on that way as adults since there were no schools.’ The chain of information between generations was weakened when Inuit families moved into settlements and sent their children to boarding schools that discouraged the use of any language other than English. Many of these children returned home with deficient Inuinaqtun abilities, a situation which hindered effective communication with parents and grandparents. This phenomenon has become more dramatic in recent years. We have documented several cases in Holman in which children are unable to understand not only the language of their grandparents, but also the language of their own parents. It is no wonder that traditional knowledge in all domains is no longer transmitted to the youngest generation of Inuit.” Page 30. | | |
| **Study limitations** | | | | | |
| N/A; none described. | | | | | |

| **Record ID** | **Title** | | | | |
| --- | --- | --- | --- | --- | --- |
| #1224 – 2014 | Premenopausal vasomotor symptoms in an ethnically diverse population | | | | |
| **Author information** | | | | | **Country** |
| Reed, S. D.; Lampe, J. W.; Qu, C.; Copeland, W. K.; Gundersen, G.; Fuller, S.; & Newton, K. M. | | | | | United States |
| **Study aims** | | | | | |
| “Objective: The prevalence of vasomotor symptoms (VMS) among women aged 45 years or older who report regular menses has not been described well. Variability by race/ethnicity is expected.” Abstract  “We explored whether VMS prevalence was associated with soy intake.” Page 154. | | | | | |
| **Study design** | **Conflicts of interest** | | **Inclusion criteria** | | **Exclusion criteria** |
| Cross-sectional study. | N/A; none listed. | | “Women were included if they were premenopausal, were 45 to 56 years old, did not use hormones, did not have intestinal or malabsorption syndromes, did not undergo colectomy, did not have cancer, and did not use antibiotics in the prior 3 months.” Page 154.  “Women were included in this analysis—and excluded from the parent study—if they reported regular menses (not having skipped a period in the past 12 mo).” Page 154. | | “They were excluded if they had a hysterectomy. Women who were unable to read English or were mentally or legally incapacitated such that an informed consent form could not be obtained were also excluded.” Page 154.  Women who were “taking hormones, antibiotics, or long-term oral steroids; were pregnant or breast-feeding; had active cancer; had a long-term medical illness, including malabsorption syndrome; or were older than 56 years.” Page 154. |
| **Population and sampling description** | | **Indigenous description** | | **Total number of participants** | |
| The population of interest was US women who were premenopausal, were 45 to 56 years old, did not use hormones, did not have intestinal or malabsorption syndromes, did not undergo colectomy, did not have cancer, and did not use antibiotics in the prior 3 months.  “This study was performed at Group Health (GH), a large, integrated, nonprofit healthcare system that coordinates care and coverage for approximately 675,000 enrollees located in the Pacific Northwest. Potentially eligible women were identified via automated databases, and eligibility was confirmed by a mailed questionnaire.” Page 154.  “A total of 18,500 women aged 45 to 58 years, identified via electronic databases, were potentially eligible for the parent study LEAVeS. Recruitment occurred through a series of direct mailings, a magazine advertisement, and rotating announcements on informational screens in GH pharmacies from March 2010 through February 2011. Study information, including the survey and an informed consent form, was mailed to potentially eligible women. An incentive of US$2 was included in the first mailing. Mailings were repeated twice, and a reminder postcard was sent between mailings.” Page 154. | | Native American.  The authors do not describe how American Indian racial/ethnic background was defined.  The percentage of the sample who were Indigenous was 1.59%. | | The total number of participants was 1513, and of those 24 were Indigenous.  “Among 18,500 women identified by automated data, 9,007 responded to the mailed survey (48.7% response rate) and were potentially eligible for LEAVeS. Of those who responded, 1,778 had not skipped menses in the prior 12 months and reported regular menses. These women were classified as pre-menopausal. Among them, 265 were excluded because they were taking hormones, antibiotics, or long-term oral steroids; were pregnant or breast-feeding; had active cancer; had a long-term medical illness, including malabsorption syndrome; or were older than 56 years, leaving 1,513 women included in these analyses.” Page 154. | |
| **Study outcomes and important results** | | | | | |
| “Premenopausal Native American and black women most commonly reported VMS ever, but this difference was not statistically significant when compared with white women.” Page 155.  “As compared with white women, Native American women, followed by black women, most commonly reported night sweats ever (66.7% and 60.0% vs 50.8%, not statistically significant)” Page 155.  “Joint pain bother in the past 3 months was most common in Native American women and differed significantly from white women (79.2% vs 45.9%, P < 0.05).” Page 155.  “Among women reporting VMS in the prior 2 weeks, the proportion reporting at least moderate VMS bother by race was as follows: Native American, 80%; Hawaiian/Pacific Islander, 80%; white, 58%; black, 48%; Asian, 38% to 50%. There were no statistically significant differences between groups (data not shown).” Page 155.  Native American results from Table 2. “Prevalence of hot flashes, night sweats, combined VMS ever, headache, or joint pain in the prior 3 months among premenopausal women, by race/ethnicity” (N=1,531) are: n=24; “Hot flashes ever (%)” = 37.5%; “Night sweats ever (%)” = 66.7%; “VMS ever (%)” = 66.7%; “Headaches (%)” = 70.8; “Joint pain (%)” =79.2%. Page 155.  “The association between obesity and increased self-report of VMS prevalence has been described, [22,23] but the relationship does not seem to be linear. Of note, two of three racial/ethnic groups of women with the highest mean BMI in our study (>30 kg/m2)—black and Native American women— also had the greatest proportion of women with VMS ever (61% and 67%, respectively), although this was not statistically different from white women (58%).” Page 156. | | | | | |
| **Use of STRAW+10** | **Menopausal transition staging of sample using STRAW+10** | | | | |
| The STRAW+10 not used. | Menopausal Transition. | | | | |
| **Author reported facilitators to treatment** | | | **Author reported barriers to treatment** | | |
| N/A. | | | N/A. | | |
| **Study limitations** | | | | | |
| “The study had limitations, however. We did not include women who had a hysterectomy, had endometrial ablation, or were currently using hormone therapy, oral contraceptives, or the levonorgestrel intrauterine system. These exclusions most probably resulted in underestimation of the true prevalence of VMS in our population. In addition, we did not query further on when the VMS occurred among women who reported VMS ever. It is well known that women can experience hot flashes in the postpartum period. We did not ask about parity or any temporal sequence of VMS with reproductive events and, therefore, could not control for this in our analyses. The findings from our survey may not be generalizable. It is possible that women who did not respond to the survey (51%) differed from the women who responded (49%). The introductory letter informed women that the survey evaluated diet and menopause, with the goal of finding out if eating certain foods affected common symptoms of menopause. It is possible that women who filled out the survey were more likely to have menopausal symptoms, as compared with those who did not complete the survey. This could have inflated findings above the true VMS prevalence in the general population.” Page 157. | | | | | |

| **Record ID** | **Title** | | | | |
| --- | --- | --- | --- | --- | --- |
| #1223 – 2012 | Reproductive cancer risk factors among Alaska Native women: The Alaska Education and Research Towards Health (EARTH) Study | | | | |
| **Author information** | | | | | **Country** |
| Redwood, D. G.; Lanier, A. P.; Johnston, J. M.; Murphy, N.; & Murtaugh, M. A. | | | | | United States |
| **Study aims** | | | | | |
| “The purpose of this study was to provide estimates for the prevalence of reproductive cancer risk factors among Alaska Native (AN) women who enrolled in the Alaska Education and Research Towards Health (EARTH) Study from 2004 to 2006.” Abstract. | | | | | |
| **Study design** | **Conflicts of interest** | | **Inclusion criteria** | | **Exclusion criteria** |
| Cross-sectional study. | N/A; none listed. | | “Inclusion criteria: AN or AI eligible for care in the Alaska Tribal Health System, age 18 years of age or older, and able to provide written, informed consent.” Page 3. | | “We asked pregnant women and chemotherapy patients to participate at a later date owing to health measurement changes caused by these two factors.” Page 3. |
| **Population and sampling description** | | **Indigenous description** | | **Total number of participants** | |
| The population of interest included AN or AI adults eligible for care in the Alaska Tribal Health System.  “We recruited participants from 26 Alaskan communities in three regions of the state (Table 1). The south central Alaska region included AN people from many different ethnic groups living in or around Alaska’s largest city with a total population of 350,000. The southeast Alaska region included mostly Tlingit, Haida, and Tsimshian Indian people living in smaller communities (ranging in population from 500 to 30,000). The southwest Alaska region included mostly Yupik people and was the most rural and remote of the three Alaskan regions, with communities ranging in population size from 200 to 5,500. All of the southeast and southwest Alaskan communities were located off the road system and were accessible only by airplane, with seasonal access by snow machine or boat. Methods of recruitment for all regions included presentations to tribal groups and healthcare providers, information tables staffed by study personnel at community events, house-to-house recruiting, brochures and flyers in public locations, and public service announcements on local radio and in newspapers. In each community, we attempted to enroll all residents of the community who met inclusion criteria.” Pages 2 and 3. | | Alaska Native.  Participants were recruited from 26 Alaskan communities in three regions of the state: south central Alaska, southeast Alaska (Tlingit, Haida, and Tsimshian Indian peoples), and southwest Alaska (which included mostly Yupik people and "was the most rural and remote of the three Alaskan regions").  The authors did not describe how Alaska Native was defined but they did include eligibility for care in the Alaska Tribal Health System as an inclusion criterion. Page 3.  The percentage of the sample who were Indigenous was 100%.  Additional sample information:  “All of the southeast and southwest Alaskan communities were located off the road system and were accessible only by airplane, with seasonal access by snow machine or boat.” Page 2. | | The total number of participants was 2315, and the total number of Indigenous participants was 2315. | |
| **Study outcomes and important results** | | | | | |
| “A total of 618 (28%) of the women reported that they had completed menopause, that is, their periods had stopped completely. Of those, a total of 140 (24%) completed menopause after age 52. Among women who did not have a hysterectomy, the mean age at menopause was 48 years. Of the women whose periods had stopped completely, 207 (34%) were because they had had a hysterectomy. A total of 231 women (10%) had had an oophorectomy. Of those who had a hysterectomy, 68% also had their ovaries removed” Page 4.  “A total of 369 women (43%) reported using hormone replacement therapy either currently or at some point in the past, with over half (62%) using hormone replacement therapy for 4 years or less (Table 3).” Page 4.  “EARTH data suggest that AN women report a younger age of menarche, age at first birth, and age of menopause than women in many other ethnic and racial groups (Dratva et al., 2009; Dratva et al., 2007; Henderson et al., 2008; Lawlor, Ebrahim, & Smith, 2003; Santoro et al., 2007).” Page 4.  “The Alaska EARTH Study found that 28% of study participants had completed menopause, and 24% reported menopause after age 52. There was a mean menopause age of 45.6 years for the whole sample, which is younger than other published reports for AI women. The Strong Heart Study found a mean age of natural menopause of 46.8 years (Cowan et al., 1997), whereas Montana Blackfeet women reported 51.2 years (Johnston, 2001).” Page 6. | | | | | |
| **Use of STRAW+10** | **Menopausal transition staging of sample using STRAW+10** | | | | |
| The STRAW+10 not used. | Reproductive, Menopausal Transition, and Postmenopause. | | | | |
| **Author reported facilitators to treatment** | | | **Author reported barriers to treatment** | | |
| N/A. | | | N/A. | | |
| **Study limitations** | | | | | |
| “A limitation of this study is the nonrandom sampling design and the inability to assess the relative contributions of these risk factors to incident reproductive cancers among AN women, specifically. Although the cohort was not randomly selected, the distribution of the study population closely resembled the demographic distributions reported by the 2000 U.S. Census for AI/AN in the respective regions (U.S. Census Bureau, 2000). All data were collected by self-report, potentially contributing to measurement error. Last, data reported here are cross-sectional from the initial study visit, and do not allow evaluation of causal relationships between variables. Further study is needed to understand whether the strength of the risk and protective factors for reproductive cancers among AN women is similar to that among other race and ethnic groups.” Page 6. | | | | | |

| **Record ID** | **Title** | | | | |
| --- | --- | --- | --- | --- | --- |
| #996 – 2000 | Women’s Stories: Ethnic Variations in Women’s Attitudes and Experiences of Menopause, Hysterectomy, and Hormone Replacement Therapy | | | | |
| **Author information** | | | | | **Country** |
| Mingo, C.; Herman, C. J.; & Jasperse, M. | | | | | United States |
| **Study aims** | | | | | |
| “To increase understanding of women’s midlife changes, 23 focus groups were held to investigate the possible ethnic variations in attitude toward and experience with menopause, hysterectomy, and hormone replacement therapy (HRT) in non-Hispanic white, Hispanic, and Navajo women in New Mexico.” Abstract.  “THE PURPOSE OF THIS QUALITATIVE PROJECT, which is part of the larger Ethnicity, Needs, and Decisions of Women (ENDOW) project, a 5-year, multisite, community-based study researching the similarities and differences in women’s attitudes, experiences, and decisions regarding menopause, hormone replacement therapy (HRT), and hysterectomy, is to gain an understanding of the ethnic variations in women’s attitudes and experiences of menopause, hysterectomy, and HRT.” Page S-27. | | | | | |
| **Study design** | **Conflicts of interest** | | **Inclusion criteria** | | **Exclusion criteria** |
| Qualitative research. | N/A; none listed. | | “Eligibility for participation in the menopause focus groups was lenient. Women were allowed to self-identify as menopausal—this included premenopause, perimenopause, and postmenopause. The self-identified, naturally menopausal participants ranged in age from 34 to 71 years (Table 1).” Page S-28. | | N/A; none listed. |
| **Population and sampling description** | | **Indigenous description** | | **Total number of participants** | |
| The population of interest was “Non-Hispanic white, Hispanic (monolingual or bilingual women whose families had lived in New Mexico for many generations), Latina (newly immigrated from Mexico, Central America, and South America, mostly Spanish-only speakers), more traditional Navajo (spoke Navajo only or were most comfortable speaking Navajo), and less traditional Navajo (English speakers but often bilingual)” Page S-28.  “Various methods were employed to recruit participants for the focus groups. After the brief article, ‘Study views changing attitudes on menopause,’ appeared in the major Albuquerque morning newspaper (the Albuquerque Journal), almost 150 women called to volunteer. Newspaper recruitment worked well in the city of 600,000, but it did not provide an effective recruitment method for the rest of the state, as the rest of New Mexico’s 1.6 million total population is scattered thinly over more than 160,000 square miles. The primary research sites outside the metropolitan area were located 212 miles, 153 miles, and 103 miles, respectively, away from the University of New Mexico. In these nonurban areas, bilingual recruiter/facilitators put up posters in community centers, healthcare centers, and grocery stores, but word of mouth was the most effective recruitment technique.” Page S-28. | | “More traditional Navajo (spoke Navajo only or were most comfortable speaking Navajo), and less traditional Navajo (English speakers but often bilingual).” Page S-28.  The authors do not describe how Navajo was defined.  The percentage of the sample who were Indigenous was 36.5% (n=57 out of 156 total participants).  Additional sample information: Table 1., Page S-29, “Age and Menopausal History and Status of Focus Group Participants for the Navajo participants.” Age (years): the 35 to 44 age category included 6 participants (11%); the 45 to 49 age category included 6 participants (11%); the 50 to 54 age category included 10 participants (17%); The age category 55 to 64 included 18 participants (32%); and, last, the 65+ age category included 17 participants (29%).  Table 2., Page S-34, “Attributes of Midlife Navajo Focus Group Participants” categorizes “Attributes” for the “Traditional,” “Transitional,” and “Modern” participants. For Traditional participants: Diet included “Corn, dried fruit and vegetables, wild game”; Physical exertion included “Tend sheep, carry water, cut and carry wood”; Healthcare included “Medicine man / women traditional herbs, healing chants”; Family support included “Listen to elders”; Housing included “Rural compound with hogan, no electricity, central heat, or running water”; Schooling included “None to <3 grade”; Language included “Navajo only”; Births included “Many, some traditional birth control”; Age (years) included “>55”; and, last, Symptoms described included “Almost none, waist pain, headache”. For Transitional participants: Diet included “Mutton, canned goods, dried fruit and vegetables”; Physical exertion included “Physically demanding job”; Healthcare included “Some traditional, some Western, some herbs”; Family support included “Less contact with elders”; Housing included “Cluster of homes, maybe with hogan, near paved road; possible heat, water, or electricity”; Schooling included “Often boarding school, at least some high school”; Language included “Navajo and English”; Births included “0-6, some birth control”; Age (years) included “45-65”; and, last, Symptoms described included “Aching back, head, fatigue”. For Modern participants: Diet included “More fast food, more sugar, fat salt”; Physical exertion included “Sedentary job, may run or do aerobics”; Healthcare included “Western medicine”; Family support included “Isolation from family”; Housing included “Single house in urban area; central heat and electricity, running water”; Schooling included “Public school, high school, some college”; Language included “Fluent English”; Births included “0-4, birth control was a choice”; Age (years) included “45-50”; and, last, Symptoms described included “More typical: hot flashes, sweats” | | The total sample size was 156, of which 57 were Navajo. | |
| **Study outcomes and important results** | | | | | |
| “Although the rhythm of hormonal fluctuations is familiar to women—mood swings, fluid retention, tender breasts—the women would like more and better information. They wished that their mothers, sisters, and friends (their primary sources of information) had prepared them for menopause. Mostly, women were surprised that their periods stopped, that they had menopausal symptoms, that it was so hard to find or get useful information, and that they and their friends and family were so reluctant to talk about menopause.” Coinciding quote from a Navajo participant: “I talked to my mom. I asked her, did you ever get like that? She said, ‘I don’t know what you are talking about.’ She never had that. She didn’t have it. —Navajo” Page S-30.  “Although the hysterectomy rate has been falling gradually since its peak at 725,000 per year in 1975, (6) we were taken by surprise at the number of women in the focus groups who had to fight to have their surgery. With debilitating symptoms, some women pleaded for surgery for up to 8 1/2 years; others were told to see a therapist. They often endured tremendous pain and discomfort for months or years before finally having a hysterectomy. There were many, many reasons for this—not the least of which was the stoicism of the women.” Coinciding quote from a Navajo participant: “I said, Lord, I don’t know how these others feel, but I’m hurting. My feelings I keep to myself. I can’t share with nobody. This is the first time I’ve talked. —Navajo” Page S-31.  “If women had mild to moderate difficulties and the healthcare provider suggested hysterectomy, they would wait and see if it would cure itself or if alternatives would work: herbs, acupuncture, diet and exercise.” Coinciding quote from a Navajo participant: “I felt weak and slept a lot again. It went on for a long time. With those problems I don’t think they can all be cured at the hospital. There are our own singers. I went to one and he held a ceremony for me. They know about herbal medicine. I was given some the herbal medicine. I became very well by drinking and using the herbal medicine. —Navajo” Page S-31.  “Many women were surprised that hysterectomy brings on menopause, either immediately or later and sometimes both.” Coinciding quote from a Navajo participant: “I talked to some ladies that had it done, and they tell you different stories. They say your sex life isn’t going to be the same, your husband isn’t going to want you anymore. I hear a lot of stories. Then I had my hysterectomy. I came home and I still felt the same. I used to bleed a lot. I kept waiting for something to happen, what the ladies were telling me. The years went on . . . then one day I was feeling hot, I was cranky, I was sad, and then I was happy. I didn’t know what was wrong with me. Things that the ladies had told me at the time I had my hysterectomy, I didn’t have that. But after all these years I started feeling weird, hot flashes, cranky, I didn’t want anything to do with mister. Later, I was reading Good Housekeeping magazine and I came to this page about menopause, and everything I was going through I read about in that book. That was menopause, and to this day, I ask myself how long is this going to last—the rest of my life? Last year I talked to the doctor and asked how long is it supposed to be, the hot flashes, crankiness, wanting to cry, and not wanting to eat. I is just awful. … I talked to the doctor about it and he didn’t answer me. Maybe he did but I don’t remember. —Navajo” Page S-32.  “What’s HRT? —Entire Navajo focus group” Page S-32.  “As women’s experiences differ, so do their responses to their bodies and the alternatives they seek to alleviate their pain and discomfort—on a continuum between meager and drastic. During the focus group analysis, we came to realize how prevalent tinkering with HRT has become.” Coinciding quote from a Navajo participant: “She put me on estrogen, but it gave me real bad headaches and I tried to use it like every other day. [laughter] I am not taking it right now. I take a lot of calcium and I take vitamin E. —Navajo” Page S-33.  “For many women, HRT worked successfully, quickly, and without side effects.” Coinciding quote from a Navajo participant: “About a year, year and a half ago, I was talking to my doctor—good thing she was a woman, I felt good talking to her. I said, ‘Something’s wrong with me. I’m so weak, I can’t sleep, I get hot, I can’t eat—everything I eat is nauseating me.’ She gave me that pill, the estrogen. I took it that evening and about half a day is over and I’m feeling myself again. It worked that fast. Overnight —Navajo” Page S-34.  “The research with Navajo women revealed only one traditional woman who had any self-identified menopausal indicators. Transitional and modern Navajo women seemed to approach the range and intensity of menopausal indicators similarly to Hispanic and non-Hispanic white women. The most easily identified factors were diet, number of births, and family support. Certainly, there were few if any phytoestrogens in the traditional diet. Many of the traditional Navajo women, but not all, supported extra weight, which manufactures estrogen. (3,p.59) Many women gave birth until the end of menses, but some had never been pregnant. The importance of extended family was prevalent, but several women lamented their aloneness. The women talked about the puberty ceremony that traditional women, many transitional women, and a few modern women had had performed. The puberty ceremony, (7) kinaaldá, took place at the time of a girl’s first menses. It was a powerful combination of spirituality, ceremony, community involvement, preparation for womanhood through mentoring, singing, feasting, running, and grinding corn. The kinaaldá is still considered a preparation for life. Traditionally, it was not only preparation for reproductive life but also preparation for the work and responsibility of caring for a home and family. The ceremony was unique among the Navajo women; Hispanic and non-Hispanic white women had experienced nothing comparable. ‘I think that’s right [that those who had kinaaldá do not experience hardship]. I never had it done ‘cuz when I got my period, one of my uncles passed away. And when someone passes away in the family, you don’t have no ceremony. The second time came, and another member [died], so I never had it done. —Navajo’ The kinaaldá is typically a 4-day ceremony and is so intricately meshed with traditional beliefs, practices, and lifestyle that isolating any individual factors is difficult. One woman, however, approached us after the Chapter House focus group because she had wanted to participate but was involved in another meeting. She discussed the life she and her sisters had lived and her kinaaldá. She summed up the puberty ceremony experience as ‘the most physically grueling experience I’ve ever had in my life.’ The contrast between the ‘physically grueling experience’ that traditional Navajo women experienced at the time of first menses while running, grinding corn, and gathering firewood for 4 days and the more common experience of Hispanic and non-Hispanic white women who often experienced the ‘poor baby, take a hot water bottle and lie down until you feel better’ approach was stark. Perhaps a life of vigorous physical work beginning with the ceremony for first menses and continuing throughout life is the best preparation for menopause. Traditional women constantly engage in physical work by carrying water, gathering firewood, baking bread or tortillas, tending fields and sheep, and walking from one place to another.” Pages S-34 through S-36. | | | | | |
| **Use of STRAW+10** | **Menopausal transition staging of sample using STRAW+10** | | | | |
| The STRAW+10 not used. | Reproductive, Menopausal Transition, and Postmenopause.  The age at last menstrual period (years): the “Still menstruating” group included 6 participants (11%*); the “<36” group included 6 participants (11%); the “36-45” group included 8 participants (14%); the “46-55” group included 20 participants (35%); and, last, the “>55” group included 2 participants (3%).  Menopausal status: the “Pre- / perimenopausal” group included 5 participants (9%); the “Natural menopause” group included 26 participants (46%); the “Surgical menopause (hysterectomy ± oophorectomy)” group included 23 participants (40%); the “Pending hysterectomy” group included 0 participants (0%); and, last, the “Status unknown” group included 3 participants (5%).  Table 2., Page S-34, “Attributes of Midlife Navajo Focus Group Participants” categorizes “Attributes” for the “Traditional,” “Transitional,” and “Modern” participants. The attributes include “Onset of menses,” “Diet,” “Physical exertion,” “Healthcare,” “Family support,” “Housing,” “Schooling,” “Language,” “Birth,” “Age,” and “Symptoms described”. For Traditional participants: Onset of menses included “Kinaaldá, at least one public celebration of menses”. For Transitional participants: Onset of menses included “Maybe kinaaldá”. For Modern participants: Onset of menses included “No kinaaldá”. | | | | |
| **Rural vs. urban differences in outcomes** | | | | | |
| The participants classified as “traditional” in this study were found to live rurally. The participants who were classified as “modern” were found to live in urban areas. Therefore, one difference between the traditional (rural) participants and those who were classified as “modern” (urban) was “Interestingly, the usual symptoms of menopause, natural or surgical, were more prevalent in all ethnic groups who were classified as modern and least prevalent or nonexistent in the women classified as traditional”. Page S-34. | | | | | |
| **Author reported facilitators to treatment** | | | **Author reported barriers to treatment** | | |
| “Women would like healthcare providers to respectfully listen and thoughtfully explain options and procedures in a nonharried, sensitive atmosphere. Books and magazines written for women, as well as bilingual oral and video presentations addressed to women who speak English as a second language, will help women make intelligent decisions for their own bodies while supporting each woman’s uniqueness. Women need to ask doctors to write down instructions, questions, and symptoms as well as medications. Most women find it difficult to understand and remember new terms, new diagnoses, or new symptoms to watch for it they are only told them.” Page S-37. | | | N/A. | | |
| **Study limitations** | | | | | |
| N/A; none described. | | | | | |

| **Record ID** | **Title** | | | | |
| --- | --- | --- | --- | --- | --- |
| #964 – 2001 | Black cohosh: Efficacy, safety, and use in clinical and preclinical applications | | | | |
| **Author information** | | | | | **Country** |
| McKenna, D. J.; Jones, K.; Humphrey, S.; & Hughes, K. | | | | | Canada |
| **Extraction note** | | | | | |
| This record was included because the abstract states that black cohosh was traditionally used among Native Americans for a variety of disorders, including menopause. It should be noted, however, that within the review’s text, the authors do not include menopause as one of the disorders for which Native Americans use/used black cohosh. We therefore do not have a citation to include and could be reinforcing a false narrative about traditional Native American use of black cohosh. | | | | | |
| **Study aims** | | | | | |
| N/A; none described. | | | | | |
| **Study design** | **Conflicts of interest** | | **Inclusion criteria** | | **Exclusion criteria** |
| Other: narrative review. | N/A; none listed. | | N/A; no methodology details are listed. | | N/A. |
| **Population and sampling description** | | **Indigenous description** | | **Total number of participants** | |
| N/A; this is a narrative review of the literature on black cohosh. | | N/A; narrative review of the literature and not a primary study. The review does reference the Algonquian language. | | N/A; this is a literature review, additionally, the authors do not describe the results of their literature review nor how many records met the final inclusion criteria. | |
| **Study outcomes and important results** | | | | | |
| “Black cohosh has a history of traditional use among Native Americans for the treatment of a variety of disorders, including various conditions unique to women such as amenorrhea and menopause.” Abstract.  “‘Cohosh’ comes from an Algonquian word meaning ‘rough’ and refers to the plant’s lumpy, blackish rhizomes.” Page 3 of 17. | | | | | |
| **Use of STRAW+10** | **Menopausal transition staging of sample using STRAW+10** | | | | |
| The STRAW+10 not used. | N/A. | | | | |
| **Author reported facilitators to treatment** | | | **Author reported barriers to treatment** | | |
| N/A. | | | N/A. | | |
| **Study limitations** | | | | | |
| N/A; none described. | | | | | |

| **Record ID** | **Title** | | | | |
| --- | --- | --- | --- | --- | --- |
| #951 – 1976 | Fertility and other demographic aspects of the Canadian Eskimo communities of Igloolik and Hall Beach | | | | |
| **Author information** | | | | | **Country** |
| McAlpine, P. J.; & Simpson, N. E. | | | | | Canada |
| **Study aims** | | | | | |
| To study reproductive histories “in two closely related Eskimo communities in the eastern Canadian Arctic: (Fig. 1) Igloolik (69°N, 82°W) and Hall beach (69°N, 81°W). This study documents fertility and population structure of these communities at a time when industrialized ‘Western civilization’ has begun to have a profound and rapid effect on the lifestyles of the Eskimos. It is hoped that the data may be used for comparisons with other Eskimo populations and with the same populations in the future at a time when ‘acculturalization’ is complete.” Page 113. | | | | | |
| **Study design** | **Conflicts of interest** | | **Inclusion criteria** | | **Exclusion criteria** |
| Cross-sectional study. | N/A; none listed. | | Mothers from the Igloolik and Hall Beach communities. | | N/A; none listed. |
| **Population and sampling description** | | **Indigenous description** | | **Total number of participants** | |
| The population of interest was all mothers from the Eskimo Igloolik and Hall Beach communities. From the abstract and page 115. | | Eskimo Igloolik. | | The total number of participants is 132 and the total number of Indigenous participants is 132. | |
| **Study outcomes and important results** | | | | | |
| “The mean age of menopause at 52 years” Abstract.  “Of the 89 mothers in Igloolik, ... 6 (6.7%) as artificially postreproductive and 11 (12.4%) as naturally postreproductive. In Hall Beach ... 4 (9.3%) , 4 (9.3%) were artificially postreproductive and 4 (9.3%) were naturally postreproductive.” Page 117.  “Age at menopause and duration of reproductive period. The mean age at menopause was 52.3 ± 1.0 yr for the 11 naturally postreproductive women in Igloolik for whom data were available and 49.3 ± 1.7 yr for the four naturally postreproductive women in Hall Beach. As the difference between these means was not statistically significant, the two groups were combined. The mean age at menopause was 51.5 ± 0.9 yr for all naturally postreproductive mothers. The period from menarche to natural menopause ranged from 28 to 43 years with a mean of 36.3 ± 1.9 yr for the eight Igloolik mothers for whom these data were available. For the four Hall Beach mothers for whom data were available, this period ranged from 33 to 41 years with a mean of 35.8 ± 1.8 years. The mean of the pooled data was 36.0 ± 1.” Pages 120 to 121.  “The mean age at menopause of all the naturally postreproductive women in the sample (51. 5 ± 0.9 yr) was significantly greater than the mean of 44.5 ± 1.1 yr for Wainwright women reported by Milan (1970) (t =4.954, p<0.001).” From the Discussion section, page 136. | | | | | |
| **Use of STRAW+10** | **Menopausal transition staging of sample using STRAW+10** | | | | |
| The STRAW+10 not used. | Postmenopause.  “The postreproductive groups were subdivided into a) artificially postreproductive, i.e. those who had had their postreproductive state induced by surgery (e.g. tubal ligation, hysterectomy) or by known disorders of the reproductive tract (e.g. Sheehan's syndrome) which prevented them from having further children and b) naturally postreproductive, i.e. those in whom the menopause had occurred naturally.” Page 116. | | | | |
| **Author reported facilitators to treatment** | | | **Author reported barriers to treatment** | | |
| N/A. | | | N/A. | | |
| **Study limitations** | | | | | |
| N/A; none described. | | | | | |

| **Record ID** | **Title** | | | | |
| --- | --- | --- | --- | --- | --- |
| #909 – 2010 | First Nations women’s knowledge of menopause: Experiences and perspectives | | | | |
| **Author information** | | | | | **Country** |
| Madden, S.; St Pierre-Hansen, N.; Kelly, L.; Cromarty, H.; Linkewich, B.; & Payne, L. | | | | | Canada |
| **Study aims** | | | | | |
| “To understand and describe the menopause experiences and perspectives of First Nations women residing in northwestern Ontario.” Abstract. | | | | | |
| **Study design** | **Conflicts of interest** | | **Inclusion criteria** | | **Exclusion criteria** |
| Qualitative research. | “None declared.” Page e337. | | “Participants who were either perimenopausal or post-menopausal by way of natural or surgical menopause were asked to participate.” Page e334. | | N/A; none listed. |
| **Population and sampling description** | | **Indigenous description** | | **Total number of participants** | |
| The population of interest was First Nations women.  “PARTICIPANTS. Eighteen perimenopausal and postmenopausal First Nations women, recruited by convenience and snowball sampling techniques.” Abstract.  “Convenience sampling techniques were used to recruit 18 First Nations women, 9 from Sioux Lookout and 9 from the 4 surrounding First Nations communities.” Page e334.  “Potential participants were identified by the research team, community health workers, community leaders, and by word of mouth.” Page e334. | | First Nations women: Sioux Lookout (n=9) and four surrounding First Nations communities (n=9). Page e334.  The authors did not describe how Indigenous background was defined in the study (e.g., self-identity, records).  The sample is 100% Indigenous. | | The total number of participants was 18; and all were indigenous. | |
| **Study outcomes and important results** | | | | | |
| “The first thing we learned from our initial consultations with our First Nations researchers and elders was that there was no consistent word for menopause in the regional languages of Ojibway and Oji-Cree. It was commonly referred to as ‘that time when periods stopped.’ We adopted this terminology and only used the word menopause when the participant introduced it. The second finding was that many participants began their narrative about menopause by discussing menarche experiences. These stories noted onset of menses with or without traditional ceremonies as well as residential school menarche experiences.” Page e334.  “Thematic analysis delivered 3 overarching themes regarding menopause among First Nations women: knowledge, communication, and experience (Figure 1).” Page e334.  Knowledge (pages e334 to e335).  “Nine of the 18 women gained some sort of information from their mothers.” Page e334.  “Other sources of information regarding menopause included books, other women (eg, older women, coworkers, and sisters), and health care providers. Some women received either no information or misinformation.” Page e335.  “Many women described the need for more information and enhanced awareness about what to expect, a greater understanding about the process, and information on relevant medications” Page e335.  Communication (pages e335 to e336).  “Most participants described menopause as something that was not discussed” Page e335.  “Some women experienced open communication with their sisters and other women” Page e336.  “Many women implicitly identified a generational change in which menopause is increasingly discussed” Page e336.  “Several informants described health care provider communication as an element of their menopause experiences. Others identified a need for more information from doctors. A couple of the women expressed a need for doctors to be more holistic. One woman identified the need for doctors to recognize language barriers” Page e336.  Experience (page e336).  “Many participants viewed menopause in a neutral or positive way. Some did not identify any value attached to the experience and others saw it simply as the cessation of menstruation” Page e336.  “It was described at times as a phase or stage in life. A couple of participants expanded on this concept as a time when women are more respected and seen as sacred” Page e336.  “Six women experienced no symptoms and 6 expressed symptoms as central to their menopause experience. Mood swings and hot flashes predominated” Page e336.  “Participants in this study told stories that referenced stages of a woman’s life. That several of the participants discussed menarche and pregnancies when asked about menopause is interesting. We had never experienced this in similar discussions with nonaboriginal patients in our clinical practices, where menopause can be discussed in isolation. This might speak to a holistic concept of ‘woman,’ in which discussion of menopause incorporates the beginning of the woman’s story. This might have implications for how clinicians begin such discussions.” Page e337.  “Caregivers would be well served to recognize the multiple influences affecting First Nations women’s experiences with menopause and to exercise caution when attaching preconceived ideas of the meaning and importance of menopause to a woman’s personal experience. Some First Nations women might be coming from backgrounds in which menopause is not discussed, yet experience a desire for information and a greater understanding of menopause; or, as one participant put it, ‘answers to questions that might be hidden somewhere, [which we] are afraid to ask.’” Page e337. | | | | | |
| **Use of STRAW+10** | **Menopausal transition staging of sample using STRAW+10** | | | | |
| The STRAW+10 not used. | Menopausal transition or postmenopause.  “perimenopausal or post-menopausal by way of natural or surgical menopause” Page e334. | | | | |
| **Author reported facilitators to treatment** | | | **Author reported barriers to treatment** | | |
| N/A. | | | N/A. | | |
| **Study limitations** | | | | | |
| “The convenience sampling technique limits the transferability our findings to other groups, especially outside of the Nishnawbe Aski Nation region of northwestern Ontario. The uniqueness of both the menopause experiences and each of the First Nations communities, respectively, means that these findings might not be generalizable. Results are also limited by the translation of words and concepts across cultures.” Page e337. | | | | | |

| **Record ID** | **Title** | | | | |
| --- | --- | --- | --- | --- | --- |
| #740 – 2003 | Menopause in Blackfeet Women – A life span perspective | | | | |
| **Author information** | | | | | **Country** |
| Johnston, S. L. | | | | | United States |
| **Extraction note** | | | | | |
| From the comparison of the cases reported in this record, it appears there are SDoH differences between the woman who had the earliest reported age at menopause and the woman with the latest. The author links this with “traditionalism” of Indigenous ways and that poverty and a traditionalism go hand-in-hand. | | | | | |
| **Study aims** | | | | | |
| “The present paper examines factors related to the timing of natural menopause in these women using a lifespan approach. Cross-sectional data are further illustrated by case studies.” Page 58. | | | | | |
| **Study design** | **Conflicts of interest** | | **Inclusion criteria** | | **Exclusion criteria** |
| Cross-sectional study. | N/A; none listed. | | Inclusion criteria not listed. The author referenced an earlier publication for methods details. Reference:  JOHNSTON, S. L., Am. J. Hum. Biol., 13 (2001) 512. | | N/A; none listed. |
| **Population and sampling description** | | **Indigenous description** | | **Total number of participants** | |
| The population of interest was Blackfeet Indian women living on the Blackfeet Reservation in northern Montana.  “An age-stratified, probability sample of 321 women was drawn from the tribal enrollment list of 2,425 eligible women 18 years and older with reservation addresses. Of these, 150 women aged 18–93 years agreed to participate in the study. Additional details of sample selection and methods are presented elsewhere. [30]” Pages 58 and 59.  Reference:  JOHNSTON, S. L., Am. J. Hum. Biol., 13 (2001) 512. | | “Blackfeet Indian women living on the Blackfeet Reservation in northern Montana.” Page 58.  Self-reported blood quantum is stated for the two qualitative cases presented in this report. (Case 1=7/8 Blackfeet blood quantum, page 60; Case 2=13/23 Blackfeet blood quantum, page 62). | | One-hundred-fifty participants (all Blackfeet women) contributed to the quantitative analysis. Thirty-three of these participants participated in in-person interviews. Page 59. | |
| **Study outcomes and important results** | | | | | |
| “Median age at natural menopause by the status quo method was 51.2 years (95% CI 50.0–53.2 yrs), n=86. The mean recalled age among the women in the sample who already had experienced natural menopause was 47.0 5.0, median 47.0 years (n=33).” Page 59.  “In this postmenopausal sample, bivariate analyses indicated age at natural menopause was associated inversely with age at menarche (p=0.07) (Table 2). In this regression there was one influential case (DFBeta=0.63) with later menarche and later menopause which, when removed, resulted in a statistically significant inverse relationship between ages at menarche and menopause at p=0.02. Age at menopause also was associated inversely with having been breastfed by one’s mother (p=0.02). Menopause age was associated positively with current household income (p=0.01), ever-use of any form of birth control (p=0.008), ever-use of oral contraceptives (p=0.007), and current or recent employment (p=0.02). Women in the latest tertile of menopause had significantly more education than women in the earliest tertile (t= –2.06, p=0.05, not shown). Age at natural menopause was not associated with current age, parity, age at first live birth, menstrual cycle length, subject’s breastfeeding history, smoking, anthropometric variables reflecting nutritional status (height, body mass index, skinfolds, arm muscle area and centripetal fat ratio), or other socio-demographic variables such as language spoken in childhood or current home, and marital status.” Page 59.  “In multivariate analysis using step-wise regression, a model including oral contraceptives, age at menarche, and household income explained the most variance (n=29, F=5.81, p=0.004, R2=0.41). However, significance levels of the household income and age at menarche variables in this regression suggested some collinearity, which was evaluated further with an analysis of joint effects (Table 3). Women with later menarche and lower household income had significantly earlier age at menopause (p=0.04). That women with later menarche and higher income also were younger at menopause (although not significantly) suggests that the main effect on age at menopause in this sample may be age at menarche, but its effect is modified in some way by household income.” Page 60.  Case 1 is the individual with the earliest age at menopause. Both women were members of the 50–69 cohort. “Ms. A was born in 1941 and was 55.0 years of age at the time of interview. She reported experiencing natural menopause at age 35 and menarche at 16.5 years. Her menstrual cycles were regular and, on average, 28 days in length. Ms. A had never married or been pregnant and had never used any form of contraception or hormone therapy. She was unsure whether she had been breastfed by her mother. Ms. A reported a 7/8 Blackfeet blood quantum and stated that during her childhood both Blackfoot and English were spoken at home. During at least 10 years of her childhood, her family lived about 80 miles from the reservation in a two- room house, without plumbing, that was heated with wood. She attended government boarding schools on the Blackfeet Reservation and in South Dakota for 11 years, nine months per year. At the time of the interview, she was living with a sister and five nieces and nephews in Browning, the main reservation town, and English was the only language in use at home. Ms. A had attained two years of post-secondary education. She was not working outside the home but had done so episodically in the past. She reported annual household income of between $1,250 and $2,499 from general assistance. The household rented the home they lived in and none of the members owned land on the reservation. Ms. A reported that the household possessed a microwave oven, a radio, a color television, a clothes washer and dryer, and one working vehicle. There was no telephone service. This household receives government commodity foods (Food Distribution Program on Indian Reservations of the U.S. Department of Agriculture) and obtains other food items at local and regional supermarkets. Ms. A described her childhood diet as more »balanced« and more plentiful in quantity than her diet at the time of interview. She was an active cigarette smoker and had been smoking for about 33 years, up to 1/2 pack per day. Ms. A’s height was 169.1 cm, BMI 21.9 kg/m2, triceps skinfold 16.5 mm, and subscapular 18 mm.” Pages 60 through 62.  Case 2 the latest age at menopause. “Mrs. B was born in 1932 and was 64.0 years of age at the time of interview. She reported natural menopause at age 58, menarche at age 11, and menstrual cycles of 25–28 days in length. She listed eight pregnancies, with the first live birth at age 17. Two of these pregnancies ended in stillbirths and one in a premature delivery of live twins who died within hours. Five deliveries were singleton live births. She breastfed all of her children for varying lengths of time and reported she was not breastfed by her own mother. She recalled having used several forms of contraception, including rhythm, oral contraceptives, and finally tubal ligation, although her husband, whom she described as older than she, »did not believe in birth control.« At the time of interview she was on hormone replacement therapy with estrogen and progestin, which she had begun after menopause. Mrs. B stated her Blackfeet blood quantum was 13/32 and the language spoken in her childhood home was English. She was raised by her grandparents and lived mostly on or near another Indian reservation about 200 miles east of Browning in north central Montana. The home she lived in for six years, the longest at that time, had two rooms, no plumbing or electrical service, was heated with wood, and was located near the Missouri River, which was their water source. She went to boarding school in Great Falls for five years, nine months per year. She completed 10 years of school and got married at age 16. At the time of interview she had been widowed for about six months and lived alone in Browning throughout most of the year; in the summer some of her family would join her. She reported working full time during the school year and stated her annual income, from wages, land lease, and summertime unemployment benefits, was in the $40,000 and $79,999 category. She owned her home and also individual trust land on the reservation. The home included a microwave oven, two radios, two color televisions, a clothes washer and dryer, two working vehicles, and telephone service. Mrs. B said most of the household food during her childhood came from a supermarket in a nearby town or was home raised (beef cattle, milk cows, vegetable garden); some food was wild (deer). She described her childhood diet as nutritious, and said she ate better then than at present (fewer »fast foods« in the past). At the time of interview, she was getting most of her food from a supermarket off the reservation and also utilized local markets, the Hutterites for seasonal fresh vegetables, and wild foods (deer, elk, berries). Mrs. B stated she was a cigarette smoker at the time of the interview, smoking one pack per day on the weekends and two cigarettes a day during the week; she had smoked intermittently since age 10 or 11. Mrs. B’s height was 161.4 cm, BMI 30.7 kg/m2, biceps skinfold 9 mm, and subscapular 29.5 mm.” Page 62.  “Among Blackfeet women who have experienced menopause, several factors are predictive of the timing of this event. Earlier menopause is associated with later menarche, lower adult income, and non-use of oral contraceptives in a multivariate model that explains almost half of the variance in age at menopause. Other associations include non-employment, lower education, and having been breastfed in infancy. Unlike many studies, this one found no association with smoking.” Pages 62 and 63.  “There was only nine years difference in age between these two women, and they were treated in the cross-sectional study as members of the same cohort; however, their lives as portrayed in the information they provided during the survey were very different. Ms. A’s history is suggestive of lifelong poverty, while Mrs. B’s appears to have been more economically secure. Ms. A never married or had children and had a relatively short reproductive span, while Mrs. B married, had children, and experienced a long reproductive period. They had in common that they both had long histories of smoking and were active smokers at the time of menopause, and that they both spent at least part of their childhood away from the Blackfeet Reservation.” Page 63. | | | | | |
| **Use of STRAW+10** | **Menopausal transition staging of sample using STRAW+10** | | | | |
| The STRAW+10 not used. | Postmenopause. | | | | |
| **Author reported facilitators to treatment** | | | **Author reported barriers to treatment** | | |
| N/A. | | | N/A. | | |
| **Study limitations** | | | | | |
| “The fact that no smoking effect was detected in the present study is curious, since many Blackfeet women smoke. [42] Perhaps the key variable to assess, if possible, would be smoking at the time of menopause, which was not evaluated here.” Page 64.  “The postmenopausal sample size was small and thus the results potentially susceptible to strong influence from the experience of a few women. Also, those who are in the sample, particularly in the older cohort, represent survivorship, and they may be different from their deceased cohort members in ways that relate to the menopause experience. Recall was an important methodological tool in this study, introducing the possibility of errors of memory.” Page 64. | | | | | |

| **Record ID** | **Title** | | | | |
| --- | --- | --- | --- | --- | --- |
| #739 – 2001 | Associations with age at natural menopause in Blackfeet Women | | | | |
| **Author information** | | | | | **Country** |
| Johnston, S. L. | | | | | United States |
| **Study aims** | | | | | |
| “This study seeks to determine the age at natural menopause and factors that may predict that age for Blackfeet women living on the reservation in Montana.” Page 513. | | | | | |
| **Study design** | **Conflicts of interest** | | **Inclusion criteria** | | **Exclusion criteria** |
| Cross-sectional study. | N/A; none listed. | | “Eligible women ≥ 18 years living on the reservation.” Page 513. | | “Women with surgical menopause or who were on hormone replacement therapy were not entered.” Page 514. |
| **Population and sampling description** | | **Indigenous description** | | **Total number of participants** | |
| The population of interest was Blackfeet women living in Montana.  “An age-stratified, probability sample of 321 women was drawn from the tribal enrollment list of 2,425 eligible women ≥18 years living on the reservation. Older age groups were oversampled to assure adequate representation.” Page 513.  “Because at least 27% of households on the reservation do not have telephones, contact was largely in person or through the mail.” Page 513. | | “Blackfeet women living in Montana”.  They used tribal enrollment lists to define their population.  The percentage of the sample who were Indigenous was 100%. | | The total sample size was150 participants, all were Indigenous. | |
| **Study outcomes and important results** | | | | | |
| “One hundred percent of the oldest cohort, 92.5% of women 50–69 years, and 19% of women 30–49 years had experienced menopause (Table 1). The surgical menopause rate, 42.9%, was high compared with the rate for the U.S. (34.5%) in the HES in 1960–1962 (MacMahon and Worcester, 1966), but was comparable to the rate (42.1%) in NHANES I in the 1970s (Hahn et al., 1997). The rate of surgical menopause for the entire sample was 20%. All surgeries occurred before natural menopause.” Pages 514 and 515.  “Median age at natural menopause by the status quo method was 51.2 years (95% CI 50.0–53.2 years), n 4 86 (Table 2). This is comparable to published data on median ages at menopause in the U.S. which range from 50.0 to 51.3 years (McKinlay et al., 1992b; MacMahon and Worcester, 1966; Hahn et al., 1997). Among the women in the sample who already had experienced natural menopause, the mean recalled age was 47.0 ± 5.0 years, median 47.0 years (n 4 33). The 70+ cohort had a mean age at natural menopause of 45.9 ± 3.9 years and a median of 45.0 years (n 4 12), and the 50–69 cohort had mean and median ages of 47.6 ± 5.5 years and 48.0 years, respectively (n 421). The difference between the cohorts was not significant. The mean and median ages at surgical menopause for the sample were 38.0 ± 8.7 years and 38.5 years, respectively(n 428); two women had missing or unreliable ages at surgery.” Page 515.  “Factors associated with age at natural menopause in the Blackfeet women who already had experienced it were subsequently considered (Tables 3 and 4). The relationship between menarche and age at menopause was examined using linear regression, and an inverse relationship (P=0.07) was found. For each year increase in age at menarche, age at menopause occurred 0.8 year earlier. There was one influential case in this regression (DFBeta=0.63) who had a later menarche and relatively late menopause; with this case removed, the relationship was significant (P=0.02). Age at menopause was associated positively with current household income. For each category increase in income, menopause occurred 1.85 years later (P 4 0.01). Women who ever used any form of birth control experienced menopause 4.52 years later than women who did not (P < 0.01); women who ever used oral contraceptives, 5.48 years later (P 0.01); women who were breastfed by their mothers experienced menopause 6.93 years earlier than women who were not (P=0.02); and recently or currently employed women had menopause 4.75 years later than unemployed women (P=0.02).” Page 515.  “There was no association with current age, parity, age at first live birth, menstrual cycle length, smoking (including current smoking, not shown), subject’s breastfeeding practices, anthropometric variables reflecting nutritional status (height, body mass index, skinfolds, arm muscle area, and centripetal fat ratio; last two not shown), or other sociodemographic variables such as education, language spoken in childhood home, and marital status (Table 4). However, when tertiles of women with the latest and earliest ages at menopause were compared, education was significantly different [P=0.04, Student’s t, mean 10.0 ± 3.6 years of education for the earliest tertile (n=15), 13.0 ± 2.2 years for the latest (n=9)].” Page 515.  “Variables with a significant bivariate association with age at natural menopause were entered into stepwise regression. The breastfeeding variable was not entered because of missing data for many women. All variables were retained except the employment variable (n=29, F=4.57, P=0.007, R2=0.43). However, and not surprisingly, the two contraceptive variables were significantly correlated with each other (r 4 −0.55 for the coefficients; r for the original variables 4 0.68, P < 0.001). For this reason, two separate stepwise regressions were run containing each of these variables and the remaining variables (age at menarche and household income). All variables were retained in each case. Of the two models, the one that explained more variance included ever use of oral contraceptives, age at menarche, and household income (n=29, F=5.81, P 4 0.004, R2=0.41). In this model, individual significance levels of household income and age at menarche were P=0.11 (r=0.31 for coefficients), suggesting some collinearity. Either age at menarche or household income, when modeled separately with ever use of oral contraceptives, was a significant predictor of age at natural menopause, with the model explaining 33%–35% of the variance (Table 5).” Pages 515 and 516.  “To further evaluate the relationship between income and age at menarche, the joint effect of the two variables on menopause age was examined using dummy variable regression (Table 6). A significant effect was found only for the group of women with later menarche and lower household income. This group’s mean age at natural menopause was 43.6 years compared with 48.8 years for the reference group (P=0.04). The effect held when oral contraceptive use was controlled in a subsequent regression.” Page 516.  “The high rate of surgical menopause may provide an explanation for the association between these variables. The remaining women ‘available’ for natural menopause may differ from those who underwent surgery. Among the Blackfeet, it is axiomatic that more ‘traditional’ women, those who adhere to indigenous beliefs and lifeways, have been historically less likely to utilize the Indian Health Service facilities. It is possible that women who have not experienced surgical menopause are more traditional. The data suggest that women in the natural menopause group who have had earlier menopause actually may be more traditional in their lifeway: they never have used birth control, they are poorer at present, they are currently and have been recently unemployed, and they were breastfed by their mothers. Given the economic and social history of the reservation, many women who currently have low income probably lived in poverty all of their lives (Johnston, 1999). Thus, whatever mediates the relationship between menarcheal age and income for this group could have occurred in childhood.” Page 516.  “The oldest cohort in this study was born between 1902 and 1925. These women were the children of the first Blackfeet confined to the reservation and required to subsist almost entirely on rations after the “starvation winter” of 1883–1884. Hunger and disease were rampant during the 1880s and 1890s when the parents of the oldest women in the study were growing and maturing. Intergenerational effects of poor growth or starvation conditions (Johnston et al., 1980; Garn et al., 1984; Lumey, 1992; Cameron, 1996) may have compromised the oldest cohort of women so that they began life at a disadvantage in terms of optimal growth and health. The period between 1902 and 1940 was a time of episodic food shortages due to drought, bad winters, and economic crises. Infectious diseases, especially tuberculosis, were the greatest causes of morbidity on the reservation, and health care was inadequate. Most of the women in this cohort grew up in rural areas of the reservation and were involved in physical work related to family ranching activities. As a whole, this cohort is significantly shorter in stature than any of the other three (Johnston, 1999). Childhood diet, disease, and activities of the oldest women in the 50–69 cohort would have been similar to those of the oldest group. Thus, history would seem to support the kind of interacting early-life effects of diet, energy balance, workloads, and disease stress that could have delayed menarche for some of the women and that potentially could have impacted ovarian function through later life.” Pages 517 and 518.  “Education and employment status may simply be alternative markers of current poverty. Although it is possible that contraceptive use is related to menopause because access to contraceptives reflects greater income, two factors argue against this. First, the two variables behaved independently in the multivariate models. Second, access to contraceptives on the reservation has not been income-dependent (federal health care is provided to all resident tribal members). On the other hand, oral contraceptives (OC) may have an effect through hormonal mechanisms (van Keep et al., 1979), or they may be a marker for some other variable that was not measured directly.” Page 519. | | | | | |
| **Use of STRAW+10** | **Menopausal transition staging of sample using STRAW+10** | | | | |
| The STRAW+10 not used. | Postmenopause.  “Menopause was defined as the age at last menstrual period if at least a 12-month interval had elapsed since that period.” Page 514. | | | | |
| **Author reported facilitators to treatment** | | | **Author reported barriers to treatment** | | |
| N/A. | | | “Among the Blackfeet, it is axiomatic that more ‘traditional’ women, those who adhere to indigenous beliefs and lifeways, have been historically less likely to utilize the Indian Health Service facilities.” Page 517. | | |
| **Study limitations** | | | | | |
| “The sample was small, and the detected effects could be influenced by the experience of just a few women.” Page 516.  “Also, women in the sample who had experienced menopause, especially in the oldest cohort, were the surviving representatives of a much larger group. Deceased women’s life histories may have differed in ways that could affect actual or expected age at menopause.” Page 516.  “Much of the data, notably age at menarche, menstrual cycle characteristics, and breastfeeding habits, were obtained through recall of earlier life events and practices, and were thus subject to error associated with recall and other factors. Unfortunately, there is no other feasible way to obtain these kinds of data from a community-based sample of women in their middle and older years.” Page 516.  “Another concern in interpreting the contraceptive data is the temporal aspects of OC usage (Stanford et al., 1987). These drugs were not available until at least 1960, so the group of older non-users in the study included women who chose not to use them as well as women who experienced menopause before OC availability.” Page 519. | | | | | |

| **Record ID** | **Title** | | | | |
| --- | --- | --- | --- | --- | --- |
| #671 – 2005 | Risk of cardiovascular disease by hysterectomy status, with and without oophorectomy: The Women’s Health Initiative observational study | | | | |
| **Author information** | | | | | **Country** |
| Howard, B. V.; Kuller, Lewis; L., Robert; Manson, J. E.; Allen, C.; Assaf, A.; Cochrane, B. B.; Larson, J. C.; Lasser, N.; Rainford, M.; Van Horn, L.; Stefanick, M. L.; Trevisan, M.; & Women’s Health Initiative | | | | | United States |
| **Study aims** | | | | | |
| “We sought to examine the hypothesis that the prevalence of CVD risk factors and rates of CVD may vary in women by hysterectomy status.” Page 1463.  “This data set provides an opportunity to examine whether hysterectomy independently increases risk for CVD in postmenopausal women. In this article, rates of CVD, total mortality, and the prevalence of CVD risk factors are compared in women who had and had not undergone hysterectomy/ oophorectomy at the time of enrollment to explore the relationship of hysterectomy with or without oophorectomy to CVD risk factors and to CVD events.” Page 1463. | | | | | |
| **Study design** | **Conflicts of interest** | | **Inclusion criteria** | | **Exclusion criteria** |
| Cohort study. | N/A; none listed. | | “Postmenopausal women ages 50 to 79 years were recruited at 40 clinical centers throughout the United States between September 1994 and December 1998.” Page 1463. | | “The only exclusions were the expectation of not residing in the area for 3 years, having a medical condition associated with a predicted survival of 3 years, and inability to provide informed consent.” Page 1463. |
| **Population and sampling description** | | **Indigenous description** | | **Total number of participants** | |
| The population of interest was postmenopausal US women aged 50 to 79.  “93,676 women who were invited to join 1 of 2 randomized, controlled clinical trials (a diet trial and a hormone trial) but who were either ineligible or were not interested after initiating screening and who were interested in contributing natural history data.” Page 1463. | | American Indian/Alaska Native.  “Ethnicity was determined by self-report.” Page 1463.  The percentage of the sample who were Indigenous was 0.42% (n=390 out of 92,368). From Table 1 on page1464. | | 89914 (92368 were enrolled in the WHI Observational Study, but 2454 had missing or equivocal data on hysterectomy or oophorectomy status and were therefore eliminated from the analysis). Page 1464. | |
| **Study outcomes and important results** | | | | | |
| From Table 1: 49.2% of the American Indian/Alaska Native sample had had a hysterectomy. Of those who had had a hysterectomy, 43.2% had a hysterectomy with oophorectomy. The American Indian/Alaska Native subsample represented the largest portion of the total sample who had had a hysterectomy at the age of 40 or younger (49.5% of the American Indian/Alaska Native subsample had had their hysterectomy at age 40 or younger).  “Black, Hispanic, and American Indian women were more likely to have undergone hysterectomy” Page 1464.  “In black, Hispanic, and American Indian women, the age distribution at the time of hysterectomy was younger than in white or Asian/Pacific Islander women, with more of those women having a hysterectomy before 40 years of age.” Page 1464.  Overall results: “The present data show that women with a hysterectomy had more obesity and diabetes and a worse CVD baseline risk factor profile than women without a hysterectomy in almost all respects. They had higher rates of hypertension and hypercholesterolemia, less physical activity, and higher saturated fat intake. In general, the adverse risk factor profile was present in women with a hysterectomy with or without oophorectomy, although risk factor profiles generally were worse in women who had also undergone oophorectomy. Although some of the differences in risk factors are small, several (hypertension, diabetes) were of clinical significance and taken together are predictive of increased CVD risk. The women who had undergone hysterectomy had lower average family incomes and less education.” Page 1468. | | | | | |
| **Use of STRAW+10** | **Menopausal transition staging of sample using STRAW+10** | | | | |
| The STRAW+10 not used. | Postmenopause.  “Postmenopausal status was defined as absence of menstrual bleeding for 1 year if under age 55 years and for 6 months if over age 55.” Page 1463. | | | | |
| **Author reported facilitators to treatment** | | | **Author reported barriers to treatment** | | |
| N/A. | | | N/A. | | |
| **Study limitations** | | | | | |
| “Some limitations of the study include that this is not a population-based sample of women, although the participants represent a wide range of ethnicity, education, socioeconomic status, and geographic distribution. In addition, hysterectomy and oophorectomy status, as well as other reproductive factors, were all obtained from self-report and not confirmed by medical record review. On the other hand, other studies have reported reasonable validity for self-report of hysterectomy. [25,26] Any inaccuracy in reporting could have obscured differences between the hysterectomy-alone and hysterectomy- plus-oophorectomy groups.” Page 1468.  “Any ethnic differences must be interpreted with caution, because the smaller number of women in the minority groups makes it less likely that they are representative of the ethnic group as a whole.” Page 1468. | | | | | |

| **Record ID** | **Title** | | | | |
| --- | --- | --- | --- | --- | --- |
| #621 – 1975 | The effect of menopause on serum cholesterol in American (Pima) Indian women | | | | |
| **Author information** | | | | | **Country** |
| Hamman, R. F.; Bennett, P. H.; & Miller, M. | | | | | United States |
| **Study aims** | | | | | |
| Serum cholesterol levels in Pima Indian women were investigated to determine if menopause caused these levels to rise. | | | | | |
| **Study design** | **Conflicts of interest** | | **Inclusion criteria** | | **Exclusion criteria** |
| Cross-sectional study. | N/A; none listed. | | N/A; none listed. | | “Twenty-nine women with a surgical menopause were excluded.” Page 165. |
| **Population and sampling description** | | **Indigenous description** | | **Total number of participants** | |
| “The Pima, a tribe of American Indians living in the arid southwestern United States have been under surveillance since 1965. [3] Those subjects who were at least four-eighths Indian, living on or near the Gila River Reservation have been examined at two-year intervals. There were 319 women aged 34-54 residing on the reservation who were examined at least once and for whom data were available. This represented 82 percent of the on-reservation population in this age group. Ninety-two additional women living off-reservation were included in the sample after analysis revealed that they did not differ significantly in weight, mean cholesterol, age at menopause and two-hour plasma glucose from those residing on the reservation. There were, therefore, 411 women available for cross- sectional analysis. Data for males and older females examined under similar circumstances were included for comparison.” Page 165. | | The Pima.  The authors do not describe how Pima racial/ethnic background was defined.  The percentage of the sample who were Indigenous was 100%. | | 411 Pima women (319 living in reservation, 92 living off-reservation). Page 165. | |
| **Study outcomes and important results** | | | | | |
| “There was no significant difference between the serum cholesterol levels of the pre- and post-menopausal women. ... Table 2 and figure 2 show that while the serum cholesterol levels were somewhat higher in the pre-menopausal women (183.3 ± 3.8 mg/100 ml, No. = 148) compared with post-menopausal levels (177.2 ± 5.0 mg/100 ml, No. = 75), this was not significant (p> 0.2).” Page 166  “The median age at menopause for these 65 women was 46 years, with a range of 34-53 years. The age at menopause for all women was not determined since recall was incomplete among the older women, and a marked digit preference was noted for ages 40 and 50, which was not present in the prospective sample. Figure 3 illustrates that as these 65 women passed through menopause, and for up to five years after, there was no change in the cholesterol level.” Page 166.  “Serum cholesterol levels in Pima Indian females showed no association or alteration with menopause.” Page 167. | | | | | |
| **Use of STRAW+10** | **Menopausal transition staging of sample using STRAW+10** | | | | |
| The STRAW+10 not used. | Unclear (may include participants that could be classified as in Reproductive, Menopausal Transition, or Postmenopause).  “Menopause was determined by asking each woman if she had noted cessation of menses, and if so, at what age.” Page 165. | | | | |
| **Author reported facilitators to treatment** | | | **Author reported barriers to treatment** | | |
| N/A. | | | N/A. | | |
| **Study limitations** | | | | | |
| “Because of possible racial or environmental differences, white female populations must be examined in a similar fashion before our findings can be considered generally applicable.” Page 169. | | | | | |

| **Record ID** | **Title** | | | | |
| --- | --- | --- | --- | --- | --- |
| #523 – 2003 | Predictors of postmenopausal body mass index and waist hip ratio in the Oklahoma Postmenopausal Health Disparities Study | | | | |
| **Author information** | | | | | **Country** |
| Gavaler, J. S.; & Rosenblum, E. | | | | | United States |
| **Study aims** | | | | | |
| “The goal of the current study has been to examine systematically the respective roles of nutrition, exercise, moderate drinking and smoking, as well as the role of postmenopausal endocrine factors including the use of ERT and hormonal status, in a setting in which race and ethnicity can simultaneously be evaluated as determinants of BMI and WHR.” Page 270. | | | | | |
| **Study design** | **Conflicts of interest** | | **Inclusion criteria** | | **Exclusion criteria** |
| Cross-sectional study. | N/A; none listed. | | Inclusion criteria are not discussed in this publication. “The study protocol has been described previously [39–45].” Page 270.  39. Gavaler JS, Bonham-Leyba M, Castro CS, Harman S: The Oklahoma postmenopausal women’s health study: Recruitment and characteristics of American Indian, Asian, Black, Hispanic and Caucasian women. Alcohol Clin Exp Res 23:220–223, 1999.  40. Gavaler JS, Galvao-Teles A, Deal S, Monteiro E: Mechanisms for the increase in postmenopausal estradiol levels among moderate drinkers: Findings in a multi-racial multi-ethnic study population. Endocrinol Metab Nutr 10:1–7, 2001.  41. Gavaler JS: Oral hormone replacement therapy: Factors which influence the estradiol level achieved in a multi-racial study population. J Clin Pharm 42:137–144, 2002.  42. Gavaler JS: Alcohol and nutrition in postmenopausal women. J Am Coll Nutr 12:347–356, 1993.  43. Gavaler JS, Deal SR, Van Thiel DH, Arria A, Allan MJ: Alcohol and estrogen levels in postmenopausal women: The spectrum of effect. Alcohol Clin Exp Res 17:786–790, 1993. 44. Gavaler JS: Effects of alcohol on endocrine function in postmeno- pausal women: A review. J Stud Alcohol, 46:495–516, 1985. 45. Gavaler JS, Love K, Van Thiel DH, Farhott S, Gluud C, Montiero E, Galvao-Teles A, Canton-Ortega T, Cuervas-Mons V: An international study of the relationship between alcohol consumption and postmenopausal estradiol levels. Alcohol Alcohol 1(Suppl): 327–330, 1991. | | N/A; none listed. |
| **Population and sampling description** | | **Indigenous description** | | **Total number of participants** | |
| “The study population was composed of 649 PMP women: 226 (34.9%) American Indians, 21 (3.2%), Asians, 78 (12.0%), Black, 54 (8.3%), Hispanics and 270 (41.6%) Whites.  Recruitment occurred between 1994 and late 1999 in Oklahoma, primarily Oklahoma City, except for American Indians.” Page 270  “The study population is a convenience sample composed of participants recruited using various methods including word of mouth, study brochures and posters as well as talks about menopause given to both small and large groups.” Page 270. | | American Indians.  No description is provided for how this is defined.  The percentage of the sample who were Indigenous is 34.9% (n=226). Page 270. | | Total number of participants was 649, and the total number of Indigenous participants was 226. | |
| **Study outcomes and important results** | | | | | |
| The percentage of no ERT use among the American Indian subsample was 46%. The percentage of the American Indian subsample who had menopausal weight gain was 83%. From Table 1 on page 271. ERT=estrogen/hormone replacement therapy  “Menopausal weight gain was significantly correlated with BMI in the total sample as well as in American Indian and Hispanic postmenopausal women” Page 272.  “Among American Indian postmenopausal women, WHR, FSH and E2, as well as menopausal weight gain and carbohydrates as percent of total calories, were found to be significant predictors of BMI. However, when WHR was not specified in the model, smoking, mean fitness, SES and age were also significant predictors of BMI” Page 272. WHR=waist hip ratio; FSH=follicle stimulating hormone; E2= Estradiol; SES=socioeconomic status.  The statistical predictors of WHR are shown in Table 4. “In the total study population with BMI in the equation, significant predictors included BMI and the neuroendocrine variables of FSH, E2, but not ERT, as well as the interaction of higher intensity exercise fitness with frequency, socioeconomic status and being American Indian or Asian postmenopausal women. When BMI was not included in the model, in addition to the neuroendocrine factors, the interaction of lower intensity exercise fitness with frequency, fat as percent of total calories, age, living alone and being American Indian or Black were significant predictors of WHR.” Page 272.  “Among American Indian postmenopausal women, in addition to BMI, both FSH and E2, as well as SES and age, were significant determinants of WHR; with BMI not in the equation, fat as percent of total calories was also a significant predictor of WHR” Page 272. | | | | | |
| **Use of STRAW+10** | **Menopausal transition staging of sample using STRAW+10** | | | | |
| The STRAW+10 not used. | Postmenopause. | | | | |
| **Author reported facilitators to treatment** | | | **Author reported barriers to treatment** | | |
| N/A. | | | N/A. | | |
| **Study limitations** | | | | | |
| N/A; none described. | | | | | |

| **Record ID** | **Title** | | | | |
| --- | --- | --- | --- | --- | --- |
| #522 – 1999 | The Oklahoma Postmenopausal Women’s Health Study: Recruitment and characteristics of American Indian, Asian, Black, Hispanic, and Caucasian women | | | | |
| **Author information** | | | | | **Country** |
| Gavaler, J. S.; Bonham-Leyba, M.; Castro, C. A.; & Harman, S. E. | | | | | United States |
| **Study aims** | | | | | |
| “The purpose of this study is to describe relevant aspects of our successful recruitment experience in Oklahoma so that they may be readily implemented by other investigators.” Page 220. | | | | | |
| **Study design** | **Conflicts of interest** | | **Inclusion criteria** | | **Exclusion criteria** |
| Cohort study. | N/A; none listed. | | Postmenopausal women from all ethnic/racial groups. | | “Initially, we were turning away volunteers who were using HRT; however, we changed the study entry criteria and allowed women who were using HRT to qualify for a substudy of women using HRT.” Page 221. |
| **Population and sampling description** | | **Indigenous description** | | **Total number of participants** | |
| The study’s population of interest included 1) American Indian (American Indian or Blend of White and American Indian) 2) Asian 3) Black 4) Hispanic 5) White.  “In the first year of the study, recruitment methods included the creation and distribution of a study brochure that described the purpose and importance of the study for postmenopausal women, what participation in the study involved, and that a $25 ‘thank-you’ payment would be made upon the participant’s completion of the study. This brochure contained a detachable postage-paid card that a potential participant could conveniently mail to us. The study brochure was placed in various areas of the Baptist Medical Center located in Oklahoma City, including the elevators.” Pages 220 and 221.  “By 1995, newspaper articles about the study published in the major daily paper increased inquiries about the study. In response to participants’ questions, we redesigned the study brochure to include more educational information about menopause; matching posters with pull-offs containing our 800 phone number were also printed. Brochures and posters were placed at many locations across the OUHSC campus, as well as at Community Centers frequented by specific minority groups, thereby expanding our population base beyond the health care setting. In the course of obtaining permission to place the study brochures and posters, our networking of community contacts increased substantially. At their request, the Primary Investigator (PI) began to present question-and-answer sessions about menopause to potential participants brought together by the directors of these community centers.” Page 221.  “In 1996, our networking efforts have reached many potential participants at outlying community centers, primarily American Indian centers. At their request, we began taking the study on-site to small groups of potential study participants. The PI and staff would travel with all necessary equipment and materials to the requesting site. These mobile clinics became possible when a trained phlebotomist joined our research staff.” Page 221.  “In 1997, we were fortunate to hire an additional minority staff member who is Hispanic. He has been instrumental in translating our study brochure, poster, and questionnaires into Spanish, as well as in establishing contacts within the Hispanic community. Also in 1997, we began a collaboration with a Professor of Nutrition who has been instrumental in recruiting and obtaining data from non-English-speaking women from the Asian community. In general, as we expanded the racial/ethnicity of our staff, a commensurate increase in the participation of minority groups of women dramatically increased. This result lends credence to the notion that cultural lines must be established between the research group and the groups we wish to study.” Page 221. | | American Indians and Blended American Indians constituted 35.6% of population. Table 3, Page 222.  Definition: “Oklahoma has a high proportion of American Indian (hind) and White/Amind Blend individuals. Health care coverage and educational benefits are associated with tribal enrollment. Because of carefully kept records, we created a separate category of White/Amind Blend, based on degree blood being less than one half. Because we are looking at biological endpoints, we decided to use this categorization to better sort out genetic influences.” Pages 221 and 222.  Table 1. “Recruitment Pattern: 1994 to 1998” includes “Terms used—Amind = American Indian: degree blood > 50%; Wham = Blend of White and American Indian: degree blood < 50%.” Page 221. | | A total of 174 Indigenous participants were included in the sample (107 American Indian and 67 White/American Indian Blend).  “For the period 1994 through 1997, 491 postmenopausal women from all racial/ethnic groups in the community have participated in this study.” Abstract. | |
| **Study outcomes and important results** | | | | | |
| “There is statistically significant heterogeneity among ethnic and racial groups with respect to sociodemographic factors (e.g., medical coverage, employment rates, and the distribution of educational levels), the prevalence of health behaviors (e.g., smoking, use of HRT, and moderate consumption of alcoholic beverages), and physical characteristics (i.e., indices of body fat distribution). Based on the overall heterogeneity among ethnic/racial groups, it is clear that any conclusions based on an all-Caucasian sample would be erroneous if applied to postmenopausal women in other ethnic/racial subgroups. ... In the AARP study, the percentages of older women who have not completed high school are: White, 2%; Black, 6%; American Indian, 12%; and Hispanic, 16%. This gradient of ‘uneducated’ women (Hispanic > American Indian > Black > White) in the AARP study is the same as in our study.” Page 222.  Table 3. “Ethnic Diversity and the Postmenopause: Health Behaviors” lists “Use of HRT” and “Ovariectomized” for the American Indian and White and American Indian subsamples. For the American Indian subsample, 46.3% of the sample used HRT and 31.5% were described as “Ovariectomized”. For the White and American Indian subsample, 59.4% of the sample used HRT and 53.6% were described as “Ovariectomized”. The Hispanic subsample had the lowest use of HRT at 37.0% and the Black subsample had the highest use of HRT at 64.4%. The Asian subsample had the lowest rate of being “Ovariectomized” at 11.1% and the White and American Indian subsample had the highest rate of being “Ovariectomized”. | | | | | |
| **Use of STRAW+10** | **Menopausal transition staging of sample using STRAW+10** | | | | |
| The STRAW+10 not used. | Postmenopause. | | | | |
| **Author reported facilitators to treatment** | | | **Author reported barriers to treatment** | | |
| N/A. | | | N/A. | | |
| **Study limitations** | | | | | |
| N/A; none described. | | | | | |

| **Record ID** | **Title** | | | | |
| --- | --- | --- | --- | --- | --- |
| #521 – 2002 | Oral hormone replacement therapy: Factors that influence the estradiol concentrations achieved in a multiracial study population | | | | |
| **Author information** | | | | | **Country** |
| Gavaler, J. S. | | | | | United States |
| **Study aims** | | | | | |
| “The purposes of this study were (1) to evaluate serum E2 concentrations in a multiracial study population of postmenopausal women already being treated chronically with HRT by their physicians and (2) to identify factors that might modulate the concentrations of E2 achieved in response to HRT treatment, with particular attention to the role of moderate alcoholic beverage consumption.” Page 137. | | | | | |
| **Study design** | **Conflicts of interest** | | **Inclusion criteria** | | **Exclusion criteria** |
| Cross-sectional study. | N/A; none listed. | | “Inclusion criteria included at least one full year since the last episode of menstrual bleeding or bilateral ovariectomy.” Page 138. | | “Exclusion criteria included unstable disease of any major organ system or a history of either cancer or alcoholism.” Page 138. |
| **Population and sampling description** | | **Indigenous description** | | **Total number of participants** | |
| “Among the 309 women in the current study population receiving HRT, 103 (33.3%) were American Indians, 8 (2.6%) were Asian Americans, 47 (15.2%) were African Americans, 16 (5.2%) were Latin Americans, and 135 (43.7%) were European Americans.” Page 138.  Sampling: “Details of the recruitment of this multiracial population have been published recently. [16]”  Reference:  Gavaler JS, Bonham-Leyba M, Castro CS, Harman S: The Oklahoma postmenopausal women’s health study: recruitment and characteristics of American Indian, Asian, Black, Hispanic and Caucasian women. Alcoholism: Clin Exp Res 1999;23:220-223 | | American Indians.  Self-report was used.  The percentage of the sample who were Indigenous was 33.3%. | | A total of 649 postmenopausal women participated in the overall study (226 of these were American Indian); 309 participated in the current study, of which, a total of 103 were American Indians. | |
| **Study outcomes and important results** | | | | | |
| “There was significant heterogeneity (p = 0.012) in the use of oral HRT among racial groups (Table I). The percentage of HRT-treated women with estradiol levels of at least 45 pg/ml (achievers) in the total study population was 51.1%. Response rates did not differ statistically among racial groups.” Page 139.  Table 1 “Study Population”: For the American Indian group, 122 (34.1%) were HRT users. Page 139.  Table 2 “Characteristics of the Study Population of Women Taking Oral Hormone Replacement Therapy (HRT)” lists “Menopause duration (years)” which was 10.4±0.8 for the American Indian subsample. The “HRT use duration (years)” for the American Indian sample was 9.4±0.8. “Bilateral ovariectomy” was reported for 58 (56.3%) of the American Indian subsample. “Natural menopause” was reported for 29 (28.2%) of the American Indian subsample. “Hysterectomy alone” was reported for 16 (15.4%) of the American Indian subsample. For “Hormone regimen” for the American Indian subsample, “Estrogen alone” was reported for 77 (74.8%), “Estrogen + progestin” was reported for 24 (23.3%), “Estrogen + testosterone” was reported for 2 (1.9%). The “Estrogen type” for the American Indian subsample was “Conjugated” type for 83 (80.6%), “Micronized” type for 13 (12.6%), “Esterified” type for 4 (3.9%), and “Estropipate” type for 3 (33.3%).  “In this snapshot of HRT-treated postmenopausal women simply living their lives, approximately half of these 309 women did not achieve circulating E2 concentrations of at least 45 pg/ml (165 pmol/l). Interestingly, there were no differences among racial/ethnic groups in achievement rates.” Page 140. | | | | | |
| **Use of STRAW+10** | **Menopausal transition staging of sample using STRAW+10** | | | | |
| The STRAW+10 not used. | Postmenopause.  “Menopausal status, particularly in women who had undergone a simple hysterectomy, was confirmed by a follicle-stimulating hormone (FSH)/luteinizing hormone (LH) ratio greater than a value of 1, as FSH levels are always higher than LH levels in the postmenopausal state.” Page 138. | | | | |
| **Author reported facilitators to treatment** | | | **Author reported barriers to treatment** | | |
| N/A. | | | N/A. | | |
| **Study limitations** | | | | | |
| “The findings of this study may be best viewed as being more seminal than definitive. At the very least, we hope that other investigators will continue to address the question of adequacy of HRT treatment, taking a broader view that incorporates the factors that modulate the E2 concentrations achieved among women treated with HRT. As E2 levels are essentially used as a surrogate marker for estrogenization, perhaps what is needed is a reliable measure of estrogenization.” Page 143. | | | | | |

| **Record ID** | **Title** | | | | |
| --- | --- | --- | --- | --- | --- |
| #469 – 2018 | The genetic underpinnings of variation in ages at menarche and natural menopause among women from the multi-ethnic Population Architecture using Genomics and Epidemiology (PAGE) Study: A trans-ethnic meta-analysis | | | | |
| **Author information** | | | | | **Country** |
| Fernandez-Rhodes, L.; Malinowski, J. R.; Wang, Y.; Tao, R.; Pankratz, N.; Jeff, J. M.; Yoneyama, S.; Carty, C. L.; Setiawan, V. W.; Le Marchand, L.; Haiman, C.; Corbett, S.; Demerath, E.; Heiss, G.; Gross, M.; Buzkova, P.; Crawford, D. C.; Hunt, S. C.; Rao, D. C.; Schwander, K.; Chakravarti, A.; Gottesman, O.; Abul-Husn, N. S.; Bottinger, E. P.; Loos, R. J. F.; Raffel, L. J.; Yao, J.; Guo, X.; Bielinski, S. J.; Rotter, J. I.; Vaidya, D.; Chen, Y.-D. I.; Castaneda, S. F.; Daviglus, M.; Kaplan, R.; Talavera, G. A.; Ryckman, K. K.; Peters, U.; Ambite, J. L.; Buyske, S.; Hindorff, L.; Kooperberg, C.; Matise, T.; Franceschini, N.; & North, K. E. | | | | | United States |
| **Study aims** | | | | | |
| “We sought to analyze the roughly 200,000 SNPs genotyped on the MetaboChip (Illumina, Inc., San Diego, CA, USA), a high-density genotyping array of primarily cardiometabolic loci [35], for association with reproductive milestones in the ancestrally diverse study participants of the PAGE Study [34]. Given the known overlap between the genetic underpinnings of AAM, and related cardiometabolic traits [22], the MetaboChip provides a densely genotyped resource to search for novel reproductive associations and broadly investigate the overlap of cardiometabolic and reproductive traits. Using race/ethnicity-stratified meta- analyses (20,398 African American, 15,856 Hispanic/Latina, 8,572 Asian American, and 538 American Indian/Alaskan Native women) and a trans-ethnic modified random-effects meta-analysis of up to 42,826 ancestrally diverse women, we sought to (i) establish how many index AAM and ANM SNPs, previously described in European-descent populations, also generalize to diverse racial/ethnic groups of women in the PAGE Study, (ii) their trans-ethnic localization, and (iii) to identify novel AAM or ANM associations on the MetaboChip.” Page 3 Abbreviations: SNPs: Single Nucleotide Polymorphisms PAGE Study: Population Architecture using Genomics and Epidemiology (PAGE) Study AAM: age at menarche ANM: age at natural menopause. | | | | | |
| **Study design** | **Conflicts of interest** | | **Inclusion criteria** | | **Exclusion criteria** |
| Cross-sectional study. | “Competing interests: Two coauthors were affiliated with commercial organizations (JRM at Write InSciTe, LLC, and JMJ at the Genotyping Arrays Division at Illumina, Inc.) at the time of the completion of this manuscript. These affiliations did not alter our adherence to PLOS ONE policies on sharing and data materials. Therefore, the coauthors collectively have declared that no competing interests exist.” Page 5. | | Inclusion criteria were based on the larger parent study (the PAGE Study). | | “S2 Text provides additional information on the following person-level exclusions. Briefly, we identified and excluded individuals with high inbreeding coefficients, F > 0.15 [40], and either excluded one woman of each 1st degree relative pair [41], or modeled relatedness using generalized estimating equations [42] and linear mixed models [43]. We generated principal components using Eigensoft for each study [44,45] and excluded ancestral outliers [46]. We excluded samples with phenotype-genotype sex discordance, low person-level call rate (<95%), or excessive heterozygosity. Only study- and race/ethnic-specific sample sizes of 50 women or more were carried for- ward to statistical analyses and summarized descriptively in S3 and S4 Tables.” Pages 4 and 5 .  “We excluded American Indian/Alaskan Native women from the trans-ethnic meta-analyses, due to their relatively small sample size as compared to the combined trans-ethnic sample of the other racial/ethnic groups (1% for both AAM and ANM).” Page 6. |
| **Population and sampling description** | | **Indigenous description** | | **Total number of participants** | |
| The population of interest was African American, Hispanic/Latina, Asian American, and American Indian/Alaskan Native women.  Participants for this meta-analysis came from the broader parent study called the PAGE study: “The PAGE Study was designed to generalize and estimate common genetic effects across multiple ancestral populations [34]. Briefly, the first phase of the PAGE study was comprised of a coordinating center, four large study sites/consortia [Causal Variants Across the Life Course (CALiCo) Consortium, including the Atherosclerosis Risk in Communities (ARIC) Study, Coronary Artery Risk Development in Young Adults (CARDIA), the Hispanic Community Health Study/Study of Latinos (HCHS/SOL); Epidemiologic Architecture for Genes Linked to Environment (EAGLE)-accessing the Vanderbilt University Medical Center’s biorepository (BioVU); Multiethnic Cohort (MEC); the Women’s Health Initiative (WHI)], and additional collaborating studies [The Hypertension Genetic Epidemiology Network (HyperGEN) Study, the MEC-Slim Initiative in Genomic Medicine for the Americas Type 2 Diabetes Consortium (MEC-SIGMA), Multi-Ethnic Study of Atherosclerosis (MESA), and Mount Sinai School of Medicine BioBank (BioME)].” Page 4.  Sampling: “Collectively, the studies represent 20,398 African American, 15,856 Hispanic/Latina, 8,572 Asian American, and 538 American Indian/Alaskan Native women who provided informed consent (or in the case of BioVU, did not opt out from the approved research), and had complete information on genetics, reproductive phenotypes and covariates.” Page 5. | | American Indian/Alaskan Native.  There are no details about how women were categorized as AI/AN.  The percentage of the sample who were American Indian/Alaska Native was 2.6% (page 5) and the percentage of the Indigenous sample who were included in the age at natural menopause analysis was 1.1%. Page 7. | | “n=45364” Abstract.  Table 1, The “Age at natural menopause” sample size was n=17,100; the AI/AN sample for natural menopause was n=184. Page 7. | |
| **Study outcomes and important results** | | | | | |
| From Table 1: The median age at natural menopause for the AI/AN subsample was 50. Note that the Median birth year for this subsample was 1935. Page 7.  “The first index SNP at MCM8, [rs236114 [19]] was in moderate LD (1000 Genomes AMR r2 = 0.36) with another SNP, rs16991615 [15,18,23], which was both Bonferroni significantly associated in Hispanic/Latinas (TDAF 94%, p-value = 7.14x10-6) in the trans-ethnic sample (TDAF 97%, p-value = 1.91x10-6; S2 Fig) and associated with ANM in a directionally-consistent manner across all race/ethnic groups. Yet within the 9 Hispanic/Latina studies this SNP association exhibited evidence of effect heterogeneity (p-value heterogeneity = 3.43x10-4), which in S3 Fig appeared to be driven by ANM-increasing effects among the MEC and MEC SIGMA Type 2 Diabetes cases, which was inconsistent with the previously reported ANM reducing allele and with the observed direction of effect in the WHI Native American/American Indian subsample. Although this apparent effect heterogeneity could be due to chance, it is also possible it could reflect differences in relevant pre-menopausal environments/health statuses of these specific MEC subsamples (e.g. gene-environment interactions).” Page 9.  “Our trans-ethnic meta-analysis of reproductive traits has expanded our understanding of the transferability of reproductive trait loci discovered in women of European descent to other race/ethnic groups. First, we observed more directionally consistent trans-ethnic associations than we expected by chance across all 32 AAM and 14 ANM loci on the MetaboChip (p-valuesbinomial of 1.2x10-6 and 0.01, respectively). Second, we generalized six AAM loci (NUCKS1, LIN28B TMEM38; SEC16B, BDNF, FTO), and two ANM loci (BRSK1, MCM8) to African, Hispanic/Latina, Asian or American Indian/Alaskan Native women, observing at each locus directional consistency between our trans-ethnic risk alleles and previous reports among European descent women [13,15,17–19]. This suggests that much of the currently known genetic architecture for AAM and ANM appears to be transferable across ancestrally diverse racial/ethnic groups.” Pages 13 and 14. | | | | | |
| **Use of STRAW+10** | **Menopausal transition staging of sample using STRAW+10** | | | | |
| The STRAW+10 not used. | Postmenopause.  Menopause was defined cessation of regular menses: “ANM (cessation of regular menses) in years were collected by questionnaire or via medical record” Page 4 Abbreviation ANM: Age at natural menopause.  Genotyping: “Forty-six index SNPs had been previously associated with either AAM or ANM (or if unavailable on the MetaboChip, a proxy SNP r2 ≥ 0.8 in 1000 Genomes CEU sample) and rep- resented distinct genetic loci (r2<0.2) (S1 Table). These SNPs included all two of the known AAM and five of the known ANM loci as of when the MetaboChip was designed, including the two strongest and most widely-generalizable AAM and ANM signals to date (LIN28B and MCM8) [22,23]. Additionally, seven of the previously associated SNPs were located within six densely-genotyped loci (S2 Table) that were associated with AAM or ANM after the initial design of the MetaboChip.” Page 4. | | | | |
| **Author reported facilitators to treatment** | | | **Author reported barriers to treatment** | | |
| N/A. | | | N/A. | | |
| **Study limitations** | | | | | |
| “Our results, however, are limited by the imbalance in sample sizes available for the AAM and ANM analyses, and the relatively low proportion of established AAM and ANM loci from studies of European descent women to date available on the MetaboChip (9% and 32%, respectively). Although our exclusion of extreme values restricted our analytic sample size, it allowed us to report on the common genetic causes of normal variation in AAM and ANM. Additionally, our use of the MetaboChip as a common dense-genotyping array to all PAGE studies, and with consistent genotype calling and quality control applied to each study, is a key strength of this study. Nonetheless our ascertainment of AAM and ANM did rely primarily on self-report. Lastly, the available sample size of racial/ethnic minority women who experienced these reproductive milestones is only a fraction of the samples of European descent women published on previously [22]. Due to relative paucity of genetic data on minority women, we did not have sufficient sample sizes to achieve statistical power to identify genetic associations of low frequency variants (MAF<5%) in most of our analyses, to seek independent replication of our novel significant findings (Table 4), or to systematically explore the role of gene-environment interactions on our findings. Yet, our observation of enrichment of directional consistency (S1 Table) suggests that given sufficient power or more comprehensive genotyping arrays, additional AAM and ANM loci may be significantly associated with reproductive traits in minority women. Larger samples of diverse women are needed to investigate all currently known AAM and ANM loci, establish statistical significance and describe the magnitude of the novel genetic effects on reproductive traits with more precision.” Page 14. | | | | | |

| **Record ID** | **Title** | | | | |
| --- | --- | --- | --- | --- | --- |
| #376 – 2023 | Treatment and resource utilization for menopausal symptoms in the United States: a retrospective review of real-world evidence from US electronic health records | | | | |
| **Author information** | | | | | **Country** |
| DePree, B.; Houghton, K.; Shiozawa, A.; Esterberg, E.; King, D. D.; Kim, J.; & Mancuso, S. | | | | | United States |
| **Study aims** | | | | | |
| “The current noninterventional, retrospective, observational cohort study aimed to identify and characterize the use of prescription and nonprescription therapies for VMS recorded by HCPs, bridge gaps in our knowledge of the treatment of VMS, and describe the broad spectrum of prescription and nonprescription therapies (including complementary therapies and behavioral/lifestyle modifications) used by women who experience VMS related to menopause.” Page 71. | | | | | |
| **Study design** | **Conflicts of interest** | | **Inclusion criteria** | | **Exclusion criteria** |
| Cohort study. | “Financial disclosure/conflicts of interest: B.D. serves on an advisory council for Astellas Pharma, Inc. K.H. and E.E. are employees of RTI Health Solutions, which provides consulting and other research services to pharmaceutical, device, governmental, and nongovernmental organizations. K.H. and E.E. work with many companies and do not receive payment or honoraria directly from these companies for services rendered. RTI Health Solutions received funding from Astellas Pharma, Inc., for the design and conduct of this study, statistical analysis, and study report preparation. A.S., D.D.K., J.K., and S.M. are employees of Astellas Pharma, Inc.” Page 70. | | “Electronic health records from women initially presenting with menopausal complaints between January 1, 2016, and December 31, 2019, who were 40 to 60 years of age at the time of initial presentation and were residing in the United States were included in the analysis. The women also must have been experiencing bothersome hot flashes (defined as daytime or nighttime sensation of heat with sweating) two or more times within a 24-hour period.” Page 70. | | “Women who experienced hot flashes for reasons other than menopause were excluded.” Page 70. |
| **Population and sampling description** | | **Indigenous description** | | **Total number of participants** | |
| US women aged 40 to 60 who had perimenopausal symptoms.  “The study was conducted by RTI Health Solutions (RTI-HS; Research Triangle Park, NC), in collaboration with the study sponsor (Astellas Pharma, Inc, Northbrook, IL). Participating HCPs were recruited from local medical association directories or from listings of previously conducted research (if they consented to be contacted for future studies) and were able to identify eligible medical records. Health care providers were gynecologists, internal medicine or family physicians, or advanced practice providers who typically consulted or treated at least three women per week presenting with menopausal complaints (including hot flashes), had prescriptive authority and access to and permission to use electronic health records (EHR) via the electronic system used by their practice, and practiced in the United States. Approximately 450 to 500 HCPs were invited to participate. Recruitment was guided by soft quotas to ensure that approximately 20% of the HCPs came from each US region (ie, Northeast, Southwest, West, Southeast, and Midwest) and that approximately 50% were practicing in gynecology and 50% in primary care.” Page 71. | | “Asian, Native American or Alaska Native”  This category is not defined. No rationale is listed for the combining of the Asian with the Native American or Alaska Native racial category.  The percentage of the sample who were “Asian, Native American or Alaska Native” was 8.2%. | | The total number of participants was1016, of which 83 were “Asian, Native American, or Alaska Native”. | |
| **Study outcomes and important results** | | | | | |
| “A greater percentage of White (88.1%) and Asian/Native American/Alaska Native women (84.3%) had private insurance compared with Black women (67.7%), whereas a higher proportion of Black women (22.2%) had Medicaid (vs 5.5% of White and 6.0% Asian/Native American/Alaska Native women); 1.1% of White, 0.6% of Black, and 7.2% of Asian/Native American/Alaska Native women were uninsured. Black women were more likely to have comorbidities than White and Asian/Native American/Alaska Native women (83.8% vs 59.9% and 49.4%, respectively).” Pages 73-74.  “At the initial visit, the most commonly reported symptoms of menopause for women of all races were hot flashes, followed by sleep problems, vaginal dryness, mood changes, reduced libido, dyspareunia, and weight gain/slowed metabolism (Table 3).” Page 74.  “Asian/Native American/Alaska Native women reported more thinning/dry hair (21.7% vs 9.0%-14.8%).” Page 74.  “Asian/Native American/Alaska Native women less commonly had prescription medications documented (49.4%) compared with women in all other groups (ie, White, 61.2%; Black, 62.9%; and other, 54.3%) and more commonly had nonprescription therapies documented (71.1%) compared with all other groups (White, 62.6%; Black, 58.1%; and other, 62.9%).” Page 74.  “Among women receiving prescription medication, 73.9% and 11.3% of White women, 60.0% and 18.1% of Black women, and 68.3% and 9.8% of Asian/Native American/Alaska Native women received HT and compounded HT, respectively (Table 3). Use of SSRIs was lowest among Asian/Native American/Alaska Native women (7.3%) compared with all other groups (14.5%-16.2%).” Page 74.  Table 3 Asian/Native American/Alaska Native results (Page 77): Menopausal symptoms—Hot flashes 73 (88.0); Vaginal dryness 38 (45.8); Vaginal atrophy 13 (15.7); Dyspareunia 21 (25.3); Mood changes 39 (47.0); Sleep problems 37 (44.6); Urinary problems 11 (13.3); Irregular periods 25 (30.1); Weight gain or slowed metabolism 16 (19.3); Thinning/dry hair 18 (21.7); Dry skin 11 (13.3); Reduced libido 20 (24.1); Difficulty concentrating 7 (8.4) Duration of symptoms—≤3 mon 20 (24.1); 3-6 mon 21 (25.3); 6-12 mon 22 (26.5); >12 mon 19 (22.9); Not documented 1 (1.2) Documented therapy—Prescription only 13 (15.7); Prescription and nonprescription 28 (33.7); Nonprescription only 31 (37.3) Prescription treatments—Hormone therapy (any) 28 (68.3); Compounded hormone therapy (any) 4 (9.8); SSRIs 3 (7.3); SNRIs 2 (4.9); Clonidine 2 (4.9); Gabapentin or pregabalin 4 (9.8) SERMs 0; Other 0 Nonprescription treatments documented—Black cohosh 12 (20.3); Estroven 9 (15.3); Vitamin E 10 (16.9); Exercise 21 (35.6); Sleep alterations 11 (18.6); Food or alcohol avoidance 10 (16.9).  “We also looked at symptoms and treatment by patient ethnicity. Whereas experience with menopausal symptoms was similar across ethnic groups, some differences were observed in treatment use. ... women who were Asian, Native American, or Alaska Natives were more likely to take nonprescription therapies and less likely to have documentation of prescription medications.” Page 78. | | | | | |
| **Use of STRAW+10** | **Menopausal transition staging of sample using STRAW+10** | | | | |
| The STRAW+10 not used. | Unclear; it is difficult to determine using the information available in the record.  Participants were aged 40 to 60. They were documented experiencing bothersome hot flashes related to menopause. | | | | |
| **Author reported facilitators to treatment** | | | **Author reported barriers to treatment** | | |
| “Research associated with the management of VMS suggests that the use of prescription and nonprescription therapies for VMS associated with menopause is not well understood. The current study provides strong evidence on the current management of VMS, the extent to which nonprescription therapy is documented in the EHR, and the HCRU associated with VMS in the United States. These data can help HCPs improve the management of VMS related to menopause and help fill the knowledge gaps to improve the care of women in the menopausal transition.” Page 78. | | | N/A. | | |
| **Study limitations** | | | | | |
| “The results of this study must be interpreted with caution as retrospective EHR review studies have a number of limitations. The women selected in this study may not represent the general population of women experiencing VMS, and the HCPs who participated in the study may have been more willing to diagnose and treat VMS. Demographic characteristics of our HCP and patient populations generally align with available US population statistics; however, our sample may have had a lower proportion of female HCPs (42% in the current study vs 46% of PCPs and 85.2% of gynecologists) and fewer women who were uninsured (2% in the current study vs 13% in women aged 18-64 years nationally). [19-26] In addition, the data were limited to information available in the EHR, and therefore, any information on nonprescription medications that were not reported to HCPs or not recorded by HCPs was unavailable for this study. Consequently, the proportion of women initiating nonprescription therapy may be underestimated. In addition, the inclusion criteria of two or more bothersome hot flashes in 24 hours led to the inclusion of women with milder symptoms. The number of patients receiving prescription or nonprescription treatment may be higher in women with more frequent or severe VMS. Symptom severity was noted as a key factor for HCP treatment selection in the qualitative interviews conducted as part of this study. [12] Data entered may also be subject to entry errors, resulting in inaccuracies in the EHR. Although data checks were conducted to assess internal consistency of the entered data, the responses were not validated by an independent reviewer.” Page 78. | | | | | |

| **Record ID** | **Title** | | | | |
| --- | --- | --- | --- | --- | --- |
| #330 – 1997 | Parity, postmenopausal estrogen use, and cardiovascular disease risk factors in American Indian women: The Strong Heart Study | | | | |
| **Author information** | | | | | **Country** |
| Cowan, L. D.; Go, O. T.; Howard, B. V.; Devereux, R. B.; Pettitt, D. J.; Fabsitz, R. R.; Lee, E. T.; & Welty, T. K. | | | | | United States |
| **Study aims** | | | | | |
| “We examined the relation of parity, adjusted for estrogen use, with selected CVD risk factors among a large population of American Indian women. Independent associations of postmenopausal estrogen use and CVD risk factors were also assessed.” Page 442 | | | | | |
| **Study design** | **Conflicts of interest** | | **Inclusion criteria** | | **Exclusion criteria** |
| Cross-sectional study. | N/A; none listed. | | Specific details about the parent study’s inclusion/exclusion criteria can be found in the methodology paper:  Lee ET, Welty TK, Fabsitz RR, et al. The Strong Heart Study. A study of cardiovascular disease in American Indians: Design and methods. Am J Epidemiol 1990;132:1141 | | “Excluded from the glucose tolerance test were participants with diabetes who were treated with insulin or oral hypoglycemic agents and any participant with a fasting glucose >=225 mg/dl as determined by an Accu-Check II (Baxter Healthcare Corporation, Grand Prairie, TX).” Page 442.  “Gravidity and parity information were available for all but seven of the 1982 ever-married, postmenopausal women in the present study. ‘Ever-married’ was used as an indirect measure of having had the opportunity to become pregnant. A total of 125 women was excluded from these analyses because they were never married and 2 because their marital status was unknown.” Page 443. |
| **Population and sampling description** | | **Indigenous description** | | **Total number of participants** | |
| The population of interest for this study was American Indian women participating in the Heart Strong Study.  “The three study centers include the Oglala and Cheyenne River Sioux Tribes in South Dakota and the Spirit Lake Sioux Tribe in North Dakota, the Gila River and Salt River Pima/ Maricopa and the Ak Chin Pima/Papago Indian communities near Phoenix, Arizona, and the seven Plains Indian Tribes of south-western Oklahoma (Apache, Caddo, Comanche, Delaware, Fort Sill Apache, Kiowa, and Wichita). The Dakota and Arizona Indian populations live on reservations; there are no longer reservations in Oklahoma.” Page 442.  Specific details about sampling can be found in the methodology paper:  Lee ET, Welty TK, Fabsitz RR, et al. The Strong Heart Study. A study of cardiovascular disease in American Indians: Design and methods. Am J Epidemiol 1990;132:1141 The percentage of Indigenous participants is 100%. | | “The three study centers include the Oglala and Cheyenne River Sioux Tribes in South Dakota and the Spirit Lake Sioux Tribe in North Dakota, the Gila River and Salt River Pima/ Maricopa and the Ak Chin Pima/Papago Indian communities near Phoenix, Arizona, and the seven Plains Indian Tribes of south-western Oklahoma (Apache, Caddo, Comanche, Delaware, Fort Sill Apache, Kiowa, and Wichita). The Dakota and Arizona Indian populations live on reservations; there are no longer reservations in Oklahoma.” Page 442.  The authors do not describe how racial/ethnic background was defined.  The percentage of the sample who were Indigenous is 100%. | | The total number of participants was 1982. All participants were AI women. | |
| **Study outcomes and important results** | | | | | |
| “The prevalence of surgical menopause varied across centers, with 44% of women in Oklahoma having had a surgically induced menopause, compared with 22% and 29% in the Dakotas and Arizona, respectively. The mean age at natural menopause was about 2 years earlier among Arizona women. The pattern of current use of postmenopausal estrogen reflected that for surgical menopause; use was much more common in Oklahoma (21 %) compared with the other two centers. Analyses of the prevalence of current estrogen use by type of menopause and SHS center indicated that use was highest among women at all three centers who had a hysterectomy with oophorectomy, but even among this group, the prevalence of estrogen use was nearly twice as high in Oklahoma women (44%) as in those from Arizona (22%) or the Dakotas (25%).” Page 445.  From Table 1: Postmenopausal natural vs. surgical by site— Arizona. 666 postmenopausal women. 469 (70%) had menopause as a result of natural causes. 98 (15%) had surgical menopause as a result of the removal of the uterus and ovaries. 92 (14%) had surgical menopause as a result of the removal of the uterus only. And, 7 (1%) had "Unknown type" of menopause. Mean Age at Natural Menopause (years plus or minus SD)=45.5 plus or minus 5.4. Oklahoma. 681 postmenopausal women. 366 (54%) had menopause as a result of natural causes. 179 (26%) had surgical menopause as a result of the removal of the uterus and ovaries. 126 (18%) had surgical menopause as a result of the removal of the uterus only. And, 10 (1%) had “Unknown type” of menopause. Mean Age at Natural Menopause (years plus or minus SD)=47.6 plus or minus 5.7. South Dakota, North Dakota. 635 postmenopausal women. 481 (76%) had menopause as a result of natural causes. 66 (10%) had surgical menopause as a result of the removal of the uterus and ovaries. 76 (12%) had surgical menopause as a result of the removal of the uterus only. And, 12 (2%) had “Unknown type” of menopause. Mean Age at Natural Menopause (years plus or minus SD)=47.3 plus or minus 4.9.  “The primary source of medical care at all three centers is the Indian Health Service (IHS), but more women from the Oklahoma center also receive care outside of the IHS system (unpublished SHS data).” Pages 446-447.  “Overall, 32% of postmenopausal women in the SHS reported a surgical menopause, which is similar to prevalence rates reported in other populations. 13,14 Differences among centers in the frequency of current estrogen use may also reflect extant geographic variations in the frequency of use of estrogen replacement in the menopause of 8%13 to >30%. 15-17” Page 447.  “The average age at natural menopause also varied across SHS centers, with the youngest age (45.5 years) observed in Arizona compared with 47.6 years and 47.3 years in Oklahoma and the Dakotas, respectively. The median age at natural menopause observed in SHS women (47 years) was 1-3 years younger than that reported in other population groups. 18,19 Whether this difference is due to a reporting artifact or whether it reflects true biologic differences in these populations is unknown.” Page 447. | | | | | |
| **Use of STRAW+10** | **Menopausal transition staging of sample using STRAW+10** | | | | |
| The STRAW+10 not used. | Postmenopause.  “Women were considered to be postmenopausal if their menstrual cycles had stopped completely for at least 12 months. Women whose menses had ceased were then asked whether their meno­pause was natural or surgical and, if surgical, whether only their uterus had been removed. Women who reported hysterectomy without oophorectomy or who did not know whether their ovaries had been removed were classified as postmenopausal if their age was ≥53 years, the age at which 90% of women in this cohort had a natural menopause.” Page 443. | | | | |
| **Differences between rural vs. urban outcomes** | | | | | |
| “Surgical menopause was nearly twice as prevalent among women in the Oklahoma center (44%) as in women in Arizona (29%) and in the Dakotas (22%).” Page 446.  “The difference in frequency of surgical menopause may also account for some, but not all, of the wide variation among centers in the frequency of current estrogen use, as differences in frequency of use persisted within categories of type of menopause.” Page 446. | | | | | |
| **Author reported facilitators to treatment** | | | **Author reported barriers to treatment** | | |
| N/A. | | | N/A. | | |
| **Study limitations** | | | | | |
| N/A; none described. | | | | | |

| **Record ID** | **Title** | | | | |
| --- | --- | --- | --- | --- | --- |
| #272 – 2016 | Experience of menopause in aboriginal women: A systematic review | | | | |
| **Author information** | | | | | **Country** |
| Chadha, N.; Chadha, V.; Ross, S.; & Sydora, B. C. | | | | | Canada |
| **Study aims** | | | | | |
| “The purpose of this review was twofold: to gather all literature that examines menopause experience in aboriginal women in order to (1) describe the breath of research conducted in this field, and (2) reach a greater understanding of the diverse knowledge, experience, and attitudes towards reproductive cessation and the menopause transition in these women.” Page 18. | | | | | |
| **Study design** | **Conflicts of interest** | | **Inclusion criteria** | | **Exclusion criteria** |
| Systematic review. | “Conflict of interest. The authors declare that there is no conflict of interest regarding the publication of this paper. The authors alone are responsible for the content and writing of this paper.” Page 24. | | “The following criteria were used for inclusion: the paper must study specifically aboriginal women and their menopause experience, including onset of menopause, menopause symptoms, attitude, experience, and knowledge about menopause and its management. Aboriginal was defined specifically as being native to the region or country with a characteristic biological, social, psychological, and cultural profile. No restriction was put on language or publication date.” Page 18. | | “We applied the following exclusion criteria: review articles, dissertations, and abstracts without data; papers including aboriginals, but where aboriginal women were not specifically the focus of study in the articles and did not form an identifiable group, i.e. they were only studied together with non-aboriginals in the same study group; articles describing their study group as ‘rural’ without using any of our search terms; and articles using the term ‘ethnic’ to describe their study groups. We also excluded studies that only assessed specific symptoms that are not exclusively related to menopause, for example sleep problems or osteoporosis.” Page 18. |
| **Population and sampling description** | | **Indigenous description** | | **Total number of participants** | |
| The population of interest for this study was perimenopausal aboriginal women.  Aboriginal group: 1) Indigenous of Central America (Mayan, Zapotec) 2) Indigenous of South America (Movima, Colombian Caribbean, Zenu, Wayuu, Amazone tribes, Quechua) 3) Native North American (Mohawk, Sioux, Navajo, Blackfeet, Native American) 4) Australian Aboriginals 5) New Zealand Maori 6) Rural tribes in India 7) Indigenous of Malaysia 8) Taiwanese Aboriginals. | | “Seven articles (two from Canada 27,28 and five from the US 29–33) described results from Native North American aboriginal women including Mohawk 27, Sioux 28, Navajo 29,30, Blackfeet 31,32, and Native Americans from the Pacific Northwest 33.” Pages 20-21. | | N/A; this review article included seven studies that were focused on Native North American women. (See Table 1, page 20). | |
| **Study outcomes and important results** | | | | | |
| “We were only able to collect 28 articles on this topic, published over a time frame of 33 years from 1981 to 2014.” Page 22.  “Despite substantial interest in menopause and its management in developed countries, our literature review found only a small number of original articles that studied menopause in aboriginal women. Collectively, the articles revealed considerable differences in menopause perception and attitude, onset, and symptom experiences among aboriginal groups. While many aboriginal women may lack menopause awareness and knowledge of menopause physiology, they may deal with the menopause transition period in their own cultural way. Our review highlights a need for further research into the menopause experience and management among aboriginal women. The knowledge gained can be used to help bridge possible gaps between aboriginal women and health-care professionals in order to further enhance an important dialogue about women’s health care.” Page 24.  “Onset of menopause was studied in 15 (54%) of the articles included in our review, with five articles studying solely menopause onset in their respective aboriginal women group. The data from these 15 studies revealed that, on average, the age of natural menopause onset is lower in aboriginals compared to non-aboriginals. This appears to be specifically true for aboriginal women in Central and South America and Asia, despite a generally lower age of menopause onset in these geographic regions compared to age of onset in women from Europe and North America [5]. In contrast, the menopause onset at age 50.78 years reported for indigenous women in Taiwan is higher than the range of median menopause age of 42.1–49.5 years reported for women in Asia [5].” Page 24.  “The reason for the reduced age at menopause among the aboriginal women in our selected articles was often hypothesized as due to malnutrition, harsh climate and living conditions, and lack of appropriate health care [31, 37, 38].” Page 24. | | | | | |
| **Use of STRAW+10** | **Menopausal transition staging of sample using STRAW+10** | | | | |
| The STRAW+10 not used. | Reproductive, Menopausal Transition, and Postmenopause.  “Most papers (17) included women in various stages of the menopause transition period. Four had a wide range of ages including women from age 18 on [16,31,32,34] to age 80 [16] and over 90 [31,32]. Those studies were about menopause knowledge generally and/or studied timing of menopause and therefore women of a wide age range could be included. The remaining seven articles did not specify age (Table 2). Three articles included women who had experienced surgical menopause due to ovary removal [28,34,35]. With regard to the menopause topic, the majority of the 28 studies looked at menopause symptoms in the form of symptom frequency, severity, or general experience (Tables 1 and 3); among them, one article studied urogenital symptoms only [23] and one studied vasomotor symptoms only [33]. Fifteen studies assessed menopause onset (Tables 1 and 3), of which five articles used menopause onset as the sole study topic [27,31,32,37,38]. Nine articles reported on menopause knowledge and awareness among aboriginal women (Table 1). Four studies compared aboriginal women at different menopause stages [16,17,24,40].” Page 20. | | | | |
| **Author reported facilitators to treatment** | | | **Author reported barriers to treatment** | | |
| N/A. | | | N/A. | | |
| **Study limitations** | | | | | |
| “Our literature search and final selection may not have captured all articles because of the extensive filtering that was applied for our searches in the ScienceDirect and PubMed databases. Other articles may have escaped our collection due to our limited use of keywords for indigenous/aboriginal groups. In particular, we did not use the term ‘rural population’ because of its lack of specificity. In developing our search strategy, we found this term might have identified research in other aboriginal groups, for example in India, Africa and China, but we were unable to definitively identify which were aboriginal populations. The differences we noted in papers describing the menopause experiences of distinct study populations or groups may have been as a result of specific methodological choices made by the investigators, rather than representing actual differences in women’s experience. As well, we found little evidence of culture-based treatments and practices associated with menopause, partly because these were not the focus of the studies we included, and also because aboriginal women tend to be reticent about their menopause experiences [42].” Page 24. | | | | | |

| **Record ID** | **Title** | | | | |
| --- | --- | --- | --- | --- | --- |
| #255 – 2013 | Replication of genetic loci for ages at menarche and menopause in the multi-ethnic Population Architecture using Genomics and Epidemiology (PAGE) study | | | | |
| **Author information** | | | | | **Country** |
| Carty, C. L.; Spencer, K. L.; Setiawan, V. W.; Fernandez-Rhodes, L.; Malinowski, J.; Buyske, S.; Young, A.; Jorgensen, N. W.; Cheng, I.; Carlson, C. S.; Brown-Gentry, K.; Goodloe, R.; Park, A.; Parikh, N. I.; Henderson, B.; Le Marchand, L.; Wactawski-Wende, J.; Fornage, M.; Matise, T. C.; Hindorff, L. A.; Arnold, A. M.; Haiman, C. A.; Franceschini, N.; Peters, U.; & Crawford, D. C. | | | | | United States |
| **Study aims** | | | | | |
| “We sought to replicate GWAS findings for AM and ANM in our large sample of European Americans and to investigate whether these associations are also consistent in women of diverse race/ethnicity, including American Indians, African Americans, Asians, Hispanics and Native Hawaiians from the Population Architecture using Genomics (PAGE) study” Page 1696 AM=Age at menarche ANM=Age at natural menopause | | | | | |
| **Study design** | **Conflicts of interest** | | **Inclusion criteria** | | **Exclusion criteria** |
| Cross-sectional study. | "Conflict of interest: None declared." Page 1704. | | Primary: “A total of 161 808 women aged 50 – 79 years old were recruited from 40 US clinical centers between 1993 and 1998 (The Women’s Health Initiative Study Group, 1998). A subset of women (n∼24000) from the WHI clinical trials and observational study were selected for genotyping in PAGE and are included in these analyses.” Page 1697.  Secondary: “Participants eligible for the current analysis were females who provided blood samples for nested case–control studies of breast, colorectal, ovarian, endometrial or other cancers for genetic association and biomarker studies in PAGE (n=7216).” Page 1697. | | “In studies with continuous data for AM, women reporting extreme values (AM <8 years or >19 years) were excluded from the analyses. Women with hysterectomy before onset of natural menopause or who reported menopause onset as a result of surgery (bilateral oophorectomy), radiation treatment or chemotherapy and women reporting ANM <40 years or >60 years were excluded from these analyses. In addition, in WHI and CHS, women who reported use of hormone re- placement before the onset of natural menopause were also excluded.” Page 1697. |
| **Population and sampling description** | | **Indigenous description** | | **Total number of participants** | |
| The population of interest for this study was “European American (EA), African American (AA), Hispanic, American Indian (Am Ind) and Native Hawaiian women from ARIC, CARDIA, CHS, SHS, EAGLE (NHANES III only), MEC and WHI with available genotype and AM or ANM data and who consented to use of genetic data.” Page 1697.  No details were provided about the sampling strategy. However, samples were drawn from the following studies: ARIC recruited predominantly EA and AA individuals from four communities— Forsyth County, NC; Jackson, MI; suburban areas of Minneapolis, MN and Washington County, MD. CARDIA—AA and EA adults were recruited from four US cities (Birmingham, AL; Chicago, IL; Minneapolis, MN; and Oakland, CA) CHS—recruited at four field centers (Forsyth County, NC; Sacramento County, CA; Washington County, MD; Pittsburgh, PA) EAGLE—population-based, cross-sectional survey of American MEC—Hawaii and California SHS—participating communities include seven tribes in south- western Oklahoma (Apache, Caddo, Comanche, Delaware, Fort Sill Apache, Kiowa and Wichita), three tribes in Arizona (Gila River and Salt River Pima/Maricopa and Akchin Pima/Papago) and three tribes from South/North Dakota (Oglala Sioux, Cheyenne River Sioux and Spirit Lake Communities WHI—recruited from 40 US clinical centers. Page 1697. | | “Participating communities include seven tribes in south-western Oklahoma (Apache, Caddo, Comanche, Delaware, Fort Sill Apache, Kiowa and Wichita), three tribes in Arizona (Gila River and Salt River Pima/Maricopa and Akchin Pima/Papago) and three tribes from South/North Dakota (Oglala Sioux, Cheyenne River Sioux and Spirit Lake Communities).” Page 1697.  All studies collected self-identified race/ethnicity information by questionnaire.  The percentage of the sample who were Indigenous was 0.93. Page 1698. | | The total number of participants was 25024 of which 233 were American Indian. | |
| **Study outcomes and important results** | | | | | |
| “Effect sizes for this SNP were larger, but also not significant in Native Hawaiians (P=0.02) and Am Ind (P =0.15)” Page 1699 SNP=single nucleotide polymorphisms.  Age at natural menopause: “Between-study heterogeneity (I^2^=74) was also seen for the smaller Am Ind sample, though the study-specific results were not significant.” Page 1699.  “The most robust associations were found for rs16991615 (20p12.3/ MCM8) in which we replicated the association in EA (P=8E-15), as well as observed significant associations in Hispanics (P=0.0004) and Am Ind (P=0.001). Notably, in the Am Ind population, each additional copy of the A allele was associated with a 2.6±0.8 year increase in menopause age.” Page 1699 EA=European American.  “Findings for these SNPs in AA, Hispanics and Am Ind were null, in spite of reasonable power to detect the reported association in AA (>90%) and Hispanics (>71%).” Page 1700. | | | | | |
| **Use of STRAW+10** | **Menopausal transition staging of sample using STRAW+10** | | | | |
| The STRAW+10 not used. | Postmenopause.  “ANM was defined as the age at cessation of regular menstrual periods. Women with hysterectomy before onset of natural menopause or who reported menopause onset as a result of surgery (bilateral oophorectomy), radiation treatment or chemotherapy and women reporting ANM <40 years or >60 years were excluded from these analyses. In addition, in WHI and CHS, women who reported use of hormone replacement before the onset of natural menopause were also excluded.” Page 1697 ANM=Age at natural menopause. | | | | |
| **Author reported facilitators to treatment** | | | **Author reported barriers to treatment** | | |
| N/A. | | | N/A. | | |
| **Study limitations** | | | | | |
| “Some limitations of our study deserve mention. We relied on self-report and recall of ages at menarche and menopause, requiring women to report events that in some cases occurred many years ago. However, recall of AM or age at menopause (natural or induced) has been shown to be fairly reproducible in mainly European descent populations with high overall validity (+ 1 year) compared with recorded age at the time of occurrence (Bean et al., 1979; Colditz et al., 1987; den Tonkelaar, 1997). Importantly, we would not expect any potential recall bias to be differential with respect to the studied genotypes. Our study is unique in describing genetic associations in large numbers of women differing by race/ethnicity. Yet, we acknowledge that some heterogeneity within our specified race/ ethnicity groups may exist. We also observed moderate-to-high between-study heterogeneity for a few SNPs; in particular, the ANM analyses of rs365132 showed significant between-study heterogeneity in EA and Am Ind, indicating that the meta-analysis effect size is not a good summary of the study-specific estimates. In EA, the SNP was significantly associated with ANM in MEC and WHI, but not in ARIC or CHS. Reasons for this heterogeneity are not clear; the SNP remained significant regardless of cancer status in WHI and MEC (in sensitivity analyses). In Am Ind, coded allele frequencies notably differed between WHI and SHS, as did effect sizes. However, the WHI sample size is small and although the effect size in SHS was comparable to the other groups, the SNP did not replicate in either study and thus the interpretation (null result) remains the same. A final limitation relates to study power to replicate previously reported findings in the non-European populations. While power for EA was excellent (.99% for all SNPs), we had low power to replicate findings in Am Ind or Native Hawaiians (power for all SNPs was <60%). Power for AA ranged from very high for the AM LIN28B SNPs (.90%) to modest for other SNPs, ranging from 56 to 83%. Hispanics had modest power for AM SNPs (60– 82%) and more variable power for ANM SNPs (19 –86%). Similarly, power for Asians varied highly by SNP (8– 87%) due to varying allele frequencies and sample sizes. Overall, low power for some of the SNP-race/ethnicity group analyses may have limited our ability to replicate previously published GWAS findings, which also may be inflated due to ascertainment bias, termed the winner’s curse.” Pages 1701 through 1703. | | | | | |

| **Record ID** | **Title** | | | | |
| --- | --- | --- | --- | --- | --- |
| #229 – 2016 | American Indian botanicals as possible alternatives to hormone therapy during menopause | | | | |
| **Author information** | | | | | **Country** |
| Burton, T.; Dunlap, T.; Dong, H.; Li, G.; Bolton, J.; Soejarto, D.; & Van Breemen, R. B. | | | | | United States |
| **Extraction notes** | | | | | |
| This is a conference abstract. | | | | | |
| **Study aims** | | | | | |
| To evaluate American Indian plants for potential women’s health-related benefits. “There are still numerous American Indian plants that lack scientific studies on their safety and efficacy. In collaboration with the Chicago Botanic Garden, 15 native plants were evaluated for potential women’s health-related benefits. Amorpha canescens Pursh. (Fabaceae) – Leadplant, Echinocystis lobata (Michx.) Torr. & A. Gray (Cucurbitaceae) – wild cucumber, and Silphium perfoliatum L. (Asteraceae) - cup plant inflorescent tissue were screened for estrogenicity, chemopreventive, and anti-inflammatory potential based on American Indian ethnobotany and published biological data.” | | | | | |
| **Study design** | **Conflicts of interest** | | **Inclusion criteria** | | **Exclusion criteria** |
| Other: phytochemical assessment. | N/A; none listed. | | N/A; none listed. | | N/A; none listed. |
| **Population and sampling description** | | **Indigenous description** | | **Total number of participants** | |
| N/A; these are plants: Amorpha canescens Pursh. (Fabaceae) – Leadplant, Echinocystis lobata (Michx.) Torr. & A. Gray (Cucurbitaceae) – wild cucumber, and Silphium perfoliatum L. (Asteraceae) - cup plant. | | N/A; “American Indian plants” are the focus of this study. | | N/A; this assessment focused on 15 native plants. | |
| **Study outcomes and important results** | | | | | |
| “Leadplant MeOH extract was selected as the best candidate for bioassay-guided fractionation due to dose-dependent (40, 20, 10, 5, 2.5, 1.25 µg/mL) anti-estrogenic and anti-inflammatory activity in the Ishikawa and Griess assays (n = 9). However, leadplant extract (20 µg/mL) demonstrated low activity in the NQO1 induction assay for chemopreventive potential (n = 6).” | | | | | |
| **Use of STRAW+10** | **Menopausal transition staging of sample using STRAW+10** | | | | |
| The STRAW+10 not used. | N/A. | | | | |
| **Author reported facilitators to treatment** | | | **Author reported barriers to treatment** | | |
| N/A. | | | N/A. | | |
| **Study limitations** | | | | | |
| N/A; none described. | | | | | |

| **Record ID** | **Title** | | | | |
| --- | --- | --- | --- | --- | --- |
| #21 – 2006 | Women’s health among the Chumash | | | | |
| **Author information** | | | | | **Country** |
| Adams, J. D., Jr.; & Garcia, C. | | | | | United States |
| **Study aims** | | | | | |
| “By providing the specific uses of plants for specific diseases and discussing chemistry, efficacy and safety concerns for each plant, we hope that this article gives direction to women seeking to use plants in their health care.” Abstract  “This is not an anthropological study of plant use since such studies frequently fail to humanize the people who use the plants, by inference making their health care practices seem foreign and unusual. In contrast, we wish to show the relevance of these traditional uses for the health of women today. During the course of writing this article and other works, the authors have been surprised at the paucity of information on many medically important California plants. It is our hope that along with introducing plants into women’s health care, this study will be a catalyst to more basic studies to elucidate the mechanisms of these fascinating plants.” Page 130. | | | | | |
| **Study design** | **Conflicts of interest** | | **Inclusion criteria** | | **Exclusion criteria** |
| Other: narrative review. | N/A; none listed. | | N/A; none listed. | | N/A; none listed. |
| **Population and sampling description** | | **Indigenous description** | | **Total number of participants** | |
| N/A; there were no participants, plants and herbs were described. | | Chumash people from California. | | N/A. | |
| **Study outcomes and important results** | | | | | |
| Atemisia douglasiana used in menopause, dysmenorrhea and for sleep. Page 126.  “Chumash women relied on, and still rely on, mugwort tea to treat hot flashes and other menopausal symptoms (2). The tea is described under premenstrual syndrome. Women drank the tea as needed. Their husbands were also encouraged to drink the tea, since menopause was a time that was difficult for them as well. It is a pleasant, soothing tea.” Page 128. | | | | | |
| **Use of STRAW+10** | **Menopausal transition staging of sample using STRAW+10** | | | | |
| The STRAW+10 not used. | N/A. | | | | |
| **Author reported facilitators to treatment** | | | **Author reported barriers to treatment** | | |
| N/A. | | | N/A. | | |
| **Study limitations** | | | | | |
| N/A; none described. | | | | | |

| **Record ID** | **Title** | | | | |
| --- | --- | --- | --- | --- | --- |
| #5 – 2013 | Mature women’s health | | | | |
| **Author information** | | | | | **Country** |
| None listed. | | | | | Canada |
| **Extraction note** | | | | | |
| This record has been retired. The focus of our extraction is on the analysis provided by the author of this summary statement. | | | | | |
| **Study aims** | | | | | |
| N/A; none described. | | | | | |
| **Study design** | **Conflicts of interest** | | **Inclusion criteria** | | **Exclusion criteria** |
| Text and opinion. | N/A; none listed. | | N/A; none listed. | | N/A. |
| **Population and sampling description** | | **Indigenous description** | | **Total number of participants** | |
| The population of interest for this study was First Nations, Inuit, and Métis women.  Sampling is N/A to this record. | | Aboriginal women of Canada: First Nations, Inuit, and Métis in general.  These are not further defined. | | N/A. | |
| **Study outcomes and important results** | | | | | |
| Health professionals should be aware that there is a great lack of research, resources, and programming about mature women’s health issues, including menopause, that is specific to First Nations, Inuit, and Métis. Health professionals should advocate for further research in this area. | | | | | |
| **Use of STRAW+10** | **Menopausal transition staging of sample using STRAW+10** | | | | |
| The STRAW+10 not used. | N/A. | | | | |
| **Author reported facilitators to treatment** | | | **Author reported barriers to treatment** | | |
| N/A. | | | There are limited resources available on this topic, such as Pauktuutit’s guide for mature Inuit women called As Time Goes On, which offers some suggestions on the importance of diet and nutrition for mature women’s health. | | |
| **Study limitations** | | | | | |
| N/A; none described. | | | | | |

| **Record ID** | **Title** | | | | |
| --- | --- | --- | --- | --- | --- |
| #3619 – 2023 | Cannabis use for menopause in women aged 35 and over: A cross-sectional survey on usage patterns and perceptions in Alberta, Canada | | | | |
| **Author information** | | | | | **Country** |
| Babyn, K.; Ross, S.; Makowsky, M.; Kiang, T.; & Yuksel, N. | | | | | Canada |
| **Extraction note** | | | | | |
| **Study aims** | | | | | |
| “The objective of this study was to examine the pattern of use and perceptions about cannabis for menopause in women aged 35 and over in Alberta, Canada.” Abstract.  “The objective of the current study was to characterise the use patterns and perceptions of cannabis specifically for menopause in a population of women aged 35 and over.” Page 2. | | | | | |
| **Study design** | **Conflicts of interest** | | **Inclusion criteria** | | **Exclusion criteria** |
| Cross-sectional study. | “Competing interests NY has been on the advisory board and/or speaker for Biosyent, Bayer, Amgen, Organon and Duchesnay. MM: none to disclose. TK: none to disclose. KB: none to disclose. SR: none to disclose.” Page 11. | | “The inclusion criteria were women, ages 35 years and over, living in Alberta and able to complete an online survey in English.” Page 2. | | “If a participant did not meet all three criteria for inclusion during screening, they were excluded.” Page 2. |
| **Population and sampling description** | | **Indigenous description** | | **Total number of participants** | |
| The population of interest for this study was: “woman, ages 35 and over, living in Alberta, Canada.” Abstract.  “Participants Self-selected sample of women recruited through social media (Facebook, Instagram, Twitter) between October and December 2020. Inclusion criteria included: identified as woman, ages 35 and over, living in Alberta, Canada.” Abstract.  “The inclusion criteria were women, ages 35 years and over, living in Alberta and able to complete an online survey in English. Women were recruited through research-specific social media pages on Facebook, Instagram and Twitter. Targeted advertisement campaigns or user-shared public social media posts were used to advertise the survey link and offer women the opportunity to participate.” Page 2. | | “Indigenous” women of Canada.  This are not further defined.  The percentage of the sample who were Indigenous was 3.6%. | | The total number of participants is 1485 and the total number of Indigenous participants is 54. | |
| **Study outcomes and important results** | | | | | |
| From Table 1: of the Indigenous portion of the sample (total = 54), 26 (5.2% of the sample who were currently using cannabis) were current users of Cannabis for menopause symptoms and 28 (2.9% of the sample wo were not currently using cannabis) were not current users.  “Women who reported current cannabis use were more likely to report experiencing menopause symptoms compared with non-users, specifically sleep difficulties, mood symptoms, muscle/joint achiness and urogenital symptoms. Our data add to the evidence indicating midlife women’s use of cannabis medically, as well as emphasise the high level of interest on cannabis through the large number of responses in our survey.” Page 8. | | | | | |
| **Use of STRAW+10** | **Menopausal transition staging of sample using STRAW+10** | | | | |
| The STRAW+10 not used. | Reproductive, Menopause transition, Postmenopause.  “Menopause stage was categorised through response to predefined descriptions of changes in menstrual cycle with or without presence of menopause symptoms, adapted from Marlatt et al. 16 Women were categorised as: ‘premenopause’ if periods were regular with no menopausal symptoms present; ‘perimenopause’ if periods were irregular or regular with presence of menopausal symptoms ‘postmenopause’ if it had been more than 12 months since last menstrual period, regardless of menopause symptoms being present. 16 If menopause stage could not be determined, then women were categorised into ‘other’.” Page 2. | | | | |
| **Author reported facilitators to treatment** | | | **Author reported barriers to treatment** | | |
| N/A. | | | N/A. | | |
| **Study limitations** | | | | | |
| “Conducting self-reported surveys carry risks of information biases, such as prevarication and notoriety biases, or responses influenced by social desirability.” Page 1.  “This study has several limitations to be considered when interpreting findings. First, our recruitment strategy included women who had access to the internet and were able to complete the survey in English, which may limit the generalisability to all women aged 35 or over. It is difficult to estimate our population for the survey given that this was an unrestricted and self-selected survey with women who had access to social media platforms. This was an exploratory study designed to inform further research, including the next qualitative phase of this mixed-methods study. These findings are not representative of the full population of women aged 35 and over, other than the women who completed the survey. Online data collection may impact complete representativeness of the population, however, the large number of responses collected in the short time frame suggests our survey method was effective to engage women in study participation. As well, our sample of self-selected women addressed the lack of evidence available on cannabis for self-management of symptoms in this population to justify future clinical investigations. Second, our survey cohort was highly educated and there was an underrepresentation of ethnic or racial minorities. To compare, 88% of women in our survey had some level of postsecondary education vs 65% of women in Alberta per census data. 47 Third, this study relied on self-reported data to categorise menopause stages and symptoms. All women were asked to report presence of menopause symptoms, which may have inadvertently introduced confounders and self-reporting bias. Other limitations, common to survey design include information bias, prevarication bias, notoriety bias, and social desirability which may have influenced how women may have responded to questions around cannabis or menopause. Fourth, it was beyond the scope of the study to objectively measure the positive or negative effects of cannabis use on individual symptom control, nor can this data establish causality. While we asked past users if side effects were the reason for stopping cannabis use, we did not explore perceived cannabis related adverse effects in current users. Fifth, the term cannabis in our study was defined according to Canadian regulations. 11 We recognise that cannabis definitions may differ depending on country-specific regulations. Finally, as medical cannabis is not currently covered by most Canadian healthcare plans, the population who use cannabis will be women who can afford it. Affordability of cannabis, including out of pocket expenses, to establish associations with demographic characteristics was beyond the scope of this survey. Rather, we developed a cross-sectional survey to identify cannabis usage patterns and provide insight on associations for further hypothesis generating and clinical investigations.” Pages 9 and 10. | | | | | |

| **Record ID** | **Title** | | | | |
| --- | --- | --- | --- | --- | --- |
| #3704 – 2024 | Treatment satisfaction, unmet needs, and new treatment expectations for vasomotor symptoms due to menopause: Women's and physicians' opinions | | | | |
| **Author information** | | | | | **Country** |
| DePree, B. J.; Shiozawa, A.; Kim, J.; Wang, Y.; Yang, H.; & Mancuso, S. | | | | | United States |
| **Extraction note** | | | | | |
| **Study aims** | | | | | |
| “To assess treatment satisfaction, unmet treatment needs, and new vasomotor symptom (VMS) treatment expectations among women with moderate to severe VMS and physicians treating women with VMS.” Abstract.  “This study aimed to assess VMS treatment satisfaction, perceived unmet treatment needs, and expectations for new VMS treatments in women with moderate to severe VMS due to menopause and VMS-treating physicians in the United States.” Pages 770 and 771. | | | | | |
| **Study design** | **Conflicts of interest** | | **Inclusion criteria** | | **Exclusion criteria** |
| Cross-sectional study. | “Financial disclosures/conflicts of interest: B. DePree serves on an advisory council for Astellas Pharma, Inc. A. Shiozawa, J. Kim, and S. Mancuso are employees of Astellas Pharma, Inc. Y. Wang and H. Yang are employees of Analysis Group, Inc., which received funding from Astellas Pharma, Inc., for the current study.” | | “Women eligible for inclusion in this analysis were perimenopausal or postmenopausal, aged 40 to 65 years, who experienced moderate to severe VMS in the past 12 months, received at least one treatment for VMS in the past 3 months (including  HT, non-HT, and OTC treatments), lived in the United States, spoke English, and were willing to provide consent to participate in the study. Eligible women also fell into one of the following two categories: (1) women aged >40 years with a uterus and  at least one ovary experiencing onset of intermenstrual cycle irregularities of ±7 days and other menopause-related symptoms for at least 2 months or (2) women with a uterus and at least one ovary and permanent cessation of menstruation or women  who had bilateral oophorectomy or hysterectomy with bilateral oophorectomy.” Page 771.  “Physicians eligible for inclusion in this analysis were OB-GYNs or PCPs, were licensed and actively practiced medicine in the United States, had treated at least 15 women with VMS in the past 3 months, had prescribed treatment for women with VMS in the past 3 months, spoke English, and were willing to provide consent to participate in the study.” Page 771. | | “Women were excluded from this analysis if they experienced mild VMS, had a hysterectomy without oophorectomy or with unilateral oophorectomy, had an endometrial ablation, used a hormonal intrauterine device at the time of the study, experienced VMS induced by medications (eg, endocrine therapy or chemotherapy for breast cancer), or were participating in a clinical trial for a VMS treatment or had participated in a clinical trial for a VMS treatment in the past 5 years.” Page 771.  “There were no exclusion criteria for physicians.” Page 771. |
| **Population and sampling description** | | **Indigenous description** | | **Total number of participants** | |
| The population of interest for this study was women with moderate and severe vasomotor symptoms.  “Survey participants were recruited from nationally representative women and physician panels. Dynata, a global market research firm with over 5 million profiled healthcare panelists in the United States,  helped recruit participants for the online survey questionnaires.” Page 771.  “Comparative participant groups were defined. Women were divided equally into two cohorts, those with moderate VMS and those with severe VMS, classified by self-evaluations of overall severity of hot flashes and night sweats during the past 12 months using the first item of the Menopause Rating Scale (hot flashes, sweating/episodes of sweating; 0 = none, 1 = mild, 2 = moderate, 3 = severe, 4 = extremely severe).30 Given the small number of women who were expected to report extremely severe VMS, the categories for severe and extremely severe were combined. Therefore, women who considered themselves to have extremely severe hot flashes and night sweats were included in the severe VMS cohort. Physicians were classified based on their specialties into two cohorts: OB-GYNs and PCPs.” Page 771. | | “Native-American”  This is not further defined.  The percent of the sample who were Indigenous is 2.7%. | | The total number of participants is 401 and the total number of Indigenous participants is 11. | |
| **Study outcomes and important results** | | | | | |
| “Few women of color were represented ... Native American 2.7% [moderate: 2.0%; severe: 3.5%]” (From Phase II Quantitative survey, Demographics section of the Results, page 774). | | | | | |
| **Use of STRAW+10** | **Menopausal transition staging of sample using STRAW+10** | | | | |
| The STRAW+10 not used. | Menopausal transition and Postmenopause. | | | | |
| **Author reported facilitators to treatment** | | | **Author reported barriers to treatment** | | |
| N/A. | | | N/A. | | |
| **Study limitations** | | | | | |
| “This analysis has a few limitations. First, the use of an online panel-based survey may present issues with generalizability. The panels of women and physicians may not be representative of the general population of women with moderate to severe VMS or of physicians that treat VMS. Second, selection bias may be present among the women or physicians who were willing or able to participate in the online panels owing to several factors, including Internet access, level of well-being or severity of illness, or time constraints. Third, this study relies on self-reported measures of VMS and its severity, which may be subject to recall bias for women with longer duration of disease or who have discontinued treatment for longer periods of time, and a reluctance to share information about sensitive topics. Additionally, the inclusion bias in this noninterventional online survey of women and physicians represents another limitation due to the convenience sampling method used. Another limitation of our study lies in the exclusive focus on physicians as providers; multidisciplinary teams are vital in primary care, yet our study lacks the contributions of physician assistants and nurse practitioners, who collectively represent almost 30% of all PCPs. 36,37 We did not stratify the data to include premenopausal women during the recruitment and analysis phases; our study included only women who were perimenopausal and postmenopausal, which may limit the generalizability of our findings for premenopausal populations.” Pages 778 and 779. | | | | | |

| **Record ID** | **Title** | | | | |
| --- | --- | --- | --- | --- | --- |
| #3602 – 2023 | Variation in hysterectomy prevalence and trends among U.S. States and Territories-Behavioral Risk Factor Surveillance System, 2012-2020 | | | | |
| **Author information** | | | | | **Country** |
| Gopalani, S. V.; Dasari, S. R.; Adam, E. E.; Thompson, T. D.; White, M. C.; & Saraiya, M. | | | | | United States |
| **Extraction note** | | | | | |
| **Study aims** | | | | | |
| “We estimated up-to-date state- and territory-level hysterectomy prevalence and trends, which can help correct the population at risk denominator and calculate more accurate uterine and cervical cancer rates.” Abstract.  “Due to the limited research on population-level hysterectomy trends and the implications for accurate cancer incidence rates, we provide up-to-date hysterectomy prevalence and patterns for U.S. states, the District of Columbia, Puerto Rico, and Guam.” Page 830. | | | | | |
| **Study design** | **Conflicts of interest** | | **Inclusion criteria** | | **Exclusion criteria** |
| Cross-sectional study. | “Conflict of interest The authors did not report any conflicts of interest.” Page 834. | | Not explicitly addressed: “The prevalence of respondents who reported a hysterectomy (hysterectomy prevalence) was self-reported and defined as ‘yes’ to the following question: ‘Have you had a hysterectomy?’ Approximately 8.9% (n=113,118) of respondents who answered ‘don’t know/not sure’ or ‘refused’ or were missing for the hysterectomy status question were excluded from the analyses.” Page 830. | | Not explicitly addressed: “Respondents who reported being pregnant at the time of survey administration were recoded as not having a hysterectomy.” Page 830. |
| **Population and sampling description** | | **Indigenous description** | | **Total number of participants** | |
| The population of interest for this study was US women who had had a hysterectomy.  “Data from the Behavioral Risk Factor Surveillance System (BRFSS) were analyzed for 1,267,013 respondents aged 18 years or older in even-numbered years from 2012 to 2020, representing 127,885,148 women. Based on previous work [15], we examined data from BRFSS because it included the U.S. territories and had a larger sample size (as compared to the National Health Interview Survey) to better support analyses of hysterectomy estimates for smaller subpopulations. BRFSS is a system of annual landline and cellular telephone surveys of the noninstitutionalized U.S. adult population that collects data at the state and local level. The survey uses random digit dialing techniques on both landline telephones and cell phones to conduct the interviews. BRFSS implemented methodological changes in 2011 involving the addition of cell phone interviews and a new weighting method. Consistent with BRFSS guidance, we did not compare pre-2012 data with post-redesign data [16]. The BRFSS question on hysterectomy was part of the measure of cervical cancer screening in the core component and was asked by all jurisdictions in even-numbered years from 2012 to 2020. In 2016, the question on hysterectomy status was not asked in eight states, and these data were suppressed in the public-use data set and not included in our analyses. The median survey response rates were 45.2% (2012), 47.0% (2014), 47.1% (2016), 49.9% (2018), and 47.9% (2020). Information about the survey design, sampling, instrument, and data collection methods are available elsewhere [17].” Page 830. | | “NH American Indian or Alaska Native  (AI/AN)”; NH = non-Hispanic.  This is not further defined.  The percent of the sample who were Indigenous is 21.1%. | | The total number of participants is 333,362 and the total number of Indigenous participants is 4,510. | |
| **Study outcomes and important results** | | | | | |
| “The prevalence of hysterectomy varied by race and ethnicity, with the highest estimates reported among women who were NH Black (21.3%) and NH AI/AN (21.1%).” Page 832.  “Despite a reduction in hysterectomy prevalence, estimates remained higher among Black and AI/AN women. Racial inequities in hysterectomy are often attributed to higher rates of non-cancerous gynecologic conditions [10, 24] and poorer access to quality gynecologic care and alternative treatments [25]. Hysterectomy as a method of forced surgical sterilization disproportionately impacted Black, Native American, and Hispanic women [26, 27].” Page 832.  “Hysterectomy prevalence among NH Black and NH AI/AN women was highest in the South region, especially in the East South Central and West South Central regions (Supplementary File 2). These findings highlight the importance of understanding racial patterns in hysterectomy by geography” Page 832.  “The findings from this study indicate that overall hysterectomy prevalence declined slightly in recent years but remained high among NH Black and NH AI/AN women and women from the South, especially in the East and West South Central divisions.” Page 834. | | | | | |
| **Use of STRAW+10** | **Menopausal transition staging of sample using STRAW+10** | | | | |
| The STRAW+10 not used. | Postmenopause. | | | | |
| **Rural vs. urban differences in outcomes** | | | | | |
| “Prevalence was also higher among women who were ... from the South (21.1%).” AND “Our findings reveal large variations in hysterectomy prevalence within and between each of the four census regions and by race and other sociodemographic characteristics, underscoring the importance of adjusting epidemiologic measures of uterine and cervical cancers for hysterectomy status.” Abstract. | | | | | |
| **Author reported facilitators to treatment** | | | **Author reported barriers to treatment** | | |
| N/A. | | | N/A. | | |
| **Study limitations** | | | | | |
| “The findings in this report are subject to several limitations. Hysterectomy status was self-reported; however, previous studies have found high agreement of self-reported hysterectomy history with clinical confirmation [10] and administrative records [30]. Also, nonresponse bias could stem from systematic differences between respondents and nonrespondents. To minimize the potential impact of nonresponse, BRFSS implemented raking as a weighting methodology to adjust for nonresponse [16]. Furthermore, the 95% CIs of state-and territory-specific estimates were wide due to limited sample sizes. Finally, estimates by hysterectomy indication (non-cancerous or cancerous) were not ascertained in the survey.” Pages 833 and 834. | | | | | |

| **Record ID** | **Title** | | | | |
| --- | --- | --- | --- | --- | --- |
| #3526 – 2024 | The association of race, ethnicity, and socioeconomic status on the severity of menopause symptoms: a study of 68,864 women | | | | |
| **Author information** | | | | | **Country** |
| Kochersberger, A.; Coakley, A.; Millheiser, L.; Morris, J. R.; Manneh, C.; Jackson, A.; Garrison, J. L.; & Hariton, E. | | | | | United States |
| **Extraction note** | | | | | |
| All aORs were reduced and p-values became less significant when SES was accounted for. Also: “The authors caution against attributing differences solely to race and ethnicity and highlights the need for a more nuanced understanding of this important health issue.” Page 483. | | | | | |
| **Study aims** | | | | | |
| “This study aimed to evaluate if and how race, ethnicity, and socioeconomic status (SES) are associated with the severity of menopause symptoms in a large, diverse sample of women.” Abstract.  “The objective of the current study is to evaluate if and how race, ethnicity, and socioeconomic status are associated with the severity of menopause symptoms in a large, diverse sample of women, expanding on prior research by including granularity in self-reported racial and ethnic groups, and evaluating socio-economic status as a covariate. The aim is to shed light on the complex role that these factors play in menopausal symptomatology to better inform treatment interventions to improve the quality of life of menopausal women.” Page 477. | | | | | |
| **Study design** | **Conflicts of interest** | | **Inclusion criteria** | | **Exclusion criteria** |
| Cross-sectional study. | “Financial disclosure/conflicts of interest: At the time of this study, Dr Leah Millheiser was chief medical officer of Evernow. Dr Eduardo Hariton is a paid medical advisor of Evernow. Dr Hariton holds other relationships with entities that are unrelated to the topic of this study and center around his work with startups in the fertility space. These entities include medical advisor positions at Alife Inc, Rescripted, Evernow, Fertilidad Integral, Oya Care, Cercle AI, Levy Health, and Ro/Modern Fertility. Dr Eduardo Hariton also hold board positions at Granata Bio and Azul Fertility. Alicia Jackson, PhD, is founder and CEO of Evernow. Jennifer L. Garrison, PhD, is chief science advisor of Evernow. Dr Jerrine R. Morris received consulting fees from Evernow in the past. Evernow is an online telehealth platform providing menopause treatment.” | | “Only participants living in the United States were eligible  to participate.” AND “The survey was administered to prospective participants who self-identified as experiencing one or more symptoms common to perimenopause or menopause.” Page 477. | | “Participants who did not experience a given symptom were removed from severity analysis for that symptom. … . Outliers for age (>70) and BMI (<15 and≥60 kg/m2) were excluded from analysis. In addition, those who reported a history of a hysterectomy but oophorectomy status was unknown were excluded.” Page 477. |
| **Population and sampling description** | | **Indigenous description** | | **Total number of participants** | |
| The population of interest for this study was US women who were experiencing at least one perimenopause symptom.  “Participants signed up for care at Evernow, advertised through digital marketing and via word of mouth. Participants usually pay out of pocket for treatment, which varied by the medication recommended by the provider, as well as during the time of the study. Only participants living in the United States were eligible to participate. A total of 68,864 participants were administered a clinical intake survey from March 24, 2019, to January 13, 2023. The survey was administered to prospective participants who self-identified as experiencing one or more symptoms common to perimenopause or menopause. All Evernow prospective participants consented to having their data deidentified and aggregated for the purposes of research. The data set used is deidentified with no key to reidentify participants available to the researchers participating in the analysis; thus, an institutional review board review was not required per the University of California San Francisco Institutional Review Board policies.” Page 477. | | “Indigenous/First Nations”  The authors used self-reporting for racial/ethnic data.  The percent of the sample who were Indigenous is 0.8%. | | The total number of participants is 67,867 and the total number of Indigenous participants is 531. | |
| **Study outcomes and important results** | | | | | |
| “Notably, individuals identifying as Black (hot flashes OR, 1.91; 97.5% CI, 1.75-2.09; P < 0.001), Hispanic (skin/hair changes OR, 1.58; 97.5% CI, 1.45-1.71; P < 0.001), Indigenous/First Nations (painful sex OR, 1.39; 97.5% CI, 1.19-2.75; P = 0.007), Middle Eastern (weight changes OR, 2.22; 97.5% CI, 1.25-4.37; P = 0.01), or with two or more races/ethnicities (skin/hair changes OR, 1.41; 97.5% CI, 1.26-1.58; P < 0.001) reported higher levels of symptom severity compared with their White counterparts. Conversely, Asian and South Asian participants reported lower symptom severity. Even after incorporating SES into the linear model, racial and ethnic groups with lower SES (Black, Hispanic, Indigenous, and multiple ethnicities) exhibited slight shifts in OR while maintaining high statistical significance (Black [hot flashes OR, 1.87; 97.5% CI, 1.72-2.04; P < 0.001], Hispanic [skin/hair changes OR, 1.54; 97.5% CI, 1.42-1.68; P < 0.001], Indigenous/First Nations [painful sex OR, 1.74; 97.5% CI, 1.17-2.70; P = 0.009], multiple ethnicities [skin/hair changes OR, 1.41; 97.5% CI, 1.26-1.58; P < 0.001]).” Abstract.  “Indigenous/First Nations individuals consistently demonstrated higher prevalence rates, particularly for sleep disturbances (87.3%), skin/hair changes (83.4%), and joint/muscular discomfort (84.1%).” Page 478.  “Within this analysis, participants identifying as Black, Hispanic, Indigenous/First Nations, Middle Eastern, and two or more races/ethnicities reported greater symptom severity as compared with those identifying as White for many symptoms” Page 478.  “Indigenous/First Nations participants experienced the most severe anxiety/depression (OR, 1.70; 97.5% CI, 1.30-2.24]), painful sex (OR, 1.77; 97.5% CI, 1.19-2.75), and joint/muscular discomfort (OR, 1.72; 97.5% CI, 1.28-2.34). Middle Eastern and Indigenous/First Nations participants had the highest severity for brain fog (OR, 1.76 [97.5% CI, 1.18-2.70] and OR, 1.48 [97.5% CI, 1.17-1.88]) and fatigue/low energy (OR, 1.91 [97.5% CI, 1.12-3.55] and OR, 1.64 [97.5% CI, 1.20-2.31]).” Page 478.  From page 4, TABLE 2. Percentages of participants within each race and ethnicity category experiencing symptom presence: Symptoms experienced by Indigenous/First Nations peoples—Hot flashes = 373/531 (70.2); Night sweats = 336/531 (63.3); Sleep disturbances = 441/531 (83.1); Brain fog = 445/531 (83.8); Painful sex = 211/531 (39.7); Vaginal dryness = 343/531 (64.6); Urgent bathroom trips = 198/531 (37.3); Mood swings = 353/531 (66.5); Unusual period = 99/531 (18.6); Skin/hair changes = 366/531 (68.9); Increased facial hair/acne = 189/531 (35.6); Fatigue/low energy = 473/531 (89.0); Weight changes = 437/531 (82.3); Anxiety/depression = 364/531 (68.5); Heart discomfort = 198/531 (37.3); Joint/muscular discomfort = 364/531 (68.5).  From page 5, TABLE 3. Adjusted OR for symptom severity by race and ethnicity: Symptoms experienced by Indigenous/First Nations peoples—Hot flashes = 1.27 (1.01-1.61) p = 0.04; Night sweats = 1.37 (1.07-1.77) p = 0.01; Sleep disturbances = 1.44 (1.10-1.93) p = 0.01; Brain fog = 1.48 (1.17-1.88) p = 0.001; Painful sex = 1.77 (1.19-2.75) p = 0.007; Vaginal dryness = 1.41 (1.11-1.81) p = 0.006; Urgent bathroom trips = 1.33 (0.97-1.85) p = 0.08; Mood swings = 1.35 (1.05-1.74) p = 0.02; Unusual period = 1.30 (0.83-2.11) p = 0.27; Skin/hair changes = 1.33 (1.03-1.73) p = 0.03; Increased facial hair/acne = 1.40 (1.01-1.96) p = 0.04; Fatigue/low energy = 1.64 (1.20-2.31) p = 0.003; Weight changes = 1.73 (1.20-2.60) p = 0.005; Anxiety/depression = 1.70 (1.30-2.24) p = 0.001; Heart discomfort = 1.54 (1.16-2.06) p = 0.003; Joint/muscular discomfort = 1.72 (1.28-2.34) p = 0.001.  “Of note, our analysis showed that those identifying as Black, Hispanic, and Indigenous/First Nations were more likely to experience lower affluence” Page 480.  “For the races and ethnicities that had lower socioeconomic status (Black, Hispanic, Indigenous, and two or more ethnicities), most symptoms had lowered OR when accounting the affluence score. Overall, these shifts in OR were small. The largest OR decreases were by 0.09, for night sweats in Black (OR, 1.78; 97.5% CI, 1.63-1.95) and Hispanic individuals (OR, 1.18; 97.5% CI, 1.10-1.27),  fatigue/low energy for Indigenous/First Nations women (OR, 1.55; 97.5% CI, 1.13-2.18).” Pages 480 and 481.  “After controlling for several proxies of socioeconomic status, which themselves have some limitations, self-identified Black women, Hispanic, Indigenous, or multiracial or multiethnic groups have a higher prevalence of worse menopause symptom severity. Our findings add to a growing body of evidence highlighting the strong connection between race and ethnicity, along with a lower socioeconomic status, and the severity of menopause symptoms.” Page 481.  “Limited data exist on the menopause experiences of Indigenous women, but a systematic review revealed substantial variability in knowledge and symptom experience.18 Indigenous women often experience an earlier onset of menopause attributed to factors such as poor nutrition and a harsher lifestyle. Our study suggests that Indigenous women experience more severe menopause symptoms than White women, mirroring the symptom severity of Black and Hispanic women. Given limited access to care and increased discrimination, these findings highlight the need for further research and healthcare interventions to address the disparities in menopause symptom severity experienced by Indigenous women.” Page 481.  From page 7, TABLE 4. Adjusted OR for symptom severity by race and ethnicity including affluence score: Symptoms experienced by Indigenous/First Nations peoples—Hot flashes = 1.23 (0.98-1.56) p = 0.08; Night sweats = 1.35 (1.05-1.74) p = 0.02; Sleep disturbances = 1.40 (1.06-1.87) p = 0.02; Brain fog = 1.43 (1.14-1.82) p = 0.003; Painful sex = 1.74 (1.17-2.70) p = 0.009; Vaginal dryness = 1.38 (1.09-1.77) p = 0.01; Urgent bathroom trips = 1.30 (0.62-1.81) p = 0.12; Mood swings = 1.31 (1.02-1.69) p = 0.03; Unusual period = 1.25 (0.80-2.04) p = 0.34; Skin/hair changes = 1.28 (1.00-1.68) p = 0.06; Increased facial hair/acne = 1.36 (0.99-1.91) p = 0.06; Fatigue/low energy = 1.55 (1.13-2.18) p = 0.009; Weight changes = 1.72 (1.19-2.58) p = 0.006; Anxiety/depression = 1.63 (1.25-2.16) p = 0.001; Heart discomfort = 1.50 (1.12-2.00) p = 0.006; Joint/muscular discomfort = 1.65 (1.23-2.25) p = 0.001. | | | | | |
| **Use of STRAW+10** | **Menopausal transition staging of sample using STRAW+10** | | | | |
| The STRAW+10 not used. | Unclear. The authors state: “The survey was administered to prospective participants who self-identified as experiencing one or more symptoms common to perimenopause or menopause.” Page 477. | | | | |
| **Author reported facilitators to treatment** | | | **Author reported barriers to treatment** | | |
| N/A. | | | N/A. | | |
| **Study limitations** | | | | | |
| “Limitations include the fact that all participants were English-speaking women who sought care through the Evernow platform, so the results may not represent all individuals across different racial and ethnic groups nor should be taken to represent prevalence rates of symptomatology. Participants must have had at least one Menopause Rating Scale symptom to be included in this study, which may bias the findings and generalizability of the findings. Participants self-reported their history and symptoms, which can introduce bias. Although we report it, we did not examine symptom prevalence in depth, as the focus of this study was on symptom severity. Furthermore, the use of ZIP code data to control for SES may not accurately reflect an individual's true SES and other measures of SES that were not accounted for could also be determinants of symptom severity, such as structural racism or geographic issues. Finally, it is important to note that this study only suggests associations and does not allow for the evaluation of causal relationships, necessitating further research to fully understand the findings.” Page 483. | | | | | |

| **Record ID** | **Title** | | | | |
| --- | --- | --- | --- | --- | --- |
| #3573 – 2023 | Nehiyawak (Cree) women’s strategies for aging well: Community-based participatory research in Maskwacis, Alberta, Canada, by the Sohkitehew (Strong Heart) group | | | | |
| **Author information** | | | | | **Country** |
| Listener, L.; Ross, S.; Oster, R.; Graham, B.; Heckman, S.; & Voyageur, C. | | | | | Canada |
| **Extraction note** | | | | | |
| **Study aims** | | | | | |
| “The goals of this research were to generate information honouring Indigenous ways of knowing, and gather strengths-based knowledge about aging well, to help Maskwacîs, women maintain wellness as they age.” Abstract.  “The goals of the research project were:  • To generate information respecting and honouring Indigenous ways of knowing.  • To gather valuable strengths-based knowledge about aging well.  • To help other women in the community find their own cultural ways to attain and maintain wellness as they age: physically, mentally, emotionally, and spiritually.” Page 3. | | | | | |
| **Study design** | **Conflicts of interest** | | **Inclusion criteria** | | **Exclusion criteria** |
| Qualitative research. | “The authors declare no competing interests.” | | The authors included “all community members, men and women of all ages” who were willing to discuss “aging, being healthy and living well. Anyone wishing to join the Sharing Circles was welcome to do so, including anyone who provided transport for a participant.” Page 4. | | “We did not exclude anyone who wished to attend an event.” Page 4. |
| **Population and sampling description** | | **Indigenous description** | | **Total number of participants** | |
| The population of interest for this study were community members of the Nehiyawak community of Maskwacîs, Canada.  “Community members of the Nehiyawak community of Maskwacîs, an unincorporated community in a rural setting located approximately 85 kms south of Edmonton, Alberta and situated between the City of Wetaskiwin and Town of Ponoka in the Treaty #6 region. The community consists of four Nehiyawak First Nations who share similar traditions, culture and language: Ermineskin Cree Nation, Louis Bull Tribe, Montana First Nation and Samson Cree First Nation. Each of these Nations is responsible for their own leadership and management. Together the four Nations form the Maskwacîs Tribal Council. Pigeon Lake is a community located 50 kms from Maskwacîs and includes members from all four Nations. The combined land mass of Maskwacîs and Pigeon Lake is approximately 310 square kilometres, mainly agricultural land. According to the 2021 Statistics Canada Census Profile of the communities in Maskwacîs, the combined population including all four Maskwacîs bands was 7,728, with around 33% of the population being 0 to 14 years of age, 62% aged 15 to 64 years, and 5% aged 65 years or older. https://www12. statcan.gc.ca/census-recensement/2021/dp-pd/prof/index.cfm?Lang=E.” Page 2.  “Community members were invited to join the Sharing Circles by displaying posters widely throughout the community. For example, they were posted on community notice boards, in the band offices and in the health centres. Public service notices were also broadcasted on the local community radio station, Hawk Radio. Community meeting notices were also included in Band newsletters [21]. The posters encouraged ‘all community members, men and women of all ages’ to discuss aging, being healthy and living well. Anyone wishing to join the Sharing Circles was welcome to do so, including anyone who provided transport for a participant. We did not exclude anyone who wished to attend an event.” Page 4. | | “Four Nehiyawak First Nations who share similar traditions, culture and language: Ermineskin Cree Nation, Louis Bull Tribe, Montana First Nation and Samson Cree First Nation.”  For the purpose of their research, Indigenous was defined as members of the Nehiyawak community of Maskwacîs. “Maskwacîs members are registered with, and received healthcare from, Maskwacîs Health Services (MHS) which operates health and wellness clinics in each Nation and in Pigeon Lake.” Page 2.  The percent of the sample who were Indigenous is 100%. | | The total number of participants is 36 and the total number of Indigenous participants is 36. | |
| **Study outcomes and important results** | | | | | |
| “Further, as women age, they encounter many bodily changes associated with menopause. All of the topics mentioned above must be frankly and respectfully spoken about to ensure that individuals have the information that they need to determine whether changes they are experiencing are normal and if they need to seek medical care for any abnormalities. One participant stated  that she did not have anyone to talk to about menopause because her mother has Alzheimer’s (SC3).” Page 7. | | | | | |
| **Use of STRAW+10** | **Menopausal transition staging of sample using STRAW+10** | | | | |
| The STRAW+10 not used. | Unclear. The sample included any community members interested in discussing aging well. | | | | |
| **Author reported facilitators to treatment** | | | **Author reported barriers to treatment** | | |
| N/A. | | | N/A. | | |
| **Study limitations** | | | | | |
| “Our study has several limitations. The research was carried out in one Indigenous community, and the proposed strategies may not apply directly elsewhere, however our methods could be applied elsewhere to generate community-specific findings and output. Our Sharing Circles were carried out during the day: this obviously suited some community members, however may have limited the number and type of people who were available to contribute. We consciously decided not to collect personal information such as age because this might be considered invasive: in fact, this might have been an overly protective decision, because most individuals knew the other people joining the Sharing Circle. The presence of men at the sessions may have meant that some important topics were not discussed. To maintain confidentiality our data collection method did not attribute specific comments to individual participants. As well, topics were discussed openly, so some comments resulted from several contributions, and perhaps reflect an aspirational view of aging well rather than one can be attained by everyone.” Page 13. | | | | | |

| **Record ID** | **Title** | | | | |
| --- | --- | --- | --- | --- | --- |
| #1073 – 2000 | Gravidity and parity in postmenopausal American Indian women: The Strong Heart Study | | | | |
| **Author information** | | | | | **Country** |
| North, K. E.; MacCluer, J. W.; Cowan, L. D.; & Howard, B. V. | | | | | United States |
| **Extraction note** | | | | | |
| **Study aims** | | | | | |
| “The purpose of the present study was: (1) to examine the fertility of this large sample of American Indian women participating in the Strong Heart Study in order to determine whether fertility differs among localities; and (2) to determine which factors are associated with variation in the level of completed fertility among these populations.” Page 399. | | | | | |
| **Study design** | **Conflicts of interest** | | **Inclusion criteria** | | **Exclusion criteria** |
| Cross-sectional study. | N/A; none listed. | | Women, “ever-married”, postmenopausal.  “Ever-married was used as an indirect measure of having had the opportunity to become pregnant. Women were considered to be postmenopausal if their menstrual cycles had stopped completely for at least 12 months” Page 401. | | N/A; none listed. |
| **Population and sampling description** | | **Indigenous description** | | **Total number of participants** | |
| The population of interest for this study was “American Indian women participating in the Strong Heart Study.”  “The Dakota center includes 3 Sioux Indian tribes, representing 2 linguistic dialects, Lakota and Dakota. The Lakota are represented by 2 tribes. The first, the Oglala Sioux, live on the Pine Ridge Reservation, located to the southeast of Rapid City, South Dakota (Figure 1). Of the 8 communities of the Pine Ridge Reservation, 5 were selected to participate in the SHS, about 50% of the entire population of that tribe. Also representing the Lakota dialect are people from the Cheyenne River Sioux Reservation, located in north-central South Dakota (Figure 1). The second linguistic dialect, the Dakota, is represented by 1 tribe from the Spirit Lake Community in the Fort Totten area of North Dakota. These communities were selected on the basis of 2 criteria. First, they were considered by the tribes to be similar to the total tribal populations in life-style, employment, education, and other sociodemographic variables. Second, facilities were available to conduct clinical examinations. The Oklahoma center encompasses 7 different tribes, historically Plains and Southeastern American Indians living in the Lawton-Anadarko, Oklahoma area (Apache, Caddo, Comanche, Delaware, Fort Sill Apache, Kiowa, and Wichita tribes). The 7 Oklahoma tribes represented in the SHS live primarily in 23 counties of southwestern Oklahoma (Lee et al. 1995). Several commonalities exist among these groups (Howard et al. 1996). First, several of the tribes (Apache, Comanche, Fort Sill Apache, Kiowa, and Wichita) are Plains Indians. Although the Caddo Indians were Woodland Indians, they took on much of the culture of Plains Indians. Similarly, although the Delaware Indians originated in the Northeastern United States, they now reside with the Cherokee Nation and in close association with Plains Indians. Second, unlike the tribes in Arizona and the Dakotas, members of these 7 tribes do not live on reservations (there are no reservations in Oklahoma), but in the general community. The Arizona field center is located in Phoenix. This center has enrolled primarily Pima Indians, but there are also representatives of the closely related Maricopa and Papago (Tohono O'odham) Indian tribes (Lee et al. 1990). The Indians live in 3 communities near Phoenix: the Gila River Reservation and the Salt River Reservation, which are Pima/Maricopa Indian communities, and the Ak-Chin Reservation, which is a Papago/Pima Indian community (Howard et al. 1992). The Gila River community is the largest of the 3, and consists of 7 districts. Approximately 76% of the Arizona SHS cohort resides in the Gila  River Reservation. The Salt River Pima-Maricopa Indian Community is smaller and is located in Maricopa County, bordering several major cities (Fig-  ure 1 ). The Ak-Chin Indian community has the smallest population, numbering in total 500, of which 36 were within the cohort age range (45 to 74 years of age) at the beginning of the study (Lee et al. 1990). Their community is located on the border of the Gila River Indian Community (Figure 1).” Pages 399 to 400  “All tribal members who met the  age criteria were invited to participate in the Strong Heart Study. Response  rates were 71.8% in Arizona, 61.5% in Oklahoma, and 55.3% in the Dakotas  (62% across all centers).” Page 401. | | Indigenous peoples included in the study were from Arizona, Oklahoma, and the Dakotas. Indigenous peoples from Arizona included: Papago/Pima Indians; Maricopa and Papago (Tohono O'odham) Indians. Indigenous peoples from Oklahoma included 7 different tribes: Apache, Caddo, Comanche, Delaware, Fort Sill Apache, Kiowa, and Wichita tribes. Indigenous people from the Dakotas included: 3 Sioux Indian tribes [representing 2 linguistic dialects of Lakota (representing the 2 tribes of the Oglala Sioux and the Cheyenne River Sioux) and Dakota (representing 1 tribe from the Spirit Lake Community)]  The authors do not describe how this was defined in their study.  The percent of the sample who were Indigenous is 100%. | | The total number of participants is 1,955 and the total number of Indigenous participants is 1,955.  From Arizona there were 660 participants. From Oklahoma there were 670 participants. From the Dakotas there were 625 participants. | |
| **Study outcomes and important results** | | | | | |
| From page 402, Table 1. “Reproductive Characteristics of Ever-Married, Postmenopausal American Indian Women by Strong Heart Study Center”: “Menopausal Status” was categorized by “Arizona”, “Oklahoma”, and “Dakotas” sites, and further subcategorized by “Natural” and “Surgical”. Surgical was sub-subcategorized as “(Complete)”, “(Uterus only)”, and “(Unknown)”.  At the Arizona site, 468 participants had “Natural” menopausal status, and 192 had “Surgical” menopausal status. Of the surgical menopausal status, 98 were “(Complete)”, 36 were “(Uterus only)”, and 58 were “(Unknown)”. At the Oklahoma site, 365 participants had “Natural” menopausal status, and 305 had “Surgical” menopausal status. Of the surgical menopausal status, 179 were “(Complete)”, 96 were “(Uterus only)”, and 30 were “(Unknown)”. At the Dakotas site, 480 participants had “Natural” menopausal status, and 145 had “Surgical” menopausal status. Of the surgical menopausal status, 65 were “(Complete)”, 68 were “(Uterus only)”, and 12 were “(Unknown)”. The mean age at surgical menopause was categorized by site. The Arizona site’s mean age at surgical menopause was 40.4 ± 6.8; the Oklahoma site’s mean age at surgical menopause was 39.3 ± 7.7; the Dakotas site’s mean age at surgical menopause was 41.8 ± 5.8. The mean age at surgical menopause for the entire sample was 40.2 ± 7.1.  The mean age at natural menopause was categorized by site. The Arizona site’s mean age at natural menopause was 45.6 ± 5.2; the Oklahoma site’s mean age at natural menopause was 47.7 ± 5.4; the Dakotas site’s mean age at natural menopause was 47.4 ± 4.7. The mean age at natural menopause for the entire sample was 46.9 ± 5.2.  “Age at Menopause. The age at natural menopause varied significantly among population centers (Table 1). Post hoc comparisons suggested that women from Arizona had significantly earlier natural menopause than women from the Dakotas and Oklahoma. Indeed, women from Arizona reported a mean age of natural menopause approximately 2 years earlier than that of the other two groups. The age at surgical menopause also varied across populations (Table 3). American Indian women from the Dakotas were significantly different from women in Oklahoma, who reported the earliest mean age of surgical menopause. The prevalence of surgically induced menopause varied across population centers, with 44% of women in the Oklahoma sample having had a surgical menopause, in comparison with fewer than 30% in the Dakotas and Arizona (Table 1). The prevalence of surgical menopause by birth cohort was also examined. In the women from the Dakotas and Arizona, the rate of surgical menopause remained relatively constant (at about 30%) across birth cohorts (nonsignificant) (Figure 3). In contrast, among women from Oklahoma the rate was relatively stable (about 40%) throughout the first 3 birth cohorts, with a dramatic increase to more than 60% in the last birth cohort (X2 = 7.80, p≤0.05). Because surgical menopause was so common in these populations, the fertility of those women who had a surgical menopause was examined. There was no significant differences between women with a surgical menopause and those with a natural menopause, in gravity or parity (Table 3).” Pages 403 through 405.  “We found significant differences in the age at natural menopause between the 3 American Indian populations. However, since natural menopause was defined as amennorhea for 1 year, there may be biases in these data because a varying proportion of these populations may have prolonged periods of amennorhea due to obesity and insulin resistance. However, no correlation between age at menopause and body mass index, at the time of interview, was detected.” Page 410.  “Surgical menopause was nearly twice as prevalent among women in the Oklahoma center (46%) as in women in Arizona (29%) and the Dakotas (23%). This level of surgical menopause was high compared to an overall US average of about 20% (Ross 1983). Other studies of American Indians have reported high levels of surgical menopause (more than 30%), but only among women who had had an average of 4 births. Interestingly, for SHS American Indian women there were no significant differences between women with a natural menopause and those with a surgical menopause in either measure of fertility. In addition, the average fertility measures did not vary across menopausal status, even when each population was considered separately. Thus, the prevalence of surgical menopause could not explain variation in fertility between Strong Heart centers.” Pages 410 and 411. | | | | | |
| **Use of STRAW+10** | **Menopausal transition staging of sample using STRAW+10** | | | | |
| The STRAW+10 not used. | Postmenopause. “Women were considered to be postmenopausal if their menstrual cycles had stopped completely for at least 12 months, either because of natural or surgical processes.” Abstract. “Women whose menses had ceased were asked if their menopause were natural or had been surgically induced. If surgical, they were asked if only their uterus had been removed or if their ovaries had also been removed (oophorectomy). Women who reported hysterectomy without oophorectomy or who did not know whether their ovaries had been removed were classified as postmenopausal if their age was greater than or equal to 53, the age at which 90% of the women had had a natural menopause.” Page 401. | | | | |
| **Rural vs. urban differences in outcomes** | | | | | |
| N/A; none described for menopause.  “these data have shown a linear relationship of both education and income with fertility. The lower fertility of women from the Oklahoma center compared to that of women from the Arizona and Dakota centers may be explained, in part, by the fact that, unlike participants from Arizona and the Dakotas, Oklahoma participants do not live on reservations. The hypothesis that cultural (perhaps in concert with environmental) factors may be the basis of any observed differences in fertility rates, independent of reservation experience, should also be considered. Indeed, differences in fertility rates between centers could be due to the diversity of tribes sampled in each center. Unfortunately, the distribution of sample size and the large amount of missing data on tribal status has made it impossible to test this hypothesis. Therefore, it may be that higher fertility actually characterizes the cultural practices of the tribes in Arizona and the Dakotas, rather than the reservation experience per se.” Page 412 | | | | | |
| **Author reported facilitators to treatment** | | | **Author reported barriers to treatment** | | |
| N/A. | | | N/A. | | |
| **Study limitations** | | | | | |
| “There may be some error associated with self-reported oophorectomy, as women can be confused about the exact nature of their surgery.” Page 401. | | | | | |

| **Record ID** | **Title** | | | | |
| --- | --- | --- | --- | --- | --- |
| #3577 – 2024 | Of a “certain age” - Menopause in Native American women: A cultural perspective | | | | |
| **Author information** | | | | | **Country** |
| Schweda, D.; & Bracamonte, J. | | | | | United States |
| **Extraction note** | | | | | |
| **Study aims** | | | | | |
| N/A; this is an editorial inviting healthcare providers to understand cultural nuance and to engage in culturally sensitive menopausal healthcare with their patients, such as with Dine´ women “of a certain age.” | | | | | |
| **Study design** | **Conflicts of interest** | | **Inclusion criteria** | | **Exclusion criteria** |
| Text and opinion. | “The authors declare that they have no competing interest.” | | N/A. | | N/A. |
| **Population and sampling description** | | **Indigenous description** | | **Total number of participants** | |
| The population of interest for this editorial was Native American, Navajo/Dine´ women.  Sampling details are not applicable as this is an editorial. | | N/A; this is an editorial that focuses on Native American, Navajo/Dine´ women. | | N/A. | |
| **Study outcomes and important results** | | | | | |
| As this is a one-page editorial, the entire editorial has been included here:  “Clinicians dedicated to providing care for patients from diverse cultural backgrounds strive to ensure they give those patients comprehensive support. The cultural perspectives of these patients may significantly influence their openness to discussions regarding menopausal symptoms and potential management strategies. Growing up on and around the Navajo reservation afforded me (the lead author) the chance to interact with a unique subset of Dine´ women, colloquially referred to as those “of a certain age.” Often situated as the discerning gatekeepers of administrative desks, these matrons were known for their adept deflection of inquiries, always seemingly involving unattainable deadlines and notarized copies in triplicate. With assertiveness bordering on intimidation, they showed little sympathy for tales of broken cars, missing addresses, or lost documents. Approaching these women required caution, politeness, and thorough preparation. As a Dine´ woman and physician approaching a similar life stage, I reflect on my own behavior and attitude. The women I grew up around seldom discussed hormonal changes with aging or explored options like hormone replacement therapy. In the quest to improve communication with patients, it’s crucial for clinicians to consider cultural perspectives, understanding the depth of concern or desire for treatment. Acknowledging the cultural context is imperative for delivering comprehensive care to this population. Curiously, my grandmother, on reaching a “certain age,” became an activist spotlighting governmental injustices against the Dine´. Traveling extensively and even addressing the United Nations multiple times, she was known for having no apprehensions about expressing herself. This raises questions about whether hormonal changes empowered her outspokenness. Did she have other symptoms that negatively affected her? Would she have wished to discuss this openly in the appropriate context? Her experiences undoubtedly influenced my perceptions of a woman’s evolution through life. Menopausal experiences vary with cultural influences as well as the potential for diverse symptom presentation in a diverse population [1,3]. This certainly factors into the importance of open discussion of symptoms with primary care clinicians. Some languages lack a direct concept of menopause, hindering the connection between physical and emotional issues with this life stage [2]. Limited research on menopausal symptoms in these populations underscores the necessity for a deeper understanding of their perceptions, desires, and healthcare utilization [1]. Among a diverse population of premenopausal women, evidence indicates prevalent vasomotor symptoms, with lower reporting rates among Asian and Hispanic women compared with their white counterparts. Native Americans, constituting approximately 2 % of the population of the United States, face a noticeable dearth of studies on women’s health, especially concerning menopause [3]. Cultural nuances contribute to First Nation women avoiding discussions about menopause with healthcare providers [2]. Perceptions of menopause vary among Native American communities, each with unique traditions and beliefs. Some view menopause as a natural and respected phase symbolizing wisdom and maturity, marked by specific rituals or ceremonies. Elders may play a pivotal role in passing down traditional knowledge and offering support. However, individual perspectives within Native American communities may also be shaped by modernization, acculturation, and access to healthcare [1,2]. Conversations about health, including menopause, differ widely among individuals and communities. Recognizing the diversity within Native American communities is crucial, with due consideration given to their distinct cultures, languages, and perspectives. Approaching specific questions or topics with sensitivity and respect for cultural diversity is essential for fostering better communication and understanding [1,2]. As physicians, understanding the uniqueness of indigenous communities and their perspectives on menopause is paramount. Culturally sensitive conversations could significantly impact cardiovascular health, bone health, and psychological well-being during this transitional period [1]. Embracing these opportunities for dialogue honors the vision of those who sought a better life for their people, exemplified by individuals like my grandmother.” | | | | | |
| **Use of STRAW+10** | **Menopausal transition staging of sample using STRAW+10** | | | | |
| N/A. | N/A. | | | | |
| **Author reported facilitators to treatment** | | | **Author reported barriers to treatment** | | |
| “Approaching specific questions or topics with sensitivity and respect for cultural diversity is essential for fostering better communication and understanding” | | | N/A. | | |
| **Study limitations** | | | | | |
| N/A. | | | | | |

| **Record ID** | **Title** | | | | |
| --- | --- | --- | --- | --- | --- |
| #3538 – 2024 | Developing a menopausal transition health promotion intervention with Indigenous, integrative, and biomedical health education: A community-based approach with urban American Indian/Alaska Native women | | | | |
| **Author information** | | | | | **Country** |
| Taylor-Swanson, L.; Kent-Marvick, J.; Austin, S. D.; Ellis, J.; Charles, C.; Ward, R.; Crandall, L.; Macias, S. N.; Moreno, C.; & Simonsen, S. E. | | | | | United States |
| **Extraction note** | | | | | |
| **Study aims** | | | | | |
| “The objectives of this community-based project were to form a Community Advisory Board (CAB) with whom to: 1) conduct CAB meetings (similar to a focus group) with midlife AI/AN women to understand their lived health care experiences and needs during the menopausal transition; and 2) obtain guidance in creating a tailored intervention.” Abstract.  “Recognizing the unique experiences of AI/AN women, we wanted to adapt MENOGAP to be culturally relevant and congruent, and thus acceptable. To realize this vision, we designed a community-based participatory research (CBPR) project with the main  objective of developing a Community Advisory Board (CAB) of Indigenous midlife women with whom we conducted CAB meetings (similar to a focus group) to better understand their experiences and health care needs during the MT.” Page 2. | | | | | |
| **Study design** | **Conflicts of interest** | | **Inclusion criteria** | | **Exclusion criteria** |
| Qualitative research. | “The author(s) declared no potential conflicts of interest with respect to the research, authorship, and/or publication of this article.” | | “Eligibility criteria were 1) biologically female, 2) aged 35 or older, 3) AI/AN, and 4) living in the Wasatch Front urban corridor.” Page 3. | | N/A; none listed. |
| **Population and sampling description** | | **Indigenous description** | | **Total number of participants** | |
| The population of interest for this study were “midlife AI/AN women.”  “Recruitment for a CAB for the study was carried out via community outreach, leveraging a collaborative community-academic-clinical partnership between the University of Utah’s MENOGAP AI/AN research team and the Urban Indian Center of Salt Lake (UICSL). UICSL is a non-profit that serves AI/AN who live within Davis, Salt Lake, Tooele, Utah, Cache, and Weber counties. The UICSL provides culturally competent health care programs and services for the whole family. The services are primary health care services, behavioral health care services, preventive/health screenings, and social services. ... Potential CAB members were identified and enrolled in the study by UICSL staff. Potential participants were contacted via phone calls and emails, providing them with study information and assessing their availability to complete the consent process and receive comprehensive details regarding the research.” Pages 2 and 3. | | “American Indian/Alaska Native alone”, “AI/AN and Hispanic”  AI/AN was defined by self-identification; CAB members were receiving care at the Urban Indian Center of Salt Lake.  The percent of the sample who were Indigenous is 100% (75% AI/AN alone and 25% AI/AN and Hispanic). | | The total number of participants is 8 and the total number of Indigenous participants is 8. | |
| **Study outcomes and important results** | | | | | |
| “In the analysis of the transcripts from the initial CAB meeting with 8 women, four key themes were identified: 1) lack of and desire for information about the menopause transition; 2) barriers to accessing care; 3) matriarchal priorities and resulting health outcomes; and 4) preferences for Indigenous and integrative medicine as first-line interventions, followed by conventional medicine.” Page 5.  From pages 4 and 5, Table 2. Additional Exemplar Quotes for Each Theme. Theme 1: Lack of and Desire for Information About the Menopause Transition—“I never really was told … that menopause was a thing. … Never really talked about it at home, with, with my mom.” (CAB 1a, participant 4); “I’m going through the emotions right now. And I’ve had a lot of questions. And just like everyone else, when I was growing up, it was never talked among the females. And the women that were around me, it was never mentioned. We were never taught, like, remedies or anything. But me going through the motions and the aches and pains, the colds, the hot sweats, and everything. I am beginning to understand because I have an older friend, and she kind of like guides me through it, but I don’t know what kind of remedies to take as far as medication.” (CAB 1a, participant 1); “As far as symptoms go, everything that was on your list, I am dealing with. It’s… and just trying to figure out how to manage the symptoms is kind of one of the reasons why I agreed to do this, and have these conversations, because I’m clueless. And my mom not really big at talking about things like this. So, trying to find somewhere else to get information is key." (CAB 1c, participant 1).  Theme 2: Barriers to Accessing Care—“And the other thing I noticed, too, is when you go see these doctors, they have, like, a limited time. You know, they kind of want you to hurry up and explain your situation. They take notes and away you go. But it’s, and um… I think sometimes you have to really, you know, say hey, this is an issue, you know, I need options. You know, what should I do? What, you know, what can I do? Rather than just giving you some medication” (CAB 1a, participant 5); “My biggest problem is, I can’t get into my primary care provider on a short notice. So, I mean, that’s because she is amazing, and she’s booked out several, you know, several months. So, but I see somebody that she recommends.” (CAB 1b, participant 1); “It is, especially with fuel prices as chaotic as they are and the more this area grows, the more compact the roads become. So, dealing with traffic is kind of a deterrent. It would be amazing if they had… even if they were just little satellite facilities within, like, one in each county, or parish. That’d be so great. So yeah, the traveling down there’s been a little bit of a deterrent.” (CAB 1c, participant 1); “Recently my legs have been starting to hurt. My headaches have gotten worse. And I went to the doctor. And just like every… Just like I’ve heard in previous conversations that some of the doctors they don’t understand, or they don’t see us. Native womens coming in to, you know, with these concerns, and so they disregard a lot of it.” (CAB 1a, participant 1)  Theme 3: Matriarchal Priorities and Resulting Health Outcomes—“I can’t talk to anyone about it, anybody about how I feel, or what I’m going through, the emotions I’m going through, and I feel like this, the reason why I joined this is to find, you know, some resolution on, um, education or any kind of, uh, therapy or something that’s very helpful, um, with going through, you know, menopause and everything, and how it could be helpful to me and how I should recognize how I am important, too.” (CAB 1a, participant 1)  Theme 4: Preferences for Indigenous and integrative Medicine as First-line interventions, Followed by Conventional Medicine—“I just, I just can’t do medication at all. Without my… My body doesn’t accept any type of medication. So that wasn’t really an option for me to take any medication. So yeah, just the distance and stuff like that. I was hoping that we had somebody close by that could  be an herbalist and utilize that person and be able to bless the herb and so I can start, you know, drinking it and start feeling better.” (CAB 1a, participant 5); “I feel like a lot of times like, like there’s things that I’ve lost, um, since I’ve moved to Salt Lake City as far as, like, more traditional medicines, or, like, my mom would go out and, like, pick a few things and make me some tea and, like, and I don’t have that anymore. So, yeah, there’s definitely It would absolutely be interested in more resources.” (CAB 1b, participant 1); “I think when it is my time, I would like to go a more natural, like, way instead of medication.” (CAB 1a, participant 4); “The herbal medicine thing, I’ve actually been very interested in, but I… Like, taking the time to find someone who’s educated and knows what they’re talking about is time consuming. So, I haven’t gotten there…” (CAB 1c, participant 1)  “Theme 1: Participants lacked information about the menopause transition but were very interested in learning more about it. Despite CAB members’ experiences with symptoms, which they believed might be related to the menopause transition, they felt uncertainty about the root cause of their symptoms and how to manage their symptoms. Some felt too young to be going through the menopause transition. Most were not aware of the extensive nature of symptoms experienced during the menopause transition, and they desired information about the transition that was ‘visual’ and ‘hands-on.’ Furthermore, the menopause transition was considered ‘the unknown.’ CAB members shared that their mothers and other females in their families didn’t talk about the menopause transition when they were growing up. They believed their grandmothers and mothers weren’t aware of the menopause transition and were also not taught about this stage in life, nor about remedies for treating symptoms. A participant said: ‘…I have these symptoms, like, I’ll be getting hot and I’m like going, “I’m so young, but I feel like I’m going through menopause.” I’ve never [known] the symptoms. That’s not something my mom talked about. We barely talked about periods. … It’s just, you don’t know where to get this kind of information. I didn’t. And then, my age, I go to the doctor. Tell him. [He replies], “[At] your age?” Like, “Oh, no. You can’t be going through menopause.” I don’t know. What is menopause?’ (CAB 1a, Participant 2)  Friends were sources of menopause-transition support for CAB members. A different participant said: ‘Going back to my grandmother and my mom, they had no idea. They weren’t taught and we were not taught that we’re going to go through this menopause. It’s kind of like the unknown. So, when I was going through it, I was asking friends, “Is this normal? Do you feel like this? I feel like my heart is gonna jump out. Does that happen to you at night?” So, I was just, you know, asking around what was going on with me.’ (CAB 1a, Participant 3)  Women reported being unsure about where to get information about the menopause transition. Some said they wanted to break the trend of silence and stigma surrounding menopause for their daughters and were interested in sharing the MENOGAP class materials with them. A participant shared, ‘I have two girls. … Maybe I share a little bit too much, but I like to tell because I haven’t got those talks.’ (CAB 1a, Participant 2)” Page 5.  “Theme 2: Participants described barriers to accessing care related to the menopausal transition (MT). Several CAB members shared their belief that many doctors are not familiar with and disregard the concerns of Indigenous people. They believed there was often a cultural disconnect between their perspective of health and that of their health care providers. They felt their individual needs were rarely considered during medical visits. These concerns were discussed in the context of reluctance toward Western medicines and the emphasis on medication within the Western medical model. CAB members largely believed their doctors offered few options beyond medication (e.g., massage, mental health care, herbal remedies). One participant shared: ‘I think a lot of doctors are not really, you know, experienced [in seeing], you know, Indigenous different Tribes. So, I kind of felt uncomfortable about that because a lot of times, to me, I feel like they just give you that medication. “Here take this, take this,” and I didn’t want to do that to my body because I don’t even take an aspirin.’ (CAB 1a, Participant 3)  Participants were also concerned with the rushed nature of doctor’s visits and felt pressure to explain their health concerns quickly. A few CAB members said they felt more comfortable with female providers, who were able to relate to them as women, and less comfortable with male providers. Several said they had to speak up and advocate for themselves. For example, a participant stated: ‘I’ll go into a doctor’s office, and sometimes they’ll be surprised that I know what I know. … I’ll come with a notepad and be like, “I have these questions.” And that will surprise them and they’ll treat you with a little bit more respect because of how I tried to project myself. … Sometimes I just think, “Why do I have to … do that to get something that should be available for all of us. Why does it have to be so hard?”’ (CAB 1b, Participant 2)  One woman discussed the additional barrier of weight stigma in the health care setting, requiring her to persistently advocate for her health needs. Participants also reported that traditional Indigenous treatments were difficult to access. One CAB member said she believed IHS ‘services aren’t as successful [in the Western states of the U.S.] as they are on the east coast’ (CAB 1c, Participant 1). Additional barriers to the access of care included the cost of services, travel and cost associated with transportation to appointments, insurance issues, difficulty securing referrals to specialized care, lack of access to educational resources, inability to access provider appointments on short notice, and juggling multiple responsibilities as a mother and wife that conflicted with the prioritization of health. This last barrier is relevant to the third theme, Matriarchal priorities and resulting health outcomes, where matriarchal priorities impacted CAB members’ health outcomes.” Pages 5 and 6.  “Theme 3: Balancing multiple responsibilities, including those related to work, home, and childrearing, made it difficult for CAB members to prioritize their own health, and even to talk about their health struggles. This discussion was raised in the context of cultural matriarchy, and the necessity to be strong for the family. One participant shared her experience of being strong for her family: ‘I’m [Indigenous] as well, and in my family unit, us females have to be the strong ones and the backbone of everyone in our household. And, we do have to put a lot of what, what our, what emotions and whatever we’re going through behind on the back burner, and act like it’s not happening. And that has a lot to do with, for me, I feel like it has a lot to do with, um, how we seen our grandma, you know, how we seen our mom, as we were growing up. And so, we felt like that is what us as females in my family have to do, is put a lot of our worries, symptoms, and everything on the back burner…’ (CAB 1a, Participant 1)  Another CAB member summarized this dynamic when discussing her financial and insurance difficulties in securing needed dental care: ‘Whether feeding your kids food, or a meal on the table versus getting insurance taken out of my check, I’m going to go with the food. A mom can survive. We’re always like, my god… With Native ways, I feel like… “We tough! Shake it off. (laughs) It’s alright. Smiles.” My mom always says, “Smiles every day, body’s still the same. Mommy’s still the same.”’ (CAB 1a, Participant 2)” Page 6.  “Theme 4: A majority of the CAB members said they did not like taking Western medications, and that they preferred a more natural approach to treating symptoms of the menopause transition. One participant shared negative experiences with medications: ‘I hate taking medications. I really do not have a good history with them and I just don’t want to go that route’ (CAB 1a, Participant 2). CAB members shared their preferences for integrative medicine, which included massage, nutritional and herbal supplements, nutritionist/dietician support, acupuncture, chiropractic care, yoga, Pilates, meditation, and breathing exercises. A participant stated: ‘I’ve only had one experience with acupuncture and I don’t know. I’m not familiar enough with it. It would be interesting to try. I know massage therapy is wonderful. Chiropractic care is also wonderful. … There’s so many different forms of therapy that would be really useful … Meditation practices. … Breathing exercises. That’s like, what I turn to when my heart starts racing. It’s just trying to find that… recenter myself.’ (CAB 1c, Participant 1)  Participant preferences for Indigenous treatments included remedies made by an Indigenous herbalist, and access to a medicine man/healer. Regarding a medicine man, a participant stated: ‘Someone said that they are thinking of hiring a medicine man or healer. I think that’s a great idea. I definitely see a lot of people on the Facebook group for Natives wondering when they can get one or talk to one because it sounds like there’s, there’s not many at all here in the area. And I’d love to have access to that (CAB  1b, Participant 2).’ Another participant said: ‘I did really consider going back to Navajo land to get an herbalist to get a Navajo lady to do some herbs for me. I gave it a short time, and I says, if I don’t feel better, you know, if I’m not feeling up to myself, then I was gonna go drive down there and see if somebody would get me some herbs that I can start taking’ (CAB 1a, Participant 5).” Pages 6 and 7. | | | | | |
| **Use of STRAW+10** | **Menopausal transition staging of sample using STRAW+10** | | | | |
| The STRAW+10 not used. | Reproductive, Menopausal transition, Postmenopause. | | | | |
| **Rural vs. urban differences in outcomes** | | | | | |
| Participants described access to Indigenous treatments that were available on their reservation lands but that were difficult to access in their urban/suburban areas. | | | | | |
| **Author reported facilitators to treatment** | | | **Author reported barriers to treatment** | | |
| “A few CAB members said they felt more comfortable with female providers, who were able to relate to them as women, and less comfortable with male providers. Several said they had to speak up and advocate for themselves.” Page 6. | | | “Participants described barriers to accessing care related to the menopausal transition (MT). Several CAB members shared their belief that many doctors are not familiar with and disregard the concerns of Indigenous people. They believed there was often a cultural disconnect between their perspective of health and that of their health care providers. They felt their individual needs were rarely considered during medical visits. These concerns were discussed in the context of reluctance toward Western medicines and the emphasis on medication within the Western medical model. CAB members largely believed their doctors offered few options beyond medication (e.g., massage, mental health care, herbal remedies). ... Participants were also concerned with the rushed nature of doctor’s visits and felt pressure to explain their health concerns quickly. ... One woman discussed the additional barrier of weight stigma in the health care setting, requiring her to persistently advocate for her health needs. Participants also reported that traditional Indigenous treatments were difficult to access. One CAB member said she believed IHS ‘services aren’t as successful [in the Western states of the U.S.] as they are on the east coast’ (CAB 1c, Participant 1). Additional barriers to the access of care included the cost of services, travel and cost associated with transportation to appointments, insurance issues, difficulty securing referrals to specialized care, lack of access to educational resources, inability to access provider appointments on short notice, and juggling multiple responsibilities as a mother and wife that conflicted with the prioritization of health. Pages 5 and 6. | | |
| **Study limitations** | | | | | |
| “There are several limitations. First, the AI/AN women who participated in the discussion may be different than those who did not participate as they were willing to discuss the MT. An additional limitation is that no rural or frontier-dwelling women were  enrolled and that the sample may not be generalizable to midlife Indigenous women living outside urban areas. Furthermore, women were only sampled from one area of the Intermountain West and most women were in the menopausal transition, thus have not yet experienced post-menopause and this may limit our generalizability to post-menopause. Future nationwide surveys are needed that include a sample of Indigenous women from all areas of the United States.” Page 7. | | | | | |
| **Record #1 excluded at full-text screening** | | | | | |
| **Erratum.** J Obstet Gynaecol Can. 2022;**44**(8):938–40. | | | | | |
| **Rationale for exclusion at full-text screening** | | | | | |
| Wrong record type. (Duplicate of this record also excluded at the full-text stage). | | | | | |

**Record #2 excluded at full-text screening**

Altemus M, Levine M, Cortes Y, Reame N. Musculoskeletal pain in perimenopause: a qualitative study [abstract]. Menopause. 2013;**20**(12):1333.

**Rationale for exclusion at full-text screening**

Unclear population.

**Record #3 excluded at full-text screening**

Ambizas EM, Etzel JV, Maniara B. Managing the vasomotor symptoms of menopause. US Pharm. 2016;**41**(1):8–11.

**Rationale for exclusion at full-text screening**

Wrong population.

**Record #4 excluded at full-text screening**

Smylie J. A guide for health professionals working with Aboriginal peoples: executive summary. J Soc Obstet Gynaecol Can. 2000;(100):1–6.

**Rationale for exclusion at full-text screening**

Wrong concept.

**Record #5 excluded at full-text screening**

Committee on American Indian/Alaska Native Women’s Health; Committee on Health Care for Underserved Women. Committee opinion no. 515: health care for urban American Indian and Alaska Native women. Obstet Gynecol. 2012;**119**(1):201–5.

**Rationale for exclusion at full-text screening**

Wrong concept.

**Record #6 excluded at full-text screening**

Armitage GD, Suter E, Verhoef MJ, Bockmuehl C, Bobey M. Women’s needs for CAM information to manage menopausal symptoms. Climacteric. 2007;**10**(3):215–24.

**Rationale for exclusion at full-text screening**

Unclear population.

**Record #7 excluded at full-text screening**

Averbach S, Ha D, Meadows A, Brubaker L, Gyamfi-Bannerman C. Failure to progress: structural racism in women’s healthcare. eClinicalMedicine. 2023;**57**:101885.

**Rationale for exclusion at full-text screening**

Wrong concept.

**Record #8 excluded at full-text screening**

Avis NE, Pian-Smith MCM. Acupuncture for hot flashes. Menopause. 2007;**14**(1):10–13.

**Rationale for exclusion at full-text screening**

Wrong population.

**Record #9 excluded at full-text screening**

Bagwell-Gray ME, Jen S, Schuetz N. How intimate partner violence and intersectional identities converge to influence women’s sexual health across environmental contexts. Soc Work. 2020;65(4):349–57.

**Rationale for exclusion at full-text screening**

Wrong concept.

**Record #10 excluded at full-text screening**

Bair YA, Gold EB, Greendale GA, Sternfeld B, Adler SR, Azari R, et al. Ethnic differences in use of complementary and alternative medicine at midlife: longitudinal results from SWAN participants. Am J Public Health. 2002;**92**(11):1832–40.

**Rationale for exclusion at full-text screening**

Wrong population.

**Record #11 excluded at full-text screening**

Balestrery JE. Indigenous elder insights about conventional care services in Alaska: culturally charged spaces. J Gerontol Soc Work. 2016;**59**(4):296–315.

**Rationale for exclusion at full-text screening**

Wrong concept.

**Record #12 excluded at full-text screening**

Barinas-Mitchell E, Duan C, Brooks M, El Khoudary SR, Thurston RC, Matthews KA, et al. Cardiovascular disease risk factor burden during the menopause transition and late midlife subclinical vascular disease: does race/ethnicity matter? J Am Heart Assoc. 2020;**9**(4):e013876.

**Rationale for exclusion at full-text screening**

Wrong population.

**Record #13 excluded at full-text screening**

Baron M, Fletcher C, Riva M. Aging, health and place from the perspective of elders in an Inuit community. J Cross Cult Gerontol. 2020;**35**(2):133–53.

**Rationale for exclusion at full-text screening**

Wrong concept.

**Record #14 excluded at full-text screening**

Barrett-Connor E, Siris ES, Wehren LE, Miller PD, Abbott TA, Berger ML, et al. Osteoporosis and fracture risk in women of different ethnic groups. J Bone Miner Res. 2005;**20**(2):185–94.

**Rationale for exclusion at full-text screening**

Wrong concept.

**Record #15 excluded at full-text screening**

Baskin C, Davey CJ. Grannies, elders, and friends: aging Aboriginal women in Toronto. J Gerontol Soc Work. 2015;**58**(1):46–65.

**Rationale for exclusion at full-text screening**

Wrong concept.

**Record #16 excluded at full-text screening**

Benda W. Alternative medicine research report – brief citations. J Herb Pharmacother. 2003;**3**(4):75–6.

**Rationale for exclusion at full-text screening**

Wrong context.

**Record #17 excluded at full-text screening**

Binkley N, Fidler E, Borchardt G, Krueger D. Does vitamin D metabolism differ by race? Evaluation of vitamin D metabolites in American Indians and Caucasian Americans prior to and following vitamin D₃ supplementation [abstract]. Osteoporos Int. 2015;**26**(Suppl 1):S289–90.

**Rationale for exclusion at full-text screening**

Wrong concept.

**Record #18 excluded at full-text screening**

Bishop KC, Ford AC, Kuller JA, Dotters-Katz S. Acupuncture in obstetrics and gynecology. Obstet Gynecol Surv. 2019;**74**(4):241–51.

**Rationale for exclusion at full-text screening**

Wrong population.

**Record #19 excluded at full-text screening**

Blanken A, Gibson CJ, Li Y, Huang AJ, Byers AL, Maguen S, et al. Racial/ethnic disparities in the diagnosis and management of menopause symptoms among midlife women veterans. Menopause. 2022;**29**(7):877–82.

**Rationale for exclusion at full-text screening**

Unstratified results.

**Record #20 excluded at full-text screening**

Boneva RS, Maloney EM, Lin JM, Jones JF, Wieser F, Nater UM, et al. Gynecological history in chronic fatigue syndrome: a population-based case-control study. J Womens Health (Larchmt). 2011;**20**(1):21–8.

**Rationale for exclusion at full-text screening**

Unclear population.

**Record #21 excluded at full-text screening**

Brett KM, Keenan NL. Complementary and alternative medicine use among midlife women for reasons including menopause in the United States: 2002. Menopause. 2007;**14**(2):300–7.

**Rationale for exclusion at full-text screening**

Wrong population.

**Record #22 excluded at full-text screening**

Brooks D, Johnston S, Parker C, Cox L, Brodie M, Radbourne C, et al. Elements of long-term care that promote quality of life for Indigenous and First Nations Peoples: a mixed methods systematic review. Gerontologist. 2022. Epub ahead of print.

**Rationale for exclusion at full-text screening**

Wrong concept.

**Record #23 excluded at full-text screening**

Brooks JL, Berry DC, Currin EG, Ledford A, Knafl GJ, Fredrickson BL, et al. A community-engaged approach to investigate cardiovascular-associated inflammation among American Indian women: a research protocol. Res Nurs Health. 2019;**42**(3):165–75.

**Rationale for exclusion at full-text screening**

Wrong concept.

**Record #24 excluded at full-text screening**

Brooks-Cleator LA, Lewis JP. Alaska Native Elders’ perspectives on physical activity and successful aging. Can J Aging. 2020;**39**(2):294–304.

**Rationale for exclusion at full-text screening**

Wrong concept.

**Record #25 excluded at full-text screening**

Brown HW, Wexner SD, Segall MM, Brezoczky KL, Lukacz ES. Accidental bowel leakage in the Mature Women’s Health Study: prevalence and predictors. Int J Clin Pract. 2012;**66**(11):1101–8.

**Rationale for exclusion at full-text screening**

Wrong concept.

**Record #26 excluded at full-text screening**

Buck MM, Gottlieb LN. The meaning of time: Mohawk women at midlife. Health Care Women Int. 1991;12(1):41–50.

**Rationale for exclusion at full-text screening**

Wrong concept.

**Record #27 excluded at full-text screening**

Burton T, Dunlap T, Dong H, Li G, Bolton J, Soejarto D, et al. American Indian botanicals: possible alternatives to hormone therapy during menopause [abstract]. Planta Med. 2015;**81**(11).

**Rationale for exclusion at full-text screening**

Wrong population.

**Record #28 excluded at full-text screening**

Buxton-Blake P. Menopausal treatment dilemma: a challenge for women and nurses. Contemp Nurse. 2002;**13**(2–3):117–25.

**Rationale for exclusion at full-text screening**

Wrong population.

**Record #29 excluded at full-text screening**

Camazine S, Bye RA. A study of the medical ethnobotany of the Zuni Indians of New Mexico. J Ethnopharmacol. 1980;**2**(4):365–88.

**Rationale for exclusion at full-text screening**

Wrong concept.

**Record #30 excluded at full-text screening**

Canales MK. Connecting to nativeness: the influence of women’s American Indian identity on their health-care decisions. Can J Nurs Res. 2004;**36**(4):18–44.

**Rationale for exclusion at full-text screening**

Wrong concept.

**Record #31 excluded at full-text screening**

Carmody J, Crawford S, Churchill L. A pilot study of mindfulness-based stress reduction for hot flashes. Menopause. 2006;**13**(5):760–9.

**Rationale for exclusion at full-text screening**

Unclear population.

**Record #32 excluded at full-text screening**

Cesario SK. Care of the Native American woman: strategies for practice, education, and research. J Obstet Gynecol Neonatal Nurs. 2001;**30**(1):13–9.

**Rationale for exclusion at full-text screening**

Wrong concept.

**Record #33 excluded at full-text screening**

Chang OH, Paraiso MFR. Revitalizing research in genitourinary syndrome of menopause. Am J Obstet Gynecol. 2019;**220**(3):246.e1–4.

**Rationale for exclusion at full-text screening**

Wrong population.

**Record #34 excluded at full-text screening**

Cherrington A, Lewis CE, McCreath HE, Herman CJ, Richter DL, Byrd T, et al. Association of complementary and alternative medicine use, demographic factors, and perimenopausal symptoms in a multiethnic sample of women: the ENDOW study. Fam Community Health. 2003;**26**(1):74–83.

**Rationale for exclusion at full-text screening**

Wrong population.

**Record #35 excluded at full-text screening**

Clarke GS, Douglas EB, Campos S, House MJ, Vaughn EE, Hudgins KEG. Building connection and improving health for Indigenous elders: findings from the Title VI evaluation. J Am Geriatr Soc. 2022;**70**(5):1525–37.

**Rationale for exclusion at full-text screening**

Wrong concept.

**Record #36 excluded at full-text screening**

Cohen SM, O'Connor AM, Hart J, Merel NH, Te HS. Autoimmune hepatitis associated with the use of black cohosh: a case study. Menopause. 2004;**11**(5):575–7.

**Rationale for exclusion at full-text screening**

Unclear population.

**Record #37 excluded at full-text screening**

Cordova-Marks FM, Harris R, Teufel-Shone NI, Norton B, Mastergeorge AM, Gerald L. Characteristics of American Indian female caregivers on a Southwest American Indian reservation. J Community Health. 2019;**44**(1):52–60.

**Rationale for exclusion at full-text screening**

Wrong concept.

**Record #38 excluded at full-text screening**

Cramer EH, Jones P, Keenan NL, Thompson BL. Is naturopathy as effective as conventional therapy for treatment of menopausal symptoms? J Altern Complement Med. 2003;**9**(4):529–38.

**Rationale for exclusion at full-text screening**

Wrong population.

**Record #39 excluded at full-text screening**

Deligiannidis KM, Freeman MP. Complementary and alternative medicine for the treatment of depressive disorders in women. Psychiatr Clin North Am. 2010;**33**(2):441–63.

**Rationale for exclusion at full-text screening**

Wrong population.

**Record #40 excluded at full-text screening**

Depree B, Houghton K, Dibenedetti DB, Shiozawa A, King DD, Kim J, et al. Practice patterns and perspectives regarding treatment for symptoms of menopause: qualitative interviews with US health care providers. Menopause. 2023;**30**(2):128–35.

**Rationale for exclusion at full-text screening**

Wrong population.

**Record #41 excluded at full-text screening**

Dickson G, Green KL. Participatory action research: lessons learned with Aboriginal grandmothers. Health Care Women Int. 2001;**22**(5):471–82.

**Rationale for exclusion at full-text screening**

Wrong concept.

**Record #42 excluded at full-text screening**

Dietz NA, Mijares-Cantrell T, Acevedo D, Annane D, Rodriguez R, Caralis P, et al. Women veterans and menopause: knowledge and preferences. Women Health. 2018;**58**(8):898–914.

**Rationale for exclusion at full-text screening**

Wrong population.

**Record #43 excluded at full-text screening**

Donaldson E, Huang V, Ross S, Sydora B. Impact of inflammatory bowel diseases on menopause experience [abstract]. Can J Gastroenterol Hepatol. 2016;**2016**:167.

**Rationale for exclusion at full-text screening**

Unclear population.

**Record #43 excluded at full-text screening**

Drost JL. Developing the alliances to expand traditional Indigenous healing practices within Alberta Health Services. J Altern Complement Med. 2019;**25**(Suppl 1):S69–77.

**Rationale for exclusion at full-text screening**

Wrong concept.

**Record #44 excluded at full-text screening**

El Amil ZG, Butera JN. Black cohosh induced pancytopenia: case report [abstract]. Blood. 2011;**118**(21).

**Rationale for exclusion at full-text screening**

Unclear population.

**Record #45 excluded at full-text screening**

Elkins G, Otte J, Carpenter JS, Roberts L, Jackson LS, Kekecs Z, et al. Hypnosis intervention for sleep disturbance: determination of optimal dose and method of delivery for postmenopausal women. Int J Clin Exp Hypn. 2021;**69**(3):323–45.

**Rationale for exclusion at full-text screening**

Unclear population.

**Record #46 excluded at full-text screening**

Eni R, Phillips-Beck W, Achan GK, Lavoie JG, Kinew KA, Katz A. Decolonizing health in Canada: a Manitoba First Nation perspective. Int J Equity Health. 2021;**20**(1):206.

**Rationale for exclusion at full-text screening**

Wrong concept.

**Record #47 excluded at full-text screening**

Scoglio S, Benedetti S, Canino C, Santagni S, Rattighieri E, Chierchia E, et al. Effect of a 2-month treatment with Klamin, a Klamath algae extract, on the general well-being, antioxidant profile and oxidative status of postmenopausal women. Gynecol Endocrinol. 2009;**25**(4):235–40.

**Rationale for exclusion at full-text screening**

Wrong population.

**Record #48 excluded at full-text screening**

Evers SE, Orchard JW, Haddad RG. Bone density in postmenopausal North American Indian and Caucasian females. Hum Biol. 1985;**57**(4):719–26.

**Rationale for exclusion at full-text screening**

Wrong concept.

**Record #49 excluded at full-text screening**

Genazzani AD, Chierchia E, Lanzoni C, Santagni S, Veltri F, Ricchieri F, et al. [Effects of Klamath algae extract on psychological disorders and depression in menopausal women: a pilot study]. [Journal Name]. 2010;**62**(5):381–8. Italian.

**Rationale for exclusion at full-text screening**

Non-English record.

**Record #50 excluded at full-text screening**

Fiegen MM, Benson KD. The frequency of urinary incontinence in Native American women of South Dakota [abstract]. Int Urogynecol J Pelvic Floor Dysfunct. 2009;**20**(3)(Suppl):S270–1.

**Rationale for exclusion at full-text screening**

Wrong concept.

**Record #51 excluded at full-text screening**

Figurelle M, Meyer DM, Perrinez E, Rapp K, Wells R, Guzik AK, et al. Perimenopausal women with migraine present with stroke at a younger age and with less comorbid diabetes [abstract]. Stroke. 2021;**52**(Suppl 1).

**Rationale for exclusion at full-text screening**

Wrong concept.

**Record #52 excluded at full-text screening**

Fisher WI, Elkins G, Thurston RC. Measuring hot flashes: examination of an alternate criterion for ambulatory hot flash detection in postmenopausal women [abstract]. Menopause. 2014;**21**(12):1336.

**Rationale for exclusion at full-text screening**

Unstratified results (population).

**Record #53 excluded at full-text screening**

Flint MP. Complementary and alternative medicines and menopause. Menopause. 2003;**10**(6):491–3.

**Rationale for exclusion at full-text screening**

Wrong population.

**Record #54 excluded at full-text screening**

Fugh-Berman A. CAM: asking the right questions. Menopause. 2008;**15**(1):7–8.

**Rationale for exclusion at full-text screening**

Wrong population.

**Record #55 excluded at full-text screening**

Galan D, Odlum O, Grymonpre R, Brecx M. Medical and dental status of a culture in transition: the case of the Inuit elderly of Canada. Gerodontology. 1993;**10**(1):44–50.

**Rationale for exclusion at full-text screening**

Wrong concept.

**Record #56 excluded at full-text screening**

Galavotti C, Richter DL. Talking about hysterectomy: the experiences of women from four cultural groups. J Womens Health Gend Based Med. 2000;**9**(Suppl 2):S63–7.

**Rationale for exclusion at full-text screening**

Unstratified results (population).

**Record #57 excluded at full-text screening**

Gepner AD, Haller IV, Krueger DC, Korcarz CE, Binkley N, Stein JH. A randomized controlled trial of the effects of vitamin D supplementation on arterial stiffness and aortic blood pressure in Native American women. Atherosclerosis. 2015;**240**(2):526–8.

**Rationale for exclusion at full-text screening**

Wrong concept.

**Record #58 excluded at full-text screening**

Gingrich PM, Fogel CI. Herbal therapy use by perimenopausal women. J Obstet Gynecol Neonatal Nurs. 2003;**32**(2):181–9.

**Rationale for exclusion at full-text screening**

Unclear population.

**Record #59 excluded at full-text screening**

Goins RT, Pilkerton CS. Comorbidity among older American Indians: the Native Elder Care Study. J Cross Cult Gerontol. 2010;**25**(4):343–54.

**Rationale for exclusion at full-text screening**

Wrong concept.

**Record #60 excluded at full-text screening**

Gold EB, Bair Y, Zhang G, Utts J, Greendale GA, Upchurch D, et al. Cross-sectional analysis of specific complementary and alternative medicine (CAM) use by racial/ethnic group and menopausal status: the Study of Women's Health Across the Nation (SWAN). Menopause. 2007;**14**(4):612–23.

**Rationale for exclusion at full-text screening**

Wrong population.

**Record #61 excluded at full-text screening**

Gold R, Michael YL, Whitlock EP, Hubbell FA, Mason ED, Rodriguez BL, et al. Race/ethnicity, socioeconomic status, and lifetime morbidity burden in the Women's Health Initiative: a cross-sectional analysis. J Womens Health (Larchmt). 2006;**15**(10):1161–73.

**Rationale for exclusion at full-text screening**

Wrong concept.

**Record #62 excluded at full-text screening**

Goldstein I, Simon J, Kaunitz A, Altomare C, Yoshida Y, Zhu J, et al. Randomized, double-blind, placebo-controlled multicenter study to evaluate efficacy and safety of ospemifene in patients with moderate to severe vaginal dryness, including evaluation of vulvar health by photographic images [abstract]. J Sex Med. 2019;**16**(4)(Suppl):S73–4.

**Rationale for exclusion at full-text screening**

Unclear population.

**Record #63 excluded at full-text screening**

Gordon NP, Sobel DS, Tarazona EZ. Use of and interest in alternative therapies among adult primary care clinicians and adult members in a large health maintenance organization. West J Med. 1998;**169**(3):153–61.

**Rationale for exclusion at full-text screening**

Unclear population.

**Record #64 excluded at full-text screening**

Grandbois DM, Sanders GF. The resilience of Native American elders. Issues Ment Health Nurs. 2009;**30**(9):569–80.

**Rationale for exclusion at full-text screening**

Wrong concept.

**Record #65 excluded at full-text screening**

Green SM, Haber E, McCabe RE, Soares CN. Cognitive-behavioral group treatment for menopausal symptoms: a pilot study. Arch Womens Ment Health. 2013;**16**(4):325–32.

**Rationale for exclusion at full-text screening**

Unclear population.

**Record #66 excluded at full-text screening**

Grzywacz JG, Arcury TA, Bell RA, Lang W, Suerken CK, Smith SL, et al. Ethnic differences in elders' home remedy use: sociostructural explanations. Am J Health Behav. 2006;**30**(1):39–50.

**Rationale for exclusion at full-text screening**

Wrong concept.

**Record #67 excluded at full-text screening**

Guo Y, Yin T, Wang X, Zhang F, Pan G, Lv H, et al. Traditional uses, phytochemistry, pharmacology and toxicology of the genus Cimicifuga: a review. J Ethnopharmacol. 2017;**209**:264–82.

**Rationale for exclusion at full-text screening**

Wrong population.

**Record #68 excluded at full-text screening**

Haller I, Krueger D, Engelke JA, Binkley N. Serum 25-hydroxyvitamin D in rural and urban postmenopausal women [abstract]. J Bone Miner Res. 2011;**26**.

**Rationale for exclusion at full-text screening**

Wrong context (not U.S. or Canada).

**Record #69 excluded at full-text screening**

Haozous EA, Jaramillo ET, Willging CE. Getting to know: American Indian elder health seeking in an under-funded healthcare system. SSM Qual Res Health. 2021;**1**:100009.

**Rationale for exclusion at full-text screening**

Wrong concept.

**Record #70 excluded at full-text screening**

Hatch JP, Rugh JD, Sakai S, Saunders MJ. Is use of exogenous estrogen associated with temporomandibular signs and symptoms? J Am Dent Assoc. 2001;**132**(3):319–26.

**Rationale for exclusion at full-text screening**

Wrong population.

**Record #71 excluded at full-text screening**

Healey GK, Meadows LM. Inuit women's health in Nunavut, Canada: a review of the literature. Int J Circumpolar Health. 2007;**66**(3):199–214.

**Rationale for exclusion at full-text screening**

Wrong concept.

**Record #72 excluded at full-text screening**

Helenius IM, Korenstein D, Halm EA. Changing use of hormone therapy among minority women since the Women's Health Initiative. Menopause. 2007;**14**(2):216–22.

**Rationale for exclusion at full-text screening**

Unclear population.

**Record #73 excluded at full-text screening**

Hopkins SE, Kwachka P, Lardon C, Mohatt GV. Keeping busy: a Yup'ik/Cup'ik perspective on health and aging. Int J Circumpolar Health. 2007;**66**(1):42–50.

**Rationale for exclusion at full-text screening**

Wrong concept.

**Record #74 excluded at full-text screening**

Hostettmann K. Black cohosh. Phytother Eur. 2009;(51):10–1.

**Rationale for exclusion at full-text screening**

Non-English record.

**Record #75 excluded at full-text screening**

Huang AJ, Moore EE, Boyko EJ, Scholes D, Lin F, Vittinghoff E, et al. Vaginal symptoms in postmenopausal women: self-reported severity, natural history, and risk factors. Menopause. 2010;**17**(1):121–6.

**Rationale for exclusion at full-text screening**

Unstratified results (population).

**Record #76 excluded at full-text screening**

Hubayter Z, Simon JA. Testosterone therapy for sexual dysfunction in postmenopausal women. Climacteric. 2008;**11**(3):181–91.

**Rationale for exclusion at full-text screening**

Wrong population.

**Record #77 excluded at full-text screening**

Im EO, Ko Y, Hwang H, Chee W. "Symptom-specific or holistic": menopausal symptom management. Health Care Women Int. 2012;**33**(6):575–92.

**Rationale for exclusion at full-text screening**

Wrong population.

**Record #78 excluded at full-text screening**

Innes K, D'Adamo C, Parker EA, Feinberg T. Prevalence and correlates of black cohosh consumption in the United States [abstract]. Glob Adv Health Med. 2018;**7**:121.

**Rationale for exclusion at full-text screening**

Unclear population.

**Record #79 excluded at full-text screening**

Israel D, Youngkin EQ. Herbal therapies for perimenopausal and menopausal complaints. Pharmacotherapy. 1997;**17**(5):970–84.

**Rationale for exclusion at full-text screening**

Wrong population.

**Record #80 excluded at full-text screening**

Jaramillo ET, Willging CE. Producing insecurity: healthcare access, health insurance, and wellbeing among American Indian elders. Soc Sci Med. 2021;**268**:113384.

**Rationale for exclusion at full-text screening**

Wrong concept.

**Record #81 excluded at full-text screening**

Jezewski E, Miller A, Eusebio M, Potter J. Targeted telehealth education increases interest in using telehealth among a diverse group of low-income older adults. Int J Environ Res Public Health. 2022;**19**(20):1–9.

**Rationale for exclusion at full-text screening**

Wrong concept.

**Record #82 excluded at full-text screening**

Joffe C. Working with Dr. Tiller: staff recollections of Women's Health Care Services of Wichita. Perspect Sex Reprod Health. 2011;**43**(3):199–204.

**Rationale for exclusion at full-text screening**

Wrong concept.

**Record #83 excluded at full-text screening**

Johnson MC, Harvey RE, Kruse SM, Joyner MJ, Miller VM, Barnes JN. Aortic hemodynamics following discontinuation of menopausal hormone therapy in postmenopausal women. Artery Res. 2014;**8**(4):173.

**Rationale for exclusion at full-text screening**

Unclear population.

**Record #84 excluded at full-text screening**

Jones EK, Jurgenson JR, Katzenellenbogen JM, Thompson SC. Menopause and the influence of culture: another gap for Indigenous Australian women? BMC Womens Health. 2012;**12**:43.

**Rationale for exclusion at full-text screening**

Wrong context (not U.S. or Canada).

**Record #85 excluded at full-text screening**

Kam IW, Dennehy CE, Tsourounis C. Dietary supplement use among menopausal women attending a San Francisco health conference. Menopause. 2002;**9**(1):72–8.

**Rationale for exclusion at full-text screening**

Unclear population.

**Record #86 excluded at full-text screening**

Kashani L, Nikbakhat MR, Akhondzadeh S. Herbal medicine for women's health. J Med Plants. 2015;**14**(55):1–8.

**Rationale for exclusion at full-text screening**

Wrong population.

**Record #87 excluded at full-text screening**

Keenan NL, Mark S, Fugh-Berman A, Browne D, Kaczmarczyk J, Hunter C. Severity of menopausal symptoms and use of both conventional and complementary/alternative therapies. Menopause. 2003;**10**(6):507–15.

**Rationale for exclusion at full-text screening**

Unclear population.

**Record #88 excluded at full-text screening**

Kelly JP, Kaufman DW, Rosenberg L, Kelley K, Cooper SG, Mitchell AA. Use of postmenopausal hormone therapy since the Women's Health Initiative findings. Pharmacoepidemiol Drug Saf. 2005;**14**(12):837–42.

**Rationale for exclusion at full-text screening**

Unclear population.

**Record #89 excluded at full-text screening**

Kessel B. Alternatives to estrogen for menopausal women. Proc Soc Exp Biol Med. 1998;**217**(1):38–44.

**Rationale for exclusion at full-text screening**

Wrong population.

**Record #90 excluded at full-text screening**

Kessel B, Kronenberg F. The role of complementary and alternative medicine in management of menopausal symptoms. Endocrinol Metab Clin North Am. 2004;**33**(4):717–39.

**Rationale for exclusion at full-text screening**

Wrong population.

**Record #91 excluded at full-text screening**

Kingsberg S, Amadio JM, Graham S. Women's attitudes and behaviors towards vulvar and vaginal atrophy (VVA) [abstract]. Menopause. 2015;**22**(12):1369–70.

**Rationale for exclusion at full-text screening**

Unclear population.

**Record #92 excluded at full-text screening**

Kolodziejczyk-Czepas J, Liudvytska O. Rheum rhaponticum and Rheum rhabarbarum: a review of phytochemistry, biological activities and therapeutic potential. Phytochem Rev. 2021;**20**(3):589–607.

**Rationale for exclusion at full-text screening**

Wrong population.

**Record #93 excluded at full-text screening**

Kripke DF, Jean-Louis G, Elliott JA, Klauber MR, Rex KM, Tuunainen A, et al. Ethnicity, sleep, mood, and illumination in postmenopausal women. BMC Psychiatry. 2004;**4**:8.

**Rationale for exclusion at full-text screening**

Wrong concept.

**Record #94 excluded at full-text screening**

Kupferer EM, Dormire SL, Becker H. Complementary and alternative medicine use for vasomotor symptoms among women who have discontinued hormone therapy. J Obstet Gynecol Neonatal Nurs. 2009;**38**(1):50–9.

**Rationale for exclusion at full-text screening**

Unstratified results (population).

**Record #95 excluded at full-text screening**

Lawton B, Clarke MJ, Gibson-Helm M, Boyle JA. The lives of women and babies matter: a call for action in Indigenous and First Nations women's health and wellbeing. Int J Gynaecol Obstet. 2021;**155**(2):167–9.

**Rationale for exclusion at full-text screening**

Wrong record type.

**Record #96 excluded at full-text screening**

Liddell JL, Lilly JM. Healthcare experiences of uninsured and under-insured American Indian women in the United States. Glob Health Res Policy. 2022;**7**(1):5.

**Rationale for exclusion at full-text screening**

Wrong concept.

**Record #97 excluded at full-text screening**

Lieberman LS, Stoller EP, Burg MA. Women's health care: cross-cultural encounters within the medical system. J Fla Med Assoc. 1997;**84**(6):364–73.

**Rationale for exclusion at full-text screening**

Wrong concept.

**Record #98 excluded at full-text screening**

Lind BK, Lafferty WE, Tyree PT, Diehr PK. Comparison of health care expenditures among insured users and nonusers of complementary and alternative medicine in Washington State: a cost minimization analysis. J Altern Complement Med. 2010;**16**(4):411–7.

**Rationale for exclusion at full-text screening**

Unclear population.

**Record #99 excluded at full-text screening**

Lunny CA, Fraser SN. The use of complementary and alternative medicines among a sample of Canadian menopausal-aged women. J Midwifery Womens Health. 2010;**55**(4):335–43.

**Rationale for exclusion at full-text screening**

Unclear population.

**Record #100 excluded at full-text screening**

Ma J, Drieling R, Stafford RS. US women desire greater professional guidance on hormone and alternative therapies for menopause symptom management. Menopause. 2006;**13**(3):506–16.

**Rationale for exclusion at full-text screening**

Unclear population.

**Record #101 excluded at full-text screening**

Mahady GB, Fabricant D, Chadwick LR, Dietz B. Black cohosh: an alternative therapy for menopause? Nutr Clin Care. 2002;**5**(6):283–9.

**Rationale for exclusion at full-text screening**

Wrong population.

**Record #102 excluded at full-text screening**

McCloskey L, Bernstein J, Goler-Blount L, Greiner A, Norton A, Jones E, et al. It's time to eliminate racism and fragmentation in women's health care. Womens Health Issues. 2021;**31**(3):186–9.

**Rationale for exclusion at full-text screening**

Wrong concept.

**Record #103 excluded at full-text screening**

McClung MR, Kagan R, Graham S, Bernick B, Mirkin S, Constantine G. Effects of E2/P4 oral capsules on bone turnover in women with vasomotor symptoms. Menopause. 2022;**29**(3):304–8.

**Rationale for exclusion at full-text screening**

Unclear population.

**Record #104 excluded at full-text screening**

Milligan C, Mantla R, Blake G, Zoe JB, Steinwand T, Greenland S, et al. Health system learning with Indigenous communities: a study protocol for a two-eyed seeing review and multiple case study. Health Res Policy Syst. 2022;**20**:65.

**Rationale for exclusion at full-text screening**

Wrong concept.

**Record #105 excluded at full-text screening**

Miroddi M, Calapai G, Navarra M, Minciullo PL, Gangemi S. Passiflora incarnata L.: ethnopharmacology, clinical application, safety and evaluation of clinical trials. J Ethnopharmacol. 2013;**150**(3):791–804.

**Rationale for exclusion at full-text screening**

Wrong population.

**Record #106 excluded at full-text screening**

Monchalin R, Smylie J, Nowgesic E. "I guess I shouldn't come back here": racism and discrimination as a barrier to accessing health and social services for urban Métis women in Toronto, Canada. J Racial Ethn Health Disparities. 2020;**7**(2):251–61.

**Rationale for exclusion at full-text screening**

Wrong concept.

**Record #107 excluded at full-text screening**

Nedrow A, Miller J, Walker M, Nygren P, Huffman LH, Nelson HD. Complementary and alternative therapies for the management of menopause-related symptoms: a systematic evidence review. Arch Intern Med. 2006;**166**(14):1453–65.

**Rationale for exclusion at full-text screening**

Wrong population.

**Record #108 excluded at full-text screening**

Ness J, Cirillo DJ, Weir DR, Nisly NL, Wallace RB. Use of complementary medicine in older Americans: results from the Health and Retirement Study. Gerontologist. 2005;**45**(4):516–24.

**Rationale for exclusion at full-text screening**

Unclear population.

**Record #109 excluded at full-text screening**

Newton KM, Buist DSM, Keenan NL, Anderson LA, LaCroix AZ. Use of alternative therapies for menopause symptoms: results of a population-based survey. Obstet Gynecol. 2002;**100**(1):18–25.

**Rationale for exclusion at full-text screening**

Unclear population.

**Record #110 excluded at full-text screening**

Newton KM, Reed SD, LaCroix AZ, Grothaus LC, Ehrlich K, Guiltinan J. Treatment of vasomotor symptoms of menopause with black cohosh, multibotanicals, soy, hormone therapy, or placebo: a randomized trial. Ann Intern Med. 2006;**145**(12):869–79.

**Rationale for exclusion at full-text screening**

Unclear population.

**Record #111 excluded at full-text screening**

Northington GM, De Vries HF, Bogner HR. Prolonged estrogen use and newly incident urinary incontinence in postmenopausal women: a community-based prospective longitudinal study [abstract]. J Pelvic Med Surg. 2010;**16**(5):S130.

**Rationale for exclusion at full-text screening**

Unstratified results (population).

**Record #112 excluded at full-text screening**

O’Hara DA. Herbals and supplements: Part II. Prog Anesthesiol. 2003;**17**(15):235–42.

**Rationale for exclusion at full-text screening**

Wrong population.

**Record #113 excluded at full-text screening**

Page K, Brownie S, Wohlmuth H. Natural management options for menopause. Integr Med (Encinitas). 2005;**4**(1):20–8.

**Rationale for exclusion at full-text screening**

Wrong population.

**Record #114 excluded at full-text screening**

Palacio C, Masri G, Mooradian AD. Black cohosh for the management of menopausal symptoms: a systematic review of clinical trials. Drugs Aging. 2009;**26**(1):23–36.

**Rationale for exclusion at full-text screening**

Wrong population.

**Record #115 excluded at full-text screening**

Palacios JF, Portillo CJ. Understanding Native women's health: historical legacies. J Transcult Nurs. 2009;**20**(1):15–27.

**Rationale for exclusion at full-text screening**

Wrong concept.

**Record #116 excluded at full-text screening**

Pan M, Zhou J, Pan X, Wang J, Qi Q, Wang L. Drugs for the treatment of postmenopausal symptoms: hormonal and non-hormonal therapy. Life Sci. 2023;**312**:121255.

**Rationale for exclusion at full-text screening**

Wrong population.

**Record #117 excluded at full-text screening**

Patade A, Devareddy L, Lucas EA, Korlagunta K, Daggy BP, Arjmandi BH. Flaxseed reduces total and LDL cholesterol concentrations in Native American postmenopausal women. J Womens Health (Larchmt). 2008;**17**(3):355–66.

**Rationale for exclusion at full-text screening**

Wrong concept.

**Record #118 excluded at full-text screening**

Perez MV, Wang PJ, Larson JC, Virnig BA, Cochrane B, Curb JD, et al. Effects of postmenopausal hormone therapy on incident atrial fibrillation: the Women's Health Initiative randomized controlled trials. Circ Arrhythm Electrophysiol. 2012;**5**(6):1108–16.

**Rationale for exclusion at full-text screening**

Unstratified results (population).

**Record #119 excluded at full-text screening**

Plotnikoff GA, Watanabe K, Torkelson C, La Valleur J, Radosevich DM. The TU-025 keishibukuryogan clinical trial for hot flash management in postmenopausal women: results and lessons for future research. Menopause. 2011;**18**(8):886–92.

**Rationale for exclusion at full-text screening**

Wrong population.

**Record #120 excluded at full-text screening**

Portella CFS, De Azevedo da Silva I, Dos Santos Simes R, Baracat EC, Soares JM, Sorpreso ICE. An overview of systematic reviews in medicinal plants and herbal formulations for the treatment of climacteric symptoms: a lack of research on southern hemispheric plants [abstract]. Menopause. 2022;**29**(12):1479.

**Rationale for exclusion at full-text screening**

Wrong population.

**Record #121 excluded at full-text screening**

Pothiwala P, Evans EM, Chapman-Novakofski KM. Ethnic variation in risk for osteoporosis among women: a review of biological and behavioral factors. J Womens Health (Larchmt). 2006;**15**(6):709–19.

**Rationale for exclusion at full-text screening**

Wrong concept.

**Record #122 excluded at full-text screening**

Rahimi R, Kiani S. Chemical compound and therapeutic effects of Hypericum perforatum. Der Pharm Lett. 2016;**8**(9):237–41.

**Rationale for exclusion at full-text screening**

Wrong population.

**Record #123 excluded at full-text screening**

Reutter L, Neufeld A, Harrison MJ. A review of the research on the health of low-income Canadian women. Can J Nurs Res. 2000;**32**(1):75–97.

**Rationale for exclusion at full-text screening**

Wrong concept.

**Record #124 excluded at full-text screening**

Reynolds RF, Obermeyer CM, Walker AM, Guilbert D. Side effects and sociobehavioral factors associated with the discontinuation of hormone therapy in a Massachusetts health maintenance organization. Menopause. 2001;**8**(3):189–99.

**Rationale for exclusion at full-text screening**

Wrong population.

**Record #125 excluded at full-text screening**

Richter DL, Corwin SJ, Rheaume CE, McKeown RE. Perceptions of alternative therapies available for women facing hysterectomy or menopause. J Women Aging. 2001;**13**(4):21–37.

**Rationale for exclusion at full-text screening**

Unstratified results (population).

**Record #126 excluded at full-text screening**

Rousseau P. Native-American elders: health care status. Clin Geriatr Med. 1995;**11**(1):83–95.

**Rationale for exclusion at full-text screening**

Wrong concept.

**Record #127 excluded at full-text screening**

Roussos-Ross K. Treatment of mood disorders in midlife women [abstract]. Menopause. 2017;**24**(12):1422.

**Rationale for exclusion at full-text screening**

Unclear population.

**Record #128 excluded at full-text screening**

Russell L, Hicks GS, Low AK, Shepherd JM, Brown CA. Phytoestrogens: a viable option? Am J Med Sci. 2002;324(4):185–8.

**Rationale for exclusion at full-text screening**

Wrong population.

**Record #129 excluded at full-text screening**

Ryan CA, Ghali WA, Boss RD, Moskowitz MA, Freund KM. Care during menopause: comparison of a women's health practice and traditional care. J Womens Health Gend Based Med. 1999;**8**(10):1295–302.

**Rationale for exclusion at full-text screening**

Unclear population.

**Record #130 excluded at full-text screening**

Salari S, Amiri MS, Ramezani M, Moghadam AT, Elyasi S, Sahebkar A, Emami SA. Ethnobotany, phytochemistry, traditional and modern uses of Actaea racemosa L. (black cohosh): a review. Adv Exp Med Biol. 2021;**1308**:403–49.

**Rationale for exclusion at full-text screening**

Wrong population.

**Record #131 excluded at full-text screening**

Salehian T, Dehcheshmeh FS. Effects of isoflavones on the climacteric period in women [abstract]. Avicenna J Phytomed. 2015;**5**(Suppl 1):133–4.

**Rationale for exclusion at full-text screening**

Unclear population.

**Record #132 excluded at full-text screening**

Saltiki K, Manteos E, Doukas C, Kanakakis I, Zotos P, Lazaros L, et al. Estrogen receptor beta gene variants may be associated with more favorable metabolic profile in postmenopausal women undergoing coronary angiography. Exp Clin Endocrinol Diabetes. 2009;**117**(10):610–5.

**Rationale for exclusion at full-text screening**

Wrong concept.

**Record #133 excluded at full-text screening**

Sampson A. Honouring traditional knowledge to promote healing in healthcare relationships [abstract]. Early Interv Psychiatry. 2023;**17**:156.

**Rationale for exclusion at full-text screening**

Wrong concept.

**Record #134 excluded at full-text screening**

Sarto GE, Brasileiro J, Franklin DJ. Women's health: racial and ethnic health inequities. Glob Adv Health Med. 2013;**2**(5):50–3.

**Rationale for exclusion at full-text screening**

Wrong concept.

**Record #135 excluded at full-text screening**

Sassarini J, Lumsden MA. Hot flushes: are there effective alternatives to estrogen? Menopause Int. 2010;**16**(2):81–8.

**Rationale for exclusion at full-text screening**

Wrong population.

**Record #136 excluded at full-text screening**

Saylors K. The Women's Circle comes full circle. J Psychoactive Drugs. 2003;**35**(1):59–62.

**Rationale for exclusion at full-text screening**

Wrong concept.

**Record #137 excluded at full-text screening**

Schonberg MA, Wee CC. Menopausal symptom management and prevention counseling after the Women's Health Initiative among women seen in an internal medicine practice. J Womens Health (Larchmt). 2005;**14**(6):507–14.

**Rationale for exclusion at full-text screening**

Unclear population.

**Record #138 excluded at full-text screening**

Schure MB, Conte KP, Goins RT. Unmet assistance need among older American Indians: the Native Elder Care Study. Gerontologist. 2015;**55**(6):920–8.

**Rationale for exclusion at full-text screening**

Wrong concept.

**Record #139 excluded at full-text screening**

Scoglio S, Benedetti S, Canino C, Santagni S, Rattighieri E, Chierchia E, et al. Effect of a 2-month treatment with Klamin, a Klamath algae extract, on the general well-being, antioxidant profile and oxidative status of postmenopausal women. Gynecol Endocrinol. 2009;**25**(4):235–40.

**Rationale for exclusion at full-text screening**

Wrong population.

**Record #140 excluded at full-text screening**

Scott R, Nahin RL, Sussman BJ, Feinberg T. Physician office visits that included complementary health approaches in U.S. adults: 2005–2015. J Integr Complement Med. 2022;**28**(8):641–50.

**Rationale for exclusion at full-text screening**

Unclear population.

**Record #141 excluded at full-text screening**

Shepherd-Banigan M, Goldstein KM, Coeytaux RR, McDuffie JR, Goode AP, Kosinski AS, et al. Improving vasomotor symptoms, psychological symptoms, and health-related quality of life in peri- or post-menopausal women through yoga: an umbrella systematic review and meta-analysis. Complement Ther Med. 2017;**34**:156–64.

**Rationale for exclusion at full-text screening**

Wrong population.

**Record #142 excluded at full-text screening**

**Sillett M. National Inuit women health concern [abstract].** Arctic Med Res. 1988;**47**(Suppl 1):102–3.

**Rationale for exclusion at full-text screening**

Wrong concept.

**Record #143 excluded at full-text screening**

Singh B, Liu XD, Der-Martirosian C, Hardy M, Singh VJ, Shepard N, et al. Menopausal issues: a national probability sample of US women. *Altern Ther Health Med*. 2007;**13**(3):24–9.

**Rationale for exclusion at full-text screening**

Unstratified results (population).

**Record #144 excluded at full-text screening**

Slomski A. Which postmenopausal women should use testosterone for low sexual desire? *JAMA*. 2020;**323**(6):493–5.

**Rationale for exclusion at full-text screening**

Wrong population.

**Record #145 excluded at full-text screening**

Soares CN, Prouty J, Born L, Steiner M. Treatment of menopause-related mood disturbances. *CNS Spectr*. 2005;**10**(6):489–497.

**Rationale for exclusion at full-text screening**

Wrong population.

**Record #146 excluded at full-text screening**

Sohail U, Ashraf I, Almashhrawi A, Rahman R, Nguyen D, Bechtold M. Black cohosh-induced abdominal pain: sometimes more than just a supplement [abstract]. *Am J Gastroenterol*. 2015;**110**(Suppl 1):S284–S285.

**Rationale for exclusion at full-text screening**

Unclear population.

**Record #147 excluded at full-text screening**

Sommerfeld DH, Jaramillo ET, Lujan E, Haozous E, Willging CE. Health care access and utilization for American Indian elders: a concept-mapping study. *J Gerontol B Psychol Sci Soc Sci*. 2021;**76**(1):141–151.

**Rationale for exclusion at full-text screening**

Wrong concept.

**Record #148 excluded at full-text screening**

Strauss KF, Mokdad A, Ballew C, Mendlein JM, Will JC, Goldberg HI, et al. The health of Navajo women: findings from the Navajo Health and Nutrition Survey, 1991–1992. *J Nutr*. 1997;**127**(10 Suppl):2128S–2133S.

**Rationale for exclusion at full-text screening**

Wrong concept.

**Record #149 excluded at full-text screening**

Taylor M. Alternative medicine and the perimenopause: an evidence-based review. *Obstet Gynecol Clin North Am*. 2002;**29**(3):555–73.

**Rationale for exclusion at full-text screening**

Wrong population.

**Record #150 excluded at full-text screening**

Tesch BJ. Herbs commonly used by women: an evidence-based review. *Dis Mon*. 2002;**48**(10):671–96.

**Rationale for exclusion at full-text screening**

Wrong population.

**Record #151 excluded at full-text screening**

Tetteh A. Use of complementary and alternative medicine during the menopause transition: longitudinal results from the Study of Women's Health Across the Nation [letter]. *Menopause*. 2008;**15**(5):1029.

**Rationale for exclusion at full-text screening**

Wrong record type.

**Record #152 excluded at full-text screening**

Thompson JJ, Ritenbaugh C, Nichter M. Why women choose compounded bioidentical hormone therapy: lessons from a qualitative study of menopausal decision-making. *BMC Womens Health*. 2017;**17**:97.

**Rationale for exclusion at full-text screening**

Wrong population.

**Record #153 excluded at full-text screening**

Tigner JL. Alaska Native elders' successful ageing and rural healthcare [abstract]. *J Investig Med*. 2018;**66**(1):191.

**Rationale for exclusion at full-text screening**

Wrong concept.

**Record #154 excluded at full-text screening**

Tonkin R, Freeman S, Martin J, Ward V, Skinner K. First Nations Elders’ perspectives of engagement in community programs in Nak’azdli Whut’en, British Columbia, Canada. *Can J Public Health*. 2018;**109**(5-6):810–816.

**Rationale for exclusion at full-text screening**

Wrong concept.

**Record #155 excluded at full-text screening**

Tu D, Roberta E, Hadjipavlou PG, Varcoe C, Dehoney J, Brown A. Mental health effects of partnering Indigenous Elders within primary care teams: mixed-methods prospective cohort study [abstract]. *Can Fam Physician*. 2018;**64**(Suppl 1):S1-S115.

**Rationale for exclusion at full-text screening**

Wrong concept.

**Record #156 excluded at full-text screening**

Turner NJ, Hebda RJ. Contemporary use of bark for medicine by two Salishan native elders of southeast Vancouver Island, Canada. *J Ethnopharmacol*. 1990;**29**(1):59–72.

**Rationale for exclusion at full-text screening**

Wrong concept.

**Record #157 excluded at full-text screening**

Upchurch DM, Dye CE, Chyu L, Gold EB, Greendale GA. Demographic, behavioral, and health correlates of complementary and alternative medicine and prayer use among midlife women: 2002. *J Womens Health (Larchmt)*. 2010;**19**(1):23–30.

**Rationale for exclusion at full-text screening**

Wrong population.

**Record #158 excluded at full-text screening**

Venzke L, Calvert JF Jr, Gilbertson B. A randomized trial of acupuncture for vasomotor symptoms in post-menopausal women. *Complement Ther Med*. 2010;**18**(2):59–66.

**Rationale for exclusion at full-text screening**

Unclear population.

**Record #159 excluded at full-text screening**

Wade SW, Satram-Hoang S, Nadkar A, Macarios D, Tosteson ANA. Impact of medication adherence on health care utilization and productivity: self-reported data from a cohort of postmenopausal women on osteoporosis therapy. *Clin Ther*. 2011;**33**(12):2006–15.

**Rationale for exclusion at full-text screening**

Unstratified results (population).

**Record #160 excluded at full-text screening**

Wampler NS, Chen Z, Jacobsen C, Henderson JA, Howard BV, Rossouw JE. Bone mineral density of American Indian and Alaska Native women compared with non-Hispanic white women: results from the Women's Health Initiative Study. *Menopause*. 2005;**12**(5):536–44.

**Rationale for exclusion at full-text screening**

Wrong concept.

**Record #161 excluded at full-text screening**

Wang MY, Tsai PS, Lee PH, Chang WY, Yang CM. The efficacy of reflexology: systematic review. *J Adv Nurs*. 2008;**62**(5):512–20.

**Rationale for exclusion at full-text screening**

Wrong population.

**Record #162 excluded at full-text screening**

Wathen CN. Alternatives to hormone replacement therapy: a multi-method study of women's experiences. *Complement Ther Med*. 2006;**14**(3):185–92.

**Rationale for exclusion at full-text screening**

Unclear population.

**Record #163 excluded at full-text screening**

Wathen CN. Health information seeking in context: how women make decisions regarding hormone replacement therapy. *J Health Commun*. 2006;**11**(5):477–93.

**Rationale for exclusion at full-text screening**

Unstratified results (population).

**Record #164 excluded at full-text screening**

Waxman AG, Haffner WHJ, Howe J, Wilder K, Ogburn T, Murphy N, et al. A 50-year commitment to American Indian and Alaska Native women. *Obstet Gynecol*. 2020;**136**(4):739–44.

**Rationale for exclusion at full-text screening**

Wrong concept.

**Record #165 excluded at full-text screening**

Williams RE, Kalilani L, DiBenedetti DB, Zhou X, Fehnel SE, Clark RV. Healthcare seeking and treatment for menopausal symptoms in the United States. *Maturitas*. 2007;**58**(4):348–58.

**Rationale for exclusion at full-text screening**

Unclear population.

**Record #166 excluded at full-text screening**

Willms DG, Lange P, Bayfield D, Beardy M, Lindsay EA, Cole DC, et al. A lament by women for "the people, the land" [Nishnawbi-Aski Nation]: an experience of loss. *Can J Public Health*. 1992;**83**(5):331–4.

**Rationale for exclusion at full-text screening**

Wrong concept.

**Record #167 excluded at full-text screening**

Wilson D. First Nations health in Canada [abstract]. *Aust N Z J Obstet Gynaecol*. 2022;**62**:83–84.

**Rationale for exclusion at full-text screening**

Wrong concept.

**Record #168 excluded at full-text screening**

Wortman E, Lewis J. “Grandpa speaks, nobody listens.” Gerotranscendence, a change in mindset of Alaska Native elders [abstract]. *J Investig Med*. 2019;**67**(1):190.

**Rationale for exclusion at full-text screening**

Wrong concept.

**Record #169 excluded at full-text screening**

Wortman ES, Lewis JP. Gerotranscendence and Alaska Native successful aging in the Aleutian Pribilof Islands, Alaska. *J Cross Cult Gerontol*. 2021;**36**(1):43-67.

**Rationale for exclusion at full-text screening**

Wrong concept.

**Record #170 excluded at full-text screening**

Xie L, Lewis-Beck C, Wang H, Altinbas S, Baser O. Demographic and clinical characteristics of menopausal women in the University of Michigan Women's Registry Database [abstract]. *Value Health*. 2012;**15**(7):A536.

**Rationale for exclusion at full-text screening**

Unclear population.

**Record #171 excluded at full-text screening**

Yuksel N, Evaniuk D, Huang L, Malhotra U, Blake J, Wolfman W, Fortier M. Corrigendum to: Guideline No. 422a: Menopause: Vasomotor symptoms, prescription therapeutic agents, complementary and alternative medicine, nutrition, and lifestyle [J Obstet Gynaecol Can. 2021;43(10):1188–1204.e1]. *J Obstet Gynaecol Can*. 2022;**44**(2):227.

**Rationale for exclusion at full-text screening**

Unclear population.

**Record #172 excluded at full-text screening**

**Yuksel N, Evaniuk D, Huang L, Malhotra U, Blake J, Wolfman W, Fortier M.** Corrigendum to: Guideline No. 422a: Menopause: Vasomotor symptoms, prescription therapeutic agents, complementary and alternative medicine, nutrition, and lifestyle [J Obstet Gynaecol Can. 2021;43(10):1188–1204.e1]. J Obstet Gynaecol Can. 2022;**44**(2):227.

**Rationale for exclusion at full-text screening**

Unclear population.

**Record #173 excluded at full-text screening**

Zhang Y, Shen CL, Brismée JM, Peck K, Dagda RY, Lo DF. Exploring the effects of self-managed acupressure on pain, function and osteoarthritis biomarkers among postmenopausal women with osteoarthritic knee pain: A feasibility study [abstract]. J Altern Complement Med. 2013;**19**(7):A31.

**Rationale for exclusion at full-text screening**

Unclear population.

**Record #174 excluded at full-text screening**

Ziabakhsh S, Pederson A, Prodan-Bhalla N, Middagh D, Jinkerson-Brass S. Women-centered and culturally responsive heart health promotion among Indigenous women in Canada. Health Promot Pract. 2016;**17**(6):814–826.

**Rationale for exclusion at full-text screening**

Wrong concept.

**Record #175 excluded at full-text screening**

Ayunerak P, Alstrom D, Moses C, Charlie J, Rasmus SM. Yup’ik culture and context in Southwest Alaska: Community member perspectives of tradition, social change, and prevention. Am J Community Psychol. 2014;**54**(1-2):91–99.

**Rationale for exclusion at full-text screening**

Wrong concept.

**Record #176 excluded at full-text screening**

Bird ME, Henley E. Women's Wellness Consumer Forum. IHS Prim Care Provid. 1993;**18**(7):132–3.

**Rationale for exclusion at full-text screening**

Wrong concept.

**Record #177 excluded at full-text screening**

Wise EA, Carmichael SK, Belar CD, Jordan CB, Berlant NE, Kazarian SS, et al. Women's health: a cultural perspective. In: Kazarian SS, Evans DR, editors. Handbook of cultural health psychology. San Diego (CA): Academic Press; 2001. p. 445–73.

**Rationale for exclusion at full-text screening**

Wrong concept.

**Record #178 excluded at full-text screening**

**Hudson T.** Women's health update: black cohosh: state of the science and art. Townsend Lett. 2006 May;(274):128–31.

**Rationale for exclusion at full-text screening**

Wrong population.

**Record #179 excluded at full-text screening**

**[Anonymous].** Women use 'alternative' therapies for menopause symptoms. Better Nutr. 2001 Jan;**63**(1):26.

**Rationale for exclusion at full-text screening**

Unclear population.

**Record #180 excluded at full-text screening**

**Conboy L, O'Connell E, Domar A.** Women at mid-life: symptoms, attitudes, and choices: an Internet based survey. Maturitas. 2001;**38**(2):129–136.

**Rationale for exclusion at full-text screening**

Unclear population.

**Record #181 excluded at full-text screening**

**Northrup C.**The wisdom of menopause: creating physical and emotional health and healing during the change. New York: Bantam Books; 2001. 605 p.

**Rationale for exclusion at full-text screening**

Book.

**Record #182 excluded at full-text screening**

**Burton CMA.** Wilaat hooxhl nisga'ahl {galdoo'o} {yans}: Gik'uuhl-gi, guun-sa ganhl angoo gam´ using plants the Nisga'a way: Past, present and future use [dissertation]. Victoria (BC): University of Victoria; 2015. Available from: ProQuest Dissertations & Theses.

**Rationale for exclusion at full-text screening**

Wrong concept.

**Record #183 excluded at full-text screening**

**Finke B.** What's new from the Elder Care Initiative. IHS Prim Care Provider. 2000;**25**(5):89.

**Rationale for exclusion at full-text screening**

Wrong concept.

**Record #184 excluded at full-text screening**

**Larson JP.** Wase Wakpa: A community-university connection. Home Health Care Manag Pract. 2005;**18**(1):53–6.

**Rationale for exclusion at full-text screening**

Wrong concept.

**Record #185 excluded at full-text screening**

**Baca R.** A voice for Indian elders. IHS Prim Care Provid. 1998;**23**(5):62–3.

**Rationale for exclusion at full-text screening**

Wrong concept.

**Record #186 excluded at full-text screening**

**Bowes JI.** Use of complementary and alternative medicine: urban study of mid-life women [dissertation]. Albuquerque (NM): University of New Mexico; 2001. Available from: UMI Dissertation Services, ProQuest Information and Learning.

**Rationale for exclusion at full-text screening**

Wrong population.

**Record #187 excluded at full-text screening**

**Bair YA.** Use of complementary and alternative medicine during the menopause transition [dissertation]. Davis (CA): University of California, Davis; 2003. Available from: UMI Dissertation Services, ProQuest Information and Learning.

**Rationale for exclusion at full-text screening**

Wrong population.

**Record #188 excluded at full-text screening**

**Harvard Women's Health Watch.** Update on black cohosh for hot flashes. Harv Womens Health Watch. 2004 Jan;**11**(5):2–3.

**Rationale for exclusion at full-text screening**

Wrong population.

**Record #189 excluded at full-text screening**

Seibel MM. Treating hot flushes without hormone replacement therapy. J Fam Pract. 2003;**52**(4):291–6.

**Rationale for exclusion at full-text screening**

Wrong population.

**Record #190 excluded at full-text screening**

Wuest J. Towards understanding women's health through a social determinants lens. Can J Nurs Res. 2006;**38**(1):3–5.

**Rationale for exclusion at full-text screening**

Wrong concept.

**Record #191 excluded at full-text screening**

Godfrey JR, Low Dog T. Toward optimal health: menopause as a rite of passage. J Womens Health (Larchmt). 2008;**17**(4):509–14.

**Rationale for exclusion at full-text screening**

Wrong population.

**Record #192 excluded at full-text screening**

W.K. Kellogg Foundation. Timely opportunities: what works in community care for the elderly: lessons learned from W.K. Kellogg Foundation programming. Battle Creek (MI): W.K. Kellogg Foundation; 1995. 69 p.

**Rationale for exclusion at full-text screening**

Book.

**Record #193 excluded at full-text screening**

Martin L, Jung P. Taking charge of the change: a holistic approach to the three phases of menopause. Albany (NY): Delmar/Thomson Learning; 2002. 334 p.

**Rationale for exclusion at full-text screening**

Book.

**Record #194 excluded at full-text screening**

**Indian Health Service.** Survey of Indian Health Service physicians to study issues related to recruitment and retention – survey report. Rockville (MD): Staff Office of Planning, Evaluation and Research; 1992. Report No.: SSI-16.

**Rationale for exclusion at full-text screening**

Wrong concept.

**Record #195 excluded at full-text screening**

**HerbalGram.** Survey finds women want more guidance from doctors about alternative therapies for menopause symptoms. HerbalGram. 2005 Summer;(67):68.

**Rationale for exclusion at full-text screening**

Unclear population.

**Record #196 excluded at full-text screening**

Weng HH, McBride CM, Bosworth HB, Grambow SC, Siegler IC, Bastian LA. Racial differences in physician recommendation of hormone replacement therapy. Prev Med. 2001;**33**(6):668–673.

**Rationale for exclusion at full-text screening**

Wrong population.

**Record #197 excluded at full-text screening**

Webster AD, Finstad DA, Kurzer MS, Torkelson CJ. Quality of life among postmenopausal women enrolled in the Minnesota Green Tea Trial. Maturitas. 2018;**108**:1–6.

**Rationale for exclusion at full-text screening**

Unstratified results (population).

**Record #198 excluded at full-text screening**

Castleman M. Power plants. Modern Maturity. 2003;**46**(1):50–5.

**Rationale for exclusion at full-text screening**

Unclear population.

**Record #199 excluded at full-text screening**

Nekhlyudov L, Bush T, Bonomi AE, Ludman EJ, Newton KM. Physicians’ and women’s views on hormone therapy and breast cancer risk after the WHI: a qualitative study. Women Health. 2009;**49**(4):280–3.

**Rationale for exclusion at full-text screening**

Unclear population.

**Record #200 excluded at full-text screening**

Kinch JL, Jakubec S. Out of the multiple margins: older women managing their health care. CJNR Can J Nurs Res. 2004;**36**(4):90–108.

**Rationale for exclusion at full-text screening**

Wrong concept.

**Record #201 excluded at full-text screening**

Amoss PT, Harrell S, editors. Other ways of growing old: anthropological perspectives. Stanford, CA: Stanford University Press; 1981. 270 p.

**Rationale for exclusion at full-text screening**

Book.

**Record #202 excluded at full-text screening**

Cass I, Runowicz CD. Nonhormonal alternatives to treating menopausal symptoms. Am J Manag Care. 1998;**4**(5):732–8.

**Rationale for exclusion at full-text screening**

Wrong population.

**Record #203 excluded at full-text screening**

Seaman B, Eldridge L. No-nonsense guide to menopause. 1st Simon & Schuster hardcover ed. New York (NY): Simon & Schuster; 2008. 482 p.

**Rationale for exclusion at full-text screening**

Book.

**Record #204 excluded at full-text screening**

Doress-Worters PB, Siegal DL. New ourselves, growing older: women aging with knowledge and power. New York (NY): Simon & Schuster; 1994. 549 p.

**Rationale for exclusion at full-text screening**

Book.

**Record #205 excluded at full-text screening**

Gower T. New answers to menopause? Health (Time Inc. Health). 2002 Jan-Feb;16(1):76.

**Rationale for exclusion at full-text screening**

Wrong population.

**Record #206 excluded at full-text screening**

Kagan L, Kessel B, Benson H. Mind over menopause: the complete mind/body approach to coping with menopause. 1st Free Press trade pbk. ed. New York (NY): Free Press; 2004. 363 p.

**Rationale for exclusion at full-text screening**

Book.

**Record #207 excluded at full-text screening**

**International Council for Health Freedom.** Menopause: black cohosh gets an iffy okay. Int Counc Health Freedom Newsl. 2003 Summer;7(1):16.

**Rationale for exclusion at full-text screening**

Wrong record type.

**Record #208 excluded at full-text screening**

McPhail G. Menopause as an issue for women with breast cancer. Cancer Nurs. 1999;**22**(2):164–71.

**Rationale for exclusion at full-text screening**

Wrong population.

**Record #209 excluded at full-text screening**

Brown L. Menopause. Better Nutr. 2001 Jan;**63**(1):58.

**Rationale for exclusion at full-text screening**

Wrong population.

**Record #210 excluded at full-text screening**

**Menopause [video].** Boston (MA): Fanlight Productions; 1996.

**Rationale for exclusion at full-text screening**

Wrong record type.

**Record #211 excluded at full-text screening**

**Weed SS.** Menopausal years: the wise woman way. Woodstock (NY): Ash Tree Pub; 1992. 222 p. (Wise woman ways).

**Rationale for exclusion at full-text screening**

Book.

**Record #212 excluded at full-text screening**

**Thompson JJ.** Managing menopause: an ethnographic study of women's midlife information-seeking and decision-making in the Southwest US [dissertation]. Tucson (AZ): University of Arizona; 2011. Available from: ProQuest Dissertations & Theses Global.

**Rationale for exclusion at full-text screening**

Unstratified results (population).

**Record #213 excluded at full-text screening**

**Caine-Francis D.** Managing menopause beautifully: physically, emotionally, and sexually. Westport (CT): Praeger; 2008. 206 p. (Sex, love, and psychology).

**Rationale for exclusion at full-text screening**

Book.

**Record #214 excluded at full-text screening**

**Cone FK.** Making sense of menopause: over 150 women and experts share their wisdom, experience, and commonsense advice. New York (NY): Simon & Schuster; 1993. 384 p.

**Rationale for exclusion at full-text screening**

Book.

**Record #215 excluded at full-text screening**

**Allsworth JE, Zierler S, Lapane KL, Krieger N, Hogan JW, Harlow BL.** Longitudinal study of the inception of perimenopause in relation to lifetime history of sexual or physical violence. J Epidemiol Community Health.2004;**58**(11):938–943.

**Rationale for exclusion at full-text screening**

Unclear population.

**Record #216 excluded at full-text screening**

**Ciano C, King TS, Wright RR, Perlis M, Sawyer AM.** Longitudinal study of insomnia symptoms among women during perimenopause. J Obstet Gynecol Neonatal Nurs. 2017;**46**(6):804–813.

**Rationale for exclusion at full-text screening**

Wrong population.

**Record #217 excluded at full-text screening**

**Wise LA, Krieger N, Zierler S, Harlow BL.** Lifetime socioeconomic position in relation to onset of perimenopause. J Epidemiol Community Health. 2002;**56**(11):851–860.

**Rationale for exclusion at full-text screening**

Unclear population.

**Record #218 excluded at full-text screening**

**Bradfield CD, Hinkle D.** "Let's celebrate aging": a case study of a community wide educational program. Gerontol Geriatr Educ. 1997;**17**(3):87–92.

**Rationale for exclusion at full-text screening**

Unclear population.

**Record #219 excluded at full-text screening**

**Lyon J.** The Indian elder, a forgotten American. Final report on the first National Indian Conference on Aging; Phoenix, Arizona; June 15–17, 1976. Washington (DC): National Tribal Chairmen's Association; 1978.

**Rationale for exclusion at full-text screening**

Wrong concept.

**Record #220 excluded at full-text screening**

**Collings P.** 'If you got everything, it's good enough': Perspectives on successful aging in a Canadian Inuit community. J Cross Cult Gerontol. 2001;**16**(2):127–155.

**Rationale for exclusion at full-text screening**

Wrong concept.

**Record #221 excluded at full-text screening**

Schwartz E. Hormone solution: naturally alleviate symptoms of hormone imbalance from adolescence through menopause. New York (NY): Warner Books; 2002. 280 p.

**Rationale for exclusion at full-text screening**

Book.

**Record #222 excluded at full-text screening**

Kamen B. Hormone replacement therapy yes or no?: how to make an informed decision about estrogen, progesterone, & other strategies for dealing with PMS, menopause, & osteoporosis. Novato (CA): Nutrition Encounter; 1993. 271 p.

**Rationale for exclusion at full-text screening**

Book.

**Record #223 excluded at full-text screening**

Napoli M. Holistic health care for native women: an integrated model. Am J Public Health. 2002;**92**(10):1573–5.

**Rationale for exclusion at full-text screening**

Wrong concept.

**Record #224 excluded at full-text screening**

John R, Blanchard PH, Hennessy CH, Coyle JM. Hidden lives: aging and contemporary American Indian women. In: Coyle JM, editor. Handbook on Women and Aging. Westport (CT): Greenwood Press; 1997. p. 290–315.

**Rationale for exclusion at full-text screening**

Wrong concept.

**Record #225 excluded at full-text screening**

Shulman N, Kim ES. Healthy transitions: a woman's guide to perimenopause, menopause and beyond. Amherst (NY): Prometheus Books; 2004. 354 p.

**Rationale for exclusion at full-text screening**

Book.

**Record #226 excluded at full-text screening**

Avis NE, McKinlay SM, Flint M, Kronenberg F, Utian W. Health-care utilization among mid-age women. In: Korenman SG, editor. Multidisciplinary perspectives on menopause. Vol. 592. New York (NY): Annals of the New York Academy of Sciences; 1990. p. 228–38.

**Rationale for exclusion at full-text screening**

Unclear population.

**Record #227 excluded at full-text screening**

Ward EM. Health concerns at menopause: HRT vs. natural remedies for relief. Environ Nutr. 2002 Jan;**25**(1):1.

**Rationale for exclusion at full-text screening**

Wrong population.

**Record #228 excluded at full-text screening**

Kaufert PA. Health and social profile of the menopausal woman. Exp Gerontol. 1994;**29**(3-4):343–350.

**Rationale for exclusion at full-text screening**

Wrong population.

**Record #229 excluded at full-text screening**

Harvard Women's Health Watch. Forecasting hot flashes. Harv Womens Health Watch. 1996 Dec;**4**(4):1.

**Rationale for exclusion at full-text screening**

Wrong population.

**Record #230 excluded at full-text screening**

Hudnall M. Finding natural ways to minimize the side effects of menopause [cover story]. Environ Nutr. 1998 Aug;**21**(8):1.

**Rationale for exclusion at full-text screening**

Wrong population.

**Record #231 excluded at full-text screening**

Dugger K, Shoemake K. Faye’s Story: Life’s Circle. IHS Prim Care Provid. 2010;**35**(5):125–6.

**Rationale for exclusion at full-text screening**

Wrong concept.

**Record #232 excluded at full-text screening**

Dukepoo, F. C. (1980). The elder American Indian: A cross-cultural study of minority elders in San Diego. University Center on Aging, San Diego State University, San Diego, CA.

**Rationale for exclusion at full-text screening**

Book.

**Record #233 excluded at full-text screening**

**Shams T, Setia MS, Hemmings R, McCusker J, Sewitch M, Ciampi A.** Efficacy of black cohosh-containing preparations on menopausal symptoms: a meta-analysis. Altern Ther Health Med. 2010 Jan-Feb;**16**(1):36–44.

**Rationale for exclusion at full-text screening**

Wrong population.

**Record #234 excluded at full-text screening**

**Soares CN, Taylor V.** Effects and management of the menopausal transition in women with depression and bipolar disorder. J Clin Psychiatry. 2007;**68**(Suppl 9):16–21.

**Rationale for exclusion at full-text screening**

Wrong population.

**Record #235 excluded at full-text screening**

**Chrisler JC, Hemstreet AH.** The diversity of women's health needs. In: Chrisler JC, Hemstreet AH, editors. Variations on a theme: Diversity and the psychology of women. Dubuque (IA): Kendall/Hunt Publishing Company; 1995. p. 1–28.

**Rationale for exclusion at full-text screening**

Wrong concept.

**Record #236 excluded at full-text screening**

**Speakman L.** Discourse on menopause: investigation of women's knowledge about menopause [dissertation]. Ann Arbor (MI): University of Colorado; 1999. Available from: ProQuest Dissertations & Theses.

**Rationale for exclusion at full-text screening**

Wrong population.

**Record #237 excluded at full-text screening**

**Riley EH, Inui TS, Kleinman K, Connelly MT.** Differential association of modifiable health behaviors with hot flashes in perimenopausal and postmenopausal women. J Gen Intern Med. 2004;**19**(7):740–6.

**Rationale for exclusion at full-text screening**

Wrong population.

**Record #238 excluded at full-text screening**

**Anonymous.** Cohosh update. Vegetarian Times. 2003 Nov;(315):14.

**Rationale for exclusion at full-text screening**

Wrong population.

**Record #239 excluded at full-text screening**

**Inglis S, Stout DR.** Cherokee Elder Care explores quality of life—for PACE participants. Geriatr Nurs (Minneap). 2012;**37**(2):119–20.

**Rationale for exclusion at full-text screening**

Wrong concept.

**Record #240 excluded at full-text screening**

**Greer G.** Change: women, aging and the menopause. New York (NY): A.A. Knopf; 1992. 422 p.

**Rationale for exclusion at full-text screening**

Book.

**Record #241 excluded at full-text screening**

**McGraw K.** Care for the aged—a tribal feeling. New Mexico Mag. 1980 Jan:32–3.

**Rationale for exclusion at full-text screening**

Wrong concept.

**Record #242 excluded at full-text screening**

**Murray MT.** Black cohosh: nature's best solution for hot flashes. Alive Can Nat Health Wellness Mag. 2004 Jul;(261):48–51.

**Rationale for exclusion at full-text screening**

Wrong population.

**Record #243 excluded at full-text screening**

**American Botanical Council.** Black cohosh safety. HerbalGram. 2006 Sep–Dec;(72):4.

**Rationale for exclusion at full-text screening**

Wrong population.

**Record #244 excluded at full-text screening**

**Ackerson AD.** Black cohosh. Better Nutr. 2005 Oct;**67**(10):18.

**Rationale for exclusion at full-text screening**

Wrong record type.

**Record #245 excluded at full-text screening**

**Gittleman AL.** Before the change: taking charge of your perimenopause. 2nd rev. & updated ed. San Francisco (CA): Harper San Francisco; 2003. 295 p.

**Rationale for exclusion at full-text screening**

Book.

**Record #246 excluded at full-text screening**

**Joffe H, Soares CN, Cohen LS.** Assessment and treatment of hot flushes and menopausal mood disturbance. Psychiatr Clin North Am. 2003;**26**(3):563–580.

**Rationale for exclusion at full-text screening**

Wrong population.

**Record #247 excluded at full-text screening**

**Ho C.** Ask the experts. Is black cohosh safe to use for PMS or menopause if breast cancer runs in my family? Alive. 2014 Jul;(381):20.

**Rationale for exclusion at full-text screening**

Wrong record type.

**Record #248 excluded at full-text screening**

**Boothe-Kepple S.** American Indian women's health perceptions and health promotion behavior [dissertation]. San Diego (CA): University of San Diego; 2016. Available from: ProQuest Information & Learning.

**Rationale for exclusion at full-text screening**

Wrong concept.

**Record #249 excluded at full-text screening**

**John R.** American Indian and Alaska Native elders: an assessment of their current status and provision of services. Washington (DC): US Department of Health and Human Services, Public Health Service, Indian Health Service, Office of Health Programs; 1994.

**Rationale for exclusion at full-text screening**

Wrong concept.

**Record #250 excluded at full-text screening**

Cheung T, Balen AH. All-natural menopause diet: the drug-free, natural way to beat your symptoms and lose weight. 1st Pegasus Books ed. New York (NY): Pegasus Books; 2008. 229 p.

**Rationale for exclusion at full-text screening**

Book.

**Record #251 excluded at full-text screening**

Collings P. Aging and life course development in an Inuit community. Arctic Anthropol. 2000;**37**(2):111–125.

**Rationale for exclusion at full-text screening**

Wrong concept.

**Record #252 excluded at full-text screening**

Baldridge D. Advocating for Native American elders. IHS Prim Care Provider. 1993;**18**(5):86–90.

**Rationale for exclusion at full-text screening**

Wrong concept.

**Record #253 excluded at full-text screening**

Stout MD, Kipling GD, Stout R. Aboriginal women's health research. Netw Mag Can Women’s Health Netw. 2001;4/5(4/1):11–12.

**Rationale for exclusion at full-text screening**

Wrong concept.

**Record #254 excluded at full-text screening**

Nettles SM. Aging women of color: Engagement and place. Women Ther. 2016 Jul;39(3-4):337–53.

**Rationale for exclusion at full-text screening**

Wrong concept.

**Record #255 excluded at full-text screening**

Stenchever MA. Alternative therapies for menopause symptoms. ACOG Clin Rev. 2002;**7**(9):10.

**Rationale for exclusion at full-text screening**

Unclear population.

**Record #256 excluded at full-text screening**

Buxton-Blake PL. Alternative therapies selected by women for menopausal symptomatology [dissertation]. Washington (DC): Catholic University of America; 2001. 179 p.

**Rationale for exclusion at full-text screening**

Unclear population.

**Record #257 excluded at full-text screening**

**Antidepressant eyed to reduce hot flashes.** Contracept Technol Update. 2011;**32**(5):56–7.

**Rationale for exclusion at full-text screening**

Unclear population.

**Record #258 excluded at full-text screening**

Forbes A, Ritchie S, Walker J, Young N. Applications of Two-Eyed Seeing in primary research focused on Indigenous health: a scoping review. Int J Qual Methods. 2020 Jun;19:18.

**Rationale for exclusion at full-text screening**

Wrong concept.

**Record #259 excluded at full-text screening**

Kling JM, Kelly M, Rullo J, Kapoor E, Kuhle CL, Vegunta S, et al. Association between menopausal symptoms and relationship distress. Maturitas. 2019;**130**:1–5.

**Rationale for exclusion at full-text screening**

Unclear population.

**Record #260 excluded at full-text screening**

Betz JM, Anderson L, Avigan MI, Barnes J, Farnsworth NR, Gerdén B, et al. Black cohosh: consideration of safety and benefit. Nutr Today. 2009;**44**(4):155–62.

**Rationale for exclusion at full-text screening**

Wrong population.

**Record #261 excluded at full-text screening**

Geller SE, Studee L. Botanical and dietary supplements for mood and anxiety in menopausal women. Menopause. 2007;**14**(3 Pt 1):541–9.

**Rationale for exclusion at full-text screening**

Unstratified results (population).

**Record #262 excluded at full-text screening**

**Hudson T.** Botanicals for managing menopause-related symptoms: state of the science. Integr Med (Encinitas). 2009;**8**(6):30–7.

**Rationale for exclusion at full-text screening**

Wrong population.

**Record #263 excluded at full-text screening**

**Tonob D, Melby MK.** Broadening our perspectives on complementary and alternative medicine for menopause: a narrative review. Maturitas. 2017 May;**99**:79–85.

**Rationale for exclusion at full-text screening**

Wrong population.

**Record #264 excluded at full-text screening**

**[No author listed.]** A call for action in Indigenous and First Nations women's health and wellbeing. FIGO Newsletter. 2021 Oct;(1):1.

**Rationale for exclusion at full-text screening**

Wrong record type.

**Record #265 excluded at full-text screening**

**[No author listed.]** Canada needs a health and healing strategy for First Nations, Inuit and Métis women: health system fails most Aboriginal women across the lifespan. Network Mag Can Women’s Health Netw. 2005 Fall/Winter;**8**(1/2):30–1.

**Rationale for exclusion at full-text screening**

Wrong concept.

**Record #266 excluded at full-text screening**

**Penson RT, Sahani D, Bell DA.** Case records of the Massachusetts General Hospital. Case 37-2004. A 52-year-old woman with postmenopausal bleeding and a cystic ovarian mass. N Engl J Med. 2004;**351**(24):2531–8.

**Rationale for exclusion at full-text screening**

Unclear population.

**Record #267 excluded at full-text screening**

**Jaramillo ET, Sommerfeld DH, Haozous EA, Brunner A, Willging CE.** Causes and consequences of not having a personal healthcare provider among American Indian elders: a mixed-method study. Front Public Health. 2022 Mar;**10**:9.

**Rationale for exclusion at full-text screening**

Wrong concept.

**Record #268 excluded at full-text screening**

**Markham R, Hunt M, Huebert S, Graham S.** Co-planning for collaborative rural and First Nations health care system transformation: developing a pattern for partnership and social accountability. BC Med J. 2021 Dec;**63**(10):416–21.

**Rationale for exclusion at full-text screening**

Wrong concept.

**Record #269 excluded at full-text screening**

**Arcury TA, Quandt SA, Bell RA, Vitolins MZ.** Complementary and alternative medicine use among rural older adults. Complement Health Pract Rev. 2002 Spring–Summer;**7**(3):167–86.

**Rationale for exclusion at full-text screening**

Wrong concept.

**Record #270 excluded at full-text screening**

**Galantino ML, Canella C, House L, Kondos L, Suydam A, Doran J, et al.** Complementary and alternative therapies for women transitioning through menopause. J Womens Health Phys Ther. 2006 Winter;**30**(3):18–26.

**Rationale for exclusion at full-text screening**

Wrong population.

**Record #271 excluded at full-text screening**

**Son JS, Hutchinson SL.** Conceptualizing leisure self-care in an exploratory study of American Indian elders’ health beliefs and behaviours. Leisure Loisir. 2009;**33**(2):479–509.

**Rationale for exclusion at full-text screening**

Wrong concept.

**Record #272 excluded at full-text screening**

**Kielich AM, Miller L.** Cultural aspects of women's health care. Patient Care. 1996;**30**(16):60–76.

**Rationale for exclusion at full-text screening**

Wrong concept.

**Record #273 excluded at full-text screening**

**Moffitt P, Vollman AR.** Designer's corner. Photovoice: picturing the health of Aboriginal women in a remote northern community. Can J Nurs Res. 2004;**36**(4):189–201.

**Rationale for exclusion at full-text screening**

Wrong concept.

**Record #274 excluded at full-text screening**

**Jull J, Stacey D, Giles A, Boyer Y, Lodge M.** Developing culturally-appropriate interventions: exploring First Nations, Métis and Inuit women's health decision-making needs. Int J Qual Methods. 2012;**11**(5):832–3.

**Rationale for exclusion at full-text screening**

Wrong concept.

**Record #275 excluded at full-text screening**

**Liddell JL, McKinley CE.** The development of the Framework of Integrated Reproductive and Sexual Health Theories (FIRSHT) to contextualize Indigenous women's health experiences. Sex Res Soc Policy. 2022 Sep;**19**(3):1020–33.

**Rationale for exclusion at full-text screening**

Wrong concept.

**Record #276 excluded at full-text screening**

**Speakman L.** Discourse on menopause: An investigation of women's knowledge about menopause [dissertation]. Denver (CO): University of Colorado Health Sciences Center; 1999. 122 p.

**Rationale for exclusion at full-text screening**

Wrong population.

**Record #277 excluded at full-text screening**

**Harlow BL, Maclehose RF, Smolenski DJ, Soares CN, Otto MW, Joffe H, et al.** Disparate rates of new-onset depression during the menopausal transition in 2 community-based populations: real, or really wrong? Am J Epidemiol. 2013;**177**(10):1148–56.

**Rationale for exclusion at full-text screening**

Unclear population.

**Record #278 excluded at full-text screening**

**Moon H, Roh S, Lee YS, Goins RT.** Disparities in health, health care access, and life experience between American Indian and white adults in South Dakota. J Racial Ethn Health Disparities. 2015;**3**(2):301–8.

**Rationale for exclusion at full-text screening**

Wrong concept.

**Record #279 excluded at full-text screening**

**Labrie F, Derogatis L, Archer DF, Koltun W, Vachon A, Young D, et al.** Effect of intravaginal prasterone on sexual dysfunction in postmenopausal women with vulvovaginal atrophy. J Sex Med. 2015 Dec;**12**(12):2401–12.

**Rationale for exclusion at full-text screening**

Unstratified results (population).

**Record #280 excluded at full-text screening**

**Johnston JL, Schneider LS.** Effects of a one-month nutrition and exercise program in Blackfoot women elders. J Can Diet Assoc. 1988 Fall;**49**(4):249–53.

**Rationale for exclusion at full-text screening**

Wrong concept.

**Record #281 excluded at full-text screening**

**Ryan-Borchers T, Chew B, Park JS, McGuire M, Fournier L, Beerman K.** Effects of dietary and supplemental forms of isoflavones on thyroid function in healthy postmenopausal women. Top Clin Nutr. 2008;**23**(1):13–22.

**Rationale for exclusion at full-text screening**

Unstratified results (population).

**Record #282 excluded at full-text screening**

Langford J, Doughty A, Wang M, Clayton L, Babich M. Effects of Morinda citrifolia on quality of life and auditory function in postmenopausal women. *J Altern Complement Med*. 2004;**10**(5):737–9.

**Rationale for exclusion at full-text screening**

Unclear population.

**Record #283 excluded at full-text screening**

Woods NF. Entering menopause increases the risk of first episode depression. *Evid Based Ment Health*. 2006;**9**(4):109.

**Rationale for exclusion at full-text screening**

Unclear population.

**Record #284 excluded at full-text screening**

Echeverria V, Echeverria F, Barreto GE, Echeverría J, Mendoza C. Estrogenic plants: to prevent neurodegeneration and memory loss and other symptoms in women after menopause. *Front Pharmacol*. 2021;**12**:644103.

**Rationale for exclusion at full-text screening**

Wrong population.

**Record #285 excluded at full-text screening**

Kupferer EM. An exploration of women's current hormone discontinuation experiences, influences, decisions, and alternatives [dissertation]. Austin (TX): University of Texas at Austin; 2007. 150 p.

**Rationale for exclusion at full-text screening**

Unstratified results (population).

**Record #286 excluded at full-text screening**

Hughes AK, Lewinson TDW. Facilitating communication about sexual health between aging women and their health care providers. *Qual Health Res*. 2015;**25**(4):540–50.

**Rationale for exclusion at full-text screening**

Unstratified results (population).

**Record #287 excluded at full-text screening**

Newton KM, Reed SD, Nekhlyudov L, Grothaus LC, Ludman EJ, Ehrlich K, et al. Factors associated with successful discontinuation of hormone therapy. *J Womens Health (Larchmt)*. 2014;**23**(5):382–8.

**Rationale for exclusion at full-text screening**

Unstratified results (population).

**Record #288 excluded at full-text screening**

North KE, MacCluer JW, Cowan LD, Howard BV. Fertility in post-menopausal American Indian women: The Strong Heart Study [abstract]. *Am J Phys Anthropol*. 1999:212.

**Rationale for exclusion at full-text screening**

Wrong concept.

**Record #289 excluded at full-text screening**

Browne AJ. First Nations women and health care services: the sociopolitical context of encounters with nurses[dissertation]. Vancouver (BC): University of British Columbia (Canada); 2003. 285 p.

**Rationale for exclusion at full-text screening**

Wrong concept.

**Record #290 excluded at full-text screening**

Nichols HB, Trentham-Dietz A, Hampton JM, Titus-Ernstoff L, Egan KM, Willett WC, et al. From menarche to menopause: trends among US women born from 1912 to 1969. Am J Epidemiol. 2006;**164**(10):1003–11.

**Rationale for exclusion at full-text screening**

Unclear population.

**Record #291 excluded at full-text screening**

Walters KL, Simoni JM. Future health needs of women of color. Reconceptualizing Native women's health: an 'indigenist' stress-coping model. Am J Public Health. 2002;**92**(4):520–4.

**Rationale for exclusion at full-text screening**

Wrong concept.

**Record #292 excluded at full-text screening**

Burnett K. The healing work and nursing care of Aboriginal women, female medical missionaries, nursing sisters, public health nurses, and female attendants in southern Alberta First Nations communities, 1880—1930 [dissertation]. Toronto (ON): York University (Canada); 2006. 280 p.

**Rationale for exclusion at full-text screening**

Wrong concept.

**Record #293 excluded at full-text screening**

Hannis P. The health of Native women in Quebec. Network Mag Can Womens Health Netw. 2001 Fall/Winter;**4/5**(4/1):13–15.

**Rationale for exclusion at full-text screening**

Wrong concept.

**Record #294 excluded at full-text screening**

Oosman S, Nisbet C, Smith L, Abonyi S. Health promotion interventions supporting Indigenous healthy ageing: a scoping review. Int J Circumpolar Health. 2021 Jan;**80**(1):27.

**Rationale for exclusion at full-text screening**

Wrong concept.

**Record #295 excluded at full-text screening**

Herbal remedies for menopausal symptoms. Nurses' Drug Alert. 2007;**31**(2):11.

**Rationale for exclusion at full-text screening**

Unclear population.

**Record #296 excluded at full-text screening**

Weil VP, Cirigliano MD, Battistini M. Herbal treatments for symptoms of menopause. Hosp Physician. 2000;**36**(11):35–44.

**Rationale for exclusion at full-text screening**

Wrong population.

**Record #297 excluded at full-text screening**

NIH Office on Women's Health. Highlights from Women of Color Health Data Book, Fourth Edition. J Womens Health (Larchmt). 2014;**23**(10):785–6.

**Rationale for exclusion at full-text screening**

Wrong concept.

**Record #298 excluded at full-text screening**

Braun KL, LaCounte C. The historic and ongoing issue of health disparities among Native elders. Generations. 2014;**38**(4):60–9.

**Rationale for exclusion at full-text screening**

Wrong concept.

**Record #299 excluded at full-text screening**

Watt PJ, Hughes RB, Rettew LB, Adams R. A holistic programmatic approach to natural hormone replacement. Fam Community Health. 2003 Jan–Mar;**26**(1):53–63.

**Rationale for exclusion at full-text screening**

Wrong population.

**Record #300 excluded at full-text screening**

**Aker A, Serghides L, Cotnam J, Jackson R, Robinson M, Gauvin H, et al.** The impact of a stress management intervention including cultural components on stress biomarker levels and mental health indicators among Indigenous women. J Behav Med. 2023;**46**:594–608.

**Rationale for exclusion at full-text screening**

Wrong concept.

**Record #301 excluded at full-text screening**

Allen L, Hatala A, Ijaz S, Courchene ED, Bushie EB. Indigenous-led health care partnerships in Canada. *Can Med Assoc J*. 2020;**192**(9):E208-E216.

**Rationale for exclusion at full-text screening**

Wrong concept.

**Record #302 excluded at full-text screening**

**Kornstein SG, Young EA, Harvey AT, Wisniewski SR, Barkin JL, Thase ME, et al.** The influence of menopause status and postmenopausal use of hormone therapy on presentation of major depression in women. Menopause. 2010;**17**(4):828-839.

**Rationale for exclusion at full-text screening**

Unclear population.

**Record #303 excluded at full-text screening**

**Suter E, Verhoef MJ, Bockmuehl C, Forest N, Bobey M, Armitage GD.** Inquiring minds: women's approaches to evaluating complementary and alternative therapies for menopausal symptoms. Can Fam Physician. 2007;**53**(1):85-90.

**Rationale for exclusion at full-text screening**

Unclear population.

**Record #304 excluded at full-text screening**

**Rand JR.** Inuit women's stories of strength: informing Inuit community-based HIV and STI prevention and sexual health promotion programming. Int J Circumpolar Health. 2016;**75**:7.

**Rationale for exclusion at full-text screening**

Wrong concept.

**Record #305 excluded at full-text screening**

**Avis NE, Stellato R, Crawford S, Johannes C, Longcope C.** Is there an association between menopause status and sexual functioning? Menopause. 2000;**7**(5):297-309.

**Rationale for exclusion at full-text screening**

Unclear population.

**Record #306 excluded at full-text screening**

**Hidaka T, Yonezawa R, Saito S.** Kami-shoyo-san, Kampo (Japanese traditional medicine), is effective for climacteric syndrome, especially in hormone-replacement-therapy-resistant patients who strongly complain of psychological symptoms. J Obstet Gynaecol Res. 2013 Jan;**39**(1):223-228.

**Rationale for exclusion at full-text screening**

Unclear population.

**Record #307 excluded at full-text screening**

**Appling SE, Allen JK, Van Zandt S, Olsen S, Brager R, Hallerdin J.** Knowledge of menopause and hormone replacement therapy use in low-income urban women. J Womens Health Gend Based Med. 2000;**9**(1):57-64.

**Rationale for exclusion at full-text screening**

Unstratified results (population).

**Record #308 excluded at full-text screening**

**Johannes CB, Crawford SL, Posner JG, McKinlay SM.** Longitudinal patterns and correlates of hormone replacement therapy use in middle-aged women. Am J Epidemiol. 1994;**140**(5):439-52.

**Rationale for exclusion at full-text screening**

Unclear population.

**Record #309 excluded at full-text screening**

**Crawford SL, Casey VA, Avis NE, McKinlay SM.** A longitudinal study of weight and the menopause transition: results from the Massachusetts Women's Health Study. Menopause. 2000;**7**(2):96-104.

**Rationale for exclusion at full-text screening**

Unclear population.

**Record #310 excluded at full-text screening**

**Leidy LE, Canali C, Callahan WE.** The medicalization of menopause: implications for recruitment of study participants. Menopause. 2000;**7**(3):193-199.

**Rationale for exclusion at full-text screening**

Unclear population.

**Record #311 excluded at full-text screening**

**Popivanov P.** Menopausal indices as criteria for the effectiveness of acupuncture treatment of the climacteric syndrome (preliminary report). Internal Diseases. 1983;**22**(6):110-113.

**Rationale for exclusion at full-text screening**

Non-English record.

**Record #312 excluded at full-text screening**

**George SA.** The menopause experience: a woman's perspective. JOGNN J Obstet Gynecol Neonatal Nurs. 2002;**31**(1):77-85.

**Rationale for exclusion at full-text screening**

Wrong population.

**Record #313 excluded at full-text screening**

**Challem J.** Menopause made easier. Delicious Living. 2013;**29**(10):51-2.

**Rationale for exclusion at full-text screening**

Wrong population.

**Record #314 excluded at full-text screening**

**Whitford L, Kunz K.** The Montana American Indian Women's Health Coalition is making a difference statewide with women's health. Pulse. 2003;**40**(1):13.

**Rationale for exclusion at full-text screening**

Wrong concept.

**Record #315 excluded at full-text screening**

**McKinlay SM, Brambilla DJ, Posner JG.** The normal menopause transition. Maturitas. 2008;**61**(1-2):4-16.

**Rationale for exclusion at full-text screening**

Unclear population.

**Record #316 excluded at full-text screening**

**Walker WA.** Papers from the Nutrition and Women's Health Symposium, June 14-15, 2002, Division of Nutrition, Harvard Medical School, Boston, Massachusetts. J Womens Health (Larchmt). 2003;**12**(2):105-192.

**Rationale for exclusion at full-text screening**

Wrong population.

**Record #317 excluded at full-text screening**

**Burhansstipanov L, Bad Wound D, Capelouto N, Goldfarb F, Harjo L, Hatathlie L, et al.** Patient resources. Culturally relevant 'navigator' patient support: the Native Sisters. Cancer Pract. 1998;**6**(3):191-194.

**Rationale for exclusion at full-text screening**

Wrong concept.

**Record #318 excluded at full-text screening**

**Carlson ES.** Perceptions, relationships, societal contexts, and previous decisions influenced women's decisions about hormone replacement therapy [commentary on Marmoreo J, Brown JB, Batty HR, et al. Hormone replacement therapy: determinants of women's decisions. Patient Educ Couns. 1998;33(3):289-98]. Evid Based Nurs. 1999;(1):30.

**Rationale for exclusion at full-text screening**

Unclear population.

**Record #319 excluded at full-text screening**

**Smolinski D, Wollner D, Orlowski J, Curcio J, Nevels J, Kim LS.** A pilot study to examine a combination botanical for the treatment of menopausal symptoms. J Altern Complement Med. 2005;**11**(3):483-9.

**Rationale for exclusion at full-text screening**

Unclear population.

**Record #320 excluded at full-text screening**

**Brooks-Cleator LA, Giles AR.** A postcolonial discourse analysis of community stakeholders' perspectives on supporting urban Indigenous older adults to age well in Ottawa, Canada. Int Indig Policy J. 2020 Jan;**11**(1):21.

**Rationale for exclusion at full-text screening**

Wrong concept.

**Record #321 excluded at full-text screening**

**Hamzelou J.** Pregnancy may affect menopause. New Sci. 2020;**245**(3267):18.

**Rationale for exclusion at full-text screening**

Unclear population.

**Record #322 excluded at full-text screening**

**Proceedings of the Annual Meeting of the International Society for the Study of Women's Sexual Health, October 25–28, 2002, Boston, Massachusetts, USA.** J Sex Marital Ther. 2002;**28**(suppl):1-244.

**Rationale for exclusion at full-text screening**

Unclear population.

**Record #323 excluded at full-text screening**

Slattery ML, Schumacher MC, Lanier AP, Edwards S, Edwards R, Murtaugh MA, et al. A prospective cohort of American Indian and Alaska Native people: Study design, methods, and implementation. Am J Epidemiol. 2007 Sep;**166**(5):606-615.

**Rationale for exclusion at full-text screening**

Wrong concept.

**Record #324 excluded at full-text screening**

Zhang C, Li L, Jiang Y. Psychotherapy and traditional Chinese medicine for perimenopausal depression: A literature review. J Chin Integr Med. 2009;**7**(8):779-783.

**Rationale for exclusion at full-text screening**

Non-English record.

**Record #325 excluded at full-text screening**

Stoffle RW, Halmo DB, Evans MJ. Puchuxwavaats uapi (To know about plants): Traditional knowledge and the cultural significance of Southern Paiute plants. Hum Organ. 1999;**58**(4):416-429.

**Rationale for exclusion at full-text screening**

Wrong concept.

**Record #326 excluded at full-text screening**

Booth NL, Piersen CE. Red clover (Trifolium pratense). In: Coates PM, Blackman MR, Cragg GM, Levine M, Moss J, White JD, editors. Encyclopedia of dietary supplements. New York: Marcel Dekker; 2004. p. 587-602.

**Rationale for exclusion at full-text screening**

Wrong concept.

**Record #327 excluded at full-text screening**

Hatala AR, Desjardins M, Bombay A. Reframing narratives of Aboriginal health inequity: exploring Cree elder resilience and well-being in contexts of historical trauma. *Qual Health Res*. 2016;**26**(14):1911-1927.

**Rationale for exclusion at full-text screening**

Wrong concept.

**Record #328 excluded at full-text screening**

Feldman K, Szappanos A, Butz H, Grolmusz V, Majnik J, Liko I, et al. The rs4844880 polymorphism in the promoter region of the HSD11B1 gene associates with bone mineral density in healthy and postmenopausal osteoporotic women. *Steroids*. 2012 Nov;**77**(13):1345-1351.

**Rationale for exclusion at full-text screening**

Wrong population.

**Record #329 excluded at full-text screening**

Tagliaferri MA, Tagliaferri MC, Creasman JM, Koltun WD. A selective estrogen receptor beta agonist for the treatment of hot flushes: Phase 2 clinical trial. *J Altern Complement Med*. 2016;**22**(9):722-728.

**Rationale for exclusion at full-text screening**

Wrong population.

**Record #330 excluded at full-text screening**

Kripke DF, Brunner R, Freeman R, Hendrix SL, Jackson RD, Masaki K, et al. Sleep complaints of postmenopausal women. *Clin J Womens Health*. 2001;**1**(5):244-252.

**Rationale for exclusion at full-text screening**

Unstratified results (population).

**Record #331 excluded at full-text screening**

Burke GL, Legault C, Anthony M, Bland DR, Morgan TM, Naughton MJ, et al. Soy protein and isoflavone effects on vasomotor symptoms in peri- and postmenopausal women: the Soy Estrogen Alternative Study. *Menopause*. 2003 Mar-Apr;**10**(2):147-153.

**Rationale for exclusion at full-text screening**

Unstratified results (population).

**Record #332 excluded at full-text screening**

Park J, Linde K, Manheimer E, Molsberger A, Sherman K, Smith C, et al. The status and future of acupuncture clinical research. *J Altern Complement Med*. 2008;**14**(7):871-881.

**Rationale for exclusion at full-text screening**

Wrong population.

**Record #333 excluded at full-text screening**

Dinges NG, Joos SK. Stress, coping, and health: models of interaction for Indian and native populations. *Am Indian Alsk Native Ment Health Res Monogr Ser*. 1988;**1**:8-64.

**Rationale for exclusion at full-text screening**

Wrong concept.

**Record #334 excluded at full-text screening**

Lewis JP. Successful aging through the eyes of Alaska Native elders: what it means to be an elder in Bristol Bay, AK. *Gerontologist*. 2011;**51**(4):540-549.

**Rationale for exclusion at full-text screening**

Wrong concept.

**Record #335 excluded at full-text screening**

Sievert LL, Obermeyer CM. Symptom clusters at midlife: a four-country comparison of checklist and qualitative responses. *Menopause*. 2012;**19**(2):133-144.

**Rationale for exclusion at full-text screening**

Unclear population.

**Record #336 excluded at full-text screening**

Obermeyer CM, Reynolds RF, Price K, Abraham A. Therapeutic decisions for menopause: results of the DAMES project in central Massachusetts. *Menopause*. 2004;**11**(4):456-465.

**Rationale for exclusion at full-text screening**

Unstratified results (population).

**Record #337 excluded at full-text screening**

Stephenson K, Neuenschwander PF, Kurdowska AK, Pinson B, Price C. Transdermal progesterone: effects on menopausal symptoms and on thrombotic, anticoagulant, and inflammatory factors in postmenopausal women. *Int J Pharm Compd*. 2008;**12**(4):294-304.

**Rationale for exclusion at full-text screening**

Unstratified results (population).

**Record #338 excluded at full-text screening**

Gordon NP, Sobel DS, Tarazona EZ. Use of and interest in alternative therapies among adult primary care clinicians and adult members in a large health maintenance organization. *West J Med*. 1998;**169**(3):153-161.

**Rationale for exclusion at full-text screening**

Wrong population.

**Record #339 excluded at full-text screening**

Bair YA, Gold EB, Azari RA, Greendale G, Sternfeld B, Harkey MR, et al. Use of conventional and complementary health care during the transition to menopause: longitudinal results from the Study of Women's Health Across the Nation (SWAN). *Menopause*. 2005;**12**(1):31-39.

**Rationale for exclusion at full-text screening**

Wrong population.

**Record #340 excluded at full-text screening**

Kelly JP, Kaufman DW, Kelley K, Rosenberg L, Mitchell AA. Use of herbal/natural supplements according to racial/ethnic group. *J Altern Complement Med*. 2006;**12**(6):555-561.

**Rationale for exclusion at full-text screening**

Wrong concept.

**Record #341 excluded at full-text screening**

Cook SJ. Use of traditional Mi'kmaq medicine among patients at a First Nations community health centre. *Can J Rural Med*. 2005;**10**(2):95-99.

**Rationale for exclusion at full-text screening**

Wrong concept.

**Record #342 excluded at full-text screening**

Napoli M. Holistic health care for Native women: an integrated model. *Am J Public Health*. 2002;**92**(10):1573-1575.

**Rationale for exclusion at full-text screening**

Wrong concept.

**Record #343 excluded at full-text screening**

Lucks BC. Vitex agnus castus essential oil and menopausal balance: a research update [Complementary Therapies in Nursing and Midwifery 8 (2003) 148–154]. *Complement Ther Nurs Midwifery*. 2003;**9**(3):157-160.

**Rationale for exclusion at full-text screening**

Unclear population.

**Record #344 excluded at full-text screening**

Rogers SF. Walking in beauty: professor prioritizes Native American women's health. *MAMM*. 2001;**1A**(2):32-33.

**Rationale for exclusion at full-text screening**

Wrong concept.

**Record #345 excluded at full-text screening**

Tchernof A, Poehlman ET. Letters to the editor. *Menopause*. 2000;**7**(6):419-421.

**Rationale for exclusion at full-text screening**

Wrong record type.

**Record #346 excluded at full-text screening**

Légaré F, Stacey D, Dodin S, O'Connor A, Richer M, Griffiths F, Leblanc A, Rousseau JL, Tapp S. Women's decision making about the use of natural health products at menopause: a needs assessment and patient decision aid. *J Altern Complement Med*. 2007;**13**(7):741-749.

**Rationale for exclusion at full-text screening**

Unclear population.

**Record #347 excluded at full-text screening**

Theroux R. Women's decision making during the menopausal transition. *J Am Acad Nurse Pract*. 2010;**22**(11):612-621.

**Rationale for exclusion at full-text screening**

Wrong population.

**Record #348 excluded at full-text screening**

Dillaway H, Byrnes M, Miller S, Rehan S. Talking "Among Us": how women from different racial-ethnic groups define and discuss menopause. *Health Care Women Int*. 2008;**29**(7):766-781.

**Rationale for exclusion at full-text screening**

Wrong population.

**Record #349 excluded at full-text screening**

Gold EB, Sternfeld B, Kelsey JL, Brown C, Mouton C, Reame N, Salamone L, Stellato R. Relation of demographic and lifestyle factors to symptoms in a multi-racial/ethnic population of women 40–55 years of age. *Am J Epidemiol*. 2000;**152**(5):463-473.

**Rationale for exclusion at full-text screening**

Wrong population.

**Record #350 excluded at full-text screening**

Obermeyer CM, Reher D, Saliba M. Symptoms, menopause status, and country differences: a comparative analysis from DAMES. *Menopause*. 2007;**14**(4):788-797.

**Rationale for exclusion at full-text screening**

Unclear population.

**Record #351 excluded at full-text screening**

Parsons MA, Obermeyer CM. Women's midlife health across cultures: DAMES comparative analysis. *Menopause*. 2007;**14**(4):760-768.

**Rationale for exclusion at full-text screening**

Unclear population.

**Record #352 excluded at full-text screening**

Newton KM, Reed SD, Grothaus L, Ehrlich K, Guiltinan J, Ludman E, LaCroix AZ. The Herbal Alternatives for Menopause (HALT) study: background and study design. *Maturitas*. 2005 Oct;**52**(2):134-146.

**Rationale for exclusion at full-text screening**

Unclear population.

**Record #353 excluded at full-text screening**

Carroll DG. Nonhormonal therapies for hot flashes in menopause. *Am Fam Physician*. 2006 Feb;**73**(3):457-464.

**Rationale for exclusion at full-text screening**

Wrong population.

**Record #354 excluded at full-text screening**

Williamson J, White A, Hart A, Ernst E. Randomised controlled trial of reflexology for menopausal symptoms. BJOG. 2002;**109**(9):1050-1055.

**Rationale for exclusion at full-text screening**

Wrong population.

**Record #355 excluded at full-text screening**

Frei-Kleiner S, Schaffner W, Rahlfs VW, Bodmer C, Birkhäuser M. Cimicifuga racemosa dried ethanolic extract in menopausal disorders: a double-blind placebo-controlled clinical trial. *Maturitas*. 2005;**51**(4):397-404.

**Rationale for exclusion at full-text screening**

Unclear population.

**Record #356 excluded at full-text screening**

Osmers R, Friede M, Liske E, Schnitker J, Freudenstein J, Henneicke-von Zepelin HH. Efficacy and safety of isopropanolic black cohosh extract for climacteric symptoms. *Obstet Gynecol*. 2005;**105**(5 Pt 1):1074-1083.

**Rationale for exclusion at full-text screening**

Wrong population.

**Record #357 excluded at full-text screening**

Pockaj BA, Loprinzi CL, Sloan JA, Novotny PJ, Barton DL, Hagenmaier A, Zhang H, Lambert GH, Reeser KA, Wisbey JA. Pilot evaluation of black cohosh for the treatment of hot flashes in women. *Cancer Invest*. 2004;**22**(4):515-521.

**Rationale for exclusion at full-text screening**

Unclear population.

**Record #358 excluded at full-text screening**

Bromberger JT, Assmann SF, Avis NE, Schocken M, Kravitz HM, Cordal A. Persistent mood symptoms in a multiethnic community cohort of pre- and perimenopausal women. *Am J Epidemiol*. 2003 Aug;**158**(4):347-356.

**Rationale for exclusion at full-text screening**

Wrong population.

**Record #359 excluded at full-text screening**

Mackenzie ER, Taylor L, Bloom BS, Hufford DJ, Johnson JC. Ethnic minority use of complementary and alternative medicine (CAM): a national probability survey of CAM utilizers. *Altern Ther Health Med*. 2003 Jul-Aug;**9**(4):50-56.

**Rationale for exclusion at full-text screening**

Wrong concept.

**Record #360 excluded at full-text screening**

Avis NE, Colvin A, Bromberger JT, Hess R, Matthews KA, Ory M, Schocken M. Change in health-related quality of life over the menopausal transition in a multiethnic cohort of middle-aged women: Study of Women's Health Across the Nation. *Menopause*. 2009 Sep-Oct;**16**(5):860-869.

**Rationale for exclusion at full-text screening**

Wrong population.

**Record #361 excluded at full-text screening**

Cohen K. Native American medicine. *Altern Ther Health Med*. 1998 Nov;**4**(6):45-57.

**Rationale for exclusion at full-text screening**

Wrong concept.

**Record #362 excluded at full-text screening**

Goldfrank ES. Observations on sexuality among the Blood Indians of Alberta, Canada. Psychoanal Soc Sci. 1951;**3**:71-98.

**Rationale for exclusion at full-text screening**

Book.

**Record #363 excluded at full-text screening**

The North American Menopause Society. The 2023 nonhormone therapy position statement of The North American Menopause Society. *Menopause*. 2023;**30**(6):573-590.

**Rationale for exclusion at full-text screening**

Wrong population.

**Record #364 excluded at full-text screening**

Akapo AO, Schultz C, Coelho D, Muffly TM. American Indian and Alaskan Native access to obstetrics and gynecology subspecialists: findings from a national mystery caller study in the United States. *Cureus*. 2023;**15**(12):e51403.

**Rationale for exclusion at full-text screening**

Wrong concept.

**Record #365 excluded at full-text screening**

Ullman KE, Diem S, Forte ML, Ensrud K, Sowerby C, Zerzan N, et al. Complementary and alternative therapies for genitourinary syndrome of menopause: an evidence map. *Ann Intern Med*. 2024;**177**(10):1389-1399.

**Rationale for exclusion at full-text screening**

Wrong population.

**Record #366 excluded at full-text screening**

Kharumnuid G, Pal RS, Pal Y, Chaitanya MNL, Srivastava P. Discovering the untouched perspective of endangered North American herb Actaea racemosa. *Comb Chem High Throughput Screen*. 2024;**11**:11.

**Rationale for exclusion at full-text screening**

Wrong population.

**Record #367 excluded at full-text screening**

Srugo SA, Ricci C, Leason J, Jiang Y, Luo W, Nelson C. Disparities in primary and emergency health care among "off-reserve" Indigenous females compared with non-Indigenous females aged 15–55 years in Canada. *CMAJ*. 2023;**195**(33):E1097-E1111.

**Rationale for exclusion at full-text screening**

Wrong concept.

**Record #368 excluded at full-text screening**

Loy MH, Fatato T. Exploring the feasibility and acceptability of telehealth Qi Gong shared medical appointments: a novel approach to expand access. *Glob Adv Integr Med Health*. 2024;**13**:27536130241283789.

**Rationale for exclusion at full-text screening**

Unclear population.

**Record #369 excluded at full-text screening**

Brown K, Clarke M. First Nations women's health 2023. *Aust N Z J Obstet Gynaecol*. 2023;**63**(3):275-277.

**Rationale for exclusion at full-text screening**

Wrong population.

**Record #370 excluded at full-text screening**

Lewis JP, Kim SM, Asquith-Heinz Z, Withrow A. Generativity as a traditional way of life: Successful aging among Unangan Elders in the Aleutian Pribilof Islands. *J Cross Cult Gerontol*. 2024;**39**(2):107-123.

**Rationale for exclusion at full-text screening**

Wrong concept.

**Record #371 excluded at full-text screening**

Good MK, Davila H, Ball D, O'Neill S, Healy H, Mengeling MA. Health and health care use of American Indian/Alaska Native women veterans: a scoping review. *Womens Health Issues*. 2024;**34**(5):455-464.

**Rationale for exclusion at full-text screening**

Wrong concept.

**Record #372 excluded at full-text screening**

Liddell JL, Stiffarm AL. "I think [Western] healthcare fails them": Qualitative perspectives of state-recognized women tribal members on elders' healthcare access experiences. *Am Indian Alsk Native Ment Health Res*. 2023;**30**(2):70-96.

**Rationale for exclusion at full-text screening**

Wrong concept.

**Record #373 excluded at full-text screening**

Evans S, Van Niekerk L, Orellana L, O'Shea M, Druitt MI, Jones S, Corrigan S, Heggen C, Arashiro Z, Baber R, Satyen L, Skvarc D, Mikocka-Walus A. The need for biopsychosocial menopause care: a narrative review. *Menopause*. 2024;**31**(12):1090-1096.

**Rationale for exclusion at full-text screening**

Wrong population.

**Record #374 excluded at full-text screening**

Fatima S, Verma M, Ansari IA. Phytochemistry and ethnopharmacological studies of genus Cimicifuga: a systematic and comprehensive review. *Fitoterapia*. 2024;**172**:105767.

**Rationale for exclusion at full-text screening**

Wrong population.

**Record #375 excluded at full-text screening**

Jacklin K, Pitawanakwat K, Blind M, Ketcher D, Jones L, Piraino E, Bretzlaff M. PIECES of my RELATIONSHIPS: the cultural adaptation of a biographical assessment tool for Indigenous older adults in Canada. *Gerontologist*. 2024;**64**(7):1-13.

**Rationale for exclusion at full-text screening**

Wrong concept.

**Record #376 excluded at full-text screening**

Kingsberg S, Banks V, Caetano C, Janssenswillen C, Moeller C, Schoof N, Harvey M, Scott M, Nappi RE. Real-world evaluation of treatment utilization by women experiencing vasomotor symptoms associated with menopause in the United States and Europe: findings from the REALISE study. *Maturitas*. 2024;**189**:108096.

**Rationale for exclusion at full-text screening**

Wrong population.

**Record #377 excluded at full-text screening**

Jaramillo ET, Haozous EA, Brechtel H, Willging CE. Trusted communicators: the role of navigation support in improving health and health care access for American Indian elders. *J Health Care Poor Underserved*. 2024;**35**(1):246-263.

**Rationale for exclusion at full-text screening**

Wrong concept.

**Record #378 excluded at full-text screening**

Watts A, Thangwaritorn P, Medina V, Khanna P. Bridging menopausal history and Alzheimer disease: a pilot study targeting sex-based disparities in Alzheimer's risk. *J Prev Alzheimers Dis*. 2019;**1**(6):50-55.

**Rationale for exclusion at full-text screening**

Wrong population.

**Record #379 excluded at full-text screening**

Smith M, Dirks L, Kramer C, Trout L, Griffin P, Harrington EK. Community-based formative research to advance sexual and reproductive health equity in Iñupiaq Alaska [abstract]. *Contraception*. 2023;**127**:110146.

**Rationale for exclusion at full-text screening**

Wrong concept.

**Record #380 excluded at full-text screening**

Ghatti SS, Garaga M, Chatterjee A, Thimmannagari S, Cherupalli SR, Hebbani NS, Krishnamani M, Soni G. A comprehensive review on Sophora japonica: ethnomedicinal, phytochemistry, and pharmacological aspects. *Combin Chem High Throughput Screen*. Forthcoming 2024:1-10.

**Rationale for exclusion at full-text screening**

Wrong population.

**Record #381 excluded at full-text screening**

Goins RT, Adamsen C, Bendixen B, Woodrich-Fernando M, Joe JR, Besse R, Zuke K. Experiences with and access to evidence-based health promotion programs for older American Indian, Alaska Native, and Native Hawaiian peoples. *Health Promot Pract*. Forthcoming 2023:15248399231201552.

**Rationale for exclusion at full-text screening**

Wrong concept.

**Record #382 excluded at full-text screening**

Akhtari E, Ram M, Zaidi SMA, Marques AM, Rahimi R, Bahramsoltani R. Fenugreek (Trigonella foenum-graecum L.) in women's health: a review of clinical evidence and traditional use. *J Herb Med*. 2024;**43**:100816.

**Rationale for exclusion at full-text screening**

Wrong population.

**Record #383 excluded at full-text screening**

Dion MHG, Fraser SL, Cookie-Brown L. Inuit wellness: a better understanding of the principles that guide actions and an overview of practices. *Transcult Psychiatry*. 2024;**61**(3):417-428.

**Rationale for exclusion at full-text screening**

Wrong concept.

**Record #384 excluded at full-text screening**

Daniel TD, Hutchinson-Colas J, Bachmann G. Menopause management: need for global perspective [abstract]. *Menopause*. 2021;**28**(12):1458.

**Rationale for exclusion at full-text screening**

Wrong population.

**Record #385 excluded at full-text screening**

Taylor-Swanson L, Stoddard K, Fritz J, Anderson B, Cortez M, Conboy L, et al. Midlife women’s menopausal transition symptom experience and access to medical and integrative health care: informing the development of MENOGAP. *Glob Adv Integr Med Health*. 2024;**13**:1-13.

**Rationale for exclusion at full-text screening**

Unstratified results (population).

**Record #386 excluded at full-text screening**

Dweck A, Patel P, Sylla S, Bernsley D, Komorowski J. A national survey study examining women's experiences with menopause [abstract]. Menopause. 2021;**28**(12):1459.

**Rationale for exclusion at full-text screening**

Unclear population.

**Record #387 excluded at full-text screening**

Cheung AM, Chaudhry R, Kapral M, Jackevicius C, Robinson G. Perimenopausal and postmenopausal health. *BMC Womens Health*. 2004;**4**(Suppl 1):S23.

**Rationale for exclusion at full-text screening**

Wrong population.

**Record #388 excluded at full-text screening**

Todorova L, Bonassi R, Guerrero Carreño FJ, Hirschberg AL, Yuksel N, Rea C, Scrine L, Kim JS. Prevalence and impact of vasomotor symptoms due to menopause among women in Brazil, Canada, Mexico, and Nordic Europe: a cross-sectional survey. *Menopause*. 2023;**30**(12):1179-1189.

**Rationale for exclusion at full-text screening**

Unclear population.

**Record #389 excluded at full-text screening**

Depree B, Houghton K, Shiozawa A, Esterberg E, King D, Kim J, Mancuso S. Real-world treatment and resource utilization for menopausal symptoms in the United States [abstract]. *Menopause*. 2021;**28**(12):1447.

**Rationale for exclusion at full-text screening**

Duplicate record.

**Record #390 excluded at full-text screening**

Shepherd JA, Shiozawa A, Schild AL, Singh D, Mancuso SA. Survey of patient experience and management of vasomotor symptoms due to menopause from the Patients Like Me community. *Menopause*. 2024;**31**(11):979-986.

**Rationale for exclusion at full-text screening**

Wrong population.

**Record #391 excluded at full-text screening**

Wheat J, Stautz H, Watson A, Carty J, Dengler K, Shvartsman K, Vaccaro C. What is normal? Using objective measurements to describe typical vulvar anatomy [abstract]. *Am J Obstet Gynecol*. 2024;**230**(4):S1243-S1244.

**Rationale for exclusion at full-text screening**

Wrong concept.

**Record #392 excluded at full-text screening**

Mikdachi H, Allen P, Dunsmoor-Su R. What women tell us about the menopause transition [abstract]. *J Neurosurg Anesthesiol*. 2023;**30**(12):52.

**Rationale for exclusion at full-text screening**

Unstratified results (population).

**Record #393 excluded at full-text screening**

Babyn K, Quintanilha M, Ross S, Makowsky M, Kiang T, Yuksel N. Women's perceptions and experiences with cannabis use in menopause: a qualitative study. *Menopause*. 2024;**31**(9):781-788.

**Rationale for exclusion at full-text screening**

Unclear population.

**Record #394 excluded at full-text screening**

Ghogare AS, Talhan TS, Madavi PB, Joshi AC, Telgote SA, Ambad RS. Beyond the antidepressant action, paroxetine in managing the hot flashes in women with menopause: a systematic review. *Glob J Med Pharm Biomed Update*. 2023;**18**(31):1-14.

**Rationale for exclusion at full-text screening**

Wrong population.

**Record #395 excluded at full-text screening**

**Campbell AR, Hill PL, Nicholson V, Lambert S, Cote HCF, Edmonds GW, Pick N, Murray MCM.** Exploring sense of purpose and conscientiousness as correlates to health and well-being with Indigenous and low socioeconomic communities on Coast Salish Territories, Vancouver, Canada. Can J Behav Sci. 2024;**56**(3):240-252.

**Rationale for exclusion at full-text screening**

Wrong concept.

**Record #396 excluded at full-text screening**

**Kelley M, Lowe J, Xie B, Shiroma K.** Native American elders' lived experiences during the COVID-19 pandemic. Int J Hum Caring. 2023;**27**(4):219-224.

**Rationale for exclusion at full-text screening**

Wrong concept.

**Record #397 excluded at full-text screening**

**Pandey M, Clark M, Beresh E, Nilson S, Kay C, Campbell T, Nicolay S, Skinner S.** Wellness Wheel Clinics: a First Nation community-partnered care model improving healthcare access, from healthcare providers' perspectives. Health Promot Int. 2023;**38**(4):1-12.

**Rationale for exclusion at full-text screening**

Wrong concept.

**Record #398 excluded at full-text screening**

**McGuire-Adams T.** Anishinaabeg Elders' land-based knowledge: enacting bagijigan for health and well-being. Int Indig Policy J. 2023 Aug;**14**(2):1-14.

**Rationale for exclusion at full-text screening**

Wrong concept.

**Record #399 excluded at full-text screening**

**Hurley ES.** The next seven generations: at the heart of care alongside Mi'kmaw women. AlterNative. 2024 Mar;**20**(1):189-193.

**Rationale for exclusion at full-text screening**

Wrong concept.

**Record #400 excluded at full-text screening**

**Oppliger K, Blair S, Price R, Nahanee M, Nahanee D, Duncan R, Lamont E, Beverly A, Dawson A, Conklin AI.** Promoting Slhanay Skwalwen (Indigenous women's heart health): findings from sharing circles with Nation. J Nutr Educ Behav. 2024 Aug;**56**(8):509-520.

**Rationale for exclusion at full-text screening**

Wrong concept.

**Record #401 excluded at full-text screening**

**Tiersma K, Keys T.** Promoting women's health screening on the Blackfeet Reservation [abstract]. J Investig Med. 2024 Jan;**72**(1):319-320.

**Rationale for exclusion at full-text screening**

Wrong concept.

**Record #402 excluded at full-text screening**

**Willis SC.** Art + culture + elders. In: Basting AD, editor. Art education and creative aging: older adults as learners, makers, and teachers of art. New York, NY: Teachers College Press; 2024. p. 171-179.

**Rationale for exclusion at full-text screening**

Book.

**Record #403 excluded at full-text screening**

**Kim SM, Lewis JP.** Protective factors in the context of successful aging in urban-dwelling Alaska Native elders. J Cross Cult Gerontol. 2024;**39**(1):53-71.

**Rationale for exclusion at full-text screening**

Wrong concept.
